# Comparative Effectiveness of Second-line Antihyperglycemic Agents for Cardiovascular Outcomes: A Large-scale, Multinational, Federated Analysis of the LEGEND-T2DM Study

**DOI:** 10.1101/2024.02.05.24302354

**Authors:** Rohan Khera, Arya Aminorroaya, Lovedeep Singh Dhingra, Phyllis M Thangaraj, Aline Pedroso Camargos, Fan Bu, Xiyu Ding, Akihiko Nishimura, Tara V Anand, Faaizah Arshad, Clair Blacketer, Yi Chai, Shounak Chattopadhyay, Michael Cook, David A Dorr, Talita Duarte-Salles, Scott L DuVall, Thomas Falconer, Tina E French, Elizabeth E Hanchrow, Guneet Kaur, Wallis CY Lau, Jing Li, Kelly Li, Yuntian Liu, Yuan Lu, Kenneth KC Man, Michael E Matheny, Nestoras Mathioudakis, Jody-Ann McLeggon, Michael F McLemore, Evan Minty, Daniel R Morales, Paul Nagy, Anna Ostropolets, Andrea Pistillo, Thanh-Phuc Phan, Nicole Pratt, Carlen Reyes, Lauren Richter, Joseph Ross, Elise Ruan, Sarah L Seager, Katherine R Simon, Benjamin Viernes, Jianxiao Yang, Can Yin, Seng Chan You, Jin J Zhou, Patrick B Ryan, Martijn J Schuemie, Harlan M Krumholz, George Hripcsak, Marc A Suchard

**Author notes:** **Correspondence to:** Marc A Suchard, MD, PhD Departments of Biomathematics, Biostatistics and Human Genetics 6558 Gonda Building, 695 Charles E. Young Drive, South Los Angeles, CA 90095-1766; @suchard_group.

## Abstract

**Background:** SGLT2 inhibitors (SGLT2is) and GLP-1 receptor agonists (GLP1-RAs) reduce major adverse cardiovascular events (MACE) in patients with type 2 diabetes mellitus (T2DM). However, their effectiveness relative to each other and other second-line antihyperglycemic agents is unknown, without any major ongoing head-to-head trials.

**Methods:** Across the LEGEND-T2DM network, we included ten federated international data sources, spanning 1992-2021. We identified 1,492,855 patients with T2DM and established cardiovascular disease (CVD) on metformin monotherapy who initiated one of four second-line agents (SGLT2is, GLP1-RAs, dipeptidyl peptidase 4 inhibitor [DPP4is], sulfonylureas [SUs]). We used large-scale propensity score models to conduct an active comparator, target trial emulation for pairwise comparisons. After evaluating empirical equipoise and population generalizability, we fit on-treatment Cox proportional hazard models for 3-point MACE (myocardial infarction, stroke, death) and 4-point MACE (3-point MACE + heart failure hospitalization) risk, and combined hazard ratio (HR) estimates in a random-effects meta-analysis.

**Findings:** Across cohorts, 16·4%, 8·3%, 27·7%, and 47·6% of individuals with T2DM initiated SGLT2is, GLP1-RAs, DPP4is, and SUs, respectively. Over 5·2 million patient-years of follow-up and 489 million patient-days of time at-risk, there were 25,982 3-point MACE and 41,447 4-point MACE events. SGLT2is and GLP1-RAs were associated with a lower risk for 3-point MACE compared with DPP4is (HR 0·89 [95% CI, 0·79-1·00] and 0·83 [0·70-0·98]), and SUs (HR 0·76 [0·65-0·89] and 0·71 [0·59-0·86]). DPP4is were associated with a lower 3-point MACE risk versus SUs (HR 0·87 [0·79-0·95]). The pattern was consistent for 4-point MACE for the comparisons above. There were no significant differences between SGLT2is and GLP1-RAs for 3-point or 4-point MACE (HR 1·06 [0·96-1·17] and 1·05 [0·97-1·13]).

**Interpretation:** In patients with T2DM and established CVD, we found comparable cardiovascular risk reduction with SGLT2is and GLP1-RAs, with both agents more effective than DPP4is, which in turn were more effective than SUs. These findings suggest that the use of GLP1-RAs and SGLT2is should be prioritized as second-line agents in those with established CVD.

**Funding:** National Institutes of Health, United States Department of Veterans Affairs

## RESEARCH IN CONTEXT

### Evidence before this study

Sodium-glucose co-transporter-2 inhibitors (SGLT2is) and glucagon-like peptide-1 receptor agonists (GLP1-RAs) exert cardiovascular benefits beyond blood glucose control, transforming the landscape of treating type 2 diabetes mellitus (T2DM) over the past decade. Large randomized clinical trials have substantiated the cardiovascular benefits of SGLT2is and GLP1-RAs in decreasing major adverse cardiovascular events. However, trials of SGLT2is and GLP1-RAs have not been designed as head-to-head comparisons with each other or older antihyperglycemic agents such as dipeptidyl peptidase-4 inhibitors (DPP4is) and sulfonylureas (SUs). Therefore, the comparative cardiovascular effectiveness of these antihyperglycemic agents is not established. Recent work also shows a substantial proportion of patients with T2DM are often initiated on DPP4is and SUs in the absence of information on their cardiovascular effectiveness. These knowledge gaps in evidence challenge the development of treatment recommendations based on cardiovascular risk profiles in T2DM.

### Added value of this study

The Large-scale Evidence Generation and Evaluation across a Network of Databases for Type 2 Diabetes Mellitus (LEGEND-T2DM) initiative is a large-scale, systematic, federated evaluation of individuals with T2DM across multiple international observational data sources, where we leveraged robust state-of-the-art methodological and analytic strategies to minimize residual confounding, publication bias, and p-hacking. In this largest study of 1·5 million patients with T2DM and established cardiovascular disease (CVD) across ten cohorts from four countries initiating a second-line antihyperglycemic agent — SGLT2is, GLP1-RAs, DPP4is, or SUs — after metformin monotherapy, SGLT2is and GLP1-RAs were associated with an 11% and 17% lower risk of cardiovascular events compared with DPP4is, respectively. All three agents were associated with lower cardiovascular events than SUs, with 24% and 28% lower risk with SGLT2is and GLP1-RAs, and 12% lower risk with DPP4is. These patterns were consistent across both ischemic events, such as MI and stroke, as well as for heart failure.

### Implications of all the available evidence

Our study defines strong empirical evidence in support of contemporary clinical practice guidelines that suggest SGLT2is and GLP1-RAs are used for those with T2DM and CVD. Our observations also provide support for exclusive second-line use of SGLT2is and GLP1-RAs among those with CVD.

## INTRODUCTION

Over the past decade, the therapeutic options for type 2 diabetes mellitus (T2DM) have undergone a significant transformation.^1,2^ Sodium-glucose co-transporter-2 inhibitors (SGLT2is) and glucagon-like peptide-1 receptor agonists (GLP1-RAs) have expanded the role of antihyperglycemic agents from managing high blood glucose to addressing the elevated cardiovascular risk in patients with T2DM.^3–6^ In several large randomized clinical trials (RCTs), SGLT2is and GLP1-RAs reduced major adverse cardiovascular events (MACE), such as myocardial infarction (MI), hospitalization for heart failure, and cardiovascular mortality.^7–14^ While older antihyperglycemic agents, like sulfonylureas (SUs), have not undergone similarly comprehensive trials to evaluate their cardiovascular efficacy or safety, some studies suggested their neutral effect on cardiovascular outcomes and risk of hypoglycemia.^15–17^ Furthermore, direct comparisons of SGLT2is and GLP1-RAs with dipeptidyl peptidase-4 inhibitors (DPP4is), which are antihyperglycemic agents with neutral effects on MACE, have not been conducted and no major trials are in progress. Nevertheless, DPP4is and SUs continue to be used in clinical practice and are recommended as second-line T2DM agents in national clinical practice guidelines.^18–22^

Despite the availability of evidence from SGLT2i and GLP1-RA trials, several gaps in evidence challenge the development of treatment recommendations in T2DM.^18,23,24^ Specifically, trials of SGLT2is and GLP1-RAs were not designed as head-to-head comparisons with older agents but rather as additive treatments on top of commonly used T2DM agents.^8,9,25^ As a result, the relative cardiovascular efficacy of newer versus older agents is not known. Moreover, the comparative cardiovascular efficacy of SGLT2is and GLP1-RAs have not been evaluated. Thus far, comparative effectiveness assessments have been based on indirect estimates from clinical trials or comparative effectiveness drawn from a limited number of data sources.^26–28^ The findings from observational studies using single data sources are challenged by our observations that the uptake of these agents, and therefore, the selective pressures on the use of these agents, varies considerably across sites,^21^ with the potential to introduce bias in the evaluation of comparative effectiveness. These evidence gaps pose a significant challenge in designing treatment algorithms that rely on the comparative effectiveness and safety of drugs.^24^ As a result, there is a large variation in clinical practice guidelines and clinical practice with regard to these medications, with many patients initiated on the newer therapies and many others treated with older regimens.^21,29^

To address this, we conducted the Large-scale Evidence Generation and Evaluation across a Network of Databases for Type 2 Diabetes Mellitus (LEGEND-T2DM) initiative,^30^ a large-scale, systematic, federated evaluation of 4·7 million patients with T2DM across multiple international observational data sources, where we leveraged robust state-of-the-art methodological and analytic strategies to minimize residual confounding, publication bias, and p-hacking. Here, we compared the cardiovascular effectiveness of four second-line antihyperglycemic agents — SGLT2is, GLP1-RAs, DPP4is, or SUs — when initiated on the background of metformin therapy in T2DM.

## METHODS

### Data Sources

In this study, we included ten real-world data sources from the LEGEND-T2DM network, including six administrative claims and four EHR databases across four countries during 1992-2021 (**figure 1**). These represent data from six national-level and four health-system datasets from the US, as well as data sources from Germany, Spain, and the United Kingdom. The vast majority of patient records span from the mid-2000s to today, covering two decades of T2DM treatment as well as the introduction of many second-line antihyperglycemic agents. All LEGEND-T2DM data sources were previously standardized to the Observational Health Data Sciences and Informatics (OHDSI)’s Observational Medical Outcomes Partnership (OMOP) common data model (CDM) version 5,^31^ which mapped international coding systems into standard vocabulary concepts. The use of the OMOP CDM allows a federated analysis of data, without patient-level data sharing, using consistent cohort definitions and study design. All data partners received institutional approval or exemption for their participation. Details of data sources are presented in **appendix pp 17-19**.

**Figure 1:**
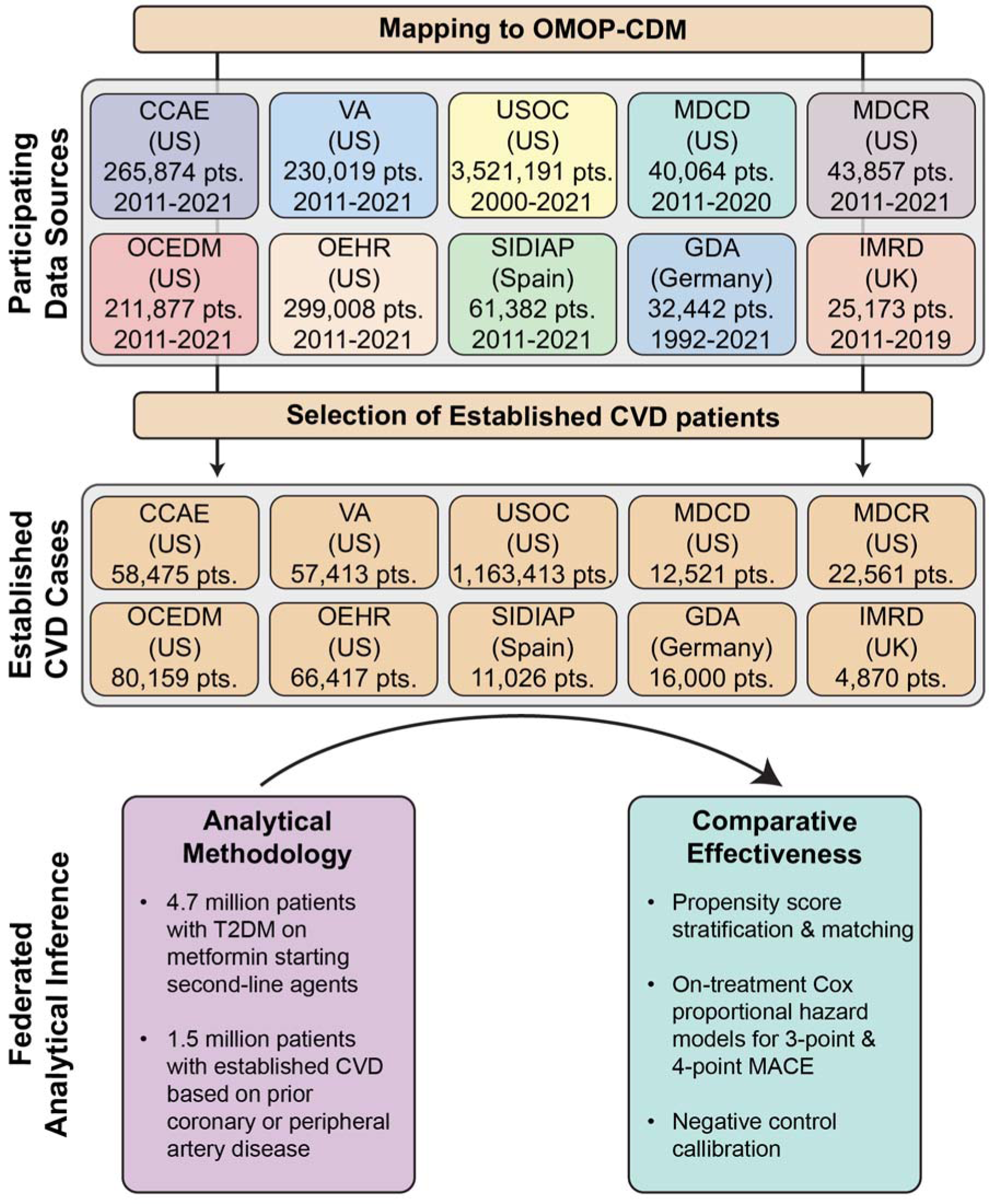
Study Design and Analytical Methodology to Evaluate the Comparative Effectiveness of SGLT2is, GLP1-RAs, DPP4is, and SUs for Cardiovascular Outcomes. **Abbreviations:** CCAE, IBM MarketScan® Commercial Claims and Encounters Data; CVD, cardiovascular disease; GDA, Germany Disease Analyser; IMRD, UK-IQVIA Medical Research Data; MACE, major adverse cardiovascular events; MDCD, IBM Health MarketScan® Multi-State Medicaid Database; MDCR, IBM Health MarketScan® Medicare Supplemental and Coordination of Benefits Database; OCEDM, Optum© Clinformatics Extended Data Mart - Date of Death; OEHR, Optum© de-identified Electronic Health Record Dataset; OMOP-CDM, Observational Medical Outcomes Partnership-common data model; SIDIAP, Information System for Research in Primary Care; T2DM, type 2 diabetes mellitus; USOC, United States Open Claims; VA, Department of Veterans Affairs Healthcare System.

### Study Design

We identified patients with T2DM and established cardiovascular disease (CVD) on metformin monotherapy who initiated on any of the drug ingredients within one of the SGLT2i, GLP1-RA, DPP4i, or SU drug classes (**appendix pp 2-10**, **20**).^21^ Each exposure cohort thus consists of new users of each drug class. We require patients with T2DM and CVD to have at least one year of prior observation in the database, with at least 3 months of prior metformin use before initiating a second-line agent, and no prior exposure to a comparator second-line or other antihyperglycemic agent or more than 30 days of insulin exposure (**appendix pp 2-10**). To ensure statistical power, we executed analyses for comparisons and data sources with at least 1,000 patients in each arm. To evaluate the comparative effectiveness of antihyperglycemic agents, we constructed target–comparator–database combinations, where we compared one antihyperglycemic agent (target) with another agent (comparator) across data sources. For this, we employed a new-user cohort design for target and comparator agents within each data source in order to emulate the hypothetical target trial.^32,33^ Methodological principles in the study design have been carefully constructed based on the evidence by experts and were leveraged previously to minimize bias and improve reproducibility.^30,34^

### Study Outcomes

Across all data sources and pairwise exposure cohorts, we assess the relative risks of two primary and four secondary cardiovascular outcomes. The two primary outcomes of interest are (1) 3-point MACE, including acute MI, stroke, and sudden cardiac death, and (2) 4-point MACE, which additionally includes hospitalization for heart failure. Secondary outcomes of interest include the four individual MACE components. We construct outcome cohorts based on previously developed phenotypes validated and tested in prior work (**appendix pp 11-14**).^34–39^

For each outcome cohort, we included patients with no events prior to treatment initiation and defined continuous drug exposure as consecutive drug prescriptions with less than 30-day prescription gaps. We considered an on-treatment time-at-risk (TAR) definition that follows a patient from treatment initiation to treatment discontinuation, which captures direct treatment effects while allowing for escalation with additional T2DM agents.

### Statistical Analysis

We employed a systematic federated analytic framework to address residual confounding, publication bias, and p-hacking.^30,40^ This framework uses data-driven, large-scale propensity score (PS) adjustment for measured confounding,^41^ a large set of negative control outcome experiments to address unmeasured confounding and systematic bias,^42–44^ study diagnostics to ensure validity and generalizability,^44,45^ and a principled meta-analysis approach to aggregate evidence across data sources. We used standardized vocabularies to construct consistent computable definitions of all study cohorts, covariates, and outcomes. We provide full disclosure of all hypotheses investigated and pre-specify and report all analytical procedures in the published protocol of LEGEND-T2DM.^30^ To promote open science and avoid publication bias, we have disseminated all results in a publicly available R ShinyApp (https://data.ohdsi.org/LegendT2dmClassEvidenceExplorer/), and all analytic code is publicly available on GitHub (https://github.com/ohdsi-studies/LegendT2dm).

Within each data source, we estimated the relative risks of all six outcomes between each pair of new-user cohorts, taking one exposure cohort as the target and the other as a comparator. For each pairwise comparison and each data source, we adjusted for measured confounding and improved balance between cohorts through both matching and stratifying on PS.^46^ We estimated the PS by a data-driven approach that adjusts for a broad range of predefined baseline patient characteristics through regularized regression.^41^ These characteristics included demographics, comorbidities, concomitant medication use, and healthcare utilization in the period before the initiation of the second-line antihyperglycemic agent. The choice of PS stratification vs. matching was based on the approach that achieves a standardized mean difference (SMD) <0·15 across all covariates.^47^ When both stratification and matching provide sufficient balance, we preferred stratification over matching and thus reported results based on stratification when available, as the former improves patient inclusion and, therefore, generalizability. We then used Cox proportional hazards models to estimate hazard ratios (HRs) of each outcome for each comparison, conditional on PS stratification or variable-ratio patient matching.

We also sought to address residual bias, which may persist in observational studies even after PS adjustment that controls for measured confounding.^42,43^ For this, we conducted negative control (falsification) outcome experiments for each comparison and outcome, where the null hypothesis of no differential effect (i.e., HR = 1) is believed to be true for each outcome. We selected 100 negative controls through a data-driven algorithm that identifies OMOP condition concept occurrences with similar prevalence to the outcomes of interest that lack evidence of association with exposures in published literature, drug–product labels, and spontaneous reports, which we then confirmed by expert review (**appendix p 15**).^48^ We list these negative controls in **appendix pp 21-22**. From these negative control experiments, we learn an empirical null distribution that informs residual study bias, i.e., a deviation from the empirical null across all outcomes represents a quantitative surrogate for the residual bias. We calibrate each original HR estimate to compute a calibrated HR estimate and 95% confidence interval (CI).^42^ We declare an HR as significantly different from the null if the calibrated p-value is <0·05 without considering multiple testing corrections.

We assessed, while blinded to the results, study diagnostics to ensure reliability and generalizability for all comparisons and only report estimates that pass the diagnostics.^44,45^ These study diagnostics included (1) minimum detectable risk ratio (MDRR) as a metric for statistical power, (2) preference score distributions between the target and comparator cohorts to evaluate empirical equipoise and population generalizability, and (3) cohort balance before and after propensity score (PS) adjustment, defined by the absolute SMDs on extensive patient characteristics. A study passed diagnostics if MDRR was less than 4, and >25% of patients had a preference score between 0·3 and 0·7 on both arms and maximum SMD <0·15 after PS adjustment. Additional diagnostics for visual examination included (4) calibration plots on negative control outcomes to examine residual bias, and (5) Kaplan-Meier plots to check proportionality assumptions for the Cox models.

We reported all HR estimates, their 95% CIs, and p-values post-calibration for studies that passed diagnostics. To aggregate evidence across non-overlapping data sources, we combined all calibrated HR estimates for each comparison using a random effects meta-analysis approach.^49^

## RESULTS

### Cohort Characteristics

Across ten federated longitudinal data sources from four countries, we identified 4,730,887 patients with T2DM. This included 1,492,855 patients with T2DM and established CVD on metformin monotherapy who initiated one of the four second-line antihyperglycemic agents and had no prior use of any other antihyperglycemic agents (**figure 1**, **appendix p 17**). Among these patients, 244,694 (16·4%) initiated SGLT2is, 123,991 (8·3%) GLP1-RAs, 413,236 (27·7%) DPP4is, and 710,934 (47·6%) SUs (**table 1**). US Open Claims (USOC) with 1,163,413 patients with T2DM and established CVD, followed by Optum Clinformatics Extended Data Mart - Date of Death (OCEDM) (N=80,159) and Optum de-identified Electronic Health Record Dataset (OEHR) (N=66,417) contributed the most patients to the study population. Median on-treatment time-at-risk for patients varied by drug class and database between 2·0 and 23·2 months. At least 25% of the patients were exposed to their first drug class for more than 12 months across a majority of databases (**table 1**).

**Table 1:** Population Size and Follow-up Time for Initiators of SGLT2is, GLP1-RAs, DPP4is, and SUs Across Databases. **Abbreviations:** CCAE, IBM MarketScan® Commercial Claims and Encounters Data; DPP4i, dipeptidyl peptidase 4 inhibitor; GDA, Germany Disease Analyzer; GLP1-RA, glucagon-like peptide-1 receptor agonist; IMRD, UK-IQVIA Medical Research Data; MDCD, IBM Health MarketScan® Multi-State Medicaid Database; IQR, interquartile range; MDCR, IBM Health MarketScan® Medicare Supplemental and Coordination of Benefits Database; OCEDM, Optum© Clinformatics Extended Data Mart - Date of Death; OEHR, Optum© de-identified Electronic Health Record Dataset; SGLT2i, sodium-glucose co-transporter-2 inhibitor; SIDIAP, Information System for Research in Primary Care; SU, Sulfonylurea; USOC, United States Open Claims; VA, Department of Veterans Affairs Healthcare System.

| Drug/Data Source | Patients, No. | On-treatment Follow-up Time (days),<br>Median [IQR] |
| --- | --- | --- |
| **SGLT2i** |  |  |
| CCAE | 10,135 | 161 [62 - 537] |
| GDA | 4,337 | 181 [97 - 537] |
| MDCD | 1,314 | 114 [30 - 383] |
| MDCR | 2,013 | 112 [34 - 541] |
| OCEDM | 11,597 | 118 [55 - 423] |
| OEHR | 8,996 | 71 [29 - 181] |
| SIDIAP | 2,281 | 322 [103 - 895] |
| USOC | 196,710 | 143 [61 - 525] |
| VA | 7,311 | 135 [71 - 443] |
| **Total** | **244,694** |  |
| **GLP1-RA** |  |  |
| CCAE | 7,446 | 116 [45 - 441] |
| MDCR | 1,120 | 94 [29 - 443] |
| OCEDM | 6,546 | 98 [36 - 368] |
| OEHR | 4,525 | 60 [29 - 173] |
| USOC | 104,354 | 112 [52 - 432] |
| **Total** | **123,991** |  |
| **DPP4i** |  |  |
| CCAE | 22,801 | 159 [63 - 597] |
| GDA | 10,428 | 221 [97 - 741] |
| IMRD | 4,024 | 354 [122 - 1,174] |
| MDCD | 4,449 | 142 [56 - 555] |
| MDCR | 10,541 | 187 [81 - 713] |
| OCEDM | 23,918 | 150 [59 - 569] |
| OEHR | 18,145 | 95 [89 - 312] |
| SIDIAP | 8,126 | 695 [204 - 1,916] |
| USOC | 299,835 | 157 [59 - 657] |
| VA | 10,969 | 220 [90 - 707] |
| **Total** | **413,236** |  |
| **SU** |  |  |
| CCAE | 18,093 | 148 [59 - 559] |
| GDA | 1,235 | 216 [119 - 684] |
| IMRD | 846 | 350 [106 - 1312] |
| MDCD | 6,758 | 124 [33 - 503] |
| MDCR | 8,887 | 159 [59 - 632] |
| OCEDM | 38,098 | 177 [85 - 652] |
| OEHR | 34,751 | 110 [89 - 386] |
| SIDIAP | 619 | 600 [158 - 1,809] |
| USOC | 562,514 | 183 [89 - 752] |
| VA | 39,133 | 186 [90 - 689] |
| **Total** | **710,934** |  |

### Addressing Confounding Across Target-Comparator-Database Combinations

PS adjustment achieved pre-specified covariate balance in patient baseline characteristics for pairwise class comparisons across all databases (**figure 2**, **appendix pp 23-108**). The maximum SMD across target–comparator–database combinations consistently decreased after PS stratification (**figure 2**). For example, in comparison of patients initiating SGLT2is (target) with patients initiating DPP4is (comparator) in the IBM MarketScan Commercial Claims and Encounters (CCAE) database, before PS adjustment, patients initiating SGLT2is were more frequently men and had obesity and heart failure relative to patients initiating DPP4is. However, after PS adjustment, the SGLT2is or DPP4is populations were well balanced on all demographic and clinical patient characteristics (**appendix pp 33-34**).

**Figure 2:**
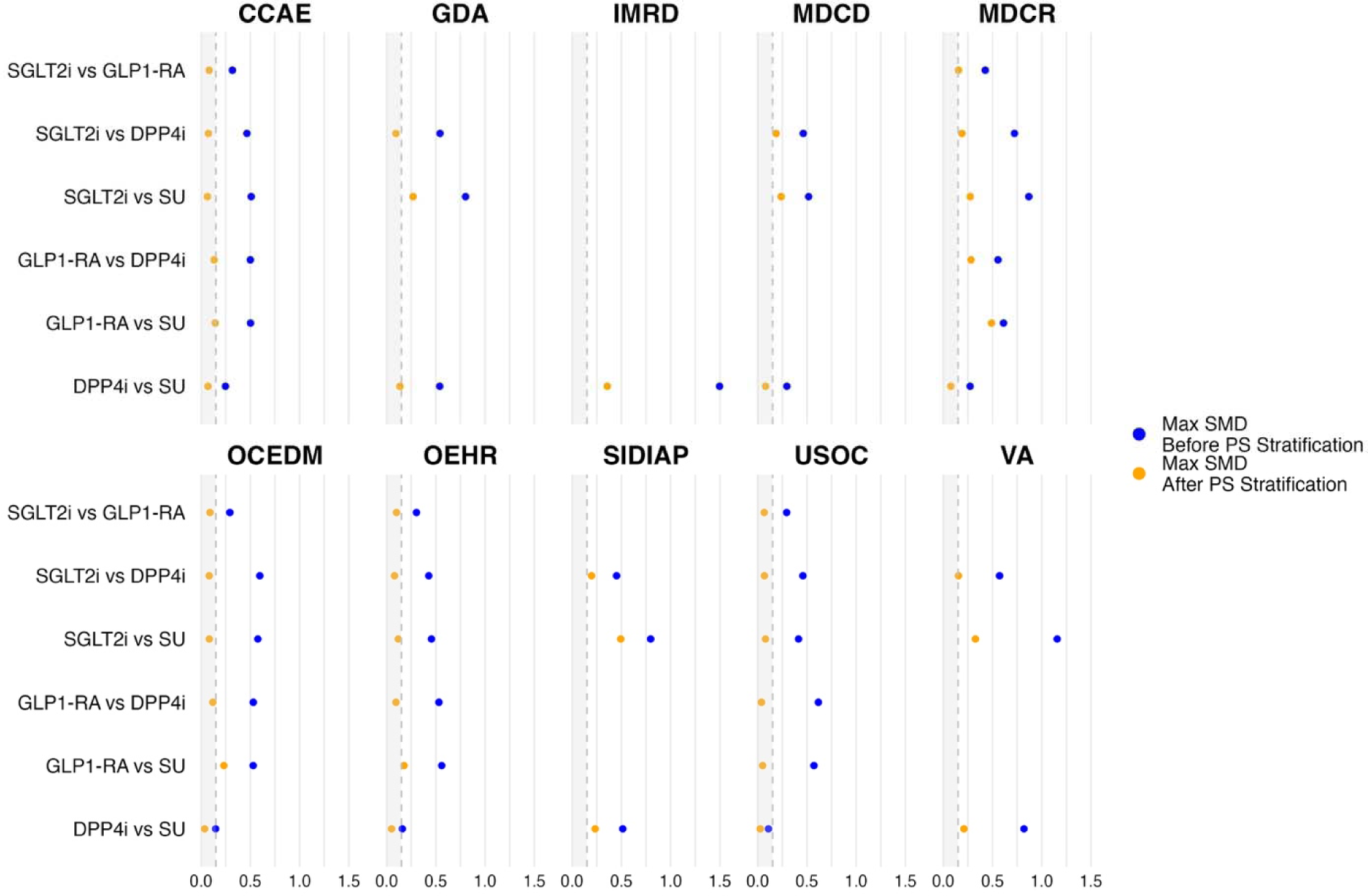
Maximum Standardized Mean Difference Before and After Propensity Score Stratification for All Covariates Across Target-Comparator-Database Combinations. **Abbreviations:** CCAE, IBM MarketScan® Commercial Claims and Encounters Data; DPP4i, dipeptidyl peptidase 4 inhibitor; GDA, Germany Disease Analyzer; GLP1-RA, glucagon-like peptide-1 receptor agonist; IMRD, UK-IQVIA Medical Research Data; MDCD, IBM Health MarketScan® Multi-State Medicaid Database; IQR, interquartile range; MDCR, IBM Health MarketScan® Medicare Supplemental and Coordination of Benefits Database; OCEDM, Optum© Clinformatics Extended Data Mart - Date of Death; OEHR, Optum© de-identified Electronic Health Record Dataset; PS, propensity score; SGLT2i, sodium-glucose co- transporter-2 inhibitor; SIDIAP, Information System for Research in Primary Care; SMD, standardized mean difference; SU, Sulfonylurea; USOC, United States Open Claims; VA, Department of Veterans Affairs Healthcare System.

### Empirical Equipoise Across Target-Comparator-Database Combinations

For most data sources, all executed class comparisons were in empirical equipoise (>25% of patients had preference scores between 0·3 and 0·7 on both arms) (**appendix pp 146-155**). GDA, SIDIAP, and VA databases showed less equipoise for comparisons involving SGLT2is and DPP4is. However, in general, PS adjustment achieved sufficient covariate balance in terms of preference score distribution to reduce concerns that measured the estimated effects of baseline confounding biases (**appendix pp 156-175**). Furthermore, the study had limited residual systematic error, where calibration of effect estimates using negative control outcomes resulted in an increase in the proportion of nominal 95% CIs that included 1 for control outcomes across a majority of comparisons and databases (**appendix pp 176-195**). For example, for SGLT2is vs DPP4is comparison in SIDIAP using PS matching, before calibration, the nominal CIs covered 75·0% of control estimates; after calibration, they covered 91·7% (**appendix p 192**).

### Comparative Effectiveness for Primary Endpoints

During 1,337,809 patient-years of follow-up, there were 25,982 3-point MACE and 41,447 4-point MACE events. SGLT2is and GLP1-RAs were associated with a lower hazard of 3-point MACE compared with DPP4is (HR 0·89 [95% CI, 0·79-1·00] and 0·83 [0·70-0·98], respectively), and with even lower HRs of 0·76 (95% CI, 0·65-0·89) and 0·72 (95% CI, 0·58-0·88) versus SUs, respectively. DPP4is were associated with a lower risk of 3-point MACE risk versus SUs (0·87 [0·79-0·95]). A consistent pattern was observed for 4-point MACE across the above-mentioned comparisons. In direct comparisons of SGLT2is and GLP1-RAs, there were no differences in either 3-point or 4-point MACE (1·06 [0·96-1·17], and 1·05 [0·97-1·13], respectively) (**appendix pp 109-121, figure 3**).

**Figure 3:**
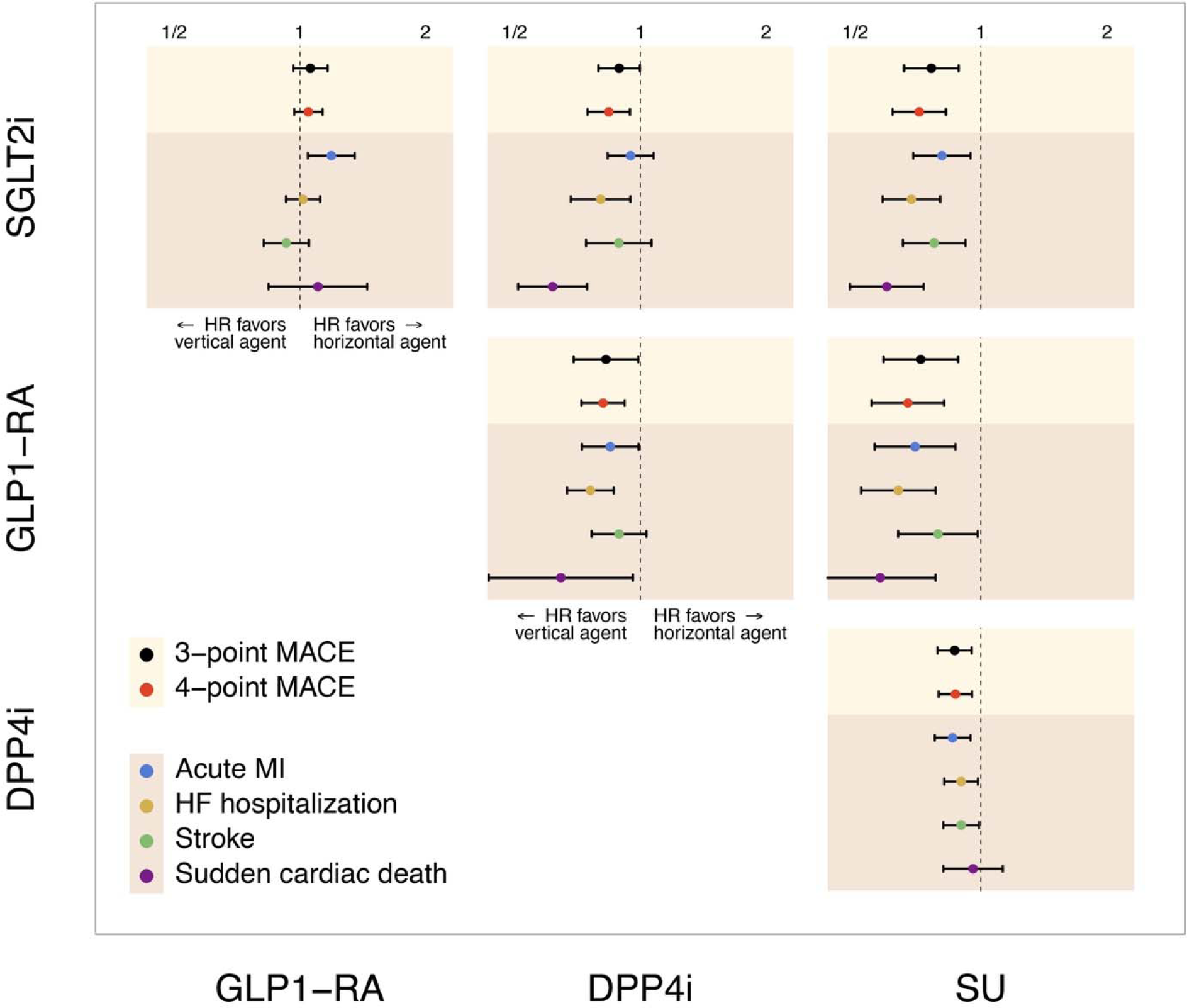
Meta-analytic Calibrated Hazard Ratio Estimates for Comparative Effectiveness of SGLT2is, GLP1-RAs, DPP4is, and SUs for Cardiovascular Outcomes. **Abbreviations:** DPP4i, dipeptidyl peptidase 4 inhibitor; GLP1-RA, glucagon-like peptide-1 receptor agonist; HF, heart failure; HR, hazard ratio; MACE, major adverse cardiovascular events; MI, myocardial infarction; SGLT2i, sodium-glucose co-transporter-2 inhibitor; SU, Sulfonylurea.

### Comparative Effectiveness for Secondary Endpoints

#### Acute MI

There were 13,536 episodes of acute MI across data sources during the follow-up. SGLT2is, GLP1-RAs, and DPP4is were associated with a lower hazard of acute MI compared with SUs (0·81 [0·69-0·95], 0·70 [0·56-0·87], and 0·86 [0·77-0·95], respectively). While GLP1-RAs were associated with a lower risk of acute MI compared with DPP4is (0·85 [0·73-0·99]), the hazard of acute MI was comparable for SGLT2is vs DPP4is (0·95 [0·83-1·08]). Compared with GLP1-RAs, SGLT2is were associated with a higher hazard of acute MI (1·19 [1·05-1·35]) (**appendix pp 109, 122-127, figure 3**).

#### Stroke

There were 13,999 episodes of stroke events across data sources during the follow-up. SGLT2is, GLP1-RAs, and DPP4is were associated with a lower hazard of stroke compared with SUs (0·77 [0·65-0·92], 0·79 [0·63-0·98], and 0·90 [0·81-0·99], respectively). Compared with DPP4is, SGLT2is and GLP1-RAs had a comparable hazard of stroke (0·89 [0·74-1·07] and 0·89 [0·77-1·03], respectively). There was no significant difference between SGLT2is and GLP1-RAs (0·93 [0·82-1·05]) regarding the hazard of stroke (**appendix pp 109, 128-133, figure 3**).

#### SCD

Over follow-up, 3,789 SCD occurred across data sources. SGLT2is and GLP1-RAs were associated with a lower hazard of SCD compared with DPP4is (0·62 (0·51-0·75), and 0·65 [0·44-0·96], respectively) and SUs (0·60 [0·48-0·73], and 0·57 [0·42-0·78], respectively). DPP4is versus SUs and SGLT2is versus GLP1-RAs had comparable hazards of SCD (0·96 [0·81-1·13], and 1·11 [0·84-1·45], respectively) (**appendix pp 109, 134-139, figure 3**).

#### HF Hospitalization

During the follow-up, 30,743 HF hospitalizations were recorded across all databases. SGLT2is and GLP1-RAs were associated with a lower risk of HF hospitalization compared with DPP4is (0·80 [0·68-0·95] and 0·76 [0·67-0·86], respectively) and SUs (0·68 [0·58-0·80] and 0·64 [0·52-0·78], respectively). DPP4is were associated with a lower hazard of HF hospitalization compared with SUs (0·90 [0·82-0·99]). Compared with GLP1-RAs, SGLT2is had a comparable hazard for HF hospitalization (1·02 [0·93-1·12]) (**appendix pp 109, 140-145, figure 3**).

### Sensitivity Analyses

Across all outcomes, 98·4% (690/701) of calibrated relative risk estimates remained within the confidence intervals of the primary meta-analysis when systematically removing each data source from the leave-one-out meta-analysis. Within the primary endpoints of 3-point MACE and 4-point MACE and secondary endpoints of acute MI and hospitalization for heart failure, none of the leave-one-out analyses were outside of the confidence intervals of the meta-analysis (**figure 4, appendix pp 196-201**).

**Figure 4:**
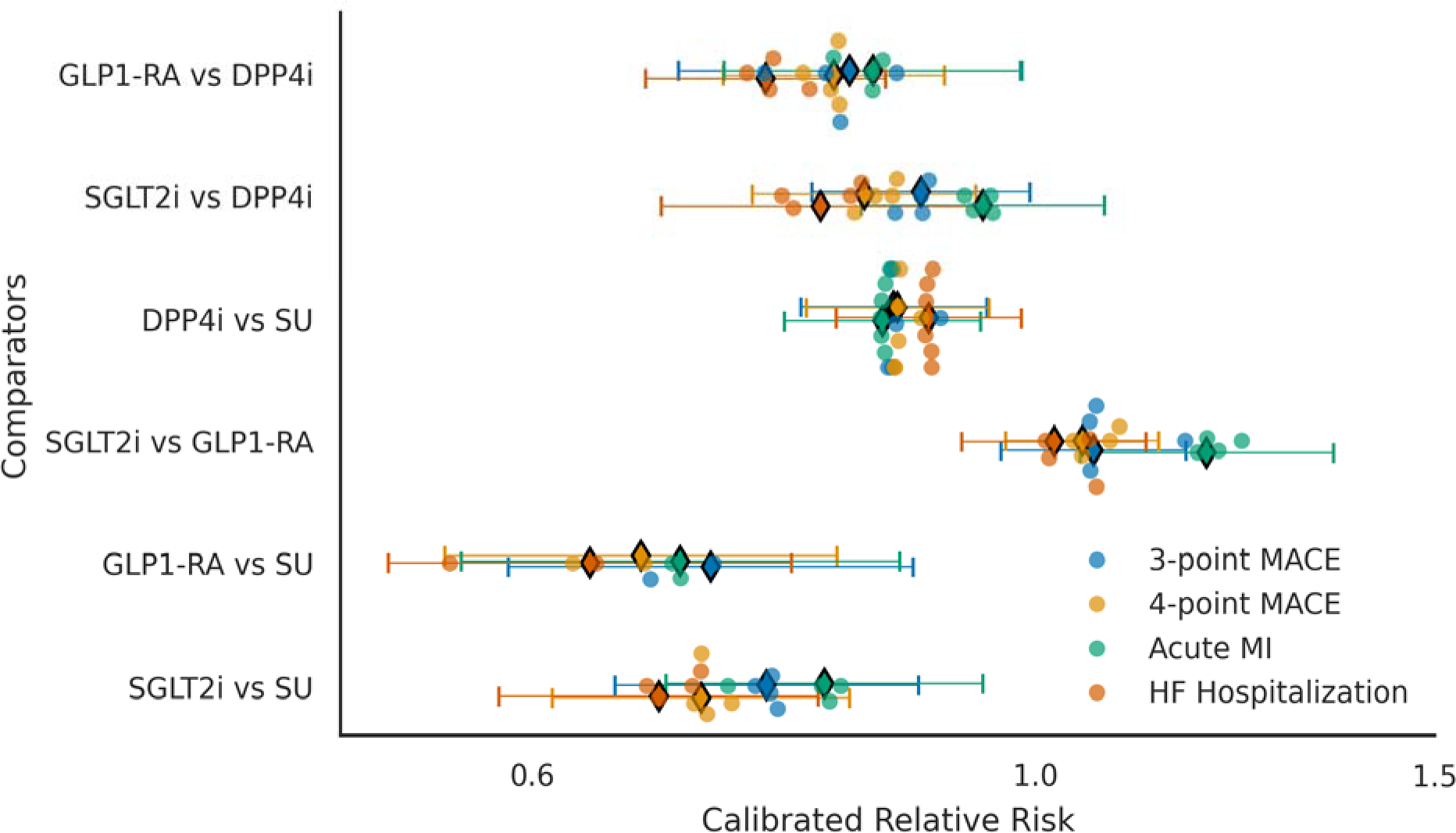
Swarm Plot of Calibrated Hazard Ratio Estimates of Major Outcome Meta-analysis and Leave-one-out Meta-analysis. Circles depict the calibrated relative risk of each leave-one-out study in which one original data source is removed from the meta-analysis. Diamonds depict the original meta-analysis with all sources. Points are color-coded by major outcomes, y-axis includes the medication comparators, and x-axis on log-scale measures the calibrated relative risk of each outcome-comparator combination. **Abbreviations:** DPP4i, dipeptidyl peptidase 4 inhibitor; GLP1-RA, glucagon-like peptide-1 receptor agonist; HF, heart failure; MACE, major adverse cardiovascular events; MI, myocardial infarction; SGLT2i, sodium-glucose co-transporter-2 inhibitor; SU, Sulfonylurea.

## DISCUSSION

In this largest, multinational, federated study of 1·5 million patients with T2DM and established CVD across ten cohorts initiating a second-line antihyperglycemic agent after metformin monotherapy, SGLT2is and GLP1-RAs were associated with an 11% and 17% lower risk of cardiovascular events compared with DPP4is, respectively, the class of drugs approved before the US FDA-mandated cardiovascular outcomes trials for antihyperglycemic agents.^50,51^ All three agents were associated with lower cardiovascular events than SUs, with 24% and 28% lower risk with SGLT2is and GLP1-RAs and 12% lower risk with DPP4is. These patterns were consistent across both ischemic events, such as MI and stroke, as well as for heart failure. Between SGLT2is and GLP1-RAs, there was no difference in composite cardiovascular effectiveness outcomes – both 3-point and 4-point MACE, but there was a lower risk of acute MI with GLP1-RAs compared with SGLT2is. The study accounted for multiple null control outcomes and calibrated all estimates for residual bias on null control outcomes, with multiple sensitivity analyses confirming the findings of the primary analysis.

This builds on the evidence of comparative cardiovascular effectiveness of currently used antihyperglycemic therapies, especially in the absence of head-to-head RCTs of any of these agents. There are no ongoing or future trials expected to fill this knowledge gap. A few observational studies have used single data sources to evaluate the cardiovascular outcomes associated with these drugs.^52–56^ Moreover, some associations observed in the present study, such as those with DPP4i, contrast with others where the smaller study population may have necessitated studying a combination of drugs directly against SU but challenges isolating an effect.^57^ These analyses are limited by local patterns of treatment use and challenges with exposure and outcome ascertainment in individual sources that may confound the observed association of these agents with outcomes. Our recent work shows that the use of antihyperglycemic agents varies substantially in populations across the world.^21^ In the present study, we developed the target trial for head-to-head of these agents, focusing explicitly on the same disease stage, i.e. when escalation to a second-line agent is being initiated. We conducted the same study across ten different data sources, ensuring that the effects of the intervention were not limited to the selective pressures or outcome differences in a single population. We ensured the empirical equipoise by comparing SMDs before and after PS stratification. Finally, we ensured that unmeasured confounding, potentially manifesting as directional effects across several negative control outcomes, were used to calibrate the PS stratified effect estimates.

The findings have important implications for clinical practice. Current clinical practice guidelines offer the use of SGLT2is and GLP1-RAs as suggested agents to be used among those with existing cardiovascular risk.^18,23,58^ We find strong evidence supportive of that suggestion. However, our recent work shows a substantial proportion of patients with T2DM are often initiated on DPP4is and SUs despite their limited cardiovascular effectiveness or known superiority on other glycemic or other diabetes-related outcomes.^59–61^ Our observations would favor stronger support for exclusive second-line use of SGLT2is and GLP1-RAs among individuals with established CVD. There is also evidence that suggests a potentially larger reduction in risk of acute MI with GLP1-RAs compared with SGLT2is, which were not replicated for stroke, questioning whether the current guideline recommendation for preferential use of GLP1-RAs in ASCVD is appropriate. Moreover, we did not observe an exclusive role of decreased HF risk with SGLT2i, as suggested in current guidelines, as there was no observed difference in HF risk among those receiving SGLT2i or GLP1-RA. The original recommendations for HF were based on inference from RCTs of individual agents and emerging evidence, such as from the STEP2-HFpEF trial that found improvements in heart failure outcomes with semaglutide support the potential role of GLP1-RAs on HF outcomes as well.^62^ Our study favors the continued evaluation of GLP1-RA as a therapy to reduce HF risk and improve outcomes of those with HF.^63,64^

There are limitations that merit consideration. First, to enable the ability to capture information across data sources spanning administrative and EHR data sources necessitated the use of administrative definitions of both exposures and outcomes. However, to ensure the information is accurately captured, we used definitions that have been validated in prior studies.^65–67^ Second, we expect the identification of the exposure and outcomes to vary across data sources. However, the consistency of effect estimates across data sources further confirms the validity of the observations. Third, capturing outcomes for EHR-based datasets, such as for recurrent hospitalizations and deaths, differs from administrative sources that track events across institutions and capture out-of-hospital death events.^68,69^ However, we find that sources that capture cross-institution out-of-hospital information did not differ and had similar outcomes as those that focused on institutionally captured death events, further supporting the robustness of the drug association with outcomes. Fourth, we do not have information on adherence to therapies and whether differential adherence of agents would confound the observations. However, these agents are widely used without substantial adverse effects.^18,19^ Moreover, lack of adherence will likely bias these observations to the null, and the observed positive association across agents with known cardiovascular benefits suggests that it is not driving the observed associations. Finally, inference on all outcomes can still be subject to residual confounding. However, our use of calibrated estimates explicitly addresses the identification of residual confounding across a series of null control outcomes.

## CONCLUSIONS

Our large, rigorous multinational network study of patients with T2DM and established CVD found cardiovascular risk reduction with SGLT2is and GLP1-RAs, with both agents more effective than DPP4is, which in turn were more effective than SUs. These findings suggest that the use of GLP1-RAs and SGLT2is should be prioritized as second-line agents in those with established CVD.

## Data Availability

To promote open science and avoid publication bias, we have disseminated all results in a publicly available R ShinyApp (https://data.ohdsi.org/LegendT2dmClassEvidenceExplorer/), and all analytic code is publicly available on GitHub (https://github.com/ohdsi-studies/LegendT2dm).

https://github.com/ohdsi-studies/LegendT2dm

## Acknowledgments

This study was partially funded through the National Institutes of Health grants K23 HL153775, R01 LM006910, R01 HG006139 and R01 HL169954, and the US Department of Veterans Affairs under the research priority to “Put VA Data to Work for Veterans” (VA ORD 22-D4V). The funders had no role in the design and conduct of the protocol; preparation, review, or approval of the manuscript; and decision to submit the manuscript for publication.

