## Supplementary Materials for "Comparative Effectiveness of Second-line Antihyperglycemic Agents for Cardiovascular Outcomes: A Large-scale, Multinational, Federated Analysis of the LEGEND-T2DM Study"

### Cohort Definitions

OHDSI's ATLAS (https://www.ohdsi.org/web/atlas) is an open-source software tool for researchers
to conduct scientific analyses on standardized observational data converted to the OMOP Common Data Model v5. Researchers can create cohorts by defining groups of people based on an exposure to a drug or diagnosis of a particular condition using healthcare claims data. ATLAS has vocabulary searching of medical concepts to identify people with specific conditions, drug exposures, etc. Patient profiles can be viewed within a specific cohort allowing visualization of a particular subject's health care records. Population effect level estimation analyses allow for comparing two different cohorts and leverage R packages. Here we have described the cohort definition for dipeptidyl peptidase-4 inhibitors (DPP4is) new users. The same method has been applied for defining cohorts of sodium-glucose co-transporter-2 inhibitors (SGLT2is), glucagon-like peptide-1 receptor agonists (GLP1-RAs), and sulfonylureas (SUs) new users.

#### I. Class-vs-Class Exposure (DPP4is New-User) Cohort / OT1

##### i. Cohort Entry Events

People with continuous observation of 365 days before the event may enter the cohort when observing any of the following:

1. drug exposure of 'DPP4 inhibitors' for the first time in the person's

history. Limit cohort entry events to the earliest event per person.

Restrict entry events to with all of the following criteria:

1. with the following event criteria: who are >= 18 years old.
2. having at least 1 condition occurrence of 'Type 2 diabetes mellitus,' starting anytime on or before cohort entry start date; allow events outside observation period.
3. having no condition occurrences of 'Type 1 diabetes mellitus,' starting anytime on or before cohort entry start date; allow events outside observation period.
4. having no condition occurrences of 'Secondary diabetes mellitus,' starting anytime on or before cohort entry start date; allow events outside observation period.

##### ii. Additional Inclusion Criteria

1. No prior GLP-1 receptor agonist exposure

Entry events having no drug exposures of 'GLP-1 receptor agonists,' starting anytime on or before cohort entry start date; allow events outside observation period.

2. No prior SGLT-2 inhibitor exposure

Entry events having no drug exposures of ‘SGLT2 inhibitors,’ starting anytime on or before cohort entry start date; allow events outside observation period.

3. No prior SU exposure

Entry events having no drug exposures of ‘Sulfonylureas,’ starting anytime on or before cohort entry start date; allow events outside observation period.

4. No prior other anti-diabetic exposure

Entry events having no drug exposures of ‘Other anti-diabetics,’ starting anytime on or before cohort entry start date; allow events outside observation period.

5. Prior metformin use

Entry events with any of the following criteria:

1. having at least 1 drug era of ‘Metformin,’ starting anytime up to 90 days before cohort entry start date; allow events outside observation period; with era length >= 90 days.
2. having at least 3 drug exposures of ‘Metformin,’ starting anytime on or before cohort entry start date; allow events outside observation period.

- No prior insulin use or combo initiation: Proxy for < 30 days drug era anytime before index and no combination use on index

Entry events with all of the following criteria:

1. having no drug eras of ‘Insulin,’ starting anytime up to 30 days before cohort entry start date; allow events outside observation period; with era length > 30 days.
2. having no drug eras of ‘Insulin,’ starting between 30 days before and 0 days after cohort entry start date; allow events outside observation period.

6. Higher cardiovascular risk

Entry events with any of the following criteria:

1. having at least 1 condition occurrence of ‘Conditions indicating established cardio- vascular disease,’ starting anytime on or before cohort entry start date; allow events outside observation period.
2. having at least 1 procedure occurrence of ‘Procedures indicating established cardio- vascular disease,’ starting anytime on or before cohort entry start date; allow events outside observation period.

**Concept: Conditions indicating established cardiovascular disease**

| Concept ID | Concept Name | Code | Vocabulary | Excluded | Descendants | Mapped |
|---|---|---|---|---|---|---|
| 319844 | Acute ischemic heart disease | 413439005 | SNOMED | NO | YES | NO |
| 321318 | Angina pectoris | 194828000 | SNOMED | NO | YES | NO |

| | | | | | | |
|---|---|---|---|---|---|---|
| 4124841 | Aortic bifurcation syndrome | 233972005 | SNOMED | YES | YES | NO |
| 312337 | Arterial embolus and thrombosis | 266262004 | SNOMED | NO | YES | NO |
| 4278217 | Arterial thrombosis | 65198009 | SNOMED | NO | YES | NO |
| 40484167 | Arteriosclerosis of artery of extremity | 443971004 | SNOMED | NO | YES | NO |
| 318443 | Arteriosclerotic vascular disease | 72092001 | SNOMED | NO | YES | NO |
| 314659 | Arteritis | 52089001 | SNOMED | NO | NO | NO |
| 40479625 | Atherosclerosis of artery | 441574008 | SNOMED | NO | YES | NO |
| 40484541 | Atherosclerosis of autologous vein bypass graft of limb | 442693003 | SNOMED | YES | YES | NO |
| 312902 | Benign intracranial hypertension | 68267002 | SNOMED | YES | YES | NO |
| 4288310 | Carotid artery obstruction | 69798007 | SNOMED | YES | YES | NO |
| 372924 | Cerebral artery occlusion | 20059004 | SNOMED | NO | YES | NO |
| 376713 | Cerebral hemorrhage | 274100004 | SNOMED | NO | YES | NO |
| 381591 | Cerebrovascular disease | 62914000 | SNOMED | NO | YES | NO |
| 316494 | Cerebrovascular disorder in the puerperium | 6594005 | SNOMED | YES | YES | NO |
| 315286 | Chronic ischemic heart disease | 413838009 | SNOMED | NO | YES | NO |
| 44782819 | Chronic occlusion of artery of extremity | 698816006 | SNOMED | NO | YES | NO |
| 4313767 | Chronic peripheral venous hypertension | 423674003 | SNOMED | YES | YES | NO |
| 372721 | Congenital anomaly of cerebrovascular system | 65587001 | SNOMED | YES | YES | NO |
| 316995 | Coronary occlusion | 63739005 | SNOMED | NO | YES | NO |
| 134057 | Disorder of cardiovascular system | 49601007 | SNOMED | NO | NO | NO |
| 40480453 | Disorder of vein of lower extremity | 441739009 | SNOMED | YES | YES | NO |
| 46272492 | Dissection of artery | 710864009 | SNOMED | YES | YES | NO |
| 4324690 | Fracture of skull | 71642004 | SNOMED | YES | YES | NO |
| 441246 | Hemangioma of intracranial structure | 93468003 | SNOMED | YES | YES | NO |
| 380113 | Hemorrhage in optic nerve sheaths | 14460007 | SNOMED | YES | YES | NO |
| 192763 | Injury of blood vessel | 57662003 | SNOMED | YES | YES | NO |
| 4275428 | Injury of vein | 64583005 | SNOMED | YES | YES | NO |
| 442774 | Intermittent claudication | 63491006 | SNOMED | NO | YES | NO |
| 439847 | Intracranial hemorrhage | 1386000 | SNOMED | NO | YES | NO |

| | | | | | | |
|---|---|---|---|---|---|---|
| 434056 | Late effects of cerebrovascular disease | 195239002 | SNOMED | NO | YES | NO |
| 4146311 | Leriche’s syndrome | 307816004 | SNOMED | NO | YES | NO |
| 4329847 | Myocardial infarction | 22298006 | SNOMED | NO | YES | NO |
| 4296029 | Periarteritis | 76805007 | SNOMED | NO | YES | NO |
| 260841 | Perinatal subarachnoid hemorrhage | 21202004 | SNOMED | YES | YES | NO |
| 317309 | Peripheral arterial occlusive disease | 399957001 | SNOMED | NO | YES | NO |
| 321822 | Peripheral vascular disorder due to diabetes mellitus | 421895002 | SNOMED | NO | YES | NO |
| 313928 | Peripheral vascular complication | 10596002 | SNOMED | NO | YES | NO |
| 321052 | Peripheral vascular disease | 400047006 | SNOMED | NO | NO | NO |
| 44782775 | Peripheral vascular disease associated with another disorder | 34881000119105 | SNOMED | NO | YES | NO |
| 318137 | Phlebitis and thrombophlebitis of intracranial sinuses | 192753009 | SNOMED | YES | YES | NO |
| 441039 | Phlebitis of lower limb vein | 312588002 | SNOMED | NO | YES | NO |
| 4067424 | Polyarteritis | 20258000 | SNOMED | NO | YES | NO |
| 320749 | Polyarteritis nodosa | 155441006 | SNOMED | YES | YES | NO |
| 443239 | Precerebral arterial occlusion | 266253001 | SNOMED | NO | YES | NO |
| 440417 | Pulmonary embolism | 59282003 | SNOMED | YES | YES | NO |
| 4318842 | Renal vasculitis | 95578000 | SNOMED | NO | YES | NO |
| 380943 | Rupture of syphilitic cerebral aneurysm | 186893003 | SNOMED | YES | YES | NO |
| 432923 | Subarachnoid hemorrhage | 21454007 | SNOMED | NO | YES | NO |
| 439040 | Subdural hemorrhage | 35486000 | SNOMED | NO | YES | NO |
| 320741 | Thrombophlebitis | 64156001 | SNOMED | YES | YES | NO |
| 4141106 | Thrombosis of arteries of the extremities | 33591000 | SNOMED | NO | YES | NO |
| 4132546 | Traumatic brain injury | 127295002 | SNOMED | YES | YES | NO |
| 4194610 | Trunk arterial embolus | 312593004 | SNOMED | NO | YES | NO |
| 318169 | Varicose veins of lower extremity | 72866009 | SNOMED | YES | YES | NO |
| 4189293 | Vascular disorder of lower extremity | 373408007 | SNOMED | NO | YES | NO |
| 443752 | Ventricular hemorrhage | 23276006 | SNOMED | YES | YES | NO |
| 432346 | Dissection of vertebral artery | 230730001 | SNOMED | YES | YES | NO |

### Concept: Procedures indicating established cardiovascular disease

| Concept ID | Concept Name | Code | Vocabulary | Excluded | Descendants | Mapped |
|---|---|---|---|---|---|---|
| 4150819 | Operative procedure on coronary artery | 31413008 | SNOMED | NO | YES | NO |
| 4331725 | Operative procedure on artery of extremity | 22701007 | SNOMED | NO | YES | NO |

#### iii. Cohort Exit

The cohort end date will be based on a continuous exposure to 'DPP4 inhibitors': allowing 30 days between exposures, adding 0 days after exposure ends, and using days supply and exposure end date for exposure duration.

#### iv. Cohort Eras

Entry events will be combined into cohort eras if they are within 0 days of each other.

#### v. Concept: DPP4 inhibitors

| Concept ID | Concept Name | Code | Vocabulary | Excluded | Descendants | Mapped |
|---|---|---|---|---|---|---|
| 43013884 | alogliptin | 1368001 | RxNorm | NO | YES | NO |
| 40239216 | linagliptin | 1100699 | RxNorm | NO | YES | NO |
| 40166035 | saxagliptin | 857974 | RxNorm | NO | YES | NO |
| 1580747 | sitagliptin | 593411 | RxNorm | NO | YES | NO |
| 19122137 | vildagliptin | 596554 | RxNorm | NO | YES | NO |

#### vi. Concept: GLP-1 receptor agonists

| Concept ID | Concept Name | Code | Vocabulary | Excluded | Descendants | Mapped |
|---|---|---|---|---|---|---|
| 44816332 | albiglutide | 1534763 | RxNorm | NO | YES | NO |
| 45774435 | dulaglutide | 1551291 | RxNorm | NO | YES | NO |
| 1583722 | exenatide | 60548 | RxNorm | NO | YES | NO |
| 40170911 | liraglutide | 475968 | RxNorm | NO | YES | NO |
| 44506754 | lixisenatide | 1440051 | RxNorm | NO | YES | NO |
| 793143 | semaglutide | 1991302 | RxNorm | NO | YES | NO |

#### vii. Concept: SGLT2 inhibitors

| Concept ID | Concept Name | Code | Vocabulary | Excluded | Descendants | Mapped |
|---|---|---|---|---|---|---|
| 43526465 | canagliflozin | 1373458 | RxNorm | NO | YES | NO |
| 44785829 | dapagliflozin | 1488564 | RxNorm | NO | YES | NO |
| 45774751 | empagliflozin | 1545653 | RxNorm | NO | YES | NO |
| 793293 | ertugliflozin | 1992672 | RxNorm | NO | YES | NO |

#### viii. Concept: Sulfonylureas

| Concept ID | Concept Name | Code | Vocabulary | Excluded | Descendants | Mapped |
|---|---|---|---|---|---|---|
| 1594973 | chlorpropamide | 2404 | RxNorm | NO | YES | NO |
| 1597756 | glimepiride | 25789 | RxNorm | NO | YES | NO |
| 1560171 | glipizide | 4821 | RxNorm | NO | YES | NO |
| 19097821 | gliquidone | 25793 | RxNorm | NO | YES | NO |
| 1559684 | glyburide | 4815 | RxNorm | NO | YES | NO |
| 1502809 | tolazamide | 10633 | RxNorm | NO | YES | NO |
| 1502855 | tolbutamide | 10635 | RxNorm | NO | YES | NO |

### ix. Concept: Other anti-diabetics

| Concept ID | Concept Name | Code | Vocabulary | Excluded | Descendants | Mapped |
|---|---|---|---|---|---|---|
| 1529331 | acarbose | 16681 | RxNorm | NO | YES | NO |
| 1530014 | acetohexamide | 173 | RxNorm | NO | YES | NO |
| 730548 | bromocriptine | 1760 | RxNorm | NO | YES | NO |
| 19033498 | carbutamide | 2068 | RxNorm | NO | YES | NO |
| 19001409 | glibornuride | 102846 | RxNorm | NO | YES | NO |
| 19059796 | gliclazide | 4816 | RxNorm | NO | YES | NO |
| 19001441 | glymidine | 102848 | RxNorm | NO | YES | NO |
| 1510202 | miglitol | 30009 | RxNorm | NO | YES | NO |
| 1502826 | nateglinide | 274332 | RxNorm | NO | YES | NO |
| 1525215 | pioglitazone | 33738 | RxNorm | NO | YES | NO |
| 1516766 | repaglinide | 73044 | RxNorm | NO | YES | NO |
| 1547504 | rosiglitazone | 84108 | RxNorm | NO | YES | NO |
| 1515249 | troglitazone | 72610 | RxNorm | NO | YES | NO |

### x. Concept: Insulin

| Concept ID | Concept Name | Code | Vocabulary | Excluded | Descendants | Mapped |
|---|---|---|---|---|---|---|
| 1596977 | insulin, regular, human | 253182 | RxNorm | NO | YES | NO |
| 1550023 | insulin lispro | 86009 | RxNorm | NO | YES | NO |
| 1567198 | insulin aspart, human | 51428 | RxNorm | NO | YES | NO |
| 1502905 | insulin glargine | 274783 | RxNorm | NO | YES | NO |
| 1513876 | insulin lispro protamine, human | 314684 | RxNorm | NO | YES | NO |
| 1531601 | insulin aspart protamine, human | 352385 | RxNorm | NO | YES | NO |
| 1586346 | insulin, regular, pork | 221109 | RxNorm | NO | YES | NO |
| 1544838 | insulin glulisine, human | 400008 | RxNorm | NO | YES | NO |
| 1516976 | insulin detemir | 139825 | RxNorm | NO | YES | NO |
| 1590165 | insulin, regular, beef-pork | 235275 | RxNorm | NO | YES | NO |
| 1513849 | lente insulin, human | 314683 | RxNorm | NO | YES | NO |
| 1562586 | lente insulin, pork | 93108 | RxNorm | NO | YES | NO |
| 1588986 | insulin human, rDNA origin | 631657 | RxNorm | NO | YES | NO |

| | | | | | | |
|---|---|---|---|---|---|---|
| 1513843 | lente insulin, beef-pork | 314682 | RxNorm | NO | YES | NO |
| 1586369 | ultralente insulin, human | 221110 | RxNorm | NO | YES | NO |
| 35605670 | insulin argine | 1740938 | RxNorm | NO | YES | NO |
| 35602717 | insulin degludec | 1670007 | RxNorm | NO | YES | NO |
| 21600713 | INSULINS AND ANALOGUES | A10A | ATC | NO | YES | NO |
| 19078608 | insulin, protamine zinc, beef-pork 100 UNT/ML Injectable Suspension | 311053 | RxNorm | NO | YES | NO |

#### xi. Concept: Metformin

| Concept ID | Concept Name | Code | Vocabulary | Excluded | Descendants | Mapped |
|---|---|---|---|---|---|---|
| 1503297 | metformin | 6809 | RxNorm | NO | YES | NO |

#### xii. Concept: Secondary diabetes mellitus

| Concept ID | Concept Name | Code | Vocabulary | Excluded | Descendants | Mapped |
|---|---|---|---|---|---|---|
| 195771 | Secondary diabetes mellitus | 8801005 | SNOMED | NO | YES | NO |

#### xiii. Concept: Type 1 diabetes mellitus

| Concept ID | Concept Name | Code | Vocabulary | Excluded | Descendants | Mapped |
|---|---|---|---|---|---|---|
| 201254 | Type 1 diabetes mellitus | 46635009 | SNOMED | NO | YES | NO |
| 435216 | Disorder due to type 1 diabetes mellitus | 420868002 | SNOMED | NO | YES | NO |
| 200687 | Renal disorder due to type 1 diabetes mellitus | 421893009 | SNOMED | NO | YES | NO |
| 377821 | Disorder of nervous system due to type 1 diabetes mellitus | 421468001 | SNOMED | NO | YES | NO |
| 318712 | Peripheral circulatory disorder due to type 1 diabetes mellitus | 421365002 | SNOMED | NO | YES | NO |

#### xiv. Concept: Type 2 diabetes mellitus

| Concept ID | Concept Name | Code | Vocabulary | Excluded | Descendants | Mapped |
|---|---|---|---|---|---|---|
| 201826 | Type 2 diabetes mellitus | 44054006 | SNOMED | NO | YES | NO |
| 443734 | Ketoacidosis due to type 2 diabetes mellitus | 421750000 | SNOMED | NO | YES | NO |
| 443767 | Disorder of eye due to diabetes mellitus | 25093002 | SNOMED | NO | YES | NO |
| 192279 | Disorder of kidney due to diabetes mellitus | 127013003 | SNOMED | NO | YES | NO |
| 443735 | Coma due to diabetes mellitus | 420662003 | SNOMED | NO | YES | NO |
| 376065 | Disorder of nervous system due to type 2 diabetes mellitus | 421326000 | SNOMED | NO | YES | NO |
| 443729 | Peripheral circulatory disorder due to type 2 diabetes mellitus | 422166005 | SNOMED | NO | YES | NO |
| 443732 | Disorder due to type 2 diabetes mellitus | 422014003 | SNOMED | NO | YES | NO |

### II. Metformin Use Modifier

#### i. No prior metformin use

Entry events having no drug eras of ‘Metformin,’ starting anytime on or before cohort entry start date; allow events outside observation period.

### III. Heterogenity Study Inclusion Criteria

#### i. Lower age group

Entry events with the following event criteria: who are < 45 years old.

#### ii. Middle age group

Entry events with all of the following criteria:

1. with the following event criteria: who are >= 45 years old.
2. with the following event criteria: who are < 65 years old.

#### iii. Older age group

Entry events with the following event criteria: who are >= 65 years old.

#### iv. Female stratum

Entry events with the following event criteria: who are female.

#### v. Male stratum

Entry events with the following event criteria: who are male.

#### vi. Race stratum

Entry events with the following event criteria: race is: “black or african american,” “black,” “african american,” “african,” “bahamian,” “barbadian,” “dominican,” “dominica islander,” “haitian,” “jamaican,” “tobagoan,” “trinidadian” or “west indian.”

### V. Escalation Exit Criteria

The cohort end date will be based on a continuous exposure to ‘DPP4 inhibitors’: allowing 30 days between exposures, adding 0 days after exposure ends, and using days supply and exposure end date for exposure duration.

The person also exits the cohort when encountering any of the following events:

1. drug exposures of ‘All alternative target exposures.’
2. drug exposures of ‘Other anti-diabetics.’
3. drug eras of ‘Insulin,’ with era length > 30 days.

#### i. Concept: All alternative target exposures

| Concept ID | Concept Name | Code | Vocabulary | Excluded | Descendants | Mapped |
|---|---|---|---|---|---|---|
| 44816332 | albiglutide | 1534763 | RxNorm | NO | YES | NO |
| 43526465 | canagliflozin | 1373458 | RxNorm | NO | YES | NO |
| 1594973 | chlorpropamide | 2404 | RxNorm | NO | YES | NO |
| 44785829 | dapagliflozin | 1488564 | RxNorm | NO | YES | NO |
| 45774435 | dulaglutide | 1551291 | RxNorm | NO | YES | NO |
| 45774751 | empagliflozin | 1545653 | RxNorm | NO | YES | NO |
| 793293 | ertugliflozin | 1992672 | RxNorm | NO | YES | NO |
| 1583722 | exenatide | 60548 | RxNorm | NO | YES | NO |
| 1597756 | glimepiride | 25789 | RxNorm | NO | YES | NO |
| 1560171 | glipizide | 4821 | RxNorm | NO | YES | NO |
| 19097821 | gliquidone | 25793 | RxNorm | NO | YES | NO |
| 1559684 | glyburide | 4815 | RxNorm | NO | YES | NO |
| 40170911 | liraglutide | 475968 | RxNorm | NO | YES | NO |
| 44506754 | lixisenatide | 1440051 | RxNorm | NO | YES | NO |
| 793143 | semaglutide | 1991302 | RxNorm | NO | YES | NO |
| 1502809 | tolazamide | 10633 | RxNorm | NO | YES | NO |
| 1502855 | tolbutamide | 10635 | RxNorm | NO | YES | NO |

### Study Outcomes

We have assessed 3-point major adverse cardiovascular events (MACE) and 4-point MACE as the primary study outcomes, where 3-point (MACE) includes acute myocardial infarction, stroke, and sudden cardiac death; and 4-point MACE additionally includes hospitalization for heart failure. Secondary outcomes of interest include the four individual MACE components as defined below.

#### Acute Myocardial Infarction

##### Cohort Entry Events

People may enter the cohort when observing any of the following:

1. condition occurrences of '[LEGEND-T2DM] Acute myocardial Infarction'.

Restrict entry events to having at least 1 visit occurrence of 'Inpatient or ER visit', starting anytime on or before cohort entry start date and ending between 0 days before and all days after cohort entry start date.

##### Cohort Exit

The cohort end date will be offset from index event's start date plus 7 days.

##### Cohort Eras

Entry events will be combined into cohort eras if they are within 180 days of each other.

##### Concept Set Definitions

1. Inpatient or ER visit

| Concept ID | Concept Name | Domain | Vocabulary | Excluded | Descendants | Mapped |
|---|---|---|---|---|---|---|
| 262 | Emergency Room and Inpatient Visit | Visit | Visit | NO | YES | NO |
| 9203 | Emergency Room Visit | Visit | Visit | NO | YES | NO |
| 9201 | Inpatient Visit | Visit | Visit | NO | YES | NO |

2. Acute myocardial infarction

| Concept ID | Concept Name | Domain | Vocabulary | Excluded | Descendants | Mapped |
|---|---|---|---|---|---|---|
| 4329847 | Myocardial infarction | Condition | SNOMED | NO | YES | NO |
| 314666 | Old myocardial infarction | Condition | SNOMED | YES | YES | NO |

#### Stroke

### Cohort Entry Events

People may enter the cohort when observing any of the following:

1. condition occurrences of '[LEGEND-T2DM] Stroke (ischemic or hemorrhagic)'.

Restrict entry events to having at least 1 visit occurrence of 'Inpatient or ER visit', starting between all days before and 1 days after cohort entry start date and ending between 0 days before and all days after cohort entry start date.

### Cohort Exit

The cohort end date will be offset from index event's start date plus 7 days.

### Cohort Eras

Entry events will be combined into cohort eras if they are within 180 days of each other.

### Concept Set Definitions

1. Inpatient or ER visit

| Concept ID | Concept Name | Domain | Vocabulary | Excluded | Descendants | Mapped |
|---|---|---|---|---|---|---|
| 262 | Emergency Room and Inpatient Visit | Visit | Visit | NO | YES | NO |
| 9203 | Emergency Room Visit | Visit | Visit | NO | YES | NO |
| 9201 | Inpatient Visit | Visit | Visit | NO | YES | NO |

2. Stroke (ischemic or hemorrhagic)

| Concept ID | Concept Name | Domain | Vocabulary | Excluded | Descendants | Mapped |
|---|---|---|---|---|---|---|
| 372924 | Cerebral artery occlusion | Condition | SNOMED | NO | NO | NO |
| 375557 | Cerebral embolism | Condition | SNOMED | NO | NO | NO |
| 376713 | Cerebral hemorrhage | Condition | SNOMED | NO | NO | NO |
| 443454 | Cerebral infarction | Condition | SNOMED | NO | YES | NO |
| 441874 | Cerebral thrombosis | Condition | SNOMED | NO | NO | NO |
| 439847 | Intracranial hemorrhage | Condition | SNOMED | NO | NO | NO |
| 432923 | Subarachnoid hemorrhage | Condition | SNOMED | NO | NO | NO |
| 43530727 | Spontaneous cerebral hemorrhage | Condition | SNOMED | NO | NO | NO |
| 4148906 | Spontaneous subarachnoid hemorrhage | Condition | SNOMED | NO | NO | NO |

#### Sudden Cardiac Death

### Cohort Entry Events

People may enter the cohort when observing any of the following:

1. condition occurrences of '[LEGEND HTN] Sudden cardiac death'.

Restrict entry events to having at least 1 visit occurrence of 'Inpatient or ER visit', starting anytime on or before cohort entry start date and ending between 0 days before and all days after cohort entry start date.

#### Cohort Exit

The cohort end date will be offset from index event's start date plus 7 days.

#### Cohort Eras

Entry events will be combined into cohort eras if they are within 180 days of each other.

#### Concept Set Definitions

1. Inpatient or ER visit

| Concept ID | Concept Name | Domain | Vocabulary | Excluded | Descendants | Mapped |
|---|---|---|---|---|---|---|
| 262 | Emergency Room and Inpatient Visit | Visit | Visit | NO | YES | NO |
| 9203 | Emergency Room Visit | Visit | Visit | NO | YES | NO |
| 9201 | Inpatient Visit | Visit | Visit | NO | YES | NO |

2. Sudden cardiac death

| Concept ID | Concept Name | Domain | Vocabulary | Excluded | Descendants | Mapped |
|---|---|---|---|---|---|---|
| 4048809 | Brainstem death | Condition | SNOMED | NO | YES | NO |
| 321042 | Cardiac arrest | Condition | SNOMED | NO | YES | NO |
| 442289 | Death in less than 24 hours from onset of symptoms | Condition | SNOMED | NO | YES | NO |
| 4317150 | Sudden cardiac death | Condition | SNOMED | NO | YES | NO |
| 4132309 | Sudden death | Condition | SNOMED | NO | YES | NO |
| 437894 | Ventricular fibrillation | Condition | SNOMED | YES | YES | NO |

### Heart Failure Hospitalization

#### Cohort Entry Events

People may enter the cohort when observing any of the following:

1. visit occurrences of 'Inpatient or ER visit'; having at least 1 condition occurrence of '[LEGEND-T2DM] Heart Failure', starting between 0 days before and all days after 'Inpatient or ER visit' start date and starting anytime on or before 'Inpatient or ER visit' end date.

### Cohort Exit

The cohort end date will be offset from index event's end date plus 0 days.

### Cohort Eras

Entry events will be combined into cohort eras if they are within 7 days of each other.

### Concept Set Definitions

1. Inpatient or ER visit

| Concept ID | Concept Name | Domain | Vocabulary | Excluded | Descendants | Mapped |
|---|---|---|---|---|---|---|
| 262 | Emergency Room and Inpatient Visit | Visit | Visit | NO | YES | NO |
| 9203 | Emergency Room Visit | Visit | Visit | NO | YES | NO |
| 9201 | Inpatient Visit | Visit | Visit | NO | YES | NO |

2. Heart Failure

| Concept ID | Concept Name | Domain | Vocabulary | Excluded | Descendants | Mapped |
|---|---|---|---|---|---|---|
| 315295 | Congestive rheumatic heart failure | Condition | SNOMED | YES | YES | NO |
| 316139 | Heart failure | Condition | SNOMED | NO | YES | NO |

### Negative Control Outcomes

We selected negative controls using a process similar to that outlined in Voss et al. [1]. We first constructed a list of all conditions that satisfy the following criteria with respect to all drug exposures in the LEGEND-HTN study [2]:

- No Medline abstract where the MeSH terms suggests a drug-condition association [3],
- No mention of the drug-condition pair on a US product label in the “Adverse Drug Reactions” or “Postmarketing” section [4],
- No US spontaneous reports suggesting that the pair is in an adverse event relationship [5, 6],
- OMOP vocabulary does not suggest that the drug is indicated for the condition,
- Vocabulary conditional concepts are usable (i.e., not too broad, not suggestive of an adverse event relationship, no pregnancy related), and
- Exact condition concept itself is used in patient level data.

We then optimized the remaining condition concepts, such that parent concepts remove children as defined by the OMOP vocabulary and performed manual review to exclude any pairs that may still be in a causal relationship or too similar to the study outcome. This process led to a candidate list of 100 negative controls (**Supplementary Table 4**).

**Supplementary Table 1:** Description of databases from the observational health data sciences and informatics network included in the study.

| Name of Database | Abbreviation | Type of Data | Country of Origin | Years of Exposure Included |
|---|---|---|---|---|
| IBM MarketScan® Commercial Claims and Encounters Data (CCAE) | CCAE | Claims Data | USA | 2011-2021 |
| Germany Disease Analyser | GDA | EHR Data | Germany | 1992-2021 |
| UK-IQVIA Medical Research Data | IMRD | EHR Data | United Kingdom | 2011-2019 |
| IBM Health MarketScan® Multi-State Medicaid Database | MDCD | Claims Data | USA | 2011-2020 |
| IBM Health MarketScan® Medicare Supplemental and Coordination of Benefits Database | MDCR | Claims Data | USA | 2011-2021 |
| Optum© Clinformatics Extended Data Mart - Date of Death (DOD) | OCEDM | Claims Data | USA | 2011-2021 |
| Optum© de-identified Electronic Health Record Dataset | OEHR | Claims Data | USA | 2011-2021 |
| Information System for Research in Primary Care | SIDIAP | EHR Data | Spain | 2011-2021 |
| US Open Claims | USOC | Claims Data | USA | 2000-2021 |
| Department of Veterans Affairs Healthcare System | VA | EHR Data | USA | 2011-2021 |

**Supplementary Table 2:** Brief descriptions of databases from the observational health data sciences and informatics network included in the study.

| Name of Database | Abbreviation | Brief Description |
|---|---|---|
| **US National Databases** | | |
| IBM MarketScan® Commercial Claims and Encounters Data | CCAE | IBM Health MarketScan® Commercial Claims and Encounters Database (CCAE) represent data from individuals enrolled in United States employer-sponsored insurance health plans. The data includes adjudicated health insurance claims (e.g. inpatient, outpatient, and outpatient pharmacy) as well as enrollment data from large employers and health plans who provide private healthcare coverage to employees, their spouses, and dependents. Additionally, it captures laboratory tests for a subset of the covered lives. This administrative claims database includes a variety of fee-for-service, preferred provider organizations, and capitated health plans. |
| IBM Health MarketScan® Multi-State Medicaid Database | MDCD | IBM MarketScan® Multi-State Medicaid Database (MDCD) adjudicated US health insurance claims for Medicaid enrollees from multiple states and includes hospital discharge diagnoses, outpatient diagnoses and procedures, and outpatient pharmacy claims as well as ethnicity and Medicare eligibility. Members maintain their same identifier even if they leave the system for a brief period however the dataset lacks lab data. |
| IBM Health MarketScan Medicare Supplemental and Coordination of Benefits Database | MDCR | IBM Health MarketScan® Medicare Supplemental and Coordination of Benefits Database (MDCR) represents health services of retirees in the United States with primary or Medicare supplemental coverage through privately insured fee-for-service, point-of-service, or capitated health plans. These data include adjudicated health insurance claims (e.g. inpatient, outpatient, and outpatient pharmacy). Additionally, it captures laboratory tests for a subset of the covered lives. |
| Optum Clinformatics Extended Data Mart - Date of Death (DOD) | OCEDM | Optum Clinformatics Extended DataMart is an adjudicated US administrative health claims database for members of private health insurance, who are fully insured in commercial plans or in administrative services only (ASOs), Legacy Medicare Choice Lives (prior to January 2006), and Medicare Advantage (Medicare Advantage Prescription Drug coverage starting January 2006). The population is primarily representative of commercial claims patients (0-65 years old) with some Medicare (65+ years old) however ages are capped at 90 years. It includes data captured from administrative claims processed from inpatient and outpatient medical services and prescriptions as dispensed, as well as results for outpatient lab tests processed by large national lab vendors who participate in data exchange with Optum. This dataset also provides date of death (month and year only) for members with both medical and pharmacy coverage from the Social Security Death Master File (however after 2011 reporting frequency changed due to changes in reporting requirements) and location information for patients is at the US state level. |
| Optum© de-identified Electronic Health Record Dataset | OEHR | Optum© de-identified Electronic Health Record Dataset represents Humedica's Electronic Health Record data a medical records database. The medical record data includes clinical information, inclusive of prescriptions as prescribed and administered, lab results, vital signs, body measurements, diagnoses, procedures, and information derived from clinical Notes using Natural Language Processing (NLP). |

| Name of Database | Abbreviation | Brief Description |
|---|---|---|
| US Open Claims | USOC | US Open Claims is a United States database of open, pre-adjudicated claims from 2000 to present. Data are reported at anonymized patient level collected from office-based physicians and specialists via office management software and clearinghouse switch sources for the purpose of reimbursement. A subset of medical claims data has adjudicated claims. |
| **US Health System Databases** | | |
| Department of Veterans Affairs health care system | VA | VA OMOP data reflects the national Department of Veterans Affairs health care system, which is the largest integrated provider of medical and mental health services in the United States. Care is provided at 170 VA Medical Centers and 1,063 outpatient sites serving more than 9 million enrolled Veterans each year. |
| **Non-US Databases** | | |
| Germany Disease Analyser | GDA | Germany Disease Analyser is collected from extracts of patient management software used by general practitioners and specialists practicing in ambulatory care settings. Data coverage includes 40.2 million distinct person records, about 48.2% population in the country and collected from 2,800 providers. Patient visiting more than one provider are not cross identified for data protection reasons and therefore recorded as separate in the system. Dates of service include from 1992 through present. Observation time is defined by the first and last consultation dates. Germany has no mandatory general practitioner system and patient have free choice of specialist. Drugs are recorded as prescriptions of marketed products. No registration or approval is required for drug utilization studies. |
| Information System for Research in Primary Care | SIDIAP | The Information System for Research in Primary Care (SIDIAP; www.sidiap.org) is a primary care records database that covers approximately 80% of the population of Catalonia, North-East Spain. Healthcare is universal and tax-payer funded in the region, and primary care physicians are gatekeepers for all care and responsible for repeat prescriptions. For this study, SIDIAP was linked to the Minimum Basic Dataset (CMBD), a national population-based registry that includes hospital discharge information of mandatory registration. |
| UK-IQVIA Medical Research Data | IMRD | The UK-IQVIA Medical Research Data (IMRD), previously known as The Health Improvement Network (THIN), contains anonymized electronic health records from over 744 general practices in the UK, covering approximately 6% of the UK population. It contains data on prescriptions, diagnoses, referrals, and patient demographics broadly representative of the UK. |

**Supplementary Table 3:** Pharmacological agents included in the drug classes.

| GLP1-RA | SGLT2i | DPP4i | SU |
|---|---|---|---|
| Albiglutide | Canagliflozin | Alogliptin | Chlorpropamide |
| Dulaglutide | Dapagliflozin | Linagliptin | Glimepiride |
| Exenatide | Empagliflozin | Saxagliptin | Glipizide |
| Liraglutide | Ertugliflozin | Sitagliptin | Gliquidone |
| Lixisenatide | | Vildagliptin | Glyburide |
| Semaglutide | | | Tolazamide |
| | | | Tolbutamide |

**Supplementary Table 4:** Negative control outcomes.

| **Condition Concept** |
|---|
| Abnormal posture |
| Abrasion and/or friction burn of multiple sites |
| Absent kidney |
| Anomaly of jaw size |
| Benign paroxysmal positional vertigo |
| Bizarre personal appearance |
| Cachexia |
| Colostomy present |
| Complication of gastrostomy |
| Developmental delay |
| Deviated nasal septum |
| Epidermoid cyst |
| Exhaustion due to excessive exertion |
| Feces contents abnormal |
| Foreign body in ear |
| Foreskin deficient |
| Galactosemia |
| Ganglion cyst |
| Genetic disorder carrier |
| Impacted cerumen |
| Inadequate sleep hygiene |
| Jellyfish poisoning |
| Lagophthalmos |
| Lipid storage disease |
| Lymphangioma |
| Malingering |
| Marfan's syndrome |
| Mechanical complication of internal orthopedic device, implant AND/OR graft |
| Minimal cognitive impairment |
| Nicotine dependence |
| Nonspecific tuberculin test reaction |
| Opioid abuse |
| Opioid intoxication |
| Physiological development failure |
| Poisoning by tranquilizer |
| Social exclusion |
| Symbolic dysfunction |
| Tooth loss |
| Toxic effect of lead compound |
| Toxic effect of tobacco and nicotine |
| Tracheostomy present |
| Unsatisfactory tooth restoration |
| Abnormal pupil |
| Abrasion and/or friction burn of trunk without infection |
| Absence of breast |
| Acquired hallux valgus |
| Acquired keratoderma |
| Anal and rectal polyp |
| Burn of forearm |

| |
|---|
| Calcaneal spur |
| Cannabis abuse |
| Changes in skin texture |
| Chondromalacia of patella |
| Cocaine abuse |
| Complication due to Crohn’s disease |
| Contact dermatitis |
| Contusion of knee |
| Crohn’s disease |
| Derangement of knee |
| Difficulty sleeping |
| Disproportion of reconstructed breast |
| Effects of hunger |
| Endometriosis |
| Feces contents abnormal |
| Foreign body in orifice |
| Ganglion cyst |
| Hammer toe |
| Hereditary thrombophilia |
| High risk sexual behavior |
| Homocystinuria |
| Impacted cerumen |
| Impingement syndrome of shoulder region |
| Ingrowing nail |
| Injury of knee |
| Kwashiorkor |
| Late effect of contusion |
| Late effect of motor vehicle accident |
| Macular drusen |
| Melena |
| Nicotine dependence |
| Noise effects on inner ear |
| Non-toxic multinodular goiter |
| Nonspecific tuberculin test reaction |
| Opioid abuse |
| Passing flatus |
| Postviral fatigue syndrome |
| Presbyopia |
| Psychalgia |
| Ptotic breast |
| Regular astigmatism |
| Senile hyperkeratosis |
| Somatic dysfunction of lumbar region |
| Splinter of face without major open wound |
| Sprain of ankle |
| Strain of rotator cuff capsule |
| Tear film insufficiency |
| Tobacco dependence syndrome |
| Verruca vulgaris |
| Wrist joint pain |
| Wristdrop |

**Supplementary Table 5:** Baseline patient characteristics for SGLT2is (T) and GLP1-RAs (C) new-users in the CCAE data source. We report proportion of initiators satisfying selected base-line characteristics and the standardized difference of population proportions (StdDiff) before and after propensity score adjustment. Less extreme StdDiffs through matching and stratification suggest improved balance between patient cohorts through adjustment.

| | Before adjustment | | | After matching | | | After stratification | | |
|---|---|---|---|---|---|---|---|---|---|
| Characteristic | T (%) | C (%) | StdDiff | T (%) | C (%) | StdDiff | T (%) | C (%) | StdDiff |
| **Age group** | | | | | | | | | |
| 20 - 24 | 0.1 | 0.2 | 0.03 | 0.2 | 0.2 | 0.01 | 0.1 | 0.1 | -0.01 |
| 25 - 29 | 0.1 | 0.2 | 0.04 | 0.1 | 0.2 | 0.02 | 0.1 | 0.2 | 0.04 |
| 30 - 34 | 0.3 | 0.9 | 0.08 | 0.4 | 0.8 | 0.05 | 0.4 | 0.8 | 0.05 |
| 35 - 39 | 1.7 | 2.6 | 0.06 | 2.0 | 2.3 | 0.02 | 1.8 | 2.0 | 0.01 |
| 40 - 44 | 4.5 | 6.3 | 0.08 | 5.2 | 5.8 | 0.03 | 4.9 | 5.3 | 0.02 |
| 45 - 49 | 10.1 | 11.5 | 0.05 | 10.8 | 11.4 | 0.02 | 10.4 | 10.8 | 0.01 |
| 50 - 54 | 18.3 | 20.2 | 0.05 | 19.3 | 19.9 | 0.02 | 19.0 | 19.5 | 0.01 |
| 55 - 59 | 29.2 | 27.0 | -0.05 | 28.1 | 27.4 | -0.02 | 28.6 | 28.4 | 0.00 |
| 60 - 64 | 33.0 | 28.6 | -0.10 | 31.5 | 29.6 | -0.04 | 32.1 | 30.2 | -0.04 |
| 65 - 69 | 2.8 | 2.4 | -0.02 | 2.5 | 2.4 | -0.01 | 2.7 | 2.7 | 0.00 |
| Gender: female | 38.1 | 53.8 | 0.32 | 48.2 | 46.1 | -0.04 | 43.0 | 41.4 | -0.03 |
| **Medical history: General** | | | | | | | | | |
| Acute respiratory disease | 29.6 | 33.5 | 0.09 | 33.6 | 32.5 | -0.02 | 31.5 | 30.2 | -0.03 |
| Attention deficit hyperactivity disorder | 1.1 | 1.5 | 0.03 | 1.6 | 1.5 | -0.01 | 1.3 | 1.2 | -0.01 |
| Chronic liver disease | 2.5 | 2.6 | 0.01 | 2.1 | 2.0 | 0.00 | 2.4 | 2.3 | 0.00 |
| Chronic obstructive lung disease | 5.2 | 5.5 | 0.01 | 5.8 | 5.1 | -0.03 | 5.5 | 4.8 | -0.03 |
| Crohn’s disease | 0.3 | 0.4 | 0.02 | 0.4 | 0.5 | 0.01 | 0.3 | 0.5 | 0.02 |
| Dementia | 0.3 | 0.2 | 0.00 | 0.3 | 0.3 | 0.00 | 0.3 | 0.3 | 0.00 |
| Depressive disorder | 11.8 | 15.7 | 0.11 | 15.5 | 14.7 | -0.02 | 13.7 | 13.4 | -0.01 |
| Gastroesophageal reflux disease | 17.2 | 18.2 | 0.03 | 19.7 | 18.3 | -0.04 | 18.5 | 17.8 | -0.02 |
| Gastrointestinal hemorrhage | 2.0 | 2.8 | 0.05 | 2.4 | 2.5 | 0.00 | 2.2 | 2.4 | 0.01 |
| Human immunodeficiency virus infection | 0.4 | 0.3 | -0.02 | 0.4 | 0.2 | -0.02 | 0.3 | 0.3 | -0.01 |
| Hyperlipidemia | 82.2 | 74.9 | -0.18 | 80.3 | 79.9 | -0.01 | 81.3 | 81.1 | 0.00 |
| Hypertensive disorder | 81.8 | 76.8 | -0.12 | 81.1 | 81.0 | 0.00 | 81.5 | 81.9 | 0.01 |
| Lesion of liver | 0.8 | 0.9 | 0.01 | 1.0 | 0.9 | -0.01 | 0.9 | 0.8 | -0.01 |
| Obesity | 31.5 | 38.0 | 0.14 | 43.7 | 40.7 | -0.06 | 37.6 | 36.0 | -0.03 |
| Osteoarthritis | 24.5 | 29.5 | 0.11 | 28.9 | 28.3 | -0.01 | 26.8 | 26.4 | -0.01 |
| Pneumonia | 3.2 | 3.3 | 0.01 | 3.7 | 3.3 | -0.02 | 3.5 | 2.8 | -0.04 |
| Psoriasis | 2.2 | 2.7 | 0.03 | 2.7 | 2.7 | 0.00 | 2.4 | 2.6 | 0.01 |
| Renal impairment | 5.3 | 5.6 | 0.01 | 6.1 | 6.6 | 0.02 | 5.7 | 6.2 | 0.02 |
| Rheumatoid arthritis | 1.6 | 2.0 | 0.03 | 2.0 | 1.7 | -0.03 | 1.7 | 1.5 | -0.02 |
| Schizophrenia | 0.1 | <0.1 | -0.03 | 0.2 | <0.1 | -0.04 | 0.2 | <0.1 | -0.04 |
| Ulcerative colitis | 0.4 | 0.5 | 0.01 | 0.4 | 0.4 | 0.00 | 0.4 | 0.4 | 0.00 |
| Urinary tract infectious disease | 6.8 | 9.2 | 0.09 | 8.6 | 7.8 | -0.03 | 7.7 | 7.5 | -0.01 |
| Viral hepatitis C | 0.4 | 0.3 | -0.01 | 0.3 | 0.2 | -0.03 | 0.3 | 0.2 | -0.02 |
| Visual system disorder | 28.1 | 30.2 | 0.05 | 30.2 | 30.2 | 0.00 | 29.2 | 29.4 | 0.00 |
| **Medical history: Cardiovascular disease** | | | | | | | | | |
| Atrial fibrillation | 5.0 | 4.8 | -0.01 | 5.0 | 5.3 | 0.01 | 5.0 | 5.3 | 0.01 |
| Cerebrovascular disease | 9.1 | 8.6 | -0.02 | 8.5 | 8.1 | -0.01 | 8.9 | 8.3 | -0.02 |
| Coronary arteriosclerosis | 34.8 | 28.0 | -0.15 | 27.6 | 28.7 | 0.02 | 31.5 | 33.0 | 0.03 |
| Heart disease | 50.8 | 47.0 | -0.08 | 45.5 | 46.8 | 0.03 | 48.5 | 50.4 | 0.04 |
| Heart failure | 6.7 | 5.7 | -0.04 | 6.1 | 5.7 | -0.01 | 6.5 | 5.9 | -0.03 |
| Ischemic heart disease | 18.2 | 16.1 | -0.06 | 17.1 | 17.4 | 0.01 | 17.1 | 17.4 | 0.01 |
| Peripheral vascular disease | 15.6 | 16.1 | 0.02 | 16.9 | 17.6 | 0.02 | 16.2 | 16.4 | 0.00 |
| Pulmonary embolism | 0.9 | 1.4 | 0.05 | 1.1 | 1.4 | 0.03 | 0.9 | 1.3 | 0.04 |
| Venous thrombosis | 1.5 | 2.0 | 0.04 | 1.7 | 1.8 | 0.01 | 1.6 | 1.7 | 0.00 |
| **Medical history: Neoplasms** | | | | | | | | | |
| Hematologic neoplasm | 1.0 | 1.0 | 0.00 | 1.2 | 1.1 | -0.01 | 1.1 | 1.0 | -0.01 |
| Malignant lymphoma | 0.4 | 0.5 | 0.02 | 0.4 | 0.7 | 0.03 | 0.4 | 0.6 | 0.03 |
| Malignant neoplasm of anorectum | 0.1 | 0.1 | -0.02 | 0.2 | <0.1 | -0.02 | 0.2 | 0.1 | -0.02 |
| Malignant neoplastic disease | 7.2 | 7.7 | 0.02 | 8.0 | 7.3 | -0.03 | 7.6 | 7.1 | -0.02 |
| Malignant tumor of breast | 1.2 | 1.5 | 0.02 | 1.6 | 1.2 | -0.04 | 1.5 | 0.9 | -0.05 |
| Malignant tumor of colon | 0.3 | 0.2 | -0.02 | 0.3 | 0.3 | -0.01 | 0.3 | 0.3 | -0.01 |
| Malignant tumor of lung | 0.3 | 0.1 | -0.04 | 0.3 | <0.1 | -0.06 | 0.3 | 0.1 | -0.04 |
| Malignant tumor of urinary bladder | 0.2 | 0.2 | 0.00 | 0.2 | 0.2 | 0.01 | 0.2 | 0.3 | 0.01 |
| Primary malignant neoplasm of prostate | 1.1 | 0.8 | -0.03 | 1.0 | 0.9 | -0.01 | 1.0 | 0.9 | -0.01 |
| **Medication use** | | | | | | | | | |
| Agents acting on the renin-angiotensin system | 74.5 | 70.6 | -0.09 | 72.0 | 71.9 | 0.00 | 73.2 | 73.8 | 0.01 |
| Antibacterials for systemic use | 61.5 | 67.5 | 0.13 | 66.0 | 64.6 | -0.03 | 63.8 | 62.8 | -0.02 |
| Antidepressants | 30.2 | 41.0 | 0.23 | 37.8 | 36.2 | -0.03 | 34.0 | 33.0 | -0.02 |
| Antiepileptics | 17.2 | 21.9 | 0.12 | 20.9 | 20.4 | -0.01 | 19.1 | 18.7 | -0.01 |

| (Continued) | Before ajustment | | | After matching | | | After stratification | | |
|---|---|---|---|---|---|---|---|---|---|
| Characteristic | T (%) | C (%) | StdDiff | T (%) | C (%) | StdDiff | T (%) | C (%) | StdDiff |
| Antiinflammatory and antirheumatic products | 39.0 | 42.3 | 0.07 | 42.4 | 41.7 | -0.01 | 40.8 | 40.0 | -0.02 |
| Antineoplastic agents | 4.5 | 6.8 | 0.10 | 5.9 | 5.5 | -0.02 | 5.3 | 5.1 | -0.01 |
| Antipsoriatics | 0.8 | 1.0 | 0.02 | 0.9 | 0.9 | 0.00 | 0.9 | 0.9 | 0.00 |
| Antithrombotic agents | 28.4 | 23.8 | -0.10 | 24.5 | 24.6 | 0.00 | 26.7 | 27.1 | 0.01 |
| Beta blocking agents | 47.3 | 43.4 | -0.08 | 43.7 | 43.6 | 0.00 | 45.7 | 46.2 | 0.01 |
| Calcium channel blockers | 27.3 | 26.5 | -0.02 | 27.3 | 27.0 | -0.01 | 27.2 | 27.1 | 0.00 |
| Diuretics | 42.5 | 49.4 | 0.14 | 46.3 | 45.2 | -0.02 | 44.3 | 43.4 | -0.02 |
| Drugs for acid related disorders | 31.2 | 35.8 | 0.10 | 33.9 | 33.0 | -0.02 | 32.6 | 32.4 | 0.00 |
| Drugs for obstructive airway diseases | 42.0 | 48.1 | 0.12 | 47.9 | 46.3 | -0.03 | 44.9 | 44.3 | -0.01 |
| Immunosuppressants | 3.2 | 4.5 | 0.07 | 4.3 | 4.1 | -0.01 | 3.8 | 3.8 | 0.00 |
| Lipid modifying agents | 80.0 | 75.1 | -0.12 | 76.3 | 76.6 | 0.01 | 78.1 | 78.7 | 0.01 |
| Opioids | 31.3 | 35.2 | 0.08 | 33.6 | 33.2 | -0.01 | 32.5 | 32.6 | 0.00 |
| Psycholeptics | 28.4 | 34.3 | 0.13 | 32.6 | 31.3 | -0.03 | 30.5 | 29.9 | -0.01 |
| Psychostimulants | 3.6 | 5.9 | 0.11 | 4.8 | 5.2 | 0.02 | 4.3 | 4.6 | 0.02 |

**Supplementary Table 6:** Baseline patient characteristics for SGLT2is (T) and GLP1-RAs (C) new-users in the OptumDOD data source. We report proportion of initiators satisfying selected base-line characteristics and the standardized difference of population proportions (StdDiff) before and after propensity score adjustment. Less extreme StdDiffs through matching and stratification suggest improved balance between patient cohorts through adjustment.

| | Before adjustment | | | After matching | | | After stratification | | |
|---|---|---|---|---|---|---|---|---|---|
| Characteristic | T (%) | C (%) | StdDiff | T (%) | C (%) | StdDiff | T (%) | C (%) | StdDiff |
| **Age group** | | | | | | | | | |
| 25 - 29 | 0.1 | 0.1 | 0.02 | 0.1 | <0.1 | -0.01 | 0.1 | 0.1 | 0.00 |
| 30 - 34 | 0.2 | 0.5 | 0.05 | 0.3 | 0.4 | 0.02 | 0.2 | 0.4 | 0.02 |
| 35 - 39 | 0.7 | 1.6 | 0.08 | 0.9 | 1.3 | 0.04 | 0.8 | 1.1 | 0.03 |
| 40 - 44 | 2.1 | 3.4 | 0.08 | 2.7 | 2.8 | 0.01 | 2.5 | 2.7 | 0.01 |
| 45 - 49 | 4.6 | 6.4 | 0.08 | 5.5 | 5.4 | 0.00 | 5.0 | 5.3 | 0.01 |
| 50 - 54 | 7.7 | 11.4 | 0.13 | 9.5 | 9.8 | 0.01 | 8.7 | 8.8 | 0.00 |
| 55 - 59 | 12.7 | 14.9 | 0.06 | 13.8 | 14.1 | 0.01 | 13.2 | 13.5 | 0.01 |
| 60 - 64 | 15.2 | 16.7 | 0.04 | 16.2 | 16.5 | 0.01 | 15.6 | 16.2 | 0.01 |
| 65 - 69 | 19.0 | 17.8 | -0.03 | 19.3 | 18.9 | -0.01 | 19.1 | 18.3 | -0.02 |
| 70 - 74 | 19.5 | 15.1 | -0.12 | 17.1 | 16.8 | -0.01 | 18.3 | 18.6 | 0.01 |
| 75 - 79 | 11.5 | 8.3 | -0.11 | 9.6 | 9.6 | 0.00 | 10.6 | 10.5 | 0.00 |
| 80 - 84 | 4.6 | 2.7 | -0.10 | 3.5 | 3.1 | -0.02 | 4.1 | 3.5 | -0.03 |
| 85 - 89 | 1.8 | 0.8 | -0.09 | 1.5 | 0.9 | -0.05 | 1.7 | 1.0 | -0.06 |
| 90 - 94 | 0.2 | <0.1 | -0.05 | 0.2 | <0.1 | -0.03 | 0.2 | <0.1 | -0.05 |
| Gender: female | 38.7 | 52.9 | 0.29 | 50.5 | 47.6 | -0.06 | 43.7 | 41.8 | -0.04 |
| **Medical history: General** | | | | | | | | | |
| Acute respiratory disease | 27.8 | 30.1 | 0.05 | 29.2 | 28.0 | -0.03 | 28.7 | 27.8 | -0.02 |
| Attention deficit hyperactivity disorder | 0.7 | 1.3 | 0.05 | 1.0 | 0.8 | -0.02 | 0.9 | 0.8 | 0.00 |
| Chronic liver disease | 2.2 | 2.7 | 0.04 | 2.1 | 2.3 | 0.01 | 2.2 | 2.1 | -0.01 |
| Chronic obstructive lung disease | 14.6 | 14.5 | 0.00 | 15.2 | 14.8 | -0.01 | 15.0 | 14.8 | -0.01 |
| Crohn’s disease | 0.4 | 0.3 | -0.02 | 0.4 | 0.2 | -0.04 | 0.4 | 0.2 | -0.03 |
| Dementia | 1.7 | 1.8 | 0.00 | 1.7 | 1.9 | 0.01 | 1.7 | 1.8 | 0.01 |
| Depressive disorder | 17.9 | 24.4 | 0.16 | 24.0 | 21.7 | -0.05 | 20.9 | 19.8 | -0.03 |
| Gastroesophageal reflux disease | 25.1 | 26.6 | 0.04 | 27.4 | 25.5 | -0.04 | 26.2 | 25.5 | -0.02 |
| Gastrointestinal hemorrhage | 2.9 | 2.8 | 0.00 | 2.9 | 2.4 | -0.03 | 2.9 | 2.6 | -0.02 |
| Human immunodeficiency virus infection | 0.3 | 0.6 | 0.04 | 0.4 | 0.6 | 0.02 | 0.4 | 0.6 | 0.03 |
| Hyperlipidemia | 87.8 | 85.2 | -0.07 | 85.2 | 85.5 | 0.01 | 86.7 | 87.1 | 0.01 |
| Hypertensive disorder | 88.4 | 86.0 | -0.07 | 87.1 | 86.8 | -0.01 | 88.0 | 88.1 | 0.00 |
| Lesion of liver | 1.6 | 1.5 | -0.01 | 1.7 | 1.5 | -0.01 | 1.7 | 1.4 | -0.03 |
| Obesity | 33.7 | 47.9 | 0.29 | 46.8 | 43.4 | -0.07 | 39.9 | 37.7 | -0.04 |
| Osteoarthritis | 33.9 | 40.2 | 0.13 | 39.5 | 37.7 | -0.04 | 36.7 | 36.2 | -0.01 |
| Pneumonia | 5.0 | 5.4 | 0.02 | 5.1 | 5.0 | 0.00 | 5.2 | 5.3 | 0.00 |
| Psoriasis | 2.2 | 2.7 | 0.04 | 2.4 | 2.8 | 0.03 | 2.3 | 2.3 | 0.01 |
| Renal impairment | 17.4 | 16.0 | -0.04 | 16.7 | 16.8 | 0.00 | 17.3 | 17.8 | 0.01 |
| Rheumatoid arthritis | 2.2 | 2.8 | 0.04 | 2.8 | 2.5 | -0.02 | 2.5 | 2.3 | -0.01 |
| Schizophrenia | 0.4 | 0.6 | 0.04 | 0.5 | 0.6 | 0.00 | 0.4 | 0.5 | 0.01 |
| Ulcerative colitis | 0.4 | 0.4 | 0.00 | 0.5 | 0.3 | -0.03 | 0.5 | 0.5 | 0.00 |
| Urinary tract infectious disease | 9.1 | 11.7 | 0.09 | 10.8 | 10.0 | -0.03 | 10.0 | 9.7 | -0.01 |
| Viral hepatitis C | 0.6 | 0.7 | 0.01 | 0.6 | 0.7 | 0.02 | 0.6 | 0.7 | 0.01 |
| Visual system disorder | 45.2 | 42.8 | -0.05 | 43.9 | 43.6 | -0.01 | 44.8 | 44.4 | -0.01 |
| **Medical history: Cardiovascular disease** | | | | | | | | | |
| Atrial fibrillation | 12.4 | 9.5 | -0.09 | 9.8 | 10.3 | 0.01 | 11.5 | 11.7 | 0.01 |
| Cerebrovascular disease | 13.3 | 11.3 | -0.06 | 11.6 | 11.4 | -0.01 | 12.7 | 12.4 | -0.01 |
| Coronary arteriosclerosis | 44.7 | 33.8 | -0.22 | 34.8 | 36.2 | 0.03 | 40.9 | 42.4 | 0.03 |
| Heart disease | 62.3 | 54.4 | -0.16 | 54.7 | 56.0 | 0.03 | 59.5 | 61.3 | 0.04 |
| Heart failure | 16.4 | 12.0 | -0.12 | 12.3 | 12.7 | 0.01 | 15.1 | 15.3 | 0.01 |
| Ischemic heart disease | 24.3 | 18.5 | -0.14 | 18.9 | 19.4 | 0.01 | 22.2 | 22.6 | 0.01 |
| Peripheral vascular disease | 27.0 | 26.7 | -0.01 | 27.2 | 27.5 | 0.01 | 27.2 | 26.9 | -0.01 |
| Pulmonary embolism | 1.3 | 1.7 | 0.03 | 1.3 | 1.5 | 0.02 | 1.3 | 1.4 | 0.00 |
| Venous thrombosis | 2.3 | 2.3 | 0.00 | 2.3 | 2.2 | -0.01 | 2.3 | 2.2 | -0.01 |
| **Medical history: Neoplasms** | | | | | | | | | |
| Hematologic neoplasm | 1.3 | 1.2 | -0.01 | 1.3 | 1.1 | -0.01 | 1.3 | 1.1 | -0.02 |
| Malignant lymphoma | 0.7 | 0.7 | 0.00 | 0.6 | 0.7 | 0.02 | 0.6 | 0.7 | 0.01 |
| Malignant neoplasm of anorectum | 0.2 | 0.2 | -0.01 | 0.2 | 0.2 | -0.01 | 0.2 | 0.2 | -0.01 |
| Malignant neoplastic disease | 12.3 | 10.9 | -0.04 | 11.6 | 11.2 | -0.01 | 11.9 | 11.8 | 0.00 |
| Malignant tumor of breast | 1.5 | 1.8 | 0.03 | 1.9 | 1.7 | -0.02 | 1.6 | 1.6 | 0.00 |
| Malignant tumor of colon | 0.6 | 0.7 | 0.01 | 0.5 | 0.6 | 0.02 | 0.6 | 0.6 | 0.00 |
| Malignant tumor of lung | 0.5 | 0.3 | -0.03 | 0.5 | 0.4 | -0.02 | 0.5 | 0.5 | 0.00 |
| Malignant tumor of urinary bladder | 0.5 | 0.5 | 0.00 | 0.3 | 0.4 | 0.02 | 0.4 | 0.4 | 0.01 |
| Primary malignant neoplasm of prostate | 2.8 | 1.8 | -0.07 | 2.1 | 2.1 | 0.01 | 2.5 | 2.6 | 0.01 |
| **Medication use** | | | | | | | | | |

| (Continued) | Before ajustment | | | After matching | | | After stratification | | |
|---|---|---|---|---|---|---|---|---|---|
| Characteristic | T (%) | C (%) | StdDiff | T (%) | C (%) | StdDiff | T (%) | C (%) | StdDiff |
| Agents acting on the renin-angiotensin system | 76.9 | 72.7 | -0.10 | 74.6 | 74.6 | 0.00 | 75.9 | 76.5 | 0.01 |
| Antibacterials for systemic use | 60.2 | 64.7 | 0.09 | 63.6 | 62.0 | -0.03 | 62.0 | 60.9 | -0.02 |
| Antidepressants | 31.8 | 43.0 | 0.23 | 41.6 | 39.2 | -0.05 | 36.2 | 34.8 | -0.03 |
| Antiepileptics | 22.5 | 29.3 | 0.16 | 28.3 | 27.2 | -0.03 | 25.2 | 24.7 | -0.01 |
| Antiinflammatory and antirheumatic products | 33.6 | 39.8 | 0.13 | 39.3 | 37.8 | -0.03 | 36.3 | 34.9 | -0.03 |
| Antineoplastic agents | 6.4 | 7.4 | 0.04 | 7.2 | 6.8 | -0.01 | 6.7 | 6.6 | -0.01 |
| Antipsoriatics | 0.5 | 0.9 | 0.04 | 0.6 | 0.8 | 0.02 | 0.6 | 0.8 | 0.03 |
| Antithrombotic agents | 35.2 | 27.4 | -0.17 | 28.5 | 29.1 | 0.01 | 32.6 | 33.3 | 0.01 |
| Beta blocking agents | 55.3 | 47.4 | -0.16 | 48.5 | 49.2 | 0.01 | 52.9 | 53.8 | 0.02 |
| Calcium channel blockers | 32.0 | 30.1 | -0.04 | 31.5 | 31.0 | -0.01 | 31.7 | 31.8 | 0.00 |
| Diuretics | 48.3 | 51.4 | 0.06 | 50.4 | 50.2 | 0.00 | 49.6 | 49.4 | 0.00 |
| Drugs for acid related disorders | 34.7 | 37.0 | 0.05 | 37.4 | 35.8 | -0.03 | 36.0 | 35.4 | -0.01 |
| Drugs for obstructive airway diseases | 43.5 | 50.7 | 0.14 | 49.5 | 48.6 | -0.02 | 46.5 | 46.1 | -0.01 |
| Immunosuppressants | 3.3 | 4.1 | 0.04 | 3.9 | 3.8 | -0.01 | 3.7 | 3.3 | -0.02 |
| Lipid modifying agents | 85.7 | 79.9 | -0.15 | 81.6 | 82.4 | 0.02 | 84.0 | 84.1 | 0.00 |
| Opioids | 32.8 | 36.3 | 0.07 | 35.2 | 34.0 | -0.03 | 34.2 | 32.9 | -0.03 |
| Psycholeptics | 29.8 | 34.1 | 0.09 | 33.1 | 31.2 | -0.04 | 31.6 | 30.0 | -0.03 |
| Psychostimulants | 2.1 | 3.8 | 0.10 | 2.8 | 2.9 | 0.01 | 2.5 | 2.7 | 0.01 |

**Supplementary Table 7:** Baseline patient characteristics for SGLT2is (T) and GLP1-RAs (C) new-users in the MDCR data source. We report proportion of initiators satisfying selected base-line characteristics and the standardized difference of population proportions (StdDiff) before and after propensity score adjustment. Less extreme StdDiffs through matching and stratification suggest improved balance between patient cohorts through adjustment.

| | Before adjustment | | | After matching | | | After stratification | | |
|---|---|---|---|---|---|---|---|---|---|
| Characteristic | T (%) | C (%) | StdDiff | T (%) | C (%) | StdDiff | T (%) | C (%) | StdDiff |
| **Age group** | | | | | | | | | |
| 50 - 54 | <0.2 | <0.4 | 0.03 | <0.2 | <0.7 | -0.01 | <0.3 | <0.5 | 0.02 |
| 55 - 59 | 1.0 | 1.1 | 0.01 | 1.2 | 1.1 | -0.01 | 1.1 | 0.7 | -0.04 |
| 60 - 64 | 2.1 | 2.5 | 0.02 | 2.4 | 1.7 | -0.04 | 2.2 | 2.3 | 0.01 |
| 65 - 69 | 27.8 | 34.6 | 0.15 | 29.1 | 32.3 | 0.07 | 28.2 | 31.3 | 0.07 |
| 70 - 74 | 36.9 | 36.8 | 0.00 | 36.7 | 36.3 | -0.01 | 36.8 | 35.5 | -0.03 |
| 75 - 79 | 21.3 | 16.2 | -0.13 | 20.4 | 16.4 | -0.10 | 21.0 | 17.7 | -0.08 |
| 80 - 84 | 7.5 | 6.1 | -0.06 | 7.1 | 8.5 | 0.05 | 7.3 | 8.1 | 0.03 |
| 85 - 89 | 2.6 | 2.0 | -0.04 | 2.3 | 2.7 | 0.03 | 2.5 | 3.3 | 0.04 |
| 90 - 94 | 0.5 | 0.6 | 0.01 | 0.5 | 0.9 | 0.06 | 0.5 | 1.0 | 0.05 |
| Gender: female | 36.1 | 49.2 | 0.27 | 48.9 | 45.7 | -0.06 | 39.8 | 38.0 | -0.04 |
| **Medical history: General** | | | | | | | | | |
| Acute respiratory disease | 26.0 | 25.8 | 0.00 | 28.5 | 26.2 | -0.05 | 26.9 | 25.0 | -0.04 |
| Attention deficit hyperactivity disorder | <0.2 | <0.4 | 0.01 | <0.2 | <0.7 | 0.01 | <0.3 | <0.5 | -0.01 |
| Chronic liver disease | 2.3 | 2.1 | -0.02 | 2.9 | 1.5 | -0.10 | 2.4 | 1.2 | -0.09 |
| Chronic obstructive lung disease | 14.2 | 15.7 | 0.04 | 16.3 | 15.3 | -0.03 | 16.3 | 15.3 | -0.03 |
| Crohn's disease | 0.5 | 0.5 | 0.00 | 0.4 | <0.7 | 0.00 | 0.5 | <0.5 | 0.01 |
| Dementia | 1.9 | 2.1 | 0.02 | 2.5 | 3.0 | 0.03 | 2.1 | 2.6 | 0.03 |
| Depressive disorder | 9.9 | 13.8 | 0.12 | 15.4 | 15.9 | 0.01 | 11.7 | 12.1 | 0.01 |
| Gastroesophageal reflux disease | 19.0 | 17.2 | -0.05 | 22.5 | 20.2 | -0.06 | 20.3 | 18.8 | -0.04 |
| Gastrointestinal hemorrhage | 3.8 | 2.9 | -0.05 | 3.8 | 2.2 | -0.10 | 3.8 | 2.2 | -0.09 |
| Human immunodeficiency virus infection | <0.2 | <0.4 | 0.00 | <0.2 | <0.7 | 0.01 | <0.3 | <0.5 | 0.02 |
| Hyperlipidemia | 84.8 | 69.0 | -0.38 | 84.6 | 83.2 | -0.04 | 84.8 | 83.1 | -0.05 |
| Hypertensive disorder | 87.7 | 81.4 | -0.17 | 88.1 | 88.6 | 0.01 | 87.8 | 88.9 | 0.03 |
| Lesion of liver | 1.7 | 0.9 | -0.07 | 1.6 | 0.9 | -0.06 | 1.7 | 0.8 | -0.08 |
| Obesity | 25.9 | 29.5 | 0.08 | 39.9 | 36.4 | -0.07 | 30.3 | 27.3 | -0.07 |
| Osteoarthritis | 36.1 | 39.4 | 0.07 | 41.1 | 41.9 | 0.02 | 37.5 | 37.9 | 0.01 |
| Pneumonia | 5.0 | 4.8 | -0.01 | 5.5 | 5.0 | -0.02 | 5.2 | 4.7 | -0.02 |
| Psoriasis | 2.5 | 2.9 | 0.03 | 2.4 | 3.5 | 0.07 | 2.5 | 3.8 | 0.08 |
| Renal impairment | 14.5 | 14.5 | 0.00 | 16.0 | 17.9 | 0.05 | 15.0 | 16.9 | 0.05 |
| Rheumatoid arthritis | 1.7 | 1.9 | 0.01 | 2.1 | 2.3 | 0.01 | 1.8 | 1.7 | 0.00 |
| Ulcerative colitis | 0.5 | <0.4 | -0.02 | 0.6 | <0.7 | -0.01 | 0.5 | 0.7 | 0.02 |
| Urinary tract infectious disease | 8.1 | 11.5 | 0.12 | 9.8 | 11.8 | 0.06 | 8.6 | 10.3 | 0.06 |
| Viral hepatitis C | 0.6 | <0.4 | -0.09 | 0.5 | <0.7 | -0.07 | 0.6 | <0.5 | -0.07 |
| Visual system disorder | 57.8 | 51.6 | -0.12 | 56.8 | 55.1 | -0.04 | 57.5 | 55.2 | -0.04 |
| **Medical history: Cardiovascular disease** | | | | | | | | | |
| Atrial fibrillation | 16.7 | 12.1 | -0.13 | 15.1 | 12.9 | -0.06 | 16.2 | 14.3 | -0.05 |
| Cerebrovascular disease | 18.9 | 17.9 | -0.03 | 18.6 | 17.9 | -0.02 | 18.7 | 17.6 | -0.03 |
| Coronary arteriosclerosis | 51.9 | 43.1 | -0.18 | 43.1 | 45.6 | 0.05 | 49.4 | 51.0 | 0.03 |
| Heart disease | 71.2 | 62.3 | -0.19 | 64.2 | 63.8 | -0.01 | 69.2 | 68.7 | -0.01 |
| Heart failure | 15.3 | 11.3 | -0.12 | 15.7 | 12.1 | -0.10 | 15.5 | 12.5 | -0.09 |
| Ischemic heart disease | 21.9 | 17.9 | -0.10 | 19.7 | 17.7 | -0.05 | 21.3 | 18.7 | -0.06 |
| Peripheral vascular disease | 24.9 | 22.5 | -0.06 | 26.6 | 23.4 | -0.07 | 25.5 | 22.6 | -0.07 |
| Pulmonary embolism | 1.0 | 1.3 | 0.02 | 1.2 | 1.3 | 0.01 | 1.1 | 1.2 | 0.01 |
| Venous thrombosis | 2.2 | 2.1 | -0.01 | 2.4 | 1.9 | -0.04 | 2.3 | 1.6 | -0.05 |
| **Medical history: Neoplasms** | | | | | | | | | |
| Hematologic neoplasm | 1.7 | 2.1 | 0.03 | 1.4 | 2.7 | 0.09 | 1.6 | 2.4 | 0.06 |
| Malignant lymphoma | 1.2 | 1.2 | 0.00 | 1.0 | 1.3 | 0.03 | 1.2 | 1.2 | 0.00 |
| Malignant neoplastic disease | 19.2 | 20.5 | 0.03 | 18.7 | 21.6 | 0.07 | 19.0 | 22.4 | 0.09 |
| Malignant tumor of breast | 2.2 | 3.8 | 0.09 | 3.3 | 3.6 | 0.02 | 2.5 | 3.1 | 0.04 |
| Malignant tumor of colon | 0.7 | 0.5 | -0.02 | 0.8 | <0.7 | -0.03 | 0.7 | 0.6 | -0.01 |
| Malignant tumor of lung | 0.4 | 0.8 | 0.04 | 0.4 | 1.1 | 0.09 | 0.4 | 1.1 | 0.08 |
| Malignant tumor of urinary bladder | 1.2 | 1.8 | 0.04 | 0.9 | 2.2 | 0.10 | 1.2 | 2.3 | 0.09 |
| Primary malignant neoplasm of prostate | 5.4 | 4.6 | -0.04 | 4.6 | 4.8 | 0.01 | 5.2 | 5.7 | 0.02 |
| **Medication use** | | | | | | | | | |
| Agents acting on the renin-angiotensin system | 77.6 | 78.2 | 0.01 | 76.6 | 76.9 | 0.01 | 77.2 | 77.7 | 0.01 |
| Antibacterials for systemic use | 64.8 | 66.7 | 0.04 | 67.5 | 66.0 | -0.03 | 65.6 | 64.4 | -0.03 |
| Antidepressants | 27.0 | 36.6 | 0.21 | 38.3 | 34.5 | -0.08 | 30.6 | 27.3 | -0.07 |
| Antiepileptics | 19.7 | 21.2 | 0.04 | 24.4 | 21.2 | -0.07 | 21.3 | 19.0 | -0.06 |
| Antiinflammatory and antirheumatic products | 29.9 | 34.6 | 0.10 | 34.1 | 34.0 | 0.00 | 31.2 | 30.4 | -0.02 |
| Antineoplastic agents | 8.4 | 10.1 | 0.06 | 9.3 | 9.9 | 0.02 | 8.7 | 10.3 | 0.06 |
| Antipsoriatics | 0.9 | 1.2 | 0.02 | 0.7 | 1.1 | 0.04 | 0.9 | 1.1 | 0.02 |

| (Continued) | Before ajustment | | | After matching | | | After stratification | | |
|---|---|---|---|---|---|---|---|---|---|
| Characteristic | T (%) | C (%) | StdDiff | T (%) | C (%) | StdDiff | T (%) | C (%) | StdDiff |
| Antithrombotic agents | 38.5 | 35.1 | -0.07 | 35.6 | 33.9 | -0.04 | 37.7 | 36.9 | -0.02 |
| Beta blocking agents | 61.7 | 55.9 | -0.12 | 55.5 | 57.7 | 0.04 | 59.8 | 62.1 | 0.05 |
| Calcium channel blockers | 35.9 | 33.7 | -0.05 | 35.8 | 32.1 | -0.08 | 36.0 | 32.7 | -0.07 |
| Diuretics | 53.1 | 56.4 | 0.07 | 55.3 | 52.6 | -0.06 | 53.7 | 50.8 | -0.06 |
| Drugs for acid related disorders | 39.1 | 39.9 | 0.02 | 43.2 | 39.0 | -0.09 | 40.5 | 37.8 | -0.05 |
| Drugs for obstructive airway diseases | 48.5 | 53.1 | 0.09 | 53.1 | 53.2 | 0.00 | 53.1 | 53.2 | 0.00 |
| Immunosuppressants | 3.9 | 4.1 | 0.01 | 4.4 | 5.0 | 0.03 | 4.0 | 4.5 | 0.02 |
| Lipid modifying agents | 89.2 | 86.7 | -0.08 | 86.4 | 87.5 | 0.03 | 88.3 | 87.7 | -0.02 |
| Opioids | 28.9 | 32.1 | 0.07 | 32.9 | 31.7 | -0.03 | 30.1 | 30.4 | 0.01 |
| Psycholeptics | 26.0 | 31.2 | 0.11 | 30.4 | 28.2 | -0.05 | 27.5 | 25.8 | -0.04 |
| Psychostimulants | 1.6 | 2.1 | 0.04 | 2.4 | 1.7 | -0.04 | 1.9 | 1.6 | -0.02 |

**Supplementary Table 8:** Baseline patient characteristics for SGLT2is (T) and GLP1-RAs (C) new-users in the Open Claims data source. We report proportion of initiators satisfying selected base-line characteristics and the standardized difference of population proportions (StdDiff) before and after propensity score adjustment. Less extreme StdDiffs through matching and stratification suggest improved balance between patient cohorts through adjustment.

| | Before adjustment | | | After matching | | | After stratification | | |
|---|---|---|---|---|---|---|---|---|---|
| Characteristic | T (%) | C (%) | StdDiff | T (%) | C (%) | StdDiff | T (%) | C (%) | StdDiff |
| **Age group** | | | | | | | | | |
| 20 - 24 | 0.1 | 0.2 | 0.02 | 0.2 | 0.2 | 0.00 | 0.1 | 0.2 | 0.00 |
| 25 - 29 | 0.2 | 0.3 | 0.03 | 0.2 | 0.3 | 0.01 | 0.2 | 0.3 | 0.01 |
| 30 - 34 | 0.4 | 0.8 | 0.05 | 0.6 | 0.6 | 0.00 | 0.5 | 0.6 | 0.01 |
| 35 - 39 | 1.0 | 1.7 | 0.06 | 1.4 | 1.5 | 0.00 | 1.2 | 1.3 | 0.01 |
| 40 - 44 | 2.4 | 3.8 | 0.08 | 2.9 | 3.0 | 0.00 | 2.9 | 3.0 | 0.00 |
| 45 - 49 | 5.2 | 7.1 | 0.08 | 6.6 | 6.4 | -0.01 | 5.9 | 5.9 | 0.00 |
| 50 - 54 | 9.4 | 11.9 | 0.08 | 11.1 | 11.0 | 0.00 | 11.1 | 11.0 | 0.00 |
| 55 - 59 | 14.5 | 16.5 | 0.06 | 16.3 | 16.0 | -0.01 | 15.3 | 15.1 | 0.00 |
| 60 - 64 | 18.0 | 18.3 | 0.01 | 18.7 | 18.5 | -0.01 | 18.2 | 17.9 | -0.01 |
| 65 - 69 | 17.5 | 16.7 | -0.02 | 17.1 | 17.4 | 0.01 | 17.2 | 17.4 | 0.01 |
| 70 - 74 | 14.4 | 12.2 | -0.06 | 12.9 | 13.1 | 0.01 | 13.6 | 13.7 | 0.00 |
| 75 - 79 | 10.0 | 6.8 | -0.12 | 7.5 | 7.6 | 0.00 | 8.9 | 8.9 | 0.00 |
| 80 - 84 | 6.1 | 3.3 | -0.13 | 3.8 | 3.8 | 0.00 | 5.2 | 5.1 | 0.00 |
| 85 - 89 | 0.5 | 0.3 | -0.04 | 0.3 | 0.3 | 0.00 | 0.4 | 0.4 | -0.01 |
| Gender: female | 39.9 | 54.4 | 0.29 | 50.7 | 50.0 | -0.01 | 44.9 | 44.6 | -0.01 |
| **Medical history: General** | | | | | | | | | |
| Acute respiratory disease | 15.6 | 17.1 | 0.04 | 16.3 | 16.1 | 0.00 | 16.3 | 16.1 | 0.00 |
| Attention deficit hyperactivity disorder | 0.5 | 0.9 | 0.05 | 0.7 | 0.7 | 0.00 | 0.6 | 0.6 | 0.00 |
| Chronic liver disease | 1.3 | 1.1 | -0.01 | 1.2 | 1.1 | -0.01 | 1.3 | 1.2 | 0.00 |
| Crohn’s disease | 0.2 | 0.2 | 0.00 | 0.2 | 0.2 | -0.01 | 0.2 | 0.2 | -0.01 |
| Dementia | 0.9 | 0.7 | -0.02 | 0.8 | 0.8 | 0.00 | 0.9 | 0.9 | 0.00 |
| Depressive disorder | 8.0 | 12.0 | 0.13 | 10.6 | 10.2 | -0.01 | 9.5 | 9.4 | 0.00 |
| Gastroesophageal reflux disease | 10.6 | 12.7 | 0.06 | 11.8 | 11.5 | -0.01 | 11.4 | 11.3 | 0.00 |
| Gastrointestinal hemorrhage | 1.8 | 1.8 | 0.00 | 1.7 | 1.7 | 0.00 | 1.7 | 1.7 | 0.00 |
| Human immunodeficiency virus infection | 0.3 | 0.4 | 0.01 | 0.3 | 0.3 | 0.00 | 0.3 | 0.3 | 0.00 |
| Hyperlipidemia | 53.2 | 50.1 | -0.06 | 50.4 | 50.2 | 0.00 | 52.3 | 52.3 | 0.00 |
| Hypertensive disorder | 60.7 | 59.4 | -0.03 | 59.1 | 58.9 | -0.01 | 60.4 | 60.6 | 0.00 |
| Lesion of liver | 0.8 | 0.9 | 0.00 | 0.9 | 0.8 | -0.01 | 0.9 | 0.8 | -0.01 |
| Obesity | 13.9 | 23.1 | 0.24 | 19.5 | 18.7 | -0.02 | 17.2 | 17.2 | 0.00 |
| Osteoarthritis | 19.0 | 23.6 | 0.11 | 21.9 | 21.6 | -0.01 | 20.6 | 20.8 | 0.00 |
| Pneumonia | 3.4 | 3.4 | 0.00 | 3.3 | 3.2 | 0.00 | 3.4 | 3.3 | 0.00 |
| Psoriasis | 1.2 | 1.5 | 0.03 | 1.3 | 1.3 | 0.00 | 1.3 | 1.3 | 0.00 |
| Renal impairment | 9.5 | 8.2 | -0.04 | 8.6 | 8.3 | -0.01 | 9.3 | 9.0 | -0.01 |
| Rheumatoid arthritis | 1.3 | 1.8 | 0.04 | 1.6 | 1.5 | 0.00 | 1.5 | 1.5 | 0.00 |
| Schizophrenia | 0.3 | 0.3 | 0.00 | 0.4 | 0.3 | -0.01 | 0.3 | 0.3 | -0.01 |
| Ulcerative colitis | 0.3 | 0.2 | 0.00 | 0.3 | 0.2 | -0.01 | 0.3 | 0.2 | -0.01 |
| Urinary tract infectious disease | 5.2 | 6.5 | 0.06 | 6.0 | 5.9 | 0.00 | 5.7 | 5.7 | 0.00 |
| Viral hepatitis C | 0.4 | 0.3 | -0.01 | 0.4 | 0.3 | -0.01 | 0.4 | 0.3 | 0.00 |
| Visual system disorder | 21.8 | 21.3 | -0.01 | 21.1 | 21.1 | 0.00 | 21.7 | 21.6 | 0.00 |
| **Medical history: Cardiovascular disease** | | | | | | | | | |
| Atrial fibrillation | 8.4 | 6.1 | -0.09 | 7.7 | 7.5 | -0.01 | 7.7 | 7.5 | -0.01 |
| Cerebrovascular disease | 7.5 | 6.4 | -0.04 | 6.6 | 6.5 | 0.00 | 7.1 | 7.1 | 0.00 |
| Coronary arteriosclerosis | 28.2 | 20.2 | -0.19 | 21.4 | 21.5 | 0.00 | 21.4 | 21.5 | 0.00 |
| Heart disease | 44.2 | 35.4 | -0.18 | 36.6 | 36.5 | 0.00 | 41.3 | 41.2 | 0.00 |
| Heart failure | 11.3 | 6.8 | -0.16 | 7.4 | 7.1 | -0.01 | 10.0 | 9.1 | -0.03 |
| Ischemic heart disease | 13.9 | 9.7 | -0.13 | 10.3 | 10.2 | 0.00 | 12.5 | 12.2 | -0.01 |
| Pulmonary embolism | 0.8 | 1.1 | 0.03 | 0.9 | 0.9 | 0.00 | 0.9 | 0.9 | 0.00 |
| Venous thrombosis | 1.3 | 1.2 | 0.00 | 1.2 | 1.2 | 0.00 | 1.3 | 1.2 | 0.00 |
| **Medical history: Neoplasms** | | | | | | | | | |
| Hematologic neoplasm | 0.8 | 0.8 | 0.00 | 0.8 | 0.8 | 0.00 | 0.8 | 0.8 | 0.00 |
| Malignant lymphoma | 0.5 | 0.4 | -0.01 | 0.5 | 0.4 | -0.01 | 0.5 | 0.5 | 0.00 |
| Malignant neoplasm of anorectum | 0.1 | 0.1 | -0.02 | 0.1 | 0.1 | -0.02 | 0.1 | 0.1 | -0.01 |
| Malignant neoplastic disease | 7.5 | 6.9 | -0.02 | 7.0 | 6.9 | 0.00 | 7.3 | 7.2 | 0.00 |
| Malignant tumor of breast | 1.1 | 1.3 | 0.02 | 1.3 | 1.3 | 0.00 | 1.2 | 1.2 | 0.00 |
| Malignant tumor of colon | 0.3 | 0.3 | 0.00 | 0.3 | 0.3 | 0.00 | 0.3 | 0.3 | 0.00 |
| Malignant tumor of lung | 0.3 | 0.3 | -0.01 | 0.3 | 0.3 | 0.00 | 0.3 | 0.3 | 0.00 |
| Malignant tumor of urinary bladder | 0.3 | 0.3 | 0.00 | 0.3 | 0.3 | 0.00 | 0.3 | 0.4 | 0.01 |
| Primary malignant neoplasm of prostate | 1.6 | 1.2 | -0.04 | 1.3 | 1.3 | 0.00 | 1.5 | 1.5 | 0.00 |
| **Medication use** | | | | | | | | | |
| Agents acting on the renin-angiotensin system | 74.6 | 68.7 | -0.13 | 70.7 | 70.6 | 0.00 | 72.9 | 72.7 | 0.00 |
| Antibacterials for systemic use | 58.9 | 63.4 | 0.09 | 62.4 | 61.5 | -0.02 | 60.7 | 60.4 | -0.01 |

| (Continued) | Before ajustment | | | After matching | | | After stratification | | |
|---|---|---|---|---|---|---|---|---|---|
| Characteristic | T (%) | C (%) | StdDiff | T (%) | C (%) | StdDiff | T (%) | C (%) | StdDiff |
| Antidepressants | 32.1 | 42.9 | 0.22 | 40.3 | 39.2 | -0.02 | 36.1 | 35.9 | 0.00 |
| Antiepileptics | 23.4 | 30.0 | 0.15 | 28.1 | 27.6 | -0.01 | 25.9 | 25.9 | 0.00 |
| Antiinflammatory and antirheumatic products | 39.5 | 43.4 | 0.08 | 42.0 | 41.6 | -0.01 | 40.9 | 40.6 | 0.00 |
| Antineoplastic agents | 5.9 | 7.2 | 0.05 | 6.7 | 6.5 | -0.01 | 6.3 | 6.2 | 0.00 |
| Antipsoriatics | 0.8 | 1.1 | 0.03 | 1.0 | 1.0 | 0.00 | 0.9 | 0.9 | 0.00 |
| Antithrombotic agents | 38.2 | 29.8 | -0.18 | 31.1 | 31.2 | 0.00 | 35.4 | 35.4 | 0.00 |
| Beta blocking agents | 55.8 | 47.1 | -0.18 | 48.8 | 48.7 | 0.00 | 52.9 | 52.9 | 0.00 |
| Calcium channel blockers | 31.9 | 30.4 | -0.03 | 31.1 | 31.2 | 0.00 | 31.4 | 31.7 | 0.00 |
| Diuretics | 47.8 | 49.1 | 0.02 | 48.8 | 48.2 | -0.01 | 48.6 | 47.9 | -0.01 |
| Drugs for acid related disorders | 40.6 | 43.3 | 0.06 | 42.4 | 42.0 | -0.01 | 41.7 | 41.6 | 0.00 |
| Drugs for obstructive airway diseases | 42.7 | 48.7 | 0.12 | 47.2 | 46.5 | -0.01 | 45.0 | 44.7 | -0.01 |
| Immunosuppressants | 3.4 | 4.4 | 0.05 | 4.0 | 3.9 | 0.00 | 3.7 | 3.7 | 0.00 |
| Lipid modifying agents | 82.5 | 76.1 | -0.16 | 78.2 | 78.3 | 0.00 | 80.5 | 80.4 | 0.00 |
| Opioids | 28.2 | 31.9 | 0.08 | 30.5 | 30.3 | -0.01 | 29.6 | 29.8 | 0.00 |
| Psycholeptics | 26.2 | 31.8 | 0.12 | 30.1 | 29.5 | -0.01 | 28.3 | 28.2 | 0.00 |
| Psychostimulants | 2.7 | 4.7 | 0.11 | 3.9 | 3.7 | -0.01 | 3.4 | 3.4 | 0.00 |

**Supplementary Table 9:** Baseline patient characteristics for SGLT2is (T) and GLP1-RAs (C) new-users in the OptumEHR data source. We report proportion of initiators satisfying selected base-line characteristics and the standardized difference of population proportions (StdDiff) before and after propensity score adjustment. Less extreme StdDiffs through matching and stratification suggest improved balance between patient cohorts through adjustment.

| | Before adjustment | | | After matching | | | After stratification | | |
|---|---|---|---|---|---|---|---|---|---|
| Characteristic | T (%) | C (%) | StdDiff | T (%) | C (%) | StdDiff | T (%) | C (%) | StdDiff |
| **Age group** | | | | | | | | | |
| 15 - 19 | <0.1 | <0.1 | 0.01 | <0.1 | <0.1 | 0.00 | <0.1 | <0.1 | 0.00 |
| 20 - 24 | <0.1 | 0.1 | 0.02 | 0.1 | <0.1 | -0.02 | 0.1 | <0.1 | -0.02 |
| 25 - 29 | 0.1 | 0.4 | 0.06 | 0.1 | 0.4 | 0.05 | 0.1 | 0.2 | 0.03 |
| 30 - 34 | 0.4 | 0.6 | 0.03 | 0.6 | 0.5 | -0.01 | 0.5 | 0.5 | 0.00 |
| 35 - 39 | 1.2 | 2.0 | 0.07 | 1.5 | 1.8 | 0.03 | 1.3 | 1.6 | 0.02 |
| 40 - 44 | 3.3 | 4.8 | 0.08 | 4.1 | 4.1 | 0.00 | 3.7 | 3.8 | 0.00 |
| 45 - 49 | 6.6 | 8.3 | 0.06 | 7.5 | 7.9 | 0.01 | 7.0 | 7.5 | 0.02 |
| 50 - 54 | 10.8 | 14.3 | 0.11 | 13.0 | 12.8 | 0.00 | 11.8 | 12.0 | 0.01 |
| 55 - 59 | 17.1 | 17.6 | 0.01 | 17.6 | 17.9 | 0.01 | 17.3 | 17.5 | 0.00 |
| 60 - 64 | 20.8 | 19.8 | -0.02 | 20.3 | 20.3 | 0.00 | 20.5 | 20.3 | 0.00 |
| 65 - 69 | 16.2 | 15.2 | -0.03 | 15.7 | 16.0 | 0.01 | 15.8 | 16.7 | 0.02 |
| 70 - 74 | 12.5 | 9.7 | -0.09 | 10.8 | 10.5 | -0.01 | 11.8 | 11.6 | -0.01 |
| 75 - 79 | 6.8 | 5.0 | -0.08 | 5.6 | 5.3 | -0.01 | 6.3 | 5.7 | -0.03 |
| 80 - 84 | 3.1 | 1.6 | -0.10 | 2.3 | 1.8 | -0.03 | 2.8 | 2.0 | -0.05 |
| 85 - 89 | 1.0 | 0.4 | -0.07 | 0.9 | 0.5 | -0.05 | 1.0 | 0.6 | -0.05 |
| Gender: female | 35.4 | 50.2 | 0.30 | 49.0 | 45.8 | -0.06 | 40.9 | 38.1 | -0.06 |
| **Race** | | | | | | | | | |
| Asian | 1.9 | 1.1 | -0.06 | 1.4 | 1.3 | -0.01 | 1.7 | 1.4 | -0.03 |
| Black or African American | 8.1 | 10.1 | 0.07 | 9.4 | 9.8 | 0.01 | 8.6 | 8.7 | 0.00 |
| White | 83.7 | 83.6 | 0.00 | 83.1 | 83.8 | 0.02 | 83.4 | 84.7 | 0.03 |
| **Ethnicity** | | | | | | | | | |
| Hispanic or Latino | 4.5 | 4.0 | -0.02 | 4.5 | 3.8 | -0.04 | 4.6 | 4.0 | -0.03 |
| Not Hispanic or Latino | 86.3 | 88.3 | 0.06 | 87.8 | 88.0 | 0.01 | 86.8 | 87.6 | 0.03 |
| **Medical history: General** | | | | | | | | | |
| Acute respiratory disease | 19.3 | 21.3 | 0.05 | 21.7 | 20.2 | -0.04 | 20.3 | 19.0 | -0.03 |
| Attention deficit hyperactivity disorder | 0.6 | 1.0 | 0.04 | 1.0 | 0.9 | -0.01 | 0.8 | 0.8 | 0.00 |
| Chronic liver disease | 2.2 | 2.7 | 0.03 | 2.6 | 2.5 | 0.00 | 2.4 | 2.5 | 0.01 |
| Chronic obstructive lung disease | 9.9 | 10.2 | 0.01 | 10.7 | 9.9 | -0.03 | 10.3 | 9.9 | -0.01 |
| Crohn’s disease | 0.3 | 0.4 | 0.02 | 0.3 | 0.4 | 0.01 | 0.3 | 0.4 | 0.01 |
| Dementia | 0.8 | 0.9 | 0.01 | 0.8 | 0.8 | 0.00 | 0.8 | 0.8 | 0.00 |
| Depressive disorder | 14.7 | 21.4 | 0.18 | 20.7 | 18.5 | -0.05 | 17.4 | 16.3 | -0.03 |
| Gastroesophageal reflux disease | 21.8 | 24.8 | 0.07 | 24.7 | 22.6 | -0.05 | 23.0 | 21.9 | -0.03 |
| Gastrointestinal hemorrhage | 1.6 | 2.0 | 0.03 | 1.7 | 1.9 | 0.01 | 1.6 | 1.9 | 0.02 |
| Human immunodeficiency virus infection | 0.2 | 0.2 | 0.01 | 0.2 | 0.3 | 0.02 | 0.2 | 0.2 | 0.00 |
| Hyperlipidemia | 78.1 | 74.6 | -0.08 | 73.9 | 75.1 | 0.03 | 76.5 | 77.1 | 0.01 |
| Hypertensive disorder | 78.9 | 77.5 | -0.03 | 77.1 | 77.0 | 0.00 | 78.4 | 78.5 | 0.00 |
| Lesion of liver | 0.9 | 1.1 | 0.02 | 1.0 | 1.1 | 0.00 | 0.9 | 1.0 | 0.01 |
| Obesity | 33.2 | 47.5 | 0.29 | 46.1 | 42.8 | -0.06 | 39.0 | 36.6 | -0.05 |
| Osteoarthritis | 20.9 | 25.3 | 0.10 | 24.6 | 23.9 | -0.02 | 22.7 | 22.0 | -0.02 |
| Pneumonia | 2.4 | 3.0 | 0.04 | 2.7 | 2.9 | 0.01 | 2.6 | 2.8 | 0.01 |
| Psoriasis | 1.7 | 2.3 | 0.04 | 1.9 | 2.0 | 0.00 | 1.8 | 2.2 | 0.03 |
| Renal impairment | 8.7 | 9.1 | 0.01 | 9.3 | 8.8 | -0.02 | 9.0 | 8.8 | 0.00 |
| Rheumatoid arthritis | 1.6 | 1.8 | 0.01 | 2.1 | 1.6 | -0.04 | 1.9 | 1.4 | -0.04 |
| Schizophrenia | 0.2 | 0.3 | 0.03 | 0.2 | 0.2 | 0.00 | 0.2 | 0.3 | 0.01 |
| Ulcerative colitis | 0.3 | 0.4 | 0.00 | 0.3 | 0.4 | 0.02 | 0.3 | 0.3 | 0.00 |
| Urinary tract infectious disease | 3.9 | 5.2 | 0.06 | 5.1 | 4.5 | -0.02 | 4.4 | 4.3 | 0.00 |
| Viral hepatitis C | 0.4 | 0.8 | 0.05 | 0.5 | 0.9 | 0.05 | 0.5 | 0.8 | 0.05 |
| Visual system disorder | 13.0 | 15.0 | 0.06 | 14.0 | 14.4 | 0.01 | 13.6 | 13.7 | 0.00 |
| **Medical history: Cardiovascular disease** | | | | | | | | | |
| Atrial fibrillation | 8.8 | 7.5 | -0.05 | 7.5 | 7.9 | 0.01 | 8.4 | 8.6 | 0.01 |
| Cerebrovascular disease | 9.1 | 8.6 | -0.02 | 8.9 | 8.4 | -0.02 | 8.9 | 8.4 | -0.02 |
| Coronary arteriosclerosis | 47.4 | 34.5 | -0.26 | 34.8 | 37.3 | 0.05 | 42.4 | 44.9 | 0.05 |
| Heart disease | 61.8 | 52.6 | -0.19 | 51.6 | 54.3 | 0.05 | 58.0 | 60.6 | 0.05 |
| Heart failure | 9.9 | 7.8 | -0.07 | 9.3 | 9.3 | 0.00 | 9.3 | 9.3 | 0.00 |
| Ischemic heart disease | 22.0 | 17.8 | -0.11 | 17.6 | 18.2 | 0.01 | 20.4 | 21.0 | 0.02 |
| Peripheral vascular disease | 20.3 | 20.9 | 0.01 | 21.7 | 20.7 | -0.03 | 20.9 | 19.7 | -0.03 |
| Pulmonary embolism | 0.9 | 1.1 | 0.03 | 1.1 | 1.1 | 0.00 | 1.1 | 1.1 | 0.00 |
| Venous thrombosis | 1.8 | 1.8 | 0.00 | 2.0 | 1.7 | -0.03 | 1.8 | 1.7 | -0.01 |
| **Medical history: Neoplasms** | | | | | | | | | |
| Hematologic neoplasm | 0.9 | 0.6 | -0.03 | 1.0 | 0.6 | -0.04 | 1.0 | 0.6 | -0.04 |
| Malignant lymphoma | 0.5 | 0.5 | 0.01 | 0.5 | 0.6 | 0.01 | 0.5 | 0.5 | 0.00 |

| (Continued) | Before ajustment | | | After matching | | | After stratification | | |
|---|---|---|---|---|---|---|---|---|---|
| Characteristic | T (%) | C (%) | StdDiff | T (%) | C (%) | StdDiff | T (%) | C (%) | StdDiff |
| Malignant neoplasm of anorectum | 0.1 | 0.2 | 0.03 | 0.1 | 0.2 | 0.02 | 0.1 | 0.1 | 0.02 |
| Malignant neoplastic disease | 7.1 | 7.2 | 0.00 | 7.5 | 7.3 | -0.01 | 7.3 | 6.9 | -0.02 |
| Malignant tumor of breast | 1.0 | 1.4 | 0.03 | 1.4 | 1.3 | -0.01 | 1.2 | 1.0 | -0.02 |
| Malignant tumor of colon | 0.4 | 0.2 | -0.03 | 0.4 | 0.2 | -0.04 | 0.4 | 0.2 | -0.04 |
| Malignant tumor of lung | 0.2 | 0.1 | -0.02 | 0.2 | 0.2 | -0.02 | 0.2 | 0.1 | -0.03 |
| Malignant tumor of urinary bladder | 0.4 | 0.3 | 0.00 | 0.3 | 0.4 | 0.01 | 0.4 | 0.5 | 0.01 |
| Primary malignant neoplasm of prostate | 1.7 | 1.0 | -0.06 | 1.3 | 1.2 | -0.01 | 1.6 | 1.4 | -0.01 |
| **Medication use** | | | | | | | | | |
| Agents acting on the renin-angiotensin system | 75.2 | 71.3 | -0.09 | 72.3 | 72.1 | 0.00 | 72.3 | 72.1 | 0.00 |
| Antibacterials for systemic use | 45.0 | 50.3 | 0.11 | 49.8 | 47.9 | -0.04 | 47.1 | 46.2 | -0.02 |
| Antidepressants | 31.9 | 42.5 | 0.22 | 41.5 | 38.8 | -0.05 | 36.0 | 34.4 | -0.03 |
| Antiepileptics | 21.1 | 26.5 | 0.13 | 25.9 | 24.0 | -0.04 | 23.3 | 22.1 | -0.03 |
| Antiinflammatory and antirheumatic products | 71.6 | 70.5 | -0.03 | 70.4 | 69.8 | -0.01 | 71.4 | 71.2 | 0.00 |
| Antineoplastic agents | 4.9 | 6.1 | 0.05 | 5.9 | 5.7 | -0.01 | 5.9 | 5.7 | -0.01 |
| Antipsoriatics | 0.4 | 0.8 | 0.06 | 0.5 | 0.8 | 0.04 | 0.4 | 0.8 | 0.05 |
| Antithrombotic agents | 68.3 | 61.2 | -0.15 | 61.7 | 62.7 | 0.02 | 65.7 | 66.8 | 0.02 |
| Beta blocking agents | 58.9 | 51.7 | -0.14 | 52.4 | 53.2 | 0.01 | 56.3 | 57.0 | 0.01 |
| Calcium channel blockers | 30.2 | 29.3 | -0.02 | 30.0 | 29.3 | -0.02 | 30.0 | 29.0 | -0.02 |
| Diuretics | 45.2 | 50.4 | 0.10 | 49.6 | 48.8 | -0.02 | 47.0 | 46.5 | -0.01 |
| Drugs for acid related disorders | 46.4 | 49.0 | 0.05 | 49.8 | 46.9 | -0.06 | 47.7 | 46.5 | -0.02 |
| Drugs for obstructive airway diseases | 46.3 | 54.1 | 0.16 | 53.2 | 51.5 | -0.04 | 49.4 | 47.4 | -0.04 |
| Immunosuppressants | 3.1 | 4.4 | 0.07 | 3.8 | 4.2 | 0.02 | 3.5 | 3.9 | 0.02 |
| Lipid modifying agents | 86.7 | 81.5 | -0.14 | 82.4 | 83.2 | 0.02 | 84.8 | 85.9 | 0.03 |
| Opioids | 31.7 | 36.6 | 0.10 | 35.1 | 34.7 | -0.01 | 33.3 | 32.9 | -0.01 |
| Psycholeptics | 33.4 | 39.4 | 0.12 | 37.6 | 36.9 | -0.02 | 35.5 | 34.7 | -0.02 |
| Psychostimulants | 3.4 | 5.1 | 0.09 | 4.8 | 4.7 | 0.00 | 4.0 | 4.0 | 0.00 |

**Supplementary Table 10:** Baseline patient characteristics for SGLT2is (T) and DPP4is (C) new-users in the CCAE data source. We report proportion of initiators satisfying selected base-line characteristics and the standardized difference of population proportions (StdDiff) before and after propensity score adjustment. Less extreme StdDiffs through matching and stratification suggest improved balance between patient cohorts through adjustment.

| | Before adjustment | | | After matching | | | After stratification | | |
|---|---|---|---|---|---|---|---|---|---|
| Characteristic | T (%) | C (%) | StdDiff | T (%) | C (%) | StdDiff | T (%) | C (%) | StdDiff |
| **Age group** | | | | | | | | | |
| 25 - 29 | 0.1 | 0.1 | 0.02 | 0.1 | 0.1 | 0.02 | 0.1 | 0.1 | 0.02 |
| 30 - 34 | 0.3 | 0.5 | 0.03 | 0.3 | 0.3 | 0.00 | 0.3 | 0.4 | 0.01 |
| 35 - 39 | 1.7 | 1.5 | -0.01 | 1.7 | 1.5 | -0.02 | 1.6 | 1.4 | -0.02 |
| 40 - 44 | 4.5 | 4.3 | -0.01 | 4.7 | 4.1 | -0.03 | 4.7 | 4.3 | -0.02 |
| 45 - 49 | 10.1 | 9.3 | -0.03 | 9.9 | 9.8 | -0.01 | 10.0 | 9.5 | -0.02 |
| 50 - 54 | 18.3 | 18.1 | 0.00 | 18.5 | 18.1 | -0.01 | 18.7 | 18.1 | -0.01 |
| 55 - 59 | 29.2 | 27.6 | -0.04 | 28.6 | 28.9 | 0.01 | 27.9 | 27.9 | 0.00 |
| 60 - 64 | 33.0 | 35.0 | 0.04 | 33.2 | 34.0 | 0.02 | 33.3 | 35.0 | 0.04 |
| 65 - 69 | 2.8 | 3.6 | 0.04 | 2.9 | 3.2 | 0.02 | 3.2 | 3.3 | 0.01 |
| Gender: female | 38.1 | 42.2 | 0.08 | 40.6 | 40.1 | -0.01 | 41.3 | 40.6 | -0.01 |
| **Medical history: General** | | | | | | | | | |
| Acute respiratory disease | 29.6 | 30.6 | 0.02 | 30.5 | 30.4 | 0.00 | 30.5 | 30.4 | 0.00 |
| Attention deficit hyperactivity disorder | 1.1 | 0.7 | -0.05 | 1.1 | 0.9 | -0.02 | 1.1 | 0.9 | -0.02 |
| Chronic liver disease | 2.5 | 4.1 | 0.09 | 2.9 | 2.6 | -0.02 | 3.5 | 3.2 | -0.02 |
| Chronic obstructive lung disease | 5.2 | 6.2 | 0.04 | 5.3 | 5.4 | 0.01 | 5.7 | 5.9 | 0.01 |
| Crohn’s disease | 0.3 | 0.4 | 0.01 | 0.3 | 0.3 | 0.00 | 0.4 | 0.4 | 0.00 |
| Dementia | 0.3 | 0.3 | 0.01 | 0.3 | 0.4 | 0.02 | 0.3 | 0.4 | 0.02 |
| Depressive disorder | 11.8 | 10.6 | -0.04 | 11.8 | 11.9 | 0.01 | 11.8 | 12.1 | 0.01 |
| Gastroesophageal reflux disease | 17.2 | 14.7 | -0.07 | 17.6 | 17.6 | 0.00 | 17.5 | 17.6 | 0.00 |
| Gastrointestinal hemorrhage | 2.0 | 2.7 | 0.04 | 2.1 | 2.1 | 0.00 | 2.2 | 2.3 | 0.01 |
| Human immunodeficiency virus infection | 0.4 | 0.3 | -0.01 | 0.3 | 0.4 | 0.01 | 0.3 | 0.4 | 0.02 |
| Hyperlipidemia | 82.2 | 75.0 | -0.18 | 81.1 | 82.0 | 0.02 | 80.7 | 81.0 | 0.01 |
| Hypertensive disorder | 81.8 | 76.5 | -0.13 | 81.2 | 81.4 | 0.00 | 81.2 | 81.4 | 0.00 |
| Lesion of liver | 0.8 | 0.9 | 0.01 | 0.8 | 0.8 | 0.00 | 0.8 | 1.0 | 0.02 |
| Obesity | 31.5 | 19.6 | -0.28 | 29.5 | 29.9 | 0.01 | 28.6 | 28.7 | 0.00 |
| Osteoarthritis | 24.5 | 23.7 | -0.02 | 24.9 | 25.2 | 0.01 | 25.5 | 25.4 | 0.00 |
| Pneumonia | 3.2 | 3.2 | 0.00 | 3.1 | 3.3 | 0.01 | 3.4 | 3.3 | -0.01 |
| Psoriasis | 2.2 | 1.7 | -0.03 | 2.2 | 1.9 | -0.02 | 2.2 | 1.9 | -0.02 |
| Renal impairment | 5.3 | 5.8 | 0.02 | 5.5 | 5.5 | 0.00 | 5.9 | 6.2 | 0.01 |
| Rheumatoid arthritis | 1.6 | 1.6 | 0.00 | 1.6 | 1.4 | -0.02 | 1.7 | 1.5 | -0.02 |
| Schizophrenia | 0.1 | 0.2 | 0.02 | 0.1 | 0.2 | 0.03 | 0.1 | 0.2 | 0.03 |
| Ulcerative colitis | 0.4 | 0.4 | -0.01 | 0.4 | 0.3 | -0.01 | 0.5 | 0.4 | -0.01 |
| Urinary tract infectious disease | 6.8 | 8.9 | 0.07 | 7.4 | 7.2 | -0.01 | 7.6 | 7.7 | 0.00 |
| Viral hepatitis C | 0.4 | 0.5 | 0.02 | 0.4 | 0.2 | -0.03 | 0.4 | 0.4 | -0.01 |
| Visual system disorder | 28.1 | 28.3 | 0.00 | 29.2 | 29.3 | 0.00 | 29.6 | 29.3 | -0.01 |
| **Medical history: Cardiovascular disease** | | | | | | | | | |
| Atrial fibrillation | 5.0 | 4.8 | -0.01 | 4.7 | 4.9 | 0.01 | 4.9 | 5.1 | 0.01 |
| Cerebrovascular disease | 9.1 | 10.4 | 0.04 | 9.4 | 9.0 | -0.01 | 9.9 | 9.3 | -0.02 |
| Coronary arteriosclerosis | 34.8 | 32.1 | -0.06 | 30.5 | 31.8 | 0.03 | 31.4 | 32.0 | 0.01 |
| Heart disease | 50.8 | 49.4 | -0.03 | 47.2 | 48.0 | 0.02 | 48.6 | 48.6 | 0.00 |
| Heart failure | 6.7 | 5.8 | -0.04 | 5.4 | 5.9 | 0.02 | 5.8 | 6.2 | 0.01 |
| Ischemic heart disease | 18.2 | 16.8 | -0.04 | 15.9 | 16.5 | 0.02 | 15.9 | 16.5 | 0.02 |
| Peripheral vascular disease | 15.6 | 13.7 | -0.05 | 14.8 | 15.2 | 0.01 | 14.8 | 15.2 | 0.01 |
| Pulmonary embolism | 0.9 | 0.8 | 0.00 | 0.8 | 1.0 | 0.02 | 0.9 | 0.9 | 0.00 |
| Venous thrombosis | 1.5 | 1.9 | 0.03 | 1.6 | 1.8 | 0.01 | 1.7 | 1.8 | 0.01 |
| **Medical history: Neoplasms** | | | | | | | | | |
| Hematologic neoplasm | 1.0 | 1.2 | 0.02 | 1.0 | 1.0 | 0.00 | 1.2 | 1.1 | -0.01 |
| Malignant lymphoma | 0.4 | 0.6 | 0.03 | 0.4 | 0.4 | 0.01 | 0.4 | 0.5 | 0.02 |
| Malignant neoplasm of anorectum | 0.1 | 0.2 | 0.01 | 0.1 | 0.1 | -0.01 | 0.1 | 0.1 | 0.00 |
| Malignant neoplastic disease | 7.2 | 8.4 | 0.04 | 7.5 | 7.6 | 0.00 | 7.5 | 7.6 | 0.00 |
| Malignant tumor of breast | 1.2 | 1.5 | 0.03 | 1.4 | 1.2 | -0.02 | 1.4 | 1.2 | -0.02 |
| Malignant tumor of colon | 0.3 | 0.4 | 0.02 | 0.3 | 0.3 | 0.00 | 0.4 | 0.4 | 0.00 |
| Malignant tumor of lung | 0.3 | 0.3 | 0.01 | 0.3 | 0.3 | -0.01 | 0.4 | 0.3 | -0.02 |
| Malignant tumor of urinary bladder | 0.2 | 0.4 | 0.03 | 0.2 | 0.3 | 0.02 | 0.2 | 0.3 | 0.03 |
| Primary malignant neoplasm of prostate | 1.1 | 1.2 | 0.01 | 1.1 | 1.1 | 0.00 | 1.1 | 1.2 | 0.01 |
| **Medication use** | | | | | | | | | |
| Agents acting on the renin-angiotensin system | 74.5 | 73.8 | -0.01 | 73.7 | 73.5 | 0.00 | 73.6 | 73.9 | 0.01 |
| Antibacterials for systemic use | 61.5 | 64.3 | 0.06 | 62.4 | 61.7 | -0.01 | 63.0 | 62.4 | -0.01 |
| Antidepressants | 30.2 | 29.3 | -0.02 | 30.1 | 29.5 | -0.01 | 30.1 | 29.7 | -0.01 |
| Antiepileptics | 17.2 | 16.1 | -0.03 | 17.8 | 17.6 | 0.00 | 17.8 | 17.6 | 0.00 |
| Antiinflammatory and antirheumatic products | 39.0 | 35.2 | -0.08 | 39.1 | 38.5 | -0.01 | 39.0 | 38.3 | -0.01 |

| (Continued) | Before ajustment | | | After matching | | | After stratification | | |
|---|---|---|---|---|---|---|---|---|---|
| Characteristic | T (%) | C (%) | StdDiff | T (%) | C (%) | StdDiff | T (%) | C (%) | StdDiff |
| Antineoplastic agents | 4.5 | 5.6 | 0.05 | 4.6 | 4.9 | 0.02 | 4.7 | 5.0 | 0.01 |
| Antipsoriatics | 0.8 | 0.7 | -0.01 | 0.8 | 0.5 | -0.03 | 0.8 | 0.6 | -0.02 |
| Antithrombotic agents | 28.4 | 26.1 | -0.05 | 25.8 | 26.6 | 0.02 | 26.4 | 26.7 | 0.01 |
| Beta blocking agents | 47.3 | 46.6 | -0.01 | 44.7 | 45.4 | 0.01 | 45.2 | 45.9 | 0.01 |
| Calcium channel blockers | 27.3 | 27.0 | -0.01 | 27.1 | 27.5 | 0.01 | 27.1 | 27.2 | 0.00 |
| Diuretics | 42.5 | 43.7 | 0.02 | 42.3 | 41.8 | -0.01 | 42.6 | 42.3 | -0.01 |
| Drugs for acid related disorders | 31.2 | 32.2 | 0.02 | 31.6 | 30.8 | -0.02 | 32.1 | 31.6 | -0.01 |
| Drugs for obstructive airway diseases | 42.0 | 41.3 | -0.01 | 42.6 | 41.8 | -0.02 | 42.9 | 42.4 | -0.01 |
| Immunosuppressants | 3.2 | 3.0 | -0.01 | 3.3 | 2.9 | -0.02 | 3.3 | 3.2 | -0.01 |
| Lipid modifying agents | 80.0 | 78.3 | -0.04 | 78.2 | 78.3 | 0.00 | 78.5 | 78.2 | -0.01 |
| Opioids | 31.3 | 32.5 | 0.03 | 31.8 | 31.0 | -0.02 | 33.3 | 31.7 | -0.03 |
| Psycholeptics | 28.4 | 28.9 | 0.01 | 28.2 | 27.8 | -0.01 | 29.1 | 28.5 | -0.01 |
| Psychostimulants | 3.6 | 3.2 | -0.02 | 3.7 | 3.3 | -0.02 | 3.6 | 3.4 | -0.01 |

**Supplementary Table 11:** Baseline patient characteristics for SGLT2is (T) and DPP4is (C) new-users in the GermanyDA data source. We report proportion of initiators satisfying selected base-line characteristics and the standardized difference of population proportions (StdDiff) before and after propensity score adjustment. Less extreme StdDiffs through matching and stratification suggest improved balance between patient cohorts through adjustment.

| | Before adjustment | | | After matching | | | After stratification | | |
|---|---|---|---|---|---|---|---|---|---|
| Characteristic | T (%) | C (%) | StdDiff | T (%) | C (%) | StdDiff | T (%) | C (%) | StdDiff |
| **Age group** | | | | | | | | | |
| 30 - 34 | 0.2 | 0.1 | -0.02 | 0.2 | <0.1 | -0.03 | 0.4 | 0.1 | -0.05 |
| 35 - 39 | 0.4 | 0.3 | -0.02 | 0.5 | 0.4 | -0.02 | 0.7 | 0.3 | -0.05 |
| 40 - 44 | 1.0 | 0.9 | 0.00 | 1.0 | 1.0 | 0.00 | 0.9 | 1.0 | 0.00 |
| 45 - 49 | 3.1 | 2.6 | -0.03 | 3.1 | 2.8 | -0.02 | 2.9 | 2.5 | -0.03 |
| 50 - 54 | 7.0 | 5.8 | -0.05 | 6.8 | 6.0 | -0.03 | 6.4 | 6.1 | -0.01 |
| 55 - 59 | 12.2 | 10.4 | -0.06 | 12.1 | 11.9 | -0.01 | 11.7 | 10.8 | -0.03 |
| 60 - 64 | 17.1 | 13.2 | -0.11 | 15.7 | 16.1 | 0.01 | 14.6 | 14.3 | -0.01 |
| 65 - 69 | 16.5 | 14.6 | -0.05 | 16.8 | 16.8 | 0.00 | 16.9 | 15.5 | -0.04 |
| 70 - 74 | 15.4 | 15.7 | 0.01 | 15.3 | 15.1 | -0.01 | 15.1 | 14.7 | -0.01 |
| 75 - 79 | 13.3 | 16.6 | 0.09 | 13.8 | 15.1 | 0.04 | 15.0 | 16.1 | 0.03 |
| 80 - 84 | 9.6 | 12.4 | 0.09 | 10.2 | 9.7 | -0.02 | 10.7 | 11.8 | 0.03 |
| 85 - 89 | 3.5 | 5.7 | 0.10 | 3.8 | 4.0 | 0.01 | 3.9 | 5.3 | 0.07 |
| 90 - 94 | 0.6 | 1.5 | 0.09 | 0.6 | 1.0 | 0.04 | 0.7 | 1.4 | 0.07 |
| Gender: female | 30.7 | 40.5 | 0.21 | 34.3 | 32.3 | -0.04 | 39.0 | 37.5 | -0.03 |
| **Medical history: General** | | | | | | | | | |
| Acute respiratory disease | 16.2 | 18.2 | 0.05 | 16.5 | 16.0 | -0.01 | 17.6 | 17.1 | -0.01 |
| Attention deficit hyperactivity disorder | <0.1 | 0.2 | 0.04 | <0.1 | <0.1 | 0.04 | <0.1 | 0.2 | 0.06 |
| Chronic liver disease | 0.7 | 0.5 | -0.02 | 0.6 | 0.5 | -0.02 | 0.5 | 0.5 | 0.01 |
| Chronic obstructive lung disease | 8.0 | 8.7 | 0.03 | 7.8 | 8.3 | 0.02 | 9.2 | 8.4 | -0.03 |
| Crohn’s disease | 0.2 | 0.2 | 0.00 | 0.1 | 0.2 | 0.01 | 0.1 | 0.2 | 0.01 |
| Dementia | 1.4 | 3.2 | 0.12 | 1.6 | 1.5 | -0.01 | 2.1 | 2.9 | 0.05 |
| Depressive disorder | 9.2 | 10.4 | 0.04 | 9.1 | 9.2 | 0.00 | 10.9 | 10.1 | -0.03 |
| Gastroesophageal reflux disease | 2.5 | 3.0 | 0.03 | 2.4 | 2.6 | 0.01 | 2.7 | 2.8 | 0.00 |
| Gastrointestinal hemorrhage | 1.0 | 1.0 | 0.00 | 0.9 | 0.9 | 0.00 | 0.8 | 1.0 | 0.02 |
| Hyperlipidemia | 28.8 | 30.4 | 0.04 | 28.0 | 27.6 | -0.01 | 27.7 | 28.5 | 0.02 |
| Hypertensive disorder | 47.8 | 54.7 | 0.14 | 47.8 | 46.8 | -0.02 | 50.0 | 50.6 | 0.01 |
| Lesion of liver | 0.7 | 0.8 | 0.01 | 0.6 | 0.8 | 0.03 | 0.6 | 0.8 | 0.03 |
| Obesity | 10.2 | 8.2 | -0.07 | 9.4 | 9.3 | 0.00 | 9.6 | 8.9 | -0.03 |
| Osteoarthritis | 11.9 | 14.1 | 0.07 | 12.2 | 12.6 | 0.01 | 12.6 | 13.3 | 0.02 |
| Pneumonia | 2.1 | 3.1 | 0.07 | 2.0 | 2.3 | 0.02 | 2.4 | 2.8 | 0.03 |
| Psoriasis | 1.7 | 1.4 | -0.02 | 1.8 | 1.5 | -0.02 | 1.6 | 1.6 | 0.00 |
| Renal impairment | 7.1 | 9.5 | 0.09 | 7.1 | 6.7 | -0.02 | 7.8 | 8.7 | 0.03 |
| Rheumatoid arthritis | 1.5 | 1.5 | 0.00 | 1.5 | 1.0 | -0.04 | 1.6 | 1.4 | -0.02 |
| Schizophrenia | 0.2 | 0.2 | 0.00 | 0.2 | 0.3 | 0.02 | 0.3 | 0.2 | -0.02 |
| Ulcerative colitis | 0.2 | 0.3 | 0.02 | 0.1 | 0.2 | 0.01 | 0.1 | 0.3 | 0.03 |
| Urinary tract infectious disease | 4.5 | 6.1 | 0.07 | 4.9 | 5.3 | 0.02 | 5.1 | 5.7 | 0.02 |
| Visual system disorder | 8.1 | 10.9 | 0.10 | 8.5 | 8.4 | -0.01 | 9.2 | 9.6 | 0.01 |
| **Medical history: Cardiovascular disease** | | | | | | | | | |
| Atrial fibrillation | 4.7 | 5.0 | 0.01 | 4.3 | 4.8 | 0.02 | 4.3 | 4.8 | 0.02 |
| Cerebrovascular disease | 8.0 | 9.6 | 0.06 | 8.2 | 8.2 | 0.00 | 8.0 | 8.9 | 0.03 |
| Coronary arteriosclerosis | 14.6 | 10.1 | -0.14 | 12.2 | 11.8 | -0.01 | 11.0 | 11.6 | 0.02 |
| Heart disease | 46.9 | 45.4 | -0.03 | 42.6 | 42.6 | 0.00 | 41.8 | 44.3 | 0.05 |
| Heart failure | 13.4 | 11.4 | -0.06 | 10.7 | 10.5 | -0.01 | 10.9 | 11.2 | 0.01 |
| Ischemic heart disease | 26.5 | 25.4 | -0.03 | 23.5 | 22.8 | -0.02 | 22.6 | 24.0 | 0.03 |
| Peripheral vascular disease | 12.0 | 12.5 | 0.01 | 11.8 | 11.0 | -0.03 | 10.9 | 12.3 | 0.04 |
| Pulmonary embolism | 0.7 | 0.9 | 0.03 | 0.5 | 0.7 | 0.03 | 0.5 | 0.9 | 0.05 |
| Venous thrombosis | 1.8 | 2.0 | 0.01 | 1.8 | 1.8 | 0.00 | 2.4 | 2.0 | -0.03 |
| **Medical history: Neoplasms** | | | | | | | | | |
| Hematologic neoplasm | 0.3 | 0.5 | 0.03 | 0.3 | 0.4 | 0.01 | 0.6 | 0.4 | -0.02 |
| Malignant lymphoma | 0.2 | 0.4 | 0.02 | 0.2 | 0.3 | 0.01 | 0.4 | 0.4 | -0.01 |
| Malignant neoplasm of anorectum | <0.1 | 0.2 | 0.04 | <0.1 | 0.2 | 0.02 | <0.1 | 0.2 | 0.04 |
| Malignant neoplastic disease | 4.7 | 6.8 | 0.09 | 5.0 | 5.3 | 0.01 | 5.8 | 6.3 | 0.02 |
| Malignant tumor of breast | 0.6 | 0.6 | 0.01 | 0.5 | 0.3 | -0.03 | 0.9 | 0.5 | -0.04 |
| Malignant tumor of colon | 0.4 | 0.5 | 0.02 | 0.4 | 0.4 | 0.00 | 0.3 | 0.5 | 0.02 |
| Malignant tumor of lung | <0.1 | 0.2 | 0.03 | <0.1 | 0.2 | 0.04 | <0.1 | 0.2 | 0.03 |
| Malignant tumor of urinary bladder | 0.2 | 0.3 | 0.01 | 0.2 | 0.1 | -0.02 | 0.4 | 0.2 | -0.04 |
| Primary malignant neoplasm of prostate | 0.8 | 1.1 | 0.03 | 0.9 | 0.9 | 0.00 | 0.7 | 0.9 | 0.03 |
| **Medication use** | | | | | | | | | |
| Agents acting on the renin-angiotensin system | 79.3 | 77.3 | -0.05 | 77.3 | 76.7 | -0.01 | 76.7 | 77.3 | 0.01 |
| Antibacterials for systemic use | 22.8 | 28.9 | 0.14 | 24.1 | 22.6 | -0.04 | 27.7 | 26.1 | -0.04 |
| Antidepressants | 11.1 | 12.5 | 0.04 | 11.8 | 11.1 | -0.02 | 13.1 | 12.2 | -0.03 |

| (Continued) | Before ajustment | | | After matching | | | After stratification | | |
|---|---|---|---|---|---|---|---|---|---|
| Characteristic | T (%) | C (%) | StdDiff | T (%) | C (%) | StdDiff | T (%) | C (%) | StdDiff |
| Antiepileptics | 5.5 | 6.6 | 0.05 | 5.7 | 5.8 | 0.01 | 6.2 | 6.7 | 0.02 |
| Antiinflammatory and antirheumatic products | 53.5 | 54.9 | 0.03 | 52.0 | 52.6 | 0.01 | 53.5 | 53.9 | 0.01 |
| Antineoplastic agents | 1.4 | 1.3 | 0.00 | 1.3 | 1.0 | -0.03 | 1.5 | 1.2 | -0.02 |
| Antipsoriatics | 0.5 | 0.6 | 0.00 | 0.5 | 0.5 | 0.00 | 0.5 | 0.6 | 0.02 |
| Antithrombotic agents | 56.3 | 53.4 | -0.06 | 52.5 | 53.6 | 0.02 | 52.4 | 54.0 | 0.03 |
| Beta blocking agents | 62.9 | 61.9 | -0.02 | 60.6 | 60.5 | 0.00 | 61.0 | 61.7 | 0.02 |
| Calcium channel blockers | 39.6 | 38.6 | -0.02 | 39.1 | 39.3 | 0.00 | 39.9 | 39.5 | -0.01 |
| Diuretics | 58.3 | 59.1 | 0.02 | 56.0 | 55.9 | 0.00 | 58.1 | 57.6 | -0.01 |
| Drugs for acid related disorders | 42.9 | 43.9 | 0.02 | 42.5 | 42.3 | 0.00 | 44.6 | 44.3 | 0.00 |
| Drugs for obstructive airway diseases | 19.8 | 20.5 | 0.02 | 19.9 | 20.1 | 0.00 | 21.0 | 20.3 | -0.02 |
| Immunosuppressants | 1.0 | 0.8 | -0.03 | 1.0 | 0.6 | -0.05 | 0.8 | 0.7 | -0.01 |
| Lipid modifying agents | 70.0 | 61.5 | -0.18 | 65.5 | 66.5 | 0.02 | 62.1 | 64.2 | 0.04 |
| Opioids | 12.5 | 14.9 | 0.07 | 12.9 | 12.8 | 0.00 | 14.5 | 14.0 | -0.02 |
| Psycholeptics | 9.3 | 11.8 | 0.08 | 9.7 | 9.7 | 0.00 | 11.7 | 11.2 | -0.02 |
| Psychostimulants | 0.1 | 0.3 | 0.04 | 0.2 | <0.1 | -0.02 | 0.3 | 0.2 | -0.03 |

**Supplementary Table 12:** Baseline patient characteristics for SGLT2is (T) and DPP4is (C) new-users in the Optum-DOD data source. We report proportion of initiators satisfying selected base-line characteristics and the standardized difference of population proportions (StdDiff) before and after propensity score adjustment. Less extreme StdDiffs through matching and stratification suggest improved balance between patient cohorts through adjustment.

| | Before adjustment | | | After matching | | | After stratification | | |
|---|---|---|---|---|---|---|---|---|---|
| Characteristic | T (%) | C (%) | StdDiff | T (%) | C (%) | StdDiff | T (%) | C (%) | StdDiff |
| **Age group** | | | | | | | | | |
| 25 - 29 | 0.1 | 0.1 | -0.01 | 0.1 | <0.1 | -0.02 | 0.1 | <0.1 | -0.02 |
| 30 - 34 | 0.2 | 0.3 | 0.01 | 0.3 | 0.3 | 0.01 | 0.3 | 0.3 | 0.01 |
| 35 - 39 | 0.7 | 0.6 | -0.01 | 0.9 | 0.6 | -0.03 | 0.9 | 0.5 | -0.05 |
| 40 - 44 | 2.1 | 1.6 | -0.04 | 1.8 | 1.6 | -0.02 | 1.8 | 1.4 | -0.03 |
| 45 - 49 | 4.6 | 3.6 | -0.05 | 4.1 | 3.7 | -0.02 | 3.7 | 3.4 | -0.01 |
| 50 - 54 | 7.7 | 6.6 | -0.04 | 7.3 | 6.9 | -0.01 | 6.7 | 6.5 | -0.01 |
| 55 - 59 | 12.7 | 10.1 | -0.08 | 11.4 | 11.7 | 0.01 | 11.4 | 10.5 | -0.03 |
| 60 - 64 | 15.2 | 12.7 | -0.07 | 13.7 | 14.0 | 0.01 | 13.1 | 12.7 | -0.01 |
| 65 - 69 | 19.0 | 16.6 | -0.06 | 18.9 | 19.0 | 0.00 | 18.0 | 17.9 | 0.00 |
| 70 - 74 | 19.5 | 18.9 | -0.01 | 20.1 | 21.0 | 0.02 | 19.1 | 20.5 | 0.04 |
| 75 - 79 | 11.5 | 14.2 | 0.08 | 13.0 | 12.7 | -0.01 | 12.6 | 13.6 | 0.03 |
| 80 - 84 | 4.6 | 9.1 | 0.18 | 7.8 | 7.8 | 0.00 | 7.8 | 7.8 | 0.00 |
| 85 - 89 | 1.8 | 5.1 | 0.18 | 2.4 | 2.1 | -0.02 | 4.2 | 4.3 | 0.01 |
| 90 - 94 | 0.2 | 0.4 | 0.03 | 0.2 | 0.5 | 0.04 | 0.2 | 0.5 | 0.06 |
| Gender: female | 38.7 | 47.3 | 0.17 | 42.8 | 42.2 | -0.01 | 45.4 | 44.4 | -0.02 |
| **Medical history: General** | | | | | | | | | |
| Acute respiratory disease | 27.8 | 29.2 | 0.03 | 28.2 | 28.0 | 0.00 | 29.7 | 28.5 | -0.03 |
| Attention deficit hyperactivity disorder | 0.7 | 0.6 | -0.02 | 0.7 | 0.8 | 0.02 | 0.6 | 0.7 | 0.01 |
| Chronic liver disease | 2.2 | 4.0 | 0.10 | 2.5 | 2.4 | -0.01 | 3.5 | 2.9 | -0.03 |
| Chronic obstructive lung disease | 14.6 | 15.5 | 0.03 | 14.8 | 14.6 | 0.00 | 16.0 | 15.8 | 0.00 |
| Crohn’s disease | 0.4 | 0.4 | 0.01 | 0.4 | 0.4 | 0.00 | 0.5 | 0.3 | -0.02 |
| Dementia | 1.7 | 4.7 | 0.17 | 2.2 | 2.2 | 0.00 | 4.2 | 4.1 | 0.00 |
| Depressive disorder | 17.9 | 17.4 | -0.01 | 18.0 | 18.1 | 0.00 | 19.4 | 18.8 | -0.02 |
| Gastroesophageal reflux disease | 25.1 | 23.6 | -0.03 | 25.1 | 24.3 | -0.02 | 26.2 | 25.3 | -0.02 |
| Gastrointestinal hemorrhage | 2.9 | 3.8 | 0.05 | 2.9 | 3.0 | 0.00 | 3.5 | 3.4 | 0.00 |
| Human immunodeficiency virus infection | 0.3 | 0.3 | 0.00 | 0.3 | 0.5 | 0.04 | 0.3 | 0.4 | 0.02 |
| Hyperlipidemia | 87.8 | 87.1 | -0.02 | 86.9 | 87.0 | 0.00 | 87.0 | 87.1 | 0.00 |
| Hypertensive disorder | 88.4 | 88.1 | -0.01 | 87.7 | 87.7 | 0.00 | 88.5 | 88.2 | -0.01 |
| Lesion of liver | 1.6 | 1.5 | 0.00 | 1.6 | 1.4 | -0.02 | 2.0 | 1.5 | -0.04 |
| Obesity | 33.7 | 23.3 | -0.23 | 30.2 | 31.1 | 0.02 | 29.1 | 29.2 | 0.00 |
| Osteoarthritis | 33.9 | 35.9 | 0.04 | 35.1 | 35.2 | 0.00 | 36.9 | 36.3 | -0.01 |
| Pneumonia | 5.0 | 6.3 | 0.06 | 4.9 | 5.1 | 0.01 | 6.1 | 6.1 | 0.00 |
| Psoriasis | 2.2 | 1.8 | -0.03 | 2.2 | 1.9 | -0.02 | 2.2 | 1.9 | -0.02 |
| Renal impairment | 17.4 | 23.7 | 0.16 | 19.2 | 18.7 | -0.01 | 22.5 | 22.7 | 0.00 |
| Rheumatoid arthritis | 2.2 | 2.5 | 0.02 | 2.4 | 2.5 | 0.01 | 2.8 | 2.6 | -0.01 |
| Schizophrenia | 0.4 | 0.5 | 0.02 | 0.4 | 0.6 | 0.03 | 0.4 | 0.6 | 0.03 |
| Ulcerative colitis | 0.4 | 0.5 | 0.00 | 0.4 | 0.3 | -0.02 | 0.6 | 0.4 | -0.02 |
| Urinary tract infectious disease | 9.1 | 15.6 | 0.20 | 10.7 | 10.4 | -0.01 | 13.6 | 13.3 | -0.01 |
| Viral hepatitis C | 0.6 | 0.7 | 0.01 | 0.6 | 0.6 | 0.00 | 0.7 | 0.7 | 0.00 |
| Visual system disorder | 45.2 | 47.7 | 0.05 | 47.1 | 46.3 | -0.02 | 48.3 | 47.7 | -0.01 |
| **Medical history: Cardiovascular disease** | | | | | | | | | |
| Atrial fibrillation | 12.4 | 13.0 | 0.02 | 11.8 | 12.0 | 0.01 | 12.6 | 13.3 | 0.02 |
| Cerebrovascular disease | 13.3 | 16.3 | 0.08 | 13.5 | 13.6 | 0.00 | 14.8 | 14.9 | 0.00 |
| Coronary arteriosclerosis | 44.7 | 38.4 | -0.13 | 39.5 | 40.2 | 0.01 | 39.1 | 40.3 | 0.02 |
| Heart disease | 62.3 | 59.4 | -0.06 | 58.1 | 58.3 | 0.01 | 59.2 | 60.0 | 0.02 |
| Heart failure | 16.4 | 14.7 | -0.05 | 13.8 | 13.9 | 0.00 | 15.3 | 15.6 | 0.01 |
| Ischemic heart disease | 24.3 | 20.5 | -0.09 | 21.1 | 21.3 | 0.00 | 21.4 | 21.5 | 0.00 |
| Peripheral vascular disease | 27.0 | 26.2 | -0.02 | 27.0 | 27.4 | 0.01 | 27.7 | 28.1 | 0.01 |
| Pulmonary embolism | 1.3 | 1.3 | 0.00 | 1.4 | 1.3 | -0.01 | 1.7 | 1.5 | -0.02 |
| Venous thrombosis | 2.3 | 2.5 | 0.01 | 2.4 | 1.9 | -0.03 | 2.8 | 2.3 | -0.03 |
| **Medical history: Neoplasms** | | | | | | | | | |
| Hematologic neoplasm | 1.3 | 1.6 | 0.02 | 1.4 | 1.3 | -0.01 | 1.4 | 1.5 | 0.01 |
| Malignant lymphoma | 0.7 | 0.8 | 0.01 | 0.7 | 0.7 | 0.00 | 0.7 | 0.8 | 0.01 |
| Malignant neoplasm of anorectum | 0.2 | 0.3 | 0.02 | 0.3 | 0.2 | 0.00 | 0.3 | 0.3 | 0.01 |
| Malignant neoplastic disease | 12.3 | 14.3 | 0.06 | 12.9 | 12.2 | -0.02 | 13.0 | 13.7 | 0.02 |
| Malignant tumor of breast | 1.5 | 2.2 | 0.05 | 1.9 | 2.0 | 0.01 | 1.9 | 2.0 | 0.01 |
| Malignant tumor of colon | 0.6 | 0.8 | 0.03 | 0.6 | 0.5 | -0.01 | 0.6 | 0.8 | 0.02 |
| Malignant tumor of lung | 0.5 | 0.7 | 0.03 | 0.5 | 0.6 | 0.02 | 0.5 | 0.7 | 0.02 |
| Malignant tumor of urinary bladder | 0.5 | 0.7 | 0.04 | 0.5 | 0.6 | 0.01 | 0.5 | 0.7 | 0.03 |
| Primary malignant neoplasm of prostate | 2.8 | 3.2 | 0.02 | 3.0 | 2.7 | -0.02 | 2.9 | 3.1 | 0.01 |
| **Medication use** | | | | | | | | | |

| (Continued) | Before ajustment | | | After matching | | | After stratification | | |
|---|---|---|---|---|---|---|---|---|---|
| Characteristic | T (%) | C (%) | StdDiff | T (%) | C (%) | StdDiff | T (%) | C (%) | StdDiff |
| Agents acting on the renin-angiotensin system | 76.9 | 75.8 | -0.03 | 75.8 | 75.9 | 0.00 | 76.0 | 76.0 | 0.00 |
| Antibacterials for systemic use | 60.2 | 63.9 | 0.08 | 61.6 | 61.2 | -0.01 | 63.2 | 62.4 | -0.02 |
| Antidepressants | 31.8 | 31.6 | 0.00 | 32.0 | 31.4 | -0.01 | 33.6 | 32.3 | -0.03 |
| Antiepileptics | 22.5 | 22.2 | -0.01 | 23.2 | 23.0 | -0.01 | 24.3 | 24.0 | -0.01 |
| Antiinflammatory and antirheumatic products | 33.6 | 32.3 | -0.03 | 33.6 | 33.8 | 0.00 | 34.4 | 33.3 | -0.02 |
| Antineoplastic agents | 6.4 | 7.2 | 0.03 | 7.0 | 7.0 | 0.00 | 7.0 | 7.0 | 0.00 |
| Antipsoriatics | 0.5 | 0.7 | 0.02 | 0.6 | 0.6 | 0.00 | 0.5 | 0.7 | 0.03 |
| Antithrombotic agents | 35.2 | 31.5 | -0.08 | 32.0 | 31.9 | 0.00 | 33.1 | 33.1 | 0.00 |
| Beta blocking agents | 55.3 | 52.3 | -0.06 | 51.9 | 52.2 | 0.00 | 53.1 | 53.2 | 0.00 |
| Calcium channel blockers | 32.0 | 33.6 | 0.04 | 32.7 | 32.2 | -0.01 | 33.7 | 33.5 | 0.00 |
| Diuretics | 48.3 | 49.6 | 0.03 | 47.4 | 47.3 | 0.00 | 49.2 | 48.6 | -0.01 |
| Drugs for acid related disorders | 34.7 | 36.2 | 0.03 | 35.5 | 35.2 | -0.01 | 37.1 | 36.8 | -0.01 |
| Drugs for obstructive airway diseases | 43.5 | 43.9 | 0.01 | 44.2 | 43.7 | -0.01 | 45.7 | 44.6 | -0.02 |
| Immunosuppressants | 3.3 | 3.3 | 0.00 | 3.6 | 3.2 | -0.02 | 3.6 | 3.3 | -0.02 |
| Lipid modifying agents | 85.7 | 82.6 | -0.08 | 84.2 | 84.5 | 0.01 | 83.6 | 84.2 | 0.01 |
| Opioids | 32.8 | 33.2 | 0.01 | 32.3 | 31.8 | -0.01 | 33.9 | 33.1 | -0.02 |
| Psycholeptics | 29.8 | 29.7 | 0.00 | 29.4 | 28.6 | -0.02 | 31.5 | 29.9 | -0.04 |
| Psychostimulants | 2.1 | 2.0 | -0.01 | 2.0 | 2.3 | 0.02 | 2.2 | 2.1 | 0.00 |

**Supplementary Table 13:** Baseline patient characteristics for SGLT2is (T) and DPP4is (C) new-users in the MDCR data source. We report proportion of initiators satisfying selected base-line characteristics and the standardized difference of population proportions (StdDiff) before and after propensity score adjustment. Less extreme StdDiffs through matching and stratification suggest improved balance between patient cohorts through adjustment.

| | Before adjustment | | | After matching | | | After stratification | | |
|---|---|---|---|---|---|---|---|---|---|
| Characteristic | T (%) | C (%) | StdDiff | T (%) | C (%) | StdDiff | T (%) | C (%) | StdDiff |
| **Age group** | | | | | | | | | |
| 50 - 54 | <0.2 | 0.1 | -0.01 | <0.2 | <0.3 | 0.01 | <0.2 | 0.2 | 0.02 |
| 55 - 59 | 1.0 | 0.4 | -0.07 | 1.1 | 0.6 | -0.05 | 1.1 | 0.5 | -0.07 |
| 60 - 64 | 2.1 | 1.5 | -0.05 | 2.2 | 1.8 | -0.03 | 2.6 | 1.6 | -0.07 |
| 65 - 69 | 27.8 | 22.1 | -0.13 | 27.2 | 27.8 | 0.01 | 22.1 | 23.3 | 0.03 |
| 70 - 74 | 36.9 | 28.8 | -0.17 | 35.9 | 32.7 | -0.07 | 34.9 | 30.3 | -0.10 |
| 75 - 79 | 21.3 | 22.1 | 0.02 | 21.0 | 22.5 | 0.04 | 21.0 | 22.1 | 0.03 |
| 80 - 84 | 7.5 | 15.0 | 0.24 | 8.4 | 9.2 | 0.03 | 10.5 | 12.7 | 0.07 |
| 85 - 89 | 2.6 | 7.3 | 0.22 | 3.2 | 3.3 | 0.01 | 5.3 | 6.4 | 0.04 |
| 90 - 94 | 0.5 | 2.3 | 0.15 | 0.6 | 1.5 | 0.09 | 2.1 | 2.4 | 0.02 |
| 95 - 99 | <0.2 | 0.4 | 0.06 | <0.2 | 0.4 | 0.05 | <0.2 | 0.5 | 0.08 |
| Gender: female | 36.1 | 44.1 | 0.16 | 38.2 | 38.5 | 0.01 | 43.7 | 42.8 | -0.02 |
| **Medical history: General** | | | | | | | | | |
| Acute respiratory disease | 26.0 | 25.4 | -0.01 | 27.1 | 25.2 | -0.04 | 29.0 | 26.2 | -0.06 |
| Attention deficit hyperactivity disorder | <0.2 | 0.1 | 0.00 | 0.2 | 0.2 | 0.01 | 0.2 | 0.2 | 0.01 |
| Chronic liver disease | 2.3 | 2.7 | 0.02 | 2.5 | 1.9 | -0.04 | 3.1 | 2.6 | -0.03 |
| Chronic obstructive lung disease | 14.2 | 14.2 | 0.00 | 14.3 | 13.3 | -0.03 | 14.9 | 15.0 | 0.00 |
| Crohn’s disease | 0.5 | 0.3 | -0.03 | 0.6 | <0.3 | -0.06 | 0.3 | 0.3 | 0.00 |
| Dementia | 1.9 | 4.1 | 0.13 | 2.1 | 3.0 | 0.06 | 2.5 | 4.4 | 0.10 |
| Depressive disorder | 9.9 | 8.6 | -0.04 | 10.0 | 10.0 | 0.00 | 11.9 | 10.7 | -0.04 |
| Gastroesophageal reflux disease | 19.0 | 14.6 | -0.12 | 19.1 | 19.7 | 0.01 | 17.8 | 19.0 | 0.03 |
| Gastrointestinal hemorrhage | 3.8 | 4.2 | 0.02 | 4.1 | 2.5 | -0.09 | 3.5 | 3.8 | 0.02 |
| Hyperlipidemia | 84.8 | 65.7 | -0.46 | 84.0 | 84.6 | 0.02 | 82.3 | 81.2 | -0.03 |
| Hypertensive disorder | 87.7 | 78.4 | -0.25 | 86.8 | 87.6 | 0.02 | 87.0 | 86.9 | 0.00 |
| Lesion of liver | 1.7 | 0.9 | -0.07 | 1.5 | 1.1 | -0.04 | 1.5 | 1.3 | -0.02 |
| Obesity | 25.9 | 11.0 | -0.39 | 22.9 | 24.1 | 0.03 | 17.8 | 18.9 | 0.03 |
| Osteoarthritis | 36.1 | 34.6 | -0.03 | 36.5 | 35.7 | -0.02 | 36.0 | 37.4 | 0.03 |
| Pneumonia | 5.0 | 6.6 | 0.07 | 4.9 | 4.3 | -0.03 | 5.5 | 6.0 | 0.02 |
| Psoriasis | 2.5 | 1.6 | -0.06 | 2.4 | 1.6 | -0.06 | 2.1 | 1.8 | -0.02 |
| Renal impairment | 14.5 | 17.8 | 0.09 | 15.1 | 13.8 | -0.04 | 18.0 | 19.8 | 0.04 |
| Rheumatoid arthritis | 1.7 | 2.0 | 0.02 | 1.6 | 1.9 | 0.02 | 1.5 | 2.0 | 0.04 |
| Ulcerative colitis | 0.5 | 0.6 | 0.02 | 0.5 | 0.7 | 0.02 | 0.4 | 0.6 | 0.03 |
| Urinary tract infectious disease | 8.1 | 12.3 | 0.14 | 8.6 | 7.9 | -0.02 | 12.3 | 11.9 | -0.01 |
| Viral hepatitis C | 0.6 | 0.3 | -0.06 | 0.6 | 0.5 | -0.01 | 0.3 | 0.3 | 0.00 |
| Visual system disorder | 57.8 | 54.7 | -0.06 | 56.9 | 58.4 | 0.03 | 57.1 | 57.5 | 0.01 |
| **Medical history: Cardiovascular disease** | | | | | | | | | |
| Atrial fibrillation | 16.7 | 16.7 | 0.00 | 16.3 | 16.1 | -0.01 | 17.1 | 18.0 | 0.02 |
| Cerebrovascular disease | 18.9 | 21.7 | 0.07 | 18.4 | 17.0 | -0.04 | 20.6 | 20.2 | -0.01 |
| Coronary arteriosclerosis | 51.9 | 43.7 | -0.16 | 47.7 | 50.1 | 0.05 | 41.8 | 45.9 | 0.08 |
| Heart disease | 71.2 | 66.6 | -0.10 | 68.1 | 68.2 | 0.00 | 66.9 | 67.5 | 0.01 |
| Heart failure | 15.3 | 14.5 | -0.02 | 13.9 | 12.5 | -0.04 | 15.5 | 14.9 | -0.02 |
| Ischemic heart disease | 21.9 | 19.4 | -0.06 | 21.4 | 20.8 | -0.01 | 21.4 | 19.9 | -0.04 |
| Peripheral vascular disease | 24.9 | 22.2 | -0.06 | 23.8 | 25.4 | 0.04 | 22.7 | 25.0 | 0.05 |
| Pulmonary embolism | 1.0 | 1.2 | 0.02 | 1.0 | 0.8 | -0.02 | 1.5 | 1.5 | 0.00 |
| Venous thrombosis | 2.2 | 2.7 | 0.03 | 2.4 | 1.6 | -0.06 | 2.1 | 2.7 | 0.04 |
| **Medical history: Neoplasms** | | | | | | | | | |
| Hematologic neoplasm | 1.7 | 2.1 | 0.03 | 1.7 | 1.7 | 0.00 | 1.2 | 2.3 | 0.09 |
| Malignant lymphoma | 1.2 | 1.1 | -0.01 | 1.1 | 1.1 | 0.00 | 0.8 | 1.1 | 0.03 |
| Malignant neoplasm of anorectum | 0.3 | 0.3 | 0.00 | 0.4 | <0.3 | -0.02 | 0.4 | 0.3 | -0.02 |
| Malignant neoplastic disease | 19.2 | 21.1 | 0.05 | 18.9 | 19.5 | 0.02 | 19.4 | 21.0 | 0.04 |
| Malignant tumor of breast | 2.2 | 2.8 | 0.04 | 2.3 | 2.2 | 0.00 | 2.5 | 2.9 | 0.02 |
| Malignant tumor of colon | 0.7 | 1.0 | 0.04 | 0.7 | 0.8 | 0.02 | 0.7 | 1.0 | 0.04 |
| Malignant tumor of lung | 0.4 | 0.8 | 0.05 | 0.5 | 1.0 | 0.06 | 0.4 | 1.0 | 0.07 |
| Malignant tumor of urinary bladder | 1.2 | 1.5 | 0.02 | 1.3 | 1.4 | 0.01 | 1.6 | 1.8 | 0.01 |
| Primary malignant neoplasm of prostate | 5.4 | 5.0 | -0.02 | 5.3 | 5.3 | 0.00 | 5.8 | 5.1 | -0.03 |
| **Medication use** | | | | | | | | | |
| Agents acting on the renin-angiotensin system | 77.6 | 77.1 | -0.01 | 77.1 | 76.5 | -0.01 | 75.6 | 76.7 | 0.03 |
| Antibacterials for systemic use | 64.8 | 66.1 | 0.03 | 65.3 | 62.8 | -0.05 | 66.0 | 65.3 | -0.01 |
| Antidepressants | 27.0 | 26.6 | -0.01 | 27.5 | 25.8 | -0.04 | 27.3 | 26.5 | -0.02 |
| Antiepileptics | 19.7 | 17.7 | -0.05 | 19.8 | 18.6 | -0.03 | 18.8 | 19.0 | 0.00 |
| Antiinflammatory and antirheumatic products | 29.9 | 29.5 | -0.01 | 30.1 | 31.9 | 0.04 | 27.2 | 30.1 | 0.06 |
| Antineoplastic agents | 8.4 | 9.3 | 0.03 | 8.7 | 8.1 | -0.02 | 9.9 | 8.3 | -0.06 |

| (Continued) | Before ajustment | | | After matching | | | After stratification | | |
|---|---|---|---|---|---|---|---|---|---|
| Characteristic | T (%) | C (%) | StdDiff | T (%) | C (%) | StdDiff | T (%) | C (%) | StdDiff |
| Antipsoriatics | 0.9 | 0.9 | 0.00 | 0.9 | 0.4 | -0.06 | 0.6 | 0.7 | 0.01 |
| Antithrombotic agents | 38.5 | 37.2 | -0.03 | 37.2 | 36.0 | -0.03 | 37.0 | 37.3 | 0.00 |
| Beta blocking agents | 61.7 | 60.9 | -0.02 | 59.7 | 60.1 | 0.01 | 58.5 | 60.9 | 0.05 |
| Calcium channel blockers | 35.9 | 38.4 | 0.05 | 35.7 | 36.2 | 0.01 | 35.3 | 38.2 | 0.06 |
| Diuretics | 53.1 | 54.1 | 0.02 | 52.2 | 50.4 | -0.04 | 51.7 | 51.9 | 0.00 |
| Drugs for acid related disorders | 39.1 | 37.7 | -0.03 | 39.1 | 38.0 | -0.02 | 39.6 | 37.9 | -0.04 |
| Drugs for obstructive airway diseases | 48.5 | 44.4 | -0.08 | 48.3 | 47.1 | -0.02 | 46.9 | 46.4 | -0.01 |
| Immunosuppressants | 3.9 | 3.5 | -0.02 | 3.8 | 3.9 | 0.01 | 3.3 | 3.8 | 0.02 |
| Lipid modifying agents | 89.2 | 84.6 | -0.14 | 87.9 | 87.3 | -0.02 | 86.3 | 85.5 | -0.02 |
| Opioids | 28.9 | 32.1 | 0.07 | 29.4 | 28.4 | -0.02 | 30.4 | 30.7 | 0.01 |
| Psycholeptics | 26.0 | 29.3 | 0.07 | 27.1 | 25.8 | -0.03 | 28.6 | 27.5 | -0.03 |
| Psychostimulants | 1.6 | 1.5 | -0.01 | 1.7 | 1.6 | -0.01 | 1.2 | 1.3 | 0.01 |

**Supplementary Table 14:** Baseline patient characteristics for SGLT2is (T) and DPP4is (C) new-users in the MDCD data source. We report proportion of initiators satisfying selected base-line characteristics and the standardized difference of population proportions (StdDiff) before and after propensity score adjustment. Less extreme StdDiffs through matching and stratification suggest improved balance between patient cohorts through adjustment.

| | Before adjustment | | | After matching | | | After stratification | | |
|---|---|---|---|---|---|---|---|---|---|
| Characteristic | T (%) | C (%) | StdDiff | T (%) | C (%) | StdDiff | T (%) | C (%) | StdDiff |
| **Age group** | | | | | | | | | |
| 20 - 24 | <0.4 | 0.2 | 0.02 | <0.4 | <0.4 | 0.05 | <0.3 | 0.2 | 0.05 |
| 25 - 29 | 1.4 | 0.5 | -0.09 | 1.5 | 0.7 | -0.08 | 1.7 | 0.5 | -0.11 |
| 30 - 34 | 2.4 | 1.6 | -0.06 | 2.5 | 1.7 | -0.05 | 2.6 | 1.6 | -0.07 |
| 35 - 39 | 5.9 | 4.2 | -0.08 | 6.0 | 5.2 | -0.03 | 5.5 | 4.6 | -0.04 |
| 40 - 44 | 8.4 | 7.0 | -0.05 | 8.4 | 7.7 | -0.03 | 7.5 | 7.5 | 0.00 |
| 45 - 49 | 14.5 | 12.1 | -0.07 | 14.4 | 13.6 | -0.02 | 15.2 | 12.5 | -0.08 |
| 50 - 54 | 17.9 | 16.3 | -0.04 | 17.4 | 16.4 | -0.03 | 17.9 | 16.0 | -0.05 |
| 55 - 59 | 22.9 | 21.2 | -0.04 | 23.1 | 22.9 | 0.00 | 23.7 | 22.1 | -0.04 |
| 60 - 64 | 18.0 | 19.4 | 0.04 | 18.1 | 18.8 | 0.02 | 17.5 | 19.7 | 0.06 |
| 65 - 69 | 4.3 | 6.8 | 0.11 | 4.3 | 5.8 | 0.07 | 4.0 | 6.2 | 0.10 |
| 70 - 74 | 2.7 | 4.2 | 0.09 | 2.7 | 2.5 | -0.01 | 2.4 | 3.6 | 0.07 |
| 75 - 79 | 0.5 | 3.2 | 0.20 | 0.6 | 1.9 | 0.12 | 0.4 | 2.7 | 0.19 |
| 80 - 84 | 0.7 | 2.2 | 0.13 | 0.7 | 1.2 | 0.05 | 0.8 | 1.7 | 0.07 |
| 85 - 89 | <0.4 | 1.1 | 0.11 | <0.4 | 1.1 | 0.10 | 0.7 | 1.0 | 0.04 |
| Gender: female | 56.4 | 61.2 | 0.10 | 56.8 | 58.7 | 0.04 | 61.4 | 60.1 | -0.03 |
| **Race** | | | | | | | | | |
| Black or African American | 22.7 | 21.9 | -0.02 | 22.9 | 20.9 | -0.05 | 23.8 | 22.9 | -0.02 |
| White | 63.6 | 62.5 | -0.02 | 63.2 | 62.8 | -0.01 | 63.1 | 61.2 | -0.04 |
| **Ethnicity** | | | | | | | | | |
| Hispanic or Latino | 1.9 | 2.0 | 0.00 | 1.8 | 2.5 | 0.05 | 1.3 | 2.2 | 0.07 |
| **Medical history: General** | | | | | | | | | |
| Acute respiratory disease | 38.7 | 39.8 | 0.02 | 38.2 | 39.4 | 0.02 | 42.1 | 39.0 | -0.06 |
| Attention deficit hyperactivity disorder | 1.9 | 1.2 | -0.06 | 1.9 | 2.1 | 0.02 | 1.8 | 1.4 | -0.03 |
| Chronic liver disease | 3.8 | 5.8 | 0.09 | 4.0 | 4.0 | 0.00 | 5.5 | 5.0 | -0.03 |
| Chronic obstructive lung disease | 29.8 | 32.3 | 0.05 | 29.6 | 29.5 | 0.00 | 31.7 | 30.7 | -0.02 |
| Crohn’s disease | 0.5 | 0.3 | -0.02 | 0.5 | <0.4 | -0.04 | 0.4 | 0.3 | -0.01 |
| Dementia | 1.6 | 3.9 | 0.14 | 1.7 | 2.9 | 0.09 | 1.4 | 3.8 | 0.15 |
| Depressive disorder | 37.3 | 36.9 | -0.01 | 36.5 | 37.8 | 0.03 | 35.8 | 37.2 | 0.03 |
| Gastroesophageal reflux disease | 31.7 | 32.2 | 0.01 | 31.9 | 32.4 | 0.01 | 31.9 | 32.4 | 0.01 |
| Gastrointestinal hemorrhage | 4.1 | 4.9 | 0.04 | 3.8 | 4.4 | 0.03 | 4.0 | 4.7 | 0.04 |
| Human immunodeficiency virus infection | 0.6 | 0.7 | 0.00 | 0.7 | 0.7 | 0.01 | 1.1 | 0.8 | -0.03 |
| Hyperlipidemia | 74.7 | 75.6 | 0.02 | 74.2 | 75.8 | 0.04 | 72.8 | 76.3 | 0.08 |
| Hypertensive disorder | 84.9 | 87.1 | 0.06 | 84.9 | 86.5 | 0.05 | 85.9 | 87.3 | 0.04 |
| Lesion of liver | 1.7 | 1.8 | 0.01 | 1.7 | 1.8 | 0.01 | 1.4 | 2.1 | 0.05 |
| Obesity | 43.6 | 36.3 | -0.15 | 42.9 | 44.2 | 0.03 | 44.4 | 40.9 | -0.07 |
| Osteoarthritis | 45.7 | 44.9 | -0.01 | 46.4 | 45.3 | -0.02 | 46.4 | 45.3 | -0.02 |
| Pneumonia | 7.6 | 10.0 | 0.08 | 7.5 | 7.7 | 0.01 | 9.8 | 9.6 | -0.01 |
| Psoriasis | 1.8 | 1.6 | -0.01 | 1.7 | 1.4 | -0.03 | 1.6 | 1.6 | 0.00 |
| Renal impairment | 12.3 | 15.2 | 0.08 | 12.5 | 12.1 | -0.01 | 13.7 | 15.4 | 0.05 |
| Rheumatoid arthritis | 2.8 | 2.7 | -0.01 | 2.8 | 2.8 | 0.00 | 4.1 | 2.7 | -0.08 |
| Schizophrenia | 3.7 | 5.0 | 0.07 | 3.7 | 3.8 | 0.01 | 4.9 | 4.6 | -0.01 |
| Ulcerative colitis | <0.4 | 0.2 | -0.01 | <0.4 | <0.4 | -0.01 | 0.9 | 0.3 | -0.08 |
| Urinary tract infectious disease | 10.6 | 14.5 | 0.12 | 10.5 | 12.0 | 0.05 | 11.9 | 13.3 | 0.04 |
| Viral hepatitis C | 2.4 | 2.6 | 0.02 | 2.3 | 2.5 | 0.01 | 2.7 | 2.7 | 0.00 |
| Visual system disorder | 48.6 | 46.6 | -0.04 | 48.5 | 48.6 | 0.00 | 48.9 | 47.1 | -0.04 |
| **Medical history: Cardiovascular disease** | | | | | | | | | |
| Atrial fibrillation | 6.5 | 7.5 | 0.04 | 6.2 | 6.5 | 0.01 | 6.4 | 7.4 | 0.04 |
| Cerebrovascular disease | 11.4 | 13.5 | 0.06 | 11.5 | 10.0 | -0.05 | 12.2 | 12.3 | 0.00 |
| Coronary arteriosclerosis | 33.6 | 34.0 | 0.01 | 32.7 | 33.7 | 0.02 | 32.1 | 33.8 | 0.04 |
| Heart disease | 55.0 | 56.9 | 0.04 | 54.0 | 55.2 | 0.03 | 54.3 | 55.5 | 0.02 |
| Heart failure | 18.1 | 17.7 | -0.01 | 17.0 | 16.8 | -0.01 | 16.4 | 17.2 | 0.02 |
| Ischemic heart disease | 22.5 | 23.2 | 0.02 | 22.2 | 23.1 | 0.02 | 20.6 | 22.8 | 0.05 |
| Peripheral vascular disease | 22.4 | 22.7 | 0.01 | 22.2 | 20.8 | -0.03 | 21.2 | 22.5 | 0.03 |
| Pulmonary embolism | 1.8 | 1.9 | 0.01 | 1.7 | 2.5 | 0.05 | 1.9 | 2.1 | 0.02 |
| Venous thrombosis | 2.4 | 3.0 | 0.03 | 2.4 | 2.0 | -0.03 | 2.6 | 2.7 | 0.01 |
| **Medical history: Neoplasms** | | | | | | | | | |
| Hematologic neoplasm | 0.8 | 1.3 | 0.05 | 0.8 | 1.1 | 0.03 | 0.8 | 1.1 | 0.03 |
| Malignant lymphoma | <0.4 | 0.7 | 0.05 | 0.5 | 0.6 | 0.01 | 0.5 | 0.6 | 0.01 |
| Malignant neoplasm of anorectum | <0.4 | 0.2 | 0.04 | <0.4 | <0.4 | 0.07 | <0.3 | 0.3 | 0.05 |
| Malignant neoplastic disease | 6.1 | 8.4 | 0.09 | 6.2 | 7.4 | 0.05 | 5.9 | 8.0 | 0.08 |
| Malignant tumor of breast | 1.6 | 2.0 | 0.03 | 1.7 | 1.7 | 0.00 | 1.7 | 1.7 | 0.00 |

| (Continued) | Before ajustment | | | After matching | | | After stratification | | |
|---|---|---|---|---|---|---|---|---|---|
| Characteristic | T (%) | C (%) | StdDiff | T (%) | C (%) | StdDiff | T (%) | C (%) | StdDiff |
| Malignant tumor of colon | <0.4 | 0.6 | 0.06 | <0.4 | 0.7 | 0.07 | <0.3 | 0.6 | 0.08 |
| Malignant tumor of lung | <0.4 | 0.7 | 0.08 | <0.4 | 0.6 | 0.07 | <0.3 | 0.6 | 0.08 |
| Malignant tumor of urinary bladder | <0.4 | 0.2 | 0.02 | <0.4 | <0.4 | -0.02 | <0.3 | 0.2 | 0.03 |
| Primary malignant neoplasm of prostate | <0.4 | 0.8 | 0.06 | <0.4 | 0.6 | 0.04 | 0.3 | 0.8 | 0.06 |
| **Medication use** | | | | | | | | | |
| Agents acting on the renin-angiotensin system | 68.7 | 71.2 | 0.05 | 68.6 | 70.0 | 0.03 | 69.8 | 69.9 | 0.00 |
| Antibacterials for systemic use | 69.6 | 71.2 | 0.04 | 69.2 | 69.2 | 0.00 | 71.4 | 69.9 | -0.03 |
| Antidepressants | 55.8 | 58.0 | 0.04 | 55.9 | 56.7 | 0.01 | 56.7 | 57.9 | 0.02 |
| Antiepileptics | 50.9 | 48.8 | -0.04 | 50.8 | 51.4 | 0.01 | 51.1 | 50.7 | -0.01 |
| Antiinflammatory and antirheumatic products | 66.3 | 59.9 | -0.13 | 66.0 | 65.8 | 0.00 | 64.7 | 63.0 | -0.04 |
| Antineoplastic agents | 5.4 | 5.5 | 0.01 | 5.3 | 6.0 | 0.03 | 5.5 | 5.2 | -0.01 |
| Antipsoriatics | 0.4 | 0.7 | 0.04 | 0.4 | 0.5 | 0.02 | <0.3 | 0.6 | 0.07 |
| Antithrombotic agents | 45.1 | 44.8 | 0.00 | 43.9 | 43.2 | -0.01 | 42.8 | 45.0 | 0.04 |
| Beta blocking agents | 52.8 | 51.4 | -0.03 | 52.3 | 49.7 | -0.05 | 51.3 | 51.1 | 0.00 |
| Calcium channel blockers | 28.2 | 32.5 | 0.09 | 28.9 | 26.8 | -0.05 | 32.1 | 31.5 | -0.01 |
| Diuretics | 53.4 | 54.6 | 0.02 | 53.0 | 51.1 | -0.04 | 55.5 | 53.0 | -0.05 |
| Drugs for acid related disorders | 55.8 | 58.2 | 0.05 | 55.8 | 56.3 | 0.01 | 57.5 | 57.3 | 0.00 |
| Drugs for obstructive airway diseases | 64.8 | 63.5 | -0.03 | 64.3 | 64.0 | -0.01 | 65.3 | 63.7 | -0.03 |
| Immunosuppressants | 3.0 | 3.2 | 0.01 | 2.8 | 4.3 | 0.08 | 3.2 | 3.5 | 0.01 |
| Lipid modifying agents | 77.6 | 78.5 | 0.02 | 77.3 | 78.6 | 0.03 | 75.5 | 78.2 | 0.06 |
| Opioids | 51.0 | 53.5 | 0.05 | 50.4 | 50.6 | 0.00 | 52.7 | 52.1 | -0.01 |
| Psycholeptics | 53.8 | 54.1 | 0.00 | 53.2 | 51.1 | -0.04 | 54.5 | 52.7 | -0.03 |
| Psychostimulants | 5.6 | 5.1 | -0.02 | 5.5 | 5.3 | -0.01 | 5.8 | 4.9 | -0.04 |

**Supplementary Table 15:** Baseline patient characteristics for SGLT2is (T) and DPP4is (C) new-users in the Open Claims data source. We report proportion of initiators satisfying selected base-line characteristics and the standardized difference of population proportions (StdDiff) before and after propensity score adjustment. Less extreme StdDiffs through matching and stratification suggest improved balance between patient cohorts through adjustment.

| | Before adjustment | | | After matching | | | After stratification | | |
|---|---|---|---|---|---|---|---|---|---|
| Characteristic | T (%) | C (%) | StdDiff | T (%) | C (%) | StdDiff | T (%) | C (%) | StdDiff |
| **Age group** | | | | | | | | | |
| 20 - 24 | 0.1 | 0.1 | 0.00 | 0.1 | 0.1 | 0.00 | 0.1 | 0.1 | 0.00 |
| 25 - 29 | 0.2 | 0.2 | 0.00 | 0.2 | 0.2 | 0.00 | 0.2 | 0.2 | 0.00 |
| 30 - 34 | 0.4 | 0.4 | -0.01 | 0.4 | 0.4 | -0.01 | 0.4 | 0.4 | -0.01 |
| 35 - 39 | 1.0 | 0.9 | -0.01 | 1.1 | 1.0 | -0.01 | 1.1 | 0.9 | -0.01 |
| 40 - 44 | 2.4 | 2.0 | -0.03 | 2.4 | 2.4 | 0.00 | 2.3 | 2.2 | -0.01 |
| 45 - 49 | 5.2 | 4.2 | -0.05 | 5.2 | 5.2 | 0.00 | 4.9 | 4.7 | -0.01 |
| 50 - 54 | 9.4 | 7.7 | -0.06 | 9.2 | 9.3 | 0.01 | 8.7 | 8.5 | 0.00 |
| 55 - 59 | 14.5 | 11.8 | -0.08 | 14.1 | 14.3 | 0.00 | 13.4 | 13.2 | -0.01 |
| 60 - 64 | 18.0 | 14.9 | -0.09 | 17.5 | 17.3 | 0.00 | 16.9 | 16.2 | -0.02 |
| 65 - 69 | 17.5 | 16.6 | -0.02 | 17.0 | 17.5 | 0.01 | 16.7 | 17.0 | 0.01 |
| 70 - 74 | 14.4 | 15.5 | 0.03 | 14.5 | 14.6 | 0.00 | 14.7 | 14.9 | 0.01 |
| 75 - 79 | 10.0 | 17.5 | 0.22 | 10.9 | 10.2 | -0.02 | 13.2 | 14.0 | 0.02 |
| 80 - 84 | 6.1 | 8.0 | 0.07 | 7.0 | 7.2 | 0.01 | 7.0 | 7.2 | 0.01 |
| 85 - 89 | 0.5 | 0.2 | -0.05 | 0.3 | 0.4 | 0.02 | 0.3 | 0.4 | 0.01 |
| Gender: female | 39.9 | 48.5 | 0.17 | 43.7 | 43.7 | 0.00 | 46.0 | 45.7 | -0.01 |
| **Medical history: General** | | | | | | | | | |
| Acute respiratory disease | 15.6 | 17.7 | 0.06 | 16.2 | 16.5 | 0.01 | 17.0 | 17.0 | 0.00 |
| Attention deficit hyperactivity disorder | 0.5 | 0.3 | -0.02 | 0.4 | 0.4 | -0.01 | 0.4 | 0.4 | -0.01 |
| Chronic liver disease | 1.3 | 1.9 | 0.05 | 1.4 | 1.4 | -0.01 | 1.8 | 1.7 | -0.01 |
| Chronic obstructive lung disease | 8.8 | 10.2 | 0.05 | 8.6 | 8.7 | 0.00 | 9.5 | 9.6 | 0.00 |
| Crohn's disease | 0.2 | 0.2 | 0.00 | 0.2 | 0.2 | 0.00 | 0.2 | 0.2 | -0.01 |
| Dementia | 0.9 | 2.3 | 0.11 | 1.7 | 1.7 | 0.00 | 1.7 | 1.7 | 0.00 |
| Depressive disorder | 8.0 | 8.8 | 0.03 | 8.3 | 8.4 | 0.00 | 8.8 | 8.6 | -0.01 |
| Gastroesophageal reflux disease | 10.6 | 11.4 | 0.03 | 10.9 | 10.9 | 0.00 | 11.5 | 11.2 | -0.01 |
| Gastrointestinal hemorrhage | 1.8 | 2.4 | 0.04 | 1.9 | 1.8 | 0.00 | 2.2 | 2.1 | 0.00 |
| Human immunodeficiency virus infection | 0.3 | 0.3 | 0.01 | 0.3 | 0.3 | 0.01 | 0.3 | 0.3 | 0.01 |
| Hyperlipidemia | 53.2 | 54.5 | 0.03 | 54.2 | 53.9 | -0.01 | 54.2 | 53.9 | -0.01 |
| Hypertensive disorder | 60.7 | 63.8 | 0.06 | 60.9 | 61.2 | 0.01 | 62.5 | 62.7 | 0.00 |
| Lesion of liver | 0.8 | 1.0 | 0.01 | 0.9 | 0.9 | 0.00 | 0.9 | 0.9 | 0.00 |
| Obesity | 13.9 | 11.3 | -0.08 | 13.2 | 13.2 | 0.00 | 12.9 | 12.6 | -0.01 |
| Osteoarthritis | 19.0 | 21.9 | 0.07 | 19.8 | 20.1 | 0.01 | 21.1 | 21.0 | 0.00 |
| Pneumonia | 3.4 | 4.2 | 0.04 | 3.3 | 3.2 | -0.01 | 3.9 | 3.8 | 0.00 |
| Psoriasis | 1.2 | 1.0 | -0.01 | 1.1 | 1.1 | 0.00 | 1.1 | 1.1 | 0.00 |
| Renal impairment | 9.5 | 12.9 | 0.11 | 9.3 | 8.8 | -0.02 | 10.9 | 11.2 | 0.01 |
| Rheumatoid arthritis | 1.3 | 1.6 | 0.03 | 1.6 | 1.5 | -0.01 | 1.6 | 1.5 | -0.01 |
| Schizophrenia | 0.3 | 0.5 | 0.03 | 0.4 | 0.4 | 0.01 | 0.4 | 0.5 | 0.01 |
| Ulcerative colitis | 0.3 | 0.3 | 0.00 | 0.3 | 0.2 | 0.00 | 0.3 | 0.2 | 0.00 |
| Urinary tract infectious disease | 5.2 | 8.6 | 0.13 | 6.0 | 6.1 | 0.00 | 7.4 | 7.3 | 0.00 |
| Viral hepatitis C | 0.4 | 0.5 | 0.02 | 0.4 | 0.4 | 0.00 | 0.4 | 0.5 | 0.00 |
| Visual system disorder | 21.8 | 25.0 | 0.07 | 22.7 | 22.7 | 0.00 | 23.8 | 23.7 | 0.00 |
| **Medical history: Cardiovascular disease** | | | | | | | | | |
| Atrial fibrillation | 8.4 | 8.8 | 0.01 | 7.5 | 7.5 | 0.00 | 8.2 | 8.3 | 0.00 |
| Cerebrovascular disease | 7.5 | 10.2 | 0.10 | 7.9 | 8.0 | 0.00 | 7.9 | 8.0 | 0.00 |
| Coronary arteriosclerosis | 28.2 | 25.3 | -0.06 | 25.1 | 24.9 | 0.00 | 25.1 | 24.9 | 0.00 |
| Heart disease | 44.2 | 42.6 | -0.03 | 40.7 | 40.4 | -0.01 | 42.3 | 42.1 | 0.00 |
| Heart failure | 11.3 | 9.6 | -0.06 | 8.5 | 8.2 | -0.01 | 9.7 | 9.3 | -0.01 |
| Ischemic heart disease | 13.9 | 12.2 | -0.05 | 11.9 | 12.0 | 0.00 | 11.9 | 12.0 | 0.00 |
| Peripheral vascular disease | 13.3 | 14.0 | 0.02 | 13.0 | 13.2 | 0.00 | 13.8 | 13.7 | 0.00 |
| Pulmonary embolism | 0.8 | 0.9 | 0.01 | 0.8 | 0.8 | 0.01 | 0.9 | 0.9 | 0.01 |
| Venous thrombosis | 1.3 | 1.6 | 0.03 | 1.3 | 1.3 | 0.00 | 1.4 | 1.5 | 0.01 |
| **Medical history: Neoplasms** | | | | | | | | | |
| Hematologic neoplasm | 0.8 | 1.1 | 0.03 | 0.9 | 0.9 | 0.00 | 1.0 | 1.0 | 0.01 |
| Malignant lymphoma | 0.5 | 0.6 | 0.01 | 0.5 | 0.5 | 0.00 | 0.6 | 0.6 | 0.00 |
| Malignant neoplasm of anorectum | 0.1 | 0.2 | 0.01 | 0.2 | 0.2 | 0.00 | 0.2 | 0.2 | 0.00 |
| Malignant neoplastic disease | 7.5 | 9.4 | 0.07 | 7.8 | 7.9 | 0.01 | 8.4 | 8.7 | 0.01 |
| Malignant tumor of breast | 1.1 | 1.6 | 0.04 | 1.3 | 1.3 | 0.00 | 1.4 | 1.4 | 0.00 |
| Malignant tumor of colon | 0.3 | 0.5 | 0.03 | 0.4 | 0.4 | 0.00 | 0.4 | 0.4 | 0.00 |
| Malignant tumor of lung | 0.3 | 0.5 | 0.03 | 0.3 | 0.4 | 0.00 | 0.4 | 0.4 | 0.01 |
| Malignant tumor of urinary bladder | 0.3 | 0.5 | 0.03 | 0.4 | 0.4 | 0.01 | 0.4 | 0.5 | 0.00 |
| Primary malignant neoplasm of prostate | 1.6 | 1.9 | 0.02 | 1.6 | 1.6 | 0.00 | 1.7 | 1.8 | 0.01 |
| **Medication use** | | | | | | | | | |

| (Continued) | Before ajustment | | | After matching | | | After stratification | | |
|---|---|---|---|---|---|---|---|---|---|
| Characteristic | T (%) | C (%) | StdDiff | T (%) | C (%) | StdDiff | T (%) | C (%) | StdDiff |
| Agents acting on the renin-angiotensin system | 74.6 | 73.8 | -0.02 | 73.1 | 73.2 | 0.00 | 73.4 | 73.6 | 0.00 |
| Antibacterials for systemic use | 58.9 | 62.5 | 0.07 | 60.4 | 60.8 | 0.01 | 61.7 | 61.4 | 0.00 |
| Antidepressants | 32.1 | 32.9 | 0.02 | 32.5 | 32.3 | 0.00 | 33.4 | 32.7 | -0.01 |
| Antiepileptics | 23.4 | 25.3 | 0.04 | 24.0 | 24.3 | 0.01 | 24.8 | 24.8 | 0.00 |
| Antiinflammatory and antirheumatic products | 39.5 | 40.2 | 0.01 | 40.0 | 40.5 | 0.01 | 40.8 | 40.3 | -0.01 |
| Antineoplastic agents | 5.9 | 6.3 | 0.02 | 5.9 | 6.0 | 0.00 | 6.2 | 6.2 | 0.00 |
| Antithrombotic agents | 38.2 | 37.2 | -0.02 | 35.6 | 35.4 | 0.00 | 36.8 | 36.7 | 0.00 |
| Beta blocking agents | 55.8 | 53.3 | -0.05 | 52.6 | 52.2 | -0.01 | 53.1 | 53.1 | 0.00 |
| Calcium channel blockers | 31.9 | 34.1 | 0.05 | 32.1 | 32.2 | 0.00 | 32.4 | 33.1 | 0.01 |
| Diuretics | 47.8 | 47.9 | 0.00 | 46.2 | 45.7 | -0.01 | 47.5 | 47.1 | -0.01 |
| Drugs for acid related disorders | 40.6 | 43.8 | 0.06 | 41.1 | 41.3 | 0.00 | 42.8 | 42.7 | 0.00 |
| Drugs for obstructive airway diseases | 42.7 | 43.8 | 0.02 | 43.1 | 43.3 | 0.00 | 43.9 | 43.7 | 0.00 |
| Immunosuppressants | 3.4 | 3.7 | 0.02 | 3.5 | 3.6 | 0.01 | 3.5 | 3.6 | 0.01 |
| Lipid modifying agents | 82.5 | 80.0 | -0.06 | 80.9 | 80.5 | -0.01 | 80.5 | 80.4 | 0.00 |
| Opioids | 28.2 | 31.6 | 0.07 | 29.3 | 29.4 | 0.00 | 30.7 | 30.4 | -0.01 |
| Psycholeptics | 26.2 | 28.6 | 0.06 | 28.2 | 27.8 | -0.01 | 28.2 | 27.8 | -0.01 |
| Psychostimulants | 2.7 | 2.5 | -0.01 | 2.7 | 2.6 | -0.01 | 2.7 | 2.6 | -0.01 |

**Supplementary Table 16:** Baseline patient characteristics for SGLT2is (T) and DPP4is (C) new-users in the OptumEHR data source. We report proportion of initiators satisfying selected base-line characteristics and the standardized difference of population proportions (StdDiff) before and after propensity score adjustment. Less extreme StdDiffs through matching and stratification suggest improved balance between patient cohorts through adjustment.

| | Before adjustment | | | After matching | | | After stratification | | |
|---|---|---|---|---|---|---|---|---|---|
| Characteristic | T (%) | C (%) | StdDiff | T (%) | C (%) | StdDiff | T (%) | C (%) | StdDiff |
| **Age group** | | | | | | | | | |
| 25 - 29 | 0.1 | 0.1 | 0.00 | 0.1 | 0.1 | -0.02 | 0.2 | 0.1 | -0.03 |
| 30 - 34 | 0.4 | 0.3 | -0.02 | 0.4 | 0.3 | -0.01 | 0.3 | 0.3 | -0.01 |
| 35 - 39 | 1.2 | 1.0 | -0.02 | 1.1 | 1.1 | 0.00 | 1.1 | 1.0 | -0.01 |
| 40 - 44 | 3.3 | 2.2 | -0.07 | 3.1 | 2.7 | -0.02 | 2.8 | 2.5 | -0.02 |
| 45 - 49 | 6.6 | 5.1 | -0.06 | 6.3 | 6.0 | -0.01 | 5.6 | 5.4 | -0.01 |
| 50 - 54 | 10.8 | 8.9 | -0.06 | 10.5 | 10.3 | -0.01 | 9.9 | 9.5 | -0.01 |
| 55 - 59 | 17.1 | 13.5 | -0.10 | 16.3 | 16.7 | 0.01 | 15.1 | 15.0 | 0.00 |
| 60 - 64 | 20.8 | 16.4 | -0.11 | 19.9 | 20.4 | 0.01 | 18.2 | 18.4 | 0.00 |
| 65 - 69 | 16.2 | 16.7 | 0.01 | 16.3 | 16.7 | 0.01 | 16.4 | 16.5 | 0.00 |
| 70 - 74 | 12.5 | 14.3 | 0.05 | 13.1 | 13.3 | 0.01 | 12.9 | 13.6 | 0.02 |
| 75 - 79 | 6.8 | 11.6 | 0.17 | 7.8 | 7.6 | -0.01 | 9.0 | 9.3 | 0.01 |
| 80 - 84 | 3.1 | 7.6 | 0.20 | 3.8 | 3.5 | -0.01 | 6.4 | 6.2 | -0.01 |
| 85 - 89 | 1.0 | 2.3 | 0.10 | 1.2 | 1.3 | 0.01 | 1.9 | 2.1 | 0.02 |
| Gender: female | 35.4 | 43.2 | 0.16 | 38.2 | 37.6 | -0.01 | 42.0 | 40.6 | -0.03 |
| **Race** | | | | | | | | | |
| Asian | 1.9 | 1.9 | 0.00 | 1.9 | 2.1 | 0.01 | 2.0 | 2.0 | 0.00 |
| Black or African American | 8.1 | 9.5 | 0.05 | 8.5 | 9.0 | 0.02 | 9.2 | 9.3 | 0.00 |
| White | 83.7 | 82.3 | -0.04 | 83.2 | 82.4 | -0.02 | 82.4 | 82.3 | 0.00 |
| **Ethnicity** | | | | | | | | | |
| Hispanic or Latino | 4.5 | 4.8 | 0.01 | 4.5 | 4.7 | 0.01 | 4.4 | 4.7 | 0.02 |
| Not Hispanic or Latino | 86.3 | 87.8 | 0.05 | 86.7 | 86.3 | -0.01 | 87.3 | 87.0 | -0.01 |
| **Medical history: General** | | | | | | | | | |
| Acute respiratory disease | 19.3 | 20.0 | 0.02 | 19.6 | 19.4 | 0.00 | 19.8 | 19.8 | 0.00 |
| Attention deficit hyperactivity disorder | 0.6 | 0.4 | -0.03 | 0.6 | 0.5 | -0.02 | 0.5 | 0.5 | -0.01 |
| Chronic liver disease | 2.2 | 3.1 | 0.06 | 2.4 | 2.2 | -0.02 | 2.8 | 2.6 | -0.02 |
| Chronic obstructive lung disease | 9.9 | 11.5 | 0.05 | 10.2 | 9.9 | -0.01 | 10.6 | 11.1 | 0.02 |
| Crohn’s disease | 0.3 | 0.3 | 0.00 | 0.3 | 0.3 | -0.01 | 0.3 | 0.3 | -0.01 |
| Dementia | 0.8 | 2.1 | 0.11 | 0.9 | 1.0 | 0.01 | 1.3 | 1.7 | 0.04 |
| Depressive disorder | 14.7 | 15.0 | 0.01 | 15.0 | 14.6 | -0.01 | 15.8 | 15.1 | -0.02 |
| Gastrointestinal hemorrhage | 1.6 | 2.2 | 0.05 | 1.6 | 2.1 | 0.04 | 1.7 | 2.1 | 0.03 |
| Human immunodeficiency virus infection | 0.2 | 0.2 | 0.00 | 0.2 | 0.2 | 0.00 | 0.2 | 0.2 | -0.01 |
| Hyperlipidemia | 78.1 | 77.3 | -0.02 | 77.2 | 77.3 | 0.00 | 77.2 | 77.3 | 0.00 |
| Hypertensive disorder | 78.9 | 79.2 | 0.01 | 78.4 | 79.0 | 0.01 | 79.7 | 79.4 | -0.01 |
| Lesion of liver | 0.9 | 1.0 | 0.01 | 0.9 | 0.8 | -0.01 | 1.1 | 1.0 | -0.01 |
| Obesity | 33.2 | 25.8 | -0.16 | 30.8 | 31.4 | 0.01 | 29.9 | 29.3 | -0.01 |
| Osteoarthritis | 20.9 | 22.6 | 0.04 | 21.1 | 21.2 | 0.00 | 22.4 | 22.2 | 0.00 |
| Pneumonia | 2.4 | 3.1 | 0.04 | 2.4 | 2.5 | 0.00 | 2.6 | 2.9 | 0.02 |
| Psoriasis | 1.7 | 1.5 | -0.02 | 1.7 | 1.7 | 0.00 | 1.7 | 1.6 | -0.01 |
| Renal impairment | 8.7 | 12.6 | 0.13 | 9.4 | 9.1 | -0.01 | 11.2 | 11.4 | 0.01 |
| Rheumatoid arthritis | 1.6 | 1.5 | -0.01 | 1.8 | 1.4 | -0.03 | 2.2 | 1.5 | -0.05 |
| Schizophrenia | 0.2 | 0.3 | 0.03 | 0.2 | 0.2 | 0.01 | 0.1 | 0.3 | 0.03 |
| Ulcerative colitis | 0.3 | 0.3 | -0.01 | 0.4 | 0.4 | 0.00 | 0.3 | 0.3 | 0.00 |
| Urinary tract infectious disease | 3.9 | 6.9 | 0.13 | 4.4 | 4.6 | 0.01 | 6.2 | 5.8 | -0.01 |
| Viral hepatitis C | 0.4 | 0.6 | 0.03 | 0.4 | 0.5 | 0.02 | 0.4 | 0.6 | 0.03 |
| Visual system disorder | 13.0 | 15.2 | 0.06 | 13.4 | 13.5 | 0.00 | 15.1 | 14.5 | -0.01 |
| **Medical history: Cardiovascular disease** | | | | | | | | | |
| Atrial fibrillation | 8.8 | 10.7 | 0.06 | 8.9 | 8.9 | 0.00 | 9.6 | 10.0 | 0.01 |
| Cerebrovascular disease | 9.1 | 11.4 | 0.07 | 9.6 | 9.1 | -0.01 | 10.6 | 10.2 | -0.01 |
| Coronary arteriosclerosis | 47.4 | 42.6 | -0.10 | 44.1 | 44.5 | 0.01 | 42.0 | 43.3 | 0.03 |
| Heart disease | 61.8 | 59.5 | -0.05 | 59.3 | 59.3 | 0.00 | 59.1 | 59.3 | 0.00 |
| Heart failure | 9.9 | 9.2 | -0.03 | 8.8 | 8.1 | -0.03 | 9.5 | 9.0 | -0.02 |
| Ischemic heart disease | 22.0 | 19.2 | -0.07 | 20.3 | 20.6 | 0.01 | 20.3 | 20.0 | -0.01 |
| Peripheral vascular disease | 20.3 | 17.7 | -0.06 | 18.9 | 19.6 | 0.02 | 19.1 | 18.9 | -0.01 |
| Pulmonary embolism | 0.9 | 0.9 | 0.00 | 1.1 | 0.9 | -0.03 | 1.1 | 0.9 | -0.03 |
| Venous thrombosis | 1.8 | 1.9 | 0.01 | 1.8 | 1.8 | 0.00 | 2.3 | 1.7 | -0.04 |
| **Medical history: Neoplasms** | | | | | | | | | |
| Hematologic neoplasm | 0.9 | 1.2 | 0.02 | 1.0 | 1.1 | 0.01 | 1.1 | 1.1 | 0.00 |
| Malignant lymphoma | 0.5 | 0.7 | 0.03 | 0.5 | 0.5 | 0.00 | 0.6 | 0.7 | 0.00 |
| Malignant neoplasm of anorectum | 0.1 | 0.1 | 0.03 | 0.1 | 0.1 | 0.01 | <0.1 | 0.1 | 0.03 |
| Malignant neoplastic disease | 7.1 | 9.5 | 0.09 | 7.5 | 7.8 | 0.01 | 8.6 | 8.7 | 0.00 |
| Malignant tumor of breast | 1.0 | 1.5 | 0.04 | 1.1 | 1.1 | 0.00 | 1.4 | 1.3 | -0.01 |

| (Continued) | Before ajustment | | | After matching | | | After stratification | | |
|---|---|---|---|---|---|---|---|---|---|
| Characteristic | T (%) | C (%) | StdDiff | T (%) | C (%) | StdDiff | T (%) | C (%) | StdDiff |
| Malignant tumor of colon | 0.4 | 0.4 | 0.01 | 0.3 | 0.4 | 0.01 | 0.4 | 0.4 | 0.00 |
| Malignant tumor of lung | 0.2 | 0.5 | 0.04 | 0.3 | 0.3 | 0.02 | 0.2 | 0.4 | 0.03 |
| Malignant tumor of urinary bladder | 0.4 | 0.5 | 0.03 | 0.4 | 0.4 | 0.00 | 0.4 | 0.5 | 0.01 |
| Primary malignant neoplasm of prostate | 1.7 | 2.0 | 0.02 | 1.7 | 1.8 | 0.01 | 1.6 | 1.8 | 0.01 |
| **Medication use** | | | | | | | | | |
| Agents acting on the renin-angiotensin system | 75.2 | 73.7 | -0.03 | 74.3 | 73.9 | -0.01 | 74.3 | 73.9 | -0.01 |
| Antibacterials for systemic use | 45.0 | 47.0 | 0.04 | 45.6 | 45.7 | 0.00 | 47.6 | 46.3 | -0.03 |
| Antidepressants | 31.9 | 32.8 | 0.02 | 32.6 | 32.5 | 0.00 | 33.6 | 32.8 | -0.02 |
| Antiepileptics | 21.1 | 22.1 | 0.03 | 21.8 | 22.2 | 0.01 | 23.1 | 22.4 | -0.02 |
| Antiinflammatory and antirheumatic products | 71.6 | 69.1 | -0.06 | 70.7 | 70.6 | 0.00 | 70.0 | 70.3 | 0.01 |
| Antineoplastic agents | 4.9 | 5.2 | 0.01 | 5.1 | 5.0 | -0.01 | 6.0 | 5.0 | -0.04 |
| Antipsoriatics | 0.4 | 0.8 | 0.05 | 0.4 | 0.6 | 0.03 | 0.5 | 0.7 | 0.04 |
| Antithrombotic agents | 68.3 | 67.1 | -0.03 | 67.3 | 67.0 | 0.00 | 67.1 | 67.3 | 0.00 |
| Beta blocking agents | 58.9 | 57.5 | -0.03 | 57.3 | 57.5 | 0.00 | 57.1 | 57.9 | 0.02 |
| Calcium channel blockers | 30.2 | 31.7 | 0.03 | 30.1 | 30.3 | 0.00 | 30.9 | 31.3 | 0.01 |
| Diuretics | 45.2 | 47.8 | 0.05 | 45.1 | 44.7 | -0.01 | 46.9 | 46.2 | -0.01 |
| Drugs for acid related disorders | 46.4 | 48.1 | 0.03 | 47.1 | 46.0 | -0.02 | 48.6 | 47.4 | -0.02 |
| Drugs for obstructive airway diseases | 46.3 | 47.3 | 0.02 | 46.8 | 46.8 | 0.00 | 47.8 | 47.5 | 0.00 |
| Immunosuppressants | 3.1 | 2.9 | -0.01 | 3.2 | 3.0 | -0.01 | 3.4 | 2.9 | -0.03 |
| Lipid modifying agents | 86.7 | 83.6 | -0.09 | 85.6 | 85.4 | -0.01 | 84.0 | 84.7 | 0.02 |
| Opioids | 31.7 | 34.8 | 0.06 | 32.2 | 32.7 | 0.01 | 34.2 | 33.8 | -0.01 |
| Psycholeptics | 33.4 | 35.6 | 0.05 | 33.8 | 33.6 | 0.00 | 35.3 | 35.1 | 0.00 |
| Psychostimulants | 3.4 | 2.5 | -0.05 | 3.4 | 3.1 | -0.02 | 3.1 | 2.8 | -0.02 |

**Supplementary Table 17:** Baseline patient characteristics for SGLT2is (T) and DPP4is (C) new-users in the SIDIAP data source. We report proportion of initiators satisfying selected base-line characteristics and the standardized difference of population proportions (StdDiff) before and after propensity score adjustment. Less extreme StdDiffs through matching and stratification suggest improved balance between patient cohorts through adjustment.

| | Before adjustment | | | After matching | | | After stratification | | |
|---|---|---|---|---|---|---|---|---|---|
| Characteristic | T (%) | C (%) | StdDiff | T (%) | C (%) | StdDiff | T (%) | C (%) | StdDiff |
| **Age group** | | | | | | | | | |
| 30 - 34 | <0.2 | <0.1 | 0.00 | <0.2 | <0.1 | -0.03 | <0.2 | <0.1 | -0.03 |
| 35 - 39 | <0.2 | 0.1 | 0.03 | <0.2 | <0.3 | 0.06 | <0.2 | 0.1 | 0.04 |
| 40 - 44 | 1.0 | 0.4 | -0.06 | 1.1 | 0.4 | -0.08 | 1.2 | 0.3 | -0.10 |
| 45 - 49 | 2.8 | 1.9 | -0.06 | 2.9 | 2.6 | -0.02 | 2.6 | 1.7 | -0.07 |
| 50 - 54 | 6.3 | 4.0 | -0.10 | 5.7 | 5.9 | 0.01 | 4.2 | 4.2 | 0.00 |
| 55 - 59 | 11.9 | 7.8 | -0.14 | 10.8 | 10.2 | -0.02 | 9.7 | 8.7 | -0.03 |
| 60 - 64 | 16.6 | 12.9 | -0.10 | 16.0 | 14.5 | -0.04 | 13.6 | 13.1 | -0.01 |
| 65 - 69 | 18.9 | 14.9 | -0.11 | 18.2 | 18.4 | 0.00 | 15.4 | 15.6 | 0.00 |
| 70 - 74 | 17.7 | 16.4 | -0.03 | 18.3 | 20.2 | 0.05 | 19.7 | 17.7 | -0.05 |
| 75 - 79 | 13.6 | 15.7 | 0.06 | 14.2 | 13.6 | -0.02 | 14.2 | 15.0 | 0.02 |
| 80 - 84 | 7.3 | 13.2 | 0.19 | 8.0 | 8.9 | 0.03 | 8.3 | 11.9 | 0.12 |
| 85 - 89 | 3.0 | 9.0 | 0.25 | 3.6 | 3.3 | -0.02 | 8.2 | 8.3 | 0.00 |
| 90 - 94 | 0.7 | 3.1 | 0.18 | 0.9 | 1.4 | 0.05 | 1.6 | 2.8 | 0.08 |
| 95 - 99 | <0.2 | 0.5 | 0.06 | 0.2 | <0.3 | 0.03 | 1.3 | 0.5 | -0.09 |
| Gender: female | 21.0 | 28.4 | 0.17 | 23.9 | 24.6 | 0.02 | 31.8 | 27.3 | -0.10 |
| **Medical history: General** | | | | | | | | | |
| Acute respiratory disease | 13.2 | 13.9 | 0.02 | 13.2 | 13.6 | 0.01 | 16.5 | 14.5 | -0.06 |
| Chronic liver disease | 0.3 | 0.3 | 0.01 | 0.3 | <0.3 | -0.04 | 0.3 | 0.2 | -0.01 |
| Chronic obstructive lung disease | 2.3 | 2.4 | 0.01 | 2.0 | 1.5 | -0.04 | 1.6 | 2.2 | 0.04 |
| Dementia | 0.3 | 1.0 | 0.08 | 0.4 | 0.6 | 0.04 | 0.3 | 0.7 | 0.05 |
| Depressive disorder | 1.0 | 2.6 | 0.11 | 1.0 | 1.1 | 0.01 | 1.4 | 2.0 | 0.05 |
| Gastroesophageal reflux disease | 1.4 | 1.3 | 0.00 | 1.4 | 1.8 | 0.03 | 1.0 | 1.8 | 0.07 |
| Gastrointestinal hemorrhage | 1.3 | 2.1 | 0.07 | 1.2 | 1.8 | 0.05 | 1.2 | 2.1 | 0.07 |
| Hyperlipidemia | 1.9 | 2.7 | 0.05 | 2.0 | 1.2 | -0.06 | 2.9 | 1.9 | -0.06 |
| Hypertensive disorder | 3.2 | 3.7 | 0.03 | 3.1 | 2.5 | -0.03 | 3.6 | 2.8 | -0.04 |
| Lesion of liver | <0.2 | 0.4 | 0.04 | <0.2 | <0.3 | 0.03 | <0.2 | 0.3 | 0.06 |
| Obesity | 5.9 | 6.7 | 0.03 | 5.9 | 5.6 | -0.01 | 7.1 | 6.0 | -0.04 |
| Osteoarthritis | 3.4 | 5.3 | 0.09 | 3.6 | 3.0 | -0.03 | 4.6 | 4.5 | -0.01 |
| Pneumonia | 2.2 | 2.3 | 0.01 | 2.1 | 1.9 | -0.02 | 2.8 | 2.1 | -0.04 |
| Psoriasis | 0.2 | 0.3 | 0.02 | 0.2 | <0.3 | 0.01 | <0.2 | 0.3 | 0.03 |
| Renal impairment | 2.0 | 6.8 | 0.23 | 2.5 | 2.1 | -0.03 | 3.0 | 5.9 | 0.14 |
| Rheumatoid arthritis | <0.2 | 0.1 | -0.01 | 0.2 | <0.3 | -0.04 | 0.4 | <0.1 | -0.07 |
| Urinary tract infectious disease | 4.8 | 7.5 | 0.11 | 5.5 | 5.9 | 0.02 | 8.0 | 7.8 | -0.01 |
| Viral hepatitis C | <0.2 | 0.1 | 0.02 | <0.2 | <0.3 | -0.02 | 0.2 | 0.1 | -0.02 |
| Visual system disorder | 13.3 | 17.8 | 0.12 | 14.8 | 15.2 | 0.01 | 15.1 | 17.3 | 0.06 |
| **Medical history: Cardiovascular disease** | | | | | | | | | |
| Atrial fibrillation | 3.3 | 2.9 | -0.02 | 2.6 | 2.6 | 0.00 | 3.5 | 2.9 | -0.04 |
| Cerebrovascular disease | 2.4 | 6.4 | 0.20 | 2.9 | 3.1 | 0.01 | 4.2 | 5.0 | 0.04 |
| Heart disease | 31.4 | 24.0 | -0.17 | 21.6 | 23.9 | 0.06 | 21.2 | 23.2 | 0.05 |
| Heart failure | 5.0 | 4.3 | -0.03 | 3.4 | 3.3 | -0.01 | 3.7 | 4.1 | 0.02 |
| Ischemic heart disease | 22.4 | 14.7 | -0.20 | 13.7 | 15.6 | 0.06 | 14.3 | 14.6 | 0.01 |
| Peripheral vascular disease | 3.5 | 5.2 | 0.08 | 4.1 | 2.9 | -0.06 | 3.8 | 4.3 | 0.03 |
| Pulmonary embolism | <0.2 | 0.2 | 0.02 | <0.2 | <0.3 | 0.01 | 0.2 | 0.2 | 0.00 |
| Venous thrombosis | 0.5 | 1.1 | 0.07 | 0.4 | 0.5 | 0.01 | 0.4 | 0.9 | 0.06 |
| **Medical history: Neoplasms** | | | | | | | | | |
| Hematologic neoplasm | <0.2 | 0.2 | 0.04 | <0.2 | <0.3 | 0.04 | <0.2 | 0.2 | 0.04 |
| Malignant neoplasm of anorectum | <0.2 | 0.1 | 0.01 | <0.2 | <0.3 | 0.02 | 0.2 | 0.1 | -0.01 |
| Malignant neoplastic disease | 2.1 | 3.9 | 0.11 | 2.4 | 2.9 | 0.03 | 3.3 | 3.5 | 0.01 |
| Malignant tumor of breast | <0.2 | 0.1 | 0.03 | <0.2 | <0.3 | 0.03 | <0.2 | 0.1 | 0.04 |
| Malignant tumor of colon | <0.2 | 0.5 | 0.06 | <0.2 | 0.4 | 0.04 | 0.2 | 0.4 | 0.05 |
| Malignant tumor of lung | <0.2 | 0.1 | 0.01 | <0.2 | <0.1 | 0.03 | <0.2 | <0.1 | 0.03 |
| Malignant tumor of urinary bladder | <0.2 | 0.5 | 0.07 | <0.2 | <0.3 | 0.00 | <0.2 | 0.4 | 0.04 |
| Primary malignant neoplasm of prostate | <0.2 | 0.4 | 0.04 | 0.3 | <0.3 | -0.03 | 0.2 | 0.3 | 0.01 |
| **Medication use** | | | | | | | | | |
| Agents acting on the renin-angiotensin system | 82.4 | 75.2 | -0.18 | 77.5 | 78.1 | 0.01 | 73.5 | 77.2 | 0.09 |
| Antibacterials for systemic use | 39.5 | 43.5 | 0.08 | 41.0 | 40.1 | -0.02 | 44.4 | 43.3 | -0.02 |
| Antidepressants | 19.1 | 23.1 | 0.10 | 21.0 | 21.9 | 0.02 | 25.1 | 23.1 | -0.05 |
| Antiepileptics | 11.0 | 11.8 | 0.03 | 12.0 | 12.9 | 0.03 | 13.3 | 12.0 | -0.04 |
| Antiinflammatory and antirheumatic products | 82.2 | 74.9 | -0.18 | 79.6 | 81.4 | 0.04 | 80.1 | 76.7 | -0.08 |
| Antineoplastic agents | 1.4 | 2.2 | 0.05 | 1.7 | 2.5 | 0.06 | 1.3 | 2.2 | 0.07 |
| Antipsoriatics | 1.7 | 2.0 | 0.02 | 1.8 | 1.6 | -0.02 | 2.1 | 2.0 | -0.01 |

| (Continued) | Before ajustment | | | After matching | | | After stratification | | |
|---|---|---|---|---|---|---|---|---|---|
| Characteristic | T (%) | C (%) | StdDiff | T (%) | C (%) | StdDiff | T (%) | C (%) | StdDiff |
| Antithrombotic agents | 91.9 | 85.5 | -0.20 | 88.6 | 89.8 | 0.04 | 83.6 | 88.1 | 0.13 |
| Beta blocking agents | 72.6 | 57.3 | -0.33 | 65.6 | 67.9 | 0.05 | 61.0 | 61.7 | 0.01 |
| Calcium channel blockers | 33.1 | 36.6 | 0.07 | 34.7 | 35.1 | 0.01 | 34.5 | 36.3 | 0.04 |
| Diuretics | 55.7 | 56.9 | 0.03 | 52.4 | 52.8 | 0.01 | 54.3 | 57.2 | 0.06 |
| Drugs for acid related disorders | 76.4 | 75.2 | -0.03 | 73.4 | 74.0 | 0.01 | 76.2 | 75.9 | -0.01 |
| Drugs for obstructive airway diseases | 32.1 | 32.3 | 0.00 | 32.4 | 31.6 | -0.02 | 32.7 | 32.8 | 0.00 |
| Immunosuppressants | 1.6 | 1.6 | 0.00 | 1.8 | 1.7 | -0.01 | 1.4 | 1.8 | 0.04 |
| Lipid modifying agents | 92.0 | 84.5 | -0.23 | 89.6 | 91.3 | 0.06 | 87.0 | 86.5 | -0.01 |
| Opioids | 20.0 | 20.5 | 0.01 | 20.5 | 21.2 | 0.02 | 21.8 | 21.0 | -0.02 |
| Psycholeptics | 30.2 | 35.9 | 0.12 | 30.9 | 31.4 | 0.01 | 36.6 | 34.2 | -0.05 |
| Psychostimulants | 24.0 | 13.7 | -0.26 | 18.8 | 19.3 | 0.01 | 16.3 | 16.6 | 0.01 |

**Supplementary Table 18:** Baseline patient characteristics for SGLT2is (T) and DPP4is (C) new-users in the VA data source. We report proportion of initiators satisfying selected base-line characteristics and the standardized difference of population proportions (StdDiff) before and after propensity score adjustment. Less extreme StdDiffs through matching and stratification suggest improved balance between patient cohorts through adjustment.

| | Before adjustment | | | After matching | | | After stratification | | |
|---|---|---|---|---|---|---|---|---|---|
| Characteristic | T (%) | C (%) | StdDiff | T (%) | C (%) | StdDiff | T (%) | C (%) | StdDiff |
| **Age group** | | | | | | | | | |
| 30 - 34 | <0.1 | 0.1 | 0.03 | <0.1 | <0.3 | 0.01 | <0.1 | <0.1 | 0.03 |
| 35 - 39 | 0.2 | 0.3 | 0.02 | 0.3 | 0.3 | 0.00 | 0.3 | 0.4 | 0.01 |
| 40 - 44 | 0.6 | 0.8 | 0.02 | 0.7 | 0.9 | 0.03 | 0.7 | 0.8 | 0.02 |
| 45 - 49 | 2.1 | 2.0 | -0.01 | 2.1 | 2.1 | 0.00 | 2.0 | 1.9 | 0.00 |
| 50 - 54 | 4.7 | 4.8 | 0.00 | 4.7 | 5.4 | 0.03 | 4.3 | 5.1 | 0.04 |
| 55 - 59 | 8.0 | 8.1 | 0.00 | 8.2 | 8.1 | 0.00 | 8.8 | 7.9 | -0.03 |
| 60 - 64 | 13.4 | 12.2 | -0.04 | 12.6 | 12.2 | -0.01 | 12.9 | 12.3 | -0.02 |
| 65 - 69 | 17.7 | 20.9 | 0.08 | 17.9 | 17.8 | 0.00 | 20.4 | 19.1 | -0.03 |
| 70 - 74 | 31.9 | 27.2 | -0.10 | 32.1 | 31.0 | -0.03 | 30.2 | 29.5 | -0.01 |
| 75 - 79 | 14.5 | 11.7 | -0.08 | 13.5 | 13.4 | 0.00 | 12.1 | 12.7 | 0.02 |
| 80 - 84 | 4.3 | 6.5 | 0.10 | 4.9 | 5.1 | 0.01 | 5.0 | 5.6 | 0.03 |
| 85 - 89 | 1.8 | 3.8 | 0.12 | 2.3 | 2.4 | 0.01 | 2.9 | 3.0 | 0.01 |
| 90 - 94 | 0.4 | 1.3 | 0.10 | 0.5 | 0.9 | 0.05 | 0.4 | 1.1 | 0.07 |
| 95 - 99 | <0.1 | 0.3 | 0.06 | 0.1 | 0.3 | 0.05 | <0.1 | 0.4 | 0.07 |
| 100 - 104 | <0.1 | <0.1 | -0.01 | <0.1 | <0.3 | -0.01 | <0.1 | <0.1 | -0.01 |
| Gender: female | 3.4 | 5.2 | 0.09 | 4.1 | 4.2 | 0.00 | 4.8 | 4.4 | -0.02 |
| **Race** | | | | | | | | | |
| Asian | 1.0 | 1.1 | 0.01 | 0.8 | 0.8 | 0.00 | 0.7 | 0.9 | 0.02 |
| Black or African American | 13.6 | 13.6 | 0.00 | 12.7 | 12.9 | 0.00 | 14.8 | 12.4 | -0.07 |
| White | 78.1 | 78.1 | 0.00 | 78.9 | 79.0 | 0.00 | 77.8 | 79.4 | 0.04 |
| Unknown | 5.5 | 5.6 | 0.00 | 5.8 | 5.4 | -0.02 | 5.2 | 5.3 | 0.01 |
| Native Hawaiian or Other Pacific Islander | 0.8 | 1.0 | 0.02 | 0.8 | 1.1 | 0.02 | 0.8 | 1.1 | 0.03 |
| American Indian or Alaska Native | 0.9 | 0.7 | -0.02 | 0.9 | 0.8 | -0.02 | 0.8 | 0.8 | 0.01 |
| **Ethnicity** | | | | | | | | | |
| Hispanic or Latino | 5.7 | 4.4 | -0.06 | 5.3 | 5.1 | -0.01 | 4.7 | 5.0 | 0.02 |
| Not Hispanic or Latino | 91.9 | 93.4 | 0.06 | 92.2 | 92.6 | 0.02 | 92.8 | 93.0 | 0.00 |
| **Medical history: General** | | | | | | | | | |
| Acute respiratory disease | 9.8 | 9.9 | 0.00 | 9.6 | 9.1 | -0.02 | 10.6 | 9.4 | -0.04 |
| Attention deficit hyperactivity disorder | 0.5 | 0.5 | 0.00 | 0.4 | 0.4 | -0.01 | 0.4 | 0.5 | 0.01 |
| Chronic liver disease | 2.4 | 2.2 | -0.01 | 2.3 | 1.8 | -0.03 | 2.6 | 2.1 | -0.03 |
| Chronic obstructive lung disease | 18.5 | 17.2 | -0.03 | 17.2 | 18.1 | 0.02 | 17.5 | 17.6 | 0.00 |
| Crohn’s disease | 0.2 | 0.2 | 0.00 | 0.3 | <0.3 | -0.03 | 0.2 | 0.1 | -0.02 |
| Dementia | 1.6 | 2.8 | 0.09 | 1.7 | 1.8 | 0.01 | 2.2 | 2.5 | 0.02 |
| Gastroesophageal reflux disease | 23.2 | 22.6 | -0.01 | 22.2 | 22.7 | 0.01 | 22.3 | 22.5 | 0.00 |
| Gastrointestinal hemorrhage | 1.6 | 1.3 | -0.03 | 1.6 | 1.3 | -0.03 | 1.7 | 1.4 | -0.03 |
| Human immunodeficiency virus infection | 0.5 | 0.4 | 0.00 | 0.4 | 0.3 | -0.03 | 0.4 | 0.4 | 0.00 |
| Hyperlipidemia | 77.1 | 75.6 | -0.03 | 74.9 | 75.9 | 0.02 | 76.8 | 76.2 | -0.01 |
| Hypertensive disorder | 82.0 | 79.5 | -0.06 | 80.3 | 80.3 | 0.00 | 81.6 | 80.3 | -0.03 |
| Lesion of liver | 2.0 | 1.5 | -0.04 | 2.0 | 1.8 | -0.02 | 2.4 | 1.7 | -0.05 |
| Obesity | 31.7 | 26.6 | -0.11 | 29.1 | 29.7 | 0.01 | 29.1 | 29.7 | 0.01 |
| Osteoarthritis | 22.7 | 23.0 | 0.01 | 22.6 | 23.1 | 0.01 | 22.6 | 23.1 | 0.01 |
| Pneumonia | 2.6 | 1.9 | -0.05 | 2.3 | 1.8 | -0.04 | 2.3 | 1.9 | -0.03 |
| Psoriasis | 1.9 | 1.9 | 0.00 | 1.8 | 1.7 | 0.00 | 1.5 | 1.7 | 0.02 |
| Renal impairment | 12.6 | 11.6 | -0.03 | 10.7 | 10.9 | 0.01 | 11.4 | 11.5 | 0.00 |
| Rheumatoid arthritis | 0.9 | 0.8 | -0.01 | 0.9 | 0.9 | 0.01 | 1.3 | 0.8 | -0.05 |
| Schizophrenia | 0.6 | 0.9 | 0.04 | 0.6 | 0.6 | 0.00 | 1.1 | 0.7 | -0.05 |
| Ulcerative colitis | 0.4 | 0.4 | 0.01 | 0.4 | 0.4 | 0.00 | 0.4 | 0.4 | 0.00 |
| Urinary tract infectious disease | 1.8 | 3.0 | 0.08 | 1.8 | 2.1 | 0.02 | 2.2 | 2.7 | 0.03 |
| Viral hepatitis C | 1.3 | 1.1 | -0.01 | 1.2 | 0.8 | -0.04 | 1.0 | 1.1 | 0.01 |
| Visual system disorder | 47.4 | 48.9 | 0.03 | 47.8 | 47.3 | -0.01 | 48.3 | 47.4 | -0.02 |
| **Medical history: Cardiovascular disease** | | | | | | | | | |
| Atrial fibrillation | 17.1 | 12.2 | -0.14 | 13.4 | 14.5 | 0.03 | 13.9 | 14.6 | 0.02 |
| Cerebrovascular disease | 7.9 | 7.5 | -0.01 | 7.9 | 7.3 | -0.02 | 7.5 | 7.5 | 0.00 |
| Coronary arteriosclerosis | 57.1 | 36.6 | -0.42 | 45.0 | 48.2 | 0.06 | 42.4 | 46.4 | 0.08 |
| Heart disease | 73.9 | 55.1 | -0.40 | 62.1 | 65.3 | 0.07 | 57.9 | 63.0 | 0.10 |
| Heart failure | 20.3 | 8.2 | -0.35 | 9.9 | 11.2 | 0.04 | 12.7 | 11.8 | -0.03 |
| Ischemic heart disease | 25.2 | 16.1 | -0.23 | 17.0 | 17.6 | 0.02 | 17.1 | 18.7 | 0.04 |
| Peripheral vascular disease | 11.4 | 10.9 | -0.01 | 11.1 | 11.1 | 0.00 | 11.4 | 11.2 | -0.01 |
| Pulmonary embolism | 1.0 | 0.9 | -0.01 | 0.8 | 0.9 | 0.01 | 0.8 | 0.9 | 0.01 |
| Venous thrombosis | 1.3 | 1.7 | 0.04 | 1.6 | 1.7 | 0.01 | 2.3 | 1.9 | -0.03 |
| **Medical history: Neoplasms** | | | | | | | | | |

| (Continued) | Before ajustment | | | After matching | | | After stratification | | |
|---|---|---|---|---|---|---|---|---|---|
| Characteristic | T (%) | C (%) | StdDiff | T (%) | C (%) | StdDiff | T (%) | C (%) | StdDiff |
| Hematologic neoplasm | 1.3 | 1.3 | 0.01 | 1.2 | 1.4 | 0.02 | 1.3 | 1.3 | 0.00 |
| Malignant lymphoma | 0.9 | 0.8 | -0.01 | 0.8 | 0.7 | -0.01 | 0.8 | 0.7 | -0.01 |
| Malignant neoplasm of anorectum | <0.1 | 0.1 | 0.00 | 0.1 | <0.3 | -0.01 | <0.1 | <0.1 | -0.01 |
| Malignant neoplastic disease | 10.9 | 11.5 | 0.02 | 11.0 | 10.7 | -0.01 | 11.2 | 11.2 | 0.00 |
| Malignant tumor of breast | <0.1 | 0.1 | 0.02 | <0.1 | <0.3 | 0.03 | 0.6 | 0.1 | -0.08 |
| Malignant tumor of colon | 0.4 | 0.6 | 0.02 | 0.4 | 0.7 | 0.03 | 0.4 | 0.5 | 0.01 |
| Malignant tumor of lung | 0.6 | 0.5 | -0.01 | 0.7 | 0.6 | -0.02 | 0.6 | 0.6 | 0.00 |
| Malignant tumor of urinary bladder | 0.8 | 0.8 | 0.00 | 0.7 | 0.8 | 0.01 | 1.2 | 0.7 | -0.04 |
| Primary malignant neoplasm of prostate | 3.8 | 4.2 | 0.02 | 3.8 | 3.6 | -0.01 | 3.9 | 4.1 | 0.01 |
| **Medication use** | | | | | | | | | |
| Agents acting on the renin-angiotensin system | 73.2 | 66.9 | -0.14 | 68.0 | 68.4 | 0.01 | 67.2 | 68.3 | 0.02 |
| Antibacterials for systemic use | 29.4 | 31.0 | 0.03 | 28.4 | 28.6 | 0.00 | 31.1 | 29.5 | -0.04 |
| Antidepressants | 35.6 | 37.0 | 0.03 | 36.3 | 35.7 | -0.01 | 38.4 | 36.5 | -0.04 |
| Antiepileptics | 28.6 | 30.2 | 0.04 | 29.5 | 28.5 | -0.02 | 29.9 | 29.5 | -0.01 |
| Antiinflammatory and antirheumatic products | 56.5 | 50.9 | -0.11 | 52.7 | 52.9 | 0.00 | 53.0 | 53.1 | 0.00 |
| Antineoplastic agents | 5.0 | 4.9 | 0.00 | 5.1 | 4.8 | -0.01 | 6.3 | 4.9 | -0.06 |
| Antipsoriatics | 1.0 | 1.0 | 0.01 | 1.0 | 1.0 | 0.00 | 1.0 | 1.0 | 0.00 |
| Antithrombotic agents | 61.5 | 50.3 | -0.23 | 54.2 | 55.1 | 0.02 | 54.8 | 55.0 | 0.00 |
| Beta blocking agents | 66.9 | 54.3 | -0.26 | 57.6 | 59.6 | 0.04 | 57.7 | 58.7 | 0.02 |
| Calcium channel blockers | 31.6 | 31.4 | 0.00 | 31.0 | 31.5 | 0.01 | 31.4 | 30.7 | -0.01 |
| Diuretics | 46.7 | 41.0 | -0.12 | 40.5 | 41.3 | 0.02 | 43.7 | 42.3 | -0.03 |
| Drugs for obstructive airway diseases | 45.4 | 44.7 | -0.01 | 46.1 | 45.1 | -0.02 | 46.1 | 45.1 | -0.02 |
| Immunosuppressants | 3.1 | 2.6 | -0.03 | 2.9 | 2.3 | -0.04 | 2.6 | 2.3 | -0.02 |
| Lipid modifying agents | 89.2 | 84.7 | -0.14 | 86.4 | 86.7 | 0.01 | 86.8 | 86.1 | -0.02 |
| Opioids | 17.3 | 19.2 | 0.05 | 16.3 | 16.1 | -0.01 | 17.7 | 17.7 | 0.00 |
| Psycholeptics | 24.8 | 25.3 | 0.01 | 24.0 | 23.7 | -0.01 | 24.6 | 24.4 | 0.00 |
| Psychostimulants | 1.2 | 1.6 | 0.03 | 1.3 | 1.4 | 0.00 | 1.6 | 1.3 | -0.02 |

**Supplementary Table 19:** Baseline patient characteristics for SGLT2is (T) and SUs (C) new-users in the CCAE data source. We report proportion of initiators satisfying selected base-line characteristics and the standardized difference of population proportions (StdDiff) before and after propensity score adjustment. Less extreme StdDiffs through matching and stratification suggest improved balance between patient cohorts through adjustment.

| | Before adjustment | | | After matching | | | After stratification | | |
|---|---|---|---|---|---|---|---|---|---|
| Characteristic | T (%) | C (%) | StdDiff | T (%) | C (%) | StdDiff | T (%) | C (%) | StdDiff |
| **Age group** | | | | | | | | | |
| 25 - 29 | 0.1 | 0.1 | -0.03 | 0.1 | 0.1 | -0.01 | 0.1 | 0.1 | -0.02 |
| 30 - 34 | 0.3 | 0.4 | -0.02 | 0.3 | 0.4 | -0.01 | 0.4 | 0.4 | -0.01 |
| 35 - 39 | 1.7 | 1.6 | 0.01 | 1.8 | 1.4 | 0.03 | 1.8 | 1.5 | 0.02 |
| 40 - 44 | 4.5 | 4.2 | 0.01 | 4.5 | 4.3 | 0.01 | 4.8 | 4.2 | 0.03 |
| 45 - 49 | 10.1 | 9.2 | 0.03 | 10.1 | 10.0 | 0.00 | 10.3 | 9.6 | 0.03 |
| 50 - 54 | 18.3 | 17.7 | 0.01 | 18.4 | 18.1 | 0.01 | 18.4 | 17.8 | 0.01 |
| 55 - 59 | 29.2 | 27.3 | 0.04 | 28.5 | 28.8 | -0.01 | 27.7 | 27.6 | 0.00 |
| 60 - 64 | 33.0 | 35.6 | -0.06 | 33.1 | 33.7 | -0.01 | 33.3 | 35.3 | -0.04 |
| 65 - 69 | 2.8 | 3.8 | -0.06 | 3.1 | 3.1 | 0.00 | 3.1 | 3.5 | -0.02 |
| Gender: female | 38.1 | 39.2 | -0.02 | 39.0 | 38.9 | 0.00 | 39.9 | 38.7 | 0.02 |
| **Medical history: General** | | | | | | | | | |
| Acute respiratory disease | 29.6 | 28.4 | 0.02 | 29.0 | 29.5 | -0.01 | 30.0 | 29.7 | 0.01 |
| Attention deficit hyperactivity disorder | 1.1 | 0.5 | 0.07 | 1.1 | 0.9 | 0.01 | 1.0 | 0.8 | 0.02 |
| Chronic liver disease | 2.5 | 3.9 | -0.08 | 2.8 | 2.3 | 0.03 | 3.3 | 2.9 | 0.02 |
| Chronic obstructive lung disease | 5.2 | 7.0 | -0.07 | 5.3 | 5.5 | 0.00 | 6.3 | 6.4 | -0.01 |
| Crohn’s disease | 0.3 | 0.3 | 0.00 | 0.3 | 0.3 | 0.00 | 0.3 | 0.3 | 0.00 |
| Dementia | 0.3 | 0.4 | -0.02 | 0.3 | 0.3 | -0.01 | 0.3 | 0.4 | -0.01 |
| Depressive disorder | 11.8 | 10.6 | 0.04 | 11.7 | 12.1 | -0.01 | 12.0 | 12.5 | -0.01 |
| Gastroesophageal reflux disease | 17.2 | 12.8 | 0.12 | 17.0 | 17.3 | -0.01 | 17.0 | 16.9 | 0.00 |
| Gastrointestinal hemorrhage | 2.0 | 2.7 | -0.05 | 2.1 | 2.3 | -0.01 | 2.2 | 2.4 | -0.02 |
| Human immunodeficiency virus infection | 0.4 | 0.3 | 0.02 | 0.3 | 0.5 | -0.03 | 0.3 | 0.4 | -0.03 |
| Hyperlipidemia | 82.2 | 68.9 | 0.31 | 80.5 | 81.4 | -0.02 | 78.5 | 79.2 | -0.02 |
| Hypertensive disorder | 81.8 | 73.1 | 0.21 | 81.4 | 81.8 | -0.01 | 81.1 | 81.2 | 0.00 |
| Lesion of liver | 0.8 | 1.2 | -0.04 | 0.8 | 1.0 | -0.02 | 0.9 | 1.2 | -0.04 |
| Obesity | 31.5 | 17.5 | 0.33 | 29.9 | 30.5 | -0.01 | 28.5 | 28.4 | 0.00 |
| Osteoarthritis | 24.5 | 21.8 | 0.06 | 24.2 | 24.7 | -0.01 | 25.3 | 24.6 | 0.02 |
| Pneumonia | 3.2 | 4.0 | -0.04 | 3.1 | 3.0 | 0.01 | 3.6 | 3.6 | 0.00 |
| Psoriasis | 2.2 | 1.5 | 0.05 | 2.1 | 2.0 | 0.01 | 2.0 | 1.9 | 0.00 |
| Renal impairment | 5.3 | 6.3 | -0.04 | 5.4 | 5.3 | 0.00 | 6.7 | 6.9 | -0.01 |
| Rheumatoid arthritis | 1.6 | 1.4 | 0.01 | 1.5 | 1.5 | 0.00 | 1.5 | 1.5 | 0.00 |
| Schizophrenia | 0.1 | 0.1 | -0.01 | 0.1 | 0.1 | -0.02 | 0.1 | 0.1 | -0.01 |
| Ulcerative colitis | 0.4 | 0.4 | 0.00 | 0.4 | 0.4 | -0.01 | 0.4 | 0.5 | -0.01 |
| Urinary tract infectious disease | 6.8 | 8.5 | -0.06 | 6.9 | 6.8 | 0.00 | 7.3 | 7.6 | -0.01 |
| Viral hepatitis C | 0.4 | 0.6 | -0.04 | 0.4 | 0.4 | 0.00 | 0.4 | 0.5 | -0.02 |
| Visual system disorder | 28.1 | 25.6 | 0.06 | 27.6 | 27.9 | -0.01 | 27.3 | 27.7 | -0.01 |
| **Medical history: Cardiovascular disease** | | | | | | | | | |
| Atrial fibrillation | 5.0 | 5.0 | 0.00 | 4.8 | 4.9 | -0.01 | 5.0 | 5.3 | -0.01 |
| Cerebrovascular disease | 9.1 | 11.0 | -0.06 | 8.9 | 9.1 | -0.01 | 9.7 | 9.8 | 0.00 |
| Coronary arteriosclerosis | 34.8 | 33.8 | 0.02 | 31.9 | 32.7 | -0.02 | 31.8 | 32.4 | -0.01 |
| Heart disease | 50.8 | 51.1 | -0.01 | 47.7 | 48.3 | -0.01 | 48.5 | 48.6 | 0.00 |
| Heart failure | 6.7 | 7.0 | -0.01 | 5.7 | 5.8 | 0.00 | 5.8 | 6.5 | -0.03 |
| Ischemic heart disease | 18.2 | 19.6 | -0.04 | 16.6 | 16.8 | -0.01 | 16.9 | 17.3 | -0.01 |
| Peripheral vascular disease | 15.6 | 12.5 | 0.09 | 15.4 | 15.3 | 0.00 | 14.7 | 14.6 | 0.00 |
| Pulmonary embolism | 0.9 | 0.9 | 0.00 | 0.9 | 0.9 | -0.01 | 0.9 | 1.1 | -0.02 |
| Venous thrombosis | 1.5 | 1.8 | -0.02 | 1.6 | 1.4 | 0.01 | 1.9 | 1.8 | 0.01 |
| **Medical history: Neoplasms** | | | | | | | | | |
| Hematologic neoplasm | 1.0 | 1.3 | -0.02 | 1.0 | 1.0 | -0.01 | 1.1 | 1.2 | -0.01 |
| Malignant lymphoma | 0.4 | 0.6 | -0.03 | 0.3 | 0.5 | -0.02 | 0.3 | 0.5 | -0.03 |
| Malignant neoplasm of anorectum | 0.1 | 0.2 | -0.03 | 0.1 | 0.1 | 0.01 | 0.1 | 0.2 | -0.01 |
| Malignant neoplastic disease | 7.2 | 8.1 | -0.03 | 7.1 | 7.1 | 0.00 | 7.1 | 7.1 | 0.00 |
| Malignant tumor of breast | 1.2 | 1.4 | -0.02 | 1.2 | 1.4 | -0.01 | 1.5 | 1.4 | 0.01 |
| Malignant tumor of colon | 0.3 | 0.5 | -0.03 | 0.4 | 0.2 | 0.03 | 0.3 | 0.3 | 0.00 |
| Malignant tumor of lung | 0.3 | 0.4 | -0.02 | 0.3 | 0.2 | 0.03 | 0.4 | 0.3 | 0.01 |
| Malignant tumor of urinary bladder | 0.2 | 0.4 | -0.03 | 0.2 | 0.2 | -0.01 | 0.2 | 0.3 | -0.02 |
| Primary malignant neoplasm of prostate | 1.1 | 1.1 | -0.01 | 1.1 | 1.0 | 0.00 | 1.2 | 1.1 | 0.00 |
| **Medication use** | | | | | | | | | |
| Agents acting on the renin-angiotensin system | 74.5 | 73.2 | 0.03 | 73.8 | 74.0 | 0.00 | 73.5 | 73.9 | -0.01 |
| Antibacterials for systemic use | 61.5 | 62.8 | -0.03 | 61.3 | 61.0 | 0.00 | 62.1 | 61.7 | 0.01 |
| Antidepressants | 30.2 | 30.2 | 0.00 | 29.8 | 30.2 | -0.01 | 30.3 | 30.4 | 0.00 |
| Antiepileptics | 17.2 | 16.5 | 0.02 | 17.4 | 17.2 | 0.01 | 18.8 | 18.0 | 0.02 |
| Antiinflammatory and antirheumatic products | 39.0 | 34.5 | 0.09 | 38.9 | 38.6 | 0.00 | 38.5 | 38.2 | 0.01 |

| (Continued) | Before ajustment | | | After matching | | | After stratification | | |
|---|---|---|---|---|---|---|---|---|---|
| Characteristic | T (%) | C (%) | StdDiff | T (%) | C (%) | StdDiff | T (%) | C (%) | StdDiff |
| Antineoplastic agents | 4.5 | 5.5 | -0.05 | 4.5 | 4.7 | -0.01 | 4.5 | 4.7 | -0.01 |
| Antipsoriatics | 0.8 | 0.7 | 0.01 | 0.7 | 0.6 | 0.01 | 0.6 | 0.6 | 0.00 |
| Antithrombotic agents | 28.4 | 27.7 | 0.02 | 26.9 | 27.1 | 0.00 | 27.5 | 28.1 | -0.01 |
| Beta blocking agents | 47.3 | 49.1 | -0.04 | 45.5 | 45.8 | 0.00 | 46.2 | 47.0 | -0.01 |
| Calcium channel blockers | 27.3 | 27.1 | 0.00 | 27.3 | 27.3 | 0.00 | 27.0 | 27.9 | -0.02 |
| Diuretics | 42.5 | 44.9 | -0.05 | 42.4 | 41.9 | 0.01 | 43.2 | 42.8 | 0.01 |
| Drugs for acid related disorders | 31.2 | 31.3 | 0.00 | 30.5 | 30.7 | 0.00 | 31.2 | 30.5 | 0.01 |
| Drugs for obstructive airway diseases | 42.0 | 38.3 | 0.07 | 41.3 | 41.1 | 0.00 | 41.7 | 41.0 | 0.01 |
| Immunosuppressants | 3.2 | 2.5 | 0.04 | 3.1 | 2.9 | 0.01 | 3.0 | 3.0 | 0.00 |
| Lipid modifying agents | 80.0 | 76.7 | 0.08 | 78.6 | 78.8 | -0.01 | 77.5 | 78.1 | -0.01 |
| Opioids | 31.3 | 33.9 | -0.06 | 31.4 | 31.3 | 0.00 | 34.7 | 33.2 | 0.03 |
| Psycholeptics | 28.4 | 28.9 | -0.01 | 27.8 | 28.1 | 0.00 | 30.2 | 29.0 | 0.03 |
| Psychostimulants | 3.6 | 2.8 | 0.04 | 3.4 | 3.2 | 0.01 | 3.5 | 3.0 | 0.03 |

**Supplementary Table 20:** Baseline patient characteristics for SGLT2is (T) and SUs (C) new-users in the GermanyDA data source. We report proportion of initiators satisfying selected base-line characteristics and the standardized difference of population proportions (StdDiff) before and after propensity score adjustment. Less extreme StdDiffs through matching and stratification suggest improved balance between patient cohorts through adjustment.

| | Before adjustment | | | After matching | | | After stratification | | |
|---|---|---|---|---|---|---|---|---|---|
| Characteristic | T (%) | C (%) | StdDiff | T (%) | C (%) | StdDiff | T (%) | C (%) | StdDiff |
| **Age group** | | | | | | | | | |
| 25 - 29 | <0.1 | <0.2 | -0.01 | <0.7 | <0.2 | 0.05 | <0.1 | <0.2 | 0.02 |
| 30 - 34 | 0.2 | <0.2 | 0.01 | <0.7 | <0.2 | 0.05 | 0.3 | <0.2 | 0.06 |
| 35 - 39 | 0.4 | 0.4 | 0.00 | 0.7 | 0.7 | 0.00 | 0.6 | 0.4 | 0.02 |
| 40 - 44 | 1.0 | 1.1 | -0.01 | 1.2 | 1.1 | 0.01 | 0.9 | 1.0 | -0.01 |
| 45 - 49 | 3.1 | 2.2 | 0.05 | 2.8 | 2.3 | 0.03 | 3.1 | 2.7 | 0.03 |
| 50 - 54 | 7.0 | 4.5 | 0.11 | 5.5 | 5.6 | 0.00 | 7.0 | 4.9 | 0.09 |
| 55 - 59 | 12.2 | 8.1 | 0.14 | 9.8 | 9.0 | 0.03 | 11.7 | 14.6 | -0.09 |
| 60 - 64 | 17.1 | 13.6 | 0.10 | 14.2 | 13.0 | 0.04 | 16.5 | 13.7 | 0.08 |
| 65 - 69 | 16.5 | 16.9 | -0.01 | 16.6 | 16.5 | 0.00 | 16.6 | 19.0 | -0.06 |
| 70 - 74 | 15.4 | 19.2 | -0.10 | 13.4 | 15.0 | -0.04 | 14.7 | 14.8 | 0.00 |
| 75 - 79 | 13.3 | 18.4 | -0.14 | 16.6 | 16.7 | 0.00 | 14.3 | 15.2 | -0.03 |
| 80 - 84 | 9.6 | 10.5 | -0.03 | 11.9 | 12.0 | 0.00 | 9.9 | 9.1 | 0.03 |
| 85 - 89 | 3.5 | 3.9 | -0.02 | 5.3 | 5.8 | -0.03 | 3.7 | 3.3 | 0.02 |
| 90 - 94 | 0.6 | 0.9 | -0.04 | 1.4 | 2.1 | -0.05 | 0.6 | 1.1 | -0.06 |
| Gender: female | 30.7 | 43.5 | -0.27 | 41.1 | 39.6 | 0.03 | 33.4 | 30.5 | 0.06 |
| **Medical history: General** | | | | | | | | | |
| Acute respiratory disease | 16.2 | 17.7 | -0.04 | 18.5 | 15.3 | 0.09 | 16.7 | 16.4 | 0.01 |
| Attention deficit hyperactivity disorder | <0.1 | 0.7 | -0.10 | <0.1 | 0.7 | -0.11 | <0.1 | 0.7 | -0.11 |
| Chronic liver disease | 0.7 | 0.3 | 0.05 | 1.1 | <0.2 | 0.12 | 0.6 | <0.2 | 0.09 |
| Chronic obstructive lung disease | 8.0 | 8.7 | -0.03 | 9.5 | 8.6 | 0.03 | 8.7 | 7.7 | 0.04 |
| Crohn's disease | 0.2 | 0.2 | 0.00 | 0.1 | 0.3 | -0.04 | 0.1 | 0.3 | -0.04 |
| Dementia | 1.4 | 3.9 | -0.16 | 2.6 | 3.1 | -0.03 | 1.6 | 1.8 | -0.02 |
| Depressive disorder | 9.2 | 11.6 | -0.08 | 12.7 | 12.2 | 0.01 | 10.2 | 15.6 | -0.16 |
| Gastroesophageal reflux disease | 2.5 | 2.5 | 0.00 | 3.6 | 2.5 | 0.07 | 2.8 | 2.5 | 0.02 |
| Gastrointestinal hemorrhage | 1.0 | 1.3 | -0.03 | 1.0 | 0.7 | 0.03 | 0.9 | 0.4 | 0.06 |
| Hyperlipidemia | 28.8 | 34.4 | -0.12 | 29.0 | 28.1 | 0.02 | 28.5 | 25.8 | 0.06 |
| Hypertensive disorder | 47.8 | 64.4 | -0.34 | 52.6 | 51.8 | 0.01 | 49.3 | 48.2 | 0.02 |
| Lesion of liver | 0.7 | 0.8 | -0.01 | 0.7 | 0.7 | 0.00 | 0.6 | 1.2 | -0.06 |
| Obesity | 10.2 | 8.4 | 0.06 | 9.7 | 9.3 | 0.01 | 10.6 | 7.6 | 0.10 |
| Osteoarthritis | 11.9 | 16.9 | -0.14 | 15.8 | 13.7 | 0.06 | 12.9 | 12.2 | 0.02 |
| Pneumonia | 2.1 | 2.6 | -0.04 | 2.2 | 2.1 | 0.01 | 2.0 | 1.7 | 0.02 |
| Psoriasis | 1.7 | 1.6 | 0.00 | 1.9 | 1.3 | 0.05 | 1.5 | 0.6 | 0.09 |
| Renal impairment | 7.1 | 7.1 | 0.00 | 6.9 | 6.7 | 0.01 | 6.7 | 5.1 | 0.07 |
| Rheumatoid arthritis | 1.5 | 1.7 | -0.02 | 1.5 | 1.4 | 0.01 | 1.5 | 0.8 | 0.06 |
| Schizophrenia | 0.2 | 0.2 | -0.01 | <0.7 | 0.2 | 0.04 | 0.2 | 0.2 | -0.01 |
| Ulcerative colitis | 0.2 | 0.2 | 0.00 | 0.1 | <0.2 | 0.02 | 0.1 | <0.2 | 0.02 |
| Urinary tract infectious disease | 4.5 | 6.1 | -0.07 | 5.1 | 5.4 | -0.01 | 4.7 | 3.2 | 0.07 |
| Visual system disorder | 8.1 | 11.7 | -0.12 | 10.2 | 10.7 | -0.01 | 8.4 | 11.5 | -0.10 |
| **Medical history: Cardiovascular disease** | | | | | | | | | |
| Atrial fibrillation | 4.7 | 3.4 | 0.06 | 4.7 | 4.0 | 0.03 | 4.3 | 2.4 | 0.11 |
| Cerebrovascular disease | 8.0 | 10.7 | -0.09 | 10.2 | 9.3 | 0.03 | 8.4 | 9.5 | -0.04 |
| Coronary arteriosclerosis | 14.6 | 7.7 | 0.22 | 8.9 | 9.2 | -0.01 | 13.3 | 13.9 | -0.02 |
| Heart disease | 46.9 | 49.3 | -0.05 | 40.5 | 37.2 | 0.07 | 44.5 | 45.1 | -0.01 |
| Heart failure | 13.4 | 13.2 | 0.00 | 10.7 | 9.1 | 0.05 | 12.8 | 9.3 | 0.11 |
| Ischemic heart disease | 26.5 | 32.4 | -0.13 | 21.7 | 20.0 | 0.04 | 24.9 | 25.8 | -0.02 |
| Peripheral vascular disease | 12.0 | 12.4 | -0.01 | 11.1 | 12.8 | -0.05 | 11.3 | 10.2 | 0.04 |
| Pulmonary embolism | 0.7 | 0.5 | 0.03 | <0.7 | 0.5 | -0.02 | 0.6 | 0.2 | 0.06 |
| Venous thrombosis | 1.8 | 1.6 | 0.02 | 1.8 | 1.7 | 0.01 | 1.9 | 2.3 | -0.03 |
| **Medical history: Neoplasms** | | | | | | | | | |
| Malignant lymphoma | 0.2 | 0.3 | -0.02 | <0.7 | <0.2 | 0.04 | 0.2 | <0.2 | 0.06 |
| Malignant neoplasm of anorectum | <0.1 | 0.2 | -0.03 | <0.1 | <0.2 | 0.01 | <0.1 | <0.2 | 0.01 |
| Malignant neoplastic disease | 4.7 | 7.4 | -0.11 | 4.4 | 5.4 | -0.05 | 4.4 | 3.7 | 0.04 |
| Malignant tumor of breast | 0.6 | 1.3 | -0.07 | 0.7 | 0.8 | -0.01 | 0.7 | 0.8 | -0.01 |
| Malignant tumor of colon | 0.4 | 0.4 | -0.01 | <0.7 | 0.3 | 0.00 | 0.4 | 0.2 | 0.03 |
| Malignant tumor of lung | <0.1 | 0.2 | -0.04 | <0.7 | 0.4 | -0.05 | <0.1 | <0.2 | -0.01 |
| Malignant tumor of urinary bladder | 0.2 | 0.3 | -0.02 | <0.7 | <0.2 | 0.05 | 0.2 | <0.2 | 0.06 |
| Primary malignant neoplasm of prostate | 0.8 | 1.4 | -0.06 | <0.7 | 1.2 | -0.09 | 0.7 | 1.0 | -0.03 |
| **Medication use** | | | | | | | | | |
| Agents acting on the renin-angiotensin system | 79.3 | 73.7 | 0.13 | 74.1 | 73.9 | 0.00 | 77.7 | 78.2 | -0.01 |
| Antibacterials for systemic use | 22.8 | 31.2 | -0.19 | 27.7 | 25.7 | 0.04 | 24.0 | 23.3 | 0.02 |
| Antidepressants | 11.1 | 11.4 | -0.01 | 12.9 | 10.5 | 0.07 | 11.6 | 12.3 | -0.02 |

| (Continued) | Before ajustment | | | After matching | | | After stratification | | |
|---|---|---|---|---|---|---|---|---|---|
| Characteristic | T (%) | C (%) | StdDiff | T (%) | C (%) | StdDiff | T (%) | C (%) | StdDiff |
| Antiepileptics | 5.5 | 4.4 | 0.05 | 4.6 | 4.0 | 0.03 | 5.2 | 4.1 | 0.05 |
| Antiinflammatory and antirheumatic products | 53.5 | 59.0 | -0.11 | 51.7 | 51.8 | 0.00 | 53.5 | 53.2 | 0.00 |
| Antineoplastic agents | 1.4 | 1.4 | 0.00 | 1.1 | 1.7 | -0.05 | 1.3 | 1.2 | 0.01 |
| Antipsoriatics | 0.5 | 0.3 | 0.03 | <0.7 | <0.2 | 0.07 | 0.6 | <0.2 | 0.09 |
| Antithrombotic agents | 56.3 | 50.9 | 0.11 | 47.6 | 48.8 | -0.02 | 53.5 | 54.1 | -0.01 |
| Beta blocking agents | 62.9 | 59.0 | 0.08 | 61.1 | 58.9 | 0.05 | 61.7 | 60.2 | 0.03 |
| Calcium channel blockers | 39.6 | 36.9 | 0.05 | 42.5 | 39.8 | 0.05 | 39.5 | 43.3 | -0.08 |
| Diuretics | 58.3 | 60.7 | -0.05 | 56.2 | 54.6 | 0.03 | 57.6 | 59.2 | -0.03 |
| Drugs for acid related disorders | 42.9 | 38.8 | 0.08 | 42.3 | 43.2 | -0.02 | 43.2 | 48.9 | -0.11 |
| Drugs for obstructive airway diseases | 19.8 | 19.2 | 0.02 | 18.5 | 18.4 | 0.00 | 19.6 | 17.5 | 0.05 |
| Immunosuppressants | 1.0 | 0.8 | 0.02 | 0.9 | 0.4 | 0.06 | 0.9 | 0.4 | 0.06 |
| Lipid modifying agents | 70.0 | 52.1 | 0.37 | 56.8 | 57.5 | -0.01 | 65.5 | 70.3 | -0.10 |
| Opioids | 12.5 | 15.8 | -0.09 | 15.1 | 14.1 | 0.03 | 13.4 | 16.9 | -0.10 |
| Psycholeptics | 9.3 | 13.4 | -0.13 | 10.7 | 11.0 | -0.01 | 9.8 | 9.8 | 0.00 |
| Psychostimulants | 0.1 | 0.7 | -0.09 | <0.7 | <0.2 | 0.05 | 0.3 | <0.2 | 0.06 |

**Supplementary Table 21:** Baseline patient characteristics for SGLT2is (T) and SUs (C) new-users in the OptumDOD data source. We report proportion of initiators satisfying selected base-line characteristics and the standardized difference of population proportions (StdDiff) before and after propensity score adjustment. Less extreme StdDiffs through matching and stratification suggest improved balance between patient cohorts through adjustment.

| | Before adjustment | | | After matching | | | After stratification | | |
|---|---|---|---|---|---|---|---|---|---|
| Characteristic | T (%) | C (%) | StdDiff | T (%) | C (%) | StdDiff | T (%) | C (%) | StdDiff |
| **Age group** | | | | | | | | | |
| 30 - 34 | 0.2 | 0.2 | 0.00 | 0.3 | 0.2 | 0.02 | 0.2 | 0.2 | 0.01 |
| 35 - 39 | 0.7 | 0.6 | 0.02 | 0.8 | 0.6 | 0.02 | 1.0 | 0.5 | 0.06 |
| 40 - 44 | 2.1 | 1.3 | 0.06 | 1.9 | 1.8 | 0.01 | 1.7 | 1.3 | 0.03 |
| 45 - 49 | 4.6 | 3.0 | 0.08 | 3.1 | 3.1 | 0.00 | 3.1 | 3.1 | 0.00 |
| 50 - 54 | 7.7 | 5.6 | 0.09 | 7.4 | 7.5 | 0.00 | 5.6 | 5.5 | 0.01 |
| 55 - 59 | 12.7 | 8.9 | 0.12 | 11.9 | 12.5 | -0.02 | 10.3 | 9.3 | 0.03 |
| 60 - 64 | 15.2 | 11.7 | 0.10 | 14.5 | 14.9 | -0.01 | 12.3 | 12.1 | 0.01 |
| 65 - 69 | 19.0 | 17.8 | 0.03 | 19.4 | 19.1 | 0.01 | 19.0 | 18.7 | 0.01 |
| 70 - 74 | 19.5 | 20.9 | -0.04 | 20.1 | 20.7 | -0.01 | 20.7 | 21.9 | -0.03 |
| 75 - 79 | 11.5 | 15.6 | -0.12 | 12.0 | 11.6 | 0.01 | 13.5 | 14.7 | -0.03 |
| 80 - 84 | 4.6 | 9.5 | -0.19 | 5.0 | 4.7 | 0.02 | 8.0 | 8.2 | -0.01 |
| 85 - 89 | 1.8 | 4.6 | -0.16 | 2.0 | 1.9 | 0.01 | 4.2 | 4.3 | 0.00 |
| 90 - 94 | 0.2 | 0.3 | -0.01 | 0.2 | 0.3 | -0.03 | 0.2 | 0.3 | -0.03 |
| Gender: female | 38.7 | 44.2 | -0.11 | 40.0 | 39.8 | 0.01 | 44.6 | 43.1 | 0.03 |
| **Medical history: General** | | | | | | | | | |
| Acute respiratory disease | 27.8 | 28.1 | -0.01 | 27.4 | 27.4 | 0.00 | 29.6 | 27.7 | 0.04 |
| Attention deficit hyperactivity disorder | 0.7 | 0.4 | 0.04 | 0.7 | 0.7 | 0.00 | 0.7 | 0.5 | 0.03 |
| Chronic liver disease | 2.2 | 3.7 | -0.09 | 2.3 | 2.3 | 0.00 | 3.8 | 2.9 | 0.05 |
| Chronic obstructive lung disease | 14.6 | 17.2 | -0.07 | 14.5 | 14.5 | 0.00 | 17.5 | 17.0 | 0.01 |
| Crohn's disease | 0.4 | 0.3 | 0.01 | 0.4 | 0.3 | 0.02 | 0.3 | 0.3 | 0.01 |
| Dementia | 1.7 | 4.4 | -0.16 | 1.9 | 1.9 | 0.00 | 4.4 | 4.0 | 0.02 |
| Depressive disorder | 17.9 | 17.1 | 0.02 | 17.9 | 17.9 | 0.00 | 21.1 | 18.6 | 0.06 |
| Gastroesophageal reflux disease | 25.1 | 22.2 | 0.07 | 24.4 | 24.3 | 0.00 | 25.7 | 24.1 | 0.04 |
| Gastrointestinal hemorrhage | 2.9 | 4.0 | -0.06 | 2.8 | 2.8 | 0.00 | 3.7 | 3.5 | 0.01 |
| Human immunodeficiency virus infection | 0.3 | 0.2 | 0.02 | 0.3 | 0.4 | 0.00 | 0.2 | 0.3 | -0.01 |
| Hyperlipidemia | 87.8 | 85.0 | 0.08 | 86.9 | 87.6 | -0.02 | 84.9 | 86.0 | -0.03 |
| Hypertensive disorder | 88.4 | 87.4 | 0.03 | 87.9 | 87.6 | 0.01 | 87.6 | 87.7 | 0.00 |
| Lesion of liver | 1.6 | 1.9 | -0.03 | 2.5 | 1.9 | 0.04 | 2.5 | 1.9 | 0.04 |
| Obesity | 33.7 | 23.4 | 0.23 | 31.8 | 32.6 | -0.02 | 29.3 | 28.3 | 0.02 |
| Osteoarthritis | 33.9 | 34.4 | -0.01 | 33.9 | 33.7 | 0.00 | 37.7 | 35.3 | 0.05 |
| Pneumonia | 5.0 | 7.2 | -0.09 | 5.0 | 4.9 | 0.00 | 6.8 | 6.6 | 0.01 |
| Psoriasis | 2.2 | 1.6 | 0.04 | 2.2 | 2.0 | 0.02 | 2.0 | 1.8 | 0.02 |
| Renal impairment | 17.4 | 24.9 | -0.18 | 17.8 | 16.7 | 0.03 | 24.5 | 24.4 | 0.00 |
| Rheumatoid arthritis | 2.2 | 2.5 | -0.02 | 2.5 | 2.5 | 0.00 | 2.5 | 2.5 | 0.00 |
| Schizophrenia | 0.4 | 0.5 | -0.02 | 0.4 | 0.4 | 0.00 | 0.4 | 0.5 | -0.01 |
| Ulcerative colitis | 0.4 | 0.4 | 0.01 | 0.5 | 0.4 | 0.01 | 0.6 | 0.4 | 0.03 |
| Urinary tract infectious disease | 9.1 | 14.7 | -0.18 | 9.4 | 9.3 | 0.00 | 13.8 | 13.1 | 0.02 |
| Viral hepatitis C | 0.6 | 0.8 | -0.02 | 0.6 | 0.7 | -0.02 | 0.8 | 0.8 | -0.01 |
| Visual system disorder | 45.2 | 47.2 | -0.04 | 45.9 | 45.4 | 0.01 | 48.4 | 47.9 | 0.01 |
| **Medical history: Cardiovascular disease** | | | | | | | | | |
| Atrial fibrillation | 12.4 | 13.1 | -0.02 | 11.6 | 11.3 | 0.01 | 12.6 | 13.0 | -0.01 |
| Cerebrovascular disease | 13.3 | 16.3 | -0.08 | 13.3 | 13.0 | 0.01 | 15.2 | 14.8 | 0.01 |
| Coronary arteriosclerosis | 44.7 | 40.0 | 0.10 | 41.8 | 42.3 | -0.01 | 38.6 | 40.3 | -0.04 |
| Heart disease | 62.3 | 60.6 | 0.04 | 59.0 | 59.9 | -0.02 | 59.0 | 59.9 | -0.02 |
| Heart failure | 16.4 | 16.0 | 0.01 | 14.4 | 14.3 | 0.00 | 15.5 | 15.8 | -0.01 |
| Ischemic heart disease | 24.3 | 22.2 | 0.05 | 22.0 | 22.7 | -0.02 | 21.3 | 21.9 | -0.02 |
| Peripheral vascular disease | 27.0 | 26.1 | 0.02 | 27.1 | 27.3 | 0.00 | 28.2 | 27.9 | 0.01 |
| Pulmonary embolism | 1.3 | 1.4 | -0.01 | 1.4 | 1.2 | 0.01 | 1.6 | 1.5 | 0.01 |
| Venous thrombosis | 2.3 | 2.8 | -0.03 | 2.3 | 2.2 | 0.00 | 2.8 | 2.7 | 0.01 |
| **Medical history: Neoplasms** | | | | | | | | | |
| Hematologic neoplasm | 1.3 | 2.0 | -0.05 | 1.4 | 1.5 | -0.01 | 1.6 | 1.9 | -0.02 |
| Malignant lymphoma | 0.7 | 0.9 | -0.02 | 0.7 | 0.8 | -0.01 | 0.8 | 0.9 | -0.01 |
| Malignant neoplasm of anorectum | 0.2 | 0.4 | -0.02 | 0.2 | 0.3 | -0.01 | 0.4 | 0.3 | 0.01 |
| Malignant neoplastic disease | 12.3 | 15.1 | -0.08 | 12.5 | 12.3 | 0.01 | 13.5 | 14.6 | -0.03 |
| Malignant tumor of breast | 1.5 | 2.1 | -0.05 | 1.6 | 1.7 | -0.01 | 1.9 | 2.0 | -0.01 |
| Malignant tumor of colon | 0.6 | 0.9 | -0.04 | 0.6 | 0.6 | 0.00 | 1.0 | 0.9 | 0.01 |
| Malignant tumor of lung | 0.5 | 0.9 | -0.05 | 0.5 | 0.5 | -0.01 | 0.6 | 0.9 | -0.03 |
| Malignant tumor of urinary bladder | 0.5 | 0.9 | -0.06 | 0.5 | 0.7 | -0.02 | 0.5 | 0.9 | -0.04 |
| Primary malignant neoplasm of prostate | 2.8 | 3.1 | -0.02 | 2.9 | 2.5 | 0.02 | 2.8 | 3.0 | -0.01 |
| **Medication use** | | | | | | | | | |
| Agents acting on the renin-angiotensin system | 76.9 | 75.8 | 0.03 | 76.4 | 75.8 | 0.01 | 76.1 | 76.2 | 0.00 |

| (Continued) | Before ajustment | | | After matching | | | After stratification | | |
|---|---|---|---|---|---|---|---|---|---|
| Characteristic | T (%) | C (%) | StdDiff | T (%) | C (%) | StdDiff | T (%) | C (%) | StdDiff |
| Antibacterials for systemic use | 60.2 | 62.4 | -0.04 | 60.2 | 59.8 | 0.01 | 63.2 | 61.4 | 0.04 |
| Antidepressants | 31.8 | 30.3 | 0.03 | 31.7 | 31.6 | 0.00 | 33.6 | 31.6 | 0.04 |
| Antiepileptics | 22.5 | 21.9 | 0.01 | 22.8 | 23.1 | -0.01 | 25.5 | 24.1 | 0.03 |
| Antiinflammatory and antirheumatic products | 33.6 | 30.4 | 0.07 | 33.2 | 33.8 | -0.01 | 34.4 | 31.9 | 0.05 |
| Antineoplastic agents | 6.4 | 6.6 | -0.01 | 6.2 | 6.2 | 0.00 | 6.6 | 6.5 | 0.00 |
| Antipsoriatics | 0.5 | 0.7 | -0.01 | 0.6 | 0.6 | -0.01 | 0.4 | 0.7 | -0.03 |
| Antithrombotic agents | 35.2 | 32.2 | 0.06 | 33.4 | 33.7 | -0.01 | 33.2 | 33.0 | 0.00 |
| Beta blocking agents | 55.3 | 54.6 | 0.02 | 53.5 | 53.4 | 0.00 | 54.1 | 54.3 | 0.00 |
| Calcium channel blockers | 32.0 | 34.2 | -0.05 | 32.3 | 32.0 | 0.01 | 32.3 | 32.0 | 0.01 |
| Diuretics | 48.3 | 50.8 | -0.05 | 47.7 | 46.9 | 0.01 | 49.8 | 49.3 | 0.01 |
| Drugs for acid related disorders | 34.7 | 35.0 | -0.01 | 34.4 | 34.2 | 0.00 | 37.4 | 35.9 | 0.03 |
| Drugs for obstructive airway diseases | 43.5 | 41.5 | 0.04 | 42.8 | 43.3 | -0.01 | 45.1 | 42.7 | 0.05 |
| Immunosuppressants | 3.3 | 2.8 | 0.03 | 3.3 | 3.0 | 0.02 | 3.2 | 3.0 | 0.01 |
| Lipid modifying agents | 85.7 | 81.1 | 0.12 | 84.8 | 85.1 | -0.01 | 82.1 | 83.1 | -0.03 |
| Opioids | 32.8 | 33.5 | -0.01 | 32.1 | 32.7 | -0.01 | 35.6 | 33.6 | 0.04 |
| Psycholeptics | 29.8 | 28.8 | 0.02 | 28.8 | 29.1 | 0.00 | 31.9 | 29.5 | 0.05 |
| Psychostimulants | 2.1 | 1.4 | 0.05 | 2.0 | 2.0 | 0.00 | 2.1 | 1.6 | 0.04 |

**Supplementary Table 22:** Baseline patient characteristics for SGLT2is (T) and SUs (C) new-users in the MDCR data source. We report proportion of initiators satisfying selected base-line characteristics and the standardized difference of population proportions (StdDiff) before and after propensity score adjustment. Less extreme StdDiffs through matching and stratification suggest improved balance between patient cohorts through adjustment.

| | Before adjustment | | | After matching | | | After stratification | | |
|---|---|---|---|---|---|---|---|---|---|
| Characteristic | T (%) | C (%) | StdDiff | T (%) | C (%) | StdDiff | T (%) | C (%) | StdDiff |
| **Age group** | | | | | | | | | |
| 50 - 54 | <0.2 | 0.1 | 0.01 | <0.3 | 0.2 | 0.00 | <0.2 | 0.1 | -0.01 |
| 55 - 59 | 1.0 | 0.5 | 0.06 | 1.0 | 0.6 | 0.05 | 0.9 | 0.5 | 0.05 |
| 60 - 64 | 2.1 | 1.3 | 0.06 | 2.1 | 1.6 | 0.04 | 2.1 | 1.6 | 0.04 |
| 65 - 69 | 27.8 | 19.8 | 0.19 | 26.9 | 27.2 | -0.01 | 20.4 | 22.3 | -0.05 |
| 70 - 74 | 36.9 | 28.4 | 0.18 | 36.4 | 35.4 | 0.02 | 31.3 | 29.9 | 0.03 |
| 75 - 79 | 21.3 | 23.3 | -0.05 | 21.3 | 22.1 | -0.02 | 21.3 | 22.1 | -0.02 |
| 80 - 84 | 7.5 | 16.4 | -0.28 | 8.3 | 8.0 | 0.01 | 13.3 | 13.8 | -0.01 |
| 85 - 89 | 2.6 | 7.6 | -0.23 | 3.1 | 3.4 | -0.02 | 8.6 | 6.9 | 0.06 |
| 90 - 94 | 0.5 | 2.2 | -0.14 | 0.6 | 1.3 | -0.08 | 0.7 | 2.2 | -0.13 |
| 95 - 99 | <0.2 | 0.3 | -0.05 | <0.3 | 0.3 | -0.03 | <0.2 | 0.3 | -0.05 |
| Gender: female | 36.1 | 42.6 | -0.13 | 37.3 | 38.0 | -0.01 | 41.8 | 40.9 | 0.02 |
| **Medical history: General** | | | | | | | | | |
| Acute respiratory disease | 26.0 | 24.4 | 0.04 | 25.7 | 26.0 | -0.01 | 30.5 | 26.4 | 0.09 |
| Attention deficit hyperactivity disorder | <0.2 | 0.1 | 0.01 | <0.3 | 0.4 | -0.05 | 0.2 | 0.3 | -0.02 |
| Chronic liver disease | 2.3 | 2.3 | 0.00 | 2.6 | 1.7 | 0.06 | 2.3 | 2.3 | 0.00 |
| Chronic obstructive lung disease | 14.2 | 15.2 | -0.03 | 14.4 | 14.7 | -0.01 | 18.3 | 16.2 | 0.06 |
| Crohn’s disease | 0.5 | 0.3 | 0.04 | 0.5 | 0.3 | 0.02 | 0.4 | 0.3 | 0.00 |
| Dementia | 1.9 | 4.5 | -0.15 | 2.1 | 2.5 | -0.03 | 1.9 | 5.3 | -0.19 |
| Depressive disorder | 9.9 | 8.2 | 0.06 | 9.7 | 11.2 | -0.05 | 17.4 | 12.2 | 0.15 |
| Gastroesophageal reflux disease | 19.0 | 11.8 | 0.20 | 18.2 | 19.1 | -0.02 | 18.4 | 18.4 | 0.00 |
| Gastrointestinal hemorrhage | 3.8 | 4.6 | -0.04 | 3.7 | 2.9 | 0.04 | 2.9 | 3.8 | -0.05 |
| Human immunodeficiency virus infection | <0.2 | 0.1 | 0.02 | <0.3 | 0.1 | -0.01 | <0.2 | 0.1 | -0.03 |
| Hyperlipidemia | 84.8 | 52.6 | 0.74 | 83.4 | 85.2 | -0.05 | 82.0 | 79.3 | 0.07 |
| Hypertensive disorder | 87.7 | 71.3 | 0.42 | 87.0 | 87.8 | -0.03 | 87.3 | 86.6 | 0.02 |
| Lesion of liver | 1.7 | 1.3 | 0.04 | 1.7 | 1.3 | 0.04 | 1.8 | 1.4 | 0.03 |
| Obesity | 25.9 | 8.2 | 0.48 | 24.0 | 24.6 | -0.01 | 15.7 | 18.3 | -0.07 |
| Osteoarthritis | 36.1 | 30.1 | 0.13 | 36.0 | 35.7 | 0.00 | 39.0 | 36.2 | 0.06 |
| Pneumonia | 5.0 | 8.3 | -0.13 | 4.9 | 5.1 | -0.01 | 7.5 | 7.3 | 0.01 |
| Psoriasis | 2.5 | 1.4 | 0.08 | 2.6 | 2.2 | 0.03 | 1.7 | 2.0 | -0.02 |
| Renal impairment | 14.5 | 16.4 | -0.05 | 15.3 | 14.7 | 0.02 | 17.1 | 21.6 | -0.11 |
| Rheumatoid arthritis | 1.7 | 1.8 | -0.01 | 1.6 | 1.8 | -0.01 | 1.4 | 2.0 | -0.04 |
| Ulcerative colitis | 0.5 | 0.4 | 0.01 | 0.5 | 0.4 | 0.01 | 0.4 | 0.4 | -0.01 |
| Urinary tract infectious disease | 8.1 | 12.3 | -0.14 | 8.5 | 9.0 | -0.02 | 14.5 | 12.6 | 0.05 |
| Viral hepatitis C | 0.6 | 0.3 | 0.06 | 0.7 | 0.3 | 0.05 | 0.3 | 0.3 | 0.00 |
| Visual system disorder | 57.8 | 50.1 | 0.16 | 55.6 | 56.8 | -0.02 | 54.7 | 54.7 | 0.00 |
| **Medical history: Cardiovascular disease** | | | | | | | | | |
| Atrial fibrillation | 16.7 | 16.4 | 0.01 | 16.4 | 16.2 | 0.00 | 17.3 | 18.0 | -0.02 |
| Cerebrovascular disease | 18.9 | 22.0 | -0.08 | 17.4 | 17.5 | 0.00 | 21.3 | 19.5 | 0.05 |
| Coronary arteriosclerosis | 51.9 | 44.3 | 0.15 | 48.5 | 50.7 | -0.04 | 40.4 | 45.5 | -0.10 |
| Heart disease | 71.2 | 66.9 | 0.09 | 65.2 | 66.6 | -0.03 | 65.2 | 66.6 | -0.03 |
| Heart failure | 15.3 | 16.6 | -0.04 | 13.6 | 12.5 | 0.03 | 17.1 | 15.6 | 0.04 |
| Peripheral vascular disease | 24.9 | 20.0 | 0.12 | 23.5 | 23.7 | -0.01 | 22.1 | 23.2 | -0.03 |
| Pulmonary embolism | 1.0 | 1.3 | -0.02 | 1.0 | 1.4 | -0.04 | 1.7 | 1.6 | 0.01 |
| Venous thrombosis | 2.2 | 3.0 | -0.05 | 2.4 | 2.5 | -0.01 | 1.8 | 3.2 | -0.09 |
| **Medical history: Neoplasms** | | | | | | | | | |
| Hematologic neoplasm | 1.7 | 2.1 | -0.03 | 1.7 | 1.9 | -0.01 | 1.0 | 2.4 | -0.11 |
| Malignant lymphoma | 1.2 | 1.0 | 0.02 | 1.1 | 1.2 | -0.01 | 0.6 | 1.3 | -0.07 |
| Malignant neoplasm of anorectum | 0.3 | 0.5 | -0.02 | 0.4 | 0.3 | 0.01 | 0.4 | 0.4 | 0.00 |
| Malignant neoplastic disease | 19.2 | 20.4 | -0.03 | 18.9 | 19.7 | -0.02 | 17.4 | 21.2 | -0.10 |
| Malignant tumor of breast | 2.2 | 2.6 | -0.03 | 2.3 | 2.3 | 0.00 | 2.2 | 2.5 | -0.02 |
| Malignant tumor of colon | 0.7 | 1.1 | -0.04 | 0.7 | 0.6 | 0.02 | 0.8 | 0.9 | -0.02 |
| Malignant tumor of lung | 0.4 | 1.1 | -0.07 | 0.5 | 0.8 | -0.04 | 0.3 | 1.2 | -0.10 |
| Malignant tumor of urinary bladder | 1.2 | 1.4 | -0.02 | 1.3 | 1.1 | 0.02 | 1.8 | 1.4 | 0.03 |
| Primary malignant neoplasm of prostate | 5.4 | 5.3 | 0.01 | 5.4 | 5.0 | 0.02 | 5.2 | 5.3 | -0.01 |
| **Medication use** | | | | | | | | | |
| Agents acting on the renin-angiotensin system | 77.6 | 74.3 | 0.08 | 77.5 | 75.7 | 0.04 | 77.0 | 75.6 | 0.03 |
| Antibacterials for systemic use | 64.8 | 65.0 | 0.00 | 64.4 | 63.9 | 0.01 | 65.9 | 65.0 | 0.02 |
| Antidepressants | 27.0 | 27.3 | -0.01 | 27.5 | 26.9 | 0.01 | 28.0 | 27.8 | 0.00 |
| Antiepileptics | 19.7 | 17.3 | 0.06 | 18.8 | 19.4 | -0.01 | 19.1 | 20.0 | -0.02 |
| Antiinflammatory and antirheumatic products | 29.9 | 27.6 | 0.05 | 30.0 | 29.2 | 0.02 | 27.7 | 28.2 | -0.01 |
| Antineoplastic agents | 8.4 | 9.6 | -0.04 | 8.4 | 8.3 | 0.00 | 8.8 | 8.4 | 0.01 |

| (Continued) | Before ajustment | | | After matching | | | After stratification | | |
|---|---|---|---|---|---|---|---|---|---|
| Characteristic | T (%) | C (%) | StdDiff | T (%) | C (%) | StdDiff | T (%) | C (%) | StdDiff |
| Antipsoriatics | 0.9 | 0.8 | 0.02 | 1.0 | 0.8 | 0.02 | 0.4 | 0.9 | -0.06 |
| Beta blocking agents | 61.7 | 61.0 | 0.02 | 60.7 | 61.1 | -0.01 | 59.6 | 61.1 | -0.03 |
| Calcium channel blockers | 35.9 | 37.1 | -0.02 | 35.8 | 35.4 | 0.01 | 34.4 | 37.2 | -0.06 |
| Diuretics | 53.1 | 56.7 | -0.07 | 52.2 | 51.0 | 0.02 | 54.9 | 52.8 | 0.04 |
| Drugs for acid related disorders | 39.1 | 36.7 | 0.05 | 37.0 | 37.4 | -0.01 | 37.4 | 36.6 | 0.02 |
| Drugs for obstructive airway diseases | 48.5 | 41.9 | 0.13 | 47.3 | 48.3 | -0.02 | 47.0 | 45.9 | 0.02 |
| Immunosuppressants | 3.9 | 2.9 | 0.05 | 3.7 | 3.9 | -0.01 | 3.0 | 3.7 | -0.04 |
| Lipid modifying agents | 89.2 | 80.2 | 0.25 | 88.0 | 88.4 | -0.01 | 87.4 | 84.2 | 0.09 |
| Opioids | 28.9 | 32.4 | -0.08 | 28.7 | 29.5 | -0.02 | 32.5 | 32.3 | 0.00 |
| Psycholeptics | 26.0 | 29.0 | -0.07 | 26.3 | 26.0 | 0.01 | 31.4 | 28.4 | 0.07 |
| Psychostimulants | 1.6 | 1.3 | 0.03 | 1.8 | 1.4 | 0.03 | 1.0 | 1.3 | -0.02 |

**Supplementary Table 23:** Baseline patient characteristics for SGLT2is (T) and SUs (C) new-users in the MDCD data source. We report proportion of initiators satisfying selected base-line characteristics and the standardized difference of population proportions (StdDiff) before and after propensity score adjustment. Less extreme StdDiffs through matching and stratification suggest improved balance between patient cohorts through adjustment.

| | Before adjustment | | | After matching | | | After stratification | | |
|---|---|---|---|---|---|---|---|---|---|
| Characteristic | T (%) | C (%) | StdDiff | T (%) | C (%) | StdDiff | T (%) | C (%) | StdDiff |
| **Age group** | | | | | | | | | |
| 15 - 19 | <0.4 | <0.1 | 0.01 | <0.4 | 0.1 | 0.01 | <0.3 | <0.1 | 0.01 |
| 20 - 24 | <0.4 | 0.3 | -0.05 | <0.4 | 0.3 | -0.05 | <0.3 | 0.3 | -0.06 |
| 25 - 29 | 1.4 | 0.7 | 0.07 | 1.5 | 0.8 | 0.07 | 2.9 | 0.7 | 0.16 |
| 30 - 34 | 2.4 | 1.9 | 0.04 | 2.5 | 1.9 | 0.04 | 3.0 | 1.8 | 0.08 |
| 35 - 39 | 5.9 | 4.1 | 0.08 | 6.1 | 4.6 | 0.06 | 5.6 | 4.5 | 0.05 |
| 40 - 44 | 8.4 | 6.8 | 0.06 | 8.3 | 8.3 | 0.00 | 7.3 | 7.1 | 0.01 |
| 45 - 49 | 14.5 | 11.9 | 0.08 | 14.0 | 13.0 | 0.03 | 12.6 | 12.0 | 0.02 |
| 50 - 54 | 17.9 | 17.0 | 0.02 | 18.0 | 17.3 | 0.02 | 19.2 | 17.1 | 0.05 |
| 55 - 59 | 22.9 | 21.4 | 0.04 | 22.8 | 22.8 | 0.00 | 22.7 | 22.6 | 0.00 |
| 60 - 64 | 18.0 | 20.2 | -0.06 | 18.2 | 19.2 | -0.03 | 19.0 | 20.8 | -0.04 |
| 65 - 69 | 4.3 | 6.8 | -0.11 | 4.4 | 5.8 | -0.06 | 4.2 | 6.1 | -0.09 |
| 70 - 74 | 2.7 | 3.9 | -0.07 | 2.6 | 3.2 | -0.04 | 1.9 | 3.3 | -0.09 |
| 75 - 79 | 0.5 | 2.4 | -0.15 | 0.5 | 1.3 | -0.08 | 0.3 | 1.8 | -0.15 |
| 80 - 84 | 0.7 | 1.6 | -0.09 | 0.7 | 1.0 | -0.03 | 0.6 | 1.1 | -0.06 |
| 85 - 89 | <0.4 | 0.9 | -0.09 | <0.4 | 0.5 | -0.05 | 0.7 | 0.7 | 0.00 |
| Gender: female | 56.4 | 58.4 | -0.04 | 56.4 | 55.0 | 0.03 | 57.4 | 56.2 | 0.03 |
| **Race** | | | | | | | | | |
| Black or African American | 22.7 | 25.5 | -0.07 | 22.8 | 20.6 | 0.05 | 25.6 | 25.4 | 0.01 |
| White | 63.6 | 58.0 | 0.11 | 63.4 | 64.3 | -0.02 | 62.3 | 57.9 | 0.09 |
| **Ethnicity** | | | | | | | | | |
| Hispanic or Latino | 1.9 | 2.1 | -0.01 | 1.8 | 2.4 | -0.04 | 1.3 | 2.3 | -0.07 |
| **Medical history: General** | | | | | | | | | |
| Acute respiratory disease | 38.7 | 40.7 | -0.04 | 38.5 | 39.8 | -0.03 | 45.6 | 40.2 | 0.11 |
| Attention deficit hyperactivity disorder | 1.9 | 1.2 | 0.06 | 1.8 | 1.8 | 0.00 | 1.9 | 1.5 | 0.03 |
| Chronic liver disease | 3.8 | 7.0 | -0.14 | 3.9 | 4.2 | -0.02 | 8.5 | 6.0 | 0.10 |
| Chronic obstructive lung disease | 29.8 | 31.2 | -0.03 | 29.9 | 29.9 | 0.00 | 33.6 | 31.2 | 0.05 |
| Crohn’s disease | 0.5 | 0.4 | 0.01 | 0.5 | 0.5 | 0.00 | 0.3 | 0.5 | -0.03 |
| Dementia | 1.6 | 3.2 | -0.11 | 1.6 | 2.1 | -0.04 | 1.5 | 3.1 | -0.11 |
| Depressive disorder | 37.3 | 36.0 | 0.03 | 36.7 | 39.0 | -0.05 | 37.6 | 38.4 | -0.02 |
| Gastroesophageal reflux disease | 31.7 | 30.7 | 0.02 | 31.5 | 33.8 | -0.05 | 35.5 | 32.8 | 0.06 |
| Gastrointestinal hemorrhage | 4.1 | 5.2 | -0.05 | 4.1 | 4.0 | 0.00 | 4.0 | 4.7 | -0.04 |
| Human immunodeficiency virus infection | 0.6 | 0.6 | 0.00 | 0.6 | 0.6 | 0.01 | 2.4 | 0.6 | 0.15 |
| Hyperlipidemia | 74.7 | 72.4 | 0.05 | 74.6 | 75.6 | -0.02 | 71.1 | 73.8 | -0.06 |
| Hypertensive disorder | 84.9 | 86.4 | -0.04 | 87.6 | 87.1 | 0.01 | 87.6 | 87.1 | 0.01 |
| Lesion of liver | 1.7 | 2.5 | -0.06 | 1.7 | 2.4 | -0.05 | 1.6 | 2.5 | -0.06 |
| Obesity | 43.6 | 35.4 | 0.17 | 43.3 | 43.0 | 0.01 | 44.8 | 40.1 | 0.10 |
| Osteoarthritis | 45.7 | 42.7 | 0.06 | 45.2 | 46.1 | -0.02 | 45.6 | 43.8 | 0.04 |
| Pneumonia | 7.6 | 10.4 | -0.10 | 7.6 | 8.2 | -0.02 | 10.1 | 9.6 | 0.02 |
| Psoriasis | 1.8 | 1.3 | 0.04 | 1.7 | 1.9 | -0.02 | 1.3 | 1.6 | -0.03 |
| Renal impairment | 12.3 | 15.7 | -0.10 | 12.4 | 11.7 | 0.02 | 14.2 | 16.2 | -0.06 |
| Rheumatoid arthritis | 2.8 | 2.5 | 0.02 | 2.7 | 2.1 | 0.04 | 4.1 | 2.3 | 0.10 |
| Schizophrenia | 3.7 | 5.3 | -0.08 | 3.5 | 4.4 | -0.05 | 6.0 | 5.2 | 0.03 |
| Ulcerative colitis | <0.4 | 0.3 | 0.00 | <0.4 | 0.3 | 0.00 | 0.8 | 0.3 | 0.07 |
| Urinary tract infectious disease | 10.6 | 15.4 | -0.14 | 10.6 | 10.8 | -0.01 | 13.2 | 13.9 | -0.02 |
| Viral hepatitis C | 2.4 | 3.5 | -0.07 | 2.4 | 2.6 | -0.01 | 2.4 | 2.6 | -0.01 |
| Visual system disorder | 48.6 | 43.6 | 0.10 | 48.3 | 47.8 | 0.01 | 43.4 | 44.8 | -0.03 |
| **Medical history: Cardiovascular disease** | | | | | | | | | |
| Atrial fibrillation | 6.5 | 7.6 | -0.04 | 6.5 | 6.6 | 0.00 | 6.7 | 7.1 | -0.01 |
| Cerebrovascular disease | 11.4 | 14.6 | -0.10 | 11.2 | 11.0 | 0.01 | 12.0 | 12.8 | -0.03 |
| Coronary arteriosclerosis | 33.6 | 33.1 | 0.01 | 33.3 | 32.9 | 0.01 | 33.6 | 32.1 | 0.03 |
| Heart disease | 55.0 | 57.8 | -0.06 | 54.6 | 54.3 | 0.00 | 55.5 | 55.8 | -0.01 |
| Heart failure | 18.1 | 19.3 | -0.03 | 17.8 | 17.2 | 0.02 | 19.3 | 18.5 | 0.02 |
| Ischemic heart disease | 22.5 | 23.8 | -0.03 | 22.3 | 23.1 | -0.02 | 22.3 | 23.1 | -0.02 |
| Peripheral vascular disease | 22.4 | 21.4 | 0.02 | 22.4 | 21.9 | 0.01 | 19.9 | 21.9 | -0.05 |
| Pulmonary embolism | 1.8 | 2.0 | -0.01 | 1.7 | 1.9 | -0.01 | 3.0 | 2.1 | 0.06 |
| Venous thrombosis | 2.4 | 3.2 | -0.05 | 2.5 | 2.9 | -0.02 | 2.2 | 3.1 | -0.06 |
| **Medical history: Neoplasms** | | | | | | | | | |
| Hematologic neoplasm | 0.8 | 1.4 | -0.06 | 0.8 | 1.2 | -0.04 | 0.7 | 1.3 | -0.06 |
| Malignant lymphoma | <0.4 | 0.5 | -0.03 | <0.4 | 0.4 | -0.01 | 0.4 | 0.5 | -0.01 |
| Malignant neoplasm of anorectum | <0.4 | 0.4 | -0.06 | <0.4 | 0.3 | -0.05 | <0.3 | 0.3 | -0.06 |
| Malignant neoplastic disease | 6.1 | 8.4 | -0.09 | 6.0 | 7.1 | -0.05 | 5.6 | 8.1 | -0.10 |

| (Continued) | Before ajustment | | | After matching | | | After stratification | | |
|---|---|---|---|---|---|---|---|---|---|
| Characteristic | T (%) | C (%) | StdDiff | T (%) | C (%) | StdDiff | T (%) | C (%) | StdDiff |
| Malignant tumor of breast | 1.6 | 1.7 | -0.01 | 1.6 | 1.3 | 0.03 | 1.5 | 1.5 | -0.01 |
| Malignant tumor of colon | <0.4 | 0.6 | -0.06 | <0.4 | 0.4 | -0.03 | <0.3 | 0.5 | -0.07 |
| Malignant tumor of lung | <0.4 | 0.7 | -0.08 | <0.4 | 0.6 | -0.07 | <0.3 | 0.7 | -0.10 |
| Malignant tumor of urinary bladder | <0.4 | 0.3 | -0.04 | <0.4 | 0.4 | -0.04 | <0.3 | 0.4 | -0.05 |
| Primary malignant neoplasm of prostate | <0.4 | 0.7 | -0.06 | <0.4 | 0.5 | -0.04 | <0.3 | 0.7 | -0.07 |
| **Medication use** | | | | | | | | | |
| Agents acting on the renin-angiotensin system | 68.7 | 71.0 | -0.05 | 68.8 | 67.5 | 0.03 | 67.8 | 69.8 | -0.04 |
| Antibacterials for systemic use | 69.6 | 70.6 | -0.02 | 72.0 | 69.7 | 0.05 | 72.0 | 69.7 | 0.05 |
| Antidepressants | 55.8 | 55.0 | 0.02 | 55.7 | 57.0 | -0.03 | 58.3 | 55.4 | 0.06 |
| Antiepileptics | 50.9 | 45.8 | 0.10 | 50.4 | 49.2 | 0.02 | 48.0 | 48.4 | -0.01 |
| Antiinflammatory and antirheumatic products | 66.3 | 58.8 | 0.15 | 66.1 | 65.9 | 0.00 | 65.3 | 62.6 | 0.06 |
| Antineoplastic agents | 5.4 | 4.9 | 0.02 | 5.3 | 4.2 | 0.05 | 4.2 | 4.5 | -0.01 |
| Antipsoriatics | 0.4 | 0.6 | -0.03 | <0.4 | 0.6 | -0.04 | <0.3 | 0.6 | -0.07 |
| Antithrombotic agents | 45.1 | 43.2 | 0.04 | 44.7 | 44.8 | 0.00 | 44.5 | 44.9 | -0.01 |
| Beta blocking agents | 52.8 | 52.6 | 0.00 | 52.5 | 51.4 | 0.02 | 52.1 | 52.5 | -0.01 |
| Calcium channel blockers | 28.2 | 33.7 | -0.12 | 28.6 | 27.4 | 0.03 | 36.3 | 32.7 | 0.08 |
| Diuretics | 53.4 | 56.8 | -0.07 | 53.2 | 51.8 | 0.03 | 56.0 | 55.0 | 0.02 |
| Drugs for acid related disorders | 55.8 | 54.8 | 0.02 | 55.4 | 55.8 | -0.01 | 59.4 | 55.2 | 0.09 |
| Drugs for obstructive airway diseases | 64.8 | 60.9 | 0.08 | 64.5 | 63.3 | 0.03 | 65.0 | 62.2 | 0.06 |
| Immunosuppressants | 3.0 | 2.6 | 0.02 | 3.0 | 2.7 | 0.01 | 3.8 | 2.7 | 0.06 |
| Lipid modifying agents | 77.6 | 74.9 | 0.06 | 77.3 | 77.5 | 0.00 | 71.8 | 76.0 | -0.10 |
| Opioids | 51.0 | 54.6 | -0.07 | 50.7 | 48.8 | 0.04 | 54.2 | 52.7 | 0.03 |
| Psycholeptics | 53.8 | 53.0 | 0.02 | 56.8 | 53.0 | 0.08 | 56.8 | 53.0 | 0.08 |
| Psychostimulants | 5.6 | 4.0 | 0.07 | 5.5 | 4.5 | 0.05 | 5.4 | 4.2 | 0.06 |

**Supplementary Table 24:** Baseline patient characteristics for SGLT2is (T) and SUs (C) new-users in the Open Claims data source. We report proportion of initiators satisfying selected base-line characteristics and the standardized difference of population proportions (StdDiff) before and after propensity score adjustment. Less extreme StdDiffs through matching and stratification suggest improved balance between patient cohorts through adjustment.

| | Before adjustment | | | After matching | | | After stratification | | |
|---|---|---|---|---|---|---|---|---|---|
| Characteristic | T (%) | C (%) | StdDiff | T (%) | C (%) | StdDiff | T (%) | C (%) | StdDiff |
| **Age group** | | | | | | | | | |
| 20 - 24 | 0.1 | 0.1 | 0.00 | 0.1 | 0.1 | 0.00 | 0.2 | 0.1 | 0.01 |
| 25 - 29 | 0.2 | 0.2 | 0.00 | 0.2 | 0.2 | 0.00 | 0.2 | 0.2 | 0.00 |
| 30 - 34 | 0.4 | 0.4 | 0.00 | 0.4 | 0.4 | 0.00 | 0.4 | 0.4 | 0.00 |
| 35 - 39 | 1.0 | 0.9 | 0.02 | 1.1 | 1.1 | 0.00 | 1.0 | 0.9 | 0.01 |
| 40 - 44 | 2.4 | 1.9 | 0.04 | 2.4 | 2.6 | -0.01 | 2.4 | 2.6 | -0.01 |
| 45 - 49 | 5.2 | 4.0 | 0.06 | 5.2 | 5.3 | 0.00 | 4.6 | 4.4 | 0.01 |
| 50 - 54 | 9.4 | 7.4 | 0.07 | 9.3 | 9.4 | 0.00 | 8.2 | 7.9 | 0.01 |
| 55 - 59 | 14.5 | 11.4 | 0.09 | 14.1 | 14.5 | -0.01 | 12.5 | 12.1 | 0.01 |
| 60 - 64 | 18.0 | 14.2 | 0.10 | 17.6 | 17.8 | 0.00 | 15.9 | 15.3 | 0.02 |
| 65 - 69 | 17.5 | 17.3 | 0.00 | 17.5 | 18.1 | -0.01 | 17.3 | 17.6 | -0.01 |
| 70 - 74 | 14.4 | 16.5 | -0.06 | 14.7 | 14.4 | 0.01 | 14.7 | 14.4 | 0.01 |
| 75 - 79 | 10.0 | 18.0 | -0.23 | 10.5 | 9.5 | 0.03 | 14.5 | 15.6 | -0.03 |
| 80 - 84 | 6.1 | 7.4 | -0.05 | 6.4 | 6.3 | 0.01 | 7.0 | 7.1 | 0.00 |
| 85 - 89 | 0.5 | 0.2 | 0.06 | 0.4 | 0.5 | 0.00 | 0.3 | 0.3 | 0.00 |
| Gender: female | 39.9 | 44.7 | -0.10 | 40.9 | 41.9 | -0.02 | 43.7 | 44.0 | -0.01 |
| **Medical history: General** | | | | | | | | | |
| Acute respiratory disease | 15.6 | 16.9 | -0.04 | 15.3 | 16.1 | -0.02 | 16.9 | 16.7 | 0.01 |
| Attention deficit hyperactivity disorder | 0.5 | 0.3 | 0.03 | 0.4 | 0.4 | 0.00 | 0.4 | 0.3 | 0.01 |
| Chronic liver disease | 1.3 | 1.8 | -0.05 | 1.3 | 1.3 | 0.00 | 1.9 | 1.7 | 0.01 |
| Chronic obstructive lung disease | 8.8 | 11.1 | -0.08 | 8.7 | 8.8 | 0.00 | 10.9 | 10.5 | 0.01 |
| Crohn's disease | 0.2 | 0.2 | 0.00 | 0.2 | 0.2 | 0.00 | 0.2 | 0.2 | 0.00 |
| Dementia | 0.9 | 1.9 | -0.09 | 1.0 | 0.9 | 0.00 | 1.7 | 1.7 | 0.00 |
| Depressive disorder | 8.0 | 8.8 | -0.03 | 8.0 | 8.4 | -0.01 | 9.0 | 8.7 | 0.01 |
| Gastroesophageal reflux disease | 10.6 | 10.4 | 0.01 | 10.4 | 10.6 | -0.01 | 11.1 | 10.6 | 0.02 |
| Gastrointestinal hemorrhage | 1.8 | 2.3 | -0.04 | 1.8 | 1.8 | 0.00 | 1.8 | 1.8 | 0.00 |
| Human immunodeficiency virus infection | 0.3 | 0.2 | 0.01 | 0.3 | 0.3 | 0.00 | 0.3 | 0.3 | 0.00 |
| Hyperlipidemia | 53.2 | 50.0 | 0.06 | 51.4 | 50.9 | 0.01 | 51.4 | 50.9 | 0.01 |
| Hypertensive disorder | 60.7 | 61.3 | -0.01 | 59.9 | 59.9 | 0.00 | 61.6 | 61.2 | 0.01 |
| Lesion of liver | 0.8 | 1.1 | -0.02 | 0.9 | 0.9 | -0.01 | 1.1 | 1.0 | 0.01 |
| Obesity | 13.9 | 11.2 | 0.08 | 13.4 | 13.6 | -0.01 | 12.6 | 12.0 | 0.02 |
| Osteoarthritis | 19.0 | 20.4 | -0.04 | 18.9 | 19.3 | -0.01 | 20.7 | 20.3 | 0.01 |
| Pneumonia | 3.4 | 4.6 | -0.06 | 3.2 | 3.4 | -0.01 | 3.2 | 3.4 | -0.01 |
| Psoriasis | 1.2 | 0.9 | 0.03 | 1.1 | 1.1 | 0.00 | 1.1 | 1.1 | 0.00 |
| Renal impairment | 9.5 | 13.2 | -0.12 | 9.2 | 8.8 | 0.01 | 11.6 | 12.2 | -0.02 |
| Rheumatoid arthritis | 1.3 | 1.4 | -0.01 | 1.3 | 1.3 | 0.00 | 1.5 | 1.4 | 0.01 |
| Schizophrenia | 0.3 | 0.5 | -0.02 | 0.3 | 0.4 | 0.00 | 0.4 | 0.4 | 0.00 |
| Ulcerative colitis | 0.3 | 0.2 | 0.00 | 0.2 | 0.2 | 0.00 | 0.3 | 0.2 | 0.00 |
| Urinary tract infectious disease | 5.2 | 7.7 | -0.10 | 5.4 | 5.4 | 0.00 | 7.7 | 7.2 | 0.02 |
| Viral hepatitis C | 0.4 | 0.5 | -0.03 | 0.4 | 0.4 | 0.00 | 0.5 | 0.5 | 0.00 |
| Visual system disorder | 21.8 | 23.4 | -0.04 | 21.8 | 21.5 | 0.01 | 23.1 | 23.0 | 0.00 |
| **Medical history: Cardiovascular disease** | | | | | | | | | |
| Atrial fibrillation | 8.4 | 8.9 | -0.02 | 7.8 | 7.5 | 0.01 | 8.5 | 8.6 | -0.01 |
| Cerebrovascular disease | 7.5 | 9.6 | -0.08 | 7.4 | 7.3 | 0.00 | 9.1 | 9.0 | 0.00 |
| Coronary arteriosclerosis | 28.2 | 25.4 | 0.06 | 26.2 | 25.1 | 0.03 | 25.9 | 25.6 | 0.01 |
| Heart disease | 44.2 | 42.3 | 0.04 | 41.7 | 40.4 | 0.03 | 42.5 | 42.1 | 0.01 |
| Heart failure | 11.3 | 10.2 | 0.04 | 9.4 | 8.7 | 0.03 | 10.1 | 9.9 | 0.01 |
| Ischemic heart disease | 13.9 | 12.6 | 0.04 | 12.8 | 12.2 | 0.02 | 12.8 | 12.6 | 0.00 |
| Peripheral vascular disease | 13.3 | 13.9 | -0.02 | 13.2 | 13.3 | 0.00 | 13.9 | 13.8 | 0.00 |
| Pulmonary embolism | 0.8 | 0.9 | -0.01 | 0.8 | 0.8 | 0.00 | 1.0 | 0.9 | 0.01 |
| Venous thrombosis | 1.3 | 1.6 | -0.03 | 1.2 | 1.3 | -0.01 | 1.2 | 1.3 | -0.01 |
| **Medical history: Neoplasms** | | | | | | | | | |
| Hematologic neoplasm | 0.8 | 1.2 | -0.04 | 0.8 | 0.9 | 0.00 | 1.1 | 1.1 | 0.00 |
| Malignant lymphoma | 0.5 | 0.6 | -0.02 | 0.5 | 0.5 | 0.00 | 0.7 | 0.6 | 0.01 |
| Malignant neoplasm of anorectum | 0.1 | 0.2 | -0.01 | 0.2 | 0.1 | 0.01 | 0.2 | 0.2 | 0.01 |
| Malignant neoplastic disease | 7.5 | 9.4 | -0.07 | 7.6 | 7.4 | 0.00 | 9.1 | 8.9 | 0.01 |
| Malignant tumor of breast | 1.1 | 1.4 | -0.02 | 1.2 | 1.2 | 0.00 | 1.3 | 1.3 | 0.00 |
| Malignant tumor of colon | 0.3 | 0.5 | -0.03 | 0.3 | 0.3 | 0.00 | 0.5 | 0.5 | 0.00 |
| Malignant tumor of lung | 0.3 | 0.6 | -0.04 | 0.3 | 0.4 | -0.01 | 0.5 | 0.5 | -0.01 |
| Malignant tumor of urinary bladder | 0.3 | 0.5 | -0.03 | 0.4 | 0.3 | 0.00 | 0.4 | 0.3 | 0.00 |
| Primary malignant neoplasm of prostate | 1.6 | 2.0 | -0.03 | 1.6 | 1.5 | 0.01 | 1.8 | 1.8 | 0.00 |
| **Medication use** | | | | | | | | | |

| (Continued) | Before ajustment | | | After matching | | | After stratification | | |
|---|---|---|---|---|---|---|---|---|---|
| Characteristic | T (%) | C (%) | StdDiff | T (%) | C (%) | StdDiff | T (%) | C (%) | StdDiff |
| Agents acting on the renin-angiotensin system | 74.6 | 73.2 | 0.03 | 73.5 | 73.0 | 0.01 | 73.3 | 73.2 | 0.00 |
| Antibacterials for systemic use | 58.9 | 60.5 | -0.03 | 58.8 | 59.6 | -0.02 | 61.4 | 60.8 | 0.01 |
| Antidepressants | 32.1 | 32.4 | -0.01 | 31.9 | 32.6 | -0.01 | 33.3 | 32.6 | 0.01 |
| Antiepileptics | 23.4 | 25.2 | -0.04 | 23.6 | 24.0 | -0.01 | 26.0 | 25.2 | 0.02 |
| Antiinflammatory and antirheumatic products | 39.5 | 37.6 | 0.04 | 39.2 | 39.4 | 0.00 | 39.4 | 38.5 | 0.02 |
| Antineoplastic agents | 5.9 | 5.4 | 0.02 | 5.6 | 5.9 | -0.01 | 5.7 | 5.6 | 0.00 |
| Antipsoriatics | 0.8 | 0.8 | 0.01 | 0.8 | 0.8 | 0.00 | 0.9 | 0.8 | 0.00 |
| Antithrombotic agents | 38.2 | 36.6 | 0.03 | 36.4 | 35.5 | 0.02 | 36.8 | 36.5 | 0.01 |
| Beta blocking agents | 55.8 | 54.5 | 0.03 | 53.9 | 52.8 | 0.02 | 53.8 | 54.3 | -0.01 |
| Calcium channel blockers | 31.9 | 33.6 | -0.04 | 31.9 | 31.8 | 0.00 | 32.4 | 33.3 | -0.02 |
| Diuretics | 47.8 | 48.6 | -0.02 | 46.6 | 46.0 | 0.01 | 48.4 | 47.9 | 0.01 |
| Drugs for acid related disorders | 40.6 | 41.1 | -0.01 | 39.9 | 40.0 | 0.00 | 42.0 | 41.0 | 0.02 |
| Drugs for obstructive airway diseases | 42.7 | 41.0 | 0.03 | 41.9 | 42.5 | -0.01 | 42.4 | 41.6 | 0.02 |
| Immunosuppressants | 3.4 | 2.9 | 0.03 | 3.2 | 3.2 | 0.00 | 3.2 | 3.0 | 0.01 |
| Lipid modifying agents | 82.5 | 77.9 | 0.12 | 81.3 | 80.7 | 0.01 | 78.8 | 78.8 | 0.00 |
| Opioids | 28.2 | 31.3 | -0.07 | 28.2 | 28.7 | -0.01 | 31.6 | 30.9 | 0.01 |
| Psycholeptics | 26.2 | 27.8 | -0.04 | 25.9 | 26.4 | -0.01 | 28.5 | 27.7 | 0.02 |
| Psychostimulants | 2.7 | 2.2 | 0.03 | 2.6 | 2.6 | 0.00 | 2.5 | 2.3 | 0.01 |

**Supplementary Table 25:** Baseline patient characteristics for SGLT2is (T) and SUs (C) new-users in the OptumEHR data source. We report proportion of initiators satisfying selected base-line characteristics and the standardized difference of population proportions (StdDiff) before and after propensity score adjustment. Less extreme StdDiffs through matching and stratification suggest improved balance between patient cohorts through adjustment.

| | Before adjustment | | | After matching | | | After stratification | | |
|---|---|---|---|---|---|---|---|---|---|
| Characteristic | T (%) | C (%) | StdDiff | T (%) | C (%) | StdDiff | T (%) | C (%) | StdDiff |
| **Age group** | | | | | | | | | |
| 20 - 24 | <0.1 | 0.1 | -0.01 | <0.1 | 0.1 | -0.02 | <0.1 | 0.1 | -0.02 |
| 30 - 34 | 0.4 | 0.3 | 0.01 | 0.4 | 0.4 | 0.01 | 0.4 | 0.3 | 0.01 |
| 35 - 39 | 1.2 | 0.9 | 0.03 | 1.2 | 1.0 | 0.01 | 1.4 | 0.9 | 0.04 |
| 40 - 44 | 3.3 | 2.0 | 0.08 | 3.1 | 3.1 | 0.00 | 2.8 | 2.3 | 0.03 |
| 45 - 49 | 6.6 | 4.6 | 0.08 | 6.3 | 6.4 | 0.00 | 5.0 | 5.1 | 0.00 |
| 50 - 54 | 10.8 | 8.2 | 0.09 | 10.6 | 10.8 | -0.01 | 8.5 | 8.8 | -0.01 |
| 55 - 59 | 17.1 | 12.6 | 0.13 | 16.6 | 16.8 | -0.01 | 13.6 | 13.7 | 0.00 |
| 60 - 64 | 20.8 | 15.6 | 0.14 | 20.2 | 20.7 | -0.01 | 17.1 | 16.9 | 0.00 |
| 65 - 69 | 16.2 | 16.3 | 0.00 | 16.4 | 16.7 | -0.01 | 17.0 | 16.3 | 0.02 |
| 70 - 74 | 12.5 | 15.4 | -0.08 | 13.0 | 12.6 | 0.01 | 13.9 | 14.7 | -0.02 |
| 75 - 79 | 6.8 | 13.2 | -0.22 | 7.4 | 6.9 | 0.02 | 9.6 | 10.6 | -0.03 |
| 80 - 84 | 3.1 | 8.7 | -0.24 | 3.4 | 3.1 | 0.02 | 8.6 | 8.0 | 0.02 |
| 85 - 89 | 1.0 | 2.0 | -0.08 | 1.2 | 1.2 | 0.00 | 1.8 | 2.2 | -0.03 |
| Gender: female | 35.4 | 39.3 | -0.08 | 36.4 | 36.2 | 0.00 | 39.9 | 38.8 | 0.02 |
| **Race** | | | | | | | | | |
| Asian | 1.9 | 1.7 | 0.01 | 1.8 | 1.9 | -0.01 | 1.7 | 1.8 | -0.01 |
| Black or African American | 8.1 | 8.0 | 0.00 | 8.2 | 8.3 | 0.00 | 7.8 | 8.3 | -0.02 |
| White | 83.7 | 84.5 | -0.02 | 83.8 | 83.5 | 0.01 | 84.6 | 83.9 | 0.02 |
| **Ethnicity** | | | | | | | | | |
| Hispanic or Latino | 4.5 | 4.9 | -0.02 | 4.4 | 4.6 | -0.01 | 4.3 | 4.7 | -0.02 |
| Not Hispanic or Latino | 86.3 | 88.4 | -0.06 | 87.6 | 87.3 | 0.01 | 89.2 | 88.2 | 0.03 |
| **Medical history: General** | | | | | | | | | |
| Acute respiratory disease | 19.3 | 18.1 | 0.03 | 19.1 | 19.2 | 0.00 | 17.7 | 18.7 | -0.03 |
| Attention deficit hyperactivity disorder | 0.6 | 0.4 | 0.03 | 0.6 | 0.6 | 0.00 | 0.5 | 0.5 | 0.00 |
| Chronic liver disease | 2.2 | 3.1 | -0.06 | 2.3 | 2.3 | 0.00 | 3.1 | 2.9 | 0.01 |
| Chronic obstructive lung disease | 9.9 | 12.1 | -0.07 | 10.2 | 10.1 | 0.01 | 12.3 | 11.9 | 0.01 |
| Crohn’s disease | 0.3 | 0.2 | 0.00 | 0.2 | 0.3 | 0.00 | 0.3 | 0.3 | 0.01 |
| Dementia | 0.8 | 2.2 | -0.12 | 0.8 | 1.0 | -0.01 | 1.4 | 1.9 | -0.04 |
| Depressive disorder | 14.7 | 15.1 | -0.01 | 14.9 | 14.7 | 0.00 | 17.0 | 15.3 | 0.05 |
| Gastroesophageal reflux disease | 21.8 | 19.8 | 0.05 | 21.5 | 21.4 | 0.00 | 22.0 | 20.9 | 0.03 |
| Gastrointestinal hemorrhage | 1.6 | 2.2 | -0.05 | 1.6 | 1.5 | 0.01 | 1.8 | 2.0 | -0.02 |
| Human immunodeficiency virus infection | 0.2 | 0.2 | 0.00 | 0.2 | 0.2 | 0.00 | 0.1 | 0.2 | -0.02 |
| Hyperlipidemia | 78.1 | 73.9 | 0.10 | 76.5 | 77.0 | -0.01 | 74.9 | 74.4 | 0.01 |
| Hypertensive disorder | 78.9 | 76.8 | 0.05 | 77.9 | 78.7 | -0.02 | 78.2 | 77.3 | 0.02 |
| Lesion of liver | 0.9 | 1.2 | -0.03 | 1.0 | 1.1 | -0.01 | 2.0 | 1.2 | 0.06 |
| Obesity | 33.2 | 23.7 | 0.21 | 31.6 | 32.8 | -0.03 | 29.4 | 27.3 | 0.05 |
| Osteoarthritis | 20.9 | 22.2 | -0.03 | 20.9 | 20.8 | 0.00 | 23.6 | 22.5 | 0.03 |
| Pneumonia | 2.4 | 3.8 | -0.08 | 2.4 | 2.6 | -0.01 | 3.3 | 3.6 | -0.01 |
| Psoriasis | 1.7 | 1.5 | 0.02 | 1.7 | 1.6 | 0.01 | 1.7 | 1.5 | 0.01 |
| Renal impairment | 8.7 | 13.9 | -0.16 | 9.0 | 8.5 | 0.02 | 14.8 | 13.4 | 0.04 |
| Rheumatoid arthritis | 1.6 | 1.5 | 0.01 | 1.6 | 1.5 | 0.01 | 1.9 | 1.5 | 0.03 |
| Schizophrenia | 0.2 | 0.3 | -0.03 | 0.2 | 0.3 | -0.02 | 0.2 | 0.3 | -0.03 |
| Ulcerative colitis | 0.3 | 0.3 | 0.01 | 0.4 | 0.3 | 0.01 | 0.3 | 0.3 | 0.00 |
| Urinary tract infectious disease | 3.9 | 6.7 | -0.13 | 4.1 | 4.2 | 0.00 | 7.7 | 6.0 | 0.07 |
| Viral hepatitis C | 0.4 | 0.7 | -0.04 | 0.5 | 0.6 | -0.01 | 0.4 | 0.7 | -0.04 |
| Visual system disorder | 13.0 | 18.0 | -0.14 | 13.2 | 13.1 | 0.01 | 18.9 | 16.8 | 0.05 |
| **Medical history: Cardiovascular disease** | | | | | | | | | |
| Atrial fibrillation | 8.8 | 10.7 | -0.06 | 8.7 | 8.5 | 0.01 | 10.6 | 10.3 | 0.01 |
| Cerebrovascular disease | 9.1 | 11.5 | -0.08 | 9.4 | 9.0 | 0.01 | 11.1 | 10.6 | 0.02 |
| Coronary arteriosclerosis | 47.4 | 42.8 | 0.09 | 44.9 | 45.4 | -0.01 | 41.4 | 42.2 | -0.02 |
| Heart disease | 61.8 | 59.1 | 0.06 | 59.6 | 59.8 | 0.00 | 59.2 | 58.4 | 0.02 |
| Heart failure | 9.9 | 10.3 | -0.01 | 9.1 | 9.2 | 0.00 | 10.8 | 10.1 | 0.02 |
| Ischemic heart disease | 22.0 | 19.6 | 0.06 | 20.8 | 21.0 | -0.01 | 19.6 | 20.0 | -0.01 |
| Peripheral vascular disease | 20.3 | 17.9 | 0.06 | 19.7 | 19.4 | 0.01 | 19.6 | 18.7 | 0.02 |
| Pulmonary embolism | 0.9 | 1.1 | -0.02 | 0.9 | 1.0 | -0.01 | 1.2 | 1.1 | 0.00 |
| Venous thrombosis | 1.8 | 1.9 | -0.01 | 1.7 | 1.5 | 0.02 | 2.3 | 1.8 | 0.03 |
| **Medical history: Neoplasms** | | | | | | | | | |
| Hematologic neoplasm | 0.9 | 1.2 | -0.02 | 1.0 | 0.9 | 0.01 | 1.2 | 1.1 | 0.00 |
| Malignant lymphoma | 0.5 | 0.7 | -0.03 | 0.5 | 0.5 | 0.01 | 0.6 | 0.6 | 0.00 |
| Malignant neoplasm of anorectum | 0.1 | 0.2 | -0.04 | <0.1 | 0.1 | -0.02 | <0.1 | 0.1 | -0.02 |
| Malignant neoplastic disease | 7.1 | 9.9 | -0.10 | 7.3 | 7.3 | 0.00 | 9.3 | 9.2 | 0.00 |

| (Continued) | Before ajustment | | | After matching | | | After stratification | | |
|---|---|---|---|---|---|---|---|---|---|
| Characteristic | T (%) | C (%) | StdDiff | T (%) | C (%) | StdDiff | T (%) | C (%) | StdDiff |
| Malignant tumor of breast | 1.0 | 1.3 | -0.03 | 1.1 | 1.0 | 0.00 | 1.0 | 1.2 | -0.02 |
| Malignant tumor of colon | 0.4 | 0.6 | -0.03 | 0.3 | 0.3 | 0.00 | 0.2 | 0.5 | -0.05 |
| Malignant tumor of lung | 0.2 | 0.5 | -0.04 | 0.2 | 0.4 | -0.03 | 0.2 | 0.5 | -0.04 |
| Malignant tumor of urinary bladder | 0.4 | 0.6 | -0.03 | 0.4 | 0.5 | -0.01 | 0.4 | 0.5 | -0.01 |
| Primary malignant neoplasm of prostate | 1.7 | 2.2 | -0.04 | 1.7 | 1.6 | 0.01 | 1.9 | 2.0 | -0.01 |
| **Medication use** | | | | | | | | | |
| Agents acting on the renin-angiotensin system | 75.2 | 73.0 | 0.05 | 74.5 | 74.3 | 0.00 | 73.0 | 73.3 | -0.01 |
| Antibacterials for systemic use | 45.0 | 46.2 | -0.02 | 44.7 | 44.9 | 0.00 | 48.8 | 46.0 | 0.06 |
| Antidepressants | 31.9 | 32.0 | 0.00 | 32.2 | 32.4 | 0.00 | 36.5 | 32.4 | 0.09 |
| Antiepileptics | 21.1 | 22.2 | -0.03 | 21.3 | 21.4 | 0.00 | 25.0 | 22.9 | 0.05 |
| Antiinflammatory and antirheumatic products | 71.6 | 69.5 | 0.05 | 70.8 | 70.7 | 0.00 | 70.1 | 70.3 | 0.00 |
| Antineoplastic agents | 4.9 | 4.8 | 0.01 | 4.8 | 4.7 | 0.00 | 5.7 | 4.9 | 0.04 |
| Antipsoriatics | 0.4 | 0.6 | -0.03 | 0.4 | 0.5 | -0.02 | 0.7 | 0.6 | 0.01 |
| Antithrombotic agents | 68.3 | 68.9 | -0.01 | 67.4 | 66.9 | 0.01 | 67.0 | 68.3 | -0.03 |
| Beta blocking agents | 58.9 | 59.5 | -0.01 | 57.7 | 57.8 | 0.00 | 58.3 | 58.5 | 0.00 |
| Calcium channel blockers | 30.2 | 30.4 | 0.00 | 29.9 | 30.0 | 0.00 | 30.9 | 30.6 | 0.01 |
| Diuretics | 45.2 | 48.4 | -0.06 | 45.0 | 44.5 | 0.01 | 47.7 | 47.1 | 0.01 |
| Drugs for acid related disorders | 46.4 | 47.8 | -0.03 | 46.2 | 45.7 | 0.01 | 49.9 | 47.8 | 0.04 |
| Drugs for obstructive airway diseases | 46.3 | 46.1 | 0.00 | 48.9 | 46.8 | 0.04 | 48.9 | 46.8 | 0.04 |
| Immunosuppressants | 3.1 | 2.8 | 0.02 | 3.0 | 3.0 | 0.00 | 3.8 | 3.0 | 0.04 |
| Lipid modifying agents | 86.7 | 82.4 | 0.12 | 85.7 | 85.5 | 0.00 | 83.1 | 83.1 | 0.00 |
| Opioids | 31.7 | 36.2 | -0.10 | 32.1 | 32.0 | 0.00 | 38.4 | 35.5 | 0.06 |
| Psycholeptics | 33.4 | 34.4 | -0.02 | 33.1 | 33.4 | -0.01 | 36.3 | 34.3 | 0.04 |
| Psychostimulants | 3.4 | 2.3 | 0.07 | 3.2 | 3.1 | 0.00 | 2.9 | 2.6 | 0.02 |

**Supplementary Table 26:** Baseline patient characteristics for SGLT2is (T) and SUs (C) new-users in the SIDIAP data source. We report proportion of initiators satisfying selected base-line characteristics and the standardized difference of population proportions (StdDiff) before and after propensity score adjustment. Less extreme StdDiffs through matching and stratification suggest improved balance between patient cohorts through adjustment.

| | Before adjustment | | | After matching | | | After stratification | | |
|---|---|---|---|---|---|---|---|---|---|
| Characteristic | T (%) | C (%) | StdDiff | T (%) | C (%) | StdDiff | T (%) | C (%) | StdDiff |
| **Age group** | | | | | | | | | |
| 35 - 39 | <0.2 | 0.2 | -0.05 | <0.2 | <0.4 | -0.01 | <0.2 | <0.4 | -0.01 |
| 40 - 44 | 1.0 | 1.1 | -0.01 | <1.6 | 1.6 | -0.03 | 1.2 | 0.8 | 0.04 |
| 45 - 49 | 2.8 | 2.5 | 0.02 | 5.6 | 3.4 | 0.11 | 4.0 | 2.7 | 0.07 |
| 50 - 54 | 6.3 | 6.2 | 0.00 | 7.8 | 5.7 | 0.09 | 6.7 | 3.6 | 0.14 |
| 55 - 59 | 11.9 | 10.9 | 0.03 | 12.5 | 9.8 | 0.09 | 12.3 | 19.0 | -0.19 |
| 60 - 64 | 16.6 | 15.2 | 0.04 | 16.9 | 16.7 | 0.01 | 16.3 | 16.3 | 0.00 |
| 65 - 69 | 18.9 | 16.3 | 0.07 | 20.4 | 20.4 | 0.00 | 18.9 | 17.3 | 0.04 |
| 70 - 74 | 17.7 | 16.2 | 0.04 | 16.9 | 17.3 | -0.01 | 17.9 | 10.8 | 0.20 |
| 75 - 79 | 13.6 | 13.9 | -0.01 | 10.0 | 11.3 | -0.04 | 12.6 | 8.5 | 0.14 |
| 80 - 84 | 7.3 | 10.5 | -0.11 | 5.3 | 8.5 | -0.12 | 6.5 | 4.9 | 0.07 |
| 85 - 89 | 3.0 | 5.6 | -0.13 | 2.2 | 4.3 | -0.12 | 2.8 | 15.2 | -0.44 |
| 90 - 94 | 0.7 | 1.0 | -0.04 | <1.6 | 0.9 | -0.08 | 0.6 | 0.8 | -0.03 |
| 95 - 99 | <0.2 | 0.2 | -0.02 | <1.6 | <0.5 | 0.06 | <0.2 | <0.4 | 0.04 |
| Gender: female | 21.0 | 28.8 | -0.18 | 30.7 | 28.8 | 0.04 | 25.4 | 21.1 | 0.10 |
| **Medical history: General** | | | | | | | | | |
| Acute respiratory disease | 13.2 | 11.5 | 0.05 | 14.7 | 16.9 | -0.06 | 14.0 | 12.7 | 0.04 |
| Chronic liver disease | 0.3 | 0.6 | -0.05 | <1.6 | <0.5 | 0.04 | 0.5 | <0.4 | 0.08 |
| Chronic obstructive lung disease | 2.3 | 2.9 | -0.04 | 1.9 | 1.8 | 0.01 | 2.1 | 1.0 | 0.09 |
| Dementia | 0.3 | 1.3 | -0.11 | 0.3 | 0.5 | -0.04 | 0.3 | 0.5 | -0.04 |
| Depressive disorder | 1.0 | 3.7 | -0.18 | <1.6 | 2.7 | -0.11 | 1.3 | 1.9 | -0.05 |
| Gastroesophageal reflux disease | 1.4 | 0.8 | 0.06 | 1.9 | 2.8 | -0.06 | 1.3 | 3.6 | -0.15 |
| Gastrointestinal hemorrhage | 1.3 | 1.4 | -0.01 | 1.9 | 1.5 | 0.03 | 1.5 | 0.7 | 0.08 |
| Hyperlipidemia | 1.9 | 4.0 | -0.13 | 1.9 | 1.3 | 0.04 | 1.7 | 0.9 | 0.07 |
| Hypertensive disorder | 3.2 | 6.0 | -0.13 | 4.1 | 3.3 | 0.04 | 3.0 | 15.7 | -0.44 |
| Lesion of liver | <0.2 | 0.6 | -0.07 | <0.2 | <0.4 | -0.02 | <0.2 | <0.4 | -0.02 |
| Obesity | 5.9 | 7.9 | -0.08 | 8.2 | 8.9 | -0.03 | 7.1 | 7.0 | 0.00 |
| Osteoarthritis | 3.4 | 5.4 | -0.10 | 4.4 | 4.4 | 0.00 | 4.1 | 2.6 | 0.08 |
| Pneumonia | 2.2 | 2.2 | 0.00 | <1.6 | 1.8 | -0.04 | 1.8 | 1.0 | 0.07 |
| Psoriasis | 0.2 | 0.7 | -0.07 | <1.6 | 0.8 | -0.07 | <0.2 | <0.4 | -0.01 |
| Renal impairment | 2.0 | 3.2 | -0.07 | 2.2 | 2.2 | 0.00 | 1.9 | 2.4 | -0.03 |
| Rheumatoid arthritis | <0.2 | <0.2 | 0.00 | <1.6 | <0.5 | 0.13 | 0.7 | <0.4 | 0.11 |
| Urinary tract infectious disease | 4.8 | 3.9 | 0.05 | 6.0 | 5.5 | 0.02 | 5.3 | 3.8 | 0.07 |
| Viral hepatitis C | <0.2 | <0.2 | -0.02 | <1.6 | <0.5 | 0.05 | 0.4 | <0.4 | 0.07 |
| Visual system disorder | 13.3 | 17.0 | -0.10 | 20.4 | 20.6 | 0.00 | 14.6 | 14.2 | 0.01 |
| **Medical history: Cardiovascular disease** | | | | | | | | | |
| Atrial fibrillation | 3.3 | 3.1 | 0.01 | <1.6 | 2.4 | -0.06 | 2.9 | 1.6 | 0.09 |
| Cerebrovascular disease | 2.4 | 8.3 | -0.26 | 5.3 | 6.3 | -0.04 | 3.4 | 3.2 | 0.01 |
| Heart disease | 31.4 | 27.7 | 0.08 | 22.3 | 23.0 | -0.02 | 28.4 | 18.6 | 0.23 |
| Heart failure | 5.0 | 3.9 | 0.05 | 2.8 | 2.6 | 0.02 | 4.2 | 1.2 | 0.19 |
| Ischemic heart disease | 22.4 | 19.3 | 0.08 | 14.7 | 13.8 | 0.03 | 20.3 | 10.9 | 0.26 |
| Peripheral vascular disease | 3.5 | 6.5 | -0.14 | 6.0 | 5.1 | 0.04 | 3.7 | 2.5 | 0.07 |
| Pulmonary embolism | <0.2 | 0.2 | -0.03 | <0.2 | <0.4 | 0.02 | <0.2 | <0.4 | 0.02 |
| Venous thrombosis | 0.5 | 1.1 | -0.06 | <1.6 | 1.6 | -0.13 | 0.5 | 0.8 | -0.05 |
| **Medical history: Neoplasms** | | | | | | | | | |
| Malignant neoplastic disease | 2.1 | 2.8 | -0.05 | 2.8 | 4.0 | -0.06 | 1.9 | 2.4 | -0.03 |
| Malignant tumor of breast | <0.2 | 0.2 | -0.04 | <0.2 | <0.4 | -0.01 | <0.2 | <0.4 | -0.01 |
| Malignant tumor of colon | <0.2 | 0.4 | -0.04 | <1.6 | <0.5 | 0.02 | <0.2 | <0.4 | 0.00 |
| Malignant tumor of urinary bladder | <0.2 | 0.5 | -0.06 | <0.2 | <0.4 | -0.03 | <0.2 | <0.4 | -0.03 |
| Primary malignant neoplasm of prostate | <0.2 | 0.3 | -0.03 | <1.6 | 1.0 | -0.08 | <0.2 | 0.9 | -0.10 |
| **Medication use** | | | | | | | | | |
| Agents acting on the renin-angiotensin system | 82.4 | 69.1 | 0.31 | 73.7 | 75.6 | -0.04 | 79.9 | 65.4 | 0.33 |
| Antibacterials for systemic use | 39.5 | 35.3 | 0.09 | 44.5 | 44.3 | 0.00 | 41.1 | 46.0 | -0.10 |
| Antidepressants | 19.1 | 20.9 | -0.04 | 23.8 | 24.5 | -0.02 | 21.4 | 16.7 | 0.12 |
| Antiepileptics | 11.0 | 9.2 | 0.06 | 13.5 | 14.1 | -0.02 | 13.6 | 9.1 | 0.14 |
| Antiinflammatory and antirheumatic products | 82.2 | 72.3 | 0.24 | 82.1 | 79.6 | 0.06 | 81.0 | 74.0 | 0.17 |
| Antineoplastic agents | 1.4 | 1.6 | -0.01 | 2.2 | 0.9 | 0.10 | 1.8 | <0.4 | 0.14 |
| Antipsoriatics | 1.7 | 1.4 | 0.03 | 1.9 | 1.1 | 0.06 | 2.0 | 0.6 | 0.12 |
| Antithrombotic agents | 91.9 | 78.3 | 0.39 | 82.8 | 83.2 | -0.01 | 87.3 | 86.4 | 0.03 |
| Beta blocking agents | 72.6 | 50.6 | 0.46 | 58.9 | 59.5 | -0.01 | 68.7 | 69.7 | -0.02 |
| Calcium channel blockers | 33.1 | 32.1 | 0.02 | 32.6 | 31.0 | 0.04 | 32.6 | 22.9 | 0.22 |
| Diuretics | 55.7 | 50.0 | 0.11 | 47.6 | 50.8 | -0.06 | 54.3 | 48.0 | 0.12 |

| (Continued) | Before ajustment | | | After matching | | | After stratification | | |
|---|---|---|---|---|---|---|---|---|---|
| Characteristic | T (%) | C (%) | StdDiff | T (%) | C (%) | StdDiff | T (%) | C (%) | StdDiff |
| Drugs for acid related disorders | 76.4 | 68.5 | 0.18 | 70.5 | 70.8 | -0.01 | 74.2 | 67.9 | 0.14 |
| Drugs for obstructive airway diseases | 32.1 | 27.3 | 0.10 | 29.8 | 29.4 | 0.01 | 31.9 | 24.7 | 0.16 |
| Immunosuppressants | 1.6 | 1.2 | 0.04 | 2.2 | 1.1 | 0.09 | 1.5 | 0.5 | 0.09 |
| Lipid modifying agents | 92.0 | 79.0 | 0.38 | 83.7 | 84.6 | -0.02 | 89.0 | 88.6 | 0.01 |
| Opioids | 20.0 | 16.5 | 0.09 | 21.3 | 23.4 | -0.05 | 21.5 | 15.5 | 0.15 |
| Psycholeptics | 30.2 | 33.4 | -0.07 | 35.4 | 37.0 | -0.03 | 32.1 | 26.5 | 0.12 |
| Psychostimulants | 24.0 | 13.7 | 0.26 | 11.6 | 13.6 | -0.06 | 21.0 | 11.7 | 0.25 |

**Supplementary Table 27:** Baseline patient characteristics for SGLT2is (T) and SUs (C) new-users in the VA data source. We report proportion of initiators satisfying selected base-line characteristics and the standardized difference of population proportions (StdDiff) before and after propensity score adjustment. Less extreme StdDiffs through matching and stratification suggest improved balance between patient cohorts through adjustment.

| | Before adjustment | | | After matching | | | After stratification | | |
|---|---|---|---|---|---|---|---|---|---|
| Characteristic | T (%) | C (%) | StdDiff | T (%) | C (%) | StdDiff | T (%) | C (%) | StdDiff |
| **Age group** | | | | | | | | | |
| 30 - 34 | <0.1 | 0.1 | -0.03 | <0.2 | 0.1 | -0.03 | <0.1 | 0.1 | -0.03 |
| 35 - 39 | 0.2 | 0.3 | -0.01 | 0.3 | 0.3 | 0.00 | 0.2 | 0.4 | -0.03 |
| 40 - 44 | 0.6 | 0.9 | -0.02 | 0.7 | 0.8 | -0.01 | 0.7 | 0.8 | -0.02 |
| 45 - 49 | 2.1 | 2.5 | -0.02 | 2.1 | 2.2 | 0.00 | 2.4 | 2.4 | 0.00 |
| 50 - 54 | 4.7 | 5.9 | -0.05 | 4.7 | 5.4 | -0.03 | 4.1 | 5.2 | -0.05 |
| 55 - 59 | 8.0 | 11.7 | -0.12 | 8.1 | 7.9 | 0.01 | 14.1 | 8.6 | 0.18 |
| 60 - 64 | 13.4 | 19.2 | -0.16 | 13.5 | 13.7 | -0.01 | 13.0 | 13.8 | -0.02 |
| 65 - 69 | 17.7 | 21.4 | -0.09 | 17.8 | 17.7 | 0.00 | 22.0 | 23.2 | -0.03 |
| 70 - 74 | 31.9 | 17.1 | 0.35 | 31.8 | 30.3 | 0.03 | 27.7 | 26.1 | 0.04 |
| 75 - 79 | 14.5 | 10.9 | 0.11 | 13.8 | 13.6 | 0.01 | 9.3 | 10.7 | -0.04 |
| 80 - 84 | 4.3 | 6.9 | -0.11 | 4.4 | 4.9 | -0.02 | 4.1 | 5.1 | -0.04 |
| 85 - 89 | 1.8 | 2.7 | -0.06 | 2.1 | 2.2 | -0.01 | 2.0 | 2.7 | -0.04 |
| 90 - 94 | 0.4 | 0.6 | -0.03 | 0.4 | 0.8 | -0.04 | 0.2 | 0.8 | -0.08 |
| 95 - 99 | <0.1 | 0.1 | -0.01 | <0.2 | 0.1 | -0.02 | <0.1 | 0.1 | -0.04 |
| Gender: female | 3.4 | 2.9 | 0.02 | 3.6 | 3.8 | -0.01 | 3.5 | 3.9 | -0.02 |
| **Race** | | | | | | | | | |
| Asian | 1.0 | 0.5 | 0.06 | 0.9 | 0.8 | 0.02 | 0.4 | 0.7 | -0.03 |
| Black or African American | 13.6 | 10.8 | 0.09 | 12.3 | 12.4 | 0.00 | 13.9 | 12.8 | 0.03 |
| White | 78.1 | 78.0 | 0.00 | 79.3 | 79.3 | 0.00 | 79.7 | 79.3 | 0.01 |
| Unknown | 5.5 | 9.3 | -0.14 | 5.7 | 5.7 | 0.00 | 5.0 | 5.6 | -0.03 |
| Native Hawaiian or Other Pacific Islander | 0.8 | 0.8 | 0.01 | 0.9 | 1.0 | 0.00 | 0.5 | 0.9 | -0.04 |
| American Indian or Alaska Native | 0.9 | 0.6 | 0.03 | 0.8 | 0.8 | 0.00 | 0.5 | 0.8 | -0.05 |
| **Ethnicity** | | | | | | | | | |
| Hispanic or Latino | 5.7 | 4.6 | 0.05 | 5.7 | 5.5 | 0.01 | 4.2 | 5.6 | -0.06 |
| Not Hispanic or Latino | 91.9 | 89.8 | 0.07 | 92.0 | 92.2 | -0.01 | 92.8 | 92.1 | 0.03 |
| **Medical history: General** | | | | | | | | | |
| Acute respiratory disease | 9.8 | 10.3 | -0.02 | 9.9 | 9.3 | 0.02 | 13.2 | 11.2 | 0.06 |
| Attention deficit hyperactivity disorder | 0.5 | 0.3 | 0.04 | 0.4 | 0.5 | -0.01 | 0.6 | 0.4 | 0.03 |
| Chronic liver disease | 2.4 | 2.4 | 0.00 | 2.3 | 2.6 | -0.02 | 1.8 | 3.0 | -0.07 |
| Chronic obstructive lung disease | 18.5 | 16.3 | 0.06 | 17.3 | 17.7 | -0.01 | 15.1 | 18.0 | -0.08 |
| Crohn’s disease | 0.2 | 0.2 | 0.00 | 0.3 | 0.2 | 0.02 | 0.1 | 0.2 | -0.02 |
| Dementia | 1.6 | 1.6 | 0.00 | 1.6 | 1.8 | -0.02 | 1.8 | 2.2 | -0.03 |
| Depressive disorder | 22.3 | 21.5 | 0.02 | 21.9 | 22.4 | -0.01 | 24.7 | 23.0 | 0.04 |
| Gastroesophageal reflux disease | 23.2 | 22.4 | 0.02 | 22.7 | 23.4 | -0.02 | 22.7 | 23.7 | -0.02 |
| Gastrointestinal hemorrhage | 1.6 | 1.7 | -0.01 | 1.8 | 1.5 | 0.02 | 1.3 | 1.6 | -0.02 |
| Human immunodeficiency virus infection | 0.5 | 0.3 | 0.03 | 0.4 | 0.5 | -0.01 | 0.2 | 0.5 | -0.04 |
| Hyperlipidemia | 77.1 | 78.1 | -0.02 | 75.5 | 76.2 | -0.02 | 77.2 | 75.3 | 0.04 |
| Hypertensive disorder | 82.0 | 82.6 | -0.02 | 81.0 | 81.0 | 0.00 | 80.7 | 80.5 | 0.00 |
| Lesion of liver | 2.0 | 1.2 | 0.07 | 2.0 | 2.3 | -0.02 | 1.9 | 2.0 | -0.01 |
| Obesity | 31.7 | 28.0 | 0.08 | 29.9 | 30.2 | -0.01 | 28.6 | 28.0 | 0.01 |
| Osteoarthritis | 22.7 | 24.2 | -0.04 | 22.6 | 22.1 | 0.01 | 22.1 | 23.3 | -0.03 |
| Pneumonia | 2.6 | 2.0 | 0.04 | 2.3 | 2.3 | 0.00 | 1.9 | 2.5 | -0.04 |
| Psoriasis | 1.9 | 1.6 | 0.02 | 2.1 | 1.7 | 0.03 | 1.2 | 1.7 | -0.04 |
| Renal impairment | 12.6 | 7.8 | 0.16 | 8.5 | 11.5 | -0.10 | 8.5 | 11.5 | -0.10 |
| Rheumatoid arthritis | 0.9 | 1.0 | -0.01 | 1.0 | 0.7 | 0.03 | 0.7 | 0.9 | -0.03 |
| Schizophrenia | 0.6 | 1.0 | -0.05 | 0.6 | 0.7 | -0.01 | 1.2 | 0.8 | 0.04 |
| Ulcerative colitis | 0.4 | 0.3 | 0.01 | 0.3 | 0.4 | -0.01 | 0.3 | 0.4 | -0.01 |
| Urinary tract infectious disease | 1.8 | 2.9 | -0.07 | 2.0 | 2.1 | -0.01 | 2.2 | 2.9 | -0.05 |
| Viral hepatitis C | 1.3 | 1.8 | -0.04 | 1.3 | 1.4 | -0.01 | 0.9 | 1.8 | -0.08 |
| Visual system disorder | 47.4 | 47.5 | 0.00 | 47.3 | 46.1 | 0.02 | 49.8 | 49.5 | 0.01 |
| **Medical history: Cardiovascular disease** | | | | | | | | | |
| Atrial fibrillation | 17.1 | 9.2 | 0.24 | 14.4 | 15.3 | -0.03 | 12.1 | 12.2 | 0.00 |
| Cerebrovascular disease | 7.9 | 10.5 | -0.09 | 7.8 | 7.3 | 0.02 | 8.1 | 7.9 | 0.01 |
| Coronary arteriosclerosis | 57.1 | 37.4 | 0.40 | 50.7 | 52.6 | -0.04 | 43.7 | 41.1 | 0.05 |
| Heart disease | 73.9 | 62.2 | 0.25 | 67.5 | 68.9 | -0.03 | 55.8 | 59.0 | -0.06 |
| Heart failure | 20.3 | 9.3 | 0.31 | 13.2 | 14.4 | -0.04 | 11.1 | 10.6 | 0.01 |
| Ischemic heart disease | 25.2 | 29.6 | -0.10 | 20.5 | 20.6 | 0.00 | 17.9 | 19.2 | -0.03 |
| Peripheral vascular disease | 11.4 | 11.9 | -0.02 | 11.2 | 11.4 | 0.00 | 13.5 | 11.4 | 0.07 |
| Pulmonary embolism | 1.0 | 0.7 | 0.03 | 1.0 | 1.2 | -0.01 | 0.8 | 1.1 | -0.03 |
| Venous thrombosis | 1.3 | 1.4 | -0.01 | 1.4 | 1.8 | -0.03 | 1.3 | 1.9 | -0.05 |
| **Medical history: Neoplasms** | | | | | | | | | |

| (Continued) | Before ajustment | | | After matching | | | After stratification | | |
|---|---|---|---|---|---|---|---|---|---|
| Characteristic | T (%) | C (%) | StdDiff | T (%) | C (%) | StdDiff | T (%) | C (%) | StdDiff |
| Hematologic neoplasm | 1.3 | 1.0 | 0.03 | 1.2 | 1.4 | -0.02 | 0.9 | 1.3 | -0.04 |
| Malignant lymphoma | 0.9 | 0.5 | 0.05 | 0.5 | 0.8 | -0.04 | 0.5 | 0.8 | -0.04 |
| Malignant neoplasm of anorectum | <0.1 | 0.2 | -0.02 | <0.2 | 0.1 | -0.01 | <0.1 | 0.1 | -0.03 |
| Malignant neoplastic disease | 10.9 | 11.4 | -0.02 | 10.8 | 10.6 | 0.01 | 9.3 | 11.3 | -0.06 |
| Malignant tumor of breast | <0.1 | 0.1 | -0.02 | <0.2 | 0.1 | 0.00 | 0.8 | 0.1 | 0.10 |
| Malignant tumor of colon | 0.4 | 0.8 | -0.04 | 0.4 | 0.5 | -0.01 | 0.3 | 0.5 | -0.04 |
| Malignant tumor of lung | 0.6 | 0.3 | 0.05 | 0.4 | 0.6 | -0.03 | 0.4 | 0.6 | -0.03 |
| Malignant tumor of urinary bladder | 0.8 | 0.9 | -0.01 | 0.8 | 0.7 | 0.01 | 1.1 | 0.9 | 0.02 |
| Primary malignant neoplasm of prostate | 3.8 | 4.6 | -0.04 | 3.7 | 3.8 | 0.00 | 2.7 | 4.0 | -0.07 |
| **Medication use** | | | | | | | | | |
| Agents acting on the renin-angiotensin system | 73.2 | 75.9 | -0.06 | 70.7 | 71.1 | -0.01 | 67.2 | 70.6 | -0.07 |
| Antibacterials for systemic use | 29.4 | 33.8 | -0.09 | 28.9 | 28.9 | 0.00 | 29.6 | 33.2 | -0.08 |
| Antidepressants | 35.6 | 36.4 | -0.02 | 36.8 | 36.6 | 0.00 | 42.2 | 38.3 | 0.08 |
| Antiepileptics | 28.6 | 23.3 | 0.12 | 29.3 | 29.2 | 0.00 | 34.1 | 30.9 | 0.07 |
| Antiinflammatory and antirheumatic products | 56.5 | 50.5 | 0.12 | 54.0 | 53.9 | 0.00 | 54.4 | 53.3 | 0.02 |
| Antineoplastic agents | 5.0 | 3.3 | 0.08 | 5.3 | 5.0 | 0.01 | 4.5 | 4.4 | 0.00 |
| Antipsoriatics | 1.0 | 0.8 | 0.02 | 1.0 | 1.0 | 0.01 | 1.1 | 0.9 | 0.02 |
| Antithrombotic agents | 61.5 | 49.9 | 0.24 | 56.8 | 57.4 | -0.01 | 57.8 | 52.2 | 0.11 |
| Beta blocking agents | 66.9 | 61.9 | 0.11 | 61.7 | 63.0 | -0.03 | 60.2 | 59.0 | 0.03 |
| Calcium channel blockers | 31.6 | 31.2 | 0.01 | 31.4 | 31.1 | 0.01 | 32.4 | 30.9 | 0.03 |
| Diuretics | 46.7 | 49.2 | -0.05 | 42.8 | 43.8 | -0.02 | 45.4 | 44.4 | 0.02 |
| Drugs for acid related disorders | 49.1 | 50.1 | -0.02 | 48.3 | 48.2 | 0.00 | 51.7 | 50.1 | 0.03 |
| Drugs for obstructive airway diseases | 45.4 | 39.1 | 0.13 | 44.6 | 44.7 | 0.00 | 48.7 | 44.6 | 0.08 |
| Immunosuppressants | 3.1 | 1.7 | 0.09 | 3.3 | 2.7 | 0.03 | 1.9 | 2.6 | -0.04 |
| Lipid modifying agents | 89.2 | 86.0 | 0.10 | 88.1 | 87.6 | 0.02 | 88.3 | 85.7 | 0.08 |
| Opioids | 17.3 | 23.8 | -0.16 | 16.7 | 16.6 | 0.00 | 20.5 | 21.3 | -0.02 |
| Psycholeptics | 24.8 | 24.1 | 0.02 | 24.2 | 23.6 | 0.01 | 27.0 | 25.2 | 0.04 |
| Psychostimulants | 1.2 | 1.4 | -0.01 | 1.2 | 1.4 | -0.01 | 1.9 | 1.5 | 0.03 |

**Supplementary Table 28:** Baseline patient characteristics for GLP1-RAs (T) and DPP4is (C) new-users in the CCAE data source. We report proportion of initiators satisfying selected base-line characteristics and the standardized difference of population proportions (StdDiff) before and after propensity score adjustment. Less extreme StdDiffs through matching and stratification suggest improved balance between patient cohorts through adjustment.

| | Before adjustment | | | After matching | | | After stratification | | |
|---|---|---|---|---|---|---|---|---|---|
| Characteristic | T (%) | C (%) | StdDiff | T (%) | C (%) | StdDiff | T (%) | C (%) | StdDiff |
| **Age group** | | | | | | | | | |
| 25 - 29 | 0.2 | 0.1 | -0.03 | 0.2 | 0.1 | -0.03 | 0.2 | 0.1 | -0.01 |
| 30 - 34 | 0.9 | 0.5 | -0.05 | 0.9 | 0.4 | -0.06 | 0.7 | 0.5 | -0.02 |
| 35 - 39 | 2.6 | 1.5 | -0.08 | 2.3 | 1.8 | -0.03 | 2.1 | 1.6 | -0.04 |
| 40 - 44 | 6.3 | 4.3 | -0.09 | 5.8 | 5.8 | 0.00 | 5.0 | 4.6 | -0.02 |
| 45 - 49 | 11.5 | 9.3 | -0.07 | 11.2 | 10.5 | -0.02 | 10.3 | 9.8 | -0.01 |
| 50 - 54 | 20.2 | 18.1 | -0.05 | 20.0 | 19.5 | -0.01 | 19.2 | 18.5 | -0.02 |
| 55 - 59 | 27.0 | 27.6 | 0.01 | 27.1 | 26.6 | -0.01 | 27.9 | 27.3 | -0.01 |
| 60 - 64 | 28.6 | 35.0 | 0.14 | 29.8 | 32.3 | 0.05 | 31.1 | 34.0 | 0.06 |
| 65 - 69 | 2.4 | 3.6 | 0.07 | 2.7 | 2.9 | 0.02 | 3.3 | 3.4 | 0.01 |
| Gender: female | 53.8 | 42.2 | -0.23 | 50.9 | 52.0 | 0.02 | 44.1 | 44.9 | 0.02 |
| **Medical history: General** | | | | | | | | | |
| Acute respiratory disease | 33.5 | 30.6 | -0.06 | 32.7 | 33.2 | 0.01 | 30.5 | 31.4 | 0.02 |
| Attention deficit hyperactivity disorder | 1.5 | 0.7 | -0.08 | 1.3 | 1.1 | -0.02 | 0.9 | 0.8 | -0.01 |
| Chronic liver disease | 2.6 | 4.1 | 0.08 | 4.1 | 3.8 | -0.02 | 4.1 | 3.8 | -0.02 |
| Chronic obstructive lung disease | 5.5 | 6.2 | 0.03 | 5.5 | 6.0 | 0.02 | 5.7 | 6.1 | 0.02 |
| Crohn’s disease | 0.4 | 0.4 | -0.01 | 0.4 | 0.3 | -0.01 | 0.6 | 0.4 | -0.03 |
| Dementia | 0.2 | 0.3 | 0.02 | 0.3 | 0.4 | 0.02 | 0.3 | 0.4 | 0.00 |
| Depressive disorder | 15.7 | 10.6 | -0.15 | 14.1 | 14.7 | 0.02 | 11.7 | 11.8 | 0.00 |
| Gastroesophageal reflux disease | 18.2 | 14.7 | -0.10 | 17.0 | 17.7 | 0.02 | 14.9 | 15.7 | 0.02 |
| Gastrointestinal hemorrhage | 2.8 | 2.7 | -0.01 | 2.7 | 2.8 | 0.01 | 2.9 | 2.7 | -0.01 |
| Human immunodeficiency virus infection | 0.3 | 0.3 | 0.01 | 0.2 | 0.4 | 0.03 | 0.2 | 0.3 | 0.02 |
| Hyperlipidemia | 74.9 | 75.0 | 0.00 | 75.7 | 76.1 | 0.01 | 76.1 | 75.2 | -0.02 |
| Hypertensive disorder | 76.8 | 76.5 | -0.01 | 77.2 | 77.3 | 0.00 | 78.3 | 76.9 | -0.03 |
| Lesion of liver | 0.9 | 0.9 | 0.00 | 0.8 | 0.9 | 0.02 | 0.7 | 0.9 | 0.03 |
| Obesity | 38.0 | 19.6 | -0.42 | 32.5 | 34.8 | 0.05 | 24.6 | 24.4 | 0.00 |
| Osteoarthritis | 29.5 | 23.7 | -0.13 | 28.3 | 28.9 | 0.01 | 24.8 | 25.2 | 0.01 |
| Pneumonia | 3.3 | 3.2 | -0.01 | 3.3 | 3.6 | 0.02 | 3.2 | 3.3 | 0.01 |
| Psoriasis | 2.7 | 1.7 | -0.06 | 2.4 | 2.1 | -0.02 | 1.8 | 1.9 | 0.00 |
| Renal impairment | 5.6 | 5.8 | 0.01 | 5.6 | 6.0 | 0.02 | 5.6 | 5.8 | 0.01 |
| Rheumatoid arthritis | 2.0 | 1.6 | -0.03 | 1.8 | 1.7 | -0.01 | 1.6 | 1.7 | 0.00 |
| Schizophrenia | <0.1 | 0.2 | 0.04 | <0.1 | 0.2 | 0.06 | <0.1 | 0.2 | 0.06 |
| Ulcerative colitis | 0.5 | 0.4 | -0.02 | 0.5 | 0.3 | -0.04 | 0.5 | 0.3 | -0.03 |
| Urinary tract infectious disease | 9.2 | 8.9 | -0.01 | 9.0 | 9.0 | 0.00 | 8.8 | 8.9 | 0.00 |
| Viral hepatitis C | 0.3 | 0.5 | 0.03 | 0.3 | 0.3 | 0.00 | 0.3 | 0.4 | 0.02 |
| Visual system disorder | 30.2 | 28.3 | -0.04 | 29.9 | 30.5 | 0.01 | 28.0 | 28.8 | 0.02 |
| **Medical history: Cardiovascular disease** | | | | | | | | | |
| Atrial fibrillation | 4.8 | 4.8 | 0.00 | 5.0 | 4.7 | -0.01 | 5.2 | 4.8 | -0.02 |
| Cerebrovascular disease | 8.6 | 10.4 | 0.06 | 8.8 | 8.3 | -0.02 | 8.9 | 9.9 | 0.03 |
| Coronary arteriosclerosis | 28.0 | 32.1 | 0.09 | 28.6 | 27.3 | -0.03 | 31.7 | 31.0 | -0.02 |
| Heart disease | 47.0 | 49.4 | 0.05 | 46.9 | 45.6 | -0.03 | 49.4 | 48.4 | -0.02 |
| Heart failure | 5.7 | 5.8 | 0.00 | 5.5 | 6.0 | 0.02 | 5.1 | 5.9 | 0.03 |
| Ischemic heart disease | 16.1 | 16.8 | 0.02 | 15.9 | 15.5 | -0.01 | 17.4 | 16.5 | -0.02 |
| Peripheral vascular disease | 16.1 | 13.7 | -0.07 | 15.5 | 15.2 | -0.01 | 14.2 | 14.2 | 0.00 |
| Pulmonary embolism | 1.4 | 0.8 | -0.05 | 1.3 | 1.1 | -0.02 | 1.0 | 0.9 | -0.01 |
| Venous thrombosis | 2.0 | 1.9 | -0.01 | 2.0 | 2.0 | 0.00 | 1.9 | 1.9 | 0.00 |
| **Medical history: Neoplasms** | | | | | | | | | |
| Hematologic neoplasm | 1.0 | 1.2 | 0.03 | 1.0 | 1.2 | 0.02 | 0.9 | 1.2 | 0.03 |
| Malignant lymphoma | 0.5 | 0.6 | 0.00 | 0.5 | 0.5 | 0.00 | 0.4 | 0.5 | 0.01 |
| Malignant neoplasm of anorectum | 0.1 | 0.2 | 0.03 | 0.1 | <0.1 | -0.01 | 0.1 | 0.2 | 0.02 |
| Malignant neoplastic disease | 7.7 | 8.4 | 0.02 | 7.8 | 8.9 | 0.04 | 8.6 | 8.4 | -0.01 |
| Malignant tumor of breast | 1.5 | 1.5 | 0.01 | 1.5 | 2.2 | 0.06 | 1.3 | 1.6 | 0.03 |
| Malignant tumor of colon | 0.2 | 0.4 | 0.03 | 0.3 | 0.3 | 0.01 | 0.3 | 0.4 | 0.02 |
| Malignant tumor of lung | 0.1 | 0.3 | 0.05 | 0.1 | 0.3 | 0.04 | 0.1 | 0.3 | 0.04 |
| Malignant tumor of urinary bladder | 0.2 | 0.4 | 0.03 | 0.2 | 0.1 | -0.02 | 0.4 | 0.3 | -0.02 |
| Primary malignant neoplasm of prostate | 0.8 | 1.2 | 0.04 | 0.8 | 0.9 | 0.01 | 1.3 | 1.1 | -0.02 |
| **Medication use** | | | | | | | | | |
| Agents acting on the renin-angiotensin system | 70.6 | 73.8 | 0.07 | 71.8 | 71.4 | -0.01 | 74.7 | 73.1 | -0.04 |
| Antibacterials for systemic use | 67.5 | 64.3 | -0.07 | 66.7 | 66.8 | 0.00 | 65.1 | 65.0 | 0.00 |
| Antidepressants | 41.0 | 29.3 | -0.25 | 38.2 | 38.9 | 0.02 | 32.2 | 32.1 | 0.00 |
| Antiepileptics | 21.9 | 16.1 | -0.15 | 20.8 | 21.1 | 0.01 | 17.2 | 17.7 | 0.01 |
| Antiinflammatory and antirheumatic products | 42.3 | 35.2 | -0.15 | 40.8 | 41.0 | 0.00 | 36.8 | 37.1 | 0.00 |

| (Continued) | Before ajustment | | | After matching | | | After stratification | | |
|---|---|---|---|---|---|---|---|---|---|
| Characteristic | T (%) | C (%) | StdDiff | T (%) | C (%) | StdDiff | T (%) | C (%) | StdDiff |
| Antineoplastic agents | 6.8 | 5.6 | -0.05 | 6.3 | 6.7 | 0.02 | 5.5 | 5.8 | 0.01 |
| Antipsoriatics | 1.0 | 0.7 | -0.03 | 0.9 | 0.8 | -0.01 | 0.7 | 0.7 | 0.01 |
| Antithrombotic agents | 23.8 | 26.1 | 0.05 | 24.3 | 24.1 | 0.00 | 24.8 | 25.5 | 0.02 |
| Beta blocking agents | 43.4 | 46.6 | 0.06 | 43.9 | 44.3 | 0.01 | 45.9 | 46.0 | 0.00 |
| Calcium channel blockers | 26.5 | 27.0 | 0.01 | 26.6 | 26.5 | 0.00 | 27.3 | 26.9 | -0.01 |
| Diuretics | 49.4 | 43.7 | -0.12 | 47.7 | 47.1 | -0.01 | 45.2 | 44.9 | -0.01 |
| Drugs for acid related disorders | 35.8 | 32.2 | -0.08 | 32.7 | 33.0 | 0.01 | 32.7 | 33.0 | 0.01 |
| Drugs for obstructive airway diseases | 48.1 | 41.3 | -0.14 | 46.5 | 46.8 | 0.01 | 43.6 | 43.1 | -0.01 |
| Immunosuppressants | 4.5 | 3.0 | -0.07 | 4.0 | 3.8 | -0.01 | 3.3 | 3.3 | 0.00 |
| Lipid modifying agents | 75.1 | 78.3 | 0.07 | 76.2 | 76.0 | 0.00 | 78.6 | 77.5 | -0.03 |
| Opioids | 35.2 | 32.5 | -0.06 | 35.1 | 34.3 | -0.02 | 32.8 | 33.1 | 0.01 |
| Psycholeptics | 34.3 | 28.9 | -0.11 | 33.2 | 33.1 | 0.00 | 29.9 | 30.2 | 0.01 |
| Psychostimulants | 5.9 | 3.2 | -0.13 | 5.1 | 4.9 | -0.01 | 4.0 | 3.8 | -0.01 |

**Supplementary Table 29:** Baseline patient characteristics for GLP1-RAs (T) and DPP4is (C) new-users in the OptumDOD data source. We report proportion of initiators satisfying selected base-line characteristics and the standardized difference of population proportions (StdDiff) before and after propensity score adjustment. Less extreme StdDiffs through matching and stratification suggest improved balance between patient cohorts through adjustment.

| | Before adjustment | | | After matching | | | After stratification | | |
|---|---|---|---|---|---|---|---|---|---|
| Characteristic | T (%) | C (%) | StdDiff | T (%) | C (%) | StdDiff | T (%) | C (%) | StdDiff |
| **Age group** | | | | | | | | | |
| 25 - 29 | 0.1 | 0.1 | -0.02 | 0.1 | <0.1 | -0.01 | 0.4 | 0.1 | -0.08 |
| 30 - 34 | 0.5 | 0.3 | -0.04 | 0.6 | 0.3 | -0.04 | 0.9 | 0.3 | -0.08 |
| 35 - 39 | 1.6 | 0.6 | -0.09 | 1.5 | 0.9 | -0.05 | 1.0 | 0.8 | -0.02 |
| 40 - 44 | 3.4 | 1.6 | -0.11 | 2.8 | 2.5 | -0.02 | 2.2 | 1.8 | -0.02 |
| 45 - 49 | 6.4 | 3.6 | -0.13 | 5.5 | 5.1 | -0.02 | 4.3 | 4.1 | -0.01 |
| 50 - 54 | 11.4 | 6.6 | -0.17 | 9.6 | 9.5 | 0.00 | 9.6 | 9.5 | 0.00 |
| 55 - 59 | 14.9 | 10.1 | -0.14 | 13.6 | 13.5 | 0.00 | 10.4 | 11.1 | 0.02 |
| 60 - 64 | 16.7 | 12.7 | -0.11 | 14.3 | 13.6 | -0.02 | 14.3 | 13.6 | -0.02 |
| 65 - 69 | 17.8 | 16.6 | -0.03 | 19.0 | 18.8 | -0.01 | 16.7 | 16.9 | 0.00 |
| 70 - 74 | 15.1 | 18.9 | 0.10 | 16.8 | 18.8 | 0.05 | 17.3 | 18.6 | 0.03 |
| 75 - 79 | 8.3 | 14.2 | 0.19 | 10.1 | 9.9 | 0.00 | 13.9 | 13.0 | -0.03 |
| 80 - 84 | 2.7 | 9.1 | 0.28 | 3.4 | 3.2 | -0.01 | 7.2 | 7.7 | 0.02 |
| 85 - 89 | 0.8 | 5.1 | 0.26 | 1.0 | 1.2 | 0.02 | 3.8 | 4.2 | 0.02 |
| 90 - 94 | <0.1 | 0.4 | 0.08 | 0.1 | 0.4 | 0.07 | 0.1 | 0.4 | 0.07 |
| Gender: female | 52.9 | 47.3 | -0.11 | 51.0 | 51.9 | 0.02 | 48.5 | 48.4 | 0.00 |
| **Medical history: General** | | | | | | | | | |
| Acute respiratory disease | 30.1 | 29.2 | -0.02 | 29.2 | 30.7 | 0.03 | 30.6 | 29.6 | -0.02 |
| Attention deficit hyperactivity disorder | 1.3 | 0.6 | -0.07 | 0.8 | 1.1 | 0.03 | 0.6 | 0.7 | 0.01 |
| Chronic liver disease | 2.7 | 4.0 | 0.07 | 3.1 | 3.0 | -0.01 | 3.5 | 3.7 | 0.01 |
| Chronic obstructive lung disease | 14.5 | 15.5 | 0.03 | 14.4 | 15.8 | 0.04 | 16.0 | 15.6 | -0.01 |
| Crohn's disease | 0.3 | 0.4 | 0.02 | 0.2 | 0.4 | 0.03 | 0.3 | 0.4 | 0.01 |
| Dementia | 1.8 | 4.7 | 0.17 | 2.0 | 2.3 | 0.02 | 3.3 | 4.1 | 0.04 |
| Depressive disorder | 24.4 | 17.4 | -0.17 | 21.6 | 22.4 | 0.02 | 19.0 | 19.1 | 0.00 |
| Gastroesophageal reflux disease | 26.6 | 23.6 | -0.07 | 25.2 | 26.2 | 0.02 | 25.8 | 24.6 | -0.03 |
| Gastrointestinal hemorrhage | 2.8 | 3.8 | 0.06 | 2.9 | 3.1 | 0.01 | 3.9 | 3.6 | -0.01 |
| Human immunodeficiency virus infection | 0.6 | 0.3 | -0.04 | 0.5 | 0.7 | 0.02 | 0.5 | 0.4 | -0.01 |
| Hyperlipidemia | 85.2 | 87.1 | 0.06 | 85.4 | 85.6 | 0.01 | 87.2 | 86.8 | -0.01 |
| Hypertensive disorder | 86.0 | 88.1 | 0.06 | 86.1 | 87.2 | 0.03 | 89.2 | 87.8 | -0.04 |
| Lesion of liver | 1.5 | 1.5 | 0.00 | 1.5 | 1.4 | -0.01 | 1.2 | 1.5 | 0.03 |
| Obesity | 47.9 | 23.3 | -0.53 | 38.8 | 42.2 | 0.07 | 28.4 | 29.0 | 0.01 |
| Osteoarthritis | 40.2 | 35.9 | -0.09 | 38.6 | 39.3 | 0.02 | 38.4 | 37.2 | -0.03 |
| Pneumonia | 5.4 | 6.3 | 0.04 | 4.9 | 5.1 | 0.00 | 7.4 | 6.1 | -0.05 |
| Psoriasis | 2.7 | 1.8 | -0.07 | 2.5 | 2.1 | -0.03 | 1.8 | 1.9 | 0.00 |
| Renal impairment | 16.0 | 23.7 | 0.19 | 17.5 | 17.8 | 0.01 | 23.6 | 22.2 | -0.03 |
| Rheumatoid arthritis | 2.8 | 2.5 | -0.02 | 2.6 | 3.1 | 0.03 | 3.1 | 2.6 | -0.03 |
| Schizophrenia | 0.6 | 0.5 | -0.01 | 0.5 | 0.6 | 0.01 | 0.4 | 0.5 | 0.02 |
| Ulcerative colitis | 0.4 | 0.5 | 0.01 | 0.4 | 0.3 | -0.01 | 0.6 | 0.4 | -0.03 |
| Urinary tract infectious disease | 11.7 | 15.6 | 0.12 | 12.0 | 12.4 | 0.01 | 16.3 | 14.7 | -0.04 |
| Viral hepatitis C | 0.7 | 0.7 | 0.00 | 0.7 | 0.5 | -0.03 | 0.7 | 0.7 | 0.00 |
| Visual system disorder | 42.8 | 47.7 | 0.10 | 44.4 | 44.9 | 0.01 | 44.4 | 44.9 | 0.01 |
| **Medical history: Cardiovascular disease** | | | | | | | | | |
| Atrial fibrillation | 9.5 | 13.0 | 0.11 | 9.7 | 9.9 | 0.01 | 11.9 | 12.3 | 0.01 |
| Cerebrovascular disease | 11.3 | 16.3 | 0.14 | 12.0 | 11.8 | 0.00 | 14.8 | 15.2 | 0.01 |
| Coronary arteriosclerosis | 33.8 | 38.4 | 0.10 | 35.1 | 35.3 | 0.00 | 37.6 | 37.6 | 0.00 |
| Heart disease | 54.4 | 59.4 | 0.10 | 55.1 | 55.5 | 0.01 | 60.2 | 58.5 | -0.03 |
| Heart failure | 12.0 | 14.7 | 0.08 | 12.0 | 12.2 | 0.01 | 14.7 | 14.3 | -0.01 |
| Ischemic heart disease | 18.5 | 20.5 | 0.05 | 18.3 | 19.0 | 0.02 | 20.4 | 20.1 | -0.01 |
| Peripheral vascular disease | 26.7 | 26.2 | -0.01 | 26.9 | 27.0 | 0.00 | 25.9 | 26.6 | 0.02 |
| Pulmonary embolism | 1.7 | 1.3 | -0.03 | 1.5 | 1.7 | 0.01 | 1.5 | 1.4 | -0.01 |
| Venous thrombosis | 2.3 | 2.5 | 0.01 | 2.3 | 1.9 | -0.03 | 3.0 | 2.5 | -0.03 |
| **Medical history: Neoplasms** | | | | | | | | | |
| Hematologic neoplasm | 1.2 | 1.6 | 0.04 | 1.2 | 1.3 | 0.01 | 1.3 | 1.5 | 0.02 |
| Malignant lymphoma | 0.7 | 0.8 | 0.01 | 0.6 | 0.7 | 0.01 | 0.6 | 0.8 | 0.02 |
| Malignant neoplasm of anorectum | 0.2 | 0.3 | 0.03 | 0.2 | 0.3 | 0.01 | 0.3 | 0.3 | 0.01 |
| Malignant neoplastic disease | 10.9 | 14.3 | 0.10 | 11.5 | 11.0 | -0.01 | 11.5 | 11.0 | -0.01 |
| Malignant tumor of breast | 1.8 | 2.2 | 0.03 | 1.9 | 1.8 | -0.01 | 2.3 | 2.1 | -0.02 |
| Malignant tumor of colon | 0.7 | 0.8 | 0.01 | 0.8 | 0.5 | -0.04 | 0.8 | 0.8 | -0.01 |
| Malignant tumor of lung | 0.3 | 0.7 | 0.05 | 0.4 | 0.5 | 0.03 | 0.9 | 0.7 | -0.03 |
| Malignant tumor of urinary bladder | 0.5 | 0.7 | 0.04 | 0.5 | 0.5 | -0.01 | 1.1 | 0.7 | -0.04 |
| Primary malignant neoplasm of prostate | 1.8 | 3.2 | 0.09 | 2.1 | 2.0 | -0.01 | 3.5 | 2.9 | -0.04 |
| **Medication use** | | | | | | | | | |

| (Continued) | Before ajustment | | | After matching | | | After stratification | | |
|---|---|---|---|---|---|---|---|---|---|
| Characteristic | T (%) | C (%) | StdDiff | T (%) | C (%) | StdDiff | T (%) | C (%) | StdDiff |
| Agents acting on the renin-angiotensin system | 72.7 | 75.8 | 0.07 | 73.6 | 72.5 | -0.03 | 76.1 | 75.0 | -0.03 |
| Antibacterials for systemic use | 64.7 | 63.9 | -0.02 | 63.4 | 65.1 | 0.04 | 64.2 | 64.1 | 0.00 |
| Antidepressants | 43.0 | 31.6 | -0.24 | 39.2 | 41.3 | 0.04 | 33.8 | 34.3 | 0.01 |
| Antiepileptics | 29.3 | 22.2 | -0.16 | 27.0 | 28.2 | 0.03 | 23.9 | 23.9 | 0.00 |
| Antiinflammatory and antirheumatic products | 39.8 | 32.3 | -0.16 | 36.9 | 38.1 | 0.03 | 31.9 | 33.9 | 0.04 |
| Antineoplastic agents | 7.4 | 7.2 | -0.01 | 7.2 | 6.9 | -0.01 | 6.8 | 7.2 | 0.01 |
| Antipsoriatics | 0.9 | 0.7 | -0.02 | 0.8 | 0.7 | 0.00 | 0.8 | 0.7 | 0.00 |
| Antithrombotic agents | 27.4 | 31.5 | 0.09 | 27.9 | 28.7 | 0.02 | 30.0 | 30.9 | 0.02 |
| Beta blocking agents | 47.4 | 52.3 | 0.10 | 48.0 | 48.2 | 0.00 | 52.5 | 51.4 | -0.02 |
| Calcium channel blockers | 30.1 | 33.6 | 0.08 | 30.8 | 31.1 | 0.00 | 30.8 | 31.1 | 0.00 |
| Diuretics | 51.4 | 49.6 | -0.04 | 50.3 | 50.7 | 0.01 | 52.5 | 50.1 | -0.05 |
| Drugs for acid related disorders | 37.0 | 36.2 | -0.02 | 36.2 | 37.2 | 0.02 | 38.6 | 36.6 | -0.04 |
| Drugs for obstructive airway diseases | 50.7 | 43.9 | -0.14 | 48.6 | 50.0 | 0.03 | 47.6 | 45.6 | -0.04 |
| Immunosuppressants | 4.1 | 3.3 | -0.05 | 3.7 | 3.9 | 0.01 | 3.4 | 3.4 | 0.00 |
| Lipid modifying agents | 79.9 | 82.6 | 0.07 | 81.1 | 80.7 | -0.01 | 80.7 | 82.2 | 0.04 |
| Opioids | 36.3 | 33.2 | -0.06 | 34.6 | 36.0 | 0.03 | 32.9 | 33.9 | 0.02 |
| Psycholeptics | 34.1 | 29.7 | -0.09 | 32.1 | 33.6 | 0.03 | 29.0 | 30.8 | 0.04 |
| Psychostimulants | 3.8 | 2.0 | -0.11 | 3.1 | 3.6 | 0.03 | 2.6 | 2.4 | -0.01 |

**Supplementary Table 30:** Baseline patient characteristics for GLP1-RAs (T) and DPP4is (C) new-users in the MDCR data source. We report proportion of initiators satisfying selected base-line characteristics and the standardized difference of population proportions (StdDiff) before and after propensity score adjustment. Less extreme StdDiffs through matching and stratification suggest improved balance between patient cohorts through adjustment.

| | Before adjustment | | | After matching | | | After stratification | | |
|---|---|---|---|---|---|---|---|---|---|
| Characteristic | T (%) | C (%) | StdDiff | T (%) | C (%) | StdDiff | T (%) | C (%) | StdDiff |
| **Age group** | | | | | | | | | |
| 50 - 54 | <0.4 | 0.1 | -0.03 | <0.4 | <0.5 | 0.03 | <0.2 | 0.1 | 0.00 |
| 55 - 59 | 1.1 | 0.4 | -0.07 | 1.1 | 0.7 | -0.04 | 0.6 | 0.5 | -0.01 |
| 60 - 64 | 2.5 | 1.5 | -0.07 | 2.5 | 1.8 | -0.05 | 2.3 | 1.5 | -0.06 |
| 65 - 69 | 34.6 | 22.1 | -0.28 | 33.8 | 35.2 | 0.03 | 23.1 | 23.2 | 0.00 |
| 70 - 74 | 36.8 | 28.8 | -0.17 | 36.5 | 32.0 | -0.10 | 33.5 | 29.1 | -0.09 |
| 75 - 79 | 16.2 | 22.1 | 0.15 | 16.2 | 17.0 | 0.02 | 21.7 | 21.6 | 0.00 |
| 80 - 84 | 6.1 | 15.0 | 0.29 | 6.8 | 6.5 | -0.01 | 10.7 | 14.2 | 0.11 |
| 85 - 89 | 2.0 | 7.3 | 0.26 | 2.2 | 3.7 | 0.09 | 7.8 | 6.9 | -0.03 |
| 90 - 94 | 0.6 | 2.3 | 0.14 | 0.7 | 2.2 | 0.12 | <0.2 | 2.3 | 0.19 |
| Gender: female | 49.2 | 44.1 | -0.10 | 48.4 | 47.1 | -0.03 | 44.6 | 44.4 | 0.00 |
| **Medical history: General** | | | | | | | | | |
| Acute respiratory disease | 25.8 | 25.4 | -0.01 | 26.2 | 24.9 | -0.03 | 28.9 | 25.5 | -0.07 |
| Attention deficit hyperactivity disorder | <0.4 | 0.1 | -0.01 | <0.4 | <0.5 | 0.06 | <0.2 | 0.2 | 0.04 |
| Chronic liver disease | 2.1 | 2.7 | 0.04 | 2.2 | 1.9 | -0.02 | 2.0 | 2.7 | 0.05 |
| Chronic obstructive lung disease | 15.7 | 14.2 | -0.04 | 15.5 | 14.9 | -0.02 | 16.7 | 14.4 | -0.06 |
| Crohn’s disease | 0.5 | 0.3 | -0.03 | <0.4 | <0.5 | -0.06 | 0.3 | 0.3 | 0.01 |
| Dementia | 2.1 | 4.1 | 0.11 | 2.2 | 3.4 | 0.07 | 2.5 | 4.1 | 0.09 |
| Depressive disorder | 13.8 | 8.6 | -0.16 | 13.0 | 12.4 | -0.02 | 10.0 | 9.0 | -0.03 |
| Gastroesophageal reflux disease | 17.2 | 14.6 | -0.07 | 17.1 | 18.5 | 0.04 | 12.2 | 15.0 | 0.08 |
| Gastrointestinal hemorrhage | 2.9 | 4.2 | 0.07 | 2.7 | 3.7 | 0.06 | 2.8 | 4.1 | 0.07 |
| Hyperlipidemia | 69.0 | 65.7 | -0.07 | 71.1 | 77.0 | 0.14 | 63.7 | 66.4 | 0.06 |
| Hypertensive disorder | 81.4 | 78.4 | -0.08 | 82.2 | 82.6 | 0.01 | 82.3 | 78.7 | -0.09 |
| Lesion of liver | 0.9 | 0.9 | 0.00 | 1.0 | 0.9 | 0.00 | 0.4 | 1.0 | 0.07 |
| Obesity | 29.5 | 11.0 | -0.47 | 27.2 | 30.5 | 0.07 | 11.2 | 12.7 | 0.05 |
| Osteoarthritis | 39.4 | 34.6 | -0.10 | 38.5 | 41.3 | 0.06 | 33.5 | 35.1 | 0.03 |
| Pneumonia | 4.8 | 6.6 | 0.08 | 5.0 | 5.1 | 0.00 | 3.5 | 6.4 | 0.13 |
| Psoriasis | 2.9 | 1.6 | -0.09 | 3.2 | 2.3 | -0.06 | 2.2 | 1.7 | -0.04 |
| Renal impairment | 14.5 | 17.8 | 0.09 | 15.0 | 18.3 | 0.09 | 17.6 | 17.7 | 0.00 |
| Rheumatoid arthritis | 1.9 | 2.0 | 0.01 | 1.8 | 2.2 | 0.03 | 1.0 | 2.0 | 0.08 |
| Schizophrenia | <0.4 | 0.2 | -0.03 | <0.4 | <0.5 | -0.02 | <0.2 | 0.2 | 0.00 |
| Ulcerative colitis | <0.4 | 0.6 | 0.04 | <0.4 | 0.8 | 0.05 | <0.2 | 0.6 | 0.07 |
| Urinary tract infectious disease | 11.5 | 12.3 | 0.02 | 11.3 | 10.0 | -0.04 | 11.6 | 12.1 | 0.02 |
| Viral hepatitis C | <0.4 | 0.3 | 0.04 | <0.4 | <0.5 | 0.06 | <0.2 | 0.3 | 0.06 |
| Visual system disorder | 51.6 | 54.7 | 0.06 | 51.8 | 55.0 | 0.06 | 59.5 | 54.6 | -0.10 |
| **Medical history: Cardiovascular disease** | | | | | | | | | |
| Atrial fibrillation | 12.1 | 16.7 | 0.13 | 11.8 | 12.6 | 0.03 | 16.6 | 16.3 | -0.01 |
| Cerebrovascular disease | 17.9 | 21.7 | 0.10 | 18.0 | 18.0 | 0.00 | 18.7 | 21.4 | 0.07 |
| Coronary arteriosclerosis | 43.1 | 43.7 | 0.01 | 42.1 | 42.2 | 0.00 | 44.3 | 43.6 | -0.01 |
| Heart disease | 62.3 | 66.6 | 0.09 | 61.5 | 62.2 | 0.01 | 69.5 | 66.2 | -0.07 |
| Heart failure | 11.3 | 14.5 | 0.09 | 11.1 | 13.5 | 0.07 | 14.8 | 14.3 | -0.01 |
| Ischemic heart disease | 17.9 | 19.4 | 0.04 | 17.5 | 16.9 | -0.02 | 18.2 | 19.3 | 0.03 |
| Peripheral vascular disease | 22.5 | 22.2 | -0.01 | 23.1 | 24.6 | 0.04 | 20.9 | 22.5 | 0.04 |
| Pulmonary embolism | 1.3 | 1.2 | 0.00 | 1.2 | 1.9 | 0.06 | 3.2 | 1.3 | -0.13 |
| Venous thrombosis | 2.1 | 2.7 | 0.04 | 2.0 | 2.3 | 0.02 | 2.5 | 2.7 | 0.01 |
| **Medical history: Neoplasms** | | | | | | | | | |
| Hematologic neoplasm | 2.1 | 2.1 | 0.01 | 2.0 | 1.8 | -0.02 | 1.4 | 2.2 | 0.06 |
| Malignant lymphoma | 1.2 | 1.1 | 0.00 | 1.1 | 0.9 | -0.01 | 0.5 | 1.1 | 0.07 |
| Malignant neoplastic disease | 20.5 | 21.1 | 0.01 | 21.1 | 18.7 | -0.06 | 24.2 | 21.1 | -0.07 |
| Malignant tumor of breast | 3.8 | 2.8 | -0.05 | 3.8 | 3.5 | -0.02 | 3.9 | 2.9 | -0.06 |
| Malignant tumor of colon | 0.5 | 1.0 | 0.06 | 0.6 | 0.9 | 0.04 | 0.2 | 1.0 | 0.10 |
| Malignant tumor of lung | 0.8 | 0.8 | 0.00 | 0.9 | 1.0 | 0.01 | 0.9 | 0.9 | 0.00 |
| Malignant tumor of urinary bladder | 1.8 | 1.5 | -0.02 | 1.9 | 1.4 | -0.04 | 3.7 | 1.5 | -0.14 |
| Primary malignant neoplasm of prostate | 4.6 | 5.0 | 0.02 | 4.5 | 4.8 | 0.01 | 7.5 | 5.0 | -0.10 |
| **Medication use** | | | | | | | | | |
| Agents acting on the renin-angiotensin system | 78.2 | 77.1 | -0.03 | 78.9 | 77.1 | -0.04 | 75.5 | 77.1 | 0.04 |
| Antibacterials for systemic use | 66.7 | 66.1 | -0.01 | 66.9 | 65.7 | -0.03 | 62.0 | 66.1 | 0.09 |
| Antidepressants | 36.6 | 26.6 | -0.22 | 34.6 | 35.1 | 0.01 | 32.2 | 27.4 | -0.10 |
| Antiepileptics | 21.2 | 17.7 | -0.09 | 20.2 | 23.6 | 0.08 | 16.3 | 18.1 | 0.05 |
| Antiinflammatory and antirheumatic products | 34.6 | 29.5 | -0.11 | 33.6 | 34.8 | 0.03 | 29.6 | 30.0 | 0.01 |
| Antineoplastic agents | 10.1 | 9.3 | -0.03 | 9.9 | 9.0 | -0.03 | 8.5 | 9.4 | 0.03 |
| Antipsoriatics | 1.2 | 0.9 | -0.03 | 1.1 | 0.7 | -0.04 | 0.9 | 0.9 | 0.00 |

| (Continued) | Before ajustment | | | After matching | | | After stratification | | |
|---|---|---|---|---|---|---|---|---|---|
| Characteristic | T (%) | C (%) | StdDiff | T (%) | C (%) | StdDiff | T (%) | C (%) | StdDiff |
| Antithrombotic agents | 35.1 | 37.2 | 0.04 | 34.9 | 35.3 | 0.01 | 40.3 | 37.0 | -0.07 |
| Beta blocking agents | 55.9 | 60.9 | 0.10 | 56.7 | 57.3 | 0.01 | 65.8 | 60.5 | -0.11 |
| Calcium channel blockers | 33.7 | 38.4 | 0.10 | 33.7 | 35.1 | 0.03 | 32.3 | 38.0 | 0.12 |
| Diuretics | 56.4 | 54.1 | -0.05 | 55.2 | 56.0 | 0.01 | 53.3 | 54.1 | 0.01 |
| Drugs for acid related disorders | 39.9 | 37.7 | -0.04 | 38.9 | 41.3 | 0.05 | 31.4 | 38.0 | 0.14 |
| Drugs for obstructive airway diseases | 53.1 | 44.4 | -0.18 | 52.2 | 52.7 | 0.01 | 46.6 | 45.1 | -0.03 |
| Immunosuppressants | 4.1 | 3.5 | -0.03 | 3.8 | 4.8 | 0.05 | 2.8 | 3.6 | 0.05 |
| Lipid modifying agents | 86.7 | 84.6 | -0.06 | 86.9 | 85.2 | -0.05 | 85.5 | 84.7 | -0.02 |
| Opioids | 32.1 | 32.1 | 0.00 | 32.1 | 33.5 | 0.03 | 31.3 | 32.1 | 0.02 |
| Psycholeptics | 31.2 | 29.3 | -0.04 | 30.8 | 30.4 | -0.01 | 33.2 | 29.4 | -0.08 |
| Psychostimulants | 2.1 | 1.5 | -0.05 | 2.1 | 1.9 | -0.01 | 1.2 | 1.5 | 0.03 |

**Supplementary Table 31:** Baseline patient characteristics for GLP1-RAs (T) and DPP4is (C) new-users in the Open Claims data source. We report proportion of initiators satisfying selected base-line characteristics and the standardized difference of population proportions (StdDiff) before and after propensity score adjustment. Less extreme StdDiffs through matching and stratification suggest improved balance between patient cohorts through adjustment.

| | Before adjustment | | | After matching | | | After stratification | | |
|---|---|---|---|---|---|---|---|---|---|
| Characteristic | T (%) | C (%) | StdDiff | T (%) | C (%) | StdDiff | T (%) | C (%) | StdDiff |
| **Age group** | | | | | | | | | |
| 20 - 24 | 0.2 | 0.1 | -0.02 | 0.2 | 0.2 | -0.01 | 0.2 | 0.2 | 0.00 |
| 25 - 29 | 0.3 | 0.2 | -0.03 | 0.3 | 0.2 | -0.02 | 0.3 | 0.2 | -0.01 |
| 30 - 34 | 0.8 | 0.4 | -0.05 | 0.6 | 0.6 | 0.00 | 0.5 | 0.5 | -0.01 |
| 35 - 39 | 1.7 | 0.9 | -0.07 | 1.4 | 1.4 | 0.00 | 1.2 | 1.1 | -0.01 |
| 40 - 44 | 3.8 | 2.0 | -0.11 | 3.1 | 3.1 | 0.00 | 2.6 | 2.4 | -0.01 |
| 45 - 49 | 7.1 | 4.2 | -0.12 | 6.2 | 6.0 | -0.01 | 5.0 | 4.9 | -0.01 |
| 50 - 54 | 11.9 | 7.7 | -0.14 | 10.5 | 10.5 | 0.00 | 8.7 | 8.7 | 0.00 |
| 55 - 59 | 16.5 | 11.7 | -0.14 | 15.3 | 15.5 | 0.00 | 13.1 | 13.0 | 0.00 |
| 60 - 64 | 18.3 | 14.9 | -0.09 | 17.7 | 17.9 | 0.00 | 16.0 | 15.9 | 0.00 |
| 65 - 69 | 16.7 | 16.6 | 0.00 | 17.3 | 17.6 | 0.01 | 16.5 | 16.7 | 0.00 |
| 70 - 74 | 12.2 | 15.5 | 0.10 | 13.8 | 13.7 | 0.00 | 14.8 | 14.7 | 0.00 |
| 75 - 79 | 6.8 | 17.5 | 0.33 | 8.5 | 8.3 | -0.01 | 13.8 | 14.7 | 0.03 |
| 80 - 84 | 3.3 | 8.0 | 0.20 | 4.5 | 4.6 | 0.00 | 7.0 | 6.8 | -0.01 |
| 85 - 89 | 0.3 | 0.2 | -0.01 | 0.3 | 0.4 | 0.02 | 0.2 | 0.2 | 0.01 |
| Gender: female | 54.4 | 48.6 | -0.12 | 51.5 | 51.9 | 0.01 | 50.1 | 49.9 | 0.00 |
| **Medical history: General** | | | | | | | | | |
| Acute respiratory disease | 17.1 | 17.7 | 0.01 | 16.6 | 17.0 | 0.01 | 17.8 | 17.7 | 0.00 |
| Attention deficit hyperactivity disorder | 0.9 | 0.3 | -0.07 | 0.6 | 0.6 | 0.00 | 0.5 | 0.4 | -0.01 |
| Chronic liver disease | 1.1 | 1.9 | 0.06 | 1.3 | 1.3 | 0.00 | 1.8 | 1.7 | -0.01 |
| Chronic obstructive lung disease | 8.8 | 10.2 | 0.05 | 9.0 | 9.2 | 0.01 | 9.9 | 10.0 | 0.00 |
| Crohn's disease | 0.2 | 0.2 | 0.00 | 0.2 | 0.2 | 0.00 | 0.2 | 0.2 | 0.00 |
| Dementia | 0.7 | 2.3 | 0.12 | 0.9 | 1.0 | 0.01 | 1.9 | 1.9 | 0.00 |
| Depressive disorder | 12.0 | 8.9 | -0.10 | 10.4 | 10.9 | 0.02 | 10.0 | 9.8 | -0.01 |
| Gastroesophageal reflux disease | 12.7 | 11.4 | -0.04 | 11.7 | 12.0 | 0.01 | 11.9 | 11.7 | 0.00 |
| Gastrointestinal hemorrhage | 1.8 | 2.4 | 0.04 | 1.8 | 1.8 | 0.00 | 2.2 | 2.2 | 0.00 |
| Human immunodeficiency virus infection | 0.4 | 0.3 | -0.01 | 0.4 | 0.4 | 0.01 | 0.3 | 0.3 | 0.00 |
| Hyperlipidemia | 50.1 | 54.4 | 0.09 | 50.2 | 50.5 | 0.01 | 53.3 | 53.5 | 0.00 |
| Hypertensive disorder | 59.4 | 63.9 | 0.09 | 59.3 | 59.4 | 0.00 | 63.0 | 62.8 | 0.00 |
| Lesion of liver | 0.9 | 1.0 | 0.01 | 0.8 | 1.0 | 0.01 | 0.8 | 1.0 | 0.01 |
| Obesity | 23.1 | 11.3 | -0.32 | 17.5 | 18.1 | 0.01 | 15.0 | 14.1 | -0.03 |
| Osteoarthritis | 23.6 | 21.9 | -0.04 | 22.2 | 22.4 | 0.01 | 22.8 | 22.4 | -0.01 |
| Pneumonia | 3.4 | 4.2 | 0.04 | 3.3 | 3.4 | 0.00 | 4.0 | 4.1 | 0.01 |
| Psoriasis | 1.5 | 1.0 | -0.04 | 1.3 | 1.3 | 0.00 | 1.3 | 1.1 | -0.01 |
| Renal impairment | 8.2 | 12.9 | 0.15 | 9.0 | 9.2 | 0.01 | 11.5 | 11.9 | 0.01 |
| Rheumatoid arthritis | 1.8 | 1.6 | -0.01 | 1.6 | 1.6 | 0.00 | 1.6 | 1.6 | 0.00 |
| Schizophrenia | 0.3 | 0.5 | 0.02 | 0.4 | 0.5 | 0.02 | 0.4 | 0.5 | 0.01 |
| Ulcerative colitis | 0.2 | 0.3 | 0.01 | 0.2 | 0.3 | 0.01 | 0.2 | 0.3 | 0.01 |
| Urinary tract infectious disease | 6.5 | 8.7 | 0.08 | 6.6 | 6.7 | 0.00 | 8.1 | 8.2 | 0.00 |
| Viral hepatitis C | 0.3 | 0.5 | 0.03 | 0.4 | 0.4 | 0.00 | 0.5 | 0.5 | 0.00 |
| Visual system disorder | 21.3 | 24.9 | 0.09 | 21.7 | 21.9 | 0.00 | 23.5 | 24.0 | 0.01 |
| **Medical history: Cardiovascular disease** | | | | | | | | | |
| Atrial fibrillation | 6.1 | 8.8 | 0.10 | 6.4 | 6.3 | -0.01 | 8.0 | 8.1 | 0.00 |
| Cerebrovascular disease | 6.4 | 10.2 | 0.14 | 6.9 | 6.8 | 0.00 | 8.8 | 9.2 | 0.01 |
| Coronary arteriosclerosis | 20.2 | 25.2 | 0.12 | 23.7 | 24.1 | 0.01 | 23.7 | 24.1 | 0.01 |
| Heart disease | 35.4 | 42.5 | 0.15 | 36.1 | 36.0 | 0.00 | 40.4 | 40.8 | 0.01 |
| Heart failure | 6.8 | 9.6 | 0.10 | 7.0 | 7.3 | 0.01 | 8.4 | 9.0 | 0.02 |
| Ischemic heart disease | 9.7 | 12.1 | 0.08 | 10.0 | 9.9 | 0.00 | 11.0 | 11.6 | 0.02 |
| Peripheral vascular disease | 13.3 | 13.9 | 0.02 | 13.0 | 13.3 | 0.01 | 13.6 | 13.8 | 0.01 |
| Pulmonary embolism | 1.1 | 0.9 | -0.01 | 1.0 | 1.0 | 0.00 | 1.0 | 0.9 | 0.00 |
| Venous thrombosis | 1.2 | 1.6 | 0.03 | 1.4 | 1.5 | 0.01 | 1.4 | 1.5 | 0.01 |
| **Medical history: Neoplasms** | | | | | | | | | |
| Hematologic neoplasm | 0.8 | 1.1 | 0.03 | 0.9 | 0.8 | -0.01 | 1.0 | 1.0 | 0.00 |
| Malignant lymphoma | 0.4 | 0.6 | 0.02 | 0.5 | 0.5 | 0.00 | 0.6 | 0.6 | 0.00 |
| Malignant neoplasm of anorectum | 0.1 | 0.2 | 0.03 | 0.1 | 0.1 | 0.02 | 0.2 | 0.2 | 0.00 |
| Malignant neoplastic disease | 6.9 | 9.4 | 0.09 | 7.3 | 7.4 | 0.00 | 8.7 | 8.8 | 0.00 |
| Malignant tumor of breast | 1.3 | 1.5 | 0.02 | 1.3 | 1.4 | 0.01 | 1.4 | 1.6 | 0.01 |
| Malignant tumor of colon | 0.3 | 0.5 | 0.03 | 0.3 | 0.3 | -0.01 | 0.4 | 0.5 | 0.01 |
| Malignant tumor of lung | 0.3 | 0.5 | 0.04 | 0.3 | 0.3 | 0.00 | 0.4 | 0.4 | 0.00 |
| Malignant tumor of urinary bladder | 0.3 | 0.5 | 0.04 | 0.4 | 0.4 | 0.00 | 0.6 | 0.5 | -0.01 |
| Primary malignant neoplasm of prostate | 1.2 | 1.9 | 0.06 | 1.4 | 1.3 | 0.00 | 1.9 | 1.7 | -0.01 |
| **Medication use** | | | | | | | | | |

| (Continued) | Before ajustment | | | After matching | | | After stratification | | |
|---|---|---|---|---|---|---|---|---|---|
| Characteristic | T (%) | C (%) | StdDiff | T (%) | C (%) | StdDiff | T (%) | C (%) | StdDiff |
| Agents acting on the renin-angiotensin system | 68.7 | 73.7 | 0.11 | 70.3 | 70.5 | 0.00 | 72.1 | 72.6 | 0.01 |
| Antibacterials for systemic use | 63.4 | 62.5 | -0.02 | 62.3 | 62.4 | 0.00 | 62.8 | 62.6 | 0.00 |
| Antidepressants | 42.9 | 32.9 | -0.21 | 38.8 | 39.6 | 0.02 | 36.5 | 35.5 | -0.02 |
| Antiepileptics | 30.0 | 25.3 | -0.11 | 27.9 | 28.6 | 0.02 | 26.9 | 26.6 | -0.01 |
| Antiinflammatory and antirheumatic products | 43.4 | 40.2 | -0.06 | 41.9 | 42.2 | 0.01 | 41.4 | 41.0 | -0.01 |
| Antineoplastic agents | 7.2 | 6.3 | -0.03 | 6.5 | 6.6 | 0.01 | 6.6 | 6.5 | 0.00 |
| Antipsoriatics | 1.1 | 0.9 | -0.02 | 1.0 | 1.0 | 0.00 | 0.9 | 1.0 | 0.00 |
| Antithrombotic agents | 29.8 | 37.2 | 0.16 | 31.4 | 31.2 | 0.00 | 34.8 | 35.4 | 0.01 |
| Beta blocking agents | 47.1 | 53.3 | 0.12 | 48.4 | 48.4 | 0.00 | 51.2 | 51.8 | 0.01 |
| Calcium channel blockers | 30.4 | 34.0 | 0.08 | 31.3 | 31.2 | 0.00 | 32.1 | 33.1 | 0.02 |
| Diuretics | 49.1 | 47.8 | -0.03 | 47.8 | 47.9 | 0.00 | 47.8 | 48.1 | 0.01 |
| Drugs for acid related disorders | 43.3 | 43.7 | 0.01 | 42.5 | 42.6 | 0.00 | 44.0 | 43.7 | -0.01 |
| Drugs for obstructive airway diseases | 48.7 | 43.8 | -0.10 | 46.6 | 47.0 | 0.01 | 45.1 | 45.2 | 0.00 |
| Immunosuppressants | 4.4 | 3.7 | -0.03 | 4.0 | 4.1 | 0.00 | 3.9 | 3.9 | 0.00 |
| Lipid modifying agents | 76.1 | 79.9 | 0.09 | 77.6 | 77.8 | 0.00 | 78.6 | 79.2 | 0.02 |
| Opioids | 31.9 | 31.5 | -0.01 | 31.0 | 31.4 | 0.01 | 31.9 | 31.8 | 0.00 |
| Psycholeptics | 31.8 | 28.6 | -0.07 | 29.7 | 30.4 | 0.01 | 30.0 | 29.6 | -0.01 |
| Psychostimulants | 4.7 | 2.5 | -0.12 | 3.6 | 3.8 | 0.01 | 3.1 | 3.0 | 0.00 |

**Supplementary Table 32:** Baseline patient characteristics for GLP1-RAs (T) and DPP4is (C) new-users in the OptumEHR data source. We report proportion of initiators satisfying selected base-line characteristics and the standardized difference of population proportions (StdDiff) before and after propensity score adjustment. Less extreme StdDiffs through matching and stratification suggest improved balance between patient cohorts through adjustment.

| | Before adjustment | | | After matching | | | After stratification | | |
|---|---|---|---|---|---|---|---|---|---|
| Characteristic | T (%) | C (%) | StdDiff | T (%) | C (%) | StdDiff | T (%) | C (%) | StdDiff |
| **Age group** | | | | | | | | | |
| 25 - 29 | 0.4 | 0.1 | -0.06 | 0.4 | <0.1 | -0.05 | 0.3 | 0.1 | -0.04 |
| 30 - 34 | 0.6 | 0.3 | -0.05 | 0.6 | 0.4 | -0.02 | 0.5 | 0.3 | -0.03 |
| 35 - 39 | 2.0 | 1.0 | -0.08 | 2.0 | 1.5 | -0.04 | 1.4 | 1.1 | -0.02 |
| 40 - 44 | 4.8 | 2.2 | -0.15 | 3.9 | 3.8 | 0.00 | 2.6 | 2.5 | 0.00 |
| 45 - 49 | 8.3 | 5.1 | -0.13 | 7.3 | 7.2 | 0.00 | 5.7 | 5.5 | -0.01 |
| 50 - 54 | 14.3 | 8.9 | -0.17 | 13.2 | 13.6 | 0.01 | 10.1 | 10.1 | 0.00 |
| 55 - 59 | 17.6 | 13.5 | -0.11 | 17.3 | 16.8 | -0.01 | 14.5 | 14.4 | 0.00 |
| 60 - 64 | 19.8 | 16.4 | -0.09 | 19.7 | 20.6 | 0.02 | 17.4 | 17.4 | 0.00 |
| 65 - 69 | 15.2 | 16.7 | 0.04 | 16.1 | 16.4 | 0.01 | 16.9 | 16.5 | -0.01 |
| 70 - 74 | 9.7 | 14.3 | 0.14 | 10.9 | 10.8 | 0.00 | 13.6 | 13.4 | 0.00 |
| 75 - 79 | 5.0 | 11.6 | 0.24 | 6.0 | 5.4 | -0.02 | 9.7 | 10.2 | 0.02 |
| 80 - 84 | 1.6 | 7.6 | 0.29 | 1.9 | 2.0 | 0.01 | 5.7 | 6.4 | 0.03 |
| 85 - 89 | 0.4 | 2.3 | 0.16 | 0.5 | 1.1 | 0.06 | 1.6 | 2.0 | 0.03 |
| Gender: female | 50.2 | 43.2 | -0.14 | 47.6 | 47.5 | 0.00 | 44.5 | 44.5 | 0.00 |
| **Race** | | | | | | | | | |
| Asian | 1.1 | 1.9 | 0.07 | 1.3 | 1.5 | 0.02 | 1.2 | 1.9 | 0.06 |
| Black or African American | 10.1 | 9.5 | -0.02 | 10.1 | 9.4 | -0.02 | 10.0 | 9.6 | -0.01 |
| White | 83.6 | 82.3 | -0.03 | 83.4 | 82.5 | -0.02 | 83.5 | 82.5 | -0.03 |
| **Ethnicity** | | | | | | | | | |
| Hispanic or Latino | 4.0 | 4.8 | 0.04 | 4.0 | 5.0 | 0.05 | 3.9 | 4.7 | 0.04 |
| Not Hispanic or Latino | 88.3 | 87.8 | -0.01 | 88.0 | 86.9 | -0.03 | 87.7 | 87.9 | 0.01 |
| **Medical history: General** | | | | | | | | | |
| Acute respiratory disease | 21.3 | 20.0 | -0.03 | 20.1 | 21.1 | 0.02 | 18.6 | 20.3 | 0.04 |
| Attention deficit hyperactivity disorder | 1.0 | 0.4 | -0.08 | 0.9 | 0.7 | -0.02 | 0.6 | 0.5 | -0.01 |
| Chronic liver disease | 2.7 | 3.1 | 0.02 | 2.8 | 2.3 | -0.03 | 3.3 | 2.9 | -0.02 |
| Chronic obstructive lung disease | 10.2 | 11.5 | 0.04 | 10.6 | 10.3 | -0.01 | 12.0 | 11.4 | -0.02 |
| Crohn’s disease | 0.4 | 0.3 | -0.02 | 0.4 | 0.3 | -0.01 | 0.3 | 0.3 | 0.00 |
| Dementia | 0.9 | 2.1 | 0.10 | 0.9 | 0.9 | 0.00 | 0.9 | 1.8 | 0.08 |
| Depressive disorder | 21.4 | 15.0 | -0.17 | 19.1 | 19.8 | 0.02 | 16.3 | 16.2 | 0.00 |
| Gastroesophageal reflux disease | 24.8 | 21.8 | -0.07 | 22.7 | 24.3 | 0.04 | 22.9 | 22.3 | -0.01 |
| Gastrointestinal hemorrhage | 2.0 | 2.2 | 0.01 | 2.1 | 1.9 | -0.02 | 3.0 | 2.2 | -0.06 |
| Human immunodeficiency virus infection | 0.2 | 0.2 | -0.01 | 0.2 | 0.3 | 0.02 | 0.1 | 0.2 | 0.03 |
| Hyperlipidemia | 74.6 | 77.3 | 0.06 | 74.4 | 74.5 | 0.00 | 78.2 | 76.7 | -0.04 |
| Hypertensive disorder | 77.5 | 79.2 | 0.04 | 76.7 | 78.0 | 0.03 | 80.2 | 78.9 | -0.03 |
| Lesion of liver | 1.1 | 1.0 | 0.00 | 1.0 | 1.0 | 0.00 | 0.8 | 1.0 | 0.03 |
| Obesity | 47.5 | 25.8 | -0.46 | 39.6 | 42.7 | 0.06 | 30.0 | 30.2 | 0.00 |
| Osteoarthritis | 25.3 | 22.6 | -0.06 | 23.4 | 23.1 | -0.01 | 22.5 | 23.1 | 0.01 |
| Pneumonia | 3.0 | 3.1 | 0.01 | 2.9 | 2.9 | 0.00 | 3.0 | 3.1 | 0.00 |
| Psoriasis | 2.3 | 1.5 | -0.06 | 2.0 | 1.9 | 0.00 | 1.8 | 1.6 | -0.02 |
| Renal impairment | 9.1 | 12.6 | 0.12 | 9.2 | 9.9 | 0.02 | 11.2 | 12.0 | 0.02 |
| Rheumatoid arthritis | 1.8 | 1.5 | -0.02 | 1.7 | 1.8 | 0.00 | 1.6 | 1.6 | 0.00 |
| Schizophrenia | 0.3 | 0.3 | 0.00 | 0.3 | 0.2 | -0.01 | 0.3 | 0.3 | 0.00 |
| Ulcerative colitis | 0.4 | 0.3 | -0.01 | 0.4 | 0.4 | -0.01 | 0.2 | 0.3 | 0.02 |
| Urinary tract infectious disease | 5.2 | 6.9 | 0.07 | 5.2 | 5.5 | 0.02 | 5.8 | 6.5 | 0.03 |
| Viral hepatitis C | 0.8 | 0.6 | -0.02 | 0.9 | 0.5 | -0.05 | 0.6 | 0.6 | 0.00 |
| Visual system disorder | 15.0 | 15.2 | 0.01 | 14.0 | 14.6 | 0.02 | 13.0 | 15.3 | 0.07 |
| **Medical history: Cardiovascular disease** | | | | | | | | | |
| Atrial fibrillation | 7.5 | 10.7 | 0.11 | 7.8 | 7.8 | 0.00 | 9.9 | 10.1 | 0.01 |
| Cerebrovascular disease | 8.6 | 11.4 | 0.09 | 8.8 | 9.1 | 0.01 | 10.4 | 10.9 | 0.02 |
| Coronary arteriosclerosis | 34.5 | 42.6 | 0.17 | 36.0 | 34.1 | -0.04 | 40.9 | 40.8 | 0.00 |
| Heart disease | 52.6 | 59.5 | 0.14 | 53.8 | 51.3 | -0.05 | 58.8 | 57.7 | -0.02 |
| Heart failure | 7.8 | 9.2 | 0.05 | 7.9 | 7.8 | -0.01 | 9.5 | 8.9 | -0.02 |
| Ischemic heart disease | 17.8 | 19.2 | 0.04 | 18.0 | 17.2 | -0.02 | 19.1 | 18.7 | -0.01 |
| Peripheral vascular disease | 20.9 | 17.7 | -0.08 | 18.9 | 21.2 | 0.06 | 16.7 | 18.4 | 0.04 |
| Pulmonary embolism | 1.1 | 0.9 | -0.02 | 1.0 | 0.9 | -0.01 | 1.0 | 0.9 | -0.01 |
| Venous thrombosis | 1.8 | 1.9 | 0.00 | 1.7 | 1.8 | 0.01 | 1.9 | 1.9 | 0.00 |
| **Medical history: Neoplasms** | | | | | | | | | |
| Hematologic neoplasm | 0.6 | 1.2 | 0.06 | 0.7 | 1.0 | 0.04 | 1.0 | 1.1 | 0.01 |
| Malignant lymphoma | 0.5 | 0.7 | 0.02 | 0.6 | 0.4 | -0.03 | 0.4 | 0.7 | 0.03 |
| Malignant neoplasm of anorectum | 0.2 | 0.1 | 0.00 | 0.1 | <0.1 | -0.01 | 0.3 | 0.1 | -0.04 |
| Malignant neoplastic disease | 7.2 | 9.5 | 0.08 | 7.3 | 7.4 | 0.00 | 9.1 | 9.0 | 0.00 |

| (Continued) | Before ajustment | | | After matching | | | After stratification | | |
|---|---|---|---|---|---|---|---|---|---|
| Characteristic | T (%) | C (%) | StdDiff | T (%) | C (%) | StdDiff | T (%) | C (%) | StdDiff |
| Malignant tumor of breast | 1.4 | 1.5 | 0.01 | 1.3 | 1.2 | -0.01 | 1.2 | 1.5 | 0.02 |
| Malignant tumor of colon | 0.2 | 0.4 | 0.04 | 0.2 | 0.3 | 0.02 | 0.1 | 0.4 | 0.05 |
| Malignant tumor of lung | 0.1 | 0.5 | 0.06 | 0.2 | 0.4 | 0.05 | 0.1 | 0.4 | 0.06 |
| Malignant tumor of urinary bladder | 0.3 | 0.5 | 0.03 | 0.4 | 0.5 | 0.01 | 0.4 | 0.5 | 0.01 |
| Primary malignant neoplasm of prostate | 1.0 | 2.0 | 0.08 | 1.1 | 1.1 | 0.00 | 2.2 | 1.8 | -0.03 |
| **Medication use** | | | | | | | | | |
| Agents acting on the renin-angiotensin system | 71.3 | 73.7 | 0.06 | 72.1 | 72.5 | 0.01 | 73.7 | 73.4 | -0.01 |
| Antibacterials for systemic use | 50.3 | 47.0 | -0.07 | 48.0 | 47.8 | 0.00 | 47.0 | 47.5 | 0.01 |
| Antidepressants | 42.5 | 32.8 | -0.20 | 39.3 | 40.5 | 0.03 | 34.6 | 34.8 | 0.00 |
| Antiepileptics | 26.5 | 22.1 | -0.10 | 24.7 | 25.7 | 0.02 | 22.3 | 23.0 | 0.02 |
| Antiinflammatory and antirheumatic products | 70.5 | 69.1 | -0.03 | 70.0 | 70.2 | 0.00 | 69.6 | 69.4 | 0.00 |
| Antineoplastic agents | 6.1 | 5.2 | -0.04 | 5.6 | 5.8 | 0.01 | 5.6 | 5.8 | 0.01 |
| Antipsoriatics | 0.8 | 0.8 | -0.01 | 0.7 | 0.9 | 0.02 | 0.7 | 0.8 | 0.01 |
| Antithrombotic agents | 61.2 | 67.1 | 0.12 | 62.9 | 62.4 | -0.01 | 66.3 | 65.9 | -0.01 |
| Beta blocking agents | 51.7 | 57.5 | 0.12 | 52.8 | 52.0 | -0.02 | 57.8 | 56.3 | -0.03 |
| Calcium channel blockers | 29.3 | 31.7 | 0.05 | 29.3 | 30.1 | 0.02 | 31.7 | 31.2 | -0.01 |
| Diuretics | 50.4 | 47.8 | -0.05 | 48.7 | 48.9 | 0.00 | 50.2 | 48.2 | -0.04 |
| Drugs for acid related disorders | 49.0 | 48.1 | -0.02 | 47.6 | 48.6 | 0.02 | 48.6 | 48.3 | -0.01 |
| Drugs for obstructive airway diseases | 54.1 | 47.3 | -0.14 | 51.3 | 51.9 | 0.01 | 46.7 | 48.8 | 0.04 |
| Immunosuppressants | 4.4 | 2.9 | -0.08 | 4.0 | 3.5 | -0.02 | 3.7 | 3.0 | -0.04 |
| Lipid modifying agents | 81.5 | 83.6 | 0.06 | 82.7 | 81.7 | -0.03 | 84.2 | 83.4 | -0.02 |
| Opioids | 36.6 | 34.8 | -0.04 | 35.4 | 34.0 | -0.03 | 36.0 | 35.0 | -0.02 |
| Psycholeptics | 39.4 | 35.6 | -0.08 | 37.4 | 37.0 | -0.01 | 35.4 | 36.4 | 0.02 |
| Psychostimulants | 5.1 | 2.5 | -0.14 | 3.1 | 2.9 | -0.01 | 3.1 | 2.9 | -0.01 |

**Supplementary Table 33:** Baseline patient characteristics for GLP1-RAs (T) and SUs (C) new-users in the CCAE data source. We report proportion of initiators satisfying selected base-line characteristics and the standardized difference of population proportions (StdDiff) before and after propensity score adjustment. Less extreme StdDiffs through matching and stratification suggest improved balance between patient cohorts through adjustment.

| | Before adjustment | | | After matching | | | After stratification | | |
|---|---|---|---|---|---|---|---|---|---|
| Characteristic | T (%) | C (%) | StdDiff | T (%) | C (%) | StdDiff | T (%) | C (%) | StdDiff |
| **Age group** | | | | | | | | | |
| 25 - 29 | 0.2 | 0.1 | 0.02 | 0.2 | 0.2 | 0.01 | 0.3 | 0.1 | 0.03 |
| 30 - 34 | 0.9 | 0.4 | 0.06 | 0.9 | 0.6 | 0.03 | 0.8 | 0.5 | 0.04 |
| 35 - 39 | 2.6 | 1.6 | 0.07 | 2.2 | 2.2 | 0.00 | 2.3 | 1.7 | 0.04 |
| 40 - 44 | 6.3 | 4.2 | 0.09 | 5.8 | 5.7 | 0.00 | 5.1 | 4.5 | 0.03 |
| 45 - 49 | 11.5 | 9.2 | 0.08 | 11.1 | 11.1 | 0.00 | 9.5 | 9.7 | -0.01 |
| 50 - 54 | 20.2 | 17.7 | 0.06 | 19.7 | 19.5 | 0.01 | 19.2 | 18.1 | 0.03 |
| 55 - 59 | 27.0 | 27.3 | -0.01 | 27.2 | 27.1 | 0.00 | 27.3 | 27.1 | 0.00 |
| 60 - 64 | 28.6 | 35.6 | -0.15 | 30.0 | 30.8 | -0.02 | 32.1 | 34.7 | -0.05 |
| 65 - 69 | 2.4 | 3.8 | -0.08 | 2.7 | 2.8 | 0.00 | 3.3 | 3.6 | -0.02 |
| Gender: female | 53.8 | 39.2 | 0.30 | 50.3 | 51.4 | -0.02 | 40.6 | 41.6 | -0.02 |
| **Medical history: General** | | | | | | | | | |
| Acute respiratory disease | 33.5 | 28.4 | 0.11 | 32.3 | 32.5 | 0.00 | 28.8 | 29.5 | -0.02 |
| Attention deficit hyperactivity disorder | 1.5 | 0.5 | 0.10 | 1.2 | 1.1 | 0.01 | 0.7 | 0.7 | 0.00 |
| Chronic liver disease | 2.6 | 3.9 | -0.07 | 3.0 | 2.9 | 0.01 | 4.6 | 3.8 | 0.04 |
| Chronic obstructive lung disease | 5.5 | 7.0 | -0.06 | 5.5 | 5.8 | -0.01 | 6.8 | 6.8 | 0.00 |
| Crohn's disease | 0.4 | 0.3 | 0.02 | 0.4 | 0.3 | 0.02 | 0.7 | 0.3 | 0.05 |
| Dementia | 0.2 | 0.4 | -0.02 | 0.2 | 0.3 | -0.01 | 0.3 | 0.4 | -0.01 |
| Depressive disorder | 15.7 | 10.6 | 0.15 | 14.0 | 15.0 | -0.03 | 12.3 | 11.8 | 0.01 |
| Gastroesophageal reflux disease | 18.2 | 12.8 | 0.15 | 16.3 | 17.1 | -0.02 | 13.6 | 14.1 | -0.01 |
| Gastrointestinal hemorrhage | 2.8 | 2.7 | 0.00 | 2.9 | 2.7 | 0.01 | 3.2 | 2.7 | 0.03 |
| Human immunodeficiency virus infection | 0.3 | 0.3 | 0.00 | 0.3 | 0.4 | -0.03 | 0.2 | 0.3 | -0.01 |
| Hyperlipidemia | 74.9 | 68.9 | 0.13 | 74.0 | 74.6 | -0.01 | 71.4 | 71.3 | 0.00 |
| Hypertensive disorder | 76.8 | 73.1 | 0.09 | 76.9 | 77.4 | -0.01 | 76.9 | 75.1 | 0.04 |
| Lesion of liver | 0.9 | 1.2 | -0.03 | 0.7 | 1.1 | -0.04 | 1.0 | 1.2 | -0.02 |
| Obesity | 38.0 | 17.5 | 0.47 | 31.8 | 34.2 | -0.05 | 22.0 | 21.9 | 0.00 |
| Osteoarthritis | 29.5 | 21.8 | 0.18 | 27.9 | 28.4 | -0.01 | 22.9 | 23.6 | -0.02 |
| Pneumonia | 3.3 | 4.0 | -0.03 | 3.2 | 3.3 | -0.01 | 3.8 | 3.9 | 0.00 |
| Psoriasis | 2.7 | 1.5 | 0.08 | 2.2 | 2.2 | 0.00 | 1.6 | 1.7 | -0.01 |
| Renal impairment | 5.6 | 6.3 | -0.03 | 5.7 | 5.4 | 0.01 | 5.6 | 6.4 | -0.03 |
| Rheumatoid arthritis | 2.0 | 1.4 | 0.04 | 1.8 | 1.9 | -0.01 | 1.7 | 1.5 | 0.01 |
| Schizophrenia | <0.1 | 0.1 | -0.04 | <0.1 | 0.2 | -0.05 | <0.1 | 0.2 | -0.05 |
| Ulcerative colitis | 0.5 | 0.4 | 0.01 | 0.5 | 0.4 | 0.01 | 0.6 | 0.4 | 0.03 |
| Urinary tract infectious disease | 9.2 | 8.5 | 0.03 | 9.0 | 9.0 | 0.00 | 8.4 | 8.6 | -0.01 |
| Viral hepatitis C | 0.3 | 0.6 | -0.05 | 0.3 | 0.4 | -0.01 | 0.3 | 0.6 | -0.04 |
| Visual system disorder | 30.2 | 25.6 | 0.10 | 29.3 | 29.2 | 0.00 | 25.5 | 26.5 | -0.02 |
| **Medical history: Cardiovascular disease** | | | | | | | | | |
| Atrial fibrillation | 4.8 | 5.0 | -0.01 | 4.9 | 4.9 | 0.00 | 5.3 | 5.1 | 0.01 |
| Cerebrovascular disease | 8.6 | 11.0 | -0.08 | 8.7 | 8.5 | 0.00 | 9.4 | 10.3 | -0.03 |
| Coronary arteriosclerosis | 28.0 | 33.8 | -0.13 | 29.0 | 28.9 | 0.00 | 32.8 | 32.3 | 0.01 |
| Heart disease | 47.0 | 51.1 | -0.08 | 47.0 | 46.8 | 0.01 | 50.4 | 49.4 | 0.02 |
| Heart failure | 5.7 | 7.0 | -0.05 | 5.5 | 5.9 | -0.01 | 6.4 | 6.7 | -0.01 |
| Ischemic heart disease | 16.1 | 19.6 | -0.09 | 16.1 | 16.1 | 0.00 | 19.5 | 18.2 | 0.04 |
| Peripheral vascular disease | 16.1 | 12.5 | 0.10 | 15.6 | 15.3 | 0.01 | 13.0 | 13.2 | -0.01 |
| Pulmonary embolism | 1.4 | 0.9 | 0.04 | 1.3 | 1.1 | 0.01 | 1.0 | 1.0 | 0.00 |
| Venous thrombosis | 2.0 | 1.8 | 0.01 | 2.0 | 1.7 | 0.02 | 1.6 | 1.8 | -0.02 |
| **Medical history: Neoplasms** | | | | | | | | | |
| Hematologic neoplasm | 1.0 | 1.3 | -0.03 | 0.9 | 1.0 | -0.02 | 0.7 | 1.2 | -0.05 |
| Malignant lymphoma | 0.5 | 0.6 | -0.01 | 0.5 | 0.5 | 0.00 | 0.3 | 0.6 | -0.03 |
| Malignant neoplasm of anorectum | 0.1 | 0.2 | -0.04 | <0.1 | 0.1 | -0.02 | 0.2 | 0.2 | -0.01 |
| Malignant neoplastic disease | 7.7 | 8.1 | -0.01 | 7.4 | 7.9 | -0.02 | 7.9 | 8.0 | 0.00 |
| Malignant tumor of breast | 1.5 | 1.4 | 0.01 | 1.4 | 1.5 | -0.01 | 1.1 | 1.4 | -0.02 |
| Malignant tumor of colon | 0.2 | 0.5 | -0.04 | 0.3 | 0.3 | -0.01 | 0.4 | 0.5 | -0.02 |
| Malignant tumor of lung | 0.1 | 0.4 | -0.06 | 0.1 | 0.2 | -0.02 | 0.2 | 0.4 | -0.03 |
| Malignant tumor of urinary bladder | 0.2 | 0.4 | -0.03 | 0.2 | 0.2 | 0.00 | 0.4 | 0.3 | 0.01 |
| Primary malignant neoplasm of prostate | 0.8 | 1.1 | -0.04 | 0.9 | 0.9 | 0.00 | 1.6 | 1.1 | 0.04 |
| **Medication use** | | | | | | | | | |
| Agents acting on the renin-angiotensin system | 70.6 | 73.2 | -0.06 | 72.7 | 71.3 | 0.03 | 74.2 | 73.3 | 0.02 |
| Antibacterials for systemic use | 67.5 | 62.8 | 0.10 | 66.6 | 66.4 | 0.00 | 64.7 | 63.6 | 0.02 |
| Antidepressants | 41.0 | 30.2 | 0.23 | 38.3 | 39.1 | -0.01 | 33.7 | 32.0 | 0.04 |
| Antiepileptics | 21.9 | 16.5 | 0.14 | 20.8 | 21.0 | 0.00 | 18.1 | 17.7 | 0.01 |
| Antiinflammatory and antirheumatic products | 42.3 | 34.5 | 0.16 | 40.6 | 41.2 | -0.01 | 36.3 | 36.2 | 0.00 |

| (Continued) | Before ajustment | | | After matching | | | After stratification | | |
|---|---|---|---|---|---|---|---|---|---|
| Characteristic | T (%) | C (%) | StdDiff | T (%) | C (%) | StdDiff | T (%) | C (%) | StdDiff |
| Antineoplastic agents | 6.8 | 5.5 | 0.05 | 6.3 | 6.5 | -0.01 | 5.3 | 5.4 | -0.01 |
| Antipsoriatics | 1.0 | 0.7 | 0.03 | 0.8 | 0.9 | -0.01 | 0.5 | 0.7 | -0.02 |
| Antithrombotic agents | 23.8 | 27.7 | -0.09 | 24.5 | 24.2 | 0.01 | 27.0 | 27.2 | 0.00 |
| Beta blocking agents | 43.4 | 49.1 | -0.12 | 44.7 | 44.4 | 0.01 | 48.4 | 48.2 | 0.00 |
| Calcium channel blockers | 26.5 | 27.1 | -0.01 | 26.3 | 26.8 | -0.01 | 27.8 | 27.2 | 0.01 |
| Diuretics | 49.4 | 44.9 | 0.09 | 48.9 | 48.7 | 0.00 | 45.8 | 45.6 | 0.00 |
| Drugs for acid related disorders | 35.8 | 31.3 | 0.09 | 34.5 | 34.6 | 0.00 | 31.0 | 31.8 | -0.02 |
| Drugs for obstructive airway diseases | 48.1 | 38.3 | 0.20 | 45.4 | 46.1 | -0.01 | 42.3 | 40.5 | 0.04 |
| Immunosuppressants | 4.5 | 2.5 | 0.11 | 3.7 | 3.6 | 0.00 | 2.7 | 2.8 | -0.01 |
| Lipid modifying agents | 75.1 | 76.7 | -0.04 | 76.3 | 75.8 | 0.01 | 77.9 | 77.0 | 0.02 |
| Opioids | 35.2 | 33.9 | 0.03 | 34.9 | 35.1 | 0.00 | 34.5 | 34.5 | 0.00 |
| Psycholeptics | 34.3 | 28.9 | 0.11 | 33.0 | 32.9 | 0.00 | 28.8 | 30.1 | -0.03 |
| Psychostimulants | 5.9 | 2.8 | 0.15 | 3.3 | 3.3 | 0.00 | 3.3 | 3.3 | 0.00 |

**Supplementary Table 34:** Baseline patient characteristics for GLP1-RAs (T) and SUs (C) new-users in the Optum-DOD data source. We report proportion of initiators satisfying selected base-line characteristics and the standardized difference of population proportions (StdDiff) before and after propensity score adjustment. Less extreme StdDiffs through matching and stratification suggest improved balance between patient cohorts through adjustment.

| | Before adjustment | | | After matching | | | After stratification | | |
|---|---|---|---|---|---|---|---|---|---|
| Characteristic | T (%) | C (%) | StdDiff | T (%) | C (%) | StdDiff | T (%) | C (%) | StdDiff |
| **Age group** | | | | | | | | | |
| 25 - 29 | 0.1 | 0.1 | 0.02 | 0.1 | 0.1 | 0.00 | 0.4 | 0.1 | 0.07 |
| 30 - 34 | 0.5 | 0.2 | 0.06 | 0.5 | 0.3 | 0.04 | 0.8 | 0.2 | 0.08 |
| 35 - 39 | 1.6 | 0.6 | 0.10 | 1.5 | 1.3 | 0.02 | 0.8 | 0.7 | 0.02 |
| 40 - 44 | 3.4 | 1.3 | 0.13 | 3.0 | 2.8 | 0.02 | 1.5 | 1.5 | 0.00 |
| 45 - 49 | 6.4 | 3.0 | 0.16 | 5.8 | 5.8 | 0.00 | 3.3 | 3.3 | 0.00 |
| 50 - 54 | 11.4 | 5.6 | 0.21 | 9.8 | 10.1 | -0.01 | 5.4 | 6.1 | -0.03 |
| 55 - 59 | 14.9 | 8.9 | 0.19 | 14.0 | 14.3 | -0.01 | 8.3 | 9.5 | -0.04 |
| 60 - 64 | 16.7 | 11.7 | 0.14 | 16.4 | 16.5 | 0.00 | 12.0 | 12.3 | -0.01 |
| 65 - 69 | 17.8 | 17.8 | 0.00 | 18.7 | 18.8 | 0.00 | 18.6 | 17.9 | 0.02 |
| 70 - 74 | 15.1 | 20.9 | -0.15 | 16.4 | 17.1 | -0.02 | 18.9 | 20.4 | -0.04 |
| 75 - 79 | 8.3 | 15.6 | -0.23 | 9.4 | 8.9 | 0.02 | 17.3 | 14.8 | 0.07 |
| 80 - 84 | 2.7 | 9.5 | -0.29 | 3.1 | 2.8 | 0.02 | 7.5 | 8.8 | -0.04 |
| 85 - 89 | 0.8 | 4.6 | -0.24 | 0.9 | 0.9 | 0.00 | 5.0 | 4.2 | 0.04 |
| 90 - 94 | <0.1 | 0.3 | -0.06 | <0.1 | 0.3 | -0.05 | 0.1 | 0.3 | -0.06 |
| Gender: female | 52.9 | 44.2 | 0.17 | 50.5 | 51.3 | -0.01 | 43.4 | 45.0 | -0.03 |
| **Medical history: General** | | | | | | | | | |
| Acute respiratory disease | 30.1 | 28.1 | 0.04 | 29.2 | 29.8 | -0.01 | 31.4 | 28.3 | 0.07 |
| Attention deficit hyperactivity disorder | 1.3 | 0.4 | 0.09 | 0.9 | 1.2 | -0.03 | 0.5 | 0.5 | 0.00 |
| Chronic liver disease | 2.7 | 3.7 | -0.05 | 2.9 | 2.7 | 0.01 | 3.1 | 3.6 | -0.03 |
| Chronic obstructive lung disease | 14.5 | 17.2 | -0.07 | 14.6 | 15.1 | -0.02 | 19.0 | 17.0 | 0.05 |
| Crohn’s disease | 0.3 | 0.3 | 0.00 | 0.2 | 0.3 | -0.02 | 0.5 | 0.3 | 0.03 |
| Dementia | 1.8 | 4.4 | -0.15 | 1.9 | 2.0 | -0.01 | 4.0 | 4.1 | -0.01 |
| Depressive disorder | 24.4 | 17.1 | 0.18 | 22.2 | 23.9 | -0.04 | 17.9 | 18.0 | 0.00 |
| Gastroesophageal reflux disease | 26.6 | 22.2 | 0.10 | 25.2 | 26.5 | -0.03 | 24.3 | 22.8 | 0.04 |
| Gastrointestinal hemorrhage | 2.8 | 4.0 | -0.06 | 2.8 | 3.0 | -0.01 | 4.2 | 3.9 | 0.02 |
| Human immunodeficiency virus infection | 0.6 | 0.2 | 0.06 | 0.5 | 0.3 | 0.02 | 0.3 | 0.2 | 0.01 |
| Hyperlipidemia | 85.2 | 85.0 | 0.01 | 84.9 | 85.3 | -0.01 | 87.4 | 85.0 | 0.07 |
| Hypertensive disorder | 86.0 | 87.4 | -0.04 | 86.6 | 86.4 | 0.00 | 89.4 | 87.3 | 0.06 |
| Lesion of liver | 1.5 | 1.9 | -0.03 | 1.5 | 1.8 | -0.02 | 1.3 | 1.9 | -0.05 |
| Obesity | 47.9 | 23.4 | 0.53 | 42.7 | 45.6 | -0.06 | 25.9 | 26.0 | 0.00 |
| Osteoarthritis | 40.2 | 34.4 | 0.12 | 39.0 | 39.6 | -0.01 | 37.2 | 35.0 | 0.05 |
| Pneumonia | 5.4 | 7.2 | -0.07 | 5.0 | 5.3 | -0.02 | 10.0 | 7.0 | 0.11 |
| Psoriasis | 2.7 | 1.6 | 0.08 | 2.6 | 2.3 | 0.02 | 1.4 | 1.7 | -0.02 |
| Renal impairment | 16.0 | 24.9 | -0.22 | 17.0 | 16.7 | 0.01 | 29.6 | 24.0 | 0.13 |
| Rheumatoid arthritis | 2.8 | 2.5 | 0.02 | 2.4 | 2.8 | -0.02 | 2.6 | 2.5 | 0.01 |
| Schizophrenia | 0.6 | 0.5 | 0.02 | 0.6 | 0.5 | 0.01 | 0.4 | 0.5 | -0.01 |
| Ulcerative colitis | 0.4 | 0.4 | 0.00 | 0.4 | 0.3 | 0.01 | 0.7 | 0.4 | 0.04 |
| Urinary tract infectious disease | 11.7 | 14.7 | -0.09 | 11.5 | 11.2 | 0.01 | 19.8 | 14.3 | 0.14 |
| Viral hepatitis C | 0.7 | 0.8 | -0.01 | 0.7 | 0.7 | 0.00 | 0.7 | 0.8 | 0.00 |
| Visual system disorder | 42.8 | 47.2 | -0.09 | 43.7 | 43.8 | 0.00 | 47.6 | 46.8 | 0.01 |
| **Medical history: Cardiovascular disease** | | | | | | | | | |
| Atrial fibrillation | 9.5 | 13.1 | -0.11 | 9.6 | 9.5 | 0.01 | 14.1 | 12.7 | 0.04 |
| Cerebrovascular disease | 11.3 | 16.3 | -0.15 | 11.7 | 11.4 | 0.01 | 16.6 | 15.8 | 0.02 |
| Coronary arteriosclerosis | 33.8 | 40.0 | -0.13 | 35.1 | 34.8 | 0.01 | 41.3 | 39.4 | 0.04 |
| Heart disease | 54.4 | 60.6 | -0.12 | 54.9 | 54.2 | 0.01 | 62.6 | 59.9 | 0.06 |
| Heart failure | 12.0 | 16.0 | -0.11 | 12.1 | 12.1 | 0.00 | 18.6 | 15.6 | 0.08 |
| Ischemic heart disease | 18.5 | 22.2 | -0.09 | 18.5 | 18.8 | -0.01 | 24.3 | 21.8 | 0.06 |
| Peripheral vascular disease | 26.7 | 26.1 | 0.01 | 27.1 | 27.6 | -0.01 | 27.6 | 26.3 | 0.03 |
| Pulmonary embolism | 1.7 | 1.4 | 0.02 | 1.6 | 1.4 | 0.01 | 2.0 | 1.4 | 0.04 |
| Venous thrombosis | 2.3 | 2.8 | -0.03 | 2.3 | 2.4 | 0.00 | 2.5 | 2.8 | -0.01 |
| **Medical history: Neoplasms** | | | | | | | | | |
| Hematologic neoplasm | 1.2 | 2.0 | -0.06 | 1.1 | 1.3 | -0.02 | 1.5 | 1.9 | -0.03 |
| Malignant lymphoma | 0.7 | 0.9 | -0.02 | 0.6 | 0.7 | -0.01 | 0.5 | 0.8 | -0.04 |
| Malignant neoplasm of anorectum | 0.2 | 0.4 | -0.03 | 0.2 | 0.2 | -0.01 | 0.2 | 0.4 | -0.03 |
| Malignant neoplastic disease | 10.9 | 15.1 | -0.12 | 11.1 | 11.1 | 0.00 | 15.1 | 14.6 | 0.01 |
| Malignant tumor of breast | 1.8 | 2.1 | -0.02 | 1.9 | 2.0 | -0.01 | 1.8 | 2.1 | -0.03 |
| Malignant tumor of colon | 0.7 | 0.9 | -0.02 | 0.8 | 0.6 | 0.02 | 1.1 | 0.9 | 0.02 |
| Malignant tumor of lung | 0.3 | 0.9 | -0.08 | 0.3 | 0.4 | 0.00 | 1.7 | 0.9 | 0.08 |
| Malignant tumor of urinary bladder | 0.5 | 0.9 | -0.06 | 0.5 | 0.5 | 0.00 | 0.9 | 0.9 | 0.00 |
| Primary malignant neoplasm of prostate | 1.8 | 3.1 | -0.09 | 2.0 | 1.6 | 0.03 | 4.3 | 3.0 | 0.07 |
| **Medication use** | | | | | | | | | |

| (Continued) | Before ajustment | | | After matching | | | After stratification | | |
|---|---|---|---|---|---|---|---|---|---|
| Characteristic | T (%) | C (%) | StdDiff | T (%) | C (%) | StdDiff | T (%) | C (%) | StdDiff |
| Agents acting on the renin-angiotensin system | 72.7 | 75.8 | -0.07 | 74.2 | 73.6 | 0.01 | 78.6 | 75.5 | 0.07 |
| Antibacterials for systemic use | 64.7 | 62.4 | 0.05 | 63.4 | 63.5 | 0.00 | 63.2 | 62.6 | 0.01 |
| Antidepressants | 43.0 | 30.3 | 0.26 | 40.0 | 41.8 | -0.04 | 32.4 | 31.7 | 0.01 |
| Antiepileptics | 29.3 | 21.9 | 0.17 | 27.5 | 28.7 | -0.03 | 23.7 | 22.8 | 0.02 |
| Antiinflammatory and antirheumatic products | 39.8 | 30.4 | 0.20 | 37.7 | 38.9 | -0.03 | 28.2 | 31.4 | -0.07 |
| Antineoplastic agents | 7.4 | 6.6 | 0.03 | 7.0 | 6.6 | 0.02 | 5.8 | 6.6 | -0.03 |
| Antipsoriatics | 0.9 | 0.7 | 0.03 | 0.8 | 0.9 | -0.01 | 0.6 | 0.7 | -0.01 |
| Antithrombotic agents | 27.4 | 32.2 | -0.10 | 28.0 | 28.4 | -0.01 | 34.3 | 31.8 | 0.05 |
| Beta blocking agents | 47.4 | 54.6 | -0.14 | 48.7 | 48.1 | 0.01 | 55.6 | 53.9 | 0.04 |
| Calcium channel blockers | 30.1 | 34.2 | -0.09 | 31.1 | 30.8 | 0.01 | 35.3 | 33.8 | 0.03 |
| Diuretics | 51.4 | 50.8 | 0.01 | 50.7 | 51.3 | -0.01 | 54.4 | 50.8 | 0.07 |
| Drugs for acid related disorders | 37.0 | 35.0 | 0.04 | 35.8 | 36.7 | -0.02 | 40.7 | 35.2 | 0.11 |
| Drugs for obstructive airway diseases | 50.7 | 41.5 | 0.18 | 48.2 | 49.8 | -0.03 | 46.0 | 42.5 | 0.07 |
| Immunosuppressants | 4.1 | 2.8 | 0.07 | 3.6 | 3.4 | 0.01 | 3.6 | 2.9 | 0.04 |
| Lipid modifying agents | 79.9 | 81.1 | -0.03 | 80.8 | 81.1 | -0.01 | 81.6 | 81.1 | 0.01 |
| Opioids | 36.3 | 33.5 | 0.06 | 34.9 | 36.0 | -0.02 | 31.9 | 33.9 | -0.04 |
| Psycholeptics | 34.1 | 28.8 | 0.11 | 32.3 | 33.2 | -0.02 | 27.1 | 29.3 | -0.05 |
| Psychostimulants | 3.8 | 1.4 | 0.15 | 3.0 | 3.0 | 0.00 | 1.9 | 1.6 | 0.02 |

**Supplementary Table 35:** Baseline patient characteristics for GLP1-RAs (T) and SUs (C) new-users in the MDCR data source. We report proportion of initiators satisfying selected base-line characteristics and the standardized difference of population proportions (StdDiff) before and after propensity score adjustment. Less extreme StdDiffs through matching and stratification suggest improved balance between patient cohorts through adjustment.

| | Before adjustment | | | After matching | | | After stratification | | |
|---|---|---|---|---|---|---|---|---|---|
| Characteristic | T (%) | C (%) | StdDiff | T (%) | C (%) | StdDiff | T (%) | C (%) | StdDiff |
| **Age group** | | | | | | | | | |
| 50 - 54 | <0.4 | 0.1 | 0.04 | <0.5 | 0.2 | 0.02 | <0.1 | 0.1 | 0.01 |
| 55 - 59 | 1.1 | 0.5 | 0.07 | 1.1 | 0.7 | 0.04 | 0.3 | 0.5 | -0.03 |
| 60 - 64 | 2.5 | 1.3 | 0.09 | 2.6 | 1.8 | 0.06 | 2.2 | 1.4 | 0.06 |
| 65 - 69 | 34.6 | 19.8 | 0.34 | 33.3 | 35.4 | -0.04 | 19.9 | 20.9 | -0.03 |
| 70 - 74 | 36.8 | 28.4 | 0.18 | 37.0 | 36.3 | 0.02 | 34.3 | 28.7 | 0.12 |
| 75 - 79 | 16.2 | 23.3 | -0.18 | 16.6 | 15.9 | 0.02 | 19.6 | 22.6 | -0.07 |
| 80 - 84 | 6.1 | 16.4 | -0.33 | 6.4 | 5.8 | 0.02 | 17.3 | 15.8 | 0.04 |
| 85 - 89 | 2.0 | 7.6 | -0.27 | 2.1 | 2.6 | -0.03 | 6.1 | 7.5 | -0.05 |
| 90 - 94 | 0.6 | 2.2 | -0.14 | 0.7 | 1.2 | -0.05 | <0.1 | 2.2 | -0.20 |
| Gender: female | 49.2 | 42.6 | 0.13 | 48.7 | 48.2 | 0.01 | 45.2 | 42.6 | 0.05 |
| **Medical history: General** | | | | | | | | | |
| Acute respiratory disease | 25.8 | 24.4 | 0.03 | 25.8 | 25.2 | 0.01 | 33.9 | 25.0 | 0.20 |
| Attention deficit hyperactivity disorder | <0.4 | 0.1 | 0.02 | <0.5 | 0.5 | -0.07 | <0.1 | 0.1 | -0.04 |
| Chronic liver disease | 2.1 | 2.3 | -0.02 | 2.2 | 2.0 | 0.01 | 1.1 | 2.4 | -0.10 |
| Chronic obstructive lung disease | 15.7 | 15.2 | 0.01 | 15.8 | 15.4 | 0.01 | 25.2 | 15.6 | 0.24 |
| Crohn's disease | 0.5 | 0.3 | 0.04 | 0.6 | 0.4 | 0.03 | 0.5 | 0.3 | 0.03 |
| Dementia | 2.1 | 4.5 | -0.13 | 2.2 | 2.6 | -0.02 | 4.0 | 4.5 | -0.03 |
| Depressive disorder | 13.8 | 8.2 | 0.18 | 12.7 | 14.9 | -0.06 | 17.5 | 9.2 | 0.24 |
| Gastroesophageal reflux disease | 17.2 | 11.8 | 0.15 | 16.5 | 16.9 | -0.01 | 7.9 | 13.0 | -0.16 |
| Gastrointestinal hemorrhage | 2.9 | 4.6 | -0.09 | 2.8 | 3.1 | -0.02 | 3.1 | 4.4 | -0.07 |
| Human immunodeficiency virus infection | <0.4 | 0.1 | 0.01 | <0.5 | 0.1 | 0.01 | <0.1 | 0.1 | -0.02 |
| Hyperlipidemia | 69.0 | 52.6 | 0.34 | 67.2 | 71.1 | -0.08 | 64.7 | 57.4 | 0.15 |
| Hypertensive disorder | 81.4 | 71.3 | 0.24 | 80.5 | 82.1 | -0.04 | 79.2 | 74.4 | 0.12 |
| Lesion of liver | 0.9 | 1.3 | -0.04 | 1.0 | 1.5 | -0.05 | 0.3 | 1.4 | -0.12 |
| Obesity | 29.5 | 8.2 | 0.56 | 25.8 | 28.8 | -0.07 | 8.8 | 10.0 | -0.04 |
| Osteoarthritis | 39.4 | 30.1 | 0.20 | 38.5 | 37.5 | 0.02 | 36.9 | 31.7 | 0.11 |
| Pneumonia | 4.8 | 8.3 | -0.14 | 5.0 | 5.8 | -0.03 | 12.7 | 8.1 | 0.15 |
| Psoriasis | 2.9 | 1.4 | 0.11 | 2.9 | 2.1 | 0.05 | 2.0 | 1.5 | 0.04 |
| Renal impairment | 14.5 | 16.4 | -0.05 | 13.9 | 15.4 | -0.04 | 26.2 | 17.7 | 0.20 |
| Rheumatoid arthritis | 1.9 | 1.8 | 0.00 | 1.7 | 2.2 | -0.03 | 0.6 | 1.9 | -0.12 |
| Schizophrenia | <0.4 | 0.2 | 0.02 | <0.5 | 0.1 | 0.05 | <0.1 | 0.1 | 0.00 |
| Ulcerative colitis | <0.4 | 0.4 | -0.01 | <0.5 | 0.4 | 0.00 | 0.2 | 0.4 | -0.05 |
| Urinary tract infectious disease | 11.5 | 12.3 | -0.02 | 11.2 | 10.1 | 0.04 | 9.8 | 12.5 | -0.09 |
| Viral hepatitis C | <0.4 | 0.3 | -0.04 | <0.5 | 0.5 | -0.08 | <0.1 | 0.3 | -0.06 |
| Visual system disorder | 51.6 | 50.1 | 0.03 | 51.3 | 52.4 | -0.02 | 57.5 | 50.8 | 0.13 |
| **Medical history: Cardiovascular disease** | | | | | | | | | |
| Atrial fibrillation | 12.1 | 16.4 | -0.12 | 12.3 | 12.5 | 0.00 | 23.3 | 16.6 | 0.17 |
| Cerebrovascular disease | 17.9 | 22.0 | -0.10 | 17.9 | 18.0 | 0.00 | 17.1 | 21.3 | -0.10 |
| Coronary arteriosclerosis | 43.1 | 44.3 | -0.03 | 42.5 | 42.8 | -0.01 | 40.8 | 43.6 | -0.06 |
| Heart disease | 62.3 | 66.9 | -0.10 | 62.0 | 63.4 | -0.03 | 70.5 | 66.1 | 0.10 |
| Heart failure | 11.3 | 16.6 | -0.15 | 11.3 | 11.8 | -0.02 | 24.1 | 15.9 | 0.20 |
| Ischemic heart disease | 17.9 | 21.8 | -0.10 | 18.6 | 17.5 | 0.03 | 18.4 | 20.7 | -0.06 |
| Peripheral vascular disease | 22.5 | 20.0 | 0.06 | 22.5 | 23.8 | -0.03 | 17.6 | 20.3 | -0.07 |
| Pulmonary embolism | 1.3 | 1.3 | 0.00 | 1.2 | 1.7 | -0.04 | 2.7 | 1.3 | 0.10 |
| Venous thrombosis | 2.1 | 3.0 | -0.06 | 2.2 | 2.5 | -0.02 | 1.9 | 3.1 | -0.08 |
| **Medical history: Neoplasms** | | | | | | | | | |
| Hematologic neoplasm | 2.1 | 2.1 | 0.00 | 2.0 | 1.9 | 0.01 | 1.3 | 2.2 | -0.07 |
| Malignant lymphoma | 1.2 | 1.0 | 0.02 | 1.1 | 1.3 | -0.02 | 0.6 | 1.0 | -0.05 |
| Malignant neoplastic disease | 20.5 | 20.4 | 0.00 | 21.0 | 21.2 | 0.00 | 16.1 | 20.7 | -0.12 |
| Malignant tumor of breast | 3.8 | 2.6 | 0.07 | 3.8 | 4.0 | -0.01 | 1.9 | 2.6 | -0.05 |
| Malignant tumor of colon | 0.5 | 1.1 | -0.06 | 0.6 | 0.7 | -0.01 | <0.1 | 1.0 | -0.12 |
| Malignant tumor of lung | 0.8 | 1.1 | -0.03 | 0.9 | 0.9 | -0.01 | 1.2 | 1.1 | 0.01 |
| Malignant tumor of urinary bladder | 1.8 | 1.4 | 0.03 | 1.8 | 1.3 | 0.04 | 1.6 | 1.5 | 0.01 |
| Primary malignant neoplasm of prostate | 4.6 | 5.3 | -0.03 | 5.1 | 5.3 | -0.01 | 5.1 | 5.3 | -0.01 |
| **Medication use** | | | | | | | | | |
| Agents acting on the renin-angiotensin system | 78.2 | 74.3 | 0.09 | 78.5 | 77.7 | 0.02 | 80.1 | 75.9 | 0.10 |
| Antibacterials for systemic use | 66.7 | 65.0 | 0.04 | 66.2 | 64.7 | 0.03 | 63.2 | 65.7 | -0.05 |
| Antidepressants | 36.6 | 27.3 | 0.20 | 34.8 | 36.1 | -0.03 | 28.4 | 28.1 | 0.01 |
| Antiepileptics | 21.2 | 17.3 | 0.10 | 20.4 | 21.5 | -0.03 | 23.1 | 18.3 | 0.12 |
| Antiinflammatory and antirheumatic products | 34.6 | 27.6 | 0.15 | 33.7 | 33.0 | 0.01 | 26.2 | 27.9 | -0.04 |
| Antineoplastic agents | 10.1 | 9.6 | 0.01 | 10.3 | 9.1 | 0.04 | 7.6 | 9.1 | -0.06 |

| (Continued) | Before ajustment | | | After matching | | | After stratification | | |
|---|---|---|---|---|---|---|---|---|---|
| Characteristic | T (%) | C (%) | StdDiff | T (%) | C (%) | StdDiff | T (%) | C (%) | StdDiff |
| Antipsoriatics | 1.2 | 0.8 | 0.04 | 1.2 | 1.0 | 0.02 | 0.7 | 0.8 | -0.02 |
| Antithrombotic agents | 35.1 | 38.5 | -0.07 | 35.9 | 34.1 | 0.04 | 45.3 | 38.6 | 0.14 |
| Beta blocking agents | 55.9 | 61.0 | -0.10 | 56.2 | 54.6 | 0.03 | 70.0 | 61.4 | 0.18 |
| Calcium channel blockers | 33.7 | 37.1 | -0.07 | 33.9 | 32.7 | 0.03 | 28.5 | 37.4 | -0.19 |
| Diuretics | 56.4 | 56.7 | 0.00 | 56.4 | 54.3 | 0.04 | 57.5 | 56.4 | 0.02 |
| Drugs for acid related disorders | 39.9 | 36.7 | 0.06 | 39.4 | 37.9 | 0.03 | 35.9 | 36.8 | -0.02 |
| Drugs for obstructive airway diseases | 53.1 | 41.9 | 0.23 | 51.8 | 52.2 | -0.01 | 51.2 | 43.6 | 0.15 |
| Immunosuppressants | 4.1 | 2.9 | 0.07 | 4.0 | 4.0 | 0.00 | 11.8 | 3.1 | 0.33 |
| Lipid modifying agents | 86.7 | 80.2 | 0.18 | 86.7 | 86.9 | 0.00 | 78.0 | 82.4 | -0.11 |
| Opioids | 32.1 | 32.4 | -0.01 | 32.4 | 32.9 | -0.01 | 39.4 | 32.9 | 0.13 |
| Psycholeptics | 31.2 | 29.0 | 0.05 | 31.0 | 31.5 | -0.01 | 37.2 | 29.5 | 0.16 |
| Psychostimulants | 2.1 | 1.3 | 0.07 | 2.2 | 2.0 | 0.02 | 1.3 | 1.4 | -0.01 |

**Supplementary Table 36:** Baseline patient characteristics for GLP1-RAs (T) and SUs (C) new-users in the Open Claims data source. We report proportion of initiators satisfying selected base-line characteristics and the standardized difference of population proportions (StdDiff) before and after propensity score adjustment. Less extreme StdDiffs through matching and stratification suggest improved balance between patient cohorts through adjustment.

| | Before adjustment | | | After matching | | | After stratification | | |
|---|---|---|---|---|---|---|---|---|---|
| Characteristic | T (%) | C (%) | StdDiff | T (%) | C (%) | StdDiff | T (%) | C (%) | StdDiff |
| **Age group** | | | | | | | | | |
| 20 - 24 | 0.2 | 0.1 | 0.02 | 0.2 | 0.1 | 0.01 | 0.2 | 0.1 | 0.01 |
| 25 - 29 | 0.3 | 0.2 | 0.03 | 0.3 | 0.2 | 0.01 | 0.2 | 0.2 | 0.01 |
| 30 - 34 | 0.8 | 0.4 | 0.05 | 0.7 | 0.7 | -0.01 | 0.5 | 0.5 | 0.01 |
| 35 - 39 | 1.7 | 0.9 | 0.07 | 1.6 | 1.5 | 0.00 | 1.1 | 1.0 | 0.01 |
| 40 - 44 | 3.8 | 2.0 | 0.11 | 3.5 | 3.3 | 0.01 | 2.5 | 2.2 | 0.02 |
| 45 - 49 | 7.1 | 4.0 | 0.14 | 6.5 | 6.5 | 0.00 | 4.5 | 4.4 | 0.01 |
| 50 - 54 | 11.9 | 7.4 | 0.15 | 10.9 | 11.0 | 0.00 | 7.9 | 8.0 | 0.00 |
| 55 - 59 | 16.5 | 11.3 | 0.15 | 15.7 | 15.8 | 0.00 | 12.0 | 12.1 | 0.00 |
| 60 - 64 | 18.3 | 14.2 | 0.11 | 18.0 | 18.2 | 0.00 | 14.8 | 14.9 | 0.00 |
| 65 - 69 | 16.7 | 17.3 | -0.02 | 17.5 | 17.8 | -0.01 | 17.3 | 17.4 | 0.00 |
| 70 - 74 | 12.2 | 16.4 | -0.12 | 13.3 | 13.0 | 0.01 | 16.3 | 16.0 | 0.01 |
| 75 - 79 | 6.8 | 18.1 | -0.35 | 7.7 | 7.6 | 0.00 | 15.2 | 16.4 | -0.03 |
| 80 - 84 | 3.3 | 7.4 | -0.18 | 3.9 | 3.8 | 0.00 | 7.3 | 6.8 | 0.02 |
| 85 - 89 | 0.3 | 0.2 | 0.01 | 0.3 | 0.4 | -0.01 | 0.2 | 0.2 | -0.01 |
| Gender: female | 54.4 | 45.0 | 0.19 | 51.8 | 51.5 | 0.01 | 46.2 | 46.0 | 0.00 |
| **Medical history: General** | | | | | | | | | |
| Acute respiratory disease | 17.1 | 17.0 | 0.00 | 16.2 | 17.4 | -0.03 | 17.5 | 17.1 | 0.01 |
| Attention deficit hyperactivity disorder | 0.9 | 0.3 | 0.08 | 0.7 | 0.6 | 0.00 | 0.4 | 0.3 | 0.01 |
| Chronic liver disease | 1.1 | 1.9 | -0.06 | 1.2 | 1.2 | 0.00 | 1.8 | 1.7 | 0.01 |
| Chronic obstructive lung disease | 8.8 | 11.2 | -0.08 | 8.8 | 9.8 | -0.03 | 11.0 | 10.9 | 0.00 |
| Crohn’s disease | 0.2 | 0.2 | 0.00 | 0.2 | 0.2 | -0.01 | 0.1 | 0.2 | -0.02 |
| Dementia | 0.7 | 2.0 | -0.10 | 0.8 | 0.9 | -0.01 | 1.8 | 1.8 | 0.00 |
| Depressive disorder | 12.0 | 8.8 | 0.10 | 10.9 | 11.2 | -0.01 | 9.9 | 9.3 | 0.02 |
| Gastroesophageal reflux disease | 12.7 | 10.5 | 0.07 | 11.5 | 12.1 | -0.02 | 10.9 | 10.9 | 0.00 |
| Gastrointestinal hemorrhage | 1.8 | 2.3 | -0.04 | 1.7 | 1.9 | -0.01 | 2.2 | 2.3 | -0.01 |
| Human immunodeficiency virus infection | 0.4 | 0.3 | 0.02 | 0.3 | 0.3 | 0.01 | 0.3 | 0.3 | 0.00 |
| Hyperlipidemia | 50.1 | 50.0 | 0.00 | 49.0 | 49.9 | -0.02 | 50.7 | 50.2 | 0.01 |
| Hypertensive disorder | 59.4 | 61.3 | -0.04 | 58.5 | 59.4 | -0.02 | 61.5 | 61.1 | 0.01 |
| Lesion of liver | 0.9 | 1.1 | -0.03 | 0.9 | 0.9 | 0.00 | 0.9 | 1.1 | -0.02 |
| Obesity | 23.1 | 11.2 | 0.32 | 19.3 | 19.7 | -0.01 | 13.7 | 12.6 | 0.03 |
| Osteoarthritis | 23.6 | 20.5 | 0.07 | 22.1 | 22.8 | -0.02 | 21.7 | 20.9 | 0.02 |
| Pneumonia | 3.4 | 4.6 | -0.06 | 3.4 | 3.6 | -0.01 | 4.2 | 4.4 | -0.01 |
| Psoriasis | 1.5 | 0.9 | 0.05 | 1.3 | 1.4 | 0.00 | 1.1 | 1.0 | 0.01 |
| Renal impairment | 8.2 | 13.3 | -0.16 | 8.5 | 8.8 | -0.01 | 12.5 | 12.6 | 0.00 |
| Rheumatoid arthritis | 1.8 | 1.4 | 0.03 | 1.6 | 1.6 | 0.00 | 1.5 | 1.5 | 0.00 |
| Schizophrenia | 0.3 | 0.5 | -0.02 | 0.3 | 0.3 | 0.00 | 0.3 | 0.3 | 0.00 |
| Ulcerative colitis | 0.2 | 0.2 | 0.00 | 0.2 | 0.2 | 0.00 | 0.2 | 0.2 | 0.00 |
| Urinary tract infectious disease | 6.5 | 7.7 | -0.05 | 6.3 | 6.5 | -0.01 | 8.1 | 7.6 | 0.01 |
| Viral hepatitis C | 0.3 | 0.6 | -0.04 | 0.3 | 0.4 | -0.01 | 0.5 | 0.5 | -0.01 |
| Visual system disorder | 21.3 | 23.4 | -0.05 | 21.0 | 21.5 | -0.01 | 22.6 | 23.1 | -0.01 |
| **Medical history: Cardiovascular disease** | | | | | | | | | |
| Atrial fibrillation | 6.1 | 8.8 | -0.11 | 6.2 | 6.5 | -0.01 | 8.4 | 8.5 | 0.00 |
| Cerebrovascular disease | 6.4 | 9.6 | -0.12 | 6.4 | 6.5 | 0.00 | 8.8 | 9.2 | -0.01 |
| Coronary arteriosclerosis | 20.2 | 25.4 | -0.12 | 20.4 | 21.1 | -0.02 | 24.6 | 24.7 | 0.00 |
| Heart disease | 35.4 | 42.3 | -0.14 | 35.2 | 36.3 | -0.02 | 41.2 | 41.4 | 0.00 |
| Heart failure | 6.8 | 10.1 | -0.12 | 6.9 | 7.4 | -0.02 | 9.6 | 9.7 | -0.01 |
| Ischemic heart disease | 9.7 | 12.6 | -0.09 | 9.7 | 10.2 | -0.02 | 11.8 | 12.2 | -0.01 |
| Peripheral vascular disease | 13.3 | 14.0 | -0.02 | 13.2 | 13.4 | 0.00 | 13.9 | 14.0 | 0.00 |
| Pulmonary embolism | 1.1 | 1.0 | 0.01 | 1.0 | 1.0 | 0.00 | 1.1 | 1.0 | 0.01 |
| Venous thrombosis | 1.2 | 1.6 | -0.03 | 1.2 | 1.2 | 0.00 | 1.7 | 1.6 | 0.01 |
| **Medical history: Neoplasms** | | | | | | | | | |
| Hematologic neoplasm | 0.8 | 1.2 | -0.04 | 0.8 | 0.8 | 0.00 | 1.2 | 1.1 | 0.01 |
| Malignant lymphoma | 0.4 | 0.6 | -0.03 | 0.5 | 0.5 | 0.00 | 0.6 | 0.6 | 0.00 |
| Malignant neoplasm of anorectum | 0.1 | 0.2 | -0.03 | 0.1 | 0.1 | 0.00 | 0.2 | 0.2 | 0.00 |
| Malignant neoplastic disease | 6.9 | 9.4 | -0.09 | 7.0 | 7.2 | -0.01 | 9.2 | 9.0 | 0.01 |
| Malignant tumor of breast | 1.3 | 1.4 | 0.00 | 1.3 | 1.4 | -0.01 | 1.3 | 1.4 | -0.01 |
| Malignant tumor of colon | 0.3 | 0.5 | -0.04 | 0.3 | 0.3 | 0.00 | 0.5 | 0.5 | 0.01 |
| Malignant tumor of lung | 0.3 | 0.6 | -0.05 | 0.6 | 0.5 | 0.01 | 0.6 | 0.5 | 0.01 |
| Malignant tumor of urinary bladder | 0.3 | 0.5 | -0.03 | 0.3 | 0.4 | -0.01 | 0.7 | 0.5 | 0.02 |
| Primary malignant neoplasm of prostate | 1.2 | 1.9 | -0.06 | 1.3 | 1.3 | 0.00 | 2.0 | 1.9 | 0.01 |
| **Medication use** | | | | | | | | | |

(Continued)

| Characteristic | Before ajustment | | | After matching | | | After stratification | | |
|---|---|---|---|---|---|---|---|---|---|
| | T (%) | C (%) | StdDiff | T (%) | C (%) | StdDiff | T (%) | C (%) | StdDiff |
| Agents acting on the renin-angiotensin system | 68.7 | 73.2 | -0.10 | 69.5 | 70.1 | -0.01 | 72.6 | 72.7 | 0.00 |
| Antibacterials for systemic use | 63.4 | 60.7 | 0.06 | 62.0 | 62.8 | -0.01 | 61.7 | 61.2 | 0.01 |
| Antidepressants | 42.9 | 32.5 | 0.22 | 39.9 | 40.7 | -0.02 | 35.7 | 34.1 | 0.04 |
| Antiepileptics | 30.0 | 25.3 | 0.10 | 28.3 | 29.4 | -0.03 | 27.3 | 26.0 | 0.03 |
| Antiinflammatory and antirheumatic products | 43.4 | 37.7 | 0.12 | 41.8 | 42.4 | -0.01 | 39.1 | 38.6 | 0.01 |
| Antineoplastic agents | 7.2 | 5.3 | 0.07 | 6.4 | 6.7 | -0.01 | 6.1 | 5.7 | 0.02 |
| Antipsoriatics | 1.1 | 0.7 | 0.04 | 1.0 | 1.0 | 0.00 | 0.7 | 0.8 | -0.01 |
| Antithrombotic agents | 29.8 | 36.4 | -0.14 | 30.5 | 31.1 | -0.01 | 35.9 | 35.6 | 0.01 |
| Beta blocking agents | 47.1 | 54.4 | -0.15 | 47.8 | 48.3 | -0.01 | 53.7 | 53.5 | 0.00 |
| Calcium channel blockers | 30.4 | 33.6 | -0.07 | 30.8 | 31.2 | -0.01 | 32.5 | 33.1 | -0.01 |
| Diuretics | 49.1 | 48.6 | 0.01 | 48.2 | 48.4 | 0.00 | 48.9 | 48.5 | 0.01 |
| Drugs for acid related disorders | 43.3 | 41.1 | 0.04 | 41.8 | 42.7 | -0.02 | 42.0 | 41.6 | 0.01 |
| Drugs for obstructive airway diseases | 48.7 | 40.9 | 0.16 | 46.4 | 47.7 | -0.03 | 42.3 | 42.2 | 0.00 |
| Immunosuppressants | 4.4 | 2.9 | 0.08 | 4.0 | 3.9 | 0.00 | 3.3 | 3.1 | 0.01 |
| Lipid modifying agents | 76.1 | 77.7 | -0.04 | 76.5 | 77.2 | -0.02 | 77.9 | 77.7 | 0.00 |
| Opioids | 31.9 | 31.5 | 0.01 | 30.8 | 32.1 | -0.03 | 32.6 | 31.4 | 0.03 |
| Psycholeptics | 31.8 | 27.9 | 0.08 | 29.8 | 31.0 | -0.03 | 29.9 | 28.5 | 0.03 |
| Psychostimulants | 4.7 | 2.2 | 0.14 | 3.8 | 3.9 | -0.01 | 2.7 | 2.5 | 0.02 |

**Supplementary Table 37:** Baseline patient characteristics for GLP1-RAs (T) and SUs (C) new-users in the OptumEHR data source. We report proportion of initiators satisfying selected base-line characteristics and the standardized difference of population proportions (StdDiff) before and after propensity score adjustment. Less extreme StdDiffs through matching and stratification suggest improved balance between patient cohorts through adjustment.

| | Before adjustment | | | After matching | | | After stratification | | |
|---|---|---|---|---|---|---|---|---|---|
| Characteristic | T (%) | C (%) | StdDiff | T (%) | C (%) | StdDiff | T (%) | C (%) | StdDiff |
| **Age group** | | | | | | | | | |
| 20 - 24 | 0.1 | 0.1 | 0.02 | <0.1 | 0.1 | -0.01 | <0.1 | 0.1 | -0.01 |
| 25 - 29 | 0.4 | 0.1 | 0.06 | 0.4 | 0.3 | 0.02 | 0.4 | 0.1 | 0.06 |
| 30 - 34 | 0.6 | 0.3 | 0.05 | 0.7 | 0.5 | 0.02 | 0.9 | 0.3 | 0.07 |
| 35 - 39 | 2.0 | 0.9 | 0.09 | 1.9 | 1.7 | 0.01 | 1.4 | 1.0 | 0.03 |
| 40 - 44 | 4.8 | 2.0 | 0.16 | 4.2 | 4.3 | 0.00 | 2.3 | 2.3 | 0.00 |
| 45 - 49 | 8.3 | 4.6 | 0.15 | 8.0 | 7.6 | 0.01 | 4.4 | 5.0 | -0.03 |
| 50 - 54 | 14.3 | 8.2 | 0.19 | 13.3 | 13.8 | -0.01 | 9.8 | 8.8 | 0.04 |
| 55 - 59 | 17.6 | 12.6 | 0.14 | 17.5 | 17.3 | 0.00 | 12.8 | 13.1 | -0.01 |
| 60 - 64 | 19.8 | 15.6 | 0.11 | 19.7 | 19.6 | 0.00 | 15.0 | 16.0 | -0.03 |
| 65 - 69 | 15.2 | 16.3 | -0.03 | 15.8 | 16.9 | -0.03 | 15.5 | 16.3 | -0.02 |
| 70 - 74 | 9.7 | 15.4 | -0.17 | 10.5 | 10.3 | 0.01 | 15.7 | 14.8 | 0.02 |
| 75 - 79 | 5.0 | 13.2 | -0.29 | 5.5 | 5.1 | 0.02 | 11.7 | 12.4 | -0.02 |
| 80 - 84 | 1.6 | 8.7 | -0.33 | 1.8 | 1.7 | 0.00 | 8.6 | 8.0 | 0.02 |
| 85 - 89 | 0.4 | 2.0 | -0.14 | 0.5 | 0.7 | -0.03 | 1.4 | 1.9 | -0.04 |
| Gender: female | 50.2 | 39.3 | 0.22 | 47.6 | 48.4 | -0.01 | 39.0 | 40.2 | -0.02 |
| **Race** | | | | | | | | | |
| Asian | 1.1 | 1.7 | -0.05 | 1.2 | 1.4 | -0.02 | 0.9 | 1.7 | -0.06 |
| Black or African American | 10.1 | 8.0 | 0.07 | 9.8 | 9.4 | 0.01 | 9.4 | 8.2 | 0.04 |
| White | 83.6 | 84.5 | -0.02 | 83.9 | 83.7 | 0.01 | 85.1 | 84.4 | 0.02 |
| **Ethnicity** | | | | | | | | | |
| Hispanic or Latino | 4.0 | 4.9 | -0.04 | 3.9 | 4.4 | -0.02 | 5.0 | 4.8 | 0.01 |
| Not Hispanic or Latino | 88.3 | 88.4 | -0.01 | 88.5 | 88.2 | 0.01 | 89.0 | 88.4 | 0.02 |
| **Medical history: General** | | | | | | | | | |
| Acute respiratory disease | 21.3 | 18.1 | 0.08 | 20.1 | 20.6 | -0.01 | 16.7 | 18.3 | -0.04 |
| Attention deficit hyperactivity disorder | 1.0 | 0.4 | 0.07 | 0.8 | 0.8 | 0.00 | 0.4 | 0.4 | 0.00 |
| Chronic liver disease | 2.7 | 3.1 | -0.02 | 2.6 | 2.6 | 0.00 | 3.2 | 3.1 | 0.01 |
| Chronic obstructive lung disease | 10.2 | 12.1 | -0.06 | 10.6 | 10.5 | 0.00 | 13.0 | 12.0 | 0.03 |
| Crohn’s disease | 0.4 | 0.2 | 0.02 | 0.4 | 0.3 | 0.01 | 0.2 | 0.3 | -0.02 |
| Dementia | 0.9 | 2.2 | -0.11 | 0.9 | 0.9 | 0.00 | 0.8 | 2.1 | -0.11 |
| Depressive disorder | 21.4 | 15.1 | 0.16 | 19.6 | 20.4 | -0.02 | 17.1 | 15.7 | 0.04 |
| Gastroesophageal reflux disease | 24.8 | 19.8 | 0.12 | 22.9 | 24.0 | -0.03 | 21.6 | 20.3 | 0.03 |
| Gastrointestinal hemorrhage | 2.0 | 2.2 | -0.01 | 2.1 | 1.7 | 0.03 | 3.8 | 2.2 | 0.10 |
| Human immunodeficiency virus infection | 0.2 | 0.2 | 0.01 | 0.2 | 0.3 | -0.01 | 0.1 | 0.2 | -0.03 |
| Hyperlipidemia | 74.6 | 73.9 | 0.02 | 73.3 | 74.1 | -0.02 | 77.5 | 73.9 | 0.08 |
| Hypertensive disorder | 77.5 | 76.8 | 0.02 | 76.5 | 77.5 | -0.03 | 80.3 | 76.8 | 0.09 |
| Lesion of liver | 1.1 | 1.2 | -0.01 | 0.9 | 1.4 | -0.04 | 0.5 | 1.2 | -0.08 |
| Obesity | 47.5 | 23.7 | 0.51 | 41.9 | 44.8 | -0.06 | 28.8 | 25.8 | 0.07 |
| Osteoarthritis | 25.3 | 22.2 | 0.07 | 24.0 | 24.5 | -0.01 | 22.0 | 22.4 | -0.01 |
| Pneumonia | 3.0 | 3.8 | -0.05 | 2.6 | 3.7 | -0.06 | 2.6 | 3.7 | -0.06 |
| Psoriasis | 2.3 | 1.5 | 0.06 | 2.1 | 1.8 | 0.02 | 1.3 | 1.5 | -0.01 |
| Renal impairment | 9.1 | 13.9 | -0.15 | 9.3 | 9.3 | 0.00 | 11.6 | 13.4 | -0.06 |
| Rheumatoid arthritis | 1.8 | 1.5 | 0.02 | 1.7 | 1.8 | -0.01 | 2.8 | 1.5 | 0.09 |
| Schizophrenia | 0.3 | 0.3 | 0.00 | 0.3 | 0.4 | 0.00 | 0.3 | 0.3 | -0.01 |
| Ulcerative colitis | 0.4 | 0.3 | 0.01 | 0.4 | 0.3 | 0.01 | 0.1 | 0.3 | -0.03 |
| Urinary tract infectious disease | 5.2 | 6.7 | -0.06 | 5.0 | 5.4 | -0.02 | 5.2 | 6.6 | -0.06 |
| Viral hepatitis C | 0.8 | 0.7 | 0.01 | 0.8 | 0.6 | 0.03 | 0.5 | 0.7 | -0.03 |
| Visual system disorder | 15.0 | 18.0 | -0.08 | 14.5 | 14.5 | 0.00 | 13.9 | 17.6 | -0.10 |
| **Medical history: Cardiovascular disease** | | | | | | | | | |
| Atrial fibrillation | 7.5 | 10.7 | -0.11 | 7.7 | 7.5 | 0.01 | 11.5 | 10.4 | 0.04 |
| Cerebrovascular disease | 8.6 | 11.5 | -0.10 | 8.5 | 8.8 | -0.01 | 9.9 | 11.2 | -0.04 |
| Coronary arteriosclerosis | 34.5 | 42.8 | -0.17 | 35.0 | 34.6 | 0.01 | 42.7 | 42.0 | 0.01 |
| Heart disease | 52.6 | 59.1 | -0.13 | 52.7 | 51.9 | 0.02 | 58.7 | 58.3 | 0.01 |
| Heart failure | 7.8 | 10.3 | -0.09 | 8.1 | 8.0 | 0.00 | 11.6 | 10.1 | 0.05 |
| Ischemic heart disease | 17.8 | 19.6 | -0.05 | 17.9 | 17.2 | 0.02 | 19.0 | 19.4 | -0.01 |
| Peripheral vascular disease | 20.9 | 17.9 | 0.07 | 20.1 | 20.8 | -0.02 | 16.3 | 18.2 | -0.05 |
| Pulmonary embolism | 1.1 | 1.1 | 0.00 | 1.1 | 1.1 | 0.00 | 1.7 | 1.1 | 0.05 |
| Venous thrombosis | 1.8 | 1.9 | -0.01 | 1.7 | 1.5 | 0.02 | 1.5 | 1.9 | -0.03 |
| **Medical history: Neoplasms** | | | | | | | | | |
| Hematologic neoplasm | 0.6 | 1.2 | -0.06 | 0.6 | 0.9 | -0.03 | 2.2 | 1.1 | 0.08 |
| Malignant lymphoma | 0.5 | 0.7 | -0.02 | 0.5 | 0.5 | 0.01 | 0.3 | 0.7 | -0.05 |
| Malignant neoplasm of anorectum | 0.2 | 0.2 | 0.00 | 0.2 | 0.1 | 0.02 | 0.2 | 0.1 | 0.02 |

| (Continued) | Before ajustment | | | After matching | | | After stratification | | |
|---|---|---|---|---|---|---|---|---|---|
| Characteristic | T (%) | C (%) | StdDiff | T (%) | C (%) | StdDiff | T (%) | C (%) | StdDiff |
| Malignant neoplastic disease | 7.2 | 9.9 | -0.09 | 7.0 | 6.9 | 0.00 | 11.1 | 9.6 | 0.05 |
| Malignant tumor of breast | 1.4 | 1.3 | 0.01 | 1.4 | 1.2 | 0.01 | 0.9 | 1.3 | -0.04 |
| Malignant tumor of colon | 0.2 | 0.6 | -0.06 | 0.2 | 0.3 | -0.02 | 0.1 | 0.5 | -0.07 |
| Malignant tumor of lung | 0.1 | 0.5 | -0.06 | 0.2 | 0.3 | -0.03 | 0.1 | 0.4 | -0.08 |
| Malignant tumor of urinary bladder | 0.3 | 0.6 | -0.04 | 0.3 | 0.3 | -0.01 | 0.4 | 0.6 | -0.02 |
| Primary malignant neoplasm of prostate | 1.0 | 2.2 | -0.10 | 1.1 | 1.2 | 0.00 | 3.8 | 2.1 | 0.10 |
| **Medication use** | | | | | | | | | |
| Agents acting on the renin-angiotensin system | 71.3 | 73.0 | -0.04 | 72.0 | 71.5 | 0.01 | 73.7 | 72.8 | 0.02 |
| Antibacterials for systemic use | 50.3 | 46.2 | 0.08 | 48.4 | 49.1 | -0.01 | 48.0 | 46.5 | 0.03 |
| Antidepressants | 42.5 | 32.0 | 0.22 | 40.1 | 41.4 | -0.03 | 34.8 | 32.9 | 0.04 |
| Antiepileptics | 26.5 | 22.2 | 0.10 | 25.1 | 25.9 | -0.02 | 25.1 | 22.6 | 0.06 |
| Antiinflammatory and antirheumatic products | 70.5 | 69.5 | 0.02 | 70.1 | 70.3 | 0.00 | 69.8 | 69.6 | 0.00 |
| Antineoplastic agents | 6.1 | 4.8 | 0.06 | 5.6 | 5.7 | 0.00 | 5.8 | 4.8 | 0.04 |
| Antipsoriatics | 0.8 | 0.6 | 0.03 | 0.7 | 0.6 | 0.01 | 1.1 | 0.6 | 0.06 |
| Antithrombotic agents | 61.2 | 68.9 | -0.16 | 62.0 | 61.6 | 0.01 | 62.0 | 61.6 | 0.01 |
| Beta blocking agents | 51.7 | 59.5 | -0.16 | 52.4 | 51.9 | 0.01 | 59.7 | 58.7 | 0.02 |
| Calcium channel blockers | 29.3 | 30.4 | -0.02 | 28.6 | 29.6 | -0.02 | 33.5 | 30.3 | 0.07 |
| Diuretics | 50.4 | 48.4 | 0.04 | 48.9 | 49.3 | -0.01 | 51.8 | 48.5 | 0.07 |
| Drugs for acid related disorders | 49.0 | 47.8 | 0.02 | 47.8 | 48.4 | -0.01 | 50.3 | 47.8 | 0.05 |
| Drugs for obstructive airway diseases | 54.1 | 46.1 | 0.16 | 51.6 | 52.9 | -0.03 | 46.0 | 46.8 | -0.01 |
| Immunosuppressants | 4.4 | 2.8 | 0.09 | 4.3 | 4.0 | 0.01 | 4.4 | 2.9 | 0.08 |
| Lipid modifying agents | 81.5 | 82.4 | -0.02 | 81.7 | 81.3 | 0.01 | 84.0 | 82.3 | 0.05 |
| Opioids | 36.6 | 36.2 | 0.01 | 35.6 | 35.7 | 0.00 | 41.0 | 36.2 | 0.10 |
| Psycholeptics | 39.4 | 34.4 | 0.10 | 37.6 | 38.6 | -0.02 | 35.7 | 34.8 | 0.02 |
| Psychostimulants | 5.1 | 2.3 | 0.15 | 4.5 | 4.3 | 0.01 | 3.0 | 2.5 | 0.03 |

**Supplementary Table 38:** Baseline patient characteristics for DPP4is (T) and SUs (C) new-users in the CCAE data source. We report proportion of initiators satisfying selected base-line characteristics and the standardized difference of population proportions (StdDiff) before and after propensity score adjustment. Less extreme StdDiffs through matching and stratification suggest improved balance between patient cohorts through adjustment.

| | Before adjustment | | | After matching | | | After stratification | | |
|---|---|---|---|---|---|---|---|---|---|
| Characteristic | T (%) | C (%) | StdDiff | T (%) | C (%) | StdDiff | T (%) | C (%) | StdDiff |
| **Age group** | | | | | | | | | |
| 25 - 29 | 0.1 | 0.1 | -0.01 | 0.1 | 0.1 | 0.00 | 0.1 | 0.1 | -0.01 |
| 30 - 34 | 0.5 | 0.4 | 0.01 | 0.5 | 0.4 | 0.02 | 0.5 | 0.4 | 0.01 |
| 35 - 39 | 1.5 | 1.6 | 0.00 | 1.6 | 1.6 | 0.00 | 1.6 | 1.6 | 0.00 |
| 40 - 44 | 4.3 | 4.2 | 0.00 | 4.2 | 4.1 | 0.01 | 4.3 | 4.2 | 0.01 |
| 45 - 49 | 9.3 | 9.2 | 0.00 | 9.4 | 9.2 | 0.01 | 9.4 | 9.2 | 0.01 |
| 50 - 54 | 18.1 | 17.7 | 0.01 | 18.0 | 17.7 | 0.01 | 17.8 | 17.7 | 0.00 |
| 55 - 59 | 27.6 | 27.3 | 0.01 | 27.6 | 27.5 | 0.00 | 27.4 | 27.2 | 0.00 |
| 60 - 64 | 35.0 | 35.6 | -0.01 | 35.0 | 35.6 | -0.01 | 35.1 | 35.7 | -0.01 |
| 65 - 69 | 3.6 | 3.8 | -0.01 | 3.6 | 3.8 | -0.01 | 3.6 | 3.8 | -0.01 |
| Gender: female | 42.2 | 39.2 | 0.06 | 40.9 | 41.5 | -0.01 | 40.1 | 40.4 | 0.00 |
| **Medical history: General** | | | | | | | | | |
| Acute respiratory disease | 30.6 | 28.4 | 0.05 | 29.7 | 30.0 | -0.01 | 29.7 | 29.7 | 0.00 |
| Attention deficit hyperactivity disorder | 0.7 | 0.5 | 0.02 | 0.6 | 0.6 | 0.00 | 0.6 | 0.6 | 0.00 |
| Chronic liver disease | 4.1 | 3.9 | 0.01 | 4.0 | 4.0 | 0.00 | 4.1 | 4.0 | 0.00 |
| Chronic obstructive lung disease | 6.2 | 7.0 | -0.03 | 6.4 | 6.2 | 0.01 | 6.8 | 6.7 | 0.00 |
| Crohn’s disease | 0.4 | 0.3 | 0.00 | 0.3 | 0.3 | 0.00 | 0.3 | 0.3 | 0.00 |
| Dementia | 0.3 | 0.4 | 0.00 | 0.4 | 0.3 | 0.00 | 0.4 | 0.3 | 0.00 |
| Gastroesophageal reflux disease | 14.7 | 12.8 | 0.06 | 14.1 | 14.5 | -0.01 | 14.0 | 14.3 | -0.01 |
| Gastrointestinal hemorrhage | 2.7 | 2.7 | 0.00 | 2.7 | 2.6 | 0.00 | 2.6 | 2.7 | 0.00 |
| Human immunodeficiency virus infection | 0.3 | 0.3 | 0.01 | 0.3 | 0.3 | 0.00 | 0.3 | 0.3 | 0.00 |
| Hyperlipidemia | 75.0 | 68.9 | 0.14 | 73.8 | 74.1 | -0.01 | 72.8 | 72.9 | 0.00 |
| Hypertensive disorder | 76.5 | 73.1 | 0.08 | 76.0 | 76.2 | 0.00 | 75.9 | 75.7 | 0.00 |
| Lesion of liver | 0.9 | 1.2 | -0.02 | 0.9 | 1.0 | -0.01 | 1.0 | 1.1 | -0.01 |
| Obesity | 19.6 | 17.5 | 0.06 | 19.3 | 19.5 | 0.00 | 19.2 | 19.5 | -0.01 |
| Osteoarthritis | 23.7 | 21.8 | 0.05 | 23.2 | 23.5 | -0.01 | 23.3 | 23.2 | 0.00 |
| Pneumonia | 3.2 | 4.0 | -0.04 | 3.3 | 3.3 | 0.00 | 3.6 | 3.6 | 0.00 |
| Psoriasis | 1.7 | 1.5 | 0.01 | 1.7 | 1.7 | 0.00 | 1.6 | 1.6 | 0.00 |
| Renal impairment | 5.8 | 6.3 | -0.02 | 6.0 | 6.0 | 0.00 | 6.6 | 6.4 | 0.00 |
| Rheumatoid arthritis | 1.6 | 1.4 | 0.01 | 1.5 | 1.5 | -0.01 | 1.5 | 1.5 | 0.00 |
| Schizophrenia | 0.2 | 0.1 | 0.00 | 0.2 | 0.1 | 0.02 | 0.2 | 0.1 | 0.01 |
| Ulcerative colitis | 0.4 | 0.4 | 0.00 | 0.4 | 0.4 | -0.01 | 0.4 | 0.4 | -0.01 |
| Urinary tract infectious disease | 8.9 | 8.5 | 0.01 | 8.4 | 8.6 | -0.01 | 8.8 | 8.6 | 0.00 |
| Viral hepatitis C | 0.5 | 0.6 | -0.02 | 0.5 | 0.5 | -0.01 | 0.5 | 0.6 | -0.01 |
| Visual system disorder | 28.3 | 25.6 | 0.06 | 27.1 | 27.5 | -0.01 | 26.5 | 26.9 | -0.01 |
| **Medical history: Cardiovascular disease** | | | | | | | | | |
| Atrial fibrillation | 4.8 | 5.0 | -0.01 | 4.8 | 4.9 | 0.00 | 5.0 | 5.0 | 0.00 |
| Cerebrovascular disease | 10.4 | 11.0 | -0.02 | 10.2 | 10.3 | 0.00 | 10.7 | 10.6 | 0.00 |
| Coronary arteriosclerosis | 32.1 | 33.8 | -0.04 | 32.6 | 32.1 | 0.01 | 32.9 | 32.6 | 0.01 |
| Heart disease | 49.4 | 51.1 | -0.03 | 49.3 | 49.0 | 0.01 | 49.9 | 49.6 | 0.01 |
| Heart failure | 5.8 | 7.0 | -0.05 | 6.6 | 6.4 | 0.00 | 6.6 | 6.4 | 0.00 |
| Ischemic heart disease | 16.8 | 19.6 | -0.07 | 17.1 | 17.0 | 0.00 | 17.9 | 17.8 | 0.00 |
| Peripheral vascular disease | 13.7 | 12.5 | 0.03 | 13.3 | 13.4 | 0.00 | 13.3 | 13.2 | 0.00 |
| Pulmonary embolism | 0.8 | 0.9 | -0.01 | 0.9 | 0.9 | 0.00 | 0.9 | 0.9 | 0.00 |
| Venous thrombosis | 1.9 | 1.8 | 0.01 | 1.9 | 1.8 | 0.01 | 2.0 | 1.9 | 0.01 |
| **Medical history: Neoplasms** | | | | | | | | | |
| Malignant lymphoma | 0.6 | 0.6 | 0.00 | 0.5 | 0.6 | 0.00 | 0.5 | 0.6 | -0.01 |
| Malignant neoplasm of anorectum | 0.2 | 0.2 | -0.01 | 0.2 | 0.2 | 0.00 | 0.2 | 0.2 | 0.01 |
| Malignant neoplastic disease | 8.4 | 8.1 | 0.01 | 8.2 | 8.1 | 0.00 | 8.3 | 8.2 | 0.00 |
| Malignant tumor of breast | 1.5 | 1.4 | 0.01 | 1.5 | 1.5 | 0.00 | 1.5 | 1.5 | 0.00 |
| Malignant tumor of colon | 0.4 | 0.5 | -0.01 | 0.5 | 0.4 | 0.00 | 0.5 | 0.5 | 0.01 |
| Malignant tumor of lung | 0.3 | 0.4 | -0.01 | 0.3 | 0.3 | 0.01 | 0.4 | 0.3 | 0.01 |
| Malignant tumor of urinary bladder | 0.4 | 0.4 | 0.00 | 0.4 | 0.3 | 0.01 | 0.4 | 0.4 | 0.00 |
| Primary malignant neoplasm of prostate | 1.2 | 1.1 | 0.00 | 1.2 | 1.2 | 0.00 | 1.2 | 1.2 | 0.00 |
| **Medication use** | | | | | | | | | |
| Agents acting on the renin-angiotensin system | 73.8 | 73.2 | 0.01 | 73.7 | 73.4 | 0.00 | 74.2 | 73.9 | 0.01 |
| Antibacterials for systemic use | 64.3 | 62.8 | 0.03 | 63.3 | 63.8 | -0.01 | 63.6 | 63.5 | 0.00 |
| Antidepressants | 29.3 | 30.2 | -0.02 | 29.2 | 28.9 | 0.01 | 29.9 | 29.7 | 0.00 |
| Antiepileptics | 16.1 | 16.5 | -0.01 | 16.3 | 15.9 | 0.01 | 16.9 | 16.5 | 0.01 |
| Antiinflammatory and antirheumatic products | 35.2 | 34.5 | 0.01 | 34.8 | 34.9 | 0.00 | 35.1 | 35.1 | 0.00 |
| Antipsoriatics | 0.7 | 0.7 | 0.01 | 0.7 | 0.7 | 0.00 | 0.7 | 0.7 | 0.00 |
| Antithrombotic agents | 26.1 | 27.7 | -0.04 | 26.8 | 26.3 | 0.01 | 27.7 | 27.2 | 0.01 |

| (Continued) | Before ajustment | | | After matching | | | After stratification | | |
|---|---|---|---|---|---|---|---|---|---|
| Characteristic | T (%) | C (%) | StdDiff | T (%) | C (%) | StdDiff | T (%) | C (%) | StdDiff |
| Beta blocking agents | 46.6 | 49.1 | -0.05 | 47.3 | 46.7 | 0.01 | 48.4 | 48.0 | 0.01 |
| Calcium channel blockers | 27.0 | 27.1 | 0.00 | 26.9 | 26.9 | 0.00 | 27.2 | 27.2 | 0.00 |
| Diuretics | 43.7 | 44.9 | -0.03 | 43.7 | 43.4 | 0.01 | 44.7 | 44.1 | 0.01 |
| Drugs for acid related disorders | 32.2 | 31.3 | 0.02 | 31.5 | 31.4 | 0.00 | 31.4 | 31.5 | 0.00 |
| Drugs for obstructive airway diseases | 41.3 | 38.3 | 0.06 | 40.3 | 40.5 | 0.00 | 40.0 | 40.2 | 0.00 |
| Immunosuppressants | 3.0 | 2.5 | 0.03 | 2.9 | 2.8 | 0.01 | 2.9 | 2.8 | 0.01 |
| Lipid modifying agents | 78.3 | 76.7 | 0.04 | 78.0 | 77.8 | 0.00 | 77.8 | 77.8 | 0.00 |
| Opioids | 32.5 | 33.9 | -0.03 | 32.7 | 32.5 | 0.00 | 33.9 | 33.6 | 0.01 |
| Psychostimulants | 3.2 | 2.8 | 0.02 | 3.1 | 3.0 | 0.01 | 3.1 | 3.0 | 0.01 |

**Supplementary Table 39:** Baseline patient characteristics for DPP4is (T) and SUs (C) new-users in the GermanyDA data source. We report proportion of initiators satisfying selected base-line characteristics and the standardized difference of population proportions (StdDiff) before and after propensity score adjustment. Less extreme StdDiffs through matching and stratification suggest improved balance between patient cohorts through adjustment.

| | Before adjustment | | | After matching | | | After stratification | | |
|---|---|---|---|---|---|---|---|---|---|
| Characteristic | T (%) | C (%) | StdDiff | T (%) | C (%) | StdDiff | T (%) | C (%) | StdDiff |
| **Age group** | | | | | | | | | |
| 30 - 34 | 0.1 | <0.2 | -0.01 | <0.3 | 0.2 | -0.01 | 0.1 | <0.1 | 0.00 |
| 35 - 39 | 0.3 | 0.4 | -0.01 | 0.3 | 0.5 | -0.03 | 0.3 | 0.5 | -0.02 |
| 40 - 44 | 0.9 | 1.1 | -0.02 | 0.8 | 1.2 | -0.04 | 0.9 | 0.8 | 0.02 |
| 45 - 49 | 2.6 | 2.2 | 0.02 | 2.7 | 2.2 | 0.03 | 2.6 | 2.2 | 0.03 |
| 50 - 54 | 5.8 | 4.5 | 0.06 | 5.6 | 4.7 | 0.04 | 5.6 | 4.5 | 0.05 |
| 55 - 59 | 10.4 | 8.1 | 0.08 | 8.8 | 8.4 | 0.01 | 10.0 | 8.7 | 0.04 |
| 60 - 64 | 13.2 | 13.6 | -0.01 | 13.1 | 12.6 | 0.01 | 13.2 | 13.1 | 0.00 |
| 65 - 69 | 14.6 | 16.9 | -0.06 | 14.8 | 14.3 | 0.02 | 14.8 | 16.0 | -0.03 |
| 70 - 74 | 15.7 | 19.2 | -0.09 | 18.9 | 18.1 | 0.02 | 16.5 | 15.8 | 0.02 |
| 75 - 79 | 16.6 | 18.4 | -0.05 | 16.9 | 19.1 | -0.06 | 16.7 | 17.6 | -0.02 |
| 80 - 84 | 12.4 | 10.5 | 0.06 | 11.7 | 12.0 | -0.01 | 12.2 | 12.9 | -0.02 |
| 85 - 89 | 5.7 | 3.9 | 0.08 | 5.0 | 5.4 | -0.02 | 5.5 | 6.1 | -0.03 |
| 90 - 94 | 1.5 | 0.9 | 0.05 | 1.2 | 1.4 | -0.02 | 1.4 | 1.8 | -0.03 |
| Gender: female | 40.5 | 43.5 | -0.06 | 40.4 | 43.3 | -0.06 | 41.2 | 41.7 | -0.01 |
| **Medical history: General** | | | | | | | | | |
| Acute respiratory disease | 18.2 | 17.7 | 0.01 | 19.7 | 18.1 | 0.04 | 18.4 | 17.0 | 0.04 |
| Attention deficit hyperactivity disorder | 0.2 | 0.7 | -0.07 | <0.3 | 0.8 | -0.09 | 0.2 | 0.4 | -0.04 |
| Chronic liver disease | 0.5 | 0.3 | 0.04 | 0.3 | 0.2 | 0.02 | 0.5 | <0.1 | 0.07 |
| Chronic obstructive lung disease | 8.7 | 8.7 | 0.00 | 8.4 | 8.7 | -0.01 | 8.7 | 8.4 | 0.01 |
| Crohn’s disease | 0.2 | 0.2 | 0.00 | <0.3 | 0.2 | 0.00 | 0.2 | 0.3 | -0.02 |
| Dementia | 3.2 | 3.9 | -0.04 | 2.8 | 3.9 | -0.06 | 3.2 | 3.2 | 0.00 |
| Depressive disorder | 10.4 | 11.6 | -0.04 | 11.9 | 12.2 | -0.01 | 10.9 | 11.6 | -0.02 |
| Gastroesophageal reflux disease | 3.0 | 2.5 | 0.04 | 3.2 | 2.4 | 0.05 | 3.1 | 2.9 | 0.01 |
| Gastrointestinal hemorrhage | 1.0 | 1.3 | -0.03 | 0.8 | 1.0 | -0.03 | 0.9 | 0.7 | 0.03 |
| Hyperlipidemia | 30.4 | 34.4 | -0.09 | 33.5 | 33.6 | 0.00 | 31.1 | 32.2 | -0.02 |
| Hypertensive disorder | 54.7 | 64.4 | -0.20 | 61.7 | 60.9 | 0.01 | 56.4 | 56.0 | 0.01 |
| Lesion of liver | 0.8 | 0.8 | 0.00 | 0.8 | 0.7 | 0.01 | 0.8 | 0.7 | 0.02 |
| Obesity | 8.2 | 8.4 | -0.01 | 7.6 | 7.6 | 0.00 | 8.2 | 7.0 | 0.04 |
| Osteoarthritis | 14.1 | 16.9 | -0.08 | 16.1 | 15.6 | 0.01 | 14.6 | 14.8 | -0.01 |
| Pneumonia | 3.1 | 2.6 | 0.03 | 3.0 | 2.7 | 0.02 | 3.1 | 2.6 | 0.03 |
| Psoriasis | 1.4 | 1.6 | -0.02 | 1.1 | 1.7 | -0.05 | 1.4 | 1.7 | -0.02 |
| Renal impairment | 9.5 | 7.1 | 0.09 | 7.5 | 8.2 | -0.02 | 9.1 | 9.0 | 0.00 |
| Rheumatoid arthritis | 1.5 | 1.7 | -0.02 | 1.3 | 1.6 | -0.03 | 1.4 | 1.5 | -0.01 |
| Schizophrenia | 0.2 | 0.2 | -0.01 | 0.4 | 0.3 | 0.01 | 0.2 | 0.3 | -0.01 |
| Ulcerative colitis | 0.3 | 0.2 | 0.03 | 0.3 | <0.2 | 0.04 | 0.3 | 0.4 | -0.01 |
| Urinary tract infectious disease | 6.1 | 6.1 | 0.00 | 6.0 | 6.2 | -0.01 | 6.2 | 5.8 | 0.02 |
| Visual system disorder | 10.9 | 11.7 | -0.03 | 12.3 | 11.4 | 0.03 | 11.2 | 12.2 | -0.03 |
| **Medical history: Cardiovascular disease** | | | | | | | | | |
| Atrial fibrillation | 5.0 | 3.4 | 0.08 | 4.2 | 3.9 | 0.01 | 4.8 | 4.7 | 0.01 |
| Cerebrovascular disease | 9.6 | 10.7 | -0.04 | 10.0 | 9.6 | 0.02 | 9.8 | 10.4 | -0.02 |
| Coronary arteriosclerosis | 10.1 | 7.7 | 0.09 | 8.6 | 8.8 | -0.01 | 9.6 | 10.6 | -0.03 |
| Heart disease | 45.4 | 49.3 | -0.08 | 46.9 | 45.0 | 0.04 | 45.7 | 45.3 | 0.01 |
| Heart failure | 11.4 | 13.2 | -0.06 | 11.6 | 11.2 | 0.01 | 11.4 | 10.9 | 0.01 |
| Ischemic heart disease | 25.4 | 32.4 | -0.16 | 28.5 | 26.9 | 0.04 | 26.1 | 25.5 | 0.01 |
| Peripheral vascular disease | 12.5 | 12.4 | 0.00 | 12.3 | 12.8 | -0.02 | 12.4 | 12.2 | 0.00 |
| Pulmonary embolism | 0.9 | 0.5 | 0.05 | 0.9 | 0.6 | 0.03 | 0.9 | 1.3 | -0.04 |
| Venous thrombosis | 2.0 | 1.6 | 0.03 | 1.3 | 1.6 | -0.02 | 1.9 | 2.2 | -0.02 |
| **Medical history: Neoplasms** | | | | | | | | | |
| Hematologic neoplasm | 0.5 | 0.3 | 0.03 | 0.6 | 0.4 | 0.03 | 0.6 | 0.3 | 0.04 |
| Malignant lymphoma | 0.4 | 0.3 | 0.01 | 0.5 | 0.3 | 0.03 | 0.4 | <0.1 | 0.05 |
| Malignant neoplasm of anorectum | 0.2 | 0.2 | 0.01 | <0.3 | <0.2 | 0.03 | 0.3 | <0.1 | 0.05 |
| Malignant neoplastic disease | 6.8 | 7.4 | -0.02 | 6.3 | 7.3 | -0.04 | 6.8 | 6.2 | 0.03 |
| Malignant tumor of breast | 0.6 | 1.3 | -0.06 | 0.7 | 1.2 | -0.05 | 0.7 | 0.9 | -0.03 |
| Malignant tumor of colon | 0.5 | 0.4 | 0.01 | 0.4 | 0.3 | 0.00 | 0.5 | 0.2 | 0.06 |
| Malignant tumor of lung | 0.2 | 0.2 | 0.00 | <0.3 | 0.3 | -0.06 | 0.2 | 0.3 | -0.01 |
| Malignant tumor of urinary bladder | 0.3 | 0.3 | 0.00 | 0.3 | 0.3 | -0.01 | 0.3 | 0.2 | 0.02 |
| Primary malignant neoplasm of prostate | 1.1 | 1.4 | -0.04 | 0.9 | 1.4 | -0.05 | 1.1 | 1.5 | -0.04 |
| **Medication use** | | | | | | | | | |
| Agents acting on the renin-angiotensin system | 77.3 | 73.7 | 0.08 | 76.0 | 74.6 | 0.03 | 77.0 | 77.4 | -0.01 |
| Antibacterials for systemic use | 28.9 | 31.2 | -0.05 | 30.4 | 30.9 | -0.01 | 29.4 | 29.6 | 0.00 |
| Antidepressants | 12.5 | 11.4 | 0.03 | 11.5 | 11.6 | 0.00 | 12.3 | 11.5 | 0.02 |

| (Continued) | Before ajustment | | | After matching | | | After stratification | | |
|---|---|---|---|---|---|---|---|---|---|
| Characteristic | T (%) | C (%) | StdDiff | T (%) | C (%) | StdDiff | T (%) | C (%) | StdDiff |
| Antiepileptics | 6.6 | 4.4 | 0.10 | 4.0 | 4.9 | -0.05 | 6.2 | 5.6 | 0.02 |
| Antiinflammatory and antirheumatic products | 54.9 | 59.0 | -0.08 | 57.2 | 56.1 | 0.02 | 55.5 | 54.6 | 0.02 |
| Antineoplastic agents | 1.3 | 1.4 | -0.01 | 1.1 | 1.4 | -0.03 | 1.2 | 1.6 | -0.03 |
| Antipsoriatics | 0.6 | 0.3 | 0.03 | <0.3 | 0.2 | -0.01 | 0.5 | 0.2 | 0.06 |
| Antithrombotic agents | 53.4 | 50.9 | 0.05 | 49.4 | 49.4 | 0.00 | 52.6 | 52.9 | 0.00 |
| Beta blocking agents | 61.9 | 59.0 | 0.06 | 62.9 | 60.6 | 0.05 | 61.8 | 61.0 | 0.02 |
| Calcium channel blockers | 38.6 | 36.9 | 0.04 | 37.7 | 37.8 | 0.00 | 38.5 | 39.1 | -0.01 |
| Diuretics | 59.1 | 60.7 | -0.03 | 62.7 | 60.2 | 0.05 | 59.8 | 59.7 | 0.00 |
| Drugs for acid related disorders | 43.9 | 38.8 | 0.10 | 42.0 | 41.1 | 0.02 | 43.3 | 44.9 | -0.03 |
| Drugs for obstructive airway diseases | 20.5 | 19.2 | 0.03 | 18.8 | 19.0 | 0.00 | 20.1 | 19.1 | 0.03 |
| Immunosuppressants | 0.8 | 0.8 | 0.00 | 0.8 | 0.7 | 0.01 | 0.8 | 0.7 | 0.01 |
| Lipid modifying agents | 61.5 | 52.1 | 0.19 | 55.7 | 55.5 | 0.00 | 60.2 | 61.8 | -0.03 |
| Opioids | 14.9 | 15.8 | -0.02 | 14.1 | 15.5 | -0.04 | 15.0 | 15.7 | -0.02 |
| Psycholeptics | 11.8 | 13.4 | -0.05 | 12.3 | 12.2 | 0.00 | 12.1 | 12.4 | -0.01 |
| Psychostimulants | 0.3 | 0.7 | -0.06 | 0.5 | 0.2 | 0.04 | 0.3 | 0.2 | 0.04 |

**Supplementary Table 40:** Baseline patient characteristics for DPP4is (T) and SUs (C) new-users in the OptumDOD data source. We report proportion of initiators satisfying selected base-line characteristics and the standardized difference of population proportions (StdDiff) before and after propensity score adjustment. Less extreme StdDiffs through matching and stratification suggest improved balance between patient cohorts through adjustment.

| | Before adjustment | | | After matching | | | After stratification | | |
|---|---|---|---|---|---|---|---|---|---|
| Characteristic | T (%) | C (%) | StdDiff | T (%) | C (%) | StdDiff | T (%) | C (%) | StdDiff |
| **Age group** | | | | | | | | | |
| 25 - 29 | 0.1 | 0.1 | 0.00 | 0.1 | 0.1 | 0.00 | 0.1 | 0.1 | 0.00 |
| 30 - 34 | 0.3 | 0.2 | 0.01 | 0.3 | 0.2 | 0.01 | 0.3 | 0.2 | 0.01 |
| 35 - 39 | 0.6 | 0.6 | 0.01 | 0.7 | 0.5 | 0.02 | 0.7 | 0.5 | 0.02 |
| 40 - 44 | 1.6 | 1.3 | 0.02 | 1.6 | 1.4 | 0.01 | 1.5 | 1.4 | 0.01 |
| 45 - 49 | 3.6 | 3.0 | 0.04 | 3.6 | 3.5 | 0.00 | 3.2 | 3.2 | 0.00 |
| 50 - 54 | 6.6 | 5.6 | 0.04 | 6.4 | 6.5 | 0.00 | 5.9 | 5.9 | 0.00 |
| 55 - 59 | 10.1 | 8.9 | 0.04 | 9.9 | 10.0 | -0.01 | 9.2 | 9.2 | 0.00 |
| 60 - 64 | 12.7 | 11.7 | 0.03 | 12.5 | 12.5 | 0.00 | 12.0 | 12.0 | 0.00 |
| 65 - 69 | 16.6 | 17.8 | -0.03 | 16.8 | 16.7 | 0.00 | 17.6 | 17.4 | 0.00 |
| 70 - 74 | 18.9 | 20.9 | -0.05 | 19.1 | 19.9 | -0.02 | 19.5 | 20.5 | -0.03 |
| 75 - 79 | 14.2 | 15.6 | -0.04 | 14.4 | 14.2 | 0.00 | 15.2 | 15.1 | 0.00 |
| 80 - 84 | 9.1 | 9.5 | -0.01 | 9.2 | 9.0 | 0.01 | 9.6 | 9.3 | 0.01 |
| 85 - 89 | 5.1 | 4.6 | 0.02 | 4.8 | 4.7 | 0.00 | 4.8 | 4.7 | 0.00 |
| 90 - 94 | 0.4 | 0.3 | 0.02 | 0.4 | 0.3 | 0.02 | 0.5 | 0.3 | 0.02 |
| Gender: female | 47.3 | 44.2 | 0.06 | 46.7 | 47.2 | -0.01 | 45.1 | 45.3 | 0.00 |
| **Medical history: General** | | | | | | | | | |
| Acute respiratory disease | 29.2 | 28.1 | 0.03 | 28.9 | 28.8 | 0.00 | 28.7 | 28.4 | 0.01 |
| Attention deficit hyperactivity disorder | 0.6 | 0.4 | 0.02 | 0.5 | 0.5 | 0.00 | 0.5 | 0.4 | 0.00 |
| Chronic liver disease | 4.0 | 3.7 | 0.02 | 3.8 | 3.8 | 0.00 | 3.8 | 3.8 | 0.00 |
| Chronic obstructive lung disease | 15.5 | 17.2 | -0.04 | 15.8 | 15.6 | 0.00 | 16.9 | 16.7 | 0.01 |
| Crohn's disease | 0.4 | 0.3 | 0.02 | 0.4 | 0.3 | 0.02 | 0.4 | 0.3 | 0.02 |
| Dementia | 4.7 | 4.4 | 0.01 | 4.7 | 4.6 | 0.00 | 4.6 | 4.5 | 0.01 |
| Depressive disorder | 17.4 | 17.1 | 0.01 | 17.2 | 17.4 | 0.00 | 17.1 | 17.3 | 0.00 |
| Gastroesophageal reflux disease | 23.6 | 22.2 | 0.03 | 23.2 | 23.5 | -0.01 | 22.8 | 22.7 | 0.00 |
| Gastrointestinal hemorrhage | 3.8 | 4.0 | -0.01 | 3.8 | 3.9 | -0.01 | 3.8 | 3.9 | -0.01 |
| Human immunodeficiency virus infection | 0.3 | 0.2 | 0.02 | 0.3 | 0.3 | 0.01 | 0.3 | 0.2 | 0.01 |
| Hypertensive disorder | 88.1 | 87.4 | 0.02 | 87.9 | 87.7 | 0.00 | 87.9 | 87.6 | 0.01 |
| Lesion of liver | 1.5 | 1.9 | -0.03 | 1.5 | 1.6 | -0.01 | 1.7 | 1.8 | -0.01 |
| Obesity | 23.3 | 23.4 | 0.00 | 23.5 | 23.6 | 0.00 | 23.7 | 23.5 | 0.00 |
| Osteoarthritis | 35.9 | 34.4 | 0.03 | 35.5 | 35.6 | 0.00 | 35.1 | 34.9 | 0.00 |
| Pneumonia | 6.3 | 7.2 | -0.03 | 6.4 | 6.3 | 0.00 | 7.0 | 6.9 | 0.00 |
| Psoriasis | 1.8 | 1.6 | 0.01 | 1.7 | 1.7 | 0.00 | 1.7 | 1.6 | 0.01 |
| Renal impairment | 23.7 | 24.9 | -0.03 | 24.0 | 23.6 | 0.01 | 25.1 | 24.5 | 0.01 |
| Rheumatoid arthritis | 2.5 | 2.5 | 0.00 | 2.5 | 2.6 | -0.01 | 2.5 | 2.5 | 0.00 |
| Schizophrenia | 0.5 | 0.5 | 0.00 | 0.5 | 0.5 | 0.01 | 0.6 | 0.5 | 0.01 |
| Ulcerative colitis | 0.5 | 0.4 | 0.01 | 0.4 | 0.4 | 0.01 | 0.5 | 0.4 | 0.01 |
| Urinary tract infectious disease | 15.6 | 14.7 | 0.02 | 15.3 | 15.5 | 0.00 | 15.1 | 15.0 | 0.00 |
| Viral hepatitis C | 0.7 | 0.8 | -0.01 | 0.7 | 0.8 | -0.01 | 0.8 | 0.8 | 0.00 |
| Visual system disorder | 47.7 | 47.2 | 0.01 | 47.5 | 47.6 | 0.00 | 47.1 | 47.4 | 0.00 |
| **Medical history: Cardiovascular disease** | | | | | | | | | |
| Atrial fibrillation | 13.0 | 13.1 | 0.00 | 13.1 | 12.9 | 0.01 | 13.2 | 13.1 | 0.00 |
| Coronary arteriosclerosis | 38.4 | 40.0 | -0.03 | 38.6 | 37.9 | 0.01 | 39.9 | 39.3 | 0.01 |
| Heart disease | 59.4 | 60.6 | -0.02 | 59.4 | 58.7 | 0.01 | 60.7 | 60.0 | 0.01 |
| Heart failure | 14.7 | 16.0 | -0.04 | 14.8 | 14.5 | 0.01 | 15.9 | 15.5 | 0.01 |
| Ischemic heart disease | 20.5 | 22.2 | -0.04 | 20.8 | 20.5 | 0.01 | 21.9 | 21.7 | 0.01 |
| Peripheral vascular disease | 26.2 | 26.1 | 0.00 | 26.3 | 26.4 | 0.00 | 26.4 | 26.3 | 0.00 |
| Pulmonary embolism | 1.3 | 1.4 | -0.01 | 1.4 | 1.4 | 0.00 | 1.4 | 1.4 | 0.00 |
| Venous thrombosis | 2.5 | 2.8 | -0.02 | 2.6 | 2.6 | 0.00 | 2.7 | 2.8 | -0.01 |
| **Medical history: Neoplasms** | | | | | | | | | |
| Hematologic neoplasm | 1.6 | 2.0 | -0.03 | 1.6 | 1.7 | -0.01 | 1.8 | 1.9 | -0.01 |
| Malignant neoplasm of anorectum | 0.3 | 0.4 | -0.01 | 0.3 | 0.3 | 0.00 | 0.4 | 0.4 | 0.00 |
| Malignant neoplastic disease | 14.3 | 15.1 | -0.02 | 14.4 | 14.4 | 0.00 | 15.0 | 14.9 | 0.00 |
| Malignant tumor of breast | 2.2 | 2.1 | 0.00 | 2.2 | 2.3 | 0.00 | 2.2 | 2.2 | 0.00 |
| Malignant tumor of colon | 0.8 | 0.9 | -0.01 | 0.9 | 0.9 | 0.00 | 0.9 | 0.9 | 0.00 |
| Malignant tumor of lung | 0.7 | 0.9 | -0.03 | 0.7 | 0.7 | 0.00 | 0.9 | 0.9 | 0.00 |
| Malignant tumor of urinary bladder | 0.7 | 0.9 | -0.02 | 0.8 | 0.8 | 0.00 | 0.9 | 0.9 | 0.00 |
| Primary malignant neoplasm of prostate | 3.2 | 3.1 | 0.00 | 3.2 | 3.1 | 0.01 | 3.2 | 3.1 | 0.01 |
| **Medication use** | | | | | | | | | |
| Agents acting on the renin-angiotensin system | 75.8 | 75.8 | 0.00 | 75.7 | 75.8 | 0.00 | 75.9 | 75.8 | 0.00 |
| Antidepressants | 31.6 | 30.3 | 0.03 | 31.4 | 31.5 | 0.00 | 30.9 | 30.8 | 0.00 |
| Antiepileptics | 22.2 | 21.9 | 0.01 | 22.1 | 22.3 | -0.01 | 22.4 | 22.1 | 0.01 |

| (Continued) | Before ajustment | | | After matching | | | After stratification | | |
|---|---|---|---|---|---|---|---|---|---|
| Characteristic | T (%) | C (%) | StdDiff | T (%) | C (%) | StdDiff | T (%) | C (%) | StdDiff |
| Antiinflammatory and antirheumatic products | 32.3 | 30.4 | 0.04 | 31.7 | 32.0 | -0.01 | 30.7 | 31.0 | -0.01 |
| Antineoplastic agents | 7.2 | 6.6 | 0.02 | 6.9 | 7.1 | -0.01 | 6.8 | 6.8 | 0.00 |
| Antipsoriatics | 0.7 | 0.7 | 0.01 | 0.7 | 0.8 | -0.01 | 0.6 | 0.7 | -0.01 |
| Antithrombotic agents | 31.5 | 32.2 | -0.01 | 32.2 | 32.1 | 0.00 | 32.2 | 32.1 | 0.00 |
| Beta blocking agents | 52.3 | 54.6 | -0.05 | 52.5 | 52.0 | 0.01 | 54.1 | 53.7 | 0.01 |
| Calcium channel blockers | 33.6 | 34.2 | -0.01 | 33.6 | 33.8 | 0.00 | 34.1 | 34.1 | 0.00 |
| Diuretics | 49.6 | 50.8 | -0.02 | 49.6 | 49.3 | 0.01 | 50.8 | 50.4 | 0.01 |
| Drugs for acid related disorders | 36.2 | 35.0 | 0.03 | 35.7 | 36.1 | -0.01 | 35.7 | 35.5 | 0.00 |
| Drugs for obstructive airway diseases | 43.9 | 41.5 | 0.05 | 43.2 | 43.8 | -0.01 | 42.2 | 42.4 | 0.00 |
| Immunosuppressants | 3.3 | 2.8 | 0.03 | 3.1 | 3.1 | 0.00 | 3.1 | 2.9 | 0.01 |
| Lipid modifying agents | 82.6 | 81.1 | 0.04 | 82.4 | 82.5 | 0.00 | 81.3 | 81.6 | -0.01 |
| Opioids | 33.2 | 33.5 | -0.01 | 33.1 | 32.8 | 0.01 | 33.9 | 33.4 | 0.01 |
| Psycholeptics | 29.7 | 28.8 | 0.02 | 29.5 | 29.5 | 0.00 | 29.4 | 29.1 | 0.01 |
| Psychostimulants | 2.0 | 1.4 | 0.04 | 1.8 | 1.8 | 0.00 | 1.7 | 1.6 | 0.01 |

**Supplementary Table 41:** Baseline patient characteristics for DPP4is (T) and SUs (C) new-users in the MDCR data source. We report proportion of initiators satisfying selected base-line characteristics and the standardized difference of population proportions (StdDiff) before and after propensity score adjustment. Less extreme StdDiffs through matching and stratification suggest improved balance between patient cohorts through adjustment.

| | Before adjustment | | | After matching | | | After stratification | | |
|---|---|---|---|---|---|---|---|---|---|
| Characteristic | T (%) | C (%) | StdDiff | T (%) | C (%) | StdDiff | T (%) | C (%) | StdDiff |
| **Age group** | | | | | | | | | |
| 50 - 54 | 0.1 | 0.1 | 0.01 | 0.1 | 0.1 | 0.01 | 0.1 | 0.1 | 0.01 |
| 55 - 59 | 0.4 | 0.5 | -0.01 | 0.4 | 0.5 | -0.01 | 0.4 | 0.5 | -0.01 |
| 60 - 64 | 1.5 | 1.3 | 0.02 | 1.5 | 1.4 | 0.01 | 1.5 | 1.4 | 0.01 |
| 65 - 69 | 22.1 | 19.8 | 0.06 | 21.8 | 22.0 | -0.01 | 21.0 | 21.1 | 0.00 |
| 70 - 74 | 28.8 | 28.4 | 0.01 | 28.6 | 28.8 | 0.00 | 28.2 | 28.5 | -0.01 |
| 75 - 79 | 22.1 | 23.3 | -0.03 | 22.2 | 22.4 | 0.00 | 22.3 | 22.6 | -0.01 |
| 80 - 84 | 15.0 | 16.4 | -0.04 | 15.2 | 15.1 | 0.00 | 15.7 | 15.8 | 0.00 |
| 85 - 89 | 7.3 | 7.6 | -0.01 | 7.4 | 7.4 | 0.00 | 7.6 | 7.6 | 0.00 |
| 90 - 94 | 2.3 | 2.2 | 0.00 | 2.3 | 1.9 | 0.03 | 2.5 | 2.1 | 0.03 |
| 95 - 99 | 0.4 | 0.3 | 0.02 | 0.4 | 0.3 | 0.02 | 0.5 | 0.3 | 0.03 |
| Gender: female | 44.1 | 42.6 | 0.03 | 43.4 | 43.6 | 0.00 | 42.5 | 42.7 | 0.00 |
| **Medical history: General** | | | | | | | | | |
| Acute respiratory disease | 25.4 | 24.4 | 0.03 | 24.9 | 24.6 | 0.01 | 25.7 | 25.1 | 0.01 |
| Attention deficit hyperactivity disorder | 0.1 | 0.1 | 0.01 | 0.1 | 0.2 | -0.01 | 0.1 | 0.1 | 0.00 |
| Chronic liver disease | 2.7 | 2.3 | 0.03 | 2.7 | 2.6 | 0.01 | 2.6 | 2.5 | 0.00 |
| Chronic obstructive lung disease | 14.2 | 15.2 | -0.03 | 14.4 | 14.2 | 0.01 | 15.7 | 15.2 | 0.01 |
| Crohn's disease | 0.3 | 0.3 | 0.01 | 0.3 | 0.3 | 0.01 | 0.3 | 0.3 | 0.01 |
| Dementia | 4.1 | 4.5 | -0.02 | 4.3 | 4.3 | 0.00 | 4.6 | 4.6 | 0.00 |
| Depressive disorder | 8.6 | 8.2 | 0.02 | 8.7 | 9.0 | -0.01 | 9.1 | 9.2 | 0.00 |
| Gastroesophageal reflux disease | 14.6 | 11.8 | 0.08 | 14.0 | 14.5 | -0.02 | 13.3 | 13.9 | -0.02 |
| Gastrointestinal hemorrhage | 4.2 | 4.6 | -0.02 | 4.3 | 4.2 | 0.01 | 4.5 | 4.3 | 0.01 |
| Hyperlipidemia | 65.7 | 52.6 | 0.27 | 64.4 | 65.7 | -0.03 | 60.6 | 62.0 | -0.03 |
| Hypertensive disorder | 78.4 | 71.3 | 0.16 | 77.7 | 78.4 | -0.02 | 76.2 | 76.7 | -0.01 |
| Lesion of liver | 0.9 | 1.3 | -0.03 | 1.0 | 1.2 | -0.03 | 1.0 | 1.3 | -0.03 |
| Obesity | 11.0 | 8.2 | 0.10 | 10.6 | 11.2 | -0.02 | 9.9 | 10.5 | -0.02 |
| Osteoarthritis | 34.6 | 30.1 | 0.10 | 34.0 | 34.0 | 0.00 | 32.7 | 33.0 | -0.01 |
| Pneumonia | 6.6 | 8.3 | -0.06 | 6.8 | 6.3 | 0.02 | 7.9 | 7.5 | 0.01 |
| Psoriasis | 1.6 | 1.4 | 0.02 | 1.5 | 1.8 | -0.02 | 1.5 | 1.6 | -0.01 |
| Renal impairment | 17.8 | 16.4 | 0.04 | 17.9 | 17.7 | 0.00 | 18.7 | 18.3 | 0.01 |
| Rheumatoid arthritis | 2.0 | 1.8 | 0.01 | 2.0 | 2.1 | -0.01 | 2.0 | 2.1 | -0.01 |
| Schizophrenia | 0.2 | 0.2 | 0.00 | 0.2 | 0.1 | 0.00 | 0.2 | 0.2 | 0.00 |
| Ulcerative colitis | 0.6 | 0.4 | 0.03 | 0.6 | 0.4 | 0.03 | 0.6 | 0.5 | 0.03 |
| Urinary tract infectious disease | 12.3 | 12.3 | 0.00 | 12.2 | 12.3 | 0.00 | 12.5 | 12.6 | 0.00 |
| Viral hepatitis C | 0.3 | 0.3 | 0.00 | 0.2 | 0.3 | -0.01 | 0.2 | 0.3 | -0.01 |
| Visual system disorder | 54.7 | 50.1 | 0.09 | 53.8 | 54.5 | -0.01 | 52.0 | 52.5 | -0.01 |
| **Medical history: Cardiovascular disease** | | | | | | | | | |
| Atrial fibrillation | 16.7 | 16.4 | 0.01 | 16.8 | 16.3 | 0.01 | 17.6 | 16.8 | 0.02 |
| Cerebrovascular disease | 21.7 | 22.0 | -0.01 | 21.8 | 21.3 | 0.01 | 21.8 | 21.3 | 0.01 |
| Coronary arteriosclerosis | 43.7 | 44.3 | -0.01 | 43.4 | 44.2 | -0.01 | 44.0 | 43.8 | 0.00 |
| Heart disease | 66.6 | 66.9 | -0.01 | 66.0 | 66.4 | -0.01 | 66.7 | 66.4 | 0.01 |
| Heart failure | 14.5 | 16.6 | -0.06 | 14.7 | 14.1 | 0.01 | 16.4 | 15.3 | 0.03 |
| Ischemic heart disease | 19.4 | 21.8 | -0.06 | 19.4 | 19.5 | 0.00 | 20.3 | 20.2 | 0.00 |
| Peripheral vascular disease | 22.2 | 20.0 | 0.06 | 21.8 | 22.1 | -0.01 | 21.0 | 21.3 | -0.01 |
| Pulmonary embolism | 1.2 | 1.3 | 0.00 | 1.2 | 1.3 | -0.01 | 1.3 | 1.3 | 0.00 |
| Venous thrombosis | 2.7 | 3.0 | -0.02 | 2.7 | 3.0 | -0.02 | 2.9 | 3.2 | -0.01 |
| **Medical history: Neoplasms** | | | | | | | | | |
| Hematologic neoplasm | 2.1 | 2.1 | 0.00 | 2.1 | 2.2 | 0.00 | 2.2 | 2.3 | -0.01 |
| Malignant lymphoma | 1.1 | 1.0 | 0.01 | 1.1 | 1.1 | 0.00 | 1.2 | 1.1 | 0.01 |
| Malignant neoplasm of anorectum | 0.3 | 0.5 | -0.02 | 0.3 | 0.3 | 0.00 | 0.4 | 0.4 | 0.00 |
| Malignant neoplastic disease | 21.1 | 20.4 | 0.02 | 21.0 | 20.9 | 0.00 | 20.8 | 21.0 | 0.00 |
| Malignant tumor of breast | 2.8 | 2.6 | 0.01 | 2.8 | 2.8 | 0.00 | 2.6 | 2.7 | -0.01 |
| Malignant tumor of colon | 1.0 | 1.1 | 0.00 | 1.1 | 0.8 | 0.03 | 1.2 | 0.9 | 0.02 |
| Malignant tumor of lung | 0.8 | 1.1 | -0.03 | 0.9 | 0.9 | 0.00 | 1.0 | 1.0 | 0.00 |
| Malignant tumor of urinary bladder | 1.5 | 1.4 | 0.01 | 1.6 | 1.4 | 0.01 | 1.6 | 1.4 | 0.01 |
| Primary malignant neoplasm of prostate | 5.0 | 5.3 | -0.01 | 5.0 | 5.2 | -0.01 | 5.0 | 5.3 | -0.01 |
| **Medication use** | | | | | | | | | |
| Agents acting on the renin-angiotensin system | 77.1 | 74.3 | 0.06 | 77.0 | 76.6 | 0.01 | 76.9 | 76.3 | 0.01 |
| Antibacterials for systemic use | 66.1 | 65.0 | 0.02 | 65.4 | 65.3 | 0.00 | 65.7 | 65.7 | 0.00 |
| Antidepressants | 26.6 | 27.3 | -0.02 | 26.8 | 26.5 | 0.01 | 27.4 | 27.4 | 0.00 |
| Antiepileptics | 17.7 | 17.3 | 0.01 | 17.7 | 18.1 | -0.01 | 18.3 | 18.4 | 0.00 |
| Antiinflammatory and antirheumatic products | 29.5 | 27.6 | 0.04 | 28.9 | 29.0 | 0.00 | 27.9 | 28.2 | -0.01 |

| (Continued) | Before ajustment | | | After matching | | | After stratification | | |
|---|---|---|---|---|---|---|---|---|---|
| Characteristic | T (%) | C (%) | StdDiff | T (%) | C (%) | StdDiff | T (%) | C (%) | StdDiff |
| Antineoplastic agents | 9.3 | 9.6 | -0.01 | 9.3 | 9.1 | 0.01 | 9.2 | 9.0 | 0.01 |
| Antipsoriatics | 0.9 | 0.8 | 0.01 | 0.8 | 1.0 | -0.02 | 0.8 | 0.9 | -0.01 |
| Antithrombotic agents | 37.2 | 38.5 | -0.03 | 37.5 | 37.1 | 0.01 | 38.9 | 38.2 | 0.02 |
| Beta blocking agents | 60.9 | 61.0 | 0.00 | 61.0 | 60.7 | 0.01 | 61.9 | 61.6 | 0.00 |
| Calcium channel blockers | 38.4 | 37.1 | 0.03 | 38.4 | 37.9 | 0.01 | 38.1 | 37.7 | 0.01 |
| Diuretics | 54.1 | 56.7 | -0.05 | 54.4 | 53.5 | 0.02 | 55.9 | 55.1 | 0.02 |
| Drugs for acid related disorders | 37.7 | 36.7 | 0.02 | 36.8 | 36.9 | 0.00 | 36.9 | 36.9 | 0.00 |
| Drugs for obstructive airway diseases | 44.4 | 41.9 | 0.05 | 44.1 | 44.0 | 0.00 | 44.0 | 44.0 | 0.00 |
| Immunosuppressants | 3.5 | 2.9 | 0.04 | 3.5 | 3.4 | 0.00 | 3.5 | 3.3 | 0.01 |
| Lipid modifying agents | 84.6 | 80.2 | 0.12 | 84.1 | 84.2 | 0.00 | 83.0 | 83.3 | -0.01 |
| Opioids | 32.1 | 32.4 | -0.01 | 32.0 | 31.7 | 0.01 | 33.0 | 32.5 | 0.01 |
| Psycholeptics | 29.3 | 29.0 | 0.01 | 29.2 | 28.9 | 0.01 | 29.5 | 29.3 | 0.00 |
| Psychostimulants | 1.5 | 1.3 | 0.02 | 1.4 | 1.4 | 0.01 | 1.5 | 1.4 | 0.01 |

**Supplementary Table 42:** Baseline patient characteristics for DPP4is (T) and SUs (C) new-users in the MDCD data source. We report proportion of initiators satisfying selected base-line characteristics and the standardized difference of population proportions (StdDiff) before and after propensity score adjustment. Less extreme StdDiffs through matching and stratification suggest improved balance between patient cohorts through adjustment.

| | Before adjustment | | | After matching | | | After stratification | | |
|---|---|---|---|---|---|---|---|---|---|
| Characteristic | T (%) | C (%) | StdDiff | T (%) | C (%) | StdDiff | T (%) | C (%) | StdDiff |
| **Age group** | | | | | | | | | |
| 20 - 24 | 0.2 | 0.3 | -0.03 | 0.1 | 0.2 | -0.02 | 0.1 | 0.3 | -0.03 |
| 25 - 29 | 0.5 | 0.7 | -0.02 | 0.5 | 0.6 | -0.01 | 0.7 | 0.7 | 0.00 |
| 30 - 34 | 1.6 | 1.9 | -0.02 | 1.6 | 1.8 | -0.02 | 1.7 | 1.8 | -0.01 |
| 35 - 39 | 4.2 | 4.1 | 0.00 | 4.3 | 3.9 | 0.02 | 4.3 | 4.0 | 0.01 |
| 40 - 44 | 7.0 | 6.8 | 0.01 | 7.1 | 6.8 | 0.01 | 6.9 | 6.8 | 0.00 |
| 45 - 49 | 12.1 | 11.9 | 0.01 | 12.1 | 11.6 | 0.01 | 12.2 | 11.8 | 0.01 |
| 50 - 54 | 16.3 | 17.0 | -0.02 | 16.3 | 17.0 | -0.02 | 16.6 | 17.1 | -0.01 |
| 55 - 59 | 21.2 | 21.4 | 0.00 | 21.2 | 21.1 | 0.00 | 21.5 | 21.4 | 0.00 |
| 60 - 64 | 19.4 | 20.2 | -0.02 | 19.5 | 19.7 | 0.00 | 19.6 | 19.9 | -0.01 |
| 70 - 74 | 4.2 | 3.9 | 0.02 | 4.2 | 4.4 | -0.01 | 3.8 | 4.1 | -0.01 |
| 75 - 79 | 3.2 | 2.4 | 0.05 | 3.1 | 2.5 | 0.04 | 2.9 | 2.4 | 0.03 |
| 80 - 84 | 2.2 | 1.6 | 0.04 | 2.2 | 1.9 | 0.02 | 2.1 | 1.7 | 0.03 |
| 85 - 89 | 1.1 | 0.9 | 0.01 | 1.1 | 1.0 | 0.01 | 1.0 | 1.0 | 0.00 |
| 90 - 94 | <0.1 | 0.1 | 0.00 | <0.1 | 0.1 | 0.01 | <0.1 | 0.1 | 0.00 |
| Gender: female | 61.2 | 58.4 | 0.06 | 60.7 | 60.5 | 0.00 | 59.0 | 59.2 | 0.00 |
| **Race** | | | | | | | | | |
| Black or African American | 21.9 | 25.5 | -0.08 | 22.3 | 22.1 | 0.01 | 24.3 | 24.4 | 0.00 |
| White | 62.5 | 58.0 | 0.09 | 62.0 | 63.0 | -0.02 | 58.8 | 59.7 | -0.02 |
| **Ethnicity** | | | | | | | | | |
| Hispanic or Latino | 2.0 | 2.1 | -0.01 | 2.0 | 2.2 | -0.01 | 1.9 | 2.1 | -0.01 |
| **Medical history: General** | | | | | | | | | |
| Acute respiratory disease | 39.8 | 40.7 | -0.02 | 39.5 | 40.1 | -0.01 | 40.0 | 40.6 | -0.01 |
| Attention deficit hyperactivity disorder | 1.2 | 1.2 | 0.00 | 1.1 | 1.5 | -0.03 | 1.1 | 1.3 | -0.02 |
| Chronic liver disease | 5.8 | 7.0 | -0.05 | 5.8 | 6.1 | -0.02 | 6.4 | 6.8 | -0.01 |
| Chronic obstructive lung disease | 32.3 | 31.2 | 0.02 | 32.0 | 31.8 | 0.00 | 31.9 | 31.5 | 0.01 |
| Crohn’s disease | 0.3 | 0.4 | -0.01 | 0.3 | 0.3 | 0.00 | 0.3 | 0.4 | -0.01 |
| Dementia | 3.9 | 3.2 | 0.04 | 3.7 | 3.5 | 0.01 | 3.7 | 3.3 | 0.02 |
| Depressive disorder | 36.9 | 36.0 | 0.02 | 36.5 | 37.7 | -0.02 | 35.5 | 36.6 | -0.02 |
| Gastroesophageal reflux disease | 32.2 | 30.7 | 0.03 | 31.7 | 32.8 | -0.02 | 30.3 | 31.5 | -0.03 |
| Gastrointestinal hemorrhage | 4.9 | 5.2 | -0.01 | 4.8 | 4.7 | 0.00 | 4.8 | 5.1 | -0.01 |
| Human immunodeficiency virus infection | 0.7 | 0.6 | 0.01 | 0.6 | 0.4 | 0.03 | 0.7 | 0.5 | 0.03 |
| Hyperlipidemia | 75.6 | 72.4 | 0.07 | 74.9 | 75.4 | -0.01 | 74.9 | 75.4 | -0.01 |
| Hypertensive disorder | 87.1 | 86.4 | 0.02 | 87.0 | 86.4 | 0.02 | 87.1 | 86.4 | 0.02 |
| Lesion of liver | 1.8 | 2.5 | -0.04 | 1.8 | 2.1 | -0.02 | 2.2 | 2.4 | -0.01 |
| Obesity | 36.3 | 35.4 | 0.02 | 36.1 | 36.4 | -0.01 | 35.7 | 35.9 | 0.00 |
| Osteoarthritis | 44.9 | 42.7 | 0.04 | 44.3 | 45.2 | -0.02 | 43.6 | 43.7 | 0.00 |
| Pneumonia | 10.0 | 10.4 | -0.01 | 9.9 | 9.4 | 0.02 | 10.7 | 10.1 | 0.02 |
| Psoriasis | 1.6 | 1.3 | 0.02 | 1.5 | 1.6 | 0.00 | 1.6 | 1.4 | 0.02 |
| Renal impairment | 15.2 | 15.7 | -0.01 | 15.3 | 15.4 | 0.00 | 15.3 | 15.5 | -0.01 |
| Rheumatoid arthritis | 2.7 | 2.5 | 0.01 | 2.6 | 2.3 | 0.02 | 2.6 | 2.5 | 0.01 |
| Schizophrenia | 5.0 | 5.3 | -0.01 | 5.0 | 4.9 | 0.00 | 5.4 | 5.1 | 0.01 |
| Ulcerative colitis | 0.2 | 0.3 | -0.01 | 0.3 | 0.3 | 0.00 | 0.2 | 0.3 | -0.01 |
| Urinary tract infectious disease | 14.5 | 15.4 | -0.03 | 14.2 | 15.0 | -0.02 | 14.7 | 15.3 | -0.02 |
| Viral hepatitis C | 2.6 | 3.5 | -0.05 | 2.7 | 2.9 | -0.02 | 3.1 | 3.3 | -0.01 |
| Visual system disorder | 46.6 | 43.6 | 0.06 | 46.1 | 47.8 | -0.03 | 43.0 | 45.1 | -0.04 |
| **Medical history: Cardiovascular disease** | | | | | | | | | |
| Atrial fibrillation | 7.5 | 7.6 | -0.01 | 7.5 | 7.3 | 0.01 | 7.6 | 7.5 | 0.00 |
| Cerebrovascular disease | 13.5 | 14.6 | -0.03 | 13.5 | 13.8 | -0.01 | 14.0 | 14.4 | -0.01 |
| Coronary arteriosclerosis | 34.0 | 33.1 | 0.02 | 33.9 | 33.6 | 0.01 | 33.4 | 33.2 | 0.00 |
| Heart disease | 56.9 | 57.8 | -0.02 | 56.9 | 57.1 | 0.00 | 57.3 | 57.5 | 0.00 |
| Heart failure | 17.7 | 19.3 | -0.04 | 17.8 | 18.2 | -0.01 | 18.8 | 19.0 | 0.00 |
| Ischemic heart disease | 23.2 | 23.8 | -0.01 | 23.1 | 23.2 | 0.00 | 23.8 | 23.6 | 0.00 |
| Peripheral vascular disease | 22.7 | 21.4 | 0.03 | 22.1 | 21.8 | 0.01 | 22.1 | 21.8 | 0.01 |
| Pulmonary embolism | 1.9 | 2.0 | 0.00 | 1.9 | 1.9 | 0.00 | 1.9 | 1.9 | 0.00 |
| Venous thrombosis | 3.0 | 3.2 | -0.01 | 2.9 | 3.1 | -0.01 | 3.1 | 3.2 | 0.00 |
| **Medical history: Neoplasms** | | | | | | | | | |
| Hematologic neoplasm | 1.3 | 1.4 | -0.01 | 1.2 | 1.3 | -0.01 | 1.2 | 1.3 | -0.01 |
| Malignant lymphoma | 0.7 | 0.5 | 0.02 | 0.7 | 0.4 | 0.03 | 0.7 | 0.5 | 0.03 |
| Malignant neoplasm of anorectum | 0.2 | 0.4 | -0.02 | 0.3 | 0.3 | -0.01 | 0.2 | 0.3 | -0.02 |
| Malignant neoplastic disease | 8.4 | 8.4 | 0.00 | 8.5 | 8.4 | 0.00 | 8.1 | 8.4 | -0.01 |
| Malignant tumor of breast | 2.0 | 1.7 | 0.02 | 2.1 | 1.9 | 0.01 | 2.1 | 1.9 | 0.01 |

| (Continued) | Before ajustment | | | After matching | | | After stratification | | |
|---|---|---|---|---|---|---|---|---|---|
| Characteristic | T (%) | C (%) | StdDiff | T (%) | C (%) | StdDiff | T (%) | C (%) | StdDiff |
| Malignant tumor of colon | 0.6 | 0.6 | 0.00 | 0.6 | 0.6 | 0.00 | 0.6 | 0.6 | -0.01 |
| Malignant tumor of lung | 0.7 | 0.7 | 0.00 | 0.7 | 0.6 | 0.01 | 0.8 | 0.7 | 0.01 |
| Malignant tumor of urinary bladder | 0.2 | 0.3 | -0.02 | 0.2 | 0.3 | -0.02 | 0.3 | 0.3 | -0.01 |
| Primary malignant neoplasm of prostate | 0.8 | 0.7 | 0.01 | 0.8 | 0.7 | 0.01 | 0.9 | 0.7 | 0.02 |
| **Medication use** | | | | | | | | | |
| Agents acting on the renin-angiotensin system | 71.2 | 71.0 | 0.00 | 71.2 | 70.8 | 0.01 | 71.2 | 71.0 | 0.01 |
| Antibacterials for systemic use | 71.2 | 70.6 | 0.01 | 70.7 | 71.2 | -0.01 | 70.4 | 70.9 | -0.01 |
| Antidepressants | 58.0 | 55.0 | 0.06 | 57.2 | 58.3 | -0.02 | 55.4 | 56.2 | -0.02 |
| Antiepileptics | 48.8 | 45.8 | 0.06 | 48.0 | 48.6 | -0.01 | 47.0 | 46.8 | 0.00 |
| Antiinflammatory and antirheumatic products | 59.9 | 58.8 | 0.02 | 59.7 | 60.2 | -0.01 | 59.6 | 59.3 | 0.00 |
| Antineoplastic agents | 5.5 | 4.9 | 0.03 | 5.5 | 5.3 | 0.01 | 5.3 | 5.0 | 0.01 |
| Antipsoriatics | 0.7 | 0.6 | 0.01 | 0.6 | 0.8 | -0.02 | 0.6 | 0.7 | -0.01 |
| Antithrombotic agents | 44.8 | 43.2 | 0.03 | 45.0 | 44.9 | 0.00 | 44.1 | 43.7 | 0.01 |
| Beta blocking agents | 51.4 | 52.6 | -0.03 | 51.6 | 51.8 | 0.00 | 51.5 | 52.4 | -0.02 |
| Calcium channel blockers | 32.5 | 33.7 | -0.02 | 32.4 | 32.8 | -0.01 | 32.4 | 32.8 | -0.01 |
| Diuretics | 54.6 | 56.8 | -0.04 | 54.9 | 54.1 | 0.01 | 56.4 | 55.9 | 0.01 |
| Drugs for acid related disorders | 58.2 | 54.8 | 0.07 | 57.4 | 58.4 | -0.02 | 54.9 | 56.2 | -0.03 |
| Drugs for obstructive airway diseases | 63.5 | 60.9 | 0.05 | 62.8 | 63.8 | -0.02 | 61.6 | 62.0 | -0.01 |
| Immunosuppressants | 3.2 | 2.6 | 0.04 | 3.1 | 2.8 | 0.02 | 3.0 | 2.7 | 0.02 |
| Lipid modifying agents | 78.5 | 74.9 | 0.09 | 77.9 | 78.9 | -0.02 | 75.8 | 76.4 | -0.01 |
| Opioids | 53.5 | 54.6 | -0.02 | 53.4 | 52.7 | 0.01 | 54.0 | 54.1 | 0.00 |
| Psycholeptics | 54.1 | 53.0 | 0.02 | 53.7 | 54.2 | -0.01 | 53.2 | 53.5 | -0.01 |
| Psychostimulants | 5.1 | 4.0 | 0.05 | 5.0 | 4.5 | 0.03 | 4.9 | 4.2 | 0.03 |

**Supplementary Table 43:** Baseline patient characteristics for DPP4is (T) and SUs (C) new-users in the IMRD data source. We report proportion of initiators satisfying selected base-line characteristics and the standardized difference of population proportions (StdDiff) before and after propensity score adjustment. Less extreme StdDiffs through matching and stratification suggest improved balance between patient cohorts through adjustment.

| | Before adjustment | | | After matching | | | After stratification | | |
|---|---|---|---|---|---|---|---|---|---|
| Characteristic | T (%) | C (%) | StdDiff | T (%) | C (%) | StdDiff | T (%) | C (%) | StdDiff |
| **Age group** | | | | | | | | | |
| 40 - 44 | 0.9 | 1.2 | -0.02 | <1.2 | 1.0 | -0.03 | 0.8 | 2.7 | -0.14 |
| 45 - 49 | 2.6 | 2.0 | 0.04 | 3.0 | 1.6 | 0.09 | 2.7 | 0.9 | 0.13 |
| 50 - 54 | 5.7 | 5.8 | 0.00 | 6.4 | 7.2 | -0.03 | 6.1 | 5.0 | 0.05 |
| 55 - 59 | 9.9 | 9.7 | 0.01 | 11.1 | 9.6 | 0.05 | 10.2 | 10.0 | 0.01 |
| 60 - 64 | 14.0 | 14.0 | 0.00 | 13.5 | 13.3 | 0.01 | 13.7 | 14.8 | -0.03 |
| 65 - 69 | 16.9 | 19.4 | -0.07 | 20.2 | 16.5 | 0.10 | 17.8 | 22.6 | -0.12 |
| 70 - 74 | 17.1 | 19.6 | -0.06 | 18.0 | 19.3 | -0.03 | 17.1 | 20.6 | -0.09 |
| 75 - 79 | 15.4 | 14.2 | 0.03 | 12.3 | 15.7 | -0.10 | 14.6 | 10.2 | 0.13 |
| 80 - 84 | 10.5 | 9.3 | 0.04 | 8.9 | 9.6 | -0.03 | 10.6 | 6.8 | 0.14 |
| 85 - 89 | 4.9 | 3.5 | 0.07 | 3.9 | 4.3 | -0.02 | 4.3 | 5.8 | -0.07 |
| 90 - 94 | 1.7 | 1.2 | 0.04 | 1.5 | 1.9 | -0.03 | 1.6 | 0.6 | 0.10 |
| Gender: female | 31.5 | 34.8 | -0.07 | 36.7 | 35.7 | 0.02 | 32.2 | 34.7 | -0.05 |
| **Race** | | | | | | | | | |
| White | 28.6 | 32.7 | -0.09 | 38.4 | 36.2 | 0.05 | 31.5 | 34.1 | -0.06 |
| Asian Indian | 0.4 | 0.8 | -0.05 | <1.2 | 0.5 | 0.03 | 0.4 | <0.2 | 0.04 |
| Chinese | <0.1 | <0.3 | -0.01 | <1.2 | <0.3 | 0.06 | <0.1 | <0.2 | 0.02 |
| Pakistani | 0.2 | <0.3 | 0.01 | 0.2 | 0.9 | -0.09 | 0.2 | 0.9 | -0.09 |
| Black | 0.2 | <0.3 | -0.01 | 0.2 | 0.9 | -0.10 | 0.2 | 0.9 | -0.10 |
| European | 0.1 | <0.3 | 0.00 | <0.1 | <0.2 | 0.02 | <0.1 | <0.2 | 0.02 |
| Middle Eastern or North African | <0.1 | <0.3 | -0.01 | <0.1 | 0.9 | -0.12 | <0.1 | 0.9 | -0.12 |
| **Medical history: General** | | | | | | | | | |
| Acute respiratory disease | 6.3 | 7.6 | -0.05 | 10.1 | 6.8 | 0.12 | 6.9 | 4.0 | 0.13 |
| Chronic obstructive lung disease | 3.4 | 2.2 | 0.07 | 3.7 | 2.2 | 0.09 | 3.4 | 1.6 | 0.12 |
| Dementia | 0.6 | <0.3 | 0.06 | <1.2 | <0.3 | 0.03 | 0.6 | <0.2 | 0.08 |
| Depressive disorder | 1.5 | 2.7 | -0.08 | 1.3 | 0.7 | 0.06 | 1.3 | 0.7 | 0.06 |
| Gastroesophageal reflux disease | 0.3 | 0.4 | -0.01 | <1.2 | <0.3 | 0.03 | 0.5 | <0.2 | 0.08 |
| Gastrointestinal hemorrhage | 0.7 | 1.3 | -0.06 | <1.2 | 1.3 | -0.06 | 0.6 | 0.8 | -0.02 |
| Hyperlipidemia | 0.6 | 2.1 | -0.13 | <1.2 | <0.3 | 0.10 | 0.6 | <0.2 | 0.07 |
| Hypertensive disorder | 1.8 | 4.0 | -0.13 | 2.5 | 2.6 | -0.01 | 1.9 | 5.0 | -0.17 |
| Obesity | 0.4 | 1.3 | -0.10 | 2.0 | 0.4 | 0.15 | 0.8 | <0.2 | 0.08 |
| Osteoarthritis | 2.5 | 3.9 | -0.08 | 3.0 | 2.6 | 0.02 | 2.5 | 1.3 | 0.09 |
| Pneumonia | 0.6 | 0.4 | 0.03 | <1.2 | <0.3 | 0.03 | 0.7 | <0.2 | 0.09 |
| Psoriasis | 0.6 | 0.4 | 0.03 | <1.2 | 0.4 | 0.01 | 0.7 | <0.2 | 0.09 |
| Renal impairment | 5.6 | 6.0 | -0.02 | 5.2 | 6.6 | -0.06 | 5.3 | 4.3 | 0.05 |
| Ulcerative colitis | <0.1 | <0.3 | -0.05 | <0.1 | 2.3 | -0.21 | <0.1 | 2.3 | -0.21 |
| Urinary tract infectious disease | 2.5 | 2.4 | 0.01 | 3.0 | 2.6 | 0.02 | 2.7 | 1.2 | 0.11 |
| Visual system disorder | 12.0 | 12.4 | -0.01 | 13.3 | 13.0 | 0.01 | 11.9 | 20.5 | -0.24 |
| **Medical history: Cardiovascular disease** | | | | | | | | | |
| Atrial fibrillation | 2.2 | 2.6 | -0.02 | 1.2 | 2.4 | -0.09 | 1.9 | 1.4 | 0.04 |
| Cerebrovascular disease | 3.3 | 3.7 | -0.02 | 4.2 | 3.2 | 0.05 | 3.2 | 1.6 | 0.10 |
| Coronary arteriosclerosis | 0.8 | 1.0 | -0.02 | <1.2 | 1.1 | -0.06 | 0.7 | 2.5 | -0.15 |
| Heart disease | 9.8 | 15.1 | -0.16 | 7.4 | 9.3 | -0.07 | 9.3 | 8.7 | 0.02 |
| Heart failure | 1.9 | 2.9 | -0.07 | <1.2 | 1.2 | -0.05 | 1.6 | 0.8 | 0.08 |
| Ischemic heart disease | 5.4 | 10.4 | -0.19 | 4.4 | 5.8 | -0.06 | 5.2 | 6.0 | -0.04 |
| Peripheral vascular disease | 0.7 | 1.5 | -0.07 | <1.2 | 0.6 | 0.02 | 0.7 | 0.5 | 0.02 |
| Pulmonary embolism | <0.1 | <0.3 | 0.01 | <0.1 | <0.2 | 0.03 | <0.1 | <0.2 | 0.03 |
| Venous thrombosis | 0.4 | 0.6 | -0.03 | 0.3 | <0.2 | 0.02 | 0.3 | <0.2 | 0.02 |
| **Medical history: Neoplasms** | | | | | | | | | |
| Malignant lymphoma | <0.1 | <0.3 | -0.01 | <0.1 | <0.2 | 0.01 | <0.1 | <0.2 | 0.01 |
| Malignant neoplastic disease | 1.8 | 2.1 | -0.02 | 1.5 | 1.8 | -0.03 | 1.8 | 1.2 | 0.05 |
| Malignant tumor of breast | <0.1 | <0.3 | -0.01 | <1.2 | <0.3 | 0.06 | <0.1 | <0.2 | 0.03 |
| Malignant tumor of colon | <0.1 | <0.3 | -0.01 | <0.1 | <0.2 | 0.01 | <0.1 | <0.2 | 0.01 |
| Malignant tumor of lung | 0.1 | <0.3 | 0.01 | 0.1 | 0.3 | -0.04 | 0.1 | 0.3 | -0.04 |
| Malignant tumor of urinary bladder | 0.1 | <0.3 | 0.00 | <1.2 | <0.3 | 0.07 | <0.1 | <0.2 | 0.03 |
| **Medication use** | | | | | | | | | |
| Agents acting on the renin-angiotensin system | 77.2 | 68.5 | 0.20 | 75.6 | 77.2 | -0.04 | 76.5 | 75.1 | 0.03 |
| Antibacterials for systemic use | 53.2 | 50.8 | 0.05 | 53.4 | 51.7 | 0.04 | 53.3 | 51.9 | 0.03 |
| Antidepressants | 27.7 | 21.9 | 0.13 | 29.1 | 27.2 | 0.04 | 27.0 | 24.4 | 0.06 |
| Antiepileptics | 10.7 | 6.7 | 0.14 | 8.6 | 8.3 | 0.01 | 9.9 | 6.9 | 0.11 |
| Antiinflammatory and antirheumatic products | 25.1 | 27.6 | -0.06 | 23.4 | 25.4 | -0.05 | 24.7 | 20.3 | 0.10 |
| Antineoplastic agents | 1.5 | 1.0 | 0.05 | <1.2 | 0.8 | 0.01 | 1.4 | 0.4 | 0.10 |

| (Continued) | Before ajustment | | | After matching | | | After stratification | | |
|---|---|---|---|---|---|---|---|---|---|
| Characteristic | T (%) | C (%) | StdDiff | T (%) | C (%) | StdDiff | T (%) | C (%) | StdDiff |
| Antipsoriatics | 1.6 | 1.8 | -0.01 | 2.5 | 2.5 | 0.00 | 1.8 | 2.0 | -0.01 |
| Antithrombotic agents | 90.3 | 88.1 | 0.07 | 89.2 | 89.2 | 0.00 | 89.9 | 89.4 | 0.02 |
| Beta blocking agents | 58.1 | 51.5 | 0.13 | 54.4 | 56.5 | -0.04 | 58.2 | 54.5 | 0.07 |
| Calcium channel blockers | 40.6 | 40.9 | 0.00 | 42.9 | 39.4 | 0.07 | 41.1 | 37.4 | 0.07 |
| Diuretics | 41.8 | 47.6 | -0.12 | 47.5 | 45.8 | 0.04 | 42.7 | 35.8 | 0.14 |
| Drugs for acid related disorders | 56.1 | 44.5 | 0.23 | 54.7 | 53.7 | 0.02 | 55.7 | 55.9 | 0.00 |
| Drugs for obstructive airway diseases | 25.2 | 21.5 | 0.09 | 26.8 | 24.2 | 0.06 | 24.9 | 26.6 | -0.04 |
| Immunosuppressants | 1.6 | 1.2 | 0.04 | 1.7 | 1.6 | 0.01 | 1.5 | 3.1 | -0.11 |
| Lipid modifying agents | 93.4 | 84.2 | 0.29 | 94.8 | 94.5 | 0.01 | 93.7 | 93.9 | 0.00 |
| Opioids | 18.9 | 24.2 | -0.13 | 20.4 | 21.6 | -0.03 | 19.1 | 21.0 | -0.05 |
| Psycholeptics | 16.5 | 17.8 | -0.04 | 17.0 | 16.3 | 0.02 | 16.6 | 20.9 | -0.11 |

**Supplementary Table 44:** Baseline patient characteristics for DPP4is (T) and SUs (C) new-users in the Open Claims data source. We report proportion of initiators satisfying selected base-line characteristics and the standardized difference of population proportions (StdDiff) before and after propensity score adjustment. Less extreme StdDiffs through matching and stratification suggest improved balance between patient cohorts through adjustment.

| | Before adjustment | | | After matching | | | After stratification | | |
|---|---|---|---|---|---|---|---|---|---|
| Characteristic | T (%) | C (%) | StdDiff | T (%) | C (%) | StdDiff | T (%) | C (%) | StdDiff |
| **Age group** | | | | | | | | | |
| 20 - 24 | 0.1 | 0.1 | 0.00 | 0.1 | 0.1 | 0.00 | 0.1 | 0.1 | 0.00 |
| 25 - 29 | 0.2 | 0.2 | 0.00 | 0.2 | 0.2 | 0.00 | 0.2 | 0.2 | 0.00 |
| 30 - 34 | 0.4 | 0.4 | 0.00 | 0.4 | 0.4 | 0.00 | 0.4 | 0.4 | 0.00 |
| 35 - 39 | 0.9 | 0.9 | 0.00 | 0.9 | 0.9 | 0.01 | 0.9 | 0.9 | 0.01 |
| 40 - 44 | 2.0 | 2.0 | 0.00 | 2.0 | 1.9 | 0.01 | 2.0 | 2.0 | 0.00 |
| 45 - 49 | 4.2 | 4.1 | 0.01 | 4.2 | 4.2 | 0.00 | 4.1 | 4.1 | 0.00 |
| 50 - 54 | 7.7 | 7.3 | 0.01 | 7.7 | 7.7 | 0.00 | 7.4 | 7.5 | -0.01 |
| 55 - 59 | 11.8 | 11.3 | 0.02 | 11.8 | 11.9 | 0.00 | 11.5 | 11.4 | 0.00 |
| 60 - 64 | 14.9 | 14.2 | 0.02 | 14.4 | 14.3 | 0.00 | 14.4 | 14.3 | 0.00 |
| 65 - 69 | 16.6 | 17.3 | -0.02 | 16.6 | 16.7 | 0.00 | 17.0 | 17.1 | 0.00 |
| 70 - 74 | 15.5 | 16.4 | -0.02 | 15.6 | 15.3 | 0.01 | 16.3 | 16.1 | 0.01 |
| 75 - 79 | 17.4 | 18.1 | -0.02 | 17.4 | 17.6 | 0.00 | 17.7 | 17.9 | 0.00 |
| 80 - 84 | 8.0 | 7.5 | 0.02 | 7.9 | 8.0 | 0.00 | 7.6 | 7.7 | 0.00 |
| 85 - 89 | 0.2 | 0.2 | 0.00 | 0.2 | 0.1 | 0.01 | 0.3 | 0.2 | 0.01 |
| Gender: female | 48.6 | 44.9 | 0.07 | 48.1 | 48.4 | 0.00 | 48.1 | 48.4 | 0.00 |
| **Medical history: General** | | | | | | | | | |
| Acute respiratory disease | 17.7 | 16.8 | 0.02 | 17.5 | 17.6 | 0.00 | 17.3 | 17.2 | 0.00 |
| Attention deficit hyperactivity disorder | 0.3 | 0.3 | 0.00 | 0.3 | 0.3 | 0.00 | 0.3 | 0.3 | 0.00 |
| Chronic liver disease | 1.9 | 1.8 | 0.01 | 1.9 | 1.9 | 0.00 | 1.9 | 1.9 | 0.00 |
| Chronic obstructive lung disease | 10.2 | 11.1 | -0.03 | 10.2 | 10.3 | 0.00 | 10.8 | 10.8 | 0.00 |
| Crohn’s disease | 0.2 | 0.2 | 0.00 | 0.2 | 0.2 | 0.00 | 0.2 | 0.2 | -0.01 |
| Dementia | 2.2 | 2.0 | 0.02 | 2.2 | 2.2 | 0.00 | 2.1 | 2.1 | 0.00 |
| Depressive disorder | 8.8 | 8.8 | 0.00 | 9.0 | 8.8 | 0.00 | 9.0 | 8.8 | 0.00 |
| Gastroesophageal reflux disease | 11.4 | 10.4 | 0.03 | 11.2 | 11.4 | 0.00 | 10.9 | 10.8 | 0.00 |
| Gastrointestinal hemorrhage | 2.4 | 2.3 | 0.01 | 2.4 | 2.4 | -0.01 | 2.4 | 2.4 | 0.00 |
| Human immunodeficiency virus infection | 0.3 | 0.2 | 0.01 | 0.3 | 0.3 | 0.01 | 0.3 | 0.3 | 0.01 |
| Hyperlipidemia | 54.4 | 50.1 | 0.09 | 53.9 | 54.2 | -0.01 | 51.4 | 51.6 | 0.00 |
| Hypertensive disorder | 63.9 | 61.3 | 0.05 | 63.5 | 63.8 | -0.01 | 62.2 | 62.3 | 0.00 |
| Lesion of liver | 1.0 | 1.1 | -0.01 | 1.0 | 1.0 | 0.00 | 1.0 | 1.1 | 0.00 |
| Obesity | 11.4 | 11.2 | 0.01 | 11.3 | 11.4 | 0.00 | 11.3 | 11.4 | 0.00 |
| Osteoarthritis | 22.0 | 20.4 | 0.04 | 21.5 | 21.7 | 0.00 | 21.1 | 21.1 | 0.00 |
| Pneumonia | 4.2 | 4.6 | -0.02 | 4.2 | 4.3 | 0.00 | 4.5 | 4.5 | 0.00 |
| Psoriasis | 1.0 | 0.9 | 0.01 | 1.0 | 1.0 | 0.00 | 1.0 | 1.0 | 0.00 |
| Renal impairment | 12.9 | 13.3 | -0.01 | 12.9 | 13.1 | -0.01 | 13.3 | 13.3 | 0.00 |
| Rheumatoid arthritis | 1.6 | 1.4 | 0.02 | 1.6 | 1.6 | 0.00 | 1.5 | 1.5 | 0.00 |
| Schizophrenia | 0.5 | 0.5 | 0.00 | 0.5 | 0.5 | 0.00 | 0.5 | 0.5 | 0.00 |
| Ulcerative colitis | 0.3 | 0.2 | 0.01 | 0.3 | 0.2 | 0.01 | 0.3 | 0.2 | 0.01 |
| Urinary tract infectious disease | 8.7 | 7.9 | 0.03 | 8.4 | 8.4 | 0.00 | 8.2 | 8.1 | 0.01 |
| Viral hepatitis C | 0.5 | 0.5 | 0.00 | 0.5 | 0.5 | 0.00 | 0.5 | 0.5 | 0.00 |
| Visual system disorder | 24.9 | 23.4 | 0.04 | 24.1 | 23.9 | 0.00 | 24.1 | 23.9 | 0.00 |
| **Medical history: Cardiovascular disease** | | | | | | | | | |
| Atrial fibrillation | 8.8 | 8.9 | 0.00 | 8.8 | 8.9 | 0.00 | 9.0 | 8.8 | 0.00 |
| Cerebrovascular disease | 10.1 | 9.5 | 0.02 | 10.1 | 10.1 | 0.00 | 9.9 | 9.9 | 0.00 |
| Coronary arteriosclerosis | 25.3 | 25.4 | 0.00 | 25.3 | 25.4 | 0.00 | 25.3 | 25.4 | 0.00 |
| Heart disease | 42.6 | 42.3 | 0.00 | 42.4 | 42.5 | 0.00 | 42.6 | 42.4 | 0.00 |
| Heart failure | 9.6 | 10.2 | -0.02 | 9.6 | 9.6 | 0.00 | 10.1 | 9.9 | 0.01 |
| Ischemic heart disease | 12.1 | 12.5 | -0.01 | 12.1 | 12.2 | 0.00 | 12.5 | 12.4 | 0.00 |
| Peripheral vascular disease | 13.9 | 14.1 | 0.00 | 13.9 | 13.6 | 0.01 | 14.1 | 14.0 | 0.00 |
| Pulmonary embolism | 0.9 | 0.9 | 0.00 | 0.9 | 1.0 | 0.00 | 0.9 | 1.0 | 0.00 |
| Venous thrombosis | 1.6 | 1.6 | 0.00 | 1.6 | 1.6 | 0.00 | 1.6 | 1.6 | 0.00 |
| **Medical history: Neoplasms** | | | | | | | | | |
| Hematologic neoplasm | 1.1 | 1.2 | -0.01 | 1.1 | 1.1 | 0.00 | 1.2 | 1.1 | 0.00 |
| Malignant lymphoma | 0.6 | 0.6 | -0.01 | 0.6 | 0.6 | 0.00 | 0.6 | 0.6 | 0.00 |
| Malignant neoplasm of anorectum | 0.2 | 0.2 | 0.00 | 0.2 | 0.2 | 0.00 | 0.2 | 0.2 | 0.00 |
| Malignant neoplastic disease | 9.3 | 9.4 | 0.00 | 9.4 | 9.3 | 0.00 | 9.6 | 9.5 | 0.00 |
| Malignant tumor of breast | 1.5 | 1.4 | 0.01 | 1.6 | 1.5 | 0.00 | 1.5 | 1.4 | 0.00 |
| Malignant tumor of colon | 0.5 | 0.5 | 0.00 | 0.5 | 0.5 | 0.00 | 0.5 | 0.5 | 0.00 |
| Malignant tumor of lung | 0.5 | 0.6 | -0.01 | 0.5 | 0.5 | -0.01 | 0.5 | 0.5 | -0.01 |
| Primary malignant neoplasm of prostate | 1.9 | 1.9 | 0.00 | 1.9 | 1.9 | 0.00 | 1.9 | 1.9 | 0.00 |
| **Medication use** | | | | | | | | | |
| Agents acting on the renin-angiotensin system | 73.7 | 73.2 | 0.01 | 73.6 | 73.4 | 0.00 | 73.5 | 73.4 | 0.00 |

| (Continued) | Before ajustment | | | After matching | | | After stratification | | |
|---|---|---|---|---|---|---|---|---|---|
| Characteristic | T (%) | C (%) | StdDiff | T (%) | C (%) | StdDiff | T (%) | C (%) | StdDiff |
| Antibacterials for systemic use | 62.5 | 60.6 | 0.04 | 62.2 | 62.4 | 0.00 | 61.4 | 61.2 | 0.00 |
| Antidepressants | 32.9 | 32.5 | 0.01 | 32.6 | 33.1 | -0.01 | 32.6 | 33.1 | -0.01 |
| Antiepileptics | 25.3 | 25.2 | 0.00 | 25.1 | 25.0 | 0.00 | 25.1 | 25.0 | 0.00 |
| Antiinflammatory and antirheumatic products | 40.2 | 37.8 | 0.05 | 39.7 | 39.6 | 0.00 | 38.9 | 38.5 | 0.01 |
| Antineoplastic agents | 6.3 | 5.4 | 0.04 | 6.1 | 6.0 | 0.00 | 5.9 | 5.6 | 0.01 |
| Antipsoriatics | 0.9 | 0.8 | 0.02 | 0.9 | 0.9 | 0.00 | 0.8 | 0.8 | 0.00 |
| Antithrombotic agents | 37.2 | 36.5 | 0.01 | 37.0 | 36.9 | 0.00 | 37.2 | 36.6 | 0.01 |
| Beta blocking agents | 53.2 | 54.3 | -0.02 | 53.2 | 53.0 | 0.00 | 54.5 | 53.8 | 0.01 |
| Calcium channel blockers | 34.0 | 33.6 | 0.01 | 33.9 | 33.7 | 0.00 | 34.1 | 33.5 | 0.01 |
| Diuretics | 47.8 | 48.5 | -0.01 | 47.8 | 47.5 | 0.01 | 48.7 | 48.2 | 0.01 |
| Drugs for acid related disorders | 43.8 | 40.9 | 0.06 | 43.2 | 43.4 | 0.00 | 42.3 | 42.0 | 0.01 |
| Drugs for obstructive airway diseases | 43.9 | 40.7 | 0.06 | 43.3 | 43.7 | -0.01 | 42.0 | 42.1 | 0.00 |
| Immunosuppressants | 3.7 | 2.9 | 0.05 | 3.5 | 3.5 | 0.00 | 3.3 | 3.1 | 0.01 |
| Lipid modifying agents | 79.9 | 77.8 | 0.05 | 78.5 | 78.4 | 0.00 | 78.5 | 78.4 | 0.00 |
| Opioids | 31.6 | 31.4 | 0.00 | 31.4 | 31.3 | 0.00 | 31.8 | 31.5 | 0.01 |
| Psycholeptics | 28.6 | 27.9 | 0.02 | 28.3 | 28.6 | -0.01 | 28.3 | 28.2 | 0.00 |
| Psychostimulants | 2.5 | 2.2 | 0.02 | 2.4 | 2.5 | -0.01 | 2.2 | 2.3 | -0.01 |

**Supplementary Table 45:** Baseline patient characteristics for DPP4is (T) and SUs (C) new-users in the OptumEHR data source. We report proportion of initiators satisfying selected base-line characteristics and the standardized difference of population proportions (StdDiff) before and after propensity score adjustment. Less extreme StdDiffs through matching and stratification suggest improved balance between patient cohorts through adjustment.

| | Before adjustment | | | After matching | | | After stratification | | |
|---|---|---|---|---|---|---|---|---|---|
| Characteristic | T (%) | C (%) | StdDiff | T (%) | C (%) | StdDiff | T (%) | C (%) | StdDiff |
| **Age group** | | | | | | | | | |
| 25 - 29 | 0.1 | 0.1 | 0.00 | 0.1 | 0.1 | -0.01 | 0.1 | 0.1 | -0.01 |
| 30 - 34 | 0.3 | 0.3 | -0.01 | 0.3 | 0.3 | -0.01 | 0.3 | 0.3 | -0.01 |
| 35 - 39 | 1.0 | 0.9 | 0.01 | 1.0 | 1.0 | 0.00 | 1.1 | 0.9 | 0.01 |
| 40 - 44 | 2.2 | 2.0 | 0.01 | 2.3 | 2.1 | 0.01 | 2.2 | 2.0 | 0.01 |
| 45 - 49 | 5.1 | 4.6 | 0.02 | 5.1 | 5.0 | 0.01 | 4.8 | 4.7 | 0.00 |
| 50 - 54 | 8.9 | 8.2 | 0.02 | 8.8 | 8.8 | 0.00 | 8.4 | 8.4 | 0.00 |
| 55 - 59 | 13.5 | 12.6 | 0.03 | 13.6 | 13.5 | 0.00 | 12.9 | 12.9 | 0.00 |
| 60 - 64 | 16.4 | 15.6 | 0.02 | 16.3 | 16.5 | -0.01 | 15.8 | 15.8 | 0.00 |
| 65 - 69 | 16.7 | 16.3 | 0.01 | 16.5 | 17.0 | -0.01 | 16.2 | 16.5 | -0.01 |
| 70 - 74 | 14.3 | 15.4 | -0.03 | 14.4 | 14.4 | 0.00 | 14.4 | 14.4 | 0.00 |
| 75 - 79 | 11.6 | 13.2 | -0.05 | 11.7 | 11.5 | 0.01 | 12.6 | 12.7 | 0.00 |
| 80 - 84 | 7.6 | 8.7 | -0.04 | 7.7 | 7.6 | 0.00 | 7.7 | 7.6 | 0.00 |
| 85 - 89 | 2.3 | 2.0 | 0.02 | 2.3 | 2.2 | 0.01 | 2.2 | 2.0 | 0.01 |
| Gender: female | 43.2 | 39.3 | 0.08 | 42.6 | 43.2 | -0.01 | 40.4 | 40.6 | -0.01 |
| **Race** | | | | | | | | | |
| Asian | 1.9 | 1.7 | 0.02 | 1.9 | 1.9 | 0.00 | 1.7 | 1.8 | 0.00 |
| Black or African American | 9.5 | 8.0 | 0.05 | 9.4 | 9.5 | -0.01 | 8.4 | 8.5 | 0.00 |
| White | 82.3 | 84.5 | -0.06 | 82.6 | 82.5 | 0.00 | 84.0 | 83.8 | 0.01 |
| **Ethnicity** | | | | | | | | | |
| Hispanic or Latino | 4.8 | 4.9 | 0.00 | 4.8 | 4.9 | -0.01 | 4.6 | 4.9 | -0.01 |
| Not Hispanic or Latino | 87.8 | 88.4 | -0.02 | 88.1 | 88.1 | 0.00 | 88.6 | 88.3 | 0.01 |
| **Medical history: General** | | | | | | | | | |
| Acute respiratory disease | 20.0 | 18.1 | 0.05 | 19.5 | 19.8 | -0.01 | 18.8 | 18.7 | 0.00 |
| Attention deficit hyperactivity disorder | 0.4 | 0.4 | 0.00 | 0.4 | 0.4 | -0.01 | 0.4 | 0.4 | -0.01 |
| Chronic liver disease | 3.1 | 3.1 | 0.00 | 3.0 | 3.0 | 0.00 | 3.1 | 3.1 | 0.00 |
| Chronic obstructive lung disease | 11.5 | 12.1 | -0.02 | 11.4 | 11.3 | 0.00 | 12.0 | 11.9 | 0.00 |
| Crohn's disease | 0.3 | 0.2 | 0.00 | 0.3 | 0.2 | 0.00 | 0.3 | 0.2 | 0.00 |
| Dementia | 2.1 | 2.2 | -0.01 | 2.1 | 2.0 | 0.01 | 2.4 | 2.1 | 0.02 |
| Depressive disorder | 15.0 | 15.1 | 0.00 | 15.0 | 15.1 | 0.00 | 15.0 | 15.2 | -0.01 |
| Gastroesophageal reflux disease | 21.8 | 19.8 | 0.05 | 21.0 | 21.3 | -0.01 | 20.7 | 20.4 | 0.01 |
| Gastrointestinal hemorrhage | 2.2 | 2.2 | 0.00 | 2.2 | 2.1 | 0.00 | 2.1 | 2.2 | -0.01 |
| Human immunodeficiency virus infection | 0.2 | 0.2 | 0.01 | 0.2 | 0.2 | 0.00 | 0.2 | 0.2 | 0.00 |
| Hyperlipidemia | 77.3 | 73.9 | 0.08 | 76.5 | 77.1 | -0.02 | 74.6 | 75.0 | -0.01 |
| Hypertensive disorder | 79.2 | 76.8 | 0.06 | 78.7 | 79.1 | -0.01 | 77.4 | 77.6 | 0.00 |
| Lesion of liver | 1.0 | 1.2 | -0.01 | 1.0 | 1.1 | 0.00 | 1.1 | 1.2 | 0.00 |
| Obesity | 25.8 | 23.7 | 0.05 | 25.5 | 25.6 | 0.00 | 24.5 | 24.4 | 0.00 |
| Osteoarthritis | 22.6 | 22.2 | 0.01 | 22.3 | 22.4 | 0.00 | 22.4 | 22.4 | 0.00 |
| Pneumonia | 3.1 | 3.8 | -0.04 | 3.2 | 3.3 | 0.00 | 3.7 | 3.7 | 0.00 |
| Psoriasis | 1.5 | 1.5 | 0.00 | 1.5 | 1.5 | 0.00 | 1.4 | 1.5 | -0.01 |
| Rheumatoid arthritis | 1.5 | 1.5 | 0.00 | 1.4 | 1.5 | 0.00 | 1.4 | 1.5 | -0.01 |
| Schizophrenia | 0.3 | 0.3 | 0.00 | 0.3 | 0.3 | 0.00 | 0.4 | 0.3 | 0.01 |
| Ulcerative colitis | 0.3 | 0.3 | 0.00 | 0.3 | 0.3 | -0.01 | 0.3 | 0.3 | 0.00 |
| Urinary tract infectious disease | 6.9 | 6.7 | 0.00 | 6.8 | 6.8 | 0.00 | 6.9 | 6.8 | 0.01 |
| Viral hepatitis C | 0.6 | 0.7 | -0.01 | 0.6 | 0.6 | 0.00 | 0.6 | 0.7 | -0.01 |
| Visual system disorder | 15.2 | 18.0 | -0.07 | 15.4 | 15.1 | 0.01 | 17.2 | 17.1 | 0.00 |
| **Medical history: Cardiovascular disease** | | | | | | | | | |
| Atrial fibrillation | 10.7 | 10.7 | 0.00 | 10.7 | 10.5 | 0.01 | 10.9 | 10.7 | 0.01 |
| Cerebrovascular disease | 11.4 | 11.5 | 0.00 | 11.2 | 11.4 | -0.01 | 11.4 | 11.5 | 0.00 |
| Coronary arteriosclerosis | 42.6 | 42.8 | 0.00 | 42.4 | 42.6 | 0.00 | 42.4 | 42.8 | -0.01 |
| Heart disease | 59.5 | 59.1 | 0.01 | 59.2 | 59.2 | 0.00 | 59.2 | 59.1 | 0.00 |
| Heart failure | 9.2 | 10.3 | -0.04 | 9.1 | 9.3 | -0.01 | 10.2 | 10.0 | 0.00 |
| Ischemic heart disease | 19.2 | 19.6 | -0.01 | 19.1 | 19.1 | 0.00 | 19.5 | 19.5 | 0.00 |
| Peripheral vascular disease | 17.7 | 17.9 | 0.00 | 17.5 | 17.5 | 0.00 | 17.9 | 17.8 | 0.00 |
| Pulmonary embolism | 0.9 | 1.1 | -0.02 | 0.9 | 1.0 | -0.01 | 0.9 | 1.1 | -0.01 |
| Venous thrombosis | 1.9 | 1.9 | 0.00 | 1.9 | 1.7 | 0.02 | 2.1 | 1.9 | 0.02 |
| **Medical history: Neoplasms** | | | | | | | | | |
| Hematologic neoplasm | 1.2 | 1.2 | 0.00 | 1.1 | 1.2 | 0.00 | 1.2 | 1.2 | 0.00 |
| Malignant lymphoma | 0.7 | 0.7 | 0.00 | 0.7 | 0.7 | 0.00 | 0.7 | 0.7 | 0.00 |
| Malignant neoplasm of anorectum | 0.1 | 0.2 | -0.01 | 0.2 | 0.2 | 0.00 | 0.2 | 0.2 | 0.00 |
| Malignant neoplastic disease | 9.5 | 9.9 | -0.01 | 9.4 | 9.5 | 0.00 | 9.6 | 9.8 | -0.01 |
| Malignant tumor of breast | 1.5 | 1.3 | 0.02 | 1.5 | 1.5 | 0.00 | 1.4 | 1.4 | 0.00 |

| (Continued) | Before ajustment | | | After matching | | | After stratification | | |
|---|---|---|---|---|---|---|---|---|---|
| Characteristic | T (%) | C (%) | StdDiff | T (%) | C (%) | StdDiff | T (%) | C (%) | StdDiff |
| Malignant tumor of colon | 0.4 | 0.6 | -0.02 | 0.4 | 0.5 | -0.01 | 0.5 | 0.5 | -0.01 |
| Malignant tumor of lung | 0.5 | 0.5 | 0.00 | 0.4 | 0.5 | 0.00 | 0.4 | 0.5 | 0.00 |
| Malignant tumor of urinary bladder | 0.5 | 0.6 | -0.01 | 0.5 | 0.5 | 0.01 | 0.6 | 0.6 | 0.01 |
| Primary malignant neoplasm of prostate | 2.0 | 2.2 | -0.02 | 2.0 | 2.1 | -0.01 | 2.1 | 2.2 | -0.01 |
| **Medication use** | | | | | | | | | |
| Agents acting on the renin-angiotensin system | 73.7 | 73.0 | 0.02 | 73.5 | 73.4 | 0.00 | 73.5 | 73.2 | 0.01 |
| Antibacterials for systemic use | 47.0 | 46.2 | 0.02 | 46.4 | 46.8 | -0.01 | 46.6 | 46.5 | 0.00 |
| Antidepressants | 32.8 | 32.0 | 0.02 | 32.4 | 32.3 | 0.00 | 32.4 | 32.3 | 0.00 |
| Antiepileptics | 22.1 | 22.2 | 0.00 | 22.0 | 22.1 | 0.00 | 22.4 | 22.2 | 0.00 |
| Antiinflammatory and antirheumatic products | 69.1 | 69.5 | -0.01 | 68.9 | 68.8 | 0.00 | 69.4 | 69.3 | 0.00 |
| Antineoplastic agents | 5.2 | 4.8 | 0.02 | 5.1 | 5.1 | 0.00 | 5.0 | 4.9 | 0.00 |
| Antipsoriatics | 0.8 | 0.6 | 0.02 | 0.8 | 0.6 | 0.02 | 0.8 | 0.6 | 0.02 |
| Antithrombotic agents | 67.1 | 68.9 | -0.04 | 67.1 | 66.5 | 0.01 | 68.8 | 68.1 | 0.01 |
| Beta blocking agents | 57.5 | 59.5 | -0.04 | 57.5 | 57.1 | 0.01 | 59.3 | 58.7 | 0.01 |
| Calcium channel blockers | 31.7 | 30.4 | 0.03 | 31.2 | 31.7 | -0.01 | 30.9 | 30.9 | 0.00 |
| Diuretics | 47.8 | 48.4 | -0.01 | 47.6 | 47.4 | 0.00 | 48.6 | 48.2 | 0.01 |
| Drugs for acid related disorders | 48.1 | 47.8 | 0.01 | 47.6 | 47.9 | -0.01 | 47.6 | 47.9 | -0.01 |
| Drugs for obstructive airway diseases | 47.3 | 46.1 | 0.02 | 46.5 | 46.9 | -0.01 | 46.7 | 46.5 | 0.00 |
| Immunosuppressants | 2.9 | 2.8 | 0.00 | 2.8 | 2.9 | 0.00 | 2.8 | 2.9 | 0.00 |
| Lipid modifying agents | 83.6 | 82.4 | 0.03 | 82.6 | 82.7 | 0.00 | 82.6 | 82.7 | 0.00 |
| Opioids | 34.8 | 36.2 | -0.03 | 34.8 | 34.7 | 0.00 | 36.2 | 35.9 | 0.01 |
| Psycholeptics | 35.6 | 34.4 | 0.03 | 35.1 | 35.3 | 0.00 | 35.0 | 34.8 | 0.00 |
| Psychostimulants | 2.5 | 2.3 | 0.01 | 2.5 | 2.5 | 0.00 | 2.3 | 2.4 | -0.01 |

**Supplementary Table 46:** Baseline patient characteristics for DPP4is (T) and SUs (C) new-users in the SIDIAP data source. We report proportion of initiators satisfying selected base-line characteristics and the standardized difference of population proportions (StdDiff) before and after propensity score adjustment. Less extreme StdDiffs through matching and stratification suggest improved balance between patient cohorts through adjustment.

| | Before adjustment | | | After matching | | | After stratification | | |
|---|---|---|---|---|---|---|---|---|---|
| Characteristic | T (%) | C (%) | StdDiff | T (%) | C (%) | StdDiff | T (%) | C (%) | StdDiff |
| **Age group** | | | | | | | | | |
| 30 - 34 | <0.1 | <0.2 | 0.00 | <0.3 | <0.2 | 0.02 | <0.1 | <0.1 | 0.01 |
| 35 - 39 | 0.1 | 0.2 | -0.02 | <0.3 | 0.3 | -0.03 | 0.1 | 0.2 | 0.00 |
| 40 - 44 | 0.4 | 1.1 | -0.07 | 0.8 | 1.1 | -0.03 | 0.5 | 0.9 | -0.04 |
| 45 - 49 | 1.9 | 2.5 | -0.04 | 3.6 | 2.8 | 0.04 | 2.2 | 2.6 | -0.02 |
| 50 - 54 | 4.0 | 6.2 | -0.10 | 5.7 | 6.1 | -0.01 | 4.5 | 4.3 | 0.01 |
| 55 - 59 | 7.8 | 10.9 | -0.11 | 10.5 | 11.2 | -0.02 | 8.7 | 8.7 | 0.00 |
| 60 - 64 | 12.9 | 15.2 | -0.07 | 15.4 | 14.5 | 0.02 | 13.4 | 13.6 | -0.01 |
| 65 - 69 | 14.9 | 16.3 | -0.04 | 16.2 | 16.3 | 0.00 | 15.4 | 16.9 | -0.04 |
| 70 - 74 | 16.4 | 16.2 | 0.00 | 15.2 | 16.1 | -0.02 | 16.3 | 18.4 | -0.06 |
| 75 - 79 | 15.7 | 13.9 | 0.05 | 14.5 | 13.6 | 0.03 | 15.1 | 13.2 | 0.06 |
| 80 - 84 | 13.2 | 10.5 | 0.08 | 11.2 | 11.0 | 0.00 | 12.5 | 11.5 | 0.03 |
| 85 - 89 | 9.0 | 5.6 | 0.13 | 5.0 | 5.4 | -0.02 | 8.1 | 8.3 | -0.01 |
| 90 - 94 | 3.1 | 1.0 | 0.14 | 1.6 | 1.3 | 0.02 | 2.7 | 1.4 | 0.10 |
| 95 - 99 | 0.5 | 0.2 | 0.04 | <0.3 | 0.2 | -0.03 | 0.4 | <0.1 | 0.06 |
| Gender: female | 28.4 | 28.8 | -0.01 | 27.4 | 28.3 | -0.02 | 28.5 | 29.2 | -0.02 |
| **Medical history: General** | | | | | | | | | |
| Acute respiratory disease | 13.9 | 11.5 | 0.07 | 12.0 | 12.6 | -0.02 | 13.5 | 13.8 | -0.01 |
| Chronic liver disease | 0.3 | 0.6 | -0.04 | 0.6 | 0.6 | 0.00 | 0.4 | 0.3 | 0.02 |
| Chronic obstructive lung disease | 2.4 | 2.9 | -0.03 | 2.5 | 2.9 | -0.02 | 2.3 | 1.9 | 0.03 |
| Crohn's disease | <0.1 | <0.2 | -0.02 | <0.1 | <0.1 | -0.01 | <0.1 | <0.1 | -0.01 |
| Dementia | 1.0 | 1.3 | -0.03 | 1.2 | 1.3 | -0.01 | 1.0 | 1.0 | 0.00 |
| Depressive disorder | 2.6 | 3.7 | -0.07 | 3.6 | 3.9 | -0.02 | 2.8 | 3.5 | -0.04 |
| Gastroesophageal reflux disease | 1.3 | 0.8 | 0.05 | 0.6 | 1.0 | -0.05 | 1.2 | 2.0 | -0.06 |
| Gastrointestinal hemorrhage | 2.1 | 1.4 | 0.06 | 1.9 | 1.5 | 0.04 | 2.0 | 1.1 | 0.07 |
| Hyperlipidemia | 2.7 | 4.0 | -0.08 | 4.3 | 3.7 | 0.04 | 3.1 | 2.4 | 0.04 |
| Hypertensive disorder | 3.7 | 6.0 | -0.10 | 5.6 | 5.3 | 0.01 | 4.2 | 5.5 | -0.06 |
| Lesion of liver | 0.4 | 0.6 | -0.03 | 0.4 | 0.7 | -0.04 | 0.4 | 0.5 | -0.02 |
| Obesity | 6.7 | 7.9 | -0.05 | 7.6 | 8.1 | -0.02 | 7.0 | 7.6 | -0.03 |
| Osteoarthritis | 5.3 | 5.4 | -0.01 | 6.8 | 5.5 | 0.05 | 5.5 | 4.6 | 0.04 |
| Pneumonia | 2.3 | 2.2 | 0.01 | 2.2 | 2.2 | 0.00 | 2.4 | 3.0 | -0.04 |
| Psoriasis | 0.3 | 0.7 | -0.06 | 0.4 | 0.7 | -0.04 | 0.3 | 0.8 | -0.06 |
| Renal impairment | 6.8 | 3.2 | 0.17 | 3.3 | 3.6 | -0.02 | 5.9 | 5.1 | 0.04 |
| Rheumatoid arthritis | 0.1 | <0.2 | -0.01 | <0.3 | <0.2 | -0.01 | 0.1 | <0.1 | 0.00 |
| Schizophrenia | <0.1 | <0.2 | -0.02 | <0.3 | <0.2 | -0.02 | <0.1 | <0.1 | 0.00 |
| Ulcerative colitis | <0.1 | <0.2 | -0.04 | <0.1 | <0.1 | -0.03 | <0.1 | <0.1 | -0.03 |
| Urinary tract infectious disease | 7.5 | 3.9 | 0.16 | 4.7 | 4.7 | 0.00 | 6.8 | 6.2 | 0.02 |
| Viral hepatitis C | 0.1 | <0.2 | 0.00 | <0.3 | 0.2 | -0.01 | 0.2 | <0.1 | 0.03 |
| Visual system disorder | 17.8 | 17.0 | 0.02 | 17.6 | 19.0 | -0.04 | 17.6 | 19.0 | -0.04 |
| **Medical history: Cardiovascular disease** | | | | | | | | | |
| Atrial fibrillation | 2.9 | 3.1 | -0.01 | 2.2 | 2.9 | -0.04 | 2.7 | 2.4 | 0.02 |
| Cerebrovascular disease | 6.4 | 8.3 | -0.07 | 8.2 | 8.3 | 0.00 | 6.8 | 6.4 | 0.02 |
| Coronary arteriosclerosis | <0.1 | <0.2 | -0.03 | <0.1 | <0.1 | -0.01 | <0.1 | <0.1 | -0.01 |
| Heart disease | 24.0 | 27.7 | -0.09 | 29.4 | 27.2 | 0.05 | 24.8 | 25.1 | -0.01 |
| Heart failure | 4.3 | 3.9 | 0.02 | 4.2 | 4.3 | 0.00 | 4.1 | 3.5 | 0.03 |
| Ischemic heart disease | 14.7 | 19.3 | -0.12 | 21.3 | 18.5 | 0.07 | 15.8 | 14.3 | 0.04 |
| Peripheral vascular disease | 5.2 | 6.5 | -0.06 | 5.4 | 6.1 | -0.03 | 5.4 | 5.2 | 0.01 |
| Pulmonary embolism | 0.2 | 0.2 | -0.01 | <0.3 | 0.2 | -0.04 | 0.2 | 0.2 | 0.00 |
| Venous thrombosis | 1.1 | 1.1 | 0.00 | 1.2 | 1.2 | 0.00 | 1.2 | 1.2 | 0.00 |
| **Medical history: Neoplasms** | | | | | | | | | |
| Hematologic neoplasm | 0.2 | <0.2 | 0.02 | <0.3 | <0.2 | 0.01 | 0.1 | <0.1 | 0.04 |
| Malignant lymphoma | 0.1 | <0.2 | -0.01 | 0.1 | <0.1 | 0.00 | 0.1 | <0.1 | 0.00 |
| Malignant neoplasm of anorectum | 0.1 | <0.2 | 0.03 | 0.1 | <0.1 | 0.03 | 0.1 | <0.1 | 0.03 |
| Malignant neoplastic disease | 3.9 | 2.8 | 0.06 | 3.9 | 3.0 | 0.05 | 3.9 | 5.4 | -0.07 |
| Malignant tumor of breast | 0.1 | 0.2 | -0.01 | <0.3 | 0.3 | -0.02 | 0.2 | <0.1 | 0.00 |
| Malignant tumor of colon | 0.5 | 0.4 | 0.01 | 0.5 | 0.4 | 0.01 | 0.5 | 0.3 | 0.04 |
| Malignant tumor of lung | 0.1 | <0.2 | 0.01 | <0.1 | <0.1 | 0.01 | <0.1 | <0.1 | 0.01 |
| Malignant tumor of urinary bladder | 0.5 | 0.5 | 0.01 | 0.4 | 0.3 | 0.02 | 0.6 | 1.9 | -0.12 |
| Primary malignant neoplasm of prostate | 0.4 | 0.3 | 0.01 | 0.7 | 0.3 | 0.05 | 0.4 | 1.1 | -0.08 |
| **Medication use** | | | | | | | | | |
| Agents acting on the renin-angiotensin system | 75.2 | 69.1 | 0.14 | 72.7 | 72.3 | 0.01 | 74.0 | 74.4 | -0.01 |
| Antibacterials for systemic use | 43.5 | 35.3 | 0.17 | 38.7 | 38.8 | 0.00 | 42.1 | 44.8 | -0.06 |

| (Continued) | Before ajustment | | | After matching | | | After stratification | | |
|---|---|---|---|---|---|---|---|---|---|
| Characteristic | T (%) | C (%) | StdDiff | T (%) | C (%) | StdDiff | T (%) | C (%) | StdDiff |
| Antidepressants | 23.1 | 20.9 | 0.06 | 20.6 | 21.7 | -0.03 | 22.6 | 24.0 | -0.03 |
| Antiepileptics | 11.8 | 9.2 | 0.09 | 9.6 | 10.2 | -0.02 | 11.1 | 11.3 | 0.00 |
| Antiinflammatory and antirheumatic products | 74.9 | 72.3 | 0.06 | 75.1 | 74.8 | 0.01 | 74.9 | 75.1 | 0.00 |
| Antineoplastic agents | 2.2 | 1.6 | 0.04 | 1.7 | 1.7 | 0.00 | 2.1 | 1.4 | 0.06 |
| Antipsoriatics | 2.0 | 1.4 | 0.05 | 1.4 | 1.4 | 0.00 | 1.9 | 1.1 | 0.06 |
| Antithrombotic agents | 85.5 | 78.3 | 0.19 | 81.2 | 81.1 | 0.00 | 84.2 | 82.8 | 0.04 |
| Beta blocking agents | 57.3 | 50.6 | 0.13 | 53.5 | 54.2 | -0.01 | 56.0 | 58.8 | -0.06 |
| Calcium channel blockers | 36.6 | 32.1 | 0.09 | 35.8 | 33.5 | 0.05 | 35.8 | 33.5 | 0.05 |
| Diuretics | 56.9 | 50.0 | 0.14 | 51.8 | 52.4 | -0.01 | 55.2 | 56.5 | -0.03 |
| Drugs for acid related disorders | 75.2 | 68.5 | 0.15 | 71.3 | 71.4 | 0.00 | 74.0 | 75.5 | -0.04 |
| Drugs for obstructive airway diseases | 32.3 | 27.3 | 0.11 | 27.2 | 29.0 | -0.04 | 30.9 | 30.2 | 0.01 |
| Immunosuppressants | 1.6 | 1.2 | 0.04 | 0.8 | 1.3 | -0.05 | 1.4 | 1.2 | 0.01 |
| Lipid modifying agents | 84.5 | 79.0 | 0.14 | 82.1 | 82.8 | -0.02 | 83.9 | 82.7 | 0.03 |
| Opioids | 20.5 | 16.5 | 0.10 | 18.1 | 17.9 | 0.00 | 19.6 | 21.7 | -0.05 |
| Psycholeptics | 35.9 | 33.4 | 0.05 | 34.7 | 34.6 | 0.00 | 35.5 | 38.6 | -0.06 |

**Supplementary Table 47:** Baseline patient characteristics for DPP4is (T) and SUs (C) new-users in the VA data source. We report proportion of initiators satisfying selected base-line characteristics and the standardized difference of population proportions (StdDiff) before and after propensity score adjustment. Less extreme StdDiffs through matching and stratification suggest improved balance between patient cohorts through adjustment.

| | Before adjustment | | | After matching | | | After stratification | | |
|---|---|---|---|---|---|---|---|---|---|
| Characteristic | T (%) | C (%) | StdDiff | T (%) | C (%) | StdDiff | T (%) | C (%) | StdDiff |
| **Age group** | | | | | | | | | |
| 30 - 34 | 0.1 | 0.1 | 0.01 | <0.1 | 0.1 | 0.00 | <0.1 | 0.1 | 0.00 |
| 35 - 39 | 0.3 | 0.3 | 0.00 | 0.3 | 0.4 | -0.01 | 0.1 | 0.3 | -0.04 |
| 40 - 44 | 0.8 | 0.9 | -0.01 | 0.8 | 0.8 | 0.01 | 1.0 | 0.9 | 0.01 |
| 45 - 49 | 2.0 | 2.5 | -0.03 | 2.1 | 2.1 | 0.00 | 1.9 | 2.4 | -0.04 |
| 50 - 54 | 4.8 | 5.9 | -0.05 | 4.8 | 4.9 | 0.00 | 2.5 | 5.4 | -0.15 |
| 55 - 59 | 8.1 | 11.7 | -0.12 | 8.0 | 7.8 | 0.00 | 8.9 | 9.9 | -0.04 |
| 60 - 64 | 12.2 | 19.2 | -0.19 | 12.4 | 12.1 | 0.01 | 17.5 | 19.7 | -0.06 |
| 65 - 69 | 20.9 | 21.4 | -0.01 | 21.2 | 20.8 | 0.01 | 22.7 | 23.4 | -0.01 |
| 70 - 74 | 27.2 | 17.1 | 0.24 | 26.9 | 27.7 | -0.02 | 19.4 | 18.3 | 0.03 |
| 75 - 79 | 11.7 | 10.9 | 0.03 | 11.7 | 11.8 | 0.00 | 12.3 | 9.7 | 0.08 |
| 80 - 84 | 6.5 | 6.9 | -0.01 | 6.5 | 6.4 | 0.00 | 7.1 | 6.0 | 0.05 |
| 85 - 89 | 3.8 | 2.7 | 0.06 | 3.6 | 3.8 | -0.01 | 4.5 | 3.0 | 0.08 |
| 90 - 94 | 1.3 | 0.6 | 0.07 | 1.3 | 1.2 | 0.01 | 1.8 | 0.8 | 0.09 |
| 95 - 99 | 0.3 | 0.1 | 0.05 | 0.3 | 0.2 | 0.02 | 0.1 | 0.1 | 0.01 |
| Gender: female | 5.2 | 2.9 | 0.12 | 5.1 | 5.0 | 0.00 | 4.1 | 3.5 | 0.03 |
| **Race** | | | | | | | | | |
| Asian | 1.1 | 0.5 | 0.06 | 1.1 | 1.0 | 0.01 | 0.6 | 0.6 | 0.00 |
| Black or African American | 13.6 | 10.8 | 0.09 | 13.2 | 13.3 | 0.00 | 9.8 | 12.1 | -0.07 |
| White | 78.1 | 78.0 | 0.00 | 78.3 | 78.0 | 0.01 | 82.5 | 79.2 | 0.08 |
| Unknown | 5.6 | 9.3 | -0.14 | 5.7 | 5.8 | -0.01 | 5.2 | 6.5 | -0.06 |
| Native Hawaiian or Other Pacific Islander | 1.0 | 0.8 | 0.02 | 1.0 | 0.9 | 0.00 | 1.3 | 0.8 | 0.04 |
| American Indian or Alaska Native | 0.7 | 0.6 | 0.01 | 0.7 | 0.9 | -0.02 | 0.6 | 0.7 | -0.02 |
| **Ethnicity** | | | | | | | | | |
| Hispanic or Latino | 4.4 | 4.6 | -0.01 | 4.5 | 4.5 | 0.00 | 3.9 | 5.0 | -0.05 |
| Not Hispanic or Latino | 93.4 | 89.8 | 0.13 | 93.4 | 93.2 | 0.01 | 92.9 | 92.1 | 0.03 |
| **Medical history: General** | | | | | | | | | |
| Acute respiratory disease | 9.9 | 10.3 | -0.02 | 9.8 | 9.7 | 0.00 | 7.9 | 10.7 | -0.10 |
| Attention deficit hyperactivity disorder | 0.5 | 0.3 | 0.03 | 0.5 | 0.5 | 0.00 | 0.3 | 0.3 | 0.00 |
| Chronic liver disease | 2.2 | 2.4 | -0.01 | 2.2 | 2.3 | 0.00 | 3.1 | 2.8 | 0.02 |
| Chronic obstructive lung disease | 17.2 | 16.3 | 0.03 | 17.0 | 17.2 | 0.00 | 20.5 | 17.0 | 0.09 |
| Crohn’s disease | 0.2 | 0.2 | -0.01 | 0.2 | 0.2 | -0.01 | 0.1 | 0.2 | -0.03 |
| Dementia | 2.8 | 1.6 | 0.09 | 2.8 | 2.9 | -0.01 | 2.0 | 1.8 | 0.01 |
| Depressive disorder | 22.3 | 21.5 | 0.02 | 22.0 | 22.2 | 0.00 | 27.6 | 23.2 | 0.10 |
| Gastroesophageal reflux disease | 22.6 | 22.4 | 0.01 | 22.6 | 22.7 | 0.00 | 23.9 | 23.5 | 0.01 |
| Gastrointestinal hemorrhage | 1.3 | 1.7 | -0.04 | 1.3 | 1.3 | 0.00 | 0.8 | 1.7 | -0.07 |
| Human immunodeficiency virus infection | 0.4 | 0.3 | 0.03 | 0.4 | 0.5 | -0.01 | 0.4 | 0.3 | 0.01 |
| Hyperlipidemia | 75.6 | 78.1 | -0.06 | 75.5 | 75.2 | 0.01 | 78.4 | 78.9 | -0.01 |
| Hypertensive disorder | 79.5 | 82.6 | -0.08 | 79.4 | 78.9 | 0.01 | 82.3 | 82.5 | 0.00 |
| Lesion of liver | 1.5 | 1.2 | 0.03 | 1.5 | 1.7 | -0.01 | 0.9 | 1.4 | -0.04 |
| Obesity | 26.6 | 28.0 | -0.03 | 26.4 | 26.3 | 0.00 | 29.7 | 29.3 | 0.01 |
| Osteoarthritis | 23.0 | 24.2 | -0.03 | 24.6 | 24.7 | 0.00 | 24.6 | 24.7 | 0.00 |
| Pneumonia | 1.9 | 2.0 | -0.01 | 1.9 | 2.1 | -0.01 | 1.7 | 2.2 | -0.04 |
| Psoriasis | 1.9 | 1.6 | 0.02 | 1.9 | 1.7 | 0.01 | 1.9 | 1.7 | 0.01 |
| Renal impairment | 11.6 | 7.8 | 0.13 | 11.5 | 11.7 | -0.01 | 12.3 | 9.5 | 0.09 |
| Rheumatoid arthritis | 0.8 | 1.0 | -0.01 | 0.9 | 0.9 | -0.01 | 0.8 | 1.0 | -0.02 |
| Schizophrenia | 0.9 | 1.0 | -0.02 | 0.9 | 0.8 | 0.00 | 0.6 | 1.0 | -0.04 |
| Ulcerative colitis | 0.4 | 0.3 | 0.02 | 0.5 | 0.4 | 0.01 | 0.2 | 0.4 | -0.03 |
| Urinary tract infectious disease | 3.0 | 2.9 | 0.00 | 2.9 | 2.8 | 0.00 | 2.4 | 3.1 | -0.04 |
| Viral hepatitis C | 1.1 | 1.8 | -0.06 | 1.2 | 1.3 | -0.01 | 1.8 | 2.0 | -0.01 |
| Visual system disorder | 48.9 | 47.5 | 0.03 | 48.8 | 48.3 | 0.01 | 49.4 | 48.9 | 0.01 |
| **Medical history: Cardiovascular disease** | | | | | | | | | |
| Atrial fibrillation | 12.2 | 9.2 | 0.10 | 12.0 | 12.4 | -0.01 | 13.0 | 9.8 | 0.10 |
| Cerebrovascular disease | 7.5 | 10.5 | -0.10 | 7.6 | 7.6 | 0.00 | 11.3 | 9.8 | 0.05 |
| Coronary arteriosclerosis | 36.6 | 37.4 | -0.02 | 36.9 | 37.2 | -0.01 | 37.6 | 34.9 | 0.06 |
| Heart disease | 55.1 | 62.2 | -0.14 | 55.2 | 55.4 | 0.00 | 65.0 | 59.2 | 0.12 |
| Heart failure | 8.2 | 9.3 | -0.04 | 8.2 | 8.5 | -0.01 | 11.9 | 9.0 | 0.10 |
| Ischemic heart disease | 16.1 | 29.6 | -0.33 | 16.4 | 15.8 | 0.02 | 31.0 | 27.1 | 0.09 |
| Peripheral vascular disease | 10.9 | 11.9 | -0.03 | 10.8 | 11.1 | -0.01 | 10.3 | 11.3 | -0.03 |
| Pulmonary embolism | 0.9 | 0.7 | 0.01 | 0.9 | 1.0 | -0.01 | 0.9 | 0.9 | 0.00 |
| Venous thrombosis | 1.7 | 1.4 | 0.02 | 1.7 | 1.8 | 0.00 | 1.4 | 1.7 | -0.03 |
| **Medical history: Neoplasms** | | | | | | | | | |

| (Continued) | Before ajustment | | | After matching | | | After stratification | | |
|---|---|---|---|---|---|---|---|---|---|
| Characteristic | T (%) | C (%) | StdDiff | T (%) | C (%) | StdDiff | T (%) | C (%) | StdDiff |
| Hematologic neoplasm | 1.3 | 1.0 | 0.03 | 1.3 | 1.4 | -0.01 | 1.8 | 1.1 | 0.06 |
| Malignant lymphoma | 0.8 | 0.5 | 0.04 | 0.8 | 1.0 | -0.01 | 0.4 | 0.6 | -0.04 |
| Malignant neoplasm of anorectum | 0.1 | 0.2 | -0.02 | <0.1 | 0.1 | 0.00 | 0.1 | 0.1 | -0.03 |
| Malignant neoplastic disease | 11.5 | 11.4 | 0.00 | 11.3 | 11.5 | -0.01 | 14.7 | 11.7 | 0.09 |
| Malignant tumor of breast | 0.1 | 0.1 | 0.00 | 0.1 | 0.1 | 0.00 | 0.1 | 0.1 | 0.00 |
| Malignant tumor of colon | 0.6 | 0.8 | -0.02 | 0.6 | 0.6 | 0.00 | 0.7 | 0.7 | 0.00 |
| Malignant tumor of lung | 0.5 | 0.3 | 0.04 | 0.5 | 0.5 | 0.00 | 0.2 | 0.4 | -0.02 |
| Malignant tumor of urinary bladder | 0.8 | 0.9 | -0.01 | 0.8 | 0.8 | 0.00 | 0.6 | 0.9 | -0.04 |
| Primary malignant neoplasm of prostate | 4.2 | 4.6 | -0.02 | 4.1 | 4.2 | -0.01 | 6.2 | 4.6 | 0.07 |
| **Medication use** | | | | | | | | | |
| Agents acting on the renin-angiotensin system | 66.9 | 75.9 | -0.20 | 67.2 | 66.8 | 0.01 | 70.3 | 73.7 | -0.08 |
| Antibacterials for systemic use | 31.0 | 33.8 | -0.06 | 30.7 | 30.6 | 0.00 | 30.7 | 34.2 | -0.07 |
| Antidepressants | 37.0 | 36.4 | 0.01 | 36.8 | 37.0 | 0.00 | 42.5 | 37.8 | 0.10 |
| Antiepileptics | 30.2 | 23.3 | 0.16 | 29.9 | 30.4 | -0.01 | 29.3 | 27.1 | 0.05 |
| Antiinflammatory and antirheumatic products | 50.9 | 50.5 | 0.01 | 50.6 | 50.8 | 0.00 | 47.5 | 50.8 | -0.07 |
| Antineoplastic agents | 4.9 | 3.3 | 0.08 | 4.8 | 4.7 | 0.00 | 5.4 | 3.6 | 0.09 |
| Antipsoriatics | 1.0 | 0.8 | 0.03 | 1.1 | 1.0 | 0.00 | 1.4 | 0.9 | 0.05 |
| Antithrombotic agents | 50.3 | 49.9 | 0.01 | 49.8 | 50.3 | -0.01 | 50.4 | 49.4 | 0.02 |
| Beta blocking agents | 54.3 | 61.9 | -0.16 | 54.8 | 54.7 | 0.00 | 58.6 | 60.6 | -0.04 |
| Calcium channel blockers | 31.4 | 31.2 | 0.00 | 31.2 | 31.1 | 0.00 | 34.7 | 31.2 | 0.07 |
| Diuretics | 41.0 | 49.2 | -0.17 | 41.2 | 40.9 | 0.01 | 47.4 | 48.0 | -0.01 |
| Drugs for acid related disorders | 49.1 | 50.1 | -0.02 | 49.1 | 49.0 | 0.00 | 58.5 | 51.1 | 0.15 |
| Drugs for obstructive airway diseases | 44.7 | 39.1 | 0.11 | 44.2 | 44.4 | 0.00 | 41.9 | 41.5 | 0.01 |
| Immunosuppressants | 2.6 | 1.7 | 0.06 | 2.6 | 2.7 | -0.01 | 3.6 | 2.1 | 0.09 |
| Lipid modifying agents | 84.7 | 86.0 | -0.04 | 84.7 | 84.7 | 0.00 | 85.1 | 86.7 | -0.04 |
| Opioids | 19.2 | 23.8 | -0.11 | 19.2 | 18.7 | 0.01 | 24.6 | 24.2 | 0.01 |
| Psycholeptics | 25.3 | 24.1 | 0.03 | 24.8 | 24.6 | 0.01 | 27.5 | 25.5 | 0.05 |
| Psychostimulants | 1.6 | 1.4 | 0.01 | 1.5 | 1.5 | 0.00 | 1.1 | 1.5 | -0.03 |

**Supplementary Table 48:** Meta-analytic calibrated hazard ratio estimates and their 95% confidence intervals comparing the relative risk of cardiovascular effectiveness outcomes between target and comparator among new users of SGLT2is, GLP1-RAs, DPP4is, and SUs.

| Target | Comparator | 3-pt MACE | 4-pt MACE | Acute MI | HF Hospitalization | Stroke | SCD |
|---|---|---|---|---|---|---|---|
| SGLT2is | GLP1-RAs | 1.06 (0.96 - 1.17) | 1.05 (0.97 - 1.13) | 1.19 (1.05 - 1.35) | 1.02 (0.93 - 1.12) | 0.93 (0.82 - 1.05) | 1.11 (0.84 - 1.45) |
| SGLT2is | DPP4is | 0.89 (0.79 - 1.00) | 0.84 (0.75 - 0.95) | 0.95 (0.83 - 1.08) | 0.80 (0.68 - 0.95) | 0.89 (0.74 - 1.07) | 0.62 (0.51 - 0.75) |
| SGLT2is | SUs | 0.76 (0.65 - 0.89) | 0.71 (0.61 - 0.83) | 0.81 (0.69 - 0.95) | 0.68 (0.58 - 0.80) | 0.77 (0.65 - 0.92) | 0.60 (0.48 - 0.73) |
| GLP1-RAs | DPP4is | 0.83 (0.70 - 0.98) | 0.81 (0.73 - 0.91) | 0.85 (0.73 - 0.99) | 0.76 (0.67 - 0.86) | 0.89 (0.77 - 1.03) | 0.65 (0.44 - 0.96) |
| GLP1-RAs | SUs | 0.72 (0.58 - 0.88) | 0.67 (0.55 - 0.82) | 0.70 (0.56 - 0.87) | 0.64 (0.52 - 0.78) | 0.79 (0.63 - 0.98) | 0.57 (0.42 - 0.78) |
| DPP4is | SUs | 0.87 (0.79 - 0.95) | 0.87 (0.79 - 0.95) | 0.86 (0.77 - 0.95) | 0.90 (0.82 - 0.99) | 0.90 (0.81 - 0.99) | 0.96 (0.81 - 1.13) |

**Supplementary Table 49:** Exposures, outcomes and risk estimates for 3-point MACE in SGLT2is (T) vs GLP1-RAs (C) new-user cohorts under two different propensity score (PS) adjustment / time-at-risk (TAR) designs across data sources. We report total patient exposure time in 1,000 patient-year (KYr) units, uncalibrated and calibrated hazard ratios (HRs) and their 95% confidence intervals (CIs). Greyed out entries did not pass study diagnostics and are included only for completeness.

| | Patients | | Exposure (KYr) | | Outcomes | | Uncalibrated | Calibrated |
|---|---|---|---|---|---|---|---|---|
| | T | C | T | C | T | C | HR (95% CI) | HR (95% CI) |
| **PS-matching / on-treatment TAR** | | | | | | | | |
| CCAE | 8,744 | 4,024 | 6.7 | 2.7 | 86 | 24 | 0.76 (0.43 - 1.36) | 0.74 (0.42 - 1.30) |
| OCEDM | 10,015 | 4,550 | 6.1 | 2.4 | 157 | 48 | 1.24 (0.79 - 1.98) | 1.28 (0.81 - 2.03) |
| MDCR | 1,768 | 663 | 1.3 | 0.4 | 33 | 9 | 1.00 (0.38 - 2.93) | 1.27 (0.46 - 3.51) |
| USOC | 161,590 | 76,637 | 122.0 | 49.3 | 1,630 | 585 | 0.98 (0.86 - 1.12) | 1.04 (0.90 - 1.19) |
| OEHR | 8,053 | 3,448 | 2.5 | 0.9 | 41 | 16 | 1.16 (0.52 - 2.85) | 1.30 (0.56 - 3.06) |
| Meta-analysis | 188,402 | 88,659 | 137.3 | 55.4 | 1,914 | 673 | 0.99 (0.87 - 1.12) | 1.04 (0.91 - 1.18) |
| **PS-stratification / on-treatment TAR** | | | | | | | | |
| CCAE | 8,747 | 4,800 | 6.7 | 3.1 | 86 | 29 | 1.10 (0.71 - 1.75) | 1.11 (0.71 - 1.74) |
| OCEDM | 10,018 | 5,304 | 6.1 | 2.8 | 157 | 54 | 1.22 (0.88 - 1.72) | 1.24 (0.88 - 1.73) |
| MDCR | 1,775 | 692 | 1.3 | 0.4 | 33 | 9 | 1.05 (0.50 - 2.43) | 1.16 (0.52 - 2.56) |
| USOC | 161,615 | 89,631 | 122.1 | 56.0 | 1,630 | 653 | 1.00 (0.91 - 1.10) | 1.05 (0.95 - 1.16) |
| OEHR | 8,063 | 3,993 | 2.5 | 1.1 | 41 | 16 | 1.08 (0.59 - 2.05) | 1.15 (0.61 - 2.18) |
| Meta-analysis | 188,443 | 103,728 | 137.4 | 62.9 | 1,914 | 752 | 1.02 (0.93 - 1.12) | 1.06 (0.96 - 1.17) |

**Supplementary Table 50:** Exposures, outcomes and risk estimates for 3-point MACE in SGLT2is (T) vs DPP4is (C) new-user cohorts under two different propensity score (PS) adjustment / time-at-risk (TAR) designs across data sources. We report total patient exposure time in 1,000 patient-year (KYr) units, uncalibrated and calibrated hazard ratios (HRs) and their 95% confidence intervals (CIs). Greyed out entries did not pass study diagnostics and are included only for completeness.

| | Patients | | Exposure (KYr) | | Outcomes | | Uncalibrated | Calibrated |
|---|---|---|---|---|---|---|---|---|
| | T | C | T | C | T | C | HR (95% CI) | HR (95% CI) |
| **PS-matching / on-treatment TAR** | | | | | | | | |
| CCAE | 7,060 | 11,442 | 5.8 | 9.0 | 76 | 132 | 0.81 (0.53 - 1.22) | 0.76 (0.49 - 1.17) |
| GDA | 3,365 | 8,557 | 2.8 | 9.0 | 0 | 0 | | |
| OCEDM | 7,471 | 15,256 | 5.0 | 11.6 | 131 | 404 | 1.00 (0.76 - 1.31) | 0.98 (0.75 - 1.29) |
| MDCR | 1,440 | 4,901 | 1.1 | 4.2 | 28 | 169 | 0.69 (0.38 - 1.21) | 0.74 (0.41 - 1.32) |
| MDCD | 964 | 2,878 | 0.5 | 2.2 | 18 | 80 | 1.05 (0.51 - 2.05) | 0.98 (0.49 - 1.95) |
| USOC | 119,943 | 251,237 | 99.4 | 225.9 | 1,320 | 4,493 | 0.79 (0.73 - 0.87) | 0.81 (0.67 - 0.98) |
| OEHR | 6,514 | 14,283 | 2.1 | 7.8 | 34 | 159 | 0.93 (0.55 - 1.50) | 0.83 (0.50 - 1.36) |
| SIDIAP | 1,241 | 5,180 | 1.7 | 10.6 | 7 | 100 | 0.47 (0.16 - 1.14) | 0.46 (0.16 - 1.37) |
| VA | 3,435 | 8,776 | 2.5 | 8.3 | 22 | 63 | 1.00 (0.51 - 1.92) | 0.96 (0.49 - 1.87) |
| Meta-analysis | 148,068 | 313,953 | 118.1 | 279.6 | 1,636 | 5,600 | 0.81 (0.75 - 0.88) | 0.81 (0.74 - 0.89) |
| **PS-stratification / on-treatment TAR** | | | | | | | | |
| CCAE | 8,747 | 11,484 | 6.7 | 9.1 | 86 | 134 | 0.84 (0.63 - 1.13) | 0.85 (0.64 - 1.15) |
| GDA | 4,434 | 8,797 | 3.5 | 9.3 | 0 | 0 | | |
| OCEDM | 10,018 | 15,616 | 6.1 | 11.9 | 157 | 427 | 0.86 (0.70 - 1.06) | 0.87 (0.70 - 1.07) |
| MDCR | 1,775 | 4,983 | 1.3 | 4.3 | 33 | 174 | 0.77 (0.49 - 1.18) | 0.79 (0.51 - 1.22) |
| MDCD | 1,030 | 2,889 | 0.6 | 2.2 | 20 | 80 | 1.11 (0.63 - 1.87) | 0.99 (0.57 - 1.70) |
| USOC | 161,615 | 251,760 | 122.1 | 226.3 | 1,630 | 4,525 | 0.87 (0.82 - 0.93) | 0.91 (0.79 - 1.06) |
| OEHR | 8,063 | 14,463 | 2.5 | 7.9 | 41 | 166 | 1.07 (0.73 - 1.55) | 0.98 (0.67 - 1.43) |
| SIDIAP | 1,726 | 5,570 | 2.2 | 11.7 | 10 | 108 | 0.55 (0.24 - 1.12) | 0.54 (0.24 - 1.21) |
| VA | 6,127 | 9,248 | 3.9 | 9.0 | 42 | 72 | 1.01 (0.60 - 1.67) | 1.02 (0.61 - 1.70) |
| Meta-analysis | 188,443 | 293,323 | 137.4 | 255.1 | 1,914 | 5,252 | 0.87 (0.82 - 0.93) | 0.89 (0.79 - 1.00) |

**Supplementary Table 51:** Exposures, outcomes and risk estimates for 3-point MACE in SGLT2is (T) vs SUs (C) new-user cohorts under two different propensity score (PS) adjustment / time-at-risk (TAR) designs across data sources. We report total patient exposure time in 1,000 patient-year (KYr) units, uncalibrated and calibrated hazard ratios (HRs) and their 95% confidence intervals (CIs). Greyed out entries did not pass study diagnostics and are included only for completeness.

| | Patients | | Exposure (KYr) | | Outcomes | | Uncalibrated | Calibrated |
|---|---|---|---|---|---|---|---|---|
| | T | C | T | C | T | C | HR (95% CI) | HR (95% CI) |
| **PS-matching / on-treatment TAR** | | | | | | | | |
| CCAE | 7,203 | 15,151 | 5.7 | 11.6 | 76 | 216 | 0.74 (0.50 - 1.07) | 0.71 (0.49 - 1.05) |
| GDA | 4,184 | 723 | 3.4 | 0.7 | 0 | 0 | | |
| OCEDM | 8,841 | 32,054 | 5.6 | 28.2 | 139 | 1,153 | 0.87 (0.69 - 1.09) | 0.90 (0.72 - 1.14) |
| MDCR | 1,474 | 7,015 | 1.1 | 5.9 | 31 | 243 | 0.65 (0.36 - 1.11) | 0.72 (0.41 - 1.27) |
| MDCD | 1,018 | 5,034 | 0.6 | 3.4 | 20 | 144 | 0.84 (0.45 - 1.48) | 0.83 (0.46 - 1.50) |
| USOC | 144,004 | 460,852 | 112.1 | 472.7 | 1,509 | 11,428 | 0.74 (0.69 - 0.80) | 0.77 (0.62 - 0.95) |
| OEHR | 7,022 | 29,857 | 2.2 | 18.1 | 36 | 523 | 0.74 (0.47 - 1.11) | 0.68 (0.44 - 1.04) |
| SIDIAP | 1,347 | 289 | 1.9 | 0.5 | 7 | 0 | | |
| VA | 4,298 | 23,732 | 3.1 | 20.3 | 30 | 220 | 0.95 (0.56 - 1.55) | 0.97 (0.58 - 1.62) |
| Meta-analysis | 169,562 | 549,963 | 127.2 | 540.0 | 1,811 | 13,707 | 0.75 (0.70 - 0.80) | 0.76 (0.65 - 0.89) |
| **PS-stratification / on-treatment TAR** | | | | | | | | |
| CCAE | 8,747 | 15,421 | 6.7 | 11.8 | 86 | 221 | 0.76 (0.58 - 1.00) | 0.75 (0.57 - 0.98) |
| GDA | 4,337 | 1,235 | 3.5 | 1.3 | 0 | 0 | | |
| OCEDM | 10,018 | 32,592 | 6.1 | 28.6 | 157 | 1,190 | 0.83 (0.69 - 1.00) | 0.83 (0.69 - 1.00) |
| MDCR | 1,775 | 7,469 | 1.3 | 6.3 | 33 | 264 | 0.82 (0.53 - 1.22) | 0.79 (0.52 - 1.20) |
| MDCD | 1,031 | 5,327 | 0.6 | 3.7 | 20 | 152 | 1.04 (0.61 - 1.67) | 1.01 (0.61 - 1.67) |
| USOC | 161,615 | 467,651 | 122.1 | 479.0 | 1,630 | 11,794 | 0.75 (0.71 - 0.79) | 0.78 (0.65 - 0.93) |
| OEHR | 8,063 | 31,364 | 2.5 | 19.3 | 41 | 573 | 0.68 (0.48 - 0.96) | 0.64 (0.45 - 0.93) |
| SIDIAP | 1,714 | 566 | 2.2 | 1.2 | 10 | 8 | 1.16 (0.27 - 4.49) | 1.22 (0.30 - 5.01) |
| VA | 6,127 | 34,583 | 3.9 | 32.5 | 42 | 424 | 1.02 (0.67 - 1.54) | 1.05 (0.69 - 1.59) |
| Meta-analysis | 188,443 | 547,028 | 137.4 | 538.7 | 1,914 | 13,778 | 0.75 (0.71 - 0.79) | 0.76 (0.66 - 0.88) |

**Supplementary Table 52:** Exposures, outcomes and risk estimates for 3-point MACE in GLP1-RAs (T) vs DPP4is (C) new-user cohorts under two different propensity score (PS) adjustment / time-at-risk (TAR) designs across data sources. We report total patient exposure time in 1,000 patient-year (KYr) units, uncalibrated and calibrated hazard ratios (HRs) and their 95% confidence intervals (CIs). Greyed out entries did not pass study diagnostics and are included only for completeness.

| | Patients | | Exposure (KYr) | | Outcomes | | Uncalibrated | Calibrated |
|---|---|---|---|---|---|---|---|---|
| | T | C | T | C | T | C | HR (95% CI) | HR (95% CI) |
| **PS-matching / on-treatment TAR** | | | | | | | | |
| CCAE | 4,967 | 20,049 | 3.3 | 16.5 | 32 | 228 | 0.91 (0.57 - 1.42) | 0.93 (0.59 - 1.48) |
| OCEDM | 4,476 | 20,558 | 2.5 | 16.4 | 54 | 522 | 0.76 (0.52 - 1.08) | 0.75 (0.52 - 1.09) |
| MDCR | 863 | 8,495 | 0.5 | 8.1 | 12 | 299 | 0.77 (0.36 - 1.50) | 0.67 (0.33 - 1.38) |
| USOC | 63,780 | 251,578 | 44.8 | 226.7 | 554 | 4,439 | 0.90 (0.80 - 1.02) | 0.88 (0.76 - 1.01) |
| OEHR | 3,337 | 16,582 | 1.0 | 8.9 | 17 | 187 | 0.90 (0.45 - 1.65) | 0.76 (0.40 - 1.46) |
| Meta-analysis | 76,560 | 308,767 | 51.6 | 268.5 | 657 | 5,376 | 0.89 (0.80 - 0.99) | 0.87 (0.76 - 0.99) |
| **PS-stratification / on-treatment TAR** | | | | | | | | |
| CCAE | 6,289 | 20,129 | 3.9 | 16.5 | 37 | 230 | 0.83 (0.55 - 1.20) | 0.83 (0.56 - 1.23) |
| OCEDM | 5,917 | 21,000 | 3.1 | 16.8 | 61 | 551 | 0.66 (0.48 - 0.89) | 0.64 (0.47 - 0.88) |
| MDCR | 940 | 9,093 | 0.5 | 8.7 | 12 | 330 | 0.67 (0.34 - 1.18) | 0.59 (0.32 - 1.10) |
| USOC | 89,766 | 253,586 | 56.1 | 228.5 | 654 | 4,557 | 0.89 (0.81 - 0.98) | 0.88 (0.75 - 1.03) |
| OEHR | 4,156 | 16,689 | 1.2 | 8.9 | 17 | 197 | 1.06 (0.60 - 1.76) | 0.88 (0.51 - 1.51) |
| Meta-analysis | 106,128 | 311,404 | 64.2 | 270.7 | 769 | 5,535 | 0.85 (0.73 - 0.98) | 0.83 (0.69 - 0.99) |

**Supplementary Table 53:** Exposures, outcomes and risk estimates for 3-point MACE in GLP1-RAs (T) vs SUs (C) new-user cohorts under two different propensity score (PS) adjustment / time-at-risk (TAR) designs across data sources. We report total patient exposure time in 1,000 patient-year (KYr) units, uncalibrated and calibrated hazard ratios (HRs) and their 95% confidence intervals (CIs). Greyed out entries did not pass study diagnostics and are included only for completeness.

| | Patients | | Exposure (KYr) | | Outcomes | | Uncalibrated | Calibrated |
|---|---|---|---|---|---|---|---|---|
| | T | C | T | C | T | C | HR (95% CI) | HR (95% CI) |
| **PS-matching / on-treatment TAR** | | | | | | | | |
| CCAE | 5,270 | 29,862 | 3.4 | 23.2 | 36 | 399 | 0.78 (0.49 - 1.18) | 0.73 (0.47 - 1.14) |
| OCEDM | 4,967 | 39,869 | 2.7 | 35.6 | 54 | 1,294 | 0.62 (0.44 - 0.87) | 0.63 (0.45 - 0.89) |
| MDCR | 931 | 13,596 | 0.6 | 13.0 | 13 | 515 | 0.64 (0.32 - 1.18) | 0.52 (0.27 - 1.02) |
| USOC | 75,427 | 451,969 | 49.5 | 465.1 | 596 | 10,863 | 0.76 (0.68 - 0.84) | 0.74 (0.61 - 0.89) |
| OEHR | 3,547 | 34,330 | 1.0 | 20.6 | 17 | 559 | 0.74 (0.40 - 1.26) | 0.68 (0.38 - 1.20) |
| Meta-analysis | 90,142 | 569,626 | 57.2 | 557.5 | 716 | 13,630 | 0.74 (0.68 - 0.82) | 0.71 (0.59 - 0.86) |
| **PS-stratification / on-treatment TAR** | | | | | | | | |
| CCAE | 6,624 | 30,474 | 4.1 | 23.6 | 38 | 412 | 0.69 (0.47 - 0.99) | 0.68 (0.46 - 0.98) |
| OCEDM | 5,917 | 45,550 | 3.1 | 41.6 | 61 | 1,715 | 0.71 (0.53 - 0.93) | 0.71 (0.53 - 0.94) |
| MDCR | 1,007 | 17,499 | 0.6 | 16.5 | 13 | 737 | 0.59 (0.32 - 1.01) | 0.49 (0.27 - 0.87) |
| USOC | 89,766 | 473,027 | 56.1 | 486.9 | 654 | 11,988 | 0.74 (0.67 - 0.80) | 0.72 (0.58 - 0.89) |
| OEHR | 4,157 | 38,799 | 1.2 | 23.6 | 17 | 699 | 0.69 (0.40 - 1.12) | 0.57 (0.34 - 0.96) |
| Meta-analysis | 96,390 | 503,501 | 60.1 | 510.5 | 692 | 12,400 | 0.73 (0.67 - 0.80) | 0.72 (0.58 - 0.88) |

**Supplementary Table 54:** Exposures, outcomes and risk estimates for 3-point MACE in DPP4is (T) vs SUs (C) new-user cohorts under two different propensity score (PS) adjustment / time-at-risk (TAR) designs across data sources. We report total patient exposure time in 1,000 patient-year (KYr) units, uncalibrated and calibrated hazard ratios (HRs) and their 95% confidence intervals (CIs). Greyed out entries did not pass study diagnostics and are included only for completeness.

| | Patients | | Exposure (KYr) | | Outcomes | | Uncalibrated | Calibrated |
|---|---|---|---|---|---|---|---|---|
| | T | C | T | C | T | C | HR (95% CI) | HR (95% CI) |
| **PS-matching / on-treatment TAR** | | | | | | | | |
| CCAE | 17,378 | 28,723 | 14.2 | 22.1 | 209 | 383 | 0.93 (0.73 - 1.19) | 0.95 (0.74 - 1.22) |
| GDA | 10,408 | 1,974 | 11.3 | 2.0 | 0 | 0 | | |
| OCEDM | 19,834 | 45,526 | 15.8 | 41.6 | 520 | 1,712 | 0.91 (0.79 - 1.04) | 0.96 (0.84 - 1.11) |
| MDCR | 8,489 | 15,979 | 8.1 | 14.9 | 307 | 663 | 0.86 (0.71 - 1.04) | 0.82 (0.67 - 1.00) |
| MDCD | 3,453 | 7,647 | 2.6 | 5.4 | 86 | 232 | 0.83 (0.58 - 1.18) | 0.87 (0.61 - 1.24) |
| IMRD | 3,388 | 407 | 5.7 | 0.6 | 0 | 0 | | |
| USOC | 246,318 | 473,034 | 220.3 | 487.0 | 4,410 | 11,989 | 0.87 (0.83 - 0.91) | 0.87 (0.77 - 0.97) |
| OEHR | 15,736 | 38,767 | 8.3 | 23.6 | 189 | 699 | 0.92 (0.74 - 1.14) | 0.90 (0.72 - 1.11) |
| SIDIAP | 7,174 | 1,676 | 19.0 | 4.2 | 153 | 25 | 1.50 (0.85 - 2.82) | 1.46 (0.80 - 2.68) |
| VA | 9,289 | 58,157 | 9.5 | 61.0 | 79 | 748 | 0.83 (0.60 - 1.14) | 0.85 (0.62 - 1.17) |
| Meta-analysis | 327,671 | 669,509 | 297.7 | 659.7 | 5,953 | 16,451 | 0.88 (0.84 - 0.92) | 0.88 (0.79 - 0.97) |
| **PS-stratification / on-treatment TAR** | | | | | | | | |
| CCAE | 20,129 | 28,846 | 16.5 | 22.2 | 230 | 386 | 0.89 (0.75 - 1.06) | 0.90 (0.75 - 1.07) |
| GDA | 10,428 | 2,365 | 11.3 | 2.5 | 0 | 0 | | |
| OCEDM | 21,003 | 45,554 | 16.8 | 41.6 | 551 | 1,715 | 0.90 (0.81 - 0.99) | 0.91 (0.82 - 1.01) |
| MDCR | 9,093 | 16,082 | 8.7 | 15.0 | 330 | 669 | 0.92 (0.80 - 1.06) | 0.92 (0.80 - 1.06) |
| MDCD | 3,622 | 7,678 | 2.8 | 5.4 | 94 | 232 | 0.88 (0.68 - 1.14) | 0.90 (0.70 - 1.17) |
| IMRD | 4,024 | 846 | 6.5 | 1.5 | 0 | 0 | | |
| USOC | 253,598 | 473,060 | 228.5 | 487.0 | 4,557 | 11,991 | 0.86 (0.83 - 0.89) | 0.85 (0.75 - 0.97) |
| OEHR | 16,689 | 38,798 | 8.9 | 23.6 | 197 | 699 | 0.85 (0.72 - 1.00) | 0.88 (0.75 - 1.04) |
| SIDIAP | 7,322 | 2,193 | 19.1 | 5.8 | 154 | 33 | 1.21 (0.79 - 1.91) | 1.15 (0.74 - 1.78) |
| VA | 9,928 | 88,164 | 9.9 | 97.7 | 81 | 1,239 | 0.77 (0.59 - 0.98) | 0.80 (0.62 - 1.03) |
| Meta-analysis | 324,134 | 610,018 | 282.2 | 594.7 | 5,959 | 15,692 | 0.87 (0.84 - 0.89) | 0.87 (0.79 - 0.95) |

**Supplementary Table 55:** Exposures, outcomes and risk estimates for 4-point MACE in SGLT2is (T) vs GLP1-RAs (C) new-user cohorts under two different propensity score (PS) adjustment / time-at-risk (TAR) designs across data sources. We report total patient exposure time in 1,000 patient-year (KYr) units, uncalibrated and calibrated hazard ratios (HRs) and their 95% confidence intervals (CIs). Greyed out entries did not pass study diagnostics and are included only for completeness.

| | Patients | | Exposure (KYr) | | Outcomes | | Uncalibrated | Calibrated |
|---|---|---|---|---|---|---|---|---|
| | T | C | T | C | T | C | HR (95% CI) | HR (95% CI) |
| **PS-matching / on-treatment TAR** | | | | | | | | |
| CCAE | 8,362 | 3,841 | 6.4 | 2.5 | 132 | 42 | 1.13 (0.71 - 1.83) | 1.09 (0.68 - 1.76) |
| OCEDM | 9,034 | 4,169 | 5.6 | 2.2 | 239 | 88 | 1.09 (0.78 - 1.53) | 1.13 (0.81 - 1.57) |
| MDCR | 1,604 | 601 | 1.2 | 0.4 | 47 | 13 | 1.05 (0.48 - 2.50) | 1.34 (0.58 - 3.06) |
| USOC | 147,379 | 71,400 | 112.6 | 46.1 | 2,402 | 884 | 0.99 (0.89 - 1.10) | 1.05 (0.93 - 1.18) |
| OEHR | 7,602 | 3,277 | 2.3 | 0.9 | 76 | 23 | 1.70 (0.94 - 3.25) | 1.91 (1.02 - 3.55) |
| Meta-analysis | 172,377 | 82,687 | 127.0 | 51.7 | 2,849 | 1,037 | 1.03 (0.91 - 1.16) | 1.08 (0.95 - 1.23) |
| **PS-stratification / on-treatment TAR** | | | | | | | | |
| CCAE | 8,365 | 4,571 | 6.4 | 2.9 | 132 | 47 | 1.16 (0.82 - 1.68) | 1.17 (0.82 - 1.67) |
| OCEDM | 9,043 | 4,874 | 5.6 | 2.6 | 239 | 99 | 0.99 (0.77 - 1.28) | 1.00 (0.77 - 1.29) |
| MDCR | 1,610 | 631 | 1.2 | 0.4 | 47 | 13 | 1.01 (0.54 - 2.02) | 1.11 (0.58 - 2.15) |
| USOC | 147,404 | 83,802 | 112.7 | 52.3 | 2,404 | 994 | 0.99 (0.92 - 1.08) | 1.05 (0.96 - 1.14) |
| OEHR | 7,615 | 3,800 | 2.3 | 1.1 | 76 | 26 | 1.35 (0.85 - 2.22) | 1.45 (0.88 - 2.39) |
| Meta-analysis | 172,427 | 97,047 | 127.0 | 58.9 | 2,851 | 1,166 | 1.01 (0.94 - 1.08) | 1.05 (0.97 - 1.13) |

**Supplementary Table 56:** Exposures, outcomes and risk estimates for 4-point MACE in SGLT2is (T) vs DPP4is (C) new-user cohorts under two different propensity score (PS) adjustment / time-at-risk (TAR) designs across data sources. We report total patient exposure time in 1,000 patient-year (KYr) units, uncalibrated and calibrated hazard ratios (HRs) and their 95% confidence intervals (CIs). Greyed out entries did not pass study diagnostics and are included only for completeness.

| | Patients | | Exposure (KYr) | | Outcomes | | Uncalibrated | Calibrated |
|---|---|---|---|---|---|---|---|---|
| | T | C | T | C | T | C | HR (95% CI) | HR (95% CI) |
| **PS-matching / on-treatment TAR** | | | | | | | | |
| CCAE | 6,786 | 10,911 | 5.5 | 8.6 | 110 | 193 | 0.78 (0.55 - 1.08) | 0.73 (0.51 - 1.04) |
| GDA | 3,365 | 8,557 | 2.8 | 9.0 | 0 | 0 | | |
| OCEDM | 6,863 | 13,775 | 4.6 | 10.4 | 183 | 615 | 0.79 (0.63 - 1.00) | 0.78 (0.62 - 0.99) |
| MDCR | 1,323 | 4,392 | 1.0 | 3.7 | 40 | 252 | 0.58 (0.33 - 0.95) | 0.62 (0.37 - 1.04) |
| MDCD | 836 | 2,430 | 0.5 | 1.8 | 27 | 117 | 1.38 (0.78 - 2.38) | 1.28 (0.73 - 2.24) |
| USOC | 112,057 | 231,516 | 93.2 | 207.3 | 1,902 | 7,197 | 0.73 (0.68 - 0.78) | 0.75 (0.63 - 0.90) |
| OEHR | 6,202 | 13,496 | 2.0 | 7.2 | 61 | 291 | 0.94 (0.66 - 1.34) | 0.84 (0.59 - 1.21) |
| SIDIAP | 1,228 | 5,121 | 1.7 | 10.5 | 10 | 126 | 0.55 (0.23 - 1.12) | 0.54 (0.22 - 1.33) |
| VA | 3,203 | 8,277 | 2.4 | 7.7 | 39 | 150 | 0.81 (0.49 - 1.31) | 0.78 (0.48 - 1.27) |
| Meta-analysis | 138,498 | 289,918 | 110.8 | 257.2 | 2,372 | 8,941 | 0.77 (0.69 - 0.86) | 0.77 (0.68 - 0.87) |
| **PS-stratification / on-treatment TAR** | | | | | | | | |
| CCAE | 8,365 | 10,985 | 6.4 | 8.7 | 132 | 196 | 0.90 (0.71 - 1.14) | 0.91 (0.71 - 1.16) |
| GDA | 4,434 | 8,797 | 3.5 | 9.3 | 0 | 0 | | |
| OCEDM | 9,043 | 14,029 | 5.6 | 10.6 | 239 | 642 | 0.84 (0.70 - 0.99) | 0.84 (0.71 - 1.00) |
| MDCR | 1,610 | 4,436 | 1.2 | 3.8 | 47 | 254 | 0.70 (0.48 - 0.98) | 0.71 (0.50 - 1.01) |
| MDCD | 882 | 2,440 | 0.5 | 1.8 | 29 | 118 | 1.23 (0.78 - 1.89) | 1.09 (0.70 - 1.70) |
| USOC | 147,404 | 231,761 | 112.7 | 207.5 | 2,404 | 7,224 | 0.81 (0.76 - 0.85) | 0.84 (0.73 - 0.98) |
| OEHR | 7,615 | 13,660 | 2.3 | 7.3 | 76 | 305 | 1.02 (0.76 - 1.34) | 0.93 (0.70 - 1.24) |
| SIDIAP | 1,693 | 5,507 | 2.1 | 11.6 | 12 | 140 | 0.55 (0.27 - 1.05) | 0.55 (0.27 - 1.12) |
| VA | 5,379 | 8,732 | 3.5 | 8.4 | 70 | 161 | 0.68 (0.47 - 0.97) | 0.69 (0.47 - 0.99) |
| Meta-analysis | 172,427 | 270,435 | 127.0 | 234.0 | 2,851 | 8,367 | 0.82 (0.77 - 0.88) | 0.84 (0.75 - 0.94) |

**Supplementary Table 57:** Exposures, outcomes and risk estimates for 4-point MACE in SGLT2is (T) vs SUs (C) new-user cohorts under two different propensity score (PS) adjustment / time-at-risk (TAR) designs across data sources. We report total patient exposure time in 1,000 patient-year (KYr) units, uncalibrated and calibrated hazard ratios (HRs) and their 95% confidence intervals (CIs). Greyed out entries did not pass study diagnostics and are included only for completeness.

| | Patients | | Exposure (KYr) | | Outcomes | | Uncalibrated | Calibrated |
|---|---|---|---|---|---|---|---|---|
| | T | C | T | C | T | C | HR (95% CI) | HR (95% CI) |
| **PS-matching / on-treatment TAR** | | | | | | | | |
| CCAE | 6,910 | 14,382 | 5.4 | 11.0 | 114 | 310 | 0.69 (0.50 - 0.94) | 0.67 (0.49 - 0.91) |
| GDA | 4,184 | 723 | 3.4 | 0.7 | 0 | 0 | | |
| OCEDM | 8,093 | 28,804 | 5.1 | 24.9 | 205 | 1,810 | 0.70 (0.58 - 0.85) | 0.73 (0.60 - 0.89) |
| MDCR | 1,340 | 6,162 | 1.0 | 5.2 | 43 | 360 | 0.54 (0.33 - 0.86) | 0.60 (0.37 - 0.98) |
| MDCD | 873 | 4,253 | 0.5 | 2.9 | 29 | 209 | 0.94 (0.58 - 1.48) | 0.93 (0.58 - 1.48) |
| USOC | 133,349 | 422,207 | 104.5 | 430.5 | 2,218 | 18,027 | 0.68 (0.64 - 0.73) | 0.71 (0.58 - 0.87) |
| OEHR | 6,668 | 28,113 | 2.1 | 16.7 | 68 | 898 | 0.79 (0.58 - 1.07) | 0.73 (0.53 - 1.00) |
| SIDIAP | 1,331 | 287 | 1.9 | 0.5 | 10 | <5 | 1.31 (0.23 - 9.93) | 1.40 (0.21 - 9.14) |
| VA | 3,947 | 22,020 | 2.9 | 18.7 | 53 | 481 | 0.68 (0.47 - 0.97) | 0.70 (0.48 - 1.01) |
| Meta-analysis | 157,233 | 503,921 | 118.6 | 491.1 | 2,677 | 21,614 | 0.69 (0.65 - 0.73) | 0.70 (0.59 - 0.81) |
| **PS-stratification / on-treatment TAR** | | | | | | | | |
| CCAE | 8,365 | 14,619 | 6.4 | 11.1 | 132 | 318 | 0.78 (0.62 - 0.97) | 0.76 (0.61 - 0.96) |
| GDA | 4,337 | 1,235 | 3.5 | 1.3 | 0 | 0 | | |
| OCEDM | 9,043 | 29,169 | 5.6 | 25.3 | 239 | 1,853 | 0.74 (0.63 - 0.86) | 0.74 (0.63 - 0.86) |
| MDCR | 1,610 | 6,535 | 1.2 | 5.5 | 47 | 398 | 0.69 (0.48 - 0.98) | 0.67 (0.47 - 0.96) |
| MDCD | 883 | 4,455 | 0.5 | 3.0 | 29 | 220 | 0.96 (0.63 - 1.43) | 0.94 (0.62 - 1.42) |
| USOC | 147,404 | 427,514 | 112.7 | 435.4 | 2,404 | 18,602 | 0.70 (0.66 - 0.73) | 0.72 (0.60 - 0.87) |
| OEHR | 7,615 | 29,444 | 2.3 | 17.6 | 76 | 986 | 0.77 (0.60 - 0.99) | 0.73 (0.55 - 0.97) |
| SIDIAP | 1,681 | 564 | 2.1 | 1.2 | 12 | 10 | 1.17 (0.34 - 3.83) | 1.23 (0.36 - 4.16) |
| VA | 5,379 | 32,188 | 3.5 | 29.9 | 70 | 851 | 0.78 (0.57 - 1.05) | 0.79 (0.58 - 1.08) |
| Meta-analysis | 172,427 | 500,746 | 127.0 | 489.5 | 2,851 | 21,759 | 0.70 (0.67 - 0.73) | 0.71 (0.61 - 0.82) |

**Supplementary Table 58:** Exposures, outcomes and risk estimates for 4-point MACE in GLP1-RAs (T) vs DPP4is (C) new-user cohorts under two different propensity score (PS) adjustment / time-at-risk (TAR) designs across data sources. We report total patient exposure time in 1,000 patient-year (KYr) units, uncalibrated and calibrated hazard ratios (HRs) and their 95% confidence intervals (CIs). Greyed out entries did not pass study diagnostics and are included only for completeness.

| | Patients | | Exposure (KYr) | | Outcomes | | Uncalibrated | Calibrated |
|---|---|---|---|---|---|---|---|---|
| | T | C | T | C | T | C | HR (95% CI) | HR (95% CI) |
| **PS-matching / on-treatment TAR** | | | | | | | | |
| CCAE | 4,722 | 19,211 | 3.1 | 15.8 | 50 | 340 | 0.84 (0.57 - 1.21) | 0.85 (0.58 - 1.25) |
| OCEDM | 4,125 | 18,746 | 2.3 | 14.9 | 95 | 805 | 0.87 (0.65 - 1.15) | 0.86 (0.65 - 1.14) |
| MDCR | 798 | 7,678 | 0.5 | 7.4 | 20 | 455 | 0.83 (0.46 - 1.38) | 0.73 (0.42 - 1.25) |
| USOC | 59,432 | 232,281 | 41.8 | 208.4 | 846 | 7,161 | 0.86 (0.78 - 0.95) | 0.83 (0.74 - 0.95) |
| OEHR | 3,175 | 15,734 | 0.9 | 8.2 | 25 | 348 | 1.01 (0.59 - 1.63) | 0.85 (0.51 - 1.41) |
| Meta-analysis | 71,454 | 285,972 | 48.1 | 247.2 | 1,016 | 8,654 | 0.86 (0.79 - 0.94) | 0.84 (0.75 - 0.95) |
| **PS-stratification / on-treatment TAR** | | | | | | | | |
| CCAE | 5,986 | 19,270 | 3.7 | 15.8 | 59 | 344 | 0.78 (0.57 - 1.06) | 0.79 (0.58 - 1.08) |
| OCEDM | 5,463 | 19,085 | 2.9 | 15.2 | 108 | 837 | 0.80 (0.63 - 1.00) | 0.78 (0.62 - 0.99) |
| MDCR | 861 | 8,112 | 0.5 | 7.8 | 21 | 503 | 0.76 (0.46 - 1.18) | 0.67 (0.42 - 1.08) |
| USOC | 83,933 | 233,481 | 52.4 | 209.5 | 996 | 7,287 | 0.84 (0.78 - 0.90) | 0.83 (0.71 - 0.96) |
| OEHR | 3,963 | 15,807 | 1.1 | 8.2 | 28 | 358 | 0.89 (0.57 - 1.33) | 0.73 (0.48 - 1.12) |
| Meta-analysis | 99,345 | 287,643 | 60.1 | 248.7 | 1,191 | 8,826 | 0.83 (0.78 - 0.89) | 0.81 (0.72 - 0.92) |

**Supplementary Table 59:** Exposures, outcomes and risk estimates for 4-point MACE in GLP1-RAs (T) vs SUs (C) new-user cohorts under two different propensity score (PS) adjustment / time-at-risk (TAR) designs across data sources. We report total patient exposure time in 1,000 patient-year (KYr) units, uncalibrated and calibrated hazard ratios (HRs) and their 95% confidence intervals (CIs). Greyed out entries did not pass study diagnostics and are included only for completeness.

| | Patients | | Exposure (KYr) | | Outcomes | | Uncalibrated | Calibrated |
|---|---|---|---|---|---|---|---|---|
| | T | C | T | C | T | C | HR (95% CI) | HR (95% CI) |
| **PS-matching / on-treatment TAR** | | | | | | | | |
| CCAE | 5,023 | 28,402 | 3.2 | 21.9 | 54 | 601 | 0.62 (0.42 - 0.89) | 0.59 (0.40 - 0.85) |
| OCEDM | 4,576 | 36,498 | 2.5 | 32.1 | 97 | 2,122 | 0.74 (0.58 - 0.94) | 0.75 (0.59 - 0.96) |
| MDCR | 856 | 11,819 | 0.5 | 11.1 | 22 | 751 | 0.73 (0.42 - 1.20) | 0.60 (0.35 - 1.02) |
| USOC | 70,228 | 415,477 | 46.2 | 424.7 | 900 | 17,219 | 0.69 (0.64 - 0.76) | 0.68 (0.57 - 0.81) |
| OEHR | 3,381 | 32,483 | 1.0 | 19.0 | 25 | 985 | 0.62 (0.39 - 0.95) | 0.57 (0.36 - 0.90) |
| Meta-analysis | 84,064 | 524,679 | 53.4 | 508.8 | 1,098 | 21,678 | 0.69 (0.64 - 0.75) | 0.67 (0.56 - 0.80) |
| **PS-stratification / on-treatment TAR** | | | | | | | | |
| CCAE | 6,307 | 28,864 | 3.9 | 22.2 | 61 | 617 | 0.64 (0.47 - 0.85) | 0.62 (0.46 - 0.84) |
| OCEDM | 5,463 | 40,955 | 2.9 | 36.8 | 108 | 2,679 | 0.72 (0.58 - 0.89) | 0.71 (0.58 - 0.89) |
| MDCR | 924 | 15,279 | 0.6 | 14.2 | 23 | 1,142 | 0.66 (0.42 - 1.01) | 0.55 (0.35 - 0.85) |
| USOC | 83,933 | 432,544 | 52.4 | 442.7 | 996 | 18,923 | 0.69 (0.64 - 0.73) | 0.67 (0.55 - 0.82) |
| OEHR | 3,964 | 36,554 | 1.1 | 21.6 | 28 | 1,239 | 0.63 (0.42 - 0.93) | 0.52 (0.35 - 0.78) |
| Meta-analysis | 90,240 | 461,408 | 56.3 | 465.0 | 1,057 | 19,540 | 0.68 (0.64 - 0.73) | 0.67 (0.55 - 0.82) |

**Supplementary Table 60:** Exposures, outcomes and risk estimates for 4-point MACE in DPP4is (T) vs SUs (C) new-user cohorts under two different propensity score (PS) adjustment / time-at-risk (TAR) designs across data sources. We report total patient exposure time in 1,000 patient-year (KYr) units, uncalibrated and calibrated hazard ratios (HRs) and their 95% confidence intervals (CIs). Greyed out entries did not pass study diagnostics and are included only for completeness.

| | Patients | | Exposure (KYr) | | Outcomes | | Uncalibrated | Calibrated |
|---|---|---|---|---|---|---|---|---|
| | T | C | T | C | T | C | HR (95% CI) | HR (95% CI) |
| **PS-matching / on-treatment TAR** | | | | | | | | |
| CCAE | 16,582 | 27,224 | 13.6 | 20.8 | 317 | 573 | 0.92 (0.75 - 1.12) | 0.94 (0.77 - 1.14) |
| GDA | 10,408 | 1,974 | 11.3 | 2.0 | 0 | 0 | | |
| OCEDM | 17,930 | 40,945 | 14.2 | 36.8 | 793 | 2,677 | 0.82 (0.73 - 0.91) | 0.86 (0.77 - 0.97) |
| MDCR | 7,545 | 13,935 | 7.2 | 12.9 | 466 | 1,026 | 0.92 (0.79 - 1.07) | 0.88 (0.75 - 1.03) |
| MDCD | 2,925 | 6,391 | 2.2 | 4.4 | 148 | 346 | 0.78 (0.59 - 1.02) | 0.81 (0.62 - 1.07) |
| IMRD | 3,388 | 407 | 5.7 | 0.6 | 0 | 0 | | |
| USOC | 226,514 | 432,555 | 201.7 | 442.8 | 7,046 | 18,926 | 0.86 (0.83 - 0.90) | 0.86 (0.77 - 0.97) |
| OEHR | 14,880 | 36,524 | 7.6 | 21.6 | 341 | 1,239 | 0.91 (0.78 - 1.07) | 0.89 (0.76 - 1.04) |
| SIDIAP | 7,106 | 1,668 | 18.8 | 4.2 | 200 | 32 | 1.55 (0.93 - 2.73) | 1.51 (0.88 - 2.61) |
| VA | 8,765 | 54,613 | 8.9 | 56.7 | 170 | 1,485 | 0.84 (0.67 - 1.04) | 0.86 (0.69 - 1.07) |
| Meta-analysis | 302,247 | 613,855 | 274.1 | 600.2 | 9,481 | 26,304 | 0.87 (0.83 - 0.91) | 0.87 (0.78 - 0.96) |
| **PS-stratification / on-treatment TAR** | | | | | | | | |
| CCAE | 19,270 | 27,338 | 15.8 | 20.9 | 344 | 577 | 0.90 (0.78 - 1.03) | 0.90 (0.78 - 1.04) |
| GDA | 10,428 | 2,365 | 11.3 | 2.5 | 0 | 0 | | |
| OCEDM | 19,088 | 40,959 | 15.2 | 36.8 | 837 | 2,679 | 0.85 (0.78 - 0.92) | 0.86 (0.79 - 0.94) |
| MDCR | 8,112 | 14,038 | 7.8 | 13.0 | 503 | 1,041 | 0.91 (0.81 - 1.02) | 0.91 (0.81 - 1.02) |
| MDCD | 3,076 | 6,408 | 2.3 | 4.4 | 156 | 346 | 0.93 (0.76 - 1.13) | 0.95 (0.77 - 1.16) |
| IMRD | 4,024 | 846 | 6.5 | 1.5 | 0 | 0 | | |
| USOC | 233,492 | 432,576 | 209.5 | 442.8 | 7,287 | 18,928 | 0.87 (0.84 - 0.89) | 0.86 (0.76 - 0.98) |
| OEHR | 15,807 | 36,553 | 8.2 | 21.6 | 358 | 1,239 | 0.87 (0.77 - 0.99) | 0.91 (0.80 - 1.03) |
| SIDIAP | 7,248 | 2,181 | 18.9 | 5.8 | 201 | 41 | 1.18 (0.81 - 1.76) | 1.11 (0.75 - 1.65) |
| VA | 9,378 | 82,904 | 9.3 | 90.9 | 176 | 2,479 | 0.79 (0.66 - 0.94) | 0.82 (0.69 - 0.98) |
| Meta-analysis | 298,845 | 557,872 | 258.8 | 539.5 | 9,485 | 24,810 | 0.87 (0.85 - 0.89) | 0.87 (0.79 - 0.95) |

**Supplementary Table 61:** Exposures, outcomes and risk estimates for Acute myocardial infarction in SGLT2is (T) vs GLP1-RAs (C) new-user cohorts under two different propensity score (PS) adjustment / time-at-risk (TAR) designs across data sources. We report total patient exposure time in 1,000 patient-year (KYr) units, uncalibrated and calibrated hazard ratios (HRs) and their 95% confidence intervals (CIs). Greyed out entries did not pass study diagnostics and are included only for completeness.

| | Patients | | Exposure (KYr) | | Outcomes | | Uncalibrated | Calibrated |
|---|---|---|---|---|---|---|---|---|
| | T | C | T | C | T | C | HR (95% CI) | HR (95% CI) |
| **PS-matching / on-treatment TAR** | | | | | | | | |
| CCAE | 9,190 | 4,234 | 7.0 | 2.8 | 54 | 16 | 0.62 (0.31 - 1.27) | 0.60 (0.29 - 1.22) |
| OCEDM | 10,597 | 4,787 | 6.5 | 2.6 | 87 | 24 | 1.42 (0.75 - 2.83) | 1.46 (0.75 - 2.84) |
| MDCR | 1,852 | 708 | 1.3 | 0.4 | 17 | 5 | 1.76 (0.43 - 12.0) | 2.24 (0.42 - 11.9) |
| USOC | 173,455 | 82,094 | 130.5 | 52.8 | 926 | 287 | 1.03 (0.86 - 1.24) | 1.09 (0.91 - 1.32) |
| OEHR | 8,303 | 3,561 | 2.6 | 1.0 | 23 | 8 | 1.02 (0.38 - 3.00) | 1.14 (0.41 - 3.19) |
| Meta-analysis | 201,545 | 94,676 | 146.6 | 59.1 | 1,090 | 335 | 1.02 (0.87 - 1.21) | 1.07 (0.90 - 1.28) |
| **PS-stratification / on-treatment TAR** | | | | | | | | |
| CCAE | 9,193 | 5,072 | 7.0 | 3.2 | 54 | 18 | 1.02 (0.59 - 1.84) | 1.03 (0.58 - 1.81) |
| OCEDM | 10,601 | 5,571 | 6.5 | 2.9 | 87 | 25 | 1.49 (0.94 - 2.45) | 1.51 (0.93 - 2.44) |
| MDCR | 1,859 | 741 | 1.3 | 0.4 | 17 | 5 | 1.30 (0.49 - 4.13) | 1.44 (0.49 - 4.18) |
| USOC | 173,486 | 95,798 | 130.5 | 59.8 | 927 | 318 | 1.13 (0.99 - 1.30) | 1.19 (1.03 - 1.36) |
| OEHR | 8,313 | 4,129 | 2.6 | 1.2 | 23 | 8 | 1.02 (0.45 - 2.55) | 1.09 (0.45 - 2.64) |
| Meta-analysis | 201,593 | 110,570 | 146.6 | 67.1 | 1,091 | 369 | 1.14 (1.01 - 1.30) | 1.19 (1.05 - 1.35) |

**Supplementary Table 62:** Exposures, outcomes and risk estimates for Acute myocardial infarction in SGLT2is (T) vs DPP4is (C) new-user cohorts under two different propensity score (PS) adjustment / time-at-risk (TAR) designs across data sources. We report total patient exposure time in 1,000 patient-year (KYr) units, uncalibrated and calibrated hazard ratios (HRs) and their 95% confidence intervals (CIs). Greyed out entries did not pass study diagnostics and are included only for completeness.

| | Patients | | Exposure (KYr) | | Outcomes | | Uncalibrated | Calibrated |
|---|---|---|---|---|---|---|---|---|
| | T | C | T | C | T | C | HR (95% CI) | HR (95% CI) |
| **PS-matching / on-treatment TAR** | | | | | | | | |
| CCAE | 7,445 | 12,129 | 6.1 | 9.5 | 47 | 79 | 1.08 (0.63 - 1.86) | 1.02 (0.59 - 1.77) |
| GDA | 3,365 | 8,557 | 2.8 | 9.0 | 0 | 0 | | |
| OCEDM | 7,925 | 16,520 | 5.3 | 12.7 | 70 | 177 | 1.12 (0.76 - 1.64) | 1.10 (0.75 - 1.62) |
| MDCR | 1,507 | 5,342 | 1.2 | 4.6 | 13 | 85 | 0.74 (0.31 - 1.58) | 0.79 (0.35 - 1.79) |
| MDCD | 1,081 | 3,213 | 0.6 | 2.4 | 12 | 39 | 1.07 (0.44 - 2.44) | 0.99 (0.42 - 2.34) |
| USOC | 128,879 | 272,160 | 106.4 | 244.7 | 755 | 2,242 | 0.83 (0.73 - 0.93) | 0.85 (0.69 - 1.04) |
| OEHR | 6,727 | 14,808 | 2.1 | 8.1 | 16 | 87 | 0.79 (0.39 - 1.53) | 0.71 (0.36 - 1.40) |
| SIDIAP | 1,284 | 5,346 | 1.8 | 11.0 | 5 | 48 | 0.35 (0.08 - 1.06) | 0.34 (0.09 - 1.34) |
| VA | 3,561 | 9,128 | 2.6 | 8.6 | 17 | 37 | 1.33 (0.58 - 2.96) | 1.27 (0.56 - 2.89) |
| Meta-analysis | 158,409 | 338,646 | 126.1 | 301.6 | 935 | 2,794 | 0.86 (0.77 - 0.96) | 0.86 (0.76 - 0.97) |
| **PS-stratification / on-treatment TAR** | | | | | | | | |
| CCAE | 9,193 | 12,152 | 7.0 | 9.5 | 54 | 79 | 0.86 (0.59 - 1.25) | 0.88 (0.60 - 1.27) |
| GDA | 4,434 | 8,797 | 3.5 | 9.3 | 0 | 0 | | |
| OCEDM | 10,601 | 16,845 | 6.5 | 12.9 | 87 | 184 | 1.04 (0.77 - 1.39) | 1.04 (0.77 - 1.41) |
| MDCR | 1,859 | 5,406 | 1.3 | 4.7 | 17 | 88 | 0.72 (0.38 - 1.32) | 0.74 (0.40 - 1.39) |
| MDCD | 1,153 | 3,228 | 0.6 | 2.4 | 12 | 39 | 1.24 (0.59 - 2.46) | 1.10 (0.54 - 2.26) |
| USOC | 173,486 | 272,871 | 130.5 | 245.3 | 927 | 2,256 | 0.93 (0.85 - 1.01) | 0.97 (0.83 - 1.14) |
| OEHR | 8,313 | 14,995 | 2.6 | 8.2 | 23 | 90 | 0.89 (0.52 - 1.45) | 0.81 (0.49 - 1.35) |
| SIDIAP | 1,790 | 5,752 | 2.2 | 12.1 | 6 | 49 | 0.69 (0.24 - 1.68) | 0.68 (0.25 - 1.85) |
| VA | 6,402 | 9,633 | 4.0 | 9.3 | 30 | 41 | 1.24 (0.65 - 2.36) | 1.25 (0.65 - 2.40) |
| Meta-analysis | 201,593 | 316,863 | 146.6 | 275.9 | 1,091 | 2,609 | 0.93 (0.86 - 1.01) | 0.95 (0.84 - 1.08) |

**Supplementary Table 63:** Exposures, outcomes and risk estimates for Acute myocardial infarction in SGLT2is (T) vs SUs (C) new-user cohorts under two different propensity score (PS) adjustment / time-at-risk (TAR) designs across data sources. We report total patient exposure time in 1,000 patient-year (KYr) units, uncalibrated and calibrated hazard ratios (HRs) and their 95% confidence intervals (CIs). Greyed out entries did not pass study diagnostics and are included only for completeness.

| | Patients | | Exposure (KYr) | | Outcomes | | Uncalibrated | Calibrated |
|---|---|---|---|---|---|---|---|---|
| | T | C | T | C | T | C | HR (95% CI) | HR (95% CI) |
| **PS-matching / on-treatment TAR** | | | | | | | | |
| CCAE | 7,603 | 16,222 | 6.0 | 12.4 | 48 | 125 | 0.87 (0.54 - 1.38) | 0.84 (0.53 - 1.35) |
| GDA | 4,184 | 723 | 3.4 | 0.7 | 0 | 0 | | |
| OCEDM | 9,353 | 34,610 | 6.0 | 30.7 | 76 | 614 | 0.75 (0.54 - 1.02) | 0.78 (0.56 - 1.08) |
| MDCR | 1,544 | 7,647 | 1.2 | 6.5 | 16 | 120 | 1.05 (0.49 - 2.11) | 1.17 (0.56 - 2.43) |
| MDCD | 1,135 | 5,804 | 0.6 | 4.0 | 12 | 81 | 0.77 (0.30 - 1.72) | 0.76 (0.32 - 1.82) |
| USOC | 154,829 | 501,492 | 119.9 | 514.6 | 854 | 5,861 | 0.77 (0.70 - 0.85) | 0.80 (0.64 - 0.99) |
| OEHR | 7,261 | 31,099 | 2.2 | 19.0 | 18 | 303 | 0.54 (0.29 - 0.93) | 0.49 (0.27 - 0.88) |
| SIDIAP | 1,392 | 302 | 1.9 | 0.5 | <5 | <5 | 0.17 (0.01 - 8.21) | 0.18 (0.00 - 6.42) |
| VA | 4,465 | 24,654 | 3.2 | 21.1 | 21 | 143 | 1.23 (0.65 - 2.23) | 1.26 (0.68 - 2.33) |
| Meta-analysis | 181,725 | 596,874 | 135.9 | 587.3 | 1,024 | 7,104 | 0.77 (0.70 - 0.84) | 0.77 (0.65 - 0.92) |
| **PS-stratification / on-treatment TAR** | | | | | | | | |
| CCAE | 9,193 | 16,529 | 7.0 | 12.6 | 54 | 129 | 0.78 (0.55 - 1.10) | 0.76 (0.54 - 1.08) |
| GDA | 4,337 | 1,235 | 3.5 | 1.3 | 0 | 0 | | |
| OCEDM | 10,601 | 35,263 | 6.5 | 31.2 | 87 | 632 | 0.80 (0.62 - 1.02) | 0.80 (0.62 - 1.03) |
| MDCR | 1,859 | 8,152 | 1.3 | 6.9 | 17 | 133 | 0.82 (0.44 - 1.46) | 0.80 (0.44 - 1.46) |
| MDCD | 1,154 | 6,044 | 0.6 | 4.2 | 12 | 87 | 1.08 (0.54 - 2.00) | 1.05 (0.54 - 2.04) |
| USOC | 173,486 | 509,521 | 130.5 | 522.2 | 927 | 6,070 | 0.81 (0.75 - 0.87) | 0.84 (0.69 - 1.01) |
| OEHR | 8,313 | 32,721 | 2.6 | 20.2 | 23 | 332 | 0.58 (0.36 - 0.89) | 0.54 (0.34 - 0.88) |
| SIDIAP | 1,776 | 583 | 2.2 | 1.3 | 6 | 5 | 0.56 (0.07 - 3.59) | 0.59 (0.08 - 4.17) |
| VA | 6,402 | 36,055 | 4.0 | 33.9 | 30 | 273 | 1.23 (0.73 - 2.01) | 1.25 (0.75 - 2.08) |
| Meta-analysis | 201,593 | 594,034 | 146.6 | 586.3 | 1,091 | 7,163 | 0.80 (0.74 - 0.86) | 0.81 (0.69 - 0.94) |

**Supplementary Table 64:** Exposures, outcomes and risk estimates for Acute myocardial infarction in GLP1-RAs (T) vs DPP4is (C) new-user cohorts under two different propensity score (PS) adjustment / time-at-risk (TAR) designs across data sources. We report total patient exposure time in 1,000 patient-year (KYr) units, uncalibrated and calibrated hazard ratios (HRs) and their 95% confidence intervals (CIs). Greyed out entries did not pass study diagnostics and are included only for completeness.

| | Patients | | Exposure (KYr) | | Outcomes | | Uncalibrated | Calibrated |
|---|---|---|---|---|---|---|---|---|
| | T | C | T | C | T | C | HR (95% CI) | HR (95% CI) |
| **PS-matching / on-treatment TAR** | | | | | | | | |
| CCAE | 5,211 | 21,004 | 3.4 | 17.2 | 19 | 135 | 0.98 (0.51 - 1.80) | 1.00 (0.54 - 1.89) |
| OCEDM | 4,694 | 21,852 | 2.6 | 17.5 | 24 | 231 | 0.84 (0.47 - 1.44) | 0.83 (0.47 - 1.46) |
| MDCR | 918 | 9,149 | 0.6 | 8.8 | 7 | 138 | 0.59 (0.17 - 1.61) | 0.52 (0.17 - 1.61) |
| USOC | 68,496 | 272,508 | 48.0 | 245.5 | 265 | 2,216 | 0.87 (0.73 - 1.03) | 0.85 (0.70 - 1.02) |
| OEHR | 3,452 | 17,135 | 1.0 | 9.2 | 7 | 98 | 0.78 (0.26 - 1.93) | 0.66 (0.24 - 1.81) |
| Meta-analysis | 81,853 | 332,499 | 55.1 | 289.5 | 315 | 2,680 | 0.88 (0.75 - 1.02) | 0.85 (0.72 - 1.02) |
| **PS-stratification / on-treatment TAR** | | | | | | | | |
| CCAE | 6,605 | 21,112 | 4.1 | 17.3 | 22 | 137 | 0.89 (0.52 - 1.45) | 0.89 (0.54 - 1.49) |
| OCEDM | 6,199 | 22,466 | 3.3 | 18.1 | 29 | 249 | 0.81 (0.51 - 1.24) | 0.79 (0.51 - 1.23) |
| MDCR | 1,000 | 9,810 | 0.6 | 9.4 | 7 | 155 | 0.67 (0.27 - 1.45) | 0.60 (0.26 - 1.39) |
| USOC | 95,937 | 274,790 | 59.9 | 247.5 | 319 | 2,272 | 0.87 (0.76 - 1.00) | 0.86 (0.71 - 1.03) |
| OEHR | 4,293 | 17,251 | 1.2 | 9.3 | 8 | 101 | 0.78 (0.32 - 1.64) | 0.64 (0.28 - 1.45) |
| Meta-analysis | 113,034 | 335,619 | 68.4 | 292.1 | 378 | 2,759 | 0.87 (0.77 - 0.98) | 0.85 (0.72 - 0.99) |

**Supplementary Table 65:** Exposures, outcomes and risk estimates for Acute myocardial infarction in GLP1-RAs (T) vs SUs (C) new-user cohorts under two different propensity score (PS) adjustment / time-at-risk (TAR) designs across data sources. We report total patient exposure time in 1,000 patient-year (KYr) units, uncalibrated and calibrated hazard ratios (HRs) and their 95% confidence intervals (CIs). Greyed out entries did not pass study diagnostics and are included only for completeness.

| | Patients | | Exposure (KYr) | | Outcomes | | Uncalibrated | Calibrated |
|---|---|---|---|---|---|---|---|---|
| | T | C | T | C | T | C | HR (95% CI) | HR (95% CI) |
| **PS-matching / on-treatment TAR** | | | | | | | | |
| CCAE | 5,530 | 31,661 | 3.6 | 24.7 | 21 | 232 | 0.78 (0.43 - 1.36) | 0.74 (0.41 - 1.32) |
| OCEDM | 5,221 | 42,347 | 2.9 | 38.2 | 25 | 661 | 0.58 (0.34 - 0.92) | 0.58 (0.35 - 0.96) |
| MDCR | 990 | 14,802 | 0.6 | 14.3 | 8 | 253 | 0.63 (0.21 - 1.51) | 0.51 (0.19 - 1.38) |
| USOC | 80,833 | 490,831 | 52.9 | 505.5 | 292 | 5,566 | 0.74 (0.64 - 0.85) | 0.72 (0.58 - 0.89) |
| OEHR | 3,665 | 35,550 | 1.1 | 21.4 | 7 | 330 | 0.40 (0.15 - 0.87) | 0.37 (0.15 - 0.88) |
| Meta-analysis | 96,239 | 615,191 | 61.0 | 604.0 | 353 | 7,042 | 0.71 (0.62 - 0.82) | 0.69 (0.56 - 0.85) |
| **PS-stratification / on-treatment TAR** | | | | | | | | |
| CCAE | 6,948 | 32,445 | 4.3 | 25.2 | 22 | 236 | 0.70 (0.42 - 1.12) | 0.69 (0.42 - 1.13) |
| OCEDM | 6,199 | 48,951 | 3.3 | 45.1 | 29 | 875 | 0.64 (0.42 - 0.94) | 0.63 (0.42 - 0.95) |
| MDCR | 1,069 | 19,142 | 0.6 | 18.2 | 8 | 354 | 0.74 (0.32 - 1.47) | 0.61 (0.28 - 1.30) |
| USOC | 95,937 | 515,263 | 59.9 | 530.7 | 319 | 6,171 | 0.71 (0.63 - 0.80) | 0.70 (0.55 - 0.88) |
| OEHR | 4,294 | 40,322 | 1.2 | 24.7 | 8 | 407 | 0.46 (0.20 - 0.92) | 0.39 (0.18 - 0.82) |
| Meta-analysis | 102,885 | 547,708 | 64.1 | 555.9 | 341 | 6,407 | 0.71 (0.63 - 0.80) | 0.70 (0.56 - 0.87) |

**Supplementary Table 66:** Exposures, outcomes and risk estimates for Acute myocardial infarction in DPP4is (T) vs SUs (C) new-user cohorts under two different propensity score (PS) adjustment / time-at-risk (TAR) designs across data sources. We report total patient exposure time in 1,000 patient-year (KYr) units, uncalibrated and calibrated hazard ratios (HRs) and their 95% confidence intervals (CIs). Greyed out entries did not pass study diagnostics and are included only for completeness.

| | Patients | | Exposure (KYr) | | Outcomes | | Uncalibrated | Calibrated |
|---|---|---|---|---|---|---|---|---|
| | T | C | T | C | T | C | HR (95% CI) | HR (95% CI) |
| **PS-matching / on-treatment TAR** | | | | | | | | |
| CCAE | 18,304 | 30,604 | 14.9 | 23.6 | 127 | 216 | 0.95 (0.68 - 1.30) | 0.96 (0.70 - 1.33) |
| GDA | 10,408 | 1,974 | 11.3 | 2.0 | 0 | 0 | | |
| OCEDM | 21,253 | 48,925 | 17.0 | 45.1 | 232 | 873 | 0.81 (0.66 - 0.99) | 0.86 (0.70 - 1.06) |
| MDCR | 9,146 | 17,469 | 8.8 | 16.4 | 143 | 320 | 0.83 (0.63 - 1.10) | 0.79 (0.60 - 1.05) |
| MDCD | 3,852 | 8,599 | 2.9 | 6.1 | 43 | 125 | 0.73 (0.45 - 1.14) | 0.76 (0.48 - 1.22) |
| IMRD | 3,388 | 407 | 5.7 | 0.6 | 0 | 0 | | |
| USOC | 267,334 | 515,266 | 239.1 | 530.8 | 2,212 | 6,170 | 0.89 (0.83 - 0.95) | 0.89 (0.78 - 1.00) |
| OEHR | 16,282 | 40,290 | 8.6 | 24.7 | 99 | 407 | 0.79 (0.59 - 1.06) | 0.77 (0.58 - 1.04) |
| SIDIAP | 7,365 | 1,710 | 19.5 | 4.3 | 72 | 14 | 1.10 (0.54 - 2.39) | 1.07 (0.51 - 2.27) |
| VA | 9,639 | 60,093 | 9.8 | 62.9 | 44 | 461 | 0.72 (0.47 - 1.07) | 0.73 (0.49 - 1.11) |
| Meta-analysis | 353,175 | 722,956 | 320.5 | 713.9 | 2,972 | 8,586 | 0.87 (0.82 - 0.92) | 0.87 (0.78 - 0.97) |
| **PS-stratification / on-treatment TAR** | | | | | | | | |
| CCAE | 21,112 | 30,744 | 17.3 | 23.7 | 137 | 218 | 0.95 (0.76 - 1.19) | 0.96 (0.76 - 1.20) |
| GDA | 10,428 | 2,365 | 11.3 | 2.5 | 0 | 0 | | |
| OCEDM | 22,470 | 48,955 | 18.1 | 45.1 | 249 | 875 | 0.82 (0.70 - 0.94) | 0.83 (0.71 - 0.97) |
| MDCR | 9,810 | 17,584 | 9.4 | 16.5 | 155 | 322 | 0.93 (0.76 - 1.14) | 0.93 (0.76 - 1.14) |
| MDCD | 4,022 | 8,634 | 3.1 | 6.1 | 47 | 125 | 0.85 (0.59 - 1.21) | 0.87 (0.61 - 1.25) |
| IMRD | 4,024 | 846 | 6.5 | 1.5 | 0 | 0 | | |
| USOC | 274,802 | 515,297 | 247.6 | 530.8 | 2,272 | 6,172 | 0.86 (0.82 - 0.90) | 0.85 (0.74 - 0.98) |
| OEHR | 17,251 | 40,321 | 9.3 | 24.7 | 101 | 407 | 0.74 (0.58 - 0.92) | 0.77 (0.61 - 0.97) |
| SIDIAP | 7,523 | 2,232 | 19.6 | 5.9 | 74 | 19 | 1.14 (0.64 - 2.12) | 1.08 (0.59 - 1.96) |
| VA | 10,323 | 91,068 | 10.3 | 101.0 | 46 | 773 | 0.68 (0.48 - 0.95) | 0.71 (0.51 - 1.00) |
| Meta-analysis | 349,467 | 661,535 | 304.7 | 646.9 | 2,961 | 8,119 | 0.86 (0.82 - 0.89) | 0.86 (0.78 - 0.94) |

**Supplementary Table 67:** Exposures, outcomes and risk estimates for Stroke in SGLT2is (T) vs GLP1-RAs (C) new-user cohorts under two different propensity score (PS) adjustment / time-at-risk (TAR) designs across data sources. We report total patient exposure time in 1,000 patient-year (KYr) units, uncalibrated and calibrated hazard ratios (HRs) and their 95% confidence intervals (CIs). Greyed out entries did not pass study diagnostics and are included only for completeness.

| | Patients | | Exposure (KYr) | | Outcomes | | Uncalibrated | Calibrated |
|---|---|---|---|---|---|---|---|---|
| | T | C | T | C | T | C | HR (95% CI) | HR (95% CI) |
| **PS-matching / on-treatment TAR** | | | | | | | | |
| CCAE | 9,658 | 4,386 | 7.4 | 2.9 | 38 | 11 | 0.51 (0.21 - 1.30) | 0.50 (0.20 - 1.26) |
| OCEDM | 10,964 | 4,837 | 6.6 | 2.6 | 84 | 29 | 0.74 (0.42 - 1.33) | 0.77 (0.43 - 1.36) |
| MDCR | 1,911 | 700 | 1.4 | 0.4 | 14 | <5 | 0.28 (0.04 - 1.53) | 0.36 (0.06 - 2.28) |
| USOC | 183,553 | 84,139 | 138.6 | 54.1 | 831 | 338 | 0.89 (0.75 - 1.06) | 0.94 (0.78 - 1.12) |
| OEHR | 8,720 | 3,666 | 2.7 | 1.0 | 19 | 6 | 1.73 (0.51 - 7.95) | 1.95 (0.49 - 7.72) |
| Meta-analysis | 212,895 | 97,028 | 155.4 | 60.6 | 972 | 384 | 0.87 (0.74 - 1.02) | 0.91 (0.77 - 1.08) |
| **PS-stratification / on-treatment TAR** | | | | | | | | |
| CCAE | 9,662 | 5,177 | 7.4 | 3.3 | 38 | 15 | 1.07 (0.58 - 2.09) | 1.08 (0.57 - 2.05) |
| OCEDM | 10,969 | 5,621 | 6.6 | 2.9 | 85 | 32 | 0.95 (0.62 - 1.50) | 0.96 (0.62 - 1.50) |
| MDCR | 1,919 | 729 | 1.4 | 0.4 | 14 | <5 | 0.76 (0.25 - 2.88) | 0.83 (0.24 - 2.85) |
| USOC | 183,570 | 97,627 | 138.6 | 61.0 | 831 | 376 | 0.87 (0.77 - 1.00) | 0.92 (0.80 - 1.05) |
| OEHR | 8,732 | 4,211 | 2.7 | 1.2 | 19 | 6 | 1.36 (0.54 - 3.93) | 1.45 (0.53 - 3.97) |
| Meta-analysis | 212,933 | 112,636 | 155.4 | 68.4 | 973 | 429 | 0.89 (0.79 - 1.01) | 0.93 (0.82 - 1.05) |

**Supplementary Table 68:** Exposures, outcomes and risk estimates for Stroke in SGLT2is (T) vs DPP4is (C) new-user cohorts under two different propensity score (PS) adjustment / time-at-risk (TAR) designs across data sources. We report total patient exposure time in 1,000 patient-year (KYr) units, uncalibrated and calibrated hazard ratios (HRs) and their 95% confidence intervals (CIs). Greyed out entries did not pass study diagnostics and are included only for completeness.

| | Patients | | Exposure (KYr) | | Outcomes | | Uncalibrated | Calibrated |
|---|---|---|---|---|---|---|---|---|
| | T | C | T | C | T | C | HR (95% CI) | HR (95% CI) |
| **PS-matching / on-treatment TAR** | | | | | | | | |
| CCAE | 7,664 | 12,416 | 6.2 | 9.9 | 34 | 64 | 0.58 (0.30 - 1.05) | 0.54 (0.29 - 1.02) |
| GDA | 3,365 | 8,557 | 2.8 | 9.0 | 0 | 0 | | |
| OCEDM | 7,973 | 16,284 | 5.3 | 12.4 | 72 | 239 | 0.96 (0.67 - 1.35) | 0.94 (0.66 - 1.34) |
| MDCR | 1,534 | 5,251 | 1.2 | 4.5 | 12 | 93 | 0.40 (0.13 - 0.98) | 0.43 (0.16 - 1.16) |
| MDCD | 1,096 | 3,186 | 0.6 | 2.4 | 10 | 52 | 1.45 (0.60 - 3.28) | 1.34 (0.57 - 3.15) |
| USOC | 131,975 | 274,309 | 110.1 | 247.7 | 679 | 2,558 | 0.79 (0.70 - 0.89) | 0.81 (0.66 - 0.99) |
| OEHR | 6,954 | 15,073 | 2.2 | 8.3 | 18 | 72 | 1.43 (0.66 - 2.98) | 1.28 (0.60 - 2.72) |
| SIDIAP | 1,448 | 5,702 | 2.1 | 11.8 | <5 | 64 | 0.58 (0.13 - 1.72) | 0.57 (0.15 - 2.20) |
| VA | 3,766 | 9,330 | 2.8 | 8.8 | <10 | 29 | 1.09 (0.28 - 3.62) | 1.05 (0.29 - 3.76) |
| Meta-analysis | 162,410 | 341,551 | 130.5 | 305.8 | <840 | 3,171 | 0.83 (0.69 - 1.01) | 0.83 (0.68 - 1.01) |
| **PS-stratification / on-treatment TAR** | | | | | | | | |
| CCAE | 9,662 | 12,459 | 7.4 | 9.9 | 38 | 64 | 0.92 (0.59 - 1.40) | 0.93 (0.60 - 1.44) |
| GDA | 4,434 | 8,797 | 3.5 | 9.3 | 0 | 0 | | |
| OCEDM | 10,969 | 16,681 | 6.6 | 12.8 | 85 | 254 | 0.87 (0.65 - 1.15) | 0.87 (0.66 - 1.16) |
| MDCR | 1,919 | 5,346 | 1.4 | 4.6 | 14 | 95 | 0.65 (0.34 - 1.18) | 0.67 (0.36 - 1.25) |
| MDCD | 1,171 | 3,201 | 0.7 | 2.4 | 10 | 52 | 0.95 (0.44 - 1.90) | 0.85 (0.41 - 1.77) |
| USOC | 183,570 | 274,921 | 138.6 | 248.2 | 831 | 2,578 | 0.82 (0.75 - 0.89) | 0.86 (0.73 - 1.01) |
| OEHR | 8,732 | 15,259 | 2.7 | 8.4 | 19 | 75 | 1.44 (0.81 - 2.47) | 1.32 (0.75 - 2.31) |
| SIDIAP | 2,219 | 6,058 | 2.8 | 12.8 | 5 | 72 | 0.45 (0.14 - 1.18) | 0.45 (0.15 - 1.32) |
| VA | 6,992 | 9,823 | 4.4 | 9.5 | 16 | 36 | 0.97 (0.43 - 2.13) | 0.98 (0.44 - 2.19) |
| Meta-analysis | 212,933 | 319,320 | 155.4 | 279.2 | 973 | 2,971 | 0.87 (0.75 - 1.01) | 0.89 (0.74 - 1.06) |

**Supplementary Table 69:** Exposures, outcomes and risk estimates for Stroke in SGLT2is (T) vs SUs (C) new-user cohorts under two different propensity score (PS) adjustment / time-at-risk (TAR) designs across data sources. We report total patient exposure time in 1,000 patient-year (KYr) units, uncalibrated and calibrated hazard ratios (HRs) and their 95% confidence intervals (CIs). Greyed out entries did not pass study diagnostics and are included only for completeness.

| | Patients | | Exposure (KYr) | | Outcomes | | Uncalibrated | Calibrated |
|---|---|---|---|---|---|---|---|---|
| | T | C | T | C | T | C | HR (95% CI) | HR (95% CI) |
| **PS-matching / on-treatment TAR** | | | | | | | | |
| CCAE | 7,885 | 16,681 | 6.2 | 12.9 | 35 | 100 | 0.82 (0.47 - 1.41) | 0.80 (0.46 - 1.39) |
| GDA | 4,184 | 723 | 3.4 | 0.7 | 0 | 0 | | |
| OCEDM | 9,556 | 34,467 | 6.0 | 30.5 | 72 | 586 | 0.98 (0.72 - 1.33) | 1.03 (0.75 - 1.40) |
| MDCR | 1,585 | 7,548 | 1.2 | 6.5 | 13 | 145 | 0.39 (0.14 - 0.88) | 0.43 (0.17 - 1.06) |
| MDCD | 1,145 | 5,692 | 0.6 | 3.9 | 10 | 79 | 0.89 (0.37 - 1.92) | 0.88 (0.38 - 2.01) |
| USOC | 161,306 | 508,864 | 126.0 | 524.4 | 772 | 6,174 | 0.77 (0.70 - 0.85) | 0.80 (0.64 - 0.99) |
| OEHR | 7,554 | 31,707 | 2.4 | 19.5 | 19 | 226 | 1.04 (0.54 - 1.87) | 0.95 (0.51 - 1.78) |
| SIDIAP | 1,601 | 308 | 2.3 | 0.6 | <5 | 0 | | |
| VA | 4,778 | 25,940 | 3.5 | 22.2 | 12 | 106 | 0.91 (0.44 - 1.75) | 0.93 (0.46 - 1.85) |
| Meta-analysis | 189,031 | 604,959 | 142.4 | 597.7 | 921 | 7,310 | 0.80 (0.71 - 0.91) | 0.81 (0.67 - 0.98) |
| **PS-stratification / on-treatment TAR** | | | | | | | | |
| CCAE | 9,662 | 16,907 | 7.4 | 13.0 | 38 | 100 | 0.83 (0.55 - 1.24) | 0.82 (0.54 - 1.23) |
| GDA | 4,337 | 1,235 | 3.5 | 1.3 | 0 | 0 | | |
| OCEDM | 10,969 | 35,168 | 6.6 | 31.1 | 85 | 612 | 0.91 (0.70 - 1.17) | 0.91 (0.70 - 1.18) |
| MDCR | 1,919 | 8,109 | 1.4 | 6.9 | 14 | 160 | 0.68 (0.36 - 1.22) | 0.66 (0.36 - 1.22) |
| MDCD | 1,172 | 5,980 | 0.7 | 4.1 | 10 | 86 | 1.01 (0.48 - 1.93) | 0.99 (0.49 - 1.99) |
| USOC | 183,570 | 516,987 | 138.6 | 532.0 | 831 | 6,403 | 0.73 (0.67 - 0.79) | 0.76 (0.63 - 0.92) |
| OEHR | 8,732 | 33,298 | 2.7 | 20.7 | 19 | 247 | 0.90 (0.53 - 1.46) | 0.85 (0.50 - 1.43) |
| SIDIAP | 2,206 | 600 | 2.8 | 1.3 | 5 | <5 | 1.98 (0.26 - 12.4) | 2.08 (0.30 - 14.5) |
| VA | 6,992 | 37,498 | 4.4 | 35.3 | 16 | 197 | 0.68 (0.34 - 1.28) | 0.69 (0.36 - 1.34) |
| Meta-analysis | 212,933 | 602,360 | 155.4 | 596.8 | 973 | 7,362 | 0.76 (0.69 - 0.84) | 0.77 (0.65 - 0.92) |

**Supplementary Table 70:** Exposures, outcomes and risk estimates for Stroke in GLP1-RAs (T) vs DPP4is (C) new-user cohorts under two different propensity score (PS) adjustment / time-at-risk (TAR) designs across data sources. We report total patient exposure time in 1,000 patient-year (KYr) units, uncalibrated and calibrated hazard ratios (HRs) and their 95% confidence intervals (CIs). Greyed out entries did not pass study diagnostics and are included only for completeness.

| | Patients | | Exposure (KYr) | | Outcomes | | Uncalibrated | Calibrated |
|---|---|---|---|---|---|---|---|---|
| | T | C | T | C | T | C | HR (95% CI) | HR (95% CI) |
| **PS-matching / on-treatment TAR** | | | | | | | | |
| CCAE | 5,374 | 21,649 | 3.5 | 18.0 | 15 | 107 | 0.76 (0.37 - 1.44) | 0.77 (0.39 - 1.52) |
| OCEDM | 4,722 | 21,900 | 2.6 | 17.6 | 34 | 303 | 0.81 (0.51 - 1.26) | 0.80 (0.51 - 1.27) |
| MDCR | 908 | 9,007 | 0.6 | 8.6 | 6 | 177 | 0.53 (0.18 - 1.28) | 0.46 (0.17 - 1.25) |
| USOC | 69,620 | 274,462 | 48.9 | 248.3 | 325 | 2,515 | 0.97 (0.83 - 1.13) | 0.94 (0.79 - 1.12) |
| OEHR | 3,513 | 17,436 | 1.0 | 9.4 | 7 | 85 | 1.15 (0.37 - 2.89) | 0.97 (0.35 - 2.71) |
| Meta-analysis | 83,229 | 335,447 | 56.1 | 293.3 | 381 | 3,010 | 0.94 (0.82 - 1.09) | 0.92 (0.78 - 1.09) |
| **PS-stratification / on-treatment TAR** | | | | | | | | |
| CCAE | 6,757 | 21,772 | 4.1 | 18.1 | 18 | 107 | 0.86 (0.47 - 1.48) | 0.87 (0.49 - 1.53) |
| OCEDM | 6,253 | 22,330 | 3.3 | 17.9 | 36 | 317 | 0.73 (0.48 - 1.07) | 0.71 (0.48 - 1.06) |
| MDCR | 987 | 9,741 | 0.6 | 9.3 | 6 | 195 | 0.61 (0.23 - 1.31) | 0.54 (0.23 - 1.29) |
| USOC | 97,769 | 276,869 | 61.1 | 250.5 | 376 | 2,595 | 0.92 (0.81 - 1.04) | 0.91 (0.76 - 1.09) |
| OEHR | 4,377 | 17,560 | 1.3 | 9.5 | 7 | 92 | 1.31 (0.54 - 2.73) | 1.08 (0.48 - 2.45) |
| Meta-analysis | 115,156 | 338,531 | 69.8 | 296.0 | 437 | 3,111 | 0.91 (0.81 - 1.02) | 0.89 (0.77 - 1.03) |

**Supplementary Table 71:** Exposures, outcomes and risk estimates for Stroke in GLP1-RAs (T) vs SUs (C) new-user cohorts under two different propensity score (PS) adjustment / time-at-risk (TAR) designs across data sources. We report total patient exposure time in 1,000 patient-year (KYr) units, uncalibrated and calibrated hazard ratios (HRs) and their 95% confidence intervals (CIs). Greyed out entries did not pass study diagnostics and are included only for completeness.

| | Patients | | Exposure (KYr) | | Outcomes | | Uncalibrated | Calibrated |
|---|---|---|---|---|---|---|---|---|
| | T | C | T | C | T | C | HR (95% CI) | HR (95% CI) |
| **PS-matching / on-treatment TAR** | | | | | | | | |
| CCAE | 5,699 | 32,641 | 3.7 | 25.7 | 20 | 173 | 0.76 (0.39 - 1.38) | 0.72 (0.38 - 1.35) |
| OCEDM | 5,259 | 42,410 | 2.8 | 38.1 | 33 | 679 | 0.94 (0.60 - 1.43) | 0.95 (0.62 - 1.47) |
| MDCR | 978 | 14,526 | 0.6 | 14.0 | 6 | 301 | 0.53 (0.18 - 1.24) | 0.43 (0.16 - 1.14) |
| USOC | 82,560 | 497,956 | 54.1 | 514.8 | 347 | 5,843 | 0.81 (0.70 - 0.93) | 0.79 (0.64 - 0.97) |
| OEHR | 3,746 | 36,039 | 1.1 | 21.8 | 7 | 233 | 0.89 (0.33 - 1.98) | 0.82 (0.33 - 2.00) |
| Meta-analysis | 98,242 | 623,572 | 62.3 | 614.3 | 413 | 7,229 | 0.81 (0.72 - 0.92) | 0.78 (0.64 - 0.96) |
| **PS-stratification / on-treatment TAR** | | | | | | | | |
| CCAE | 7,119 | 33,346 | 4.4 | 26.1 | 20 | 185 | 0.82 (0.47 - 1.37) | 0.81 (0.48 - 1.38) |
| OCEDM | 6,253 | 48,963 | 3.3 | 45.0 | 36 | 909 | 0.90 (0.61 - 1.29) | 0.90 (0.62 - 1.30) |
| MDCR | 1,058 | 19,052 | 0.6 | 18.1 | 6 | 456 | 0.54 (0.21 - 1.16) | 0.45 (0.19 - 1.05) |
| USOC | 97,769 | 522,751 | 61.1 | 540.5 | 376 | 6,508 | 0.80 (0.72 - 0.90) | 0.79 (0.63 - 0.99) |
| OEHR | 4,378 | 40,993 | 1.3 | 25.2 | 7 | 300 | 0.78 (0.32 - 1.60) | 0.65 (0.29 - 1.44) |
| Meta-analysis | 104,888 | 556,097 | 65.5 | 566.6 | 396 | 6,693 | 0.81 (0.72 - 0.90) | 0.79 (0.63 - 0.98) |

**Supplementary Table 72:** Exposures, outcomes and risk estimates for Stroke in DPP4is (T) vs SUs (C) new-user cohorts under two different propensity score (PS) adjustment / time-at-risk (TAR) designs across data sources. We report total patient exposure time in 1,000 patient-year (KYr) units, uncalibrated and calibrated hazard ratios (HRs) and their 95% confidence intervals (CIs). Greyed out entries did not pass study diagnostics and are included only for completeness.

| | Patients | | Exposure (KYr) | | Outcomes | | Uncalibrated | Calibrated |
|---|---|---|---|---|---|---|---|---|
| | T | C | T | C | T | C | HR (95% CI) | HR (95% CI) |
| **PS-matching / on-treatment TAR** | | | | | | | | |
| CCAE | 18,883 | 31,456 | 15.7 | 24.4 | 94 | 176 | 0.90 (0.62 - 1.30) | 0.92 (0.63 - 1.33) |
| GDA | 10,408 | 1,974 | 11.3 | 2.0 | 0 | 0 | | |
| OCEDM | 21,131 | 48,934 | 16.9 | 45.0 | 304 | 906 | 1.02 (0.86 - 1.22) | 1.08 (0.90 - 1.30) |
| MDCR | 9,136 | 17,372 | 8.7 | 16.3 | 186 | 404 | 0.83 (0.65 - 1.07) | 0.79 (0.62 - 1.02) |
| MDCD | 3,826 | 8,480 | 2.9 | 6.0 | 53 | 132 | 0.88 (0.55 - 1.37) | 0.92 (0.59 - 1.45) |
| IMRD | 3,388 | 407 | 5.7 | 0.6 | 0 | 0 | | |
| USOC | 269,428 | 522,756 | 242.0 | 540.5 | 2,517 | 6,509 | 0.90 (0.85 - 0.96) | 0.90 (0.80 - 1.02) |
| OEHR | 16,561 | 40,955 | 8.8 | 25.2 | 86 | 300 | 0.94 (0.68 - 1.29) | 0.92 (0.67 - 1.26) |
| SIDIAP | 7,715 | 1,757 | 20.4 | 4.5 | 97 | 16 | 1.56 (0.71 - 3.91) | 1.52 (0.65 - 3.56) |
| VA | 9,845 | 62,310 | 10.0 | 65.4 | 40 | 339 | 0.81 (0.50 - 1.27) | 0.83 (0.52 - 1.32) |
| Meta-analysis | 356,525 | 734,020 | 325.4 | 727.4 | 3,377 | 8,782 | 0.91 (0.86 - 0.97) | 0.91 (0.82 - 1.02) |
| **PS-stratification / on-treatment TAR** | | | | | | | | |
| CCAE | 21,772 | 31,598 | 18.1 | 24.5 | 107 | 177 | 0.90 (0.70 - 1.16) | 0.91 (0.70 - 1.17) |
| GDA | 10,428 | 2,365 | 11.3 | 2.5 | 0 | 0 | | |
| OCEDM | 22,333 | 48,967 | 17.9 | 45.1 | 317 | 909 | 0.97 (0.85 - 1.11) | 0.99 (0.86 - 1.14) |
| MDCR | 9,741 | 17,495 | 9.3 | 16.4 | 195 | 407 | 0.89 (0.74 - 1.06) | 0.89 (0.74 - 1.06) |
| MDCD | 3,996 | 8,512 | 3.1 | 6.0 | 56 | 132 | 0.94 (0.67 - 1.30) | 0.96 (0.69 - 1.34) |
| IMRD | 4,024 | 846 | 6.5 | 1.5 | 0 | 0 | | |
| USOC | 276,882 | 522,785 | 250.5 | 540.5 | 2,595 | 6,509 | 0.89 (0.85 - 0.93) | 0.88 (0.77 - 1.01) |
| OEHR | 17,560 | 40,992 | 9.5 | 25.2 | 92 | 300 | 0.93 (0.73 - 1.19) | 0.97 (0.76 - 1.24) |
| SIDIAP | 7,892 | 2,289 | 20.6 | 6.1 | 97 | 20 | 1.17 (0.68 - 2.10) | 1.11 (0.63 - 1.94) |
| VA | 10,531 | 94,263 | 10.5 | 104.7 | 42 | 555 | 0.87 (0.60 - 1.24) | 0.91 (0.63 - 1.31) |
| Meta-analysis | 352,284 | 670,349 | 308.4 | 657.8 | 3,362 | 8,434 | 0.90 (0.86 - 0.94) | 0.90 (0.81 - 0.99) |

**Supplementary Table 73:** Exposures, outcomes and risk estimates for Sudden cardiac death in SGLT2is (T) vs GLP1-RAs (C) new-user cohorts under two different propensity score (PS) adjustment / time-at-risk (TAR) designs across data sources. We report total patient exposure time in 1,000 patient-year (KYr) units, uncalibrated and calibrated hazard ratios (HRs) and their 95% confidence intervals (CIs). Greyed out entries did not pass study diagnostics and are included only for completeness.

| | Patients | | Exposure (KYr) | | Outcomes | | Uncalibrated | Calibrated |
|---|---|---|---|---|---|---|---|---|
| | T | C | T | C | T | C | HR (95% CI) | HR (95% CI) |
| **PS-matching / on-treatment TAR** | | | | | | | | |
| CCAE | 10,084 | 4,579 | 7.7 | 3.0 | 6 | 5 | 1.29 (0.24 - 9.60) | 1.25 (0.20 - 7.91) |
| OCEDM | 11,546 | 5,065 | 7.1 | 2.7 | 17 | 7 | 0.74 (0.23 - 2.60) | 0.77 (0.23 - 2.58) |
| MDCR | 1,997 | 739 | 1.4 | 0.4 | 9 | <5 | | |
| USOC | 195,288 | 89,502 | 146.8 | 57.5 | 214 | 67 | 1.03 (0.72 - 1.51) | 1.09 (0.75 - 1.59) |
| OEHR | 8,953 | 3,776 | 2.8 | 1.0 | <5 | <5 | 0.64 (0.07 - 5.56) | 0.72 (0.08 - 6.26) |
| Meta-analysis | 206,834 | 94,567 | 153.8 | 60.3 | 231 | 74 | 1.00 (0.70 - 1.44) | 1.05 (0.73 - 1.51) |
| **PS-stratification / on-treatment TAR** | | | | | | | | |
| CCAE | 10,088 | 5,427 | 7.7 | 3.5 | 6 | 5 | 0.64 (0.18 - 2.35) | 0.65 (0.18 - 2.32) |
| OCEDM | 11,552 | 5,880 | 7.1 | 3.1 | 17 | 7 | 0.93 (0.38 - 2.51) | 0.94 (0.36 - 2.42) |
| MDCR | 2,005 | 771 | 1.4 | 0.4 | 9 | <5 | 2.18 (0.39 - 40.7) | 2.40 (0.24 - 24.5) |
| USOC | 195,307 | 103,692 | 146.8 | 64.7 | 215 | 75 | 1.08 (0.82 - 1.43) | 1.13 (0.85 - 1.50) |
| OEHR | 8,965 | 4,349 | 2.8 | 1.2 | <5 | <5 | 0.63 (0.13 - 3.32) | 0.67 (0.13 - 3.39) |
| Meta-analysis | 206,859 | 109,572 | 153.8 | 67.8 | 232 | 82 | 1.06 (0.81 - 1.39) | 1.11 (0.84 - 1.45) |

**Supplementary Table 74:** Exposures, outcomes and risk estimates for Sudden cardiac death in SGLT2is (T) vs DPP4is (C) new-user cohorts under two different propensity score (PS) adjustment / time-at-risk (TAR) designs across data sources. We report total patient exposure time in 1,000 patient-year (KYr) units, uncalibrated and calibrated hazard ratios (HRs) and their 95% confidence intervals (CIs). Greyed out entries did not pass study diagnostics and are included only for completeness.

| | Patients | | Exposure (KYr) | | Outcomes | | Uncalibrated | Calibrated |
|---|---|---|---|---|---|---|---|---|
| | T | C | T | C | T | C | HR (95% CI) | HR (95% CI) |
| **PS-matching / on-treatment TAR** | | | | | | | | |
| CCAE | 8,032 | 13,091 | 6.5 | 10.4 | 5 | 13 | 0.47 (0.07 - 2.14) | 0.44 (0.08 - 2.50) |
| GDA | 3,365 | 8,557 | 2.8 | 9.0 | 0 | 0 | | |
| OCEDM | 8,440 | 17,580 | 5.7 | 13.5 | 12 | 90 | 0.48 (0.20 - 1.02) | 0.47 (0.21 - 1.06) |
| MDCR | 1,607 | 5,722 | 1.3 | 4.9 | 8 | 23 | 1.76 (0.36 - 6.63) | 1.88 (0.44 - 8.01) |
| MDCD | 1,216 | 3,541 | 0.7 | 2.7 | <5 | 18 | 1.31 (0.26 - 5.38) | 1.21 (0.26 - 5.57) |
| USOC | 140,950 | 295,487 | 116.9 | 266.6 | 148 | 720 | 0.58 (0.45 - 0.75) | 0.59 (0.44 - 0.81) |
| OEHR | 7,150 | 15,593 | 2.3 | 8.6 | <5 | 21 | 0.26 (0.01 - 1.49) | 0.23 (0.02 - 2.41) |
| SIDIAP | 1,497 | 5,892 | 2.1 | 12.2 | 0 | <5 | | |
| VA | 3,908 | 9,710 | 2.9 | 9.1 | <10 | <10 | 0.88 (0.04 - 9.66) | 0.84 (0.05 - 13.2) |
| Meta-analysis | 171,303 | 360,724 | 136.3 | 315.7 | <193 | <895 | 0.59 (0.47 - 0.75) | 0.59 (0.47 - 0.75) |
| **PS-stratification / on-treatment TAR** | | | | | | | | |
| CCAE | 10,088 | 13,116 | 7.7 | 10.4 | 6 | 14 | 0.55 (0.18 - 1.50) | 0.56 (0.19 - 1.60) |
| GDA | 4,434 | 8,797 | 3.5 | 9.3 | 0 | 0 | | |
| OCEDM | 11,552 | 17,937 | 7.1 | 13.8 | 17 | 92 | 0.39 (0.21 - 0.67) | 0.39 (0.22 - 0.69) |
| MDCR | 2,005 | 5,793 | 1.4 | 5.0 | 9 | 23 | 1.81 (0.70 - 4.35) | 1.85 (0.74 - 4.61) |
| MDCD | 1,302 | 3,561 | 0.7 | 2.7 | <5 | 18 | 0.85 (0.19 - 2.77) | 0.76 (0.20 - 2.91) |
| USOC | 195,307 | 296,299 | 146.8 | 267.2 | 215 | 728 | 0.63 (0.53 - 0.75) | 0.66 (0.53 - 0.82) |
| OEHR | 8,965 | 15,785 | 2.8 | 8.7 | <5 | 23 | 0.72 (0.19 - 2.14) | 0.66 (0.20 - 2.20) |
| SIDIAP | 2,288 | 6,265 | 2.9 | 13.3 | 0 | <5 | | |
| VA | 7,274 | 10,232 | 4.5 | 9.9 | <10 | <10 | 0.92 (0.21 - 4.05) | 0.93 (0.21 - 4.12) |
| Meta-analysis | 225,912 | 343,137 | 164.3 | 300.1 | <243 | 857 | 0.61 (0.51 - 0.71) | 0.62 (0.51 - 0.75) |

**Supplementary Table 75:** Exposures, outcomes and risk estimates for Sudden cardiac death in SGLT2is (T) vs SUs (C) new-user cohorts under two different propensity score (PS) adjustment / time-at-risk (TAR) designs across data sources. We report total patient exposure time in 1,000 patient-year (KYr) units, uncalibrated and calibrated hazard ratios (HRs) and their 95% confidence intervals (CIs). Greyed out entries did not pass study diagnostics and are included only for completeness.

| | Patients | | Exposure (KYr) | | Outcomes | | Uncalibrated | Calibrated |
|---|---|---|---|---|---|---|---|---|
| | T | C | T | C | T | C | HR (95% CI) | HR (95% CI) |
| **PS-matching / on-treatment TAR** | | | | | | | | |
| CCAE | 8,285 | 17,761 | 6.5 | 13.7 | 6 | 40 | 0.29 (0.07 - 0.91) | 0.28 (0.08 - 1.04) |
| GDA | 4,184 | 723 | 3.4 | 0.7 | 0 | 0 | | |
| OCEDM | 10,076 | 37,084 | 6.4 | 33.0 | 15 | 204 | 0.55 (0.26 - 1.06) | 0.58 (0.28 - 1.17) |
| MDCR | 1,660 | 8,237 | 1.3 | 7.1 | 9 | 31 | 2.31 (0.67 - 7.39) | 2.57 (0.78 - 8.51) |
| MDCD | 1,274 | 6,483 | 0.7 | 4.5 | <5 | 25 | 0.80 (0.12 - 3.40) | 0.78 (0.14 - 4.27) |
| USOC | 172,056 | 549,964 | 133.6 | 566.3 | 186 | 1,751 | 0.56 (0.46 - 0.68) | 0.58 (0.44 - 0.77) |
| OEHR | 7,784 | 32,954 | 2.4 | 20.4 | <5 | 77 | 0.45 (0.10 - 1.42) | 0.42 (0.11 - 1.55) |
| SIDIAP | 1,648 | 320 | 2.4 | 0.6 | 0 | 0 | | |
| VA | 4,946 | 26,895 | 3.6 | 23.0 | <10 | 12 | 1.48 (0.28 - 6.58) | 1.51 (0.31 - 7.35) |
| Meta-analysis | 201,135 | 652,483 | 150.9 | 645.0 | <226 | 2,128 | 0.60 (0.42 - 0.87) | 0.61 (0.41 - 0.90) |
| **PS-stratification / on-treatment TAR** | | | | | | | | |
| CCAE | 10,088 | 18,021 | 7.7 | 13.9 | 6 | 40 | 0.29 (0.11 - 0.68) | 0.29 (0.11 - 0.72) |
| GDA | 4,337 | 1,235 | 3.5 | 1.3 | 0 | 0 | | |
| OCEDM | 11,552 | 37,919 | 7.1 | 33.7 | 17 | 217 | 0.61 (0.34 - 1.01) | 0.61 (0.35 - 1.05) |
| MDCR | 2,005 | 8,843 | 1.4 | 7.5 | 9 | 33 | 1.93 (0.75 - 4.62) | 1.87 (0.75 - 4.65) |
| MDCD | 1,304 | 6,690 | 0.7 | 4.6 | <5 | 29 | 0.63 (0.15 - 1.91) | 0.62 (0.17 - 2.24) |
| USOC | 195,307 | 559,493 | 146.8 | 575.4 | 215 | 1,822 | 0.60 (0.51 - 0.70) | 0.62 (0.49 - 0.79) |
| OEHR | 8,965 | 34,672 | 2.8 | 21.6 | <5 | 87 | 0.56 (0.16 - 1.51) | 0.53 (0.17 - 1.64) |
| SIDIAP | 2,273 | 619 | 2.9 | 1.4 | 0 | 0 | | |
| VA | 7,274 | 39,031 | 4.5 | 36.7 | <10 | 25 | 1.62 (0.37 - 6.22) | 1.65 (0.40 - 6.72) |
| Meta-analysis | 225,912 | 650,105 | 164.3 | 644.7 | <243 | 2,166 | 0.59 (0.51 - 0.68) | 0.60 (0.49 - 0.73) |

**Supplementary Table 76:** Exposures, outcomes and risk estimates for Sudden cardiac death in GLP1-RAs (T) vs DPP4is (C) new-user cohorts under two different propensity score (PS) adjustment / time-at-risk (TAR) designs across data sources. We report total patient exposure time in 1,000 patient-year (KYr) units, uncalibrated and calibrated hazard ratios (HRs) and their 95% confidence intervals (CIs). Greyed out entries did not pass study diagnostics and are included only for completeness.

| | Patients | | Exposure (KYr) | | Outcomes | | Uncalibrated | Calibrated |
|---|---|---|---|---|---|---|---|---|
| | T | C | T | C | T | C | HR (95% CI) | HR (95% CI) |
| **PS-matching / on-treatment TAR** | | | | | | | | |
| CCAE | 5,610 | 22,583 | 3.7 | 18.7 | 7 | 22 | 1.87 (0.51 - 6.43) | 1.90 (0.54 - 6.75) |
| OCEDM | 4,933 | 23,099 | 2.8 | 18.6 | 7 | 102 | 0.46 (0.17 - 1.06) | 0.45 (0.18 - 1.14) |
| MDCR | 959 | 9,661 | 0.6 | 9.3 | <5 | 41 | 0.55 (0.03 - 3.19) | 0.48 (0.05 - 5.12) |
| USOC | 74,338 | 295,582 | 52.1 | 267.2 | 65 | 706 | 0.81 (0.57 - 1.14) | 0.79 (0.56 - 1.12) |
| OEHR | 3,634 | 17,979 | 1.1 | 9.7 | <5 | 27 | 0.63 (0.09 - 2.78) | 0.53 (0.10 - 2.97) |
| Meta-analysis | 88,515 | 359,243 | 59.7 | 314.2 | <84 | 857 | 0.79 (0.55 - 1.13) | 0.77 (0.53 - 1.12) |
| **PS-stratification / on-treatment TAR** | | | | | | | | |
| CCAE | 7,047 | 22,729 | 4.3 | 18.8 | 7 | 23 | 1.34 (0.49 - 3.33) | 1.34 (0.51 - 3.52) |
| OCEDM | 6,525 | 23,819 | 3.5 | 19.2 | 8 | 110 | 0.39 (0.16 - 0.81) | 0.38 (0.17 - 0.85) |
| MDCR | 1,040 | 10,492 | 0.6 | 10.1 | <5 | 42 | 0.70 (0.04 - 3.41) | 0.63 (0.07 - 5.85) |
| USOC | 103,838 | 298,333 | 64.8 | 269.6 | 75 | 730 | 0.64 (0.48 - 0.83) | 0.63 (0.47 - 0.85) |
| OEHR | 4,516 | 18,109 | 1.3 | 9.8 | <5 | 29 | 0.88 (0.19 - 2.85) | 0.72 (0.19 - 2.78) |
| Meta-analysis | 121,926 | 362,990 | 73.9 | 317.4 | <95 | 892 | 0.66 (0.45 - 0.97) | 0.65 (0.43 - 0.96) |

**Supplementary Table 77:** Exposures, outcomes and risk estimates for Sudden cardiac death in GLP1-RAs (T) vs SUs (C) new-user cohorts under two different propensity score (PS) adjustment / time-at-risk (TAR) designs across data sources. We report total patient exposure time in 1,000 patient-year (KYr) units, uncalibrated and calibrated hazard ratios (HRs) and their 95% confidence intervals (CIs). Greyed out entries did not pass study diagnostics and are included only for completeness.

| | Patients | | Exposure (KYr) | | Outcomes | | Uncalibrated | Calibrated |
|---|---|---|---|---|---|---|---|---|
| | T | C | T | C | T | C | HR (95% CI) | HR (95% CI) |
| **PS-matching / on-treatment TAR** | | | | | | | | |
| CCAE | 5,953 | 34,454 | 3.8 | 27.1 | 6 | 71 | 1.49 (0.46 - 4.11) | 1.41 (0.47 - 4.23) |
| OCEDM | 5,502 | 44,858 | 3.0 | 40.7 | 8 | 187 | 0.92 (0.38 - 1.96) | 0.94 (0.41 - 2.12) |
| MDCR | 1,031 | 15,659 | 0.6 | 15.3 | <5 | 57 | 0.37 (0.02 - 2.25) | 0.30 (0.03 - 3.30) |
| USOC | 87,882 | 537,539 | 57.6 | 555.7 | 66 | 1,617 | 0.60 (0.44 - 0.80) | 0.59 (0.42 - 0.82) |
| OEHR | 3,870 | 37,279 | 1.1 | 22.7 | <5 | 76 | 1.39 (0.29 - 4.73) | 1.27 (0.31 - 5.15) |
| Meta-analysis | 103,207 | 654,130 | 65.5 | 646.1 | <85 | 1,951 | 0.79 (0.52 - 1.22) | 0.76 (0.48 - 1.21) |
| **PS-stratification / on-treatment TAR** | | | | | | | | |
| CCAE | 7,417 | 35,329 | 4.6 | 27.7 | 7 | 73 | 0.74 (0.28 - 1.67) | 0.72 (0.30 - 1.76) |
| OCEDM | 6,525 | 52,453 | 3.5 | 48.6 | 8 | 287 | 0.73 (0.31 - 1.49) | 0.73 (0.34 - 1.59) |
| MDCR | 1,113 | 20,776 | 0.7 | 19.9 | <5 | 91 | 0.41 (0.02 - 2.01) | 0.34 (0.04 - 3.17) |
| USOC | 103,838 | 565,620 | 64.8 | 584.5 | 75 | 1,845 | 0.58 (0.44 - 0.73) | 0.56 (0.41 - 0.77) |
| OEHR | 4,517 | 42,529 | 1.3 | 26.3 | <5 | 106 | 0.99 (0.23 - 2.91) | 0.82 (0.23 - 2.93) |
| Meta-analysis | 111,255 | 600,949 | 69.4 | 612.2 | 82 | 1,918 | 0.59 (0.46 - 0.75) | 0.57 (0.42 - 0.78) |

**Supplementary Table 78:** Exposures, outcomes and risk estimates for Sudden cardiac death in DPP4is (T) vs SUs (C) new-user cohorts under two different propensity score (PS) adjustment / time-at-risk (TAR) designs across data sources. We report total patient exposure time in 1,000 patient-year (KYr) units, uncalibrated and calibrated hazard ratios (HRs) and their 95% confidence intervals (CIs). Greyed out entries did not pass study diagnostics and are included only for completeness.

| | Patients | | Exposure (KYr) | | Outcomes | | Uncalibrated | Calibrated |
|---|---|---|---|---|---|---|---|---|
| | T | C | T | C | T | C | HR (95% CI) | HR (95% CI) |
| **PS-matching / on-treatment TAR** | | | | | | | | |
| CCAE | 19,792 | 33,348 | 16.4 | 25.9 | 21 | 71 | 0.66 (0.33 - 1.27) | 0.68 (0.35 - 1.32) |
| GDA | 10,408 | 1,974 | 11.3 | 2.0 | 0 | 0 | | |
| OCEDM | 22,570 | 52,421 | 18.1 | 48.6 | 104 | 287 | 1.06 (0.78 - 1.43) | 1.12 (0.82 - 1.52) |
| MDCR | 9,814 | 18,945 | 9.4 | 17.9 | 41 | 83 | 0.85 (0.49 - 1.42) | 0.81 (0.47 - 1.37) |
| MDCD | 4,249 | 9,440 | 3.2 | 6.7 | 22 | 41 | 0.83 (0.33 - 1.92) | 0.87 (0.36 - 2.10) |
| IMRD | 3,388 | 407 | 5.7 | 0.6 | 0 | 0 | | |
| USOC | 290,676 | 565,621 | 260.8 | 584.6 | 713 | 1,845 | 0.89 (0.79 - 1.00) | 0.89 (0.76 - 1.04) |
| OEHR | 17,092 | 42,491 | 9.1 | 26.3 | 28 | 106 | 0.99 (0.56 - 1.70) | 0.97 (0.55 - 1.69) |
| SIDIAP | 7,933 | 1,797 | 21.0 | 4.6 | <5 | 0 | | |
| VA | 10,219 | 64,285 | 10.4 | 67.3 | <10 | 50 | 1.48 (0.38 - 4.62) | 1.51 (0.44 - 5.23) |
| Meta-analysis | 374,412 | 786,551 | 327.4 | 777.3 | <939 | 2,483 | 0.91 (0.81 - 1.01) | 0.90 (0.78 - 1.04) |
| **PS-stratification / on-treatment TAR** | | | | | | | | |
| CCAE | 22,729 | 33,507 | 18.8 | 26.0 | 23 | 71 | 0.55 (0.33 - 0.88) | 0.55 (0.33 - 0.90) |
| GDA | 10,428 | 2,365 | 11.3 | 2.5 | 0 | 0 | | |
| OCEDM | 23,823 | 52,457 | 19.2 | 48.6 | 110 | 287 | 1.12 (0.88 - 1.40) | 1.14 (0.90 - 1.44) |
| MDCR | 10,492 | 19,080 | 10.1 | 18.1 | 42 | 85 | 1.01 (0.68 - 1.48) | 1.01 (0.69 - 1.49) |
| MDCD | 4,421 | 9,467 | 3.4 | 6.7 | 22 | 41 | 1.12 (0.64 - 1.91) | 1.14 (0.66 - 1.98) |
| IMRD | 4,024 | 846 | 6.5 | 1.5 | 0 | 0 | | |
| USOC | 298,346 | 565,656 | 269.6 | 584.6 | 730 | 1,846 | 0.94 (0.86 - 1.03) | 0.94 (0.80 - 1.09) |
| OEHR | 18,109 | 42,528 | 9.8 | 26.3 | 29 | 106 | 0.93 (0.60 - 1.41) | 0.97 (0.63 - 1.49) |
| SIDIAP | 8,121 | 2,335 | 21.2 | 6.3 | <5 | 0 | | |
| VA | 10,950 | 97,207 | 10.9 | 108.0 | <10 | 82 | 0.92 (0.32 - 2.33) | 0.96 (0.35 - 2.60) |
| Meta-analysis | 377,920 | 722,695 | 330.9 | 710.3 | 956 | 2,436 | 0.96 (0.84 - 1.10) | 0.96 (0.81 - 1.13) |

**Supplementary Table 79:** Exposures, outcomes and risk estimates for Hospitalization with heart failure in SGLT2is (T) vs GLP1-RAs (C) new-user cohorts under two different propensity score (PS) adjustment / time-at-risk (TAR) designs across data sources. We report total patient exposure time in 1,000 patient-year (KYr) units, uncalibrated and calibrated hazard ratios (HRs) and their 95% confidence intervals (CIs). Greyed out entries did not pass study diagnostics and are included only for completeness.

| | Patients | | Exposure (KYr) | | Outcomes | | Uncalibrated | Calibrated |
|---|---|---|---|---|---|---|---|---|
| | T | C | T | C | T | C | HR (95% CI) | HR (95% CI) |
| **PS-matching / on-treatment TAR** | | | | | | | | |
| CCAE | 9,482 | 4,327 | 7.3 | 2.9 | 80 | 27 | 1.34 (0.74 - 2.51) | 1.30 (0.70 - 2.39) |
| OCEDM | 10,071 | 4,554 | 6.3 | 2.4 | 175 | 65 | 1.08 (0.74 - 1.60) | 1.11 (0.76 - 1.65) |
| MDCR | 1,760 | 654 | 1.3 | 0.4 | 35 | 11 | 2.50 (0.79 - 11.1) | 3.18 (0.85 - 11.9) |
| USOC | 173,187 | 82,053 | 132.5 | 52.9 | 1,647 | 581 | 1.05 (0.92 - 1.19) | 1.10 (0.96 - 1.27) |
| OEHR | 8,356 | 3,557 | 2.6 | 1.0 | 54 | 16 | 1.45 (0.75 - 3.01) | 1.63 (0.81 - 3.28) |
| Meta-analysis | 201,096 | 94,491 | 148.6 | 59.1 | 1,956 | 689 | 1.07 (0.95 - 1.20) | 1.12 (0.98 - 1.27) |
| **PS-stratification / on-treatment TAR** | | | | | | | | |
| CCAE | 9,485 | 5,119 | 7.3 | 3.3 | 80 | 29 | 1.29 (0.83 - 2.06) | 1.30 (0.82 - 2.05) |
| OCEDM | 10,084 | 5,301 | 6.3 | 2.8 | 175 | 71 | 0.93 (0.69 - 1.26) | 0.94 (0.70 - 1.27) |
| MDCR | 1,767 | 690 | 1.3 | 0.4 | 35 | 11 | 1.11 (0.55 - 2.40) | 1.22 (0.58 - 2.55) |
| USOC | 173,214 | 95,533 | 132.5 | 59.7 | 1,649 | 657 | 0.96 (0.88 - 1.06) | 1.01 (0.92 - 1.12) |
| OEHR | 8,371 | 4,092 | 2.6 | 1.1 | 55 | 20 | 1.25 (0.73 - 2.21) | 1.34 (0.76 - 2.37) |
| Meta-analysis | 201,154 | 110,045 | 148.6 | 66.9 | 1,959 | 777 | 0.98 (0.90 - 1.07) | 1.02 (0.93 - 1.12) |

**Supplementary Table 80:** Exposures, outcomes and risk estimates for Hospitalization with heart failure in SGLT2is (T) vs DPP4is (C) new-user cohorts under two different propensity score (PS) adjustment / time-at-risk (TAR) designs across data sources. We report total patient exposure time in 1,000 patient-year (KYr) units, uncalibrated and calibrated hazard ratios (HRs) and their 95% confidence intervals (CIs). Greyed out entries did not pass study diagnostics and are included only for completeness.

| | Patients | | Exposure (KYr) | | Outcomes | | Uncalibrated | Calibrated |
|---|---|---|---|---|---|---|---|---|
| | T | C | T | C | T | C | HR (95% CI) | HR (95% CI) |
| **PS-matching / on-treatment TAR** | | | | | | | | |
| CCAE | 7,609 | 12,295 | 6.2 | 9.8 | 63 | 113 | 0.79 (0.50 - 1.22) | 0.74 (0.47 - 1.17) |
| GDA | 3,365 | 8,557 | 2.8 | 9.0 | 0 | 0 | | |
| OCEDM | 7,558 | 15,376 | 5.1 | 11.6 | 126 | 489 | 0.74 (0.56 - 0.98) | 0.73 (0.55 - 0.97) |
| MDCR | 1,442 | 4,929 | 1.1 | 4.3 | 29 | 183 | 0.83 (0.45 - 1.45) | 0.88 (0.49 - 1.59) |
| MDCD | 1,013 | 2,892 | 0.6 | 2.1 | 19 | 92 | 1.02 (0.50 - 1.97) | 0.94 (0.48 - 1.87) |
| USOC | 129,185 | 266,519 | 108.1 | 239.2 | 1,242 | 5,411 | 0.67 (0.61 - 0.73) | 0.69 (0.57 - 0.83) |
| OEHR | 6,729 | 14,588 | 2.1 | 7.8 | 42 | 225 | 0.99 (0.63 - 1.53) | 0.88 (0.57 - 1.38) |
| SIDIAP | 1,484 | 5,821 | 2.1 | 12.1 | <5 | 36 | 0.52 (0.12 - 1.69) | 0.51 (0.12 - 2.10) |
| VA | 3,557 | 8,994 | 2.6 | 8.4 | 32 | 129 | 0.65 (0.38 - 1.08) | 0.62 (0.37 - 1.06) |
| Meta-analysis | 158,577 | 331,414 | 128.0 | 295.3 | <1,558 | 6,678 | 0.69 (0.64 - 0.75) | 0.69 (0.63 - 0.76) |
| **PS-stratification / on-treatment TAR** | | | | | | | | |
| CCAE | 9,485 | 12,359 | 7.3 | 9.8 | 80 | 114 | 0.93 (0.68 - 1.26) | 0.94 (0.69 - 1.28) |
| GDA | 4,434 | 8,797 | 3.5 | 9.3 | 0 | 0 | | |
| OCEDM | 10,084 | 15,580 | 6.3 | 11.8 | 175 | 503 | 0.78 (0.64 - 0.95) | 0.78 (0.64 - 0.96) |
| MDCR | 1,767 | 4,968 | 1.3 | 4.3 | 35 | 185 | 0.76 (0.50 - 1.14) | 0.78 (0.52 - 1.18) |
| MDCD | 1,068 | 2,906 | 0.6 | 2.1 | 20 | 93 | 1.03 (0.60 - 1.70) | 0.92 (0.54 - 1.55) |
| USOC | 173,214 | 266,932 | 132.5 | 239.6 | 1,649 | 5,455 | 0.72 (0.68 - 0.77) | 0.76 (0.65 - 0.88) |
| OEHR | 8,371 | 14,761 | 2.6 | 7.9 | 55 | 238 | 0.97 (0.69 - 1.33) | 0.88 (0.64 - 1.23) |
| SIDIAP | 2,252 | 6,192 | 2.9 | 13.1 | <5 | 41 | 0.32 (0.06 - 1.10) | 0.32 (0.07 - 1.33) |
| VA | 6,102 | 9,499 | 3.9 | 9.1 | 63 | 133 | 0.62 (0.42 - 0.91) | 0.62 (0.42 - 0.92) |
| Meta-analysis | 201,154 | 309,632 | 148.6 | 269.1 | 1,959 | 6,310 | 0.79 (0.69 - 0.90) | 0.80 (0.68 - 0.95) |

**Supplementary Table 81:** Exposures, outcomes and risk estimates for Hospitalization with heart failure in SGLT2is (T) vs SUs (C) new-user cohorts under two different propensity score (PS) adjustment / time-at-risk (TAR) designs across data sources. We report total patient exposure time in 1,000 patient-year (KYr) units, uncalibrated and calibrated hazard ratios (HRs) and their 95% confidence intervals (CIs). Greyed out entries did not pass study diagnostics and are included only for completeness.

| | Patients | | Exposure (KYr) | | Outcomes | | Uncalibrated | Calibrated |
|---|---|---|---|---|---|---|---|---|
| | T | C | T | C | T | C | HR (95% CI) | HR (95% CI) |
| **PS-matching / on-treatment TAR** | | | | | | | | |
| CCAE | 7,827 | 16,603 | 6.1 | 12.8 | 66 | 188 | 0.63 (0.41 - 0.95) | 0.61 (0.40 - 0.93) |
| GDA | 4,184 | 723 | 3.4 | 0.7 | 0 | 0 | | |
| OCEDM | 8,986 | 32,296 | 5.8 | 28.2 | 145 | 1,384 | 0.66 (0.53 - 0.83) | 0.69 (0.55 - 0.87) |
| MDCR | 1,476 | 6,933 | 1.1 | 5.9 | 31 | 269 | 0.57 (0.31 - 0.99) | 0.64 (0.36 - 1.14) |
| MDCD | 1,054 | 5,163 | 0.6 | 3.5 | 20 | 164 | 0.90 (0.50 - 1.54) | 0.89 (0.51 - 1.56) |
| USOC | 155,896 | 492,655 | 122.4 | 503.9 | 1,469 | 13,293 | 0.63 (0.59 - 0.68) | 0.65 (0.53 - 0.80) |
| OEHR | 7,307 | 30,706 | 2.3 | 18.5 | 49 | 654 | 0.82 (0.56 - 1.16) | 0.75 (0.52 - 1.08) |
| SIDIAP | 1,634 | 317 | 2.4 | 0.6 | <5 | <5 | 0.33 (0.01 - 3.72) | 0.36 (0.02 - 5.66) |
| VA | 4,420 | 24,348 | 3.2 | 20.6 | 41 | 408 | 0.72 (0.47 - 1.07) | 0.74 (0.49 - 1.11) |
| Meta-analysis | 182,546 | 584,356 | 138.3 | 572.8 | 1,780 | 15,952 | 0.64 (0.60 - 0.68) | 0.65 (0.55 - 0.76) |
| **PS-stratification / on-treatment TAR** | | | | | | | | |
| CCAE | 9,485 | 16,807 | 7.3 | 12.9 | 80 | 195 | 0.77 (0.57 - 1.02) | 0.75 (0.56 - 1.01) |
| GDA | 4,337 | 1,235 | 3.5 | 1.3 | 0 | 0 | | |
| OCEDM | 10,084 | 32,769 | 6.3 | 28.6 | 175 | 1,433 | 0.69 (0.58 - 0.82) | 0.69 (0.58 - 0.83) |
| MDCR | 1,767 | 7,419 | 1.3 | 6.3 | 35 | 302 | 0.75 (0.49 - 1.12) | 0.73 (0.49 - 1.10) |
| MDCD | 1,069 | 5,365 | 0.6 | 3.6 | 20 | 170 | 0.86 (0.51 - 1.36) | 0.84 (0.51 - 1.37) |
| USOC | 173,214 | 499,973 | 132.5 | 511.0 | 1,649 | 13,876 | 0.65 (0.62 - 0.69) | 0.68 (0.57 - 0.82) |
| OEHR | 8,371 | 32,179 | 2.6 | 19.5 | 55 | 732 | 0.82 (0.60 - 1.10) | 0.78 (0.56 - 1.08) |
| SIDIAP | 2,237 | 616 | 2.9 | 1.4 | <5 | <5 | 1.70 (0.11 - 24.3) | 1.79 (0.12 - 26.0) |
| VA | 6,102 | 35,626 | 3.9 | 33.1 | 63 | 705 | 0.76 (0.54 - 1.04) | 0.77 (0.55 - 1.07) |
| Meta-analysis | 201,154 | 581,728 | 148.6 | 572.0 | 1,959 | 16,236 | 0.67 (0.63 - 0.73) | 0.68 (0.58 - 0.80) |

**Supplementary Table 82:** Exposures, outcomes and risk estimates for Hospitalization with heart failure in GLP1-RAs (T) vs DPP4is (C) new-user cohorts under two different propensity score (PS) adjustment / time-at-risk (TAR) designs across data sources. We report total patient exposure time in 1,000 patient-year (KYr) units, uncalibrated and calibrated hazard ratios (HRs) and their 95% confidence intervals (CIs). Greyed out entries did not pass study diagnostics and are included only for completeness.

| | Patients | | Exposure (KYr) | | Outcomes | | Uncalibrated | Calibrated |
|---|---|---|---|---|---|---|---|---|
| | T | C | T | C | T | C | HR (95% CI) | HR (95% CI) |
| **PS-matching / on-treatment TAR** | | | | | | | | |
| CCAE | 5,272 | 21,388 | 3.5 | 17.7 | 28 | 187 | 0.59 (0.35 - 0.97) | 0.60 (0.36 - 1.01) |
| OCEDM | 4,470 | 20,568 | 2.5 | 16.4 | 68 | 594 | 0.94 (0.66 - 1.32) | 0.94 (0.66 - 1.32) |
| MDCR | 871 | 8,539 | 0.5 | 8.2 | 16 | 310 | 0.87 (0.44 - 1.60) | 0.77 (0.40 - 1.47) |
| USOC | 68,153 | 267,295 | 47.9 | 240.4 | 552 | 5,383 | 0.78 (0.69 - 0.88) | 0.76 (0.66 - 0.87) |
| OEHR | 3,418 | 16,895 | 1.0 | 8.9 | 17 | 267 | 0.96 (0.51 - 1.69) | 0.81 (0.45 - 1.47) |
| Meta-analysis | 81,313 | 326,146 | 54.8 | 283.4 | 665 | 6,431 | 0.79 (0.71 - 0.88) | 0.77 (0.68 - 0.88) |
| **PS-stratification / on-treatment TAR** | | | | | | | | |
| CCAE | 6,644 | 21,496 | 4.1 | 17.8 | 34 | 193 | 0.67 (0.44 - 1.00) | 0.68 (0.45 - 1.02) |
| OCEDM | 5,916 | 21,034 | 3.1 | 16.8 | 76 | 630 | 0.87 (0.65 - 1.14) | 0.85 (0.64 - 1.12) |
| MDCR | 938 | 9,068 | 0.6 | 8.7 | 17 | 365 | 0.88 (0.50 - 1.45) | 0.78 (0.46 - 1.33) |
| USOC | 95,674 | 268,831 | 59.8 | 241.8 | 658 | 5,510 | 0.77 (0.70 - 0.84) | 0.76 (0.65 - 0.89) |
| OEHR | 4,259 | 16,984 | 1.2 | 8.9 | 21 | 273 | 0.87 (0.52 - 1.38) | 0.72 (0.44 - 1.17) |
| Meta-analysis | 112,493 | 328,345 | 68.2 | 285.3 | 789 | 6,606 | 0.78 (0.71 - 0.85) | 0.76 (0.67 - 0.86) |

**Supplementary Table 83:** Exposures, outcomes and risk estimates for Hospitalization with heart failure in GLP1-RAs (T) vs SUs (C) new-user cohorts under two different propensity score (PS) adjustment / time-at-risk (TAR) designs across data sources. We report total patient exposure time in 1,000 patient-year (KYr) units, uncalibrated and calibrated hazard ratios (HRs) and their 95% confidence intervals (CIs). Greyed out entries did not pass study diagnostics and are included only for completeness.

| | Patients | | Exposure (KYr) | | Outcomes | | Uncalibrated | Calibrated |
|---|---|---|---|---|---|---|---|---|
| | T | C | T | C | T | C | HR (95% CI) | HR (95% CI) |
| **PS-matching / on-treatment TAR** | | | | | | | | |
| CCAE | 5,606 | 32,248 | 3.6 | 25.3 | 29 | 373 | 0.57 (0.34 - 0.91) | 0.54 (0.33 - 0.88) |
| OCEDM | 4,978 | 40,094 | 2.7 | 35.7 | 72 | 1,588 | 0.80 (0.59 - 1.06) | 0.81 (0.61 - 1.09) |
| MDCR | 934 | 13,239 | 0.6 | 12.6 | 18 | 550 | 0.78 (0.40 - 1.38) | 0.64 (0.34 - 1.19) |
| USOC | 80,521 | 483,889 | 52.9 | 496.1 | 604 | 12,582 | 0.65 (0.59 - 0.72) | 0.63 (0.52 - 0.76) |
| OEHR | 3,645 | 35,006 | 1.1 | 20.7 | 18 | 705 | 0.73 (0.42 - 1.19) | 0.67 (0.40 - 1.13) |
| Meta-analysis | 95,684 | 604,476 | 60.8 | 590.4 | 741 | 15,798 | 0.66 (0.61 - 0.73) | 0.64 (0.53 - 0.77) |
| **PS-stratification / on-treatment TAR** | | | | | | | | |
| CCAE | 6,996 | 33,018 | 4.3 | 25.8 | 35 | 388 | 0.56 (0.37 - 0.81) | 0.55 (0.37 - 0.81) |
| OCEDM | 5,916 | 45,669 | 3.1 | 41.5 | 76 | 2,045 | 0.69 (0.53 - 0.89) | 0.69 (0.53 - 0.89) |
| MDCR | 1,006 | 17,464 | 0.6 | 16.5 | 19 | 893 | 0.79 (0.47 - 1.26) | 0.65 (0.40 - 1.07) |
| USOC | 95,674 | 505,613 | 59.8 | 519.2 | 658 | 14,102 | 0.65 (0.60 - 0.71) | 0.64 (0.52 - 0.79) |
| OEHR | 4,260 | 39,634 | 1.2 | 23.7 | 21 | 929 | 0.70 (0.43 - 1.08) | 0.58 (0.36 - 0.92) |
| Meta-analysis | 102,670 | 538,631 | 64.1 | 545.0 | 693 | 14,490 | 0.65 (0.60 - 0.70) | 0.64 (0.52 - 0.78) |

**Supplementary Table 84:** Exposures, outcomes and risk estimates for Hospitalization with heart failure in DPP4is (T) vs SUs (C) new-user cohorts under two different propensity score (PS) adjustment / time-at-risk (TAR) designs across data sources. We report total patient exposure time in 1,000 patient-year (KYr) units, uncalibrated and calibrated hazard ratios (HRs) and their 95% confidence intervals (CIs). Greyed out entries did not pass study diagnostics and are included only for completeness.

| | Patients | | Exposure (KYr) | | Outcomes | | Uncalibrated | Calibrated |
|---|---|---|---|---|---|---|---|---|
| | T | C | T | C | T | C | HR (95% CI) | HR (95% CI) |
| **PS-matching / on-treatment TAR** | | | | | | | | |
| CCAE | 18,650 | 31,183 | 15.4 | 24.2 | 181 | 361 | 1.00 (0.77 - 1.28) | 1.02 (0.79 - 1.31) |
| GDA | 10,408 | 1,974 | 11.3 | 2.0 | 0 | 0 | | |
| OCEDM | 19,817 | 45,657 | 15.7 | 41.5 | 600 | 2,043 | 0.84 (0.74 - 0.95) | 0.88 (0.78 - 1.01) |
| MDCR | 8,439 | 15,905 | 8.1 | 14.8 | 340 | 806 | 0.83 (0.70 - 1.00) | 0.79 (0.66 - 0.95) |
| MDCD | 3,460 | 7,581 | 2.6 | 5.2 | 121 | 268 | 0.81 (0.60 - 1.10) | 0.85 (0.63 - 1.16) |
| IMRD | 3,388 | 407 | 5.7 | 0.6 | 0 | 0 | | |
| USOC | 261,555 | 505,623 | 233.6 | 519.3 | 5,340 | 14,104 | 0.89 (0.85 - 0.93) | 0.89 (0.79 - 0.99) |
| OEHR | 16,001 | 39,601 | 8.3 | 23.7 | 259 | 929 | 0.93 (0.78 - 1.11) | 0.91 (0.76 - 1.08) |
| SIDIAP | 7,853 | 1,788 | 20.8 | 4.6 | 56 | 8 | 0.86 (0.31 - 2.79) | 0.84 (0.28 - 2.51) |
| VA | 9,482 | 59,199 | 9.6 | 61.4 | 136 | 1,182 | 0.81 (0.63 - 1.03) | 0.83 (0.64 - 1.06) |
| Meta-analysis | 345,257 | 706,537 | 314.1 | 694.6 | 7,033 | 19,701 | 0.88 (0.85 - 0.92) | 0.88 (0.79 - 0.97) |
| **PS-stratification / on-treatment TAR** | | | | | | | | |
| CCAE | 21,496 | 31,324 | 17.8 | 24.3 | 193 | 363 | 0.87 (0.72 - 1.04) | 0.87 (0.72 - 1.05) |
| GDA | 10,428 | 2,365 | 11.3 | 2.5 | 0 | 0 | | |
| OCEDM | 21,038 | 45,673 | 16.8 | 41.5 | 630 | 2,045 | 0.85 (0.78 - 0.94) | 0.87 (0.79 - 0.96) |
| MDCR | 9,068 | 16,035 | 8.7 | 15.0 | 365 | 819 | 0.90 (0.79 - 1.03) | 0.90 (0.79 - 1.03) |
| MDCD | 3,626 | 7,602 | 2.7 | 5.2 | 127 | 269 | 0.93 (0.74 - 1.17) | 0.95 (0.76 - 1.20) |
| IMRD | 4,024 | 846 | 6.5 | 1.5 | 0 | 0 | | |
| USOC | 268,843 | 505,648 | 241.8 | 519.3 | 5,510 | 14,106 | 0.90 (0.87 - 0.93) | 0.90 (0.79 - 1.02) |
| OEHR | 16,984 | 39,633 | 8.9 | 23.7 | 273 | 929 | 0.91 (0.79 - 1.05) | 0.95 (0.82 - 1.10) |
| SIDIAP | 8,035 | 2,321 | 21.0 | 6.2 | 56 | 9 | 1.04 (0.49 - 2.44) | 0.98 (0.44 - 2.19) |
| VA | 10,169 | 90,050 | 10.1 | 98.8 | 143 | 2,004 | 0.78 (0.64 - 0.95) | 0.81 (0.67 - 0.99) |
| Meta-analysis | 341,055 | 645,915 | 296.8 | 629.0 | 7,098 | 18,531 | 0.90 (0.87 - 0.92) | 0.90 (0.82 - 0.98) |

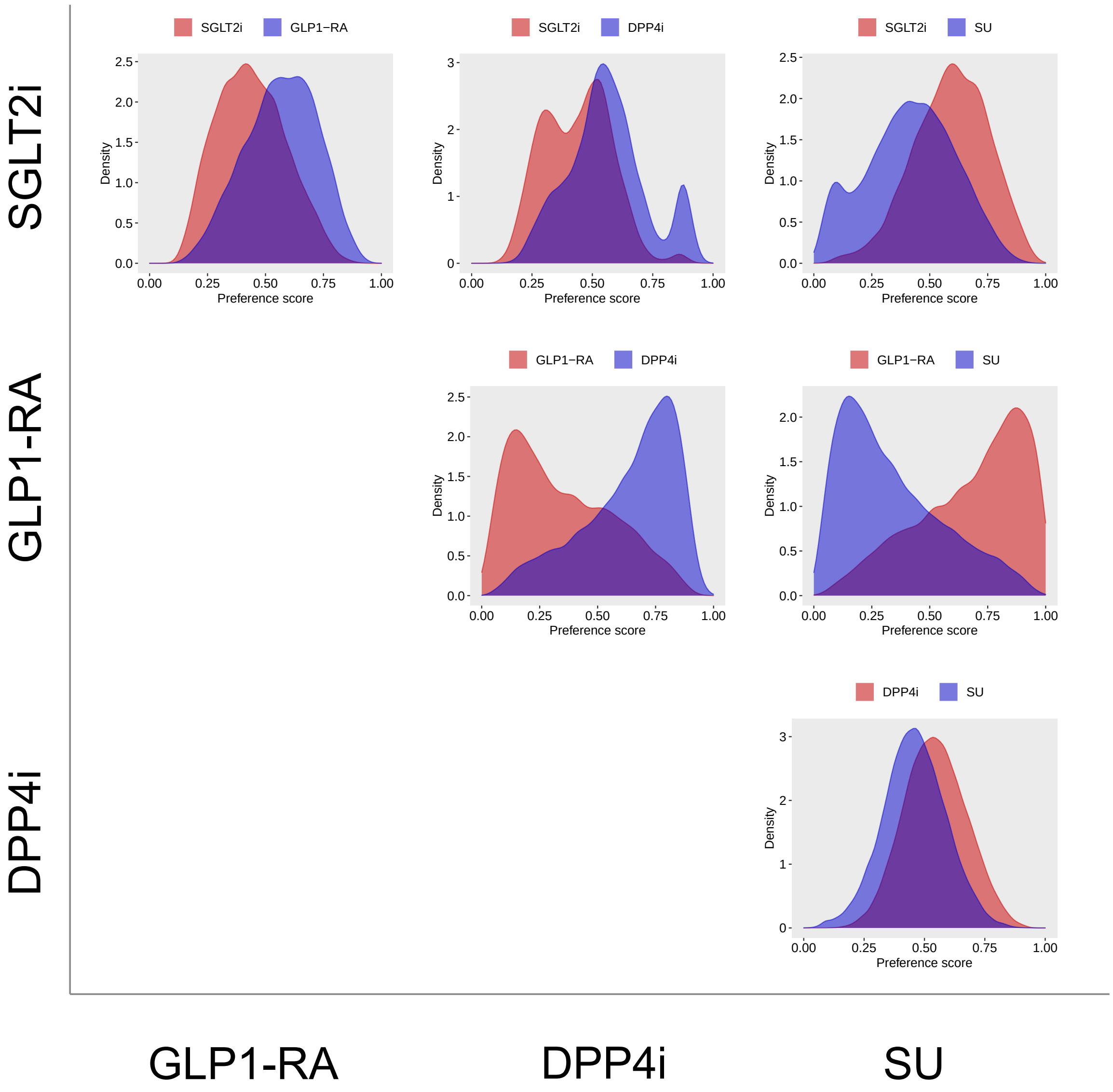


**Supplementary Figure 1: Preference score distributions comparing SGLT2is, GLP1-RAs, DPP4is, and SUs new-users with established CVD in CCAE.** The preference score is a transformation of the propensity score that adjusts for prevalence differences between populations. For each class pair, we fit a large-scale propensity score model using >40,000 baseline patient characteristics. A higher overlap between score distributions indicates that patients in the target (left, red) and compartor (bottom, blue) populations are more similar in terms of their predicted probability of receiving one treatment over the other. We define clinical empirical equipoise when a majority of patients in both populations carry preference scores between 0.3 and 0.7.

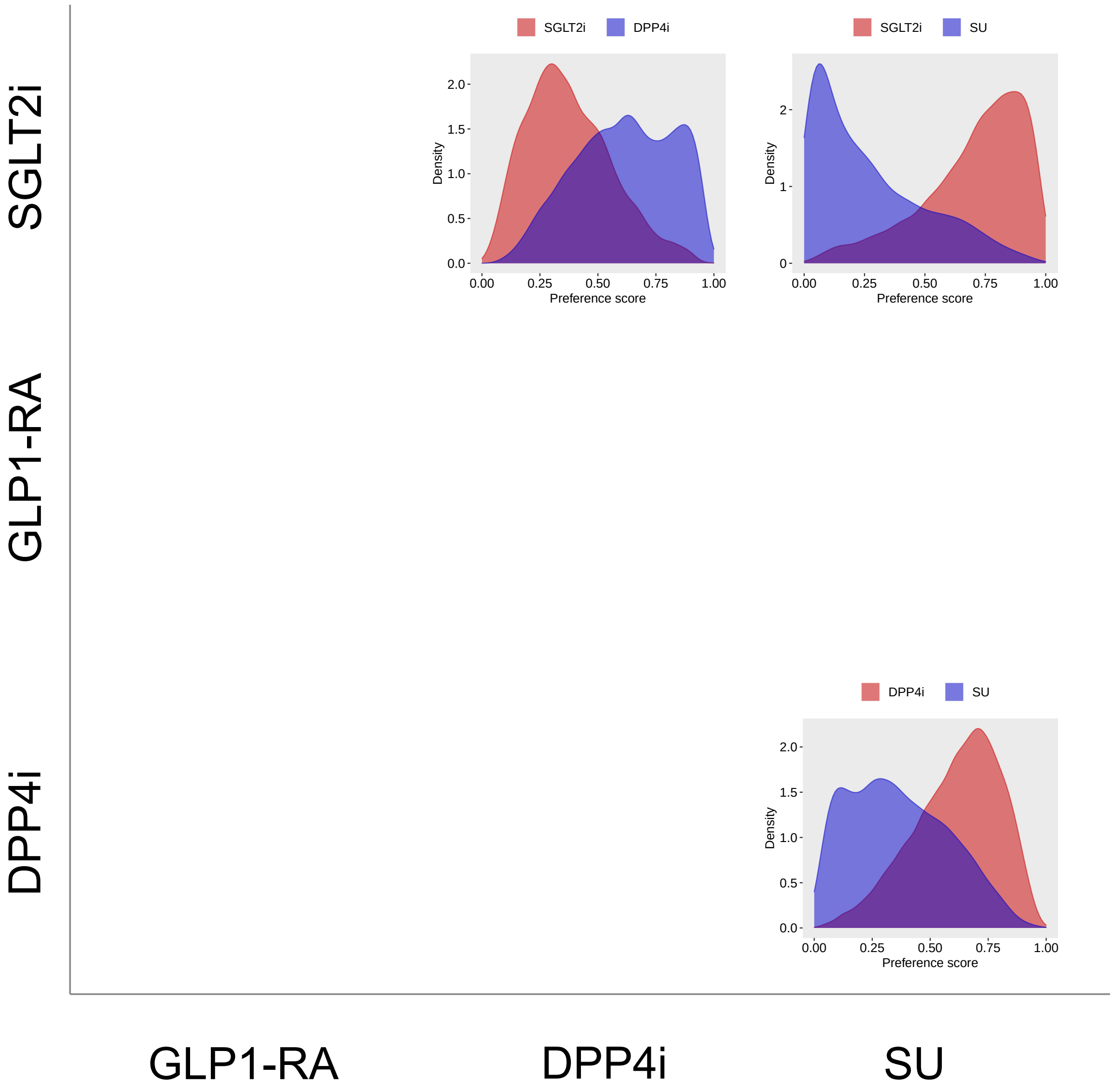


**Supplementary Figure 2: Preference score distributions comparing SGLT2is, GLP1-RAs, DPP4is, and SUs new-users with established CVD in GDA.** The preference score is a transformation of the propensity score that adjusts for prevalence differences between populations. For each class pair, we fit a large-scale propensity score model using >40,000 baseline patient characteristics. A higher overlap between score distributions indicates that patients in the target (left, red) and compartor (bottom, blue) populations are more similar in terms of their predicted probability of receiving one treatment over the other. We define clinical empirical equipoise when a majority of patients in both populations carry preference scores between 0.3 and 0.7.

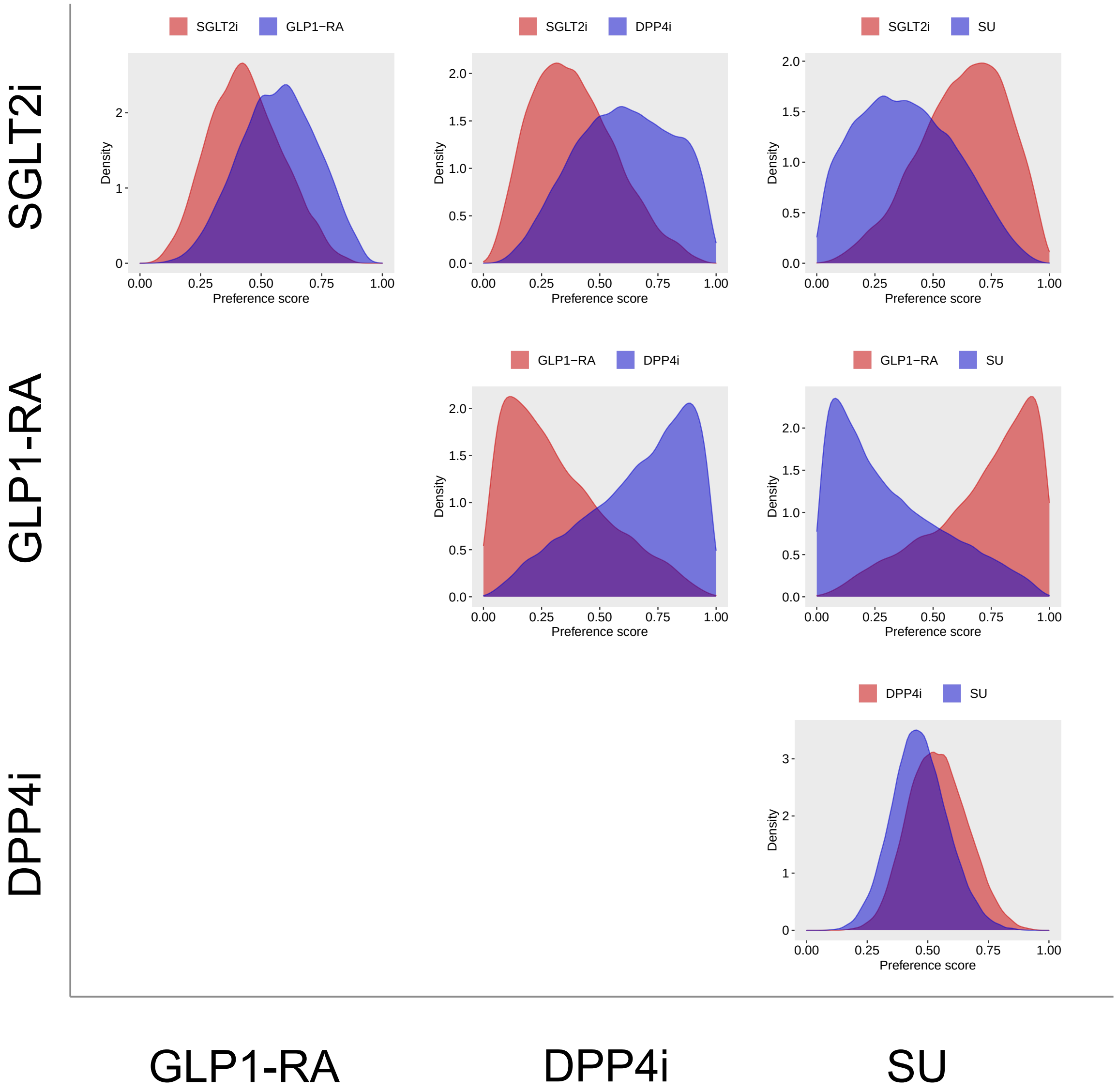


**Supplementary Figure 3: Preference score distributions comparing SGLT2is, GLP1-RAs, DPP4is, and SUs new-users with established CVD in OptumDOD.** The preference score is a transformation of the propensity score that adjusts for prevalence differences between populations. For each class pair, we fit a large-scale propensity score model using >40,000 baseline patient characteristics. A higher overlap between score distributions indicates that patients in the target (left, red) and compartor (bottom, blue) populations are more similar in terms of their predicted probability of receiving one treatment over the other. We define clinical empirical equipoise when a majority of patients in both populations carry preference scores between 0.3 and 0.7.

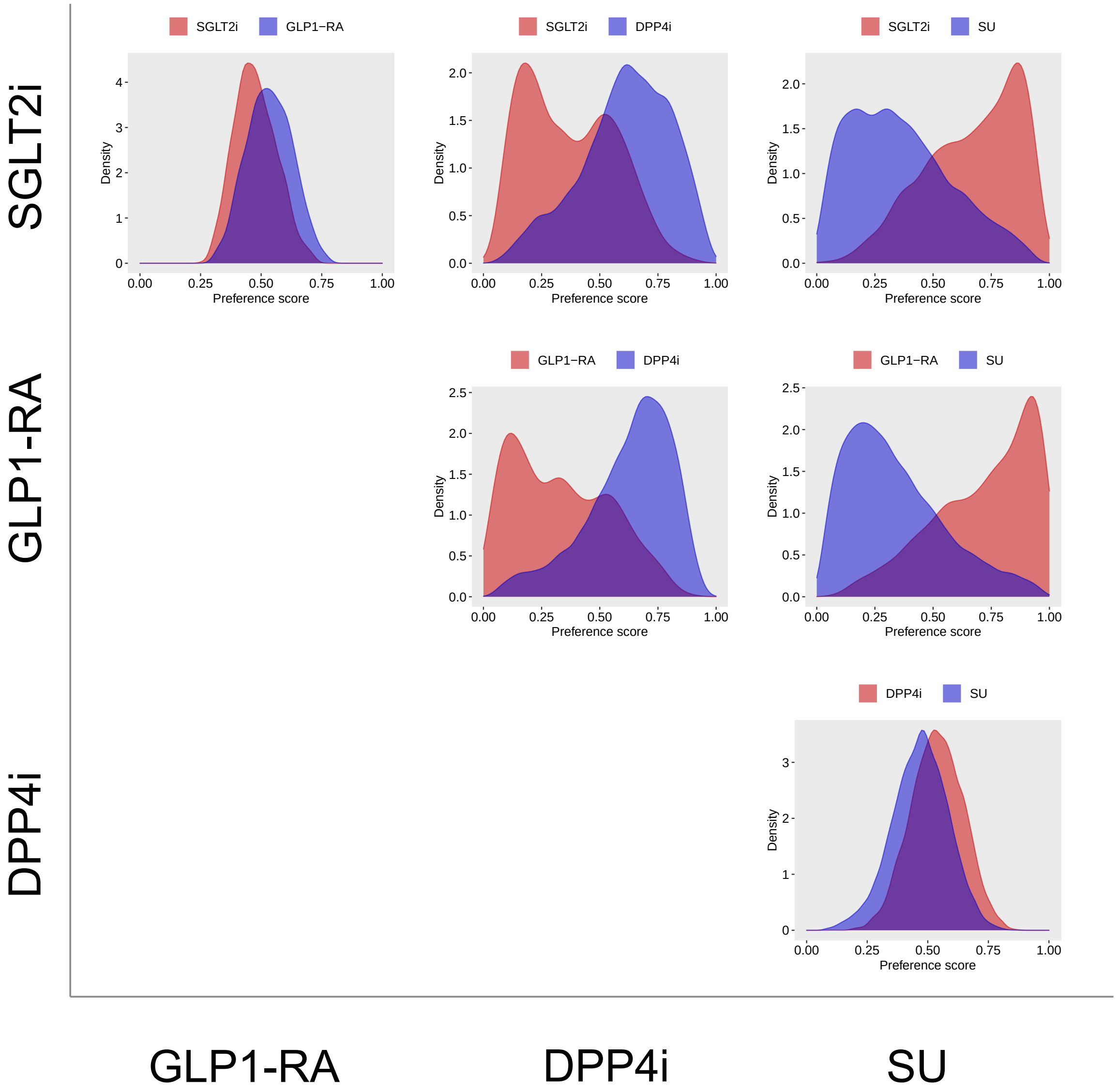


**Supplementary Figure 4: Preference score distributions comparing SGLT2is, GLP1-RAs, DPP4is, and SUs new-users with established CVD in MDCR.** The preference score is a transformation of the propensity score that adjusts for prevalence differences between populations. For each class pair, we fit a large-scale propensity score model using >40,000 baseline patient characteristics. A higher overlap between score distributions indicates that patients in the target (left, red) and compartor (bottom, blue) populations are more similar in terms of their predicted probability of receiving one treatment over the other. We define clinical empirical equipoise when a majority of patients in both populations carry preference scores between 0.3 and 0.7.

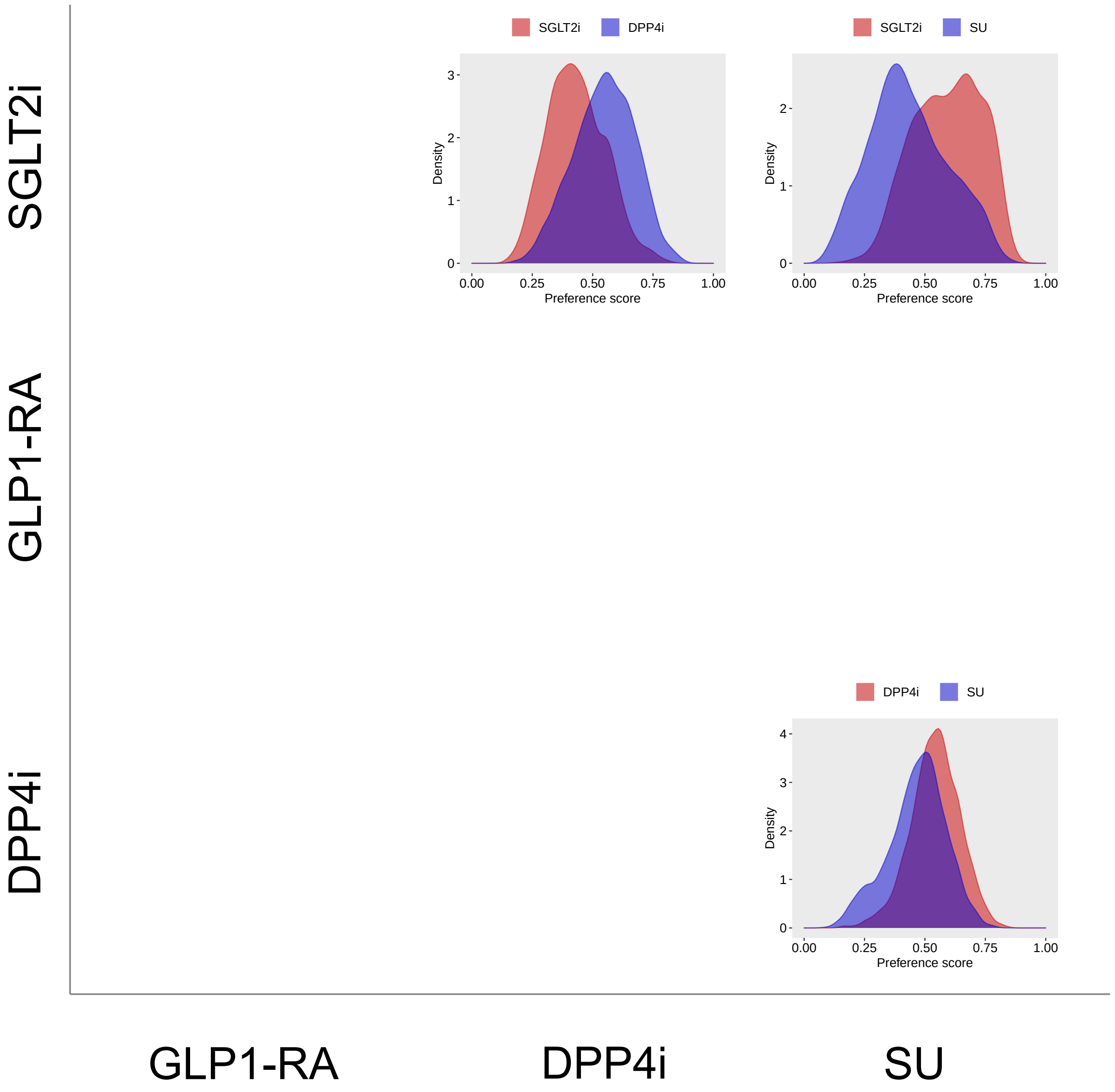


**Supplementary Figure 5: Preference score distributions comparing SGLT2is, GLP1-RAs, DPP4is, and SUs new-users with established CVD in MDCD.** The preference score is a transformation of the propensity score that adjusts for prevalence differences between populations. For each class pair, we fit a large-scale propensity score model using >40,000 baseline patient characteristics. A higher overlap between score distributions indicates that patients in the target (left, red) and compartor (bottom, blue) populations are more similar in terms of their predicted probability of receiving one treatment over the other. We define clinical empirical equipoise when a majority of patients in both populations carry preference scores between 0.3 and 0.7.

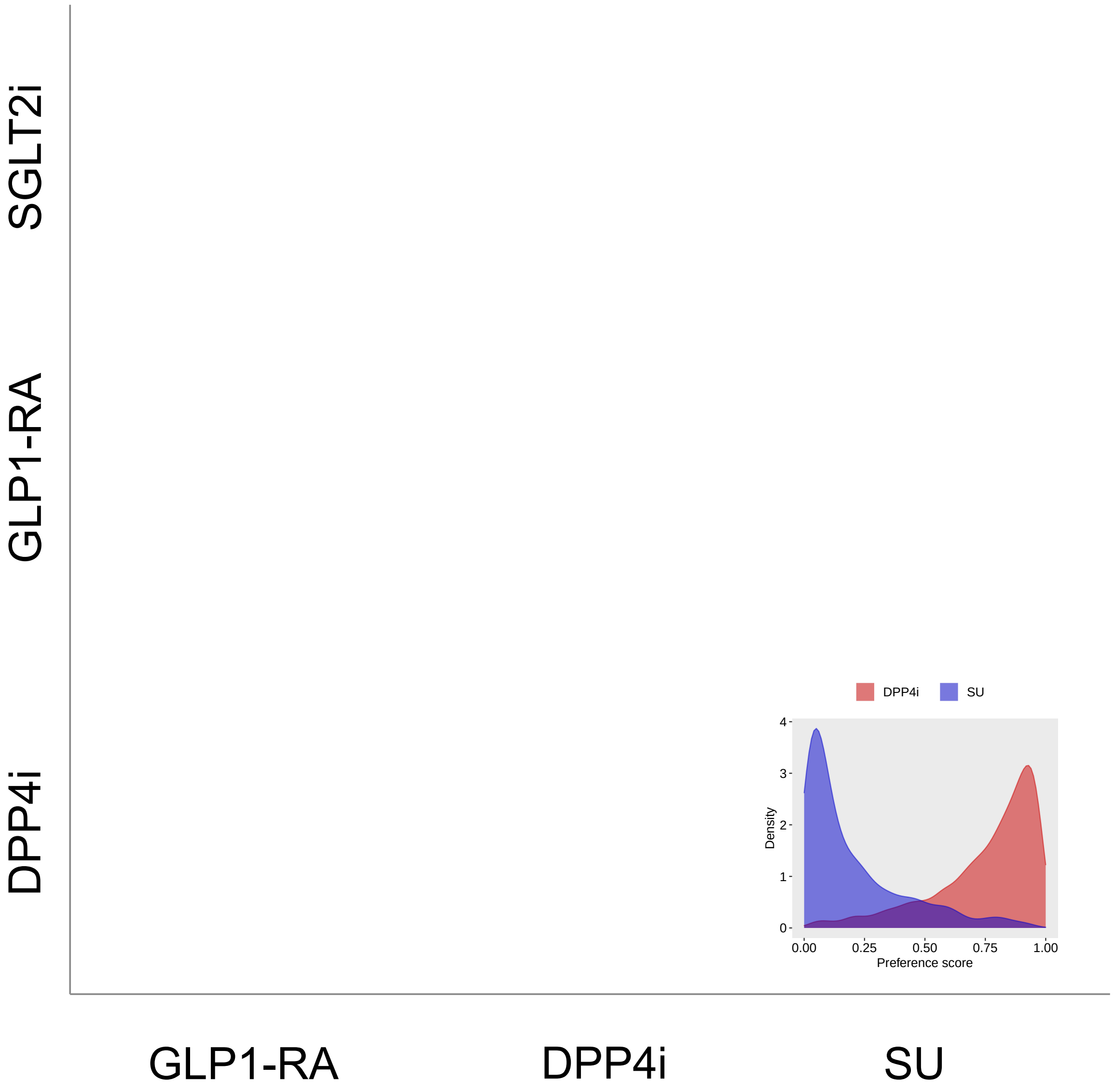


**Supplementary Figure 6: Preference score distributions comparing SGLT2is, GLP1-RAs, DPP4is, and SUs new-users with established CVD in IMRD.** The preference score is a transformation of the propensity score that adjusts for prevalence differences between populations. For each class pair, we fit a large-scale propensity score model using >40,000 baseline patient characteristics. A higher overlap between score distributions indicates that patients in the target (left, red) and compartor (bottom, blue) populations are more similar in terms of their predicted probability of receiving one treatment over the other. We define clinical empirical equipoise when a majority of patients in both populations carry preference scores between 0.3 and 0.7.

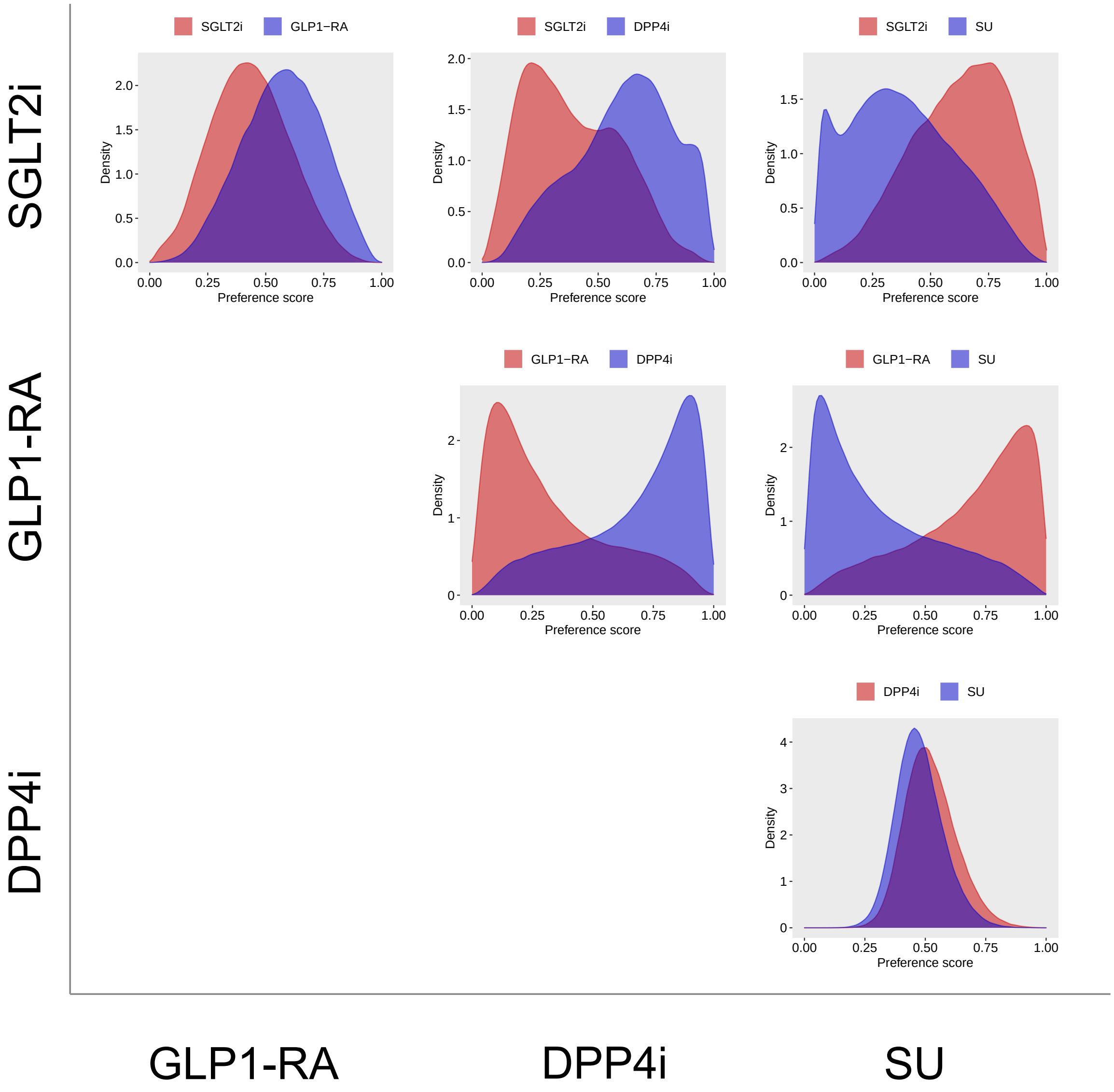


**Supplementary Figure 7: Preference score distributions comparing SGLT2is, GLP1-RAs, DPP4is, and SUs new-users with established CVD in US Open Claims.** The preference score is a transformation of the propensity score that adjusts for prevalence differences between populations. For each class pair, we fit a large-scale propensity score model using $>$40,000 baseline patient characteristics. A higher overlap between score distributions indicates that patients in the target (left, red) and compartor (bottom, blue) populations are more similar in terms of their predicted probability of receiving one treatment over the other. We define clinical empirical equipoise when a majority of patients in both populations carry preference scores between 0.3 and 0.7.

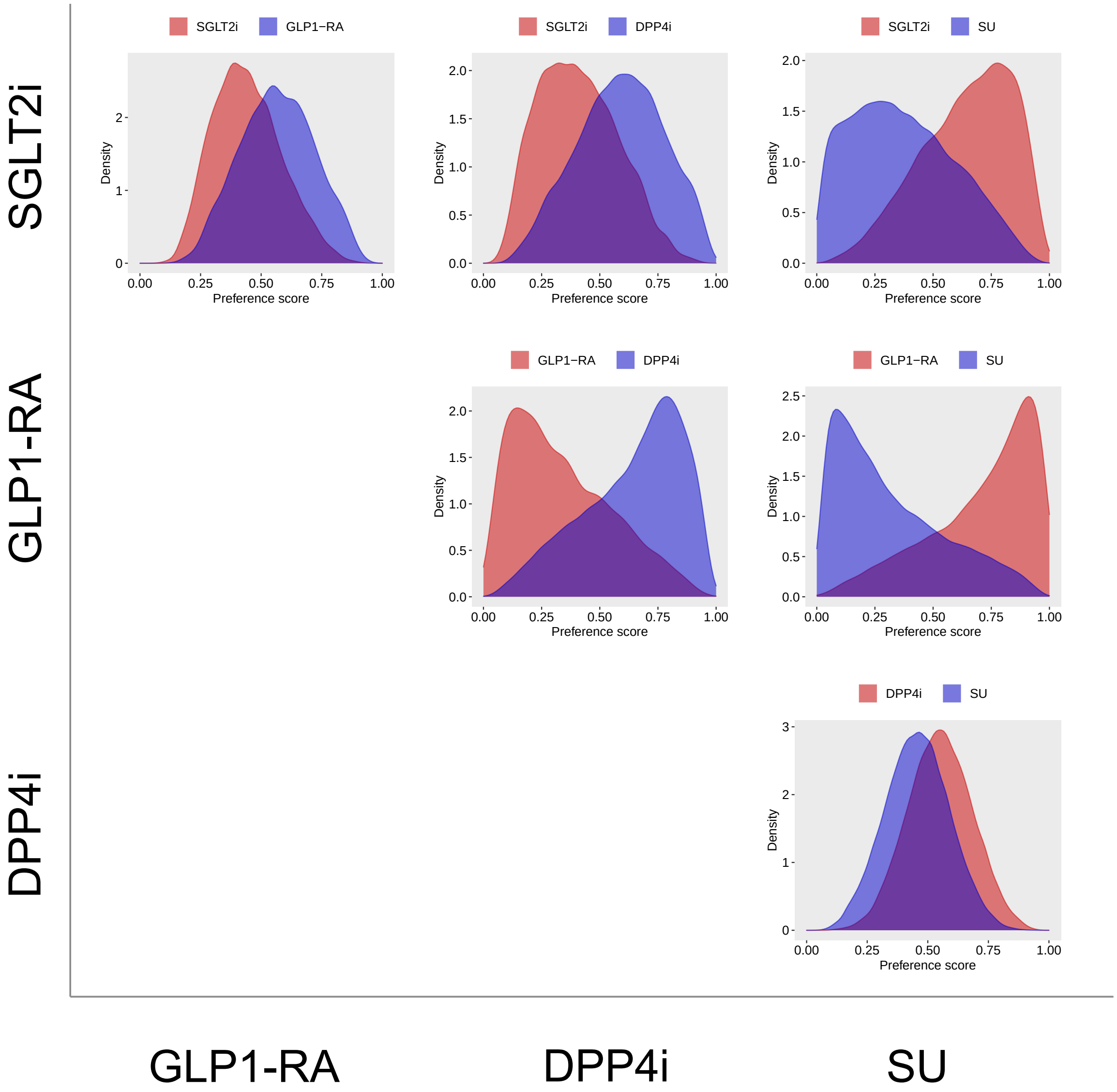


**Supplementary Figure 8: Preference score distributions comparing SGLT2is, GLP1-RAs, DPP4is, and SUs new-users with established CVD in OptumEHR.** The preference score is a transformation of the propensity score that adjusts for prevalence differences between populations. For each class pair, we fit a large-scale propensity score model using >40,000 baseline patient characteristics. A higher overlap between score distributions indicates that patients in the target (left, red) and compartor (bottom, blue) populations are more similar in terms of their predicted probability of receiving one treatment over the other. We define clinical empirical equipoise when a majority of patients in both populations carry preference scores between 0.3 and 0.7.

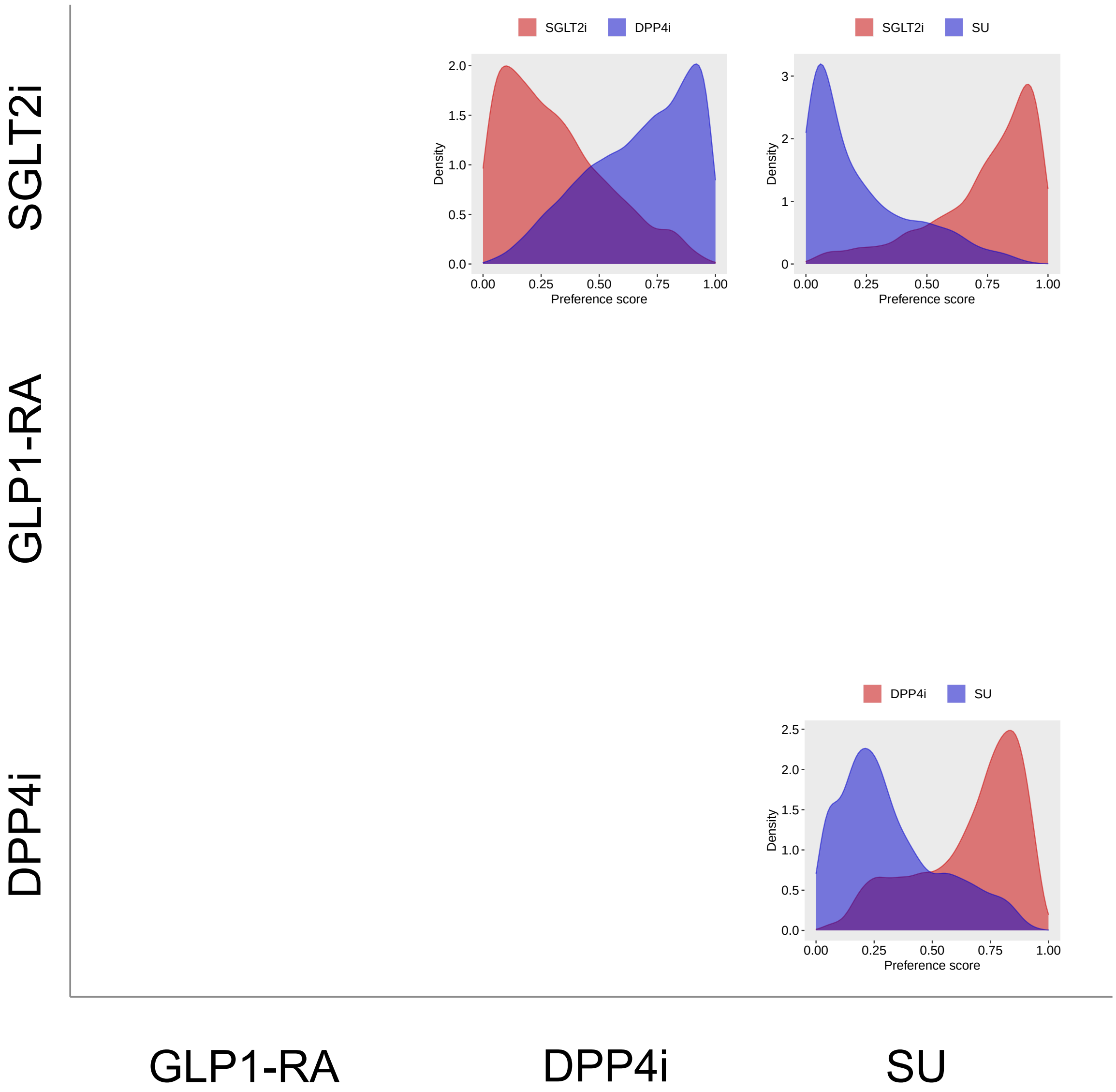


**Supplementary Figure 9: Preference score distributions comparing SGLT2is, GLP1-RAs, DPP4is, and SUs new-users with established CVD in SIDIAP.** The preference score is a transformation of the propensity score that adjusts for prevalence differences between populations. For each class pair, we fit a large-scale propensity score model using >40,000 baseline patient characteristics. A higher overlap between score distributions indicates that patients in the target (left, red) and compartor (bottom, blue) populations are more similar in terms of their predicted probability of receiving one treatment over the other. We define clinical empirical equipoise when a majority of patients in both populations carry preference scores between 0.3 and 0.7.

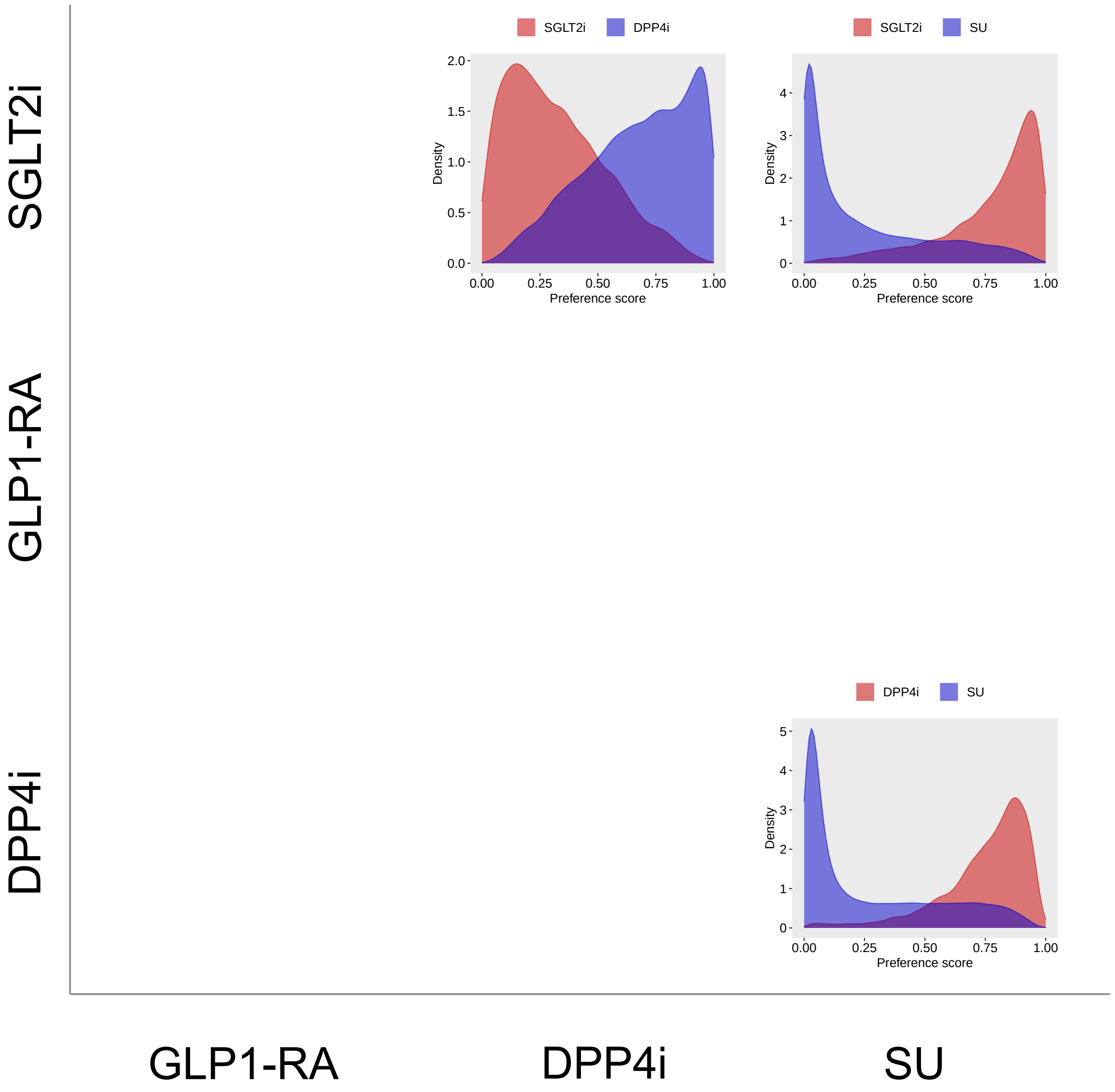


**Supplementary Figure 10: Preference score distributions comparing SGLT2is, GLP1-RAs, DPP4is, and SUs new-users with established CVD in VA.** The preference score is a transformation of the propensity score that adjusts for prevalence differences between populations. For each class pair, we fit a large-scale propensity score model using >40,000 baseline patient characteristics. A higher overlap between score distributions indicates that patients in the target (left, red) and compartor (bottom, blue) populations are more similar in terms of their predicted probability of receiving one treatment over the other. We define clinical empirical equipoise when a majority of patients in both populations carry preference scores between 0.3 and 0.7.

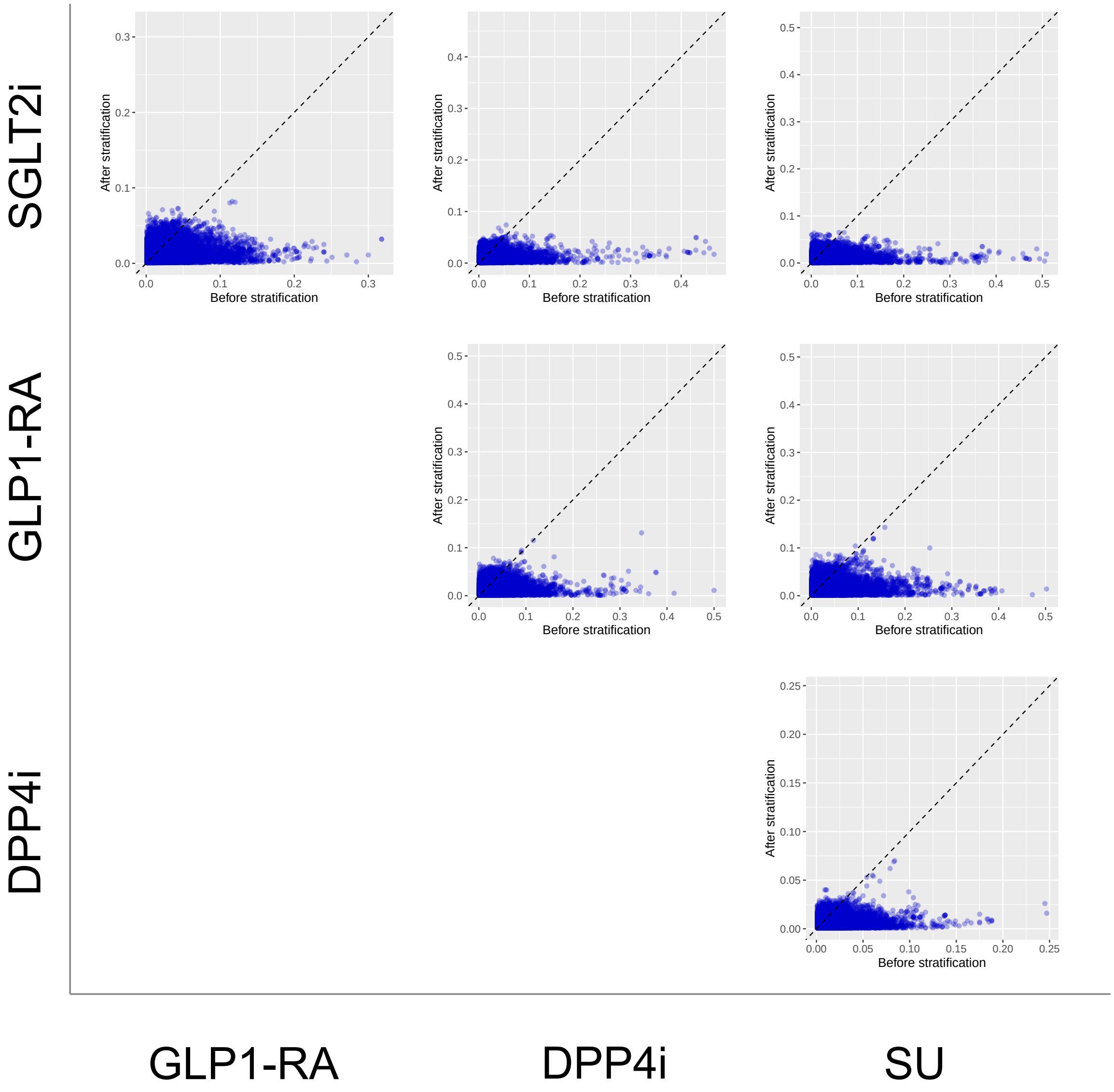


**Supplementary Figure 11: Covariate balance comparing SGLT2is, GLP1-RAs, DPP4is, and SUs new-users with established CVD in CCAE before/after PS stratification.** Covariate balance is measured by standardized mean differences (SMDs) between covariates in target and comparator cohorts. We show SMDs on >40,000 baseline patient characteristics before and after PS stratification.

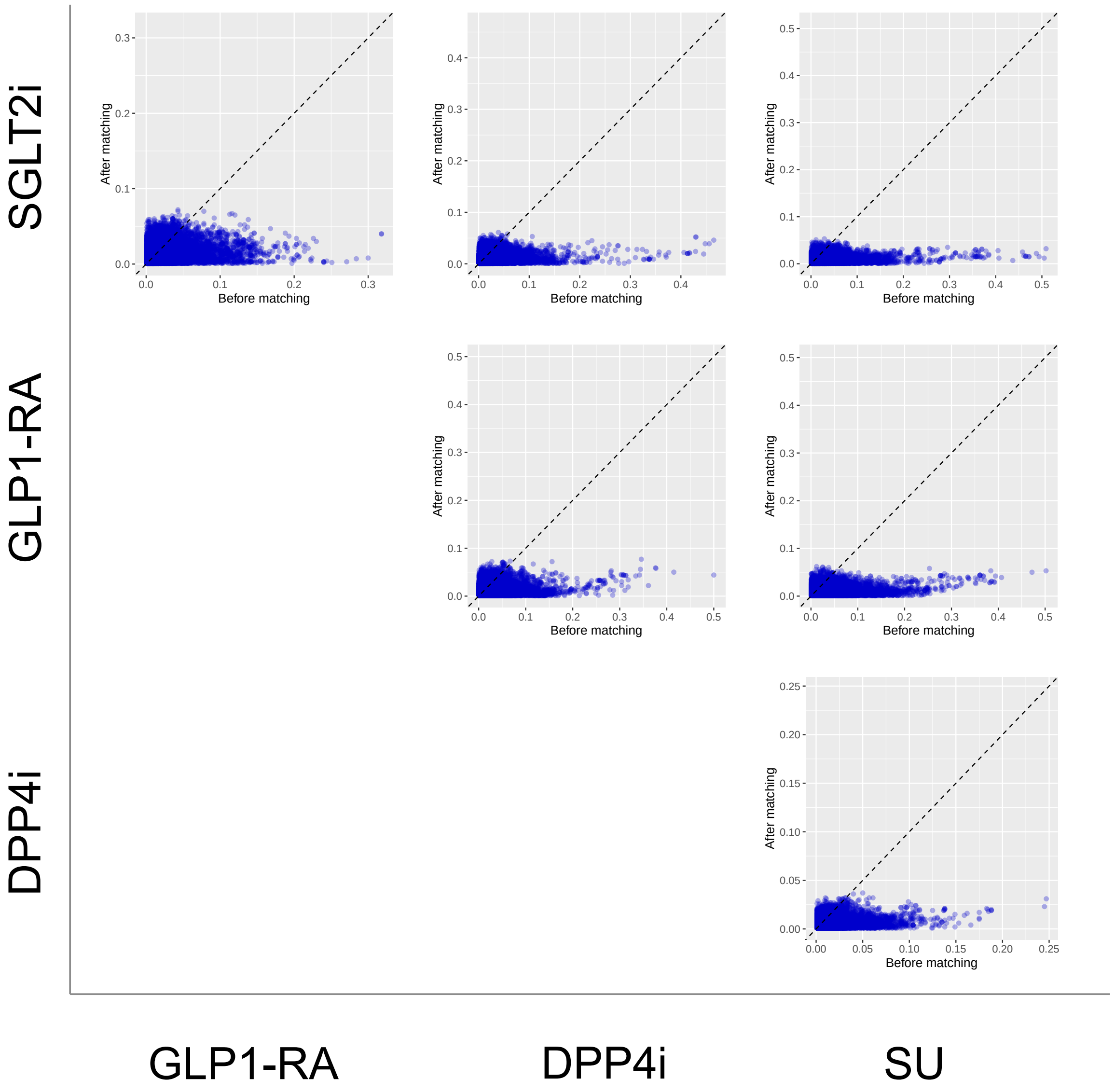


**Supplementary Figure 12: Covariate balance comparing SGLT2is, GLP1-RAs, DPP4is, and SUs new-users with established CVD in CCAE before/after PS matching.** Covariate balance is measured by standardized mean differences (SMDs) between covariates in target and comparator cohorts. We show SMDs on $>$40,000 baseline patient characteristics before and after PS matching.

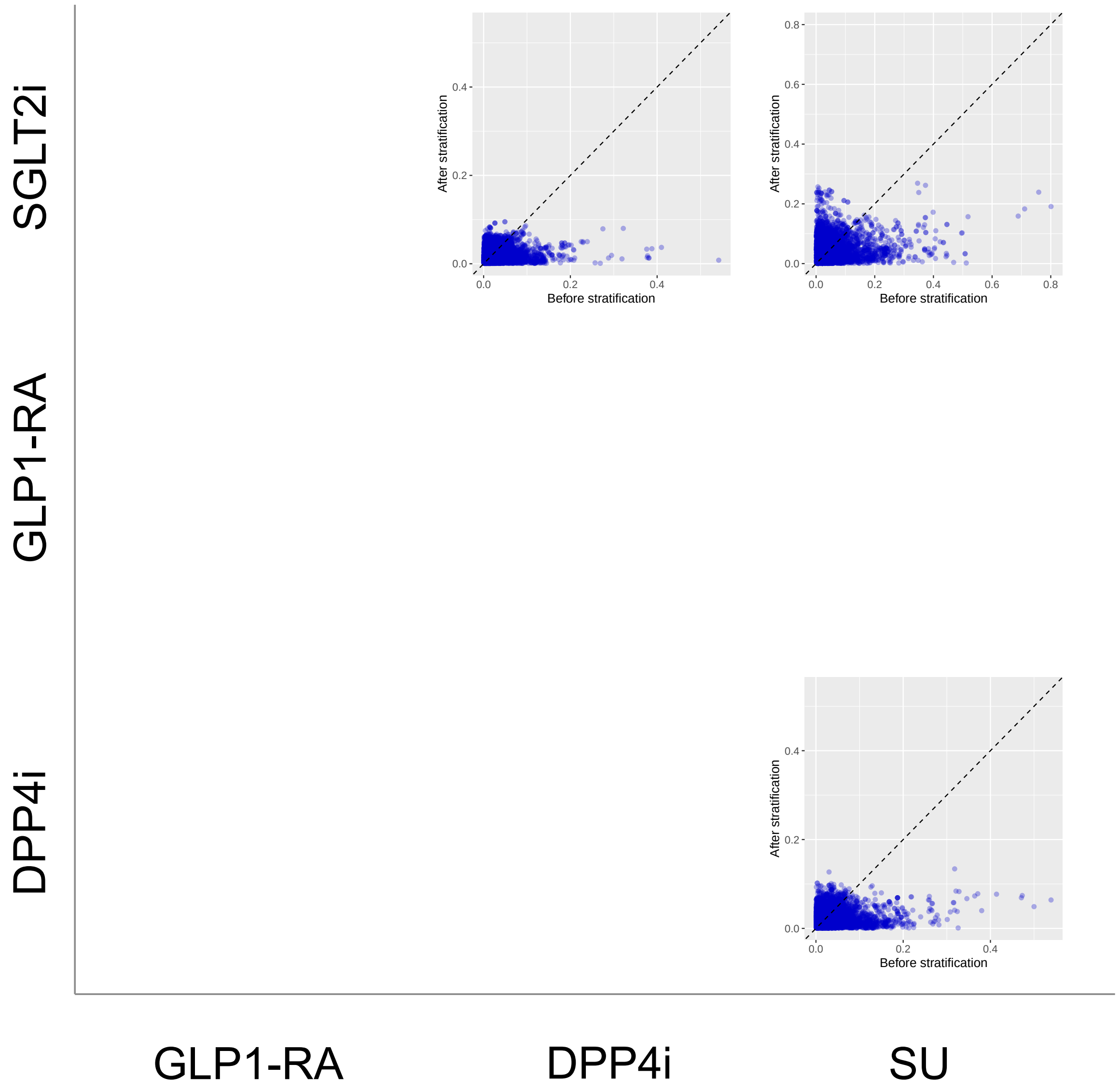


**Supplementary Figure 13: Covariate balance comparing SGLT2is, GLP1-RAs, DPP4is, and SUs new-users with established CVD in GDA before/after PS stratification.** Covariate balance is measured by standardized mean differences (SMDs) between covariates in target and comparator cohorts. We show SMDs on >40,000 baseline patient characteristics before and after PS stratification.

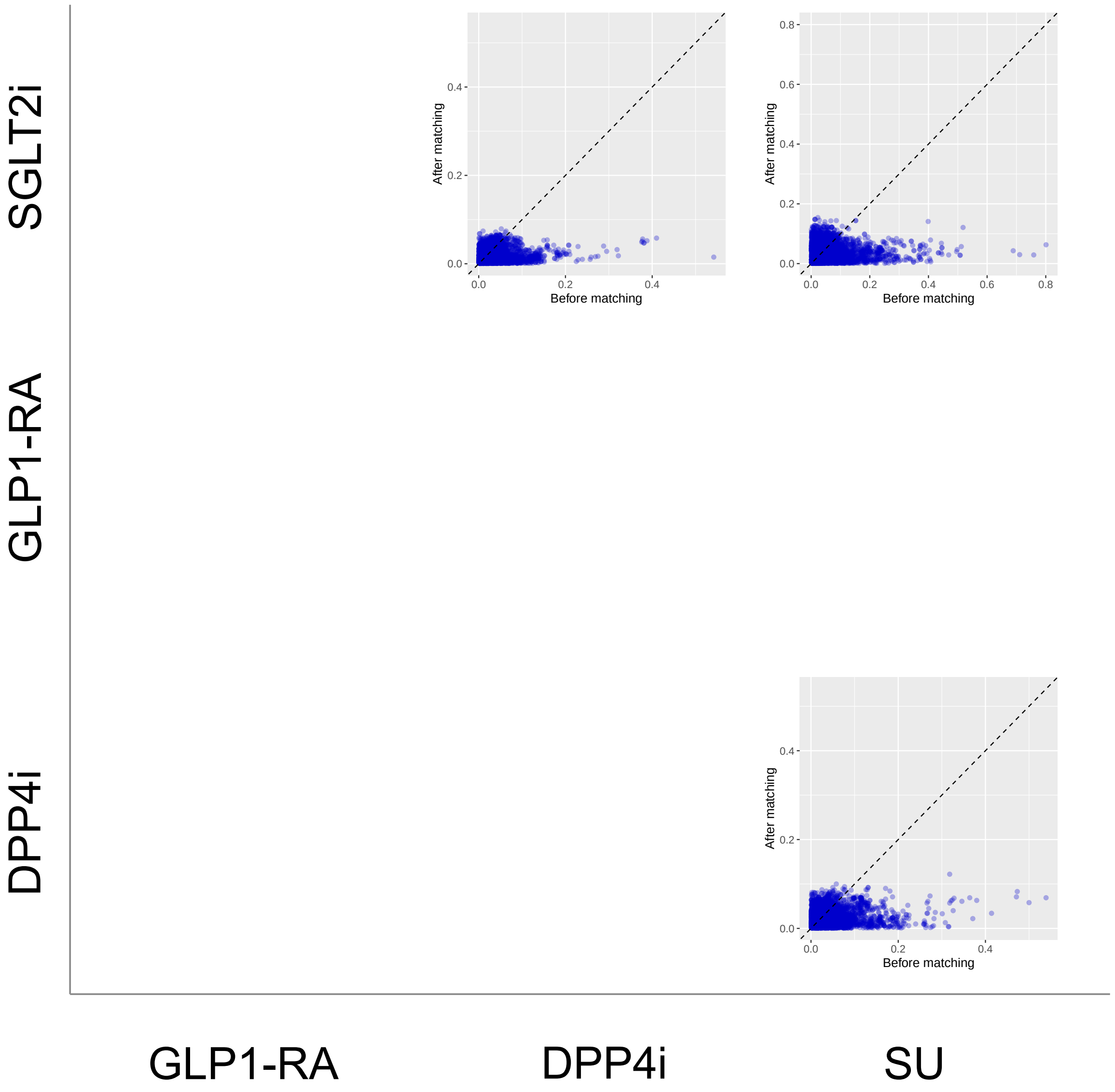


**Supplementary Figure 14: Covariate balance comparing SGLT2is, GLP1-RAs, DPP4is, and SUs new-users with established CVD in GDA before/after PS matching.** Covariate balance is measured by standardized mean differences (SMDs) between covariates in target and comparator cohorts. We show SMDs on >40,000 baseline patient characteristics before and after PS matching.

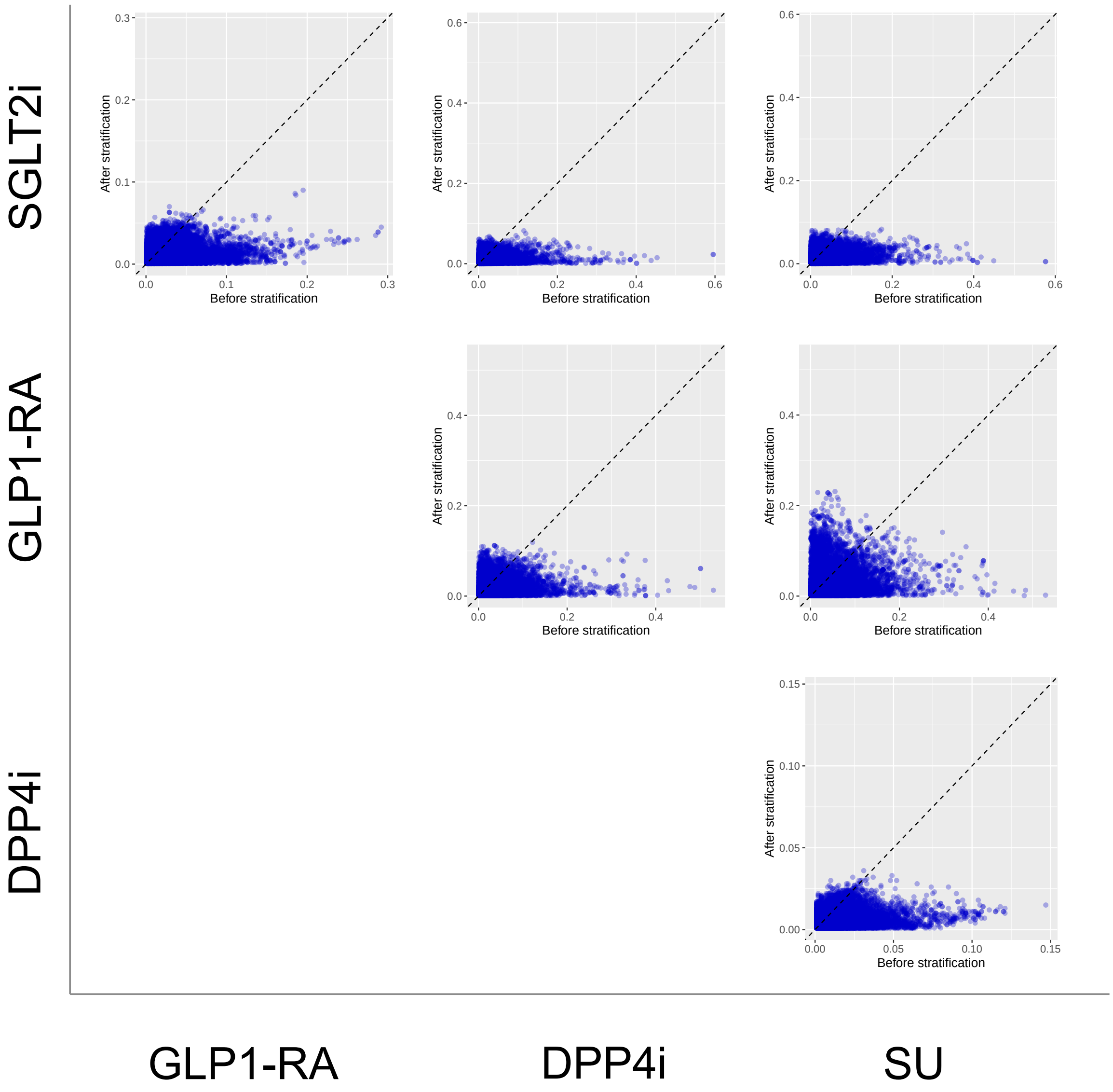


**Supplementary Figure 15: Covariate balance comparing SGLT2is, GLP1-RAs, DPP4is, and SUs new-users with established CVD in OptumDOD before/after PS stratification.** Covariate balance is measured by standardized mean differences (SMDs) between covariates in target and comparator cohorts. We show SMDs on $>$40,000 baseline patient characteristics before and after PS stratification.

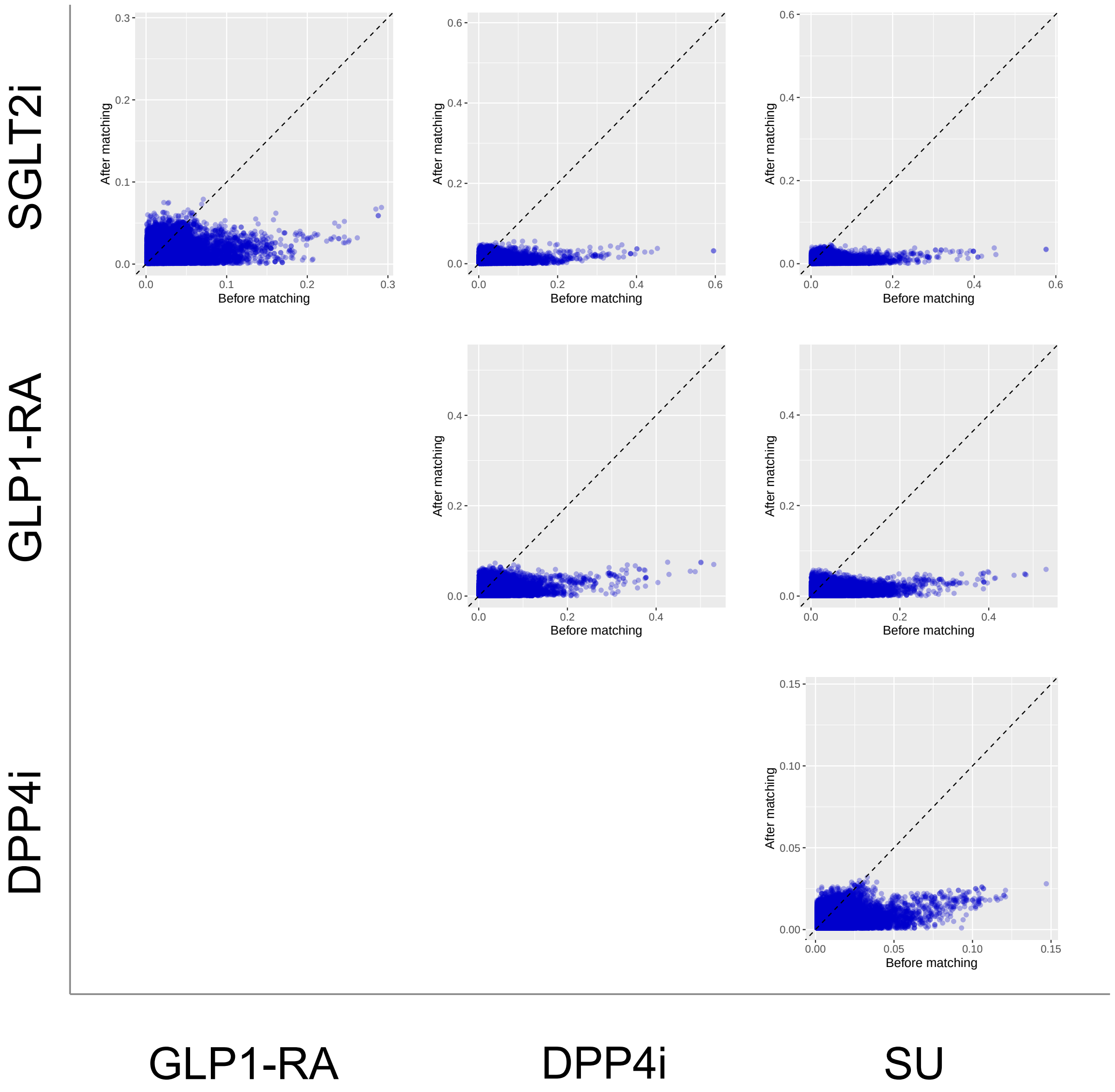


**Supplementary Figure 16: Covariate balance comparing SGLT2is, GLP1-RAs, DPP4is, and SUs new-users with established CVD in OptumDOD before/after PS matching.** Covariate balance is measured by standardized mean differences (SMDs) between covariates in target and comparator cohorts. We show SMDs on >40,000 baseline patient characteristics before and after PS matching.

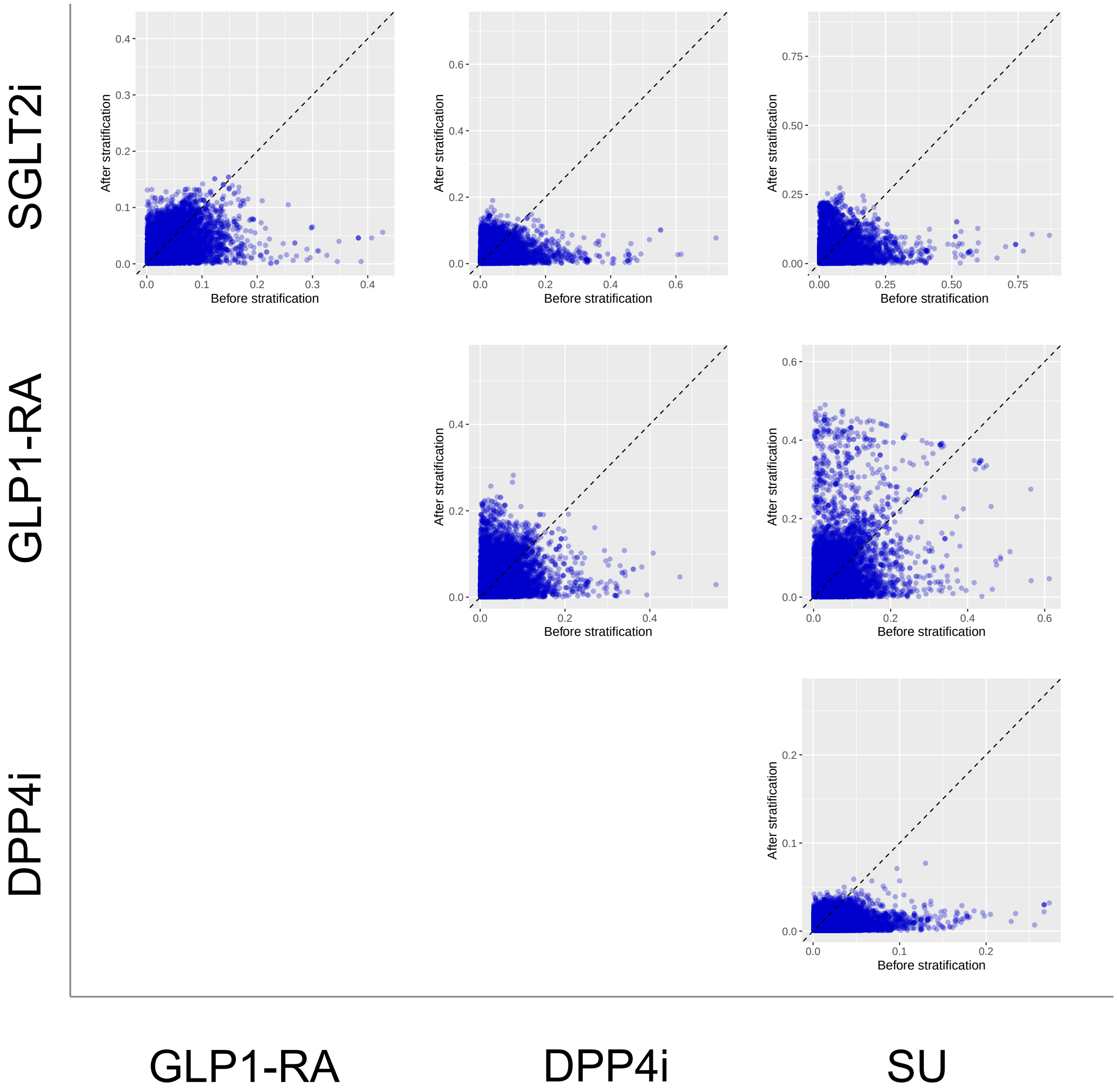


**Supplementary Figure 17: Covariate balance comparing SGLT2is, GLP1-RAs, DPP4is, and SUs new-users with established CVD in MDCR before/after PS stratification.** Covariate balance is measured by standardized mean differences (SMDs) between covariates in target and comparator cohorts. We show SMDs on >40,000 baseline patient characteristics before and after PS stratification.

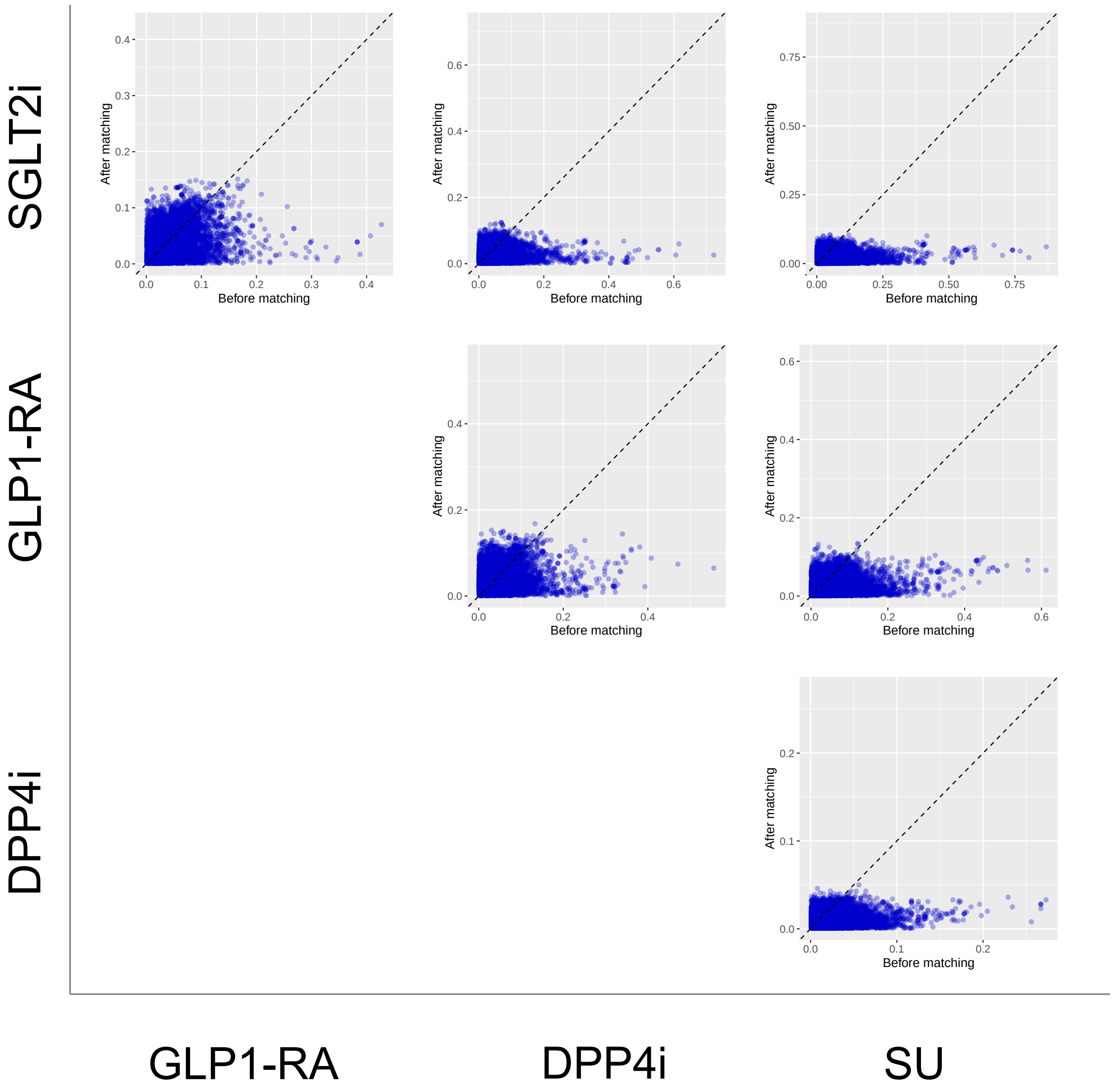


**Supplementary Figure 18: Covariate balance comparing SGLT2is, GLP1-RAs, DPP4is, and SUs new-users with established CVD in MDCR before/after PS matching.** Covariate balance is measured by standardized mean differences (SMDs) between covariates in target and comparator cohorts. We show SMDs on >40,000 baseline patient characteristics before and after PS matching.

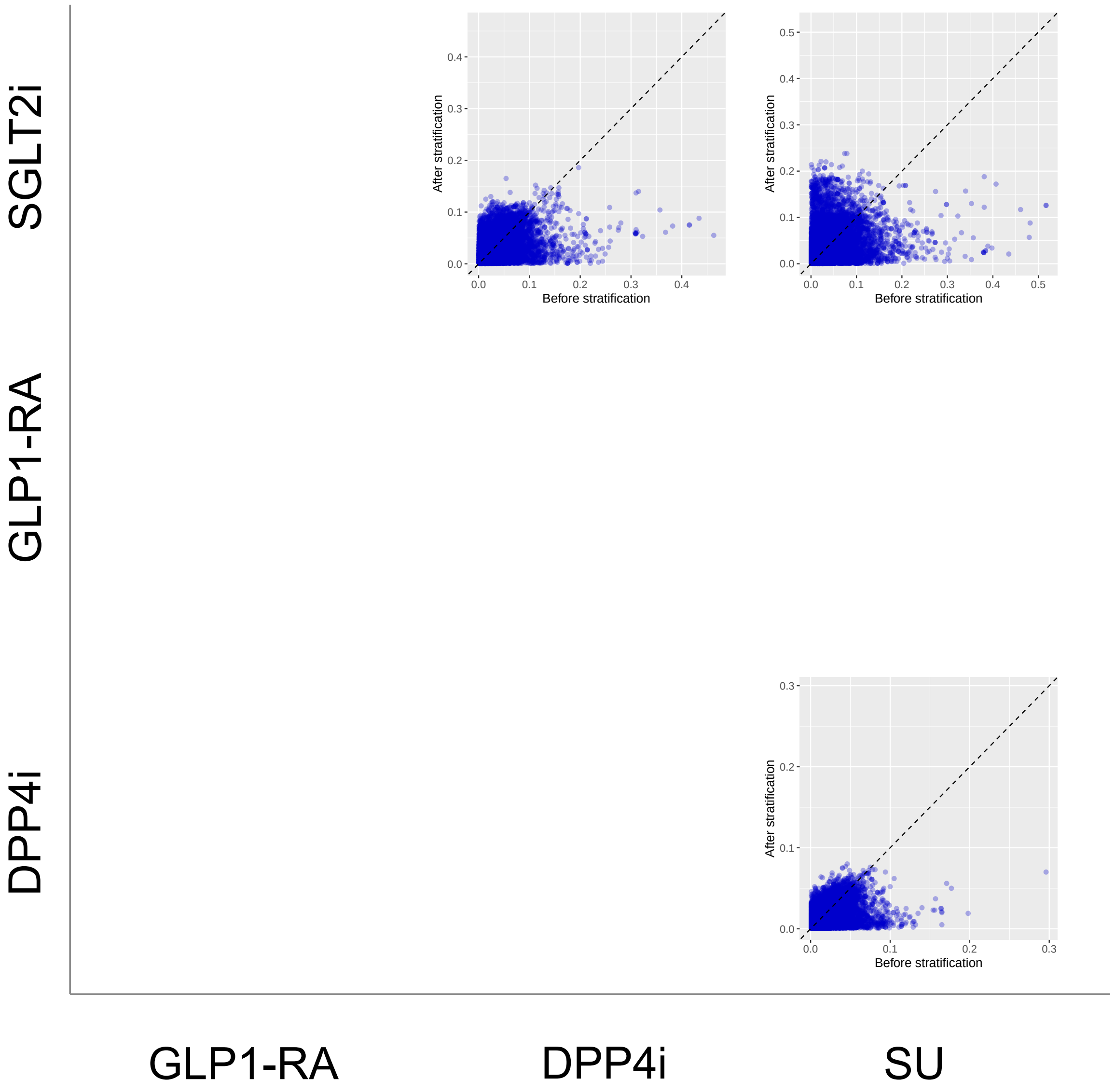


**Supplementary Figure 19: Covariate balance comparing SGLT2is, GLP1-RAs, DPP4is, and SUs new-users with established CVD in MDCD before/after PS stratification.** Covariate balance is measured by standardized mean differences (SMDs) between covariates in target and comparator cohorts. We show SMDs on >40,000 baseline patient characteristics before and after PS stratification.

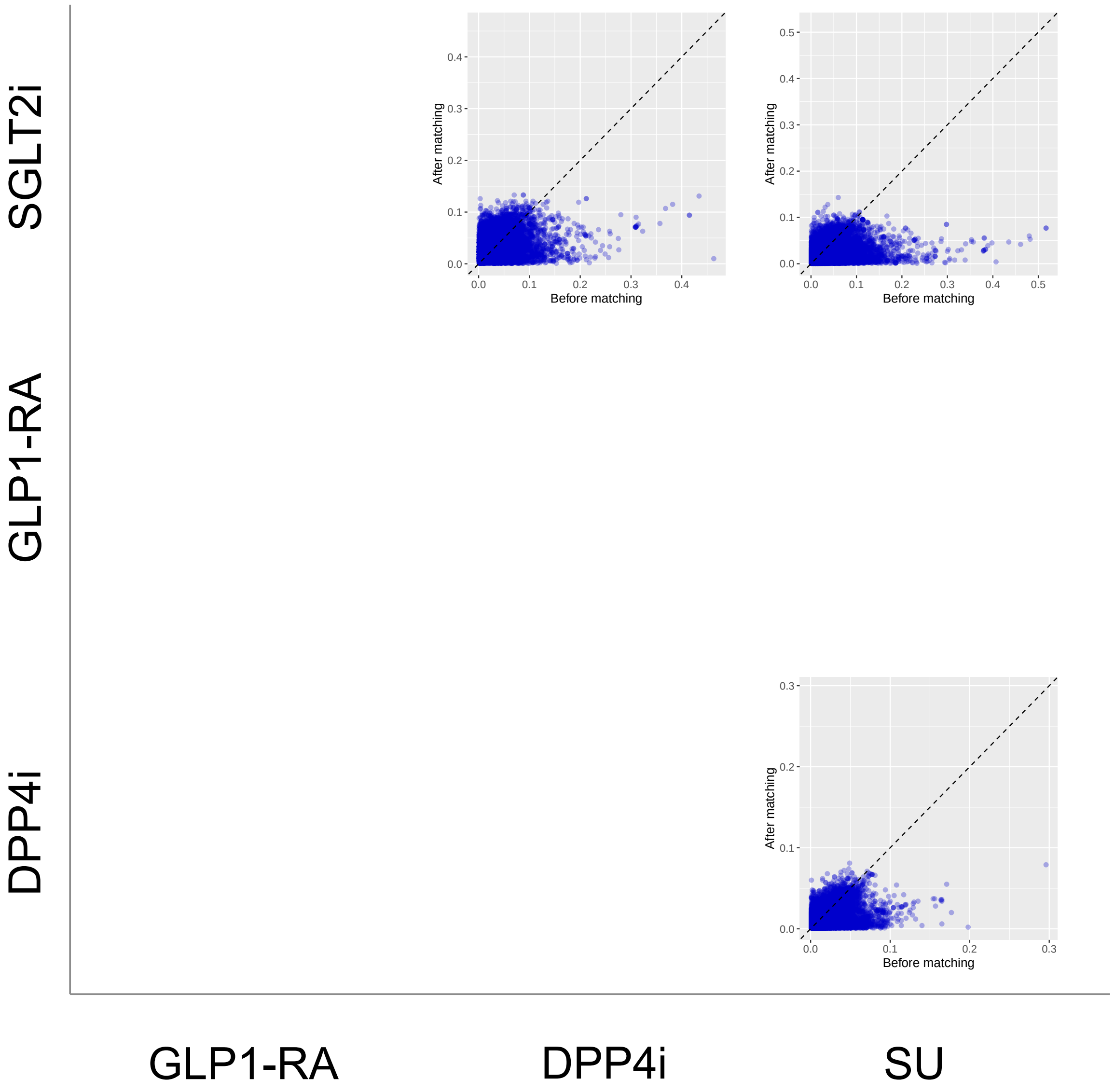


**Supplementary Figure 20: Covariate balance comparing SGLT2is, GLP1-RAs, DPP4is, and SUs new-users with established CVD in MDCD before/after PS matching.** Covariate balance is measured by standardized mean differences (SMDs) between covariates in target and comparator cohorts. We show SMDs on $>$40,000 baseline patient characteristics before and after PS matching.

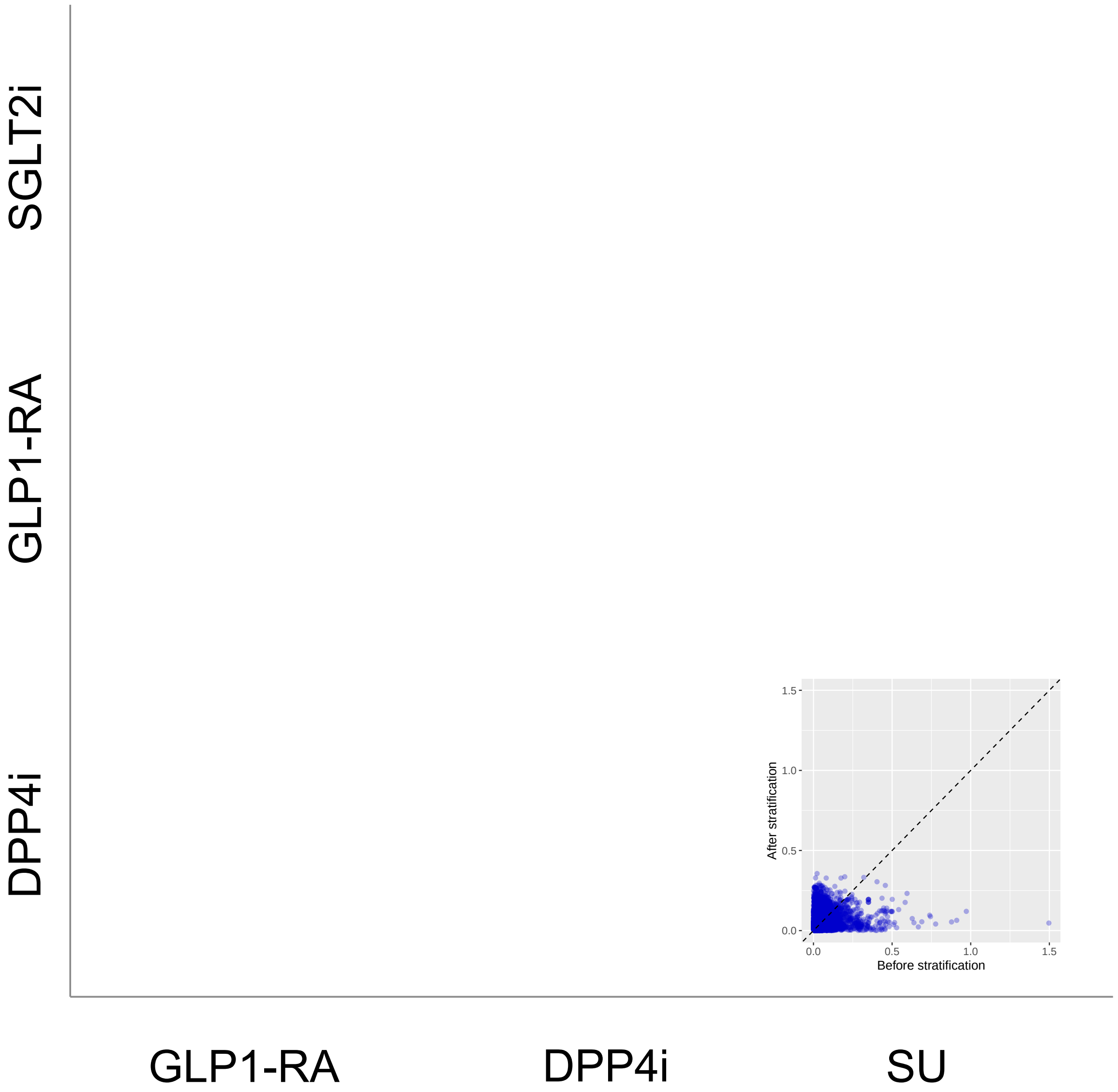


**Supplementary Figure 21: Covariate balance comparing SGLT2is, GLP1-RAs, DPP4is, and SUs new-users with established CVD in IMRD before/after PS stratification.** Covariate balance is measured by standardized mean differences (SMDs) between covariates in target and comparator cohorts. We show SMDs on $>$40,000 baseline patient characteristics before and after PS stratification.

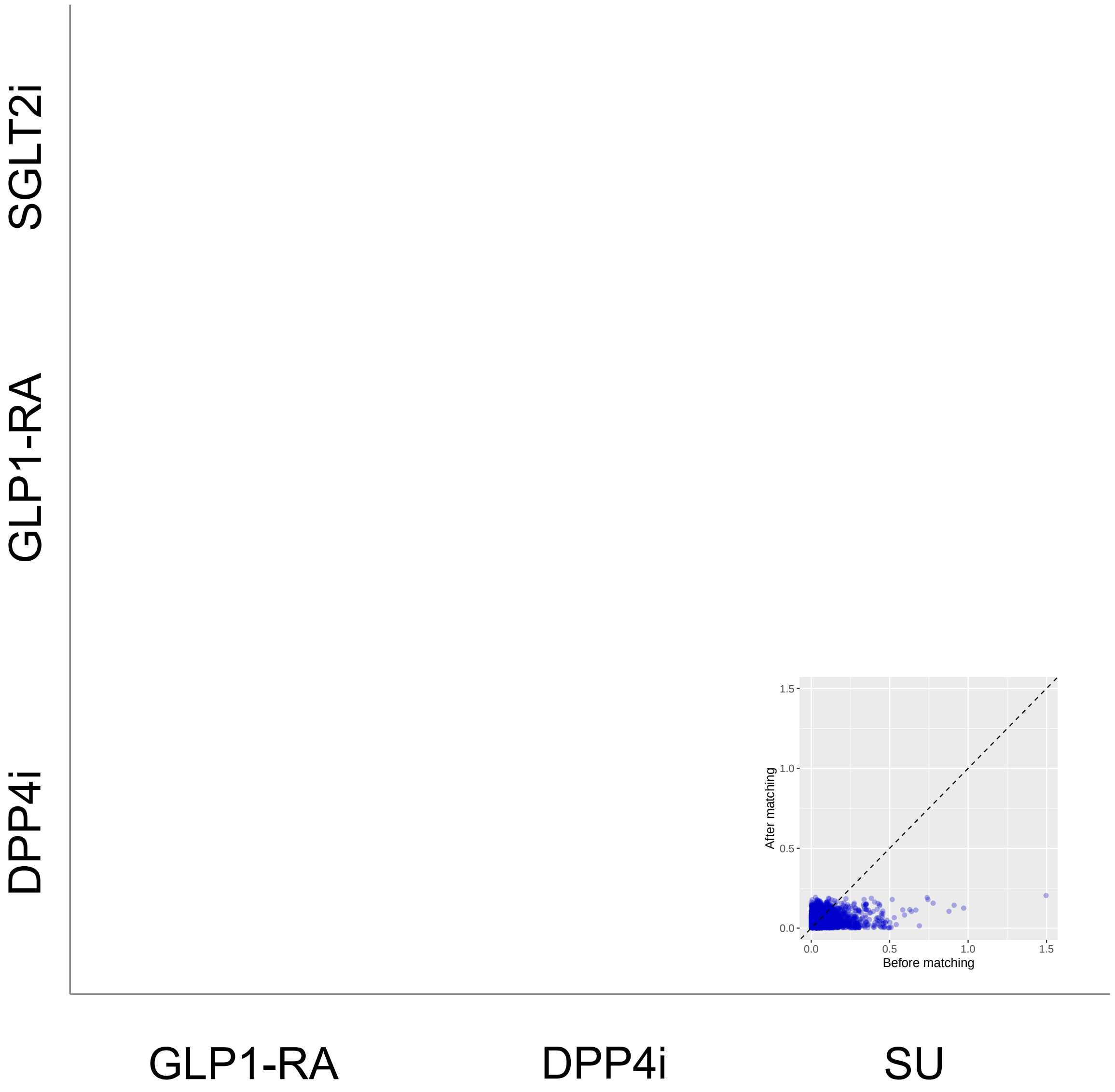


**Supplementary Figure 22: Covariate balance comparing SGLT2is, GLP1-RAs, DPP4is, and SUs new-users with established CVD in IMRD before/after PS matching.** Covariate balance is measured by standardized mean differences (SMDs) between covariates in target and comparator cohorts. We show SMDs on >40,000 baseline patient characteristics before and after PS matching.

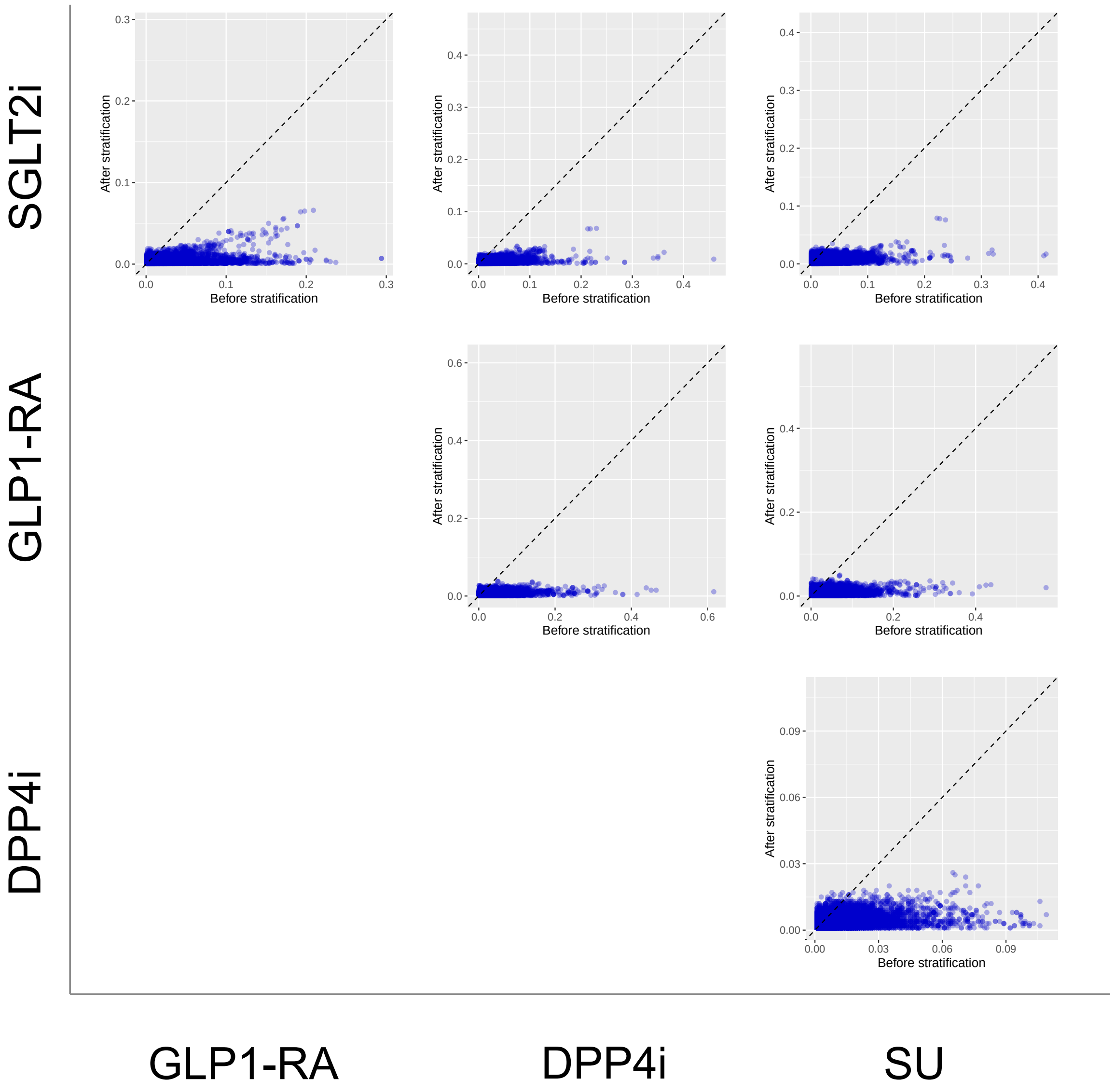


**Supplementary Figure 23: Covariate balance comparing SGLT2is, GLP1-RAs, DPP4is, and SUs new-users with established CVD in US Open Claims before/after PS stratification.** Covariate balance is measured by standardized mean differences (SMDs) between covariates in target and comparator cohorts. We show SMDs on $>$40,000 baseline patient characteristics before and after PS stratification.

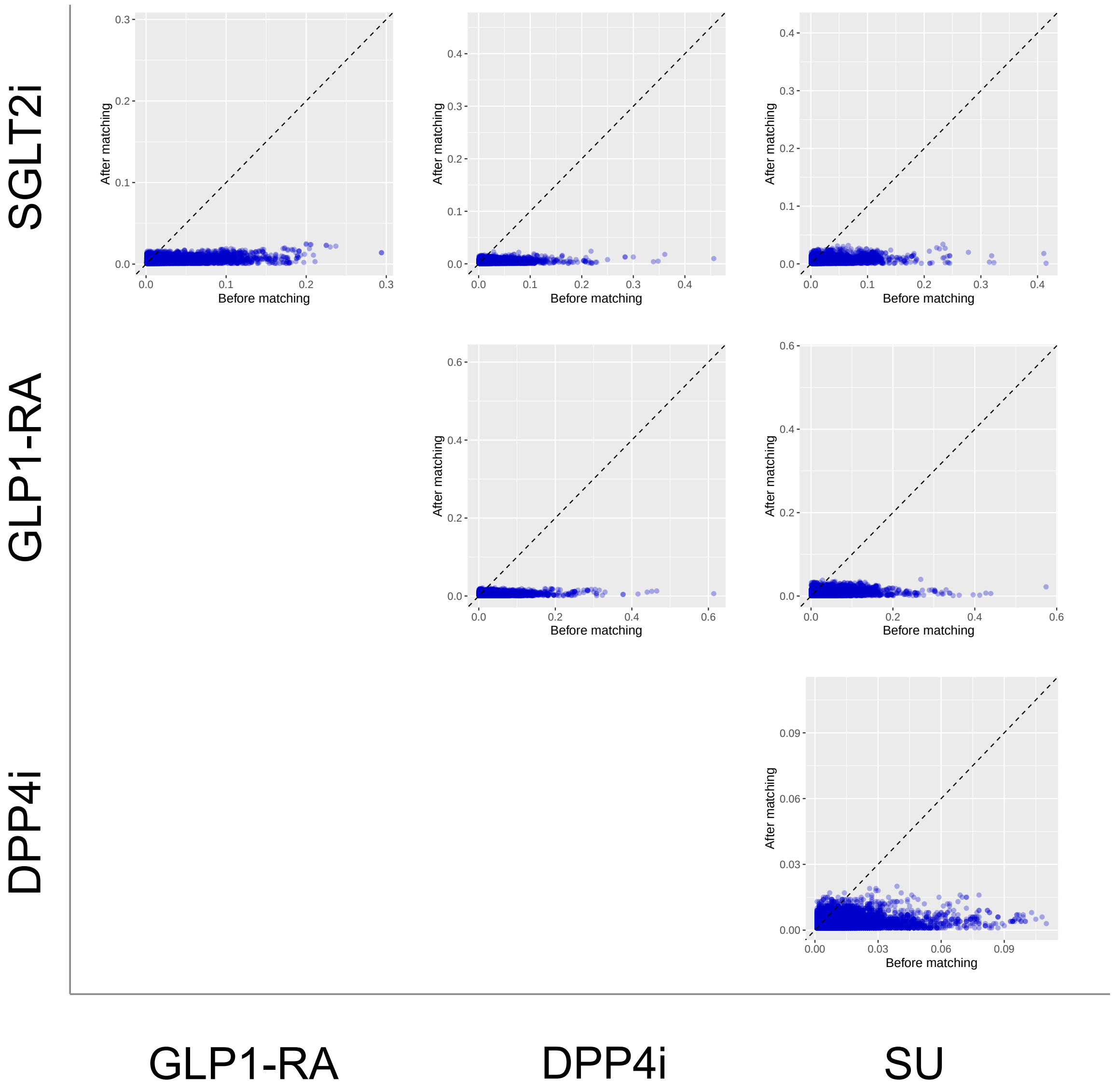


**Supplementary Figure 24: Covariate balance comparing SGLT2is, GLP1-RAs, DPP4is, and SUs new-users with established CVD in US Open Claims before/after PS matching.** Covariate balance is measured by standardized mean differences (SMDs) between covariates in target and comparator cohorts. We show SMDs on >40,000 baseline patient characteristics before and after PS matching.

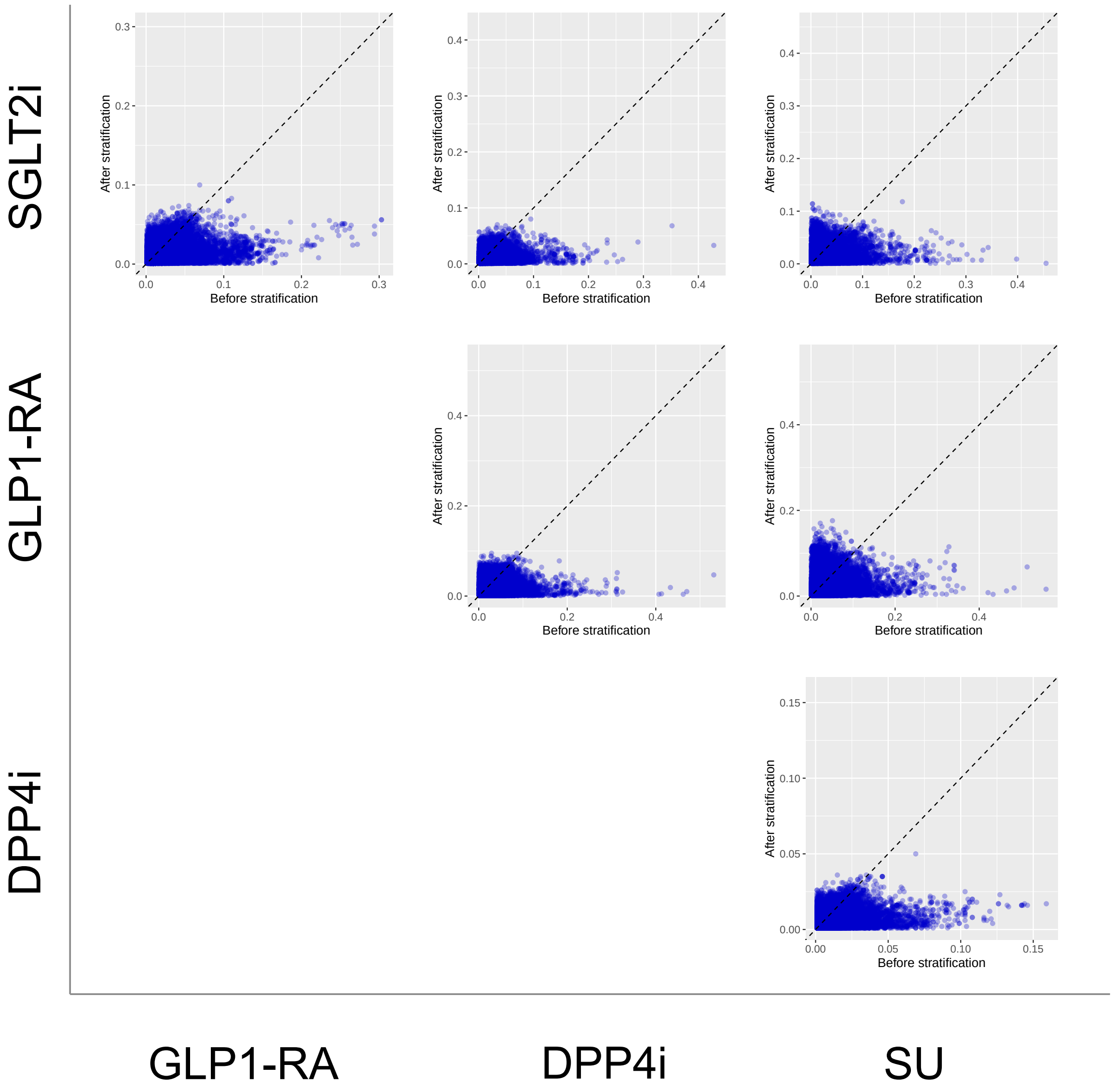


**Supplementary Figure 25: Covariate balance comparing SGLT2is, GLP1-RAs, DPP4is, and SUs new-users with established CVD in OptumEHR before/after PS stratification.** Covariate balance is measured by standardized mean differences (SMDs) between covariates in target and comparator cohorts. We show SMDs on >40,000 baseline patient characteristics before and after PS stratification.

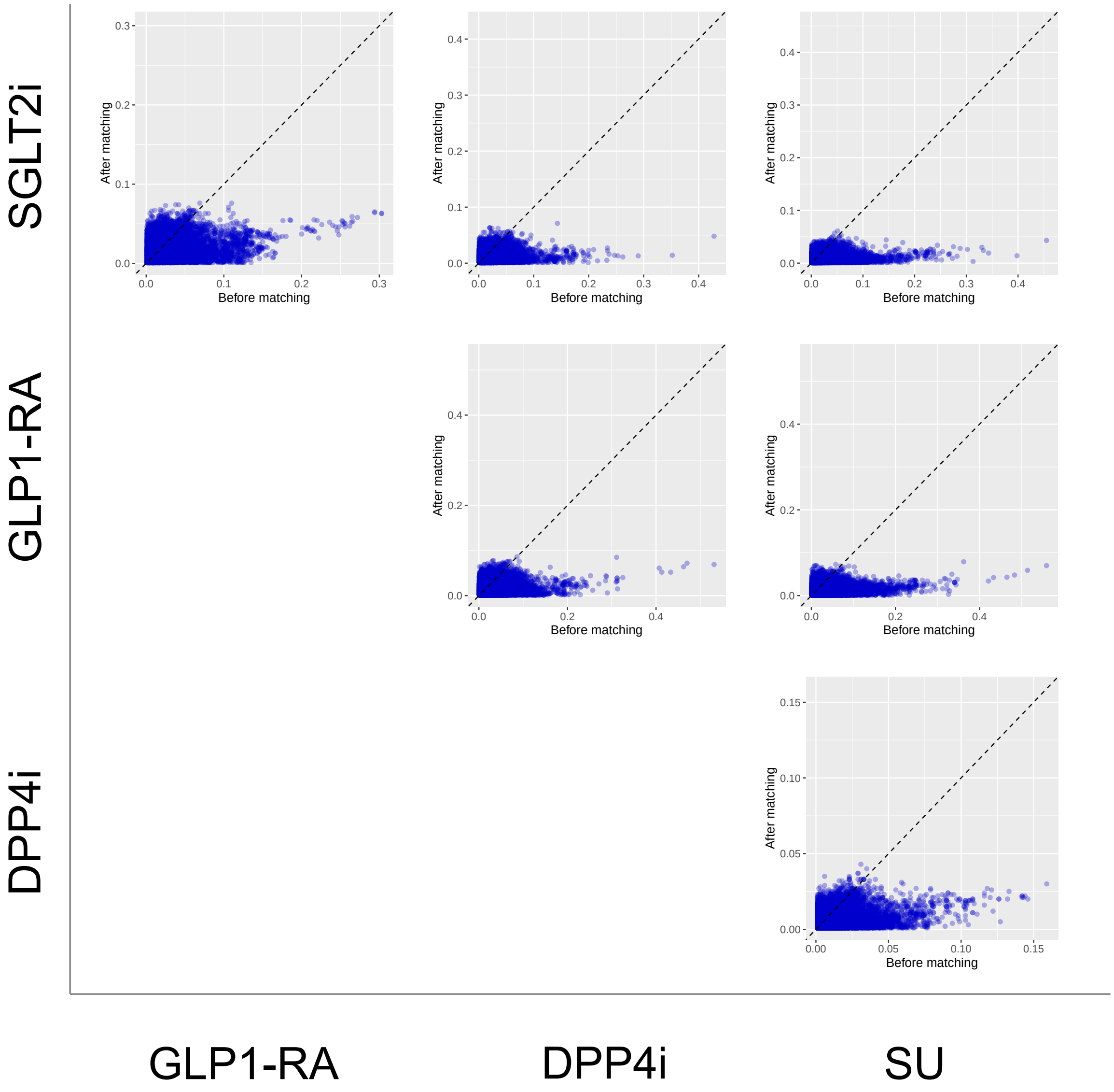


**Supplementary Figure 26: Covariate balance comparing SGLT2is, GLP1-RAs, DPP4is, and SUs new-users with established CVD in OptumEHR before/after PS matching.** Covariate balance is measured by standardized mean differences (SMDs) between covariates in target and comparator cohorts. We show SMDs on >40,000 baseline patient characteristics before and after PS matching.

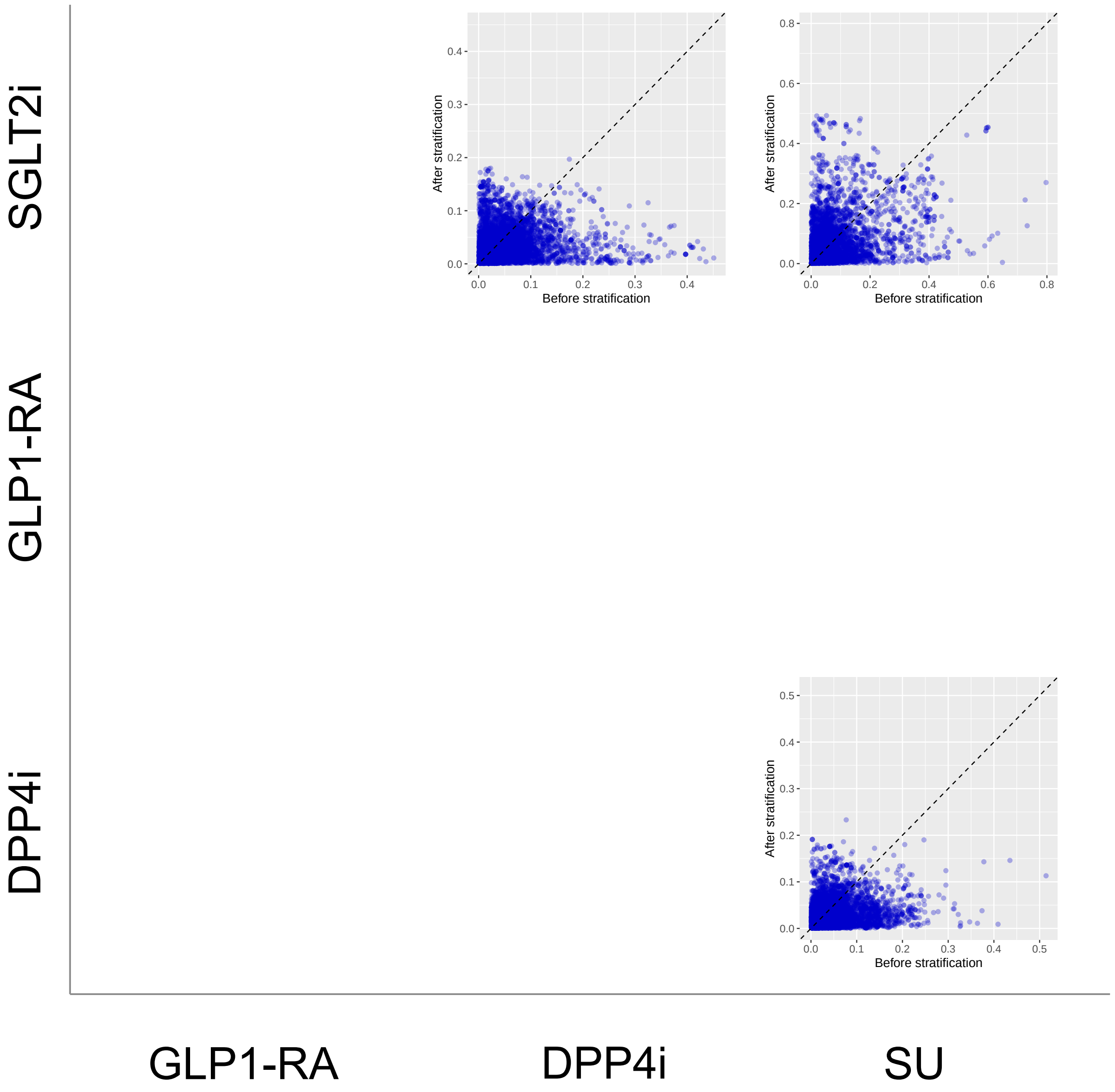


**Supplementary Figure 27: Covariate balance comparing SGLT2is, GLP1-RAs, DPP4is, and SUs new-users with established CVD in SIDIAP before/after PS stratification.** Covariate balance is measured by standardized mean differences (SMDs) between covariates in target and comparator cohorts. We show SMDs on >40,000 baseline patient characteristics before and after PS stratification.

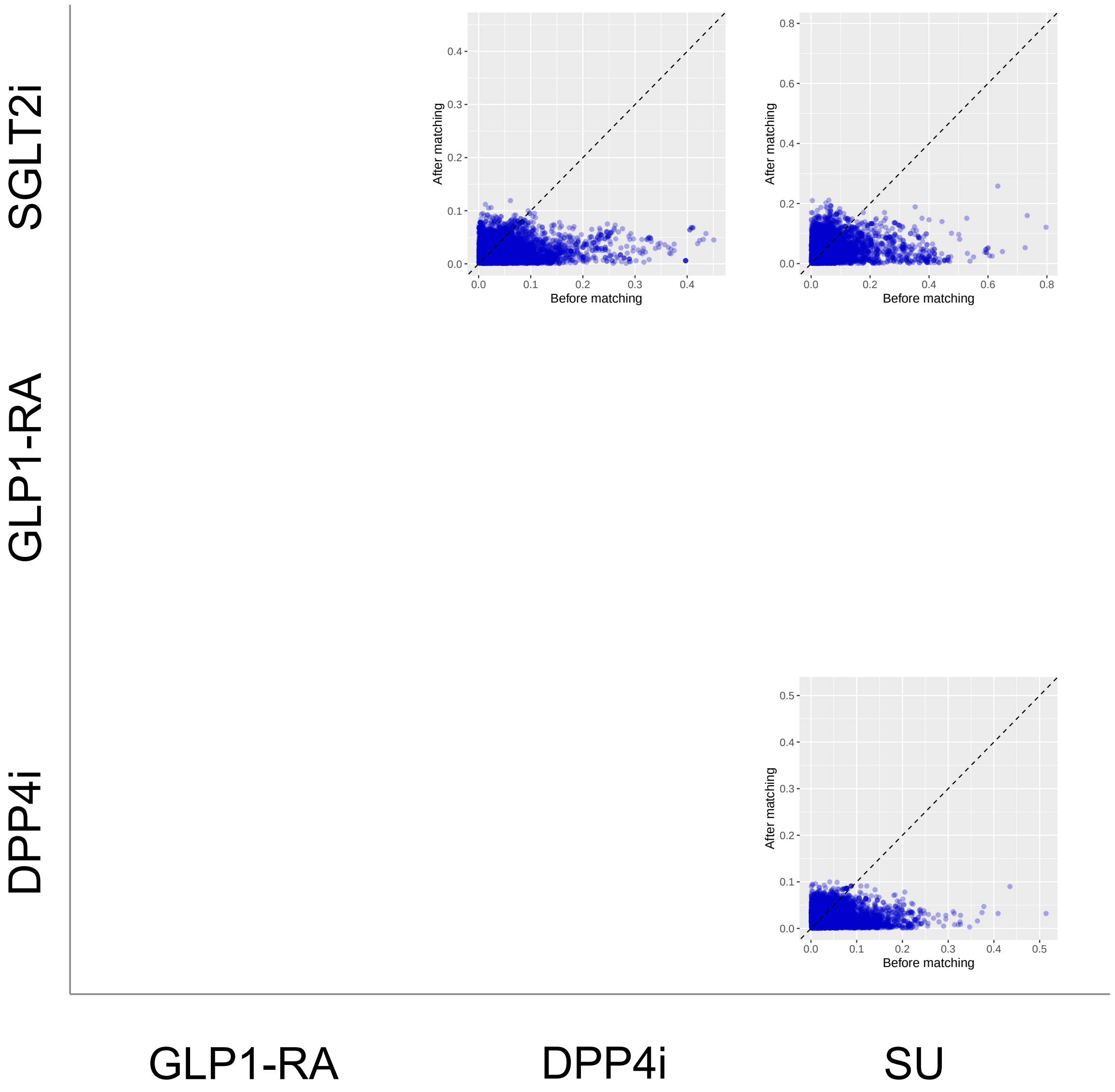


**Supplementary Figure 28: Covariate balance comparing SGLT2is, GLP1-RAs, DPP4is, and SUs new-users with established CVD in SIDIAP before/after PS matching.** Covariate balance is measured by standardized mean differences (SMDs) between covariates in target and comparator cohorts. We show SMDs on >40,000 baseline patient characteristics before and after PS matching.

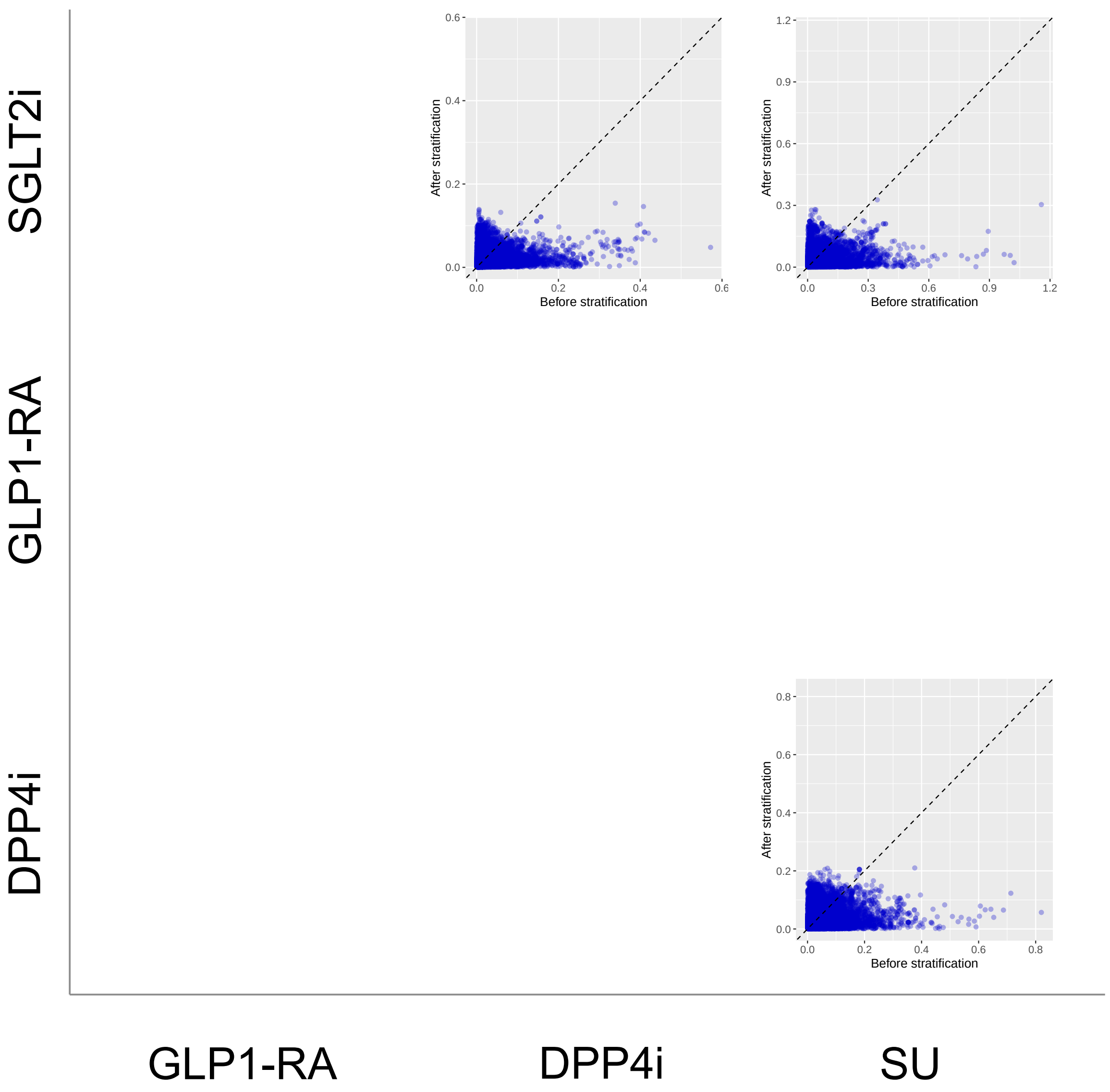


**Supplementary Figure 29: Covariate balance comparing SGLT2is, GLP1-RAs, DPP4is, and SUs new-users with established CVD in VA before/after PS stratification.** Covariate balance is measured by standardized mean differences (SMDs) between covariates in target and comparator cohorts. We show SMDs on $>$40,000 baseline patient characteristics before and after PS stratification.

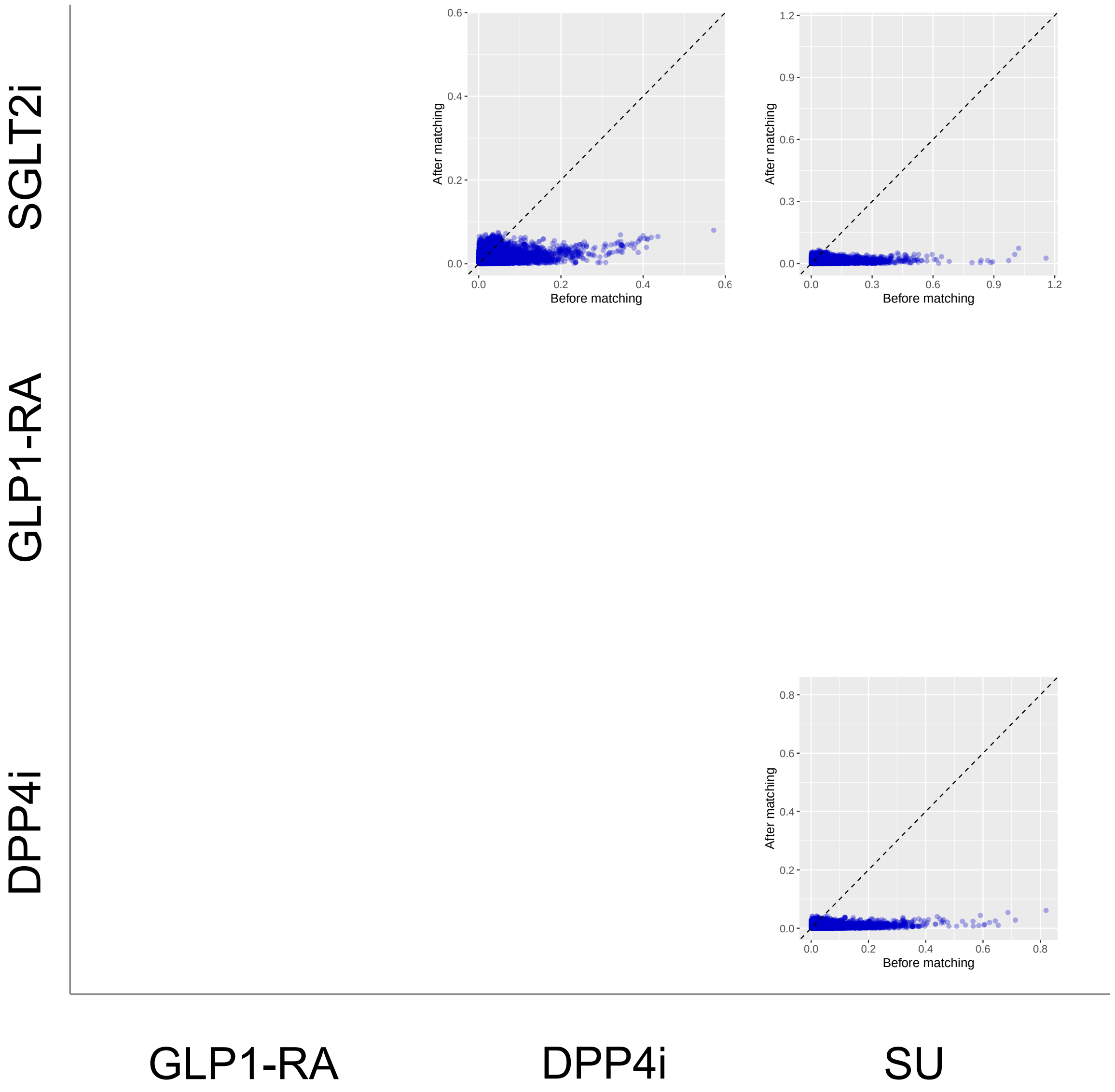


**Supplementary Figure 30: Covariate balance comparing SGLT2is, GLP1-RAs, DPP4is, and SUs new-users with established CVD in VA before/after PS matching.** Covariate balance is measured by standardized mean differences (SMDs) between covariates in target and comparator cohorts. We show SMDs on >40,000 baseline patient characteristics before and after PS matching.

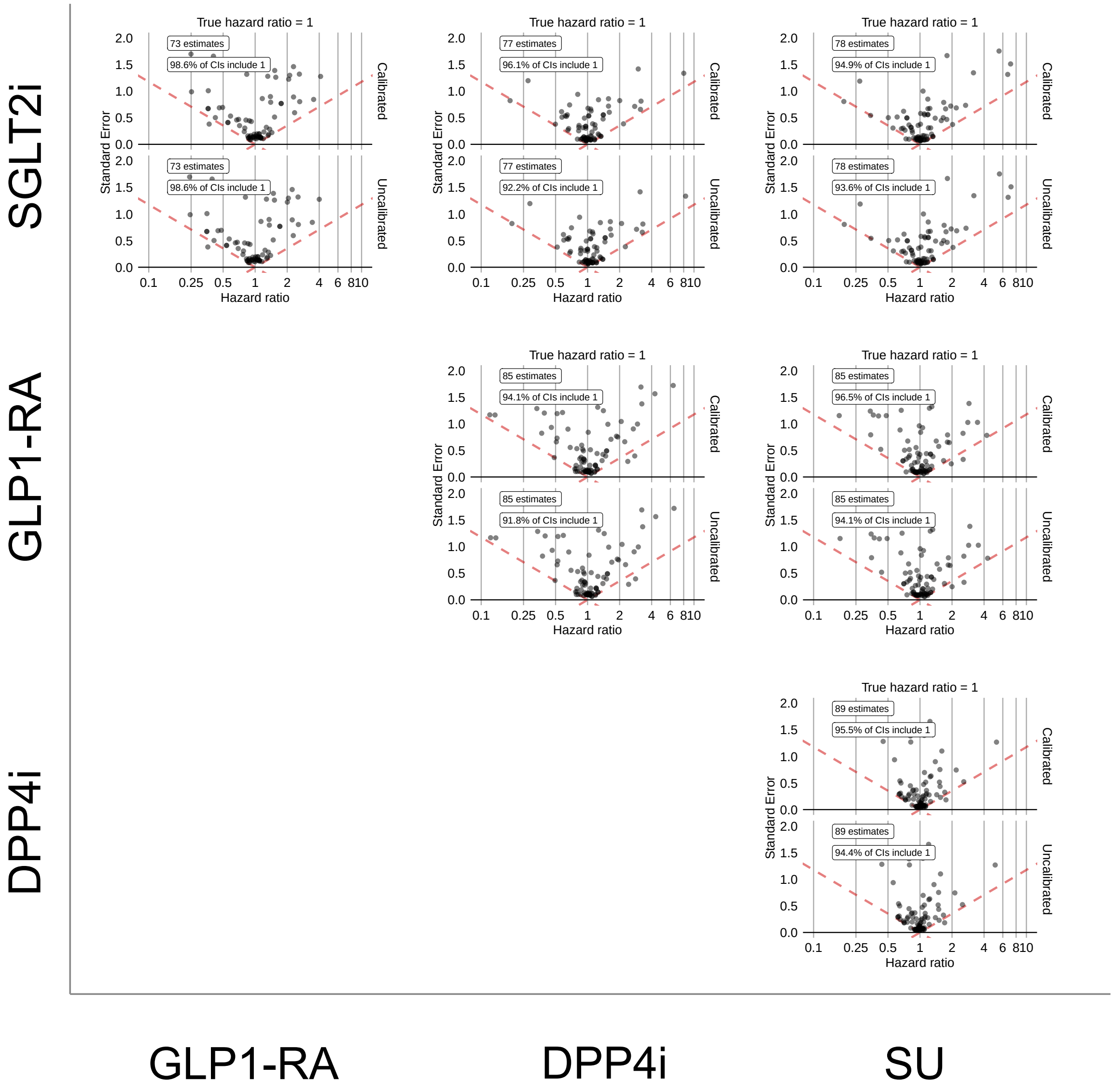


**Supplementary Figure 31: Systematic error distributions comparing SGLT2is, GLP1-RAs, DPP4is, and SUs new-users with established CVD in CCAE using PS stratification.** Systematic error distributions are measured by effect estimates and 95% confidence intervals for negative control outcomes, presented before (bottom) and after (top) empirical calibration. Results are shown using PS stratification.

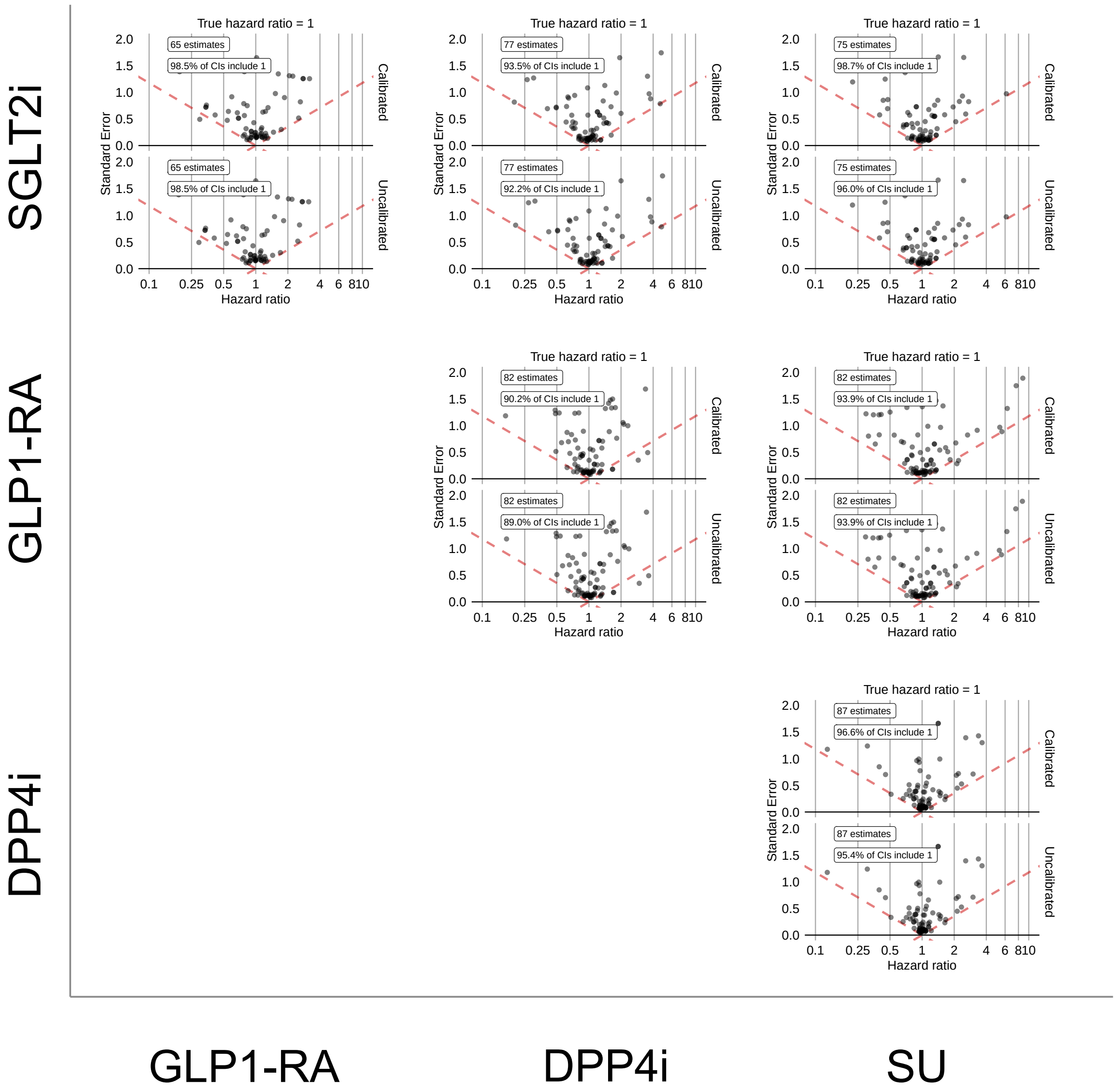


**Supplementary Figure 32: Systematic error distributions comparing SGLT2is, GLP1-RAs, DPP4is, and SUs new-users with established CVD in CCAE using PS matching.** Systematic error distributions are measured by effect estimates and 95% confidence intervals for negative control outcomes, presented before (bottom) and after (top) empirical calibration. Results are shown using PS matching.

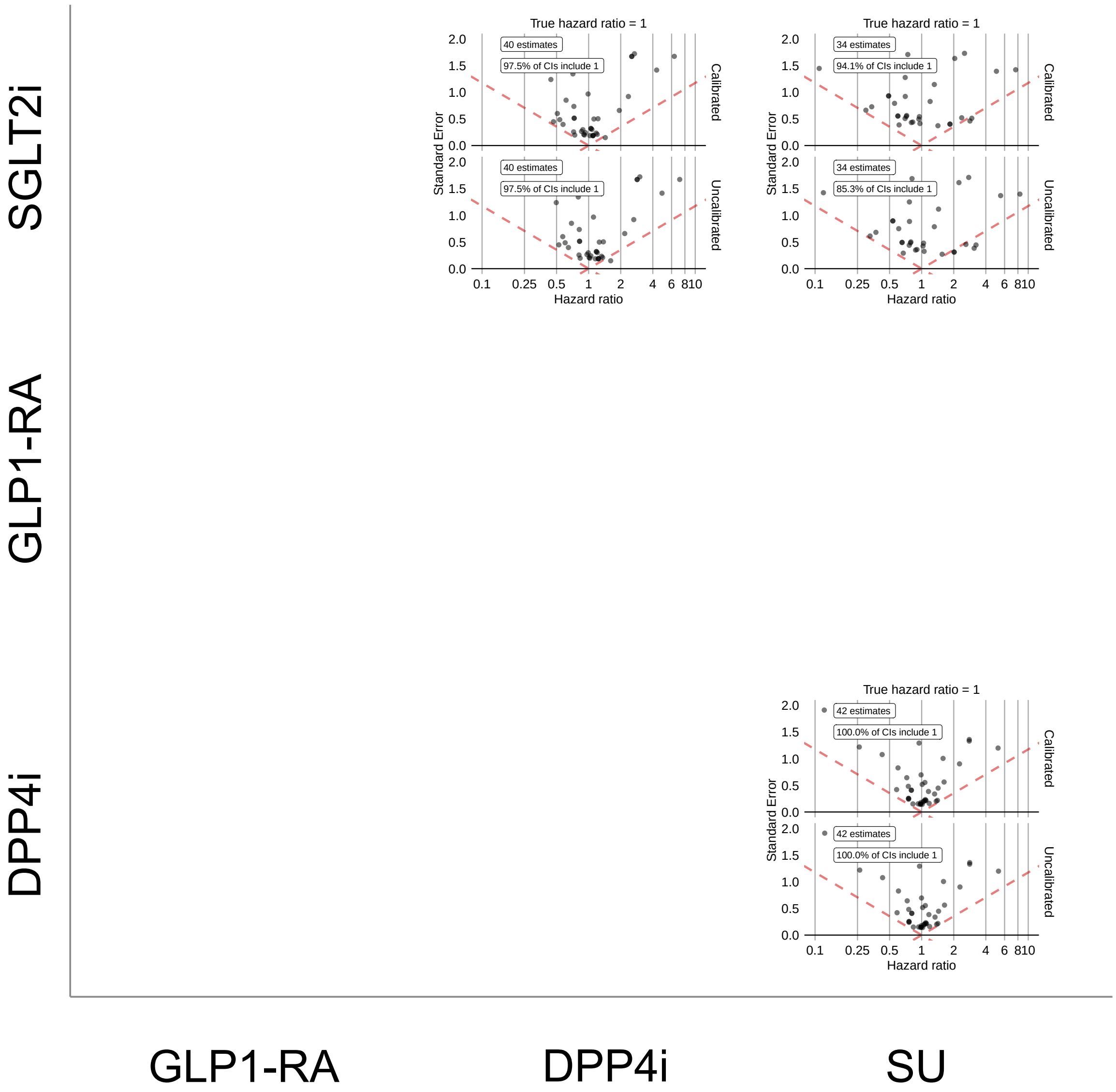


**Supplementary Figure 33: Systematic error distributions comparing SGLT2is, GLP1-RAs, DPP4is, and SUs new-users with established CVD in GDA using PS stratification.** Systematic error distributions are measured by effect estimates and 95% confidence intervals for negative control outcomes, presented before (bottom) and after (top) empirical calibration. Results are shown using PS stratification.

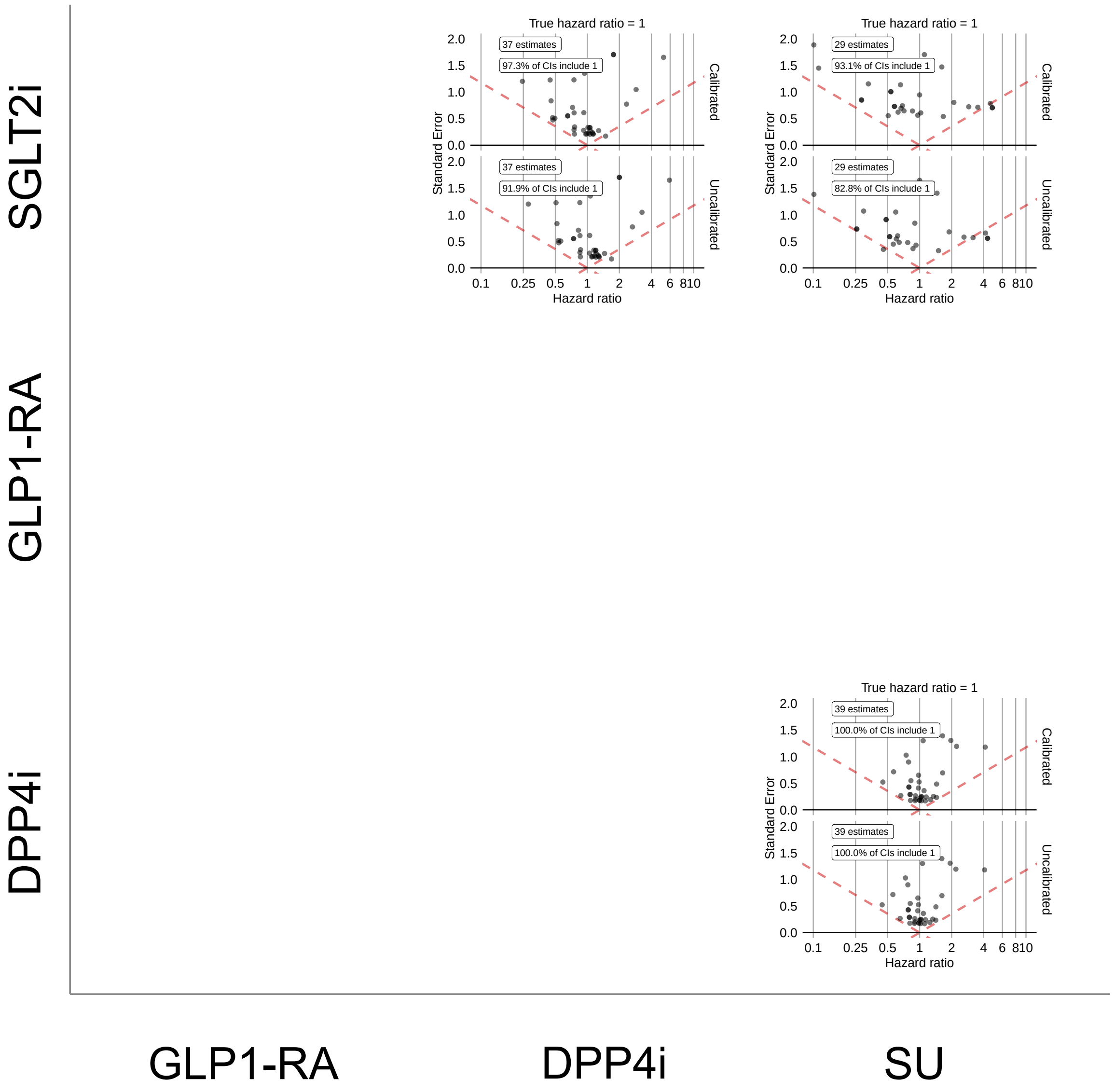


**Supplementary Figure 34: Systematic error distributions comparing SGLT2is, GLP1-RAs, DPP4is, and SUs new-users with established CVD in GDA using PS matching.** Systematic error distributions are measured by effect estimates and 95% confidence intervals for negative control outcomes, presented before (bottom) and after (top) empirical calibration. Results are shown using PS matching.

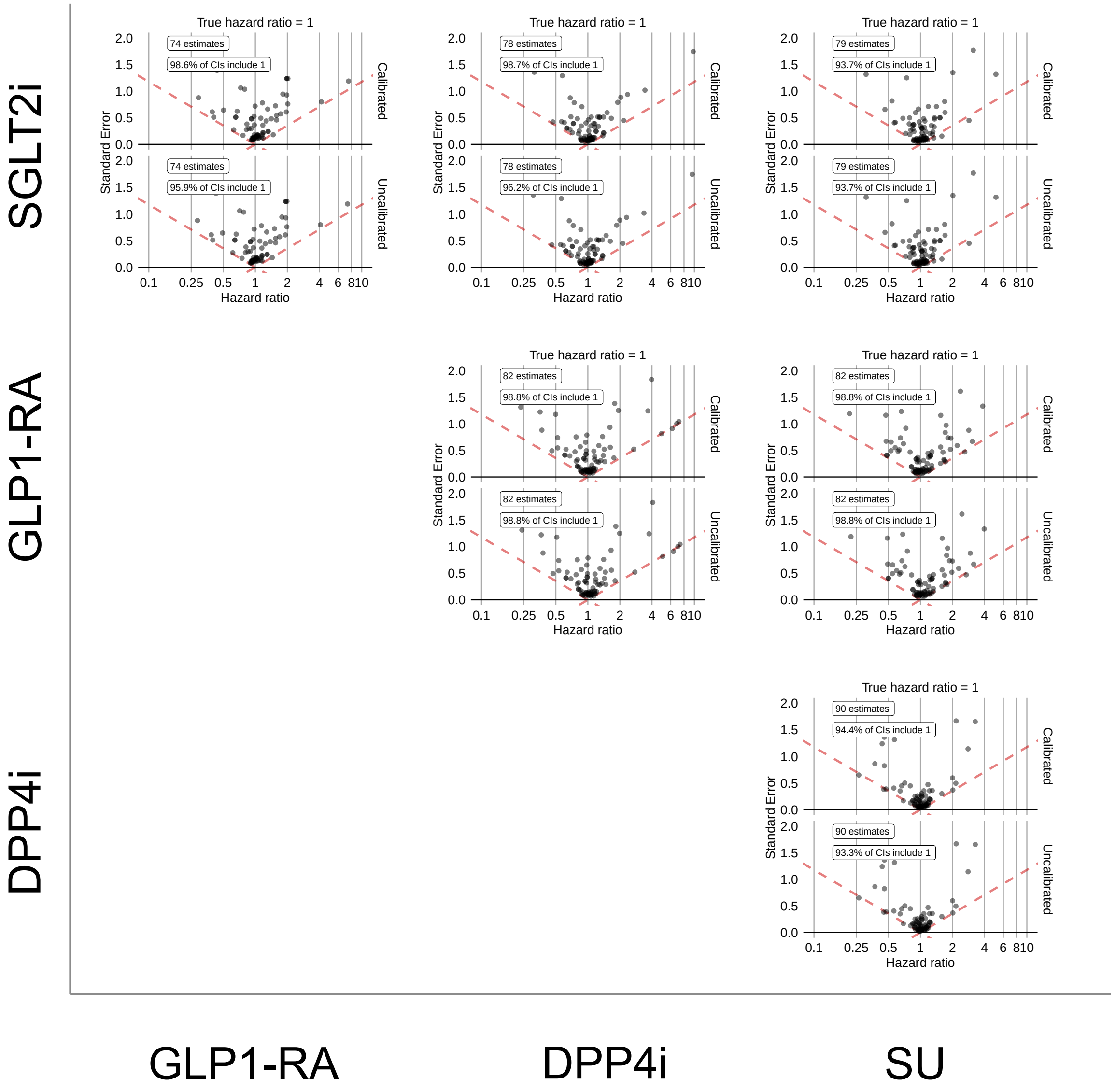


**Supplementary Figure 35: Systematic error distributions comparing SGLT2is, GLP1-RAs, DPP4is, and SUs new-users with established CVD in OptumDOD using PS stratification.** Systematic error distributions are measured by effect estimates and 95% confidence intervals for negative control outcomes, presented before (bottom) and after (top) empirical calibration. Results are shown using PS stratification.

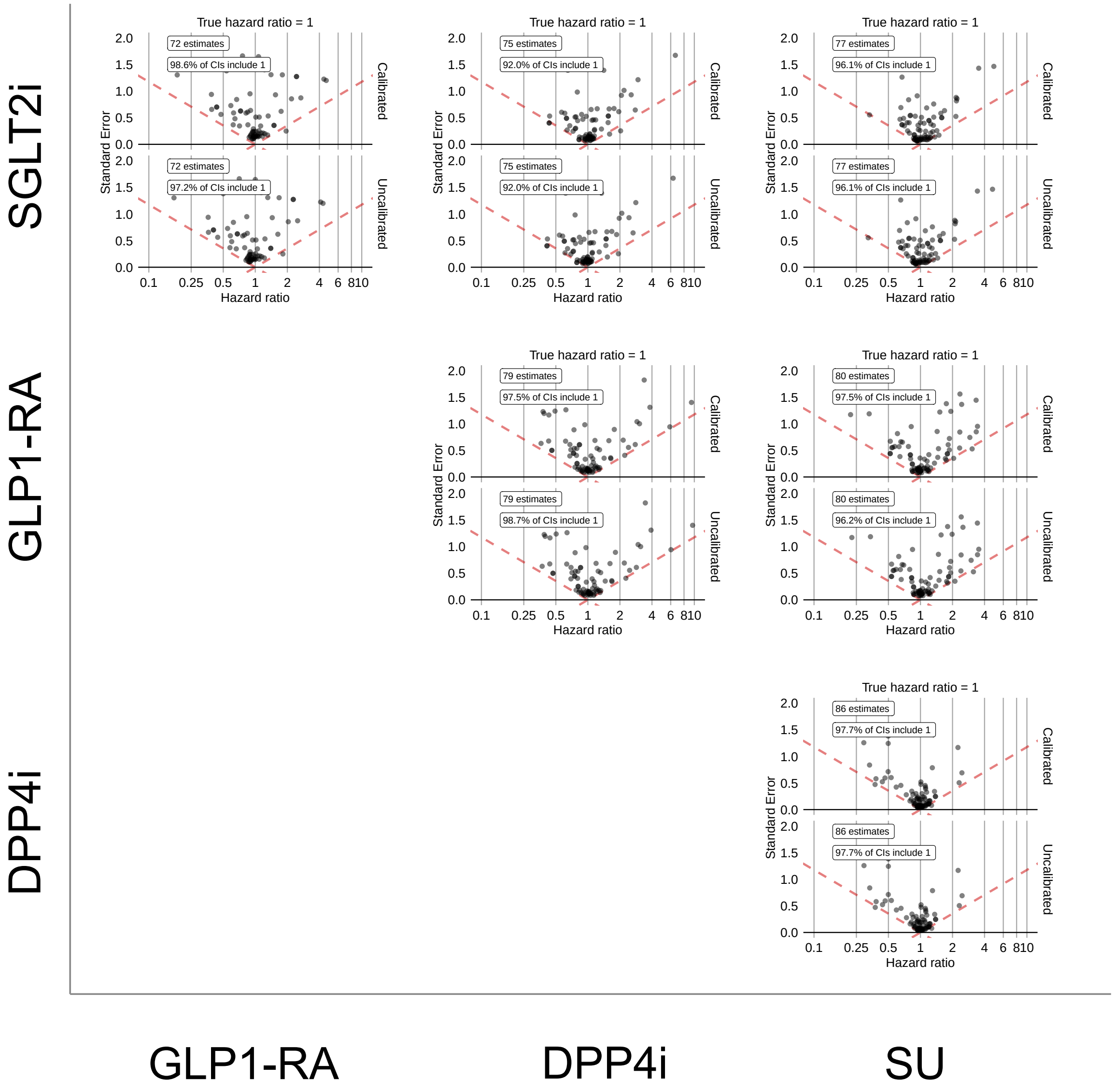


**Supplementary Figure 36: Systematic error distributions comparing SGLT2is, GLP1-RAs, DPP4is, and SUs new-users with established CVD in OptumDOD using PS matching.** Systematic error distributions are measured by effect estimates and 95% confidence intervals for negative control outcomes, presented before (bottom) and after (top) empirical calibration. Results are shown using PS matching.

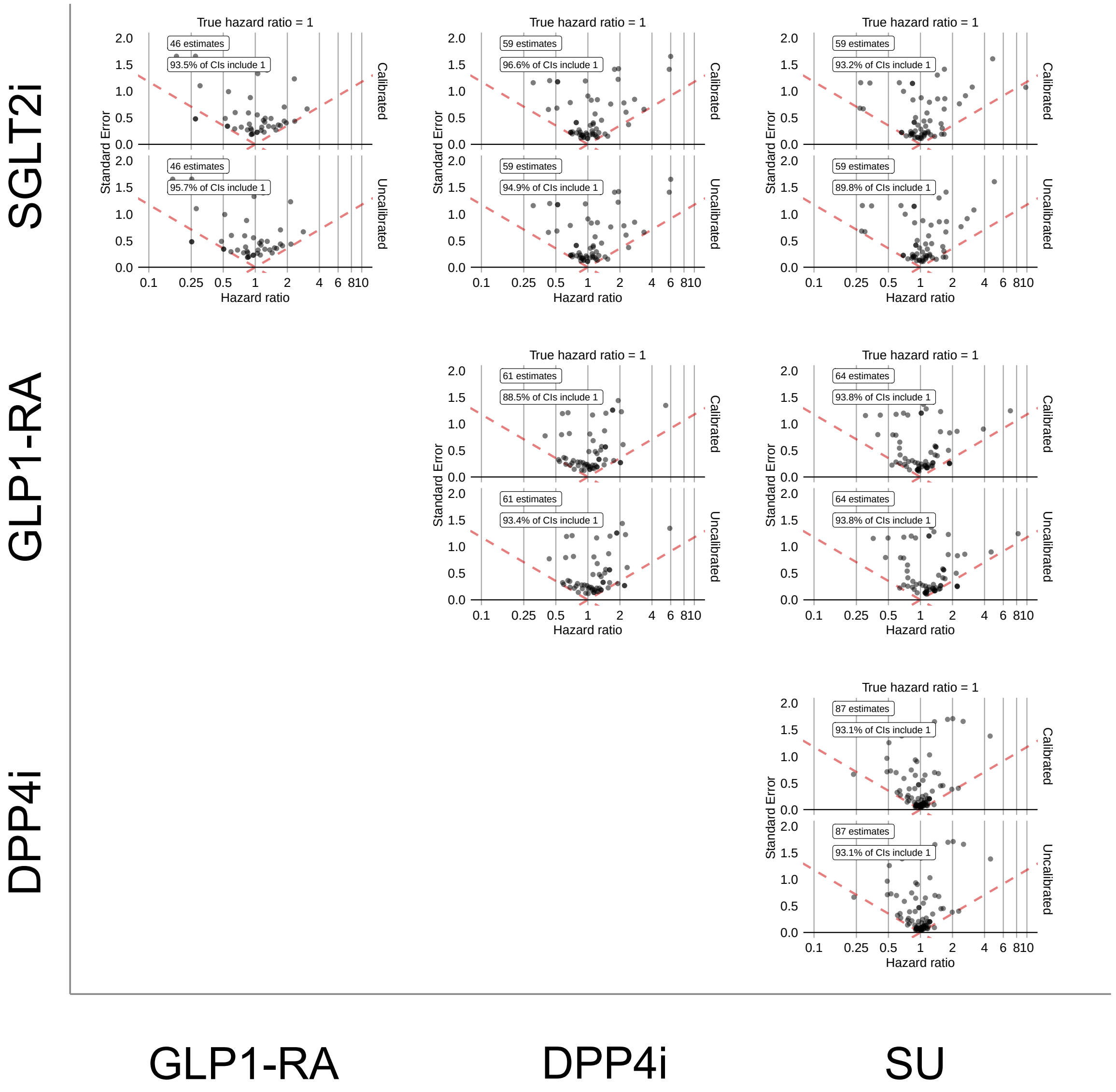


**Supplementary Figure 37: Systematic error distributions comparing SGLT2is, GLP1-RAs, DPP4is, and SUs new-users with established CVD in MDCR using PS stratification.** Systematic error distributions are measured by effect estimates and 95% confidence intervals for negative control outcomes, presented before (bottom) and after (top) empirical calibration. Results are shown using PS stratification.

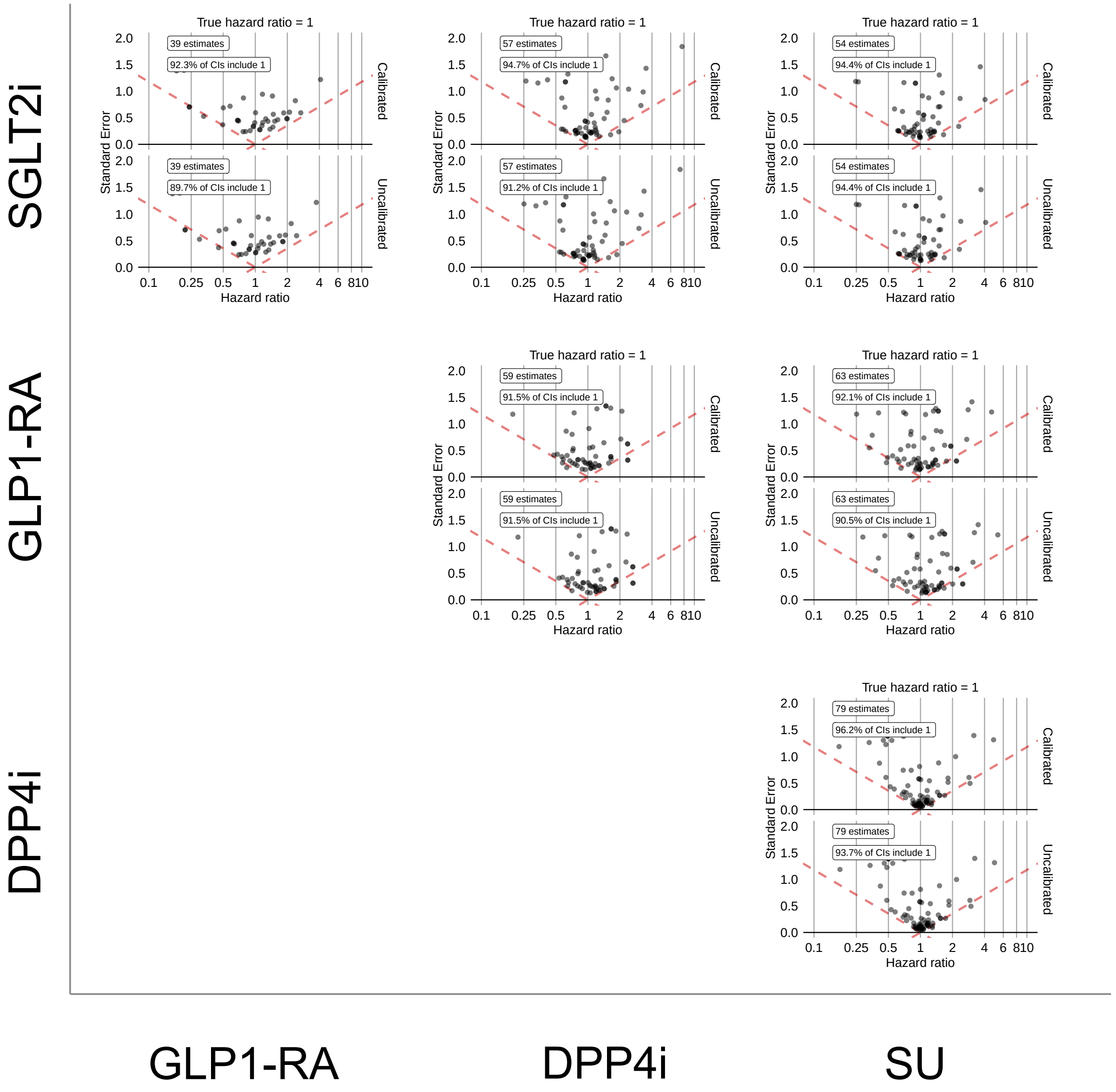


**Supplementary Figure 38: Systematic error distributions comparing SGLT2is, GLP1-RAs, DPP4is, and SUs new-users with established CVD in MDCR using PS matching.** Systematic error distributions are measured by effect estimates and 95% confidence intervals for negative control outcomes, presented before (bottom) and after (top) empirical calibration. Results are shown using PS matching.

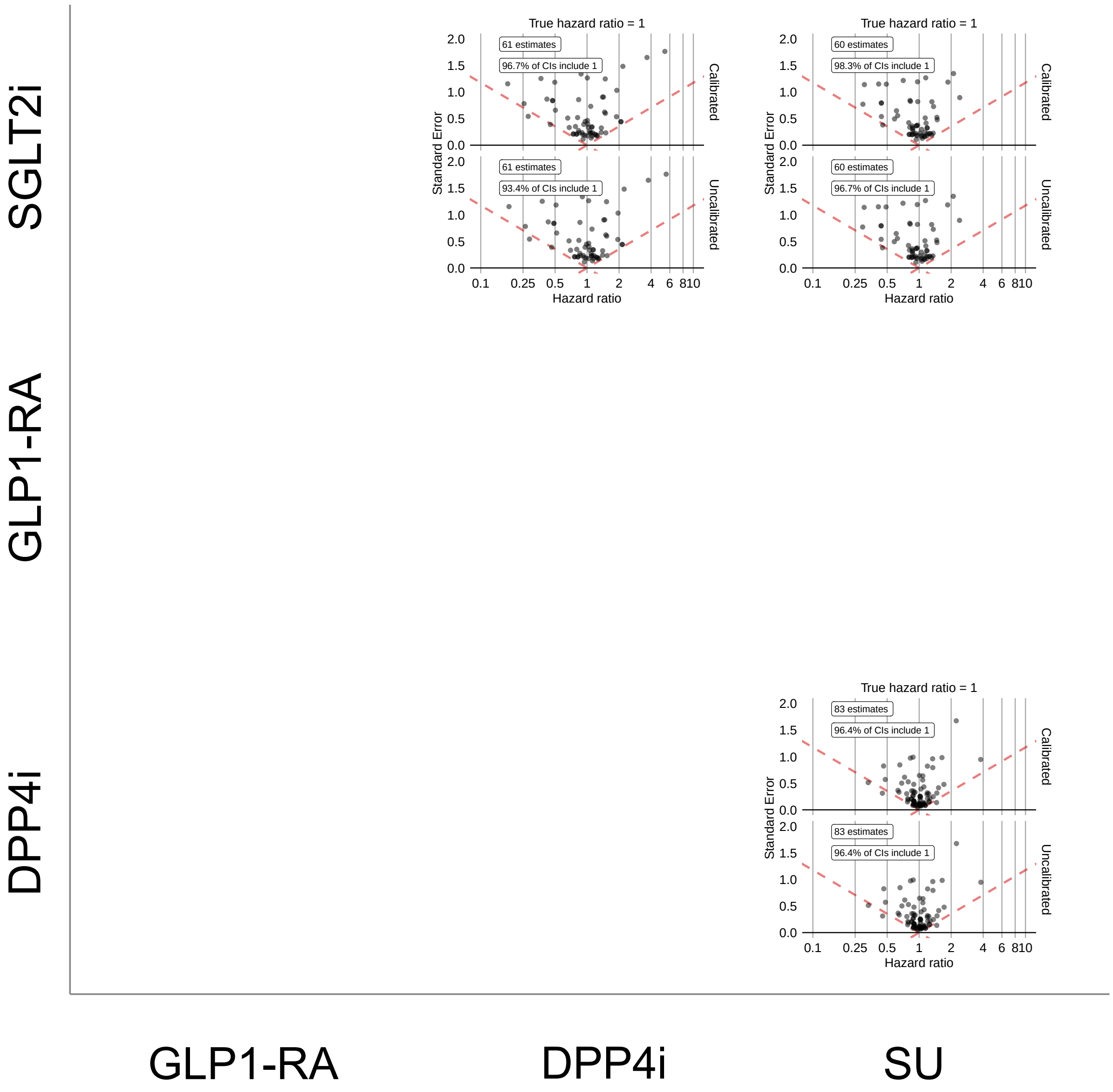


**Supplementary Figure 39: Systematic error distributions comparing SGLT2is, GLP1-RAs, DPP4is, and SUs new-users with established CVD in MDCD using PS stratification.** Systematic error distributions are measured by effect estimates and 95% confidence intervals for negative control outcomes, presented before (bottom) and after (top) empirical calibration. Results are shown using PS stratification.

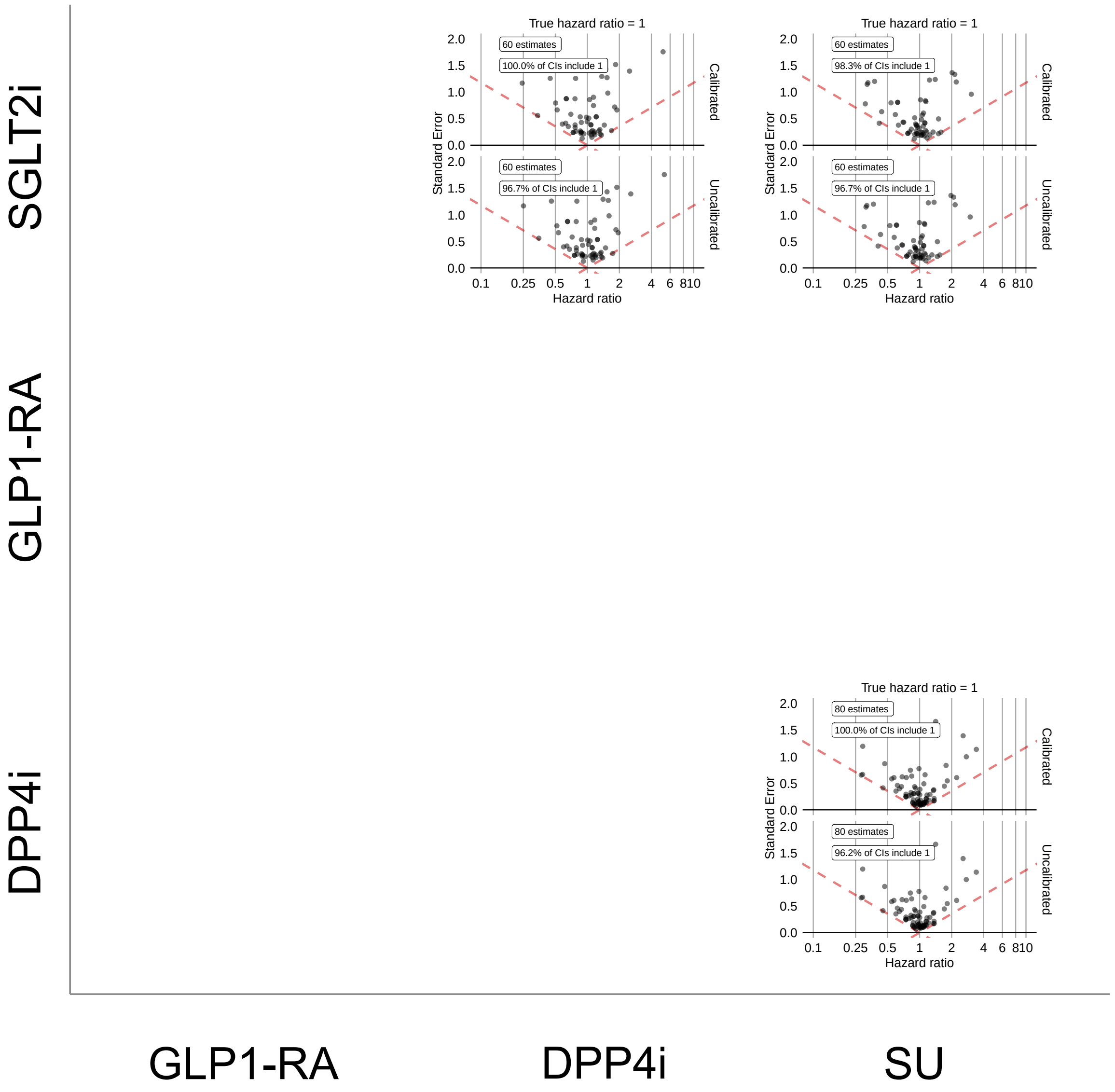


**Supplementary Figure 40: Systematic error distributions comparing SGLT2is, GLP1-RAs, DPP4is, and SUs new-users with established CVD in MDCD using PS matching.** Systematic error distributions are measured by effect estimates and 95% confidence intervals for negative control outcomes, presented before (bottom) and after (top) empirical calibration. Results are shown using PS matching.

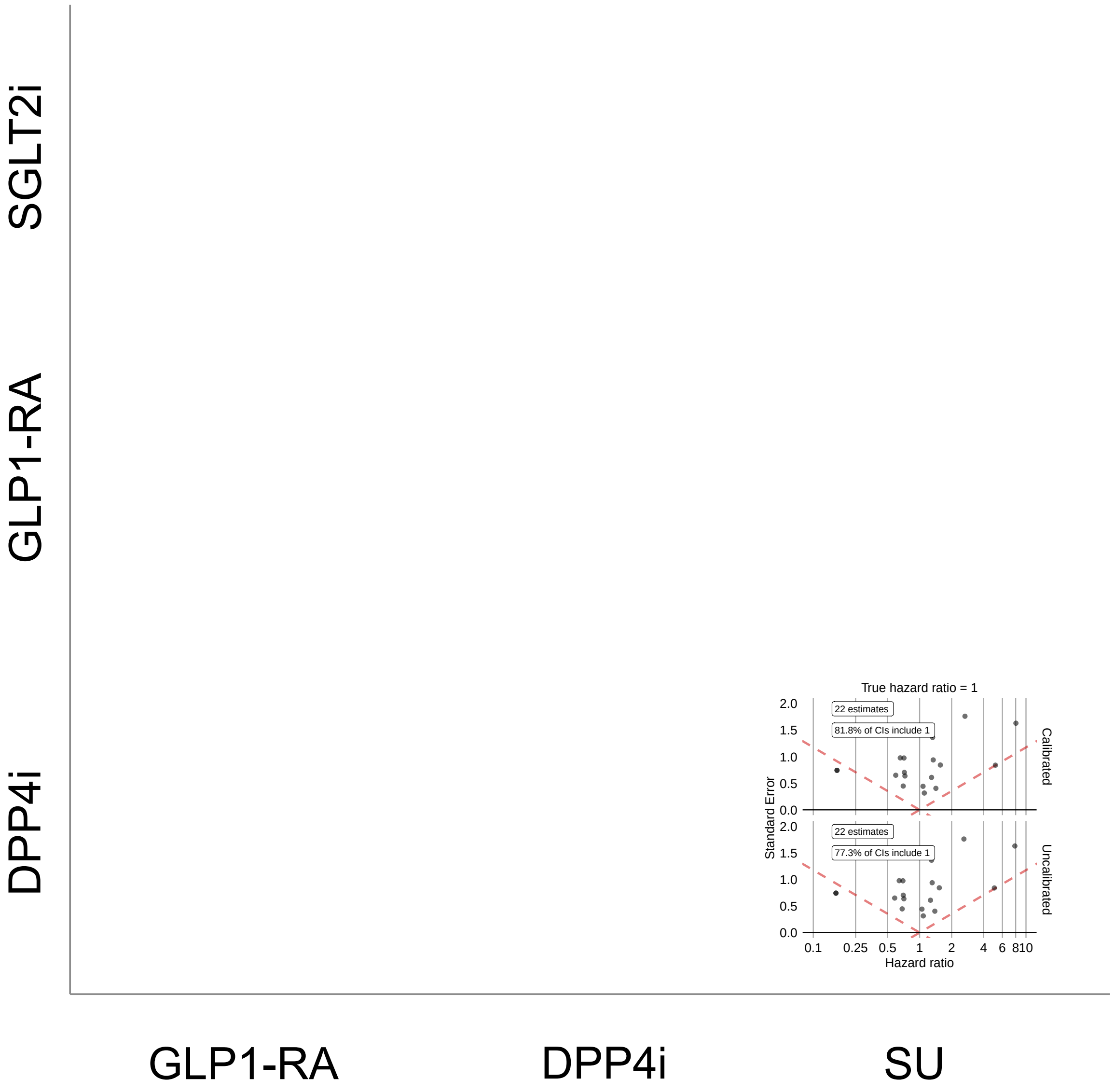


**Supplementary Figure 41: Systematic error distributions comparing SGLT2is, GLP1-RAs, DPP4is, and SUs new-users with established CVD in IMRD using PS stratification.** Systematic error distributions are measured by effect estimates and 95% confidence intervals for negative control outcomes, presented before (bottom) and after (top) empirical calibration. Results are shown using PS stratification.

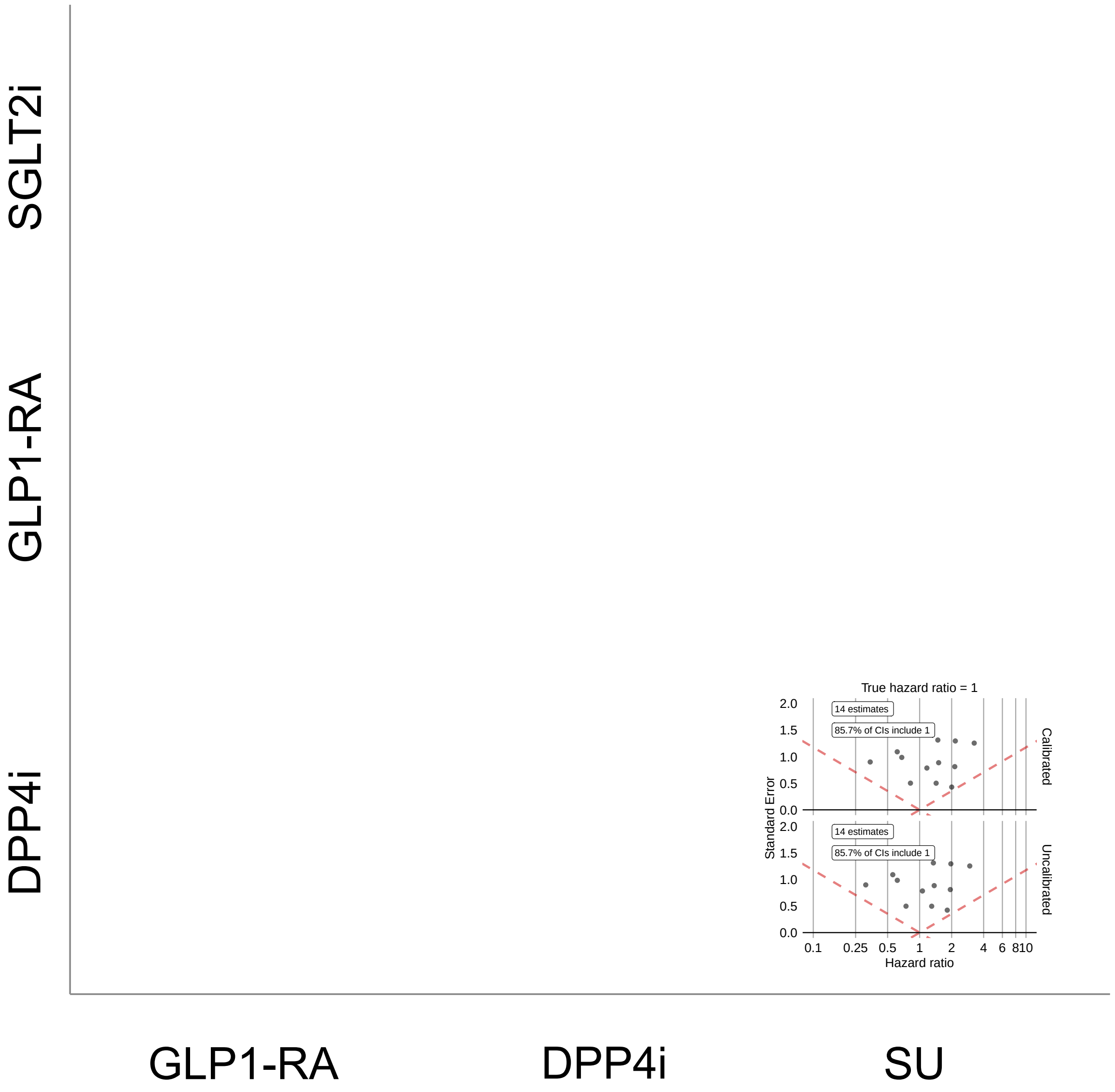


**Supplementary Figure 42: Systematic error distributions comparing SGLT2is, GLP1-RAs, DPP4is, and SUs new-users with established CVD in IMRD using PS matching.** Systematic error distributions are measured by effect estimates and 95% confidence intervals for negative control outcomes, presented before (bottom) and after (top) empirical calibration. Results are shown using PS matching.

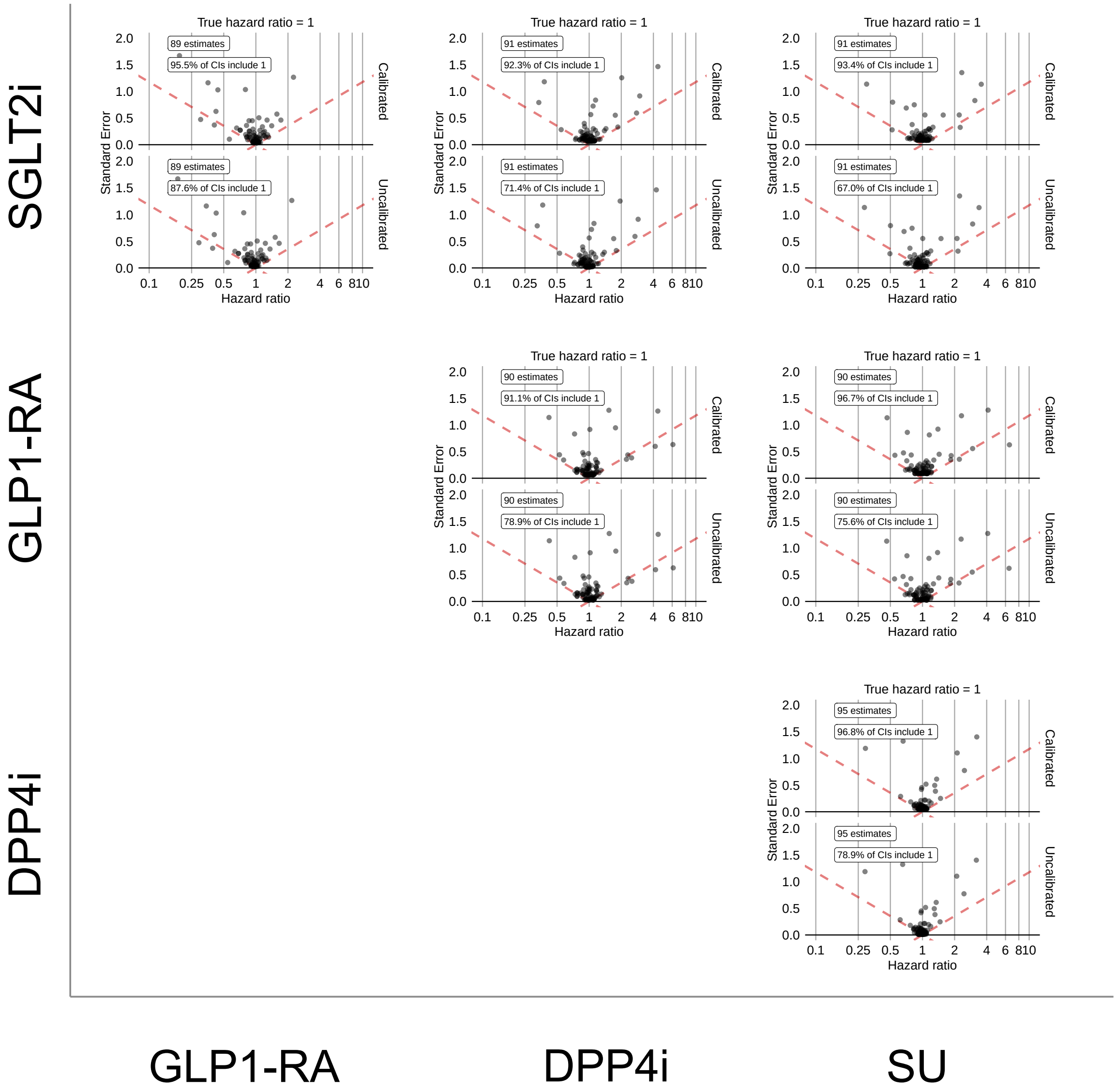


**Supplementary Figure 43: Systematic error distributions comparing SGLT2is, GLP1-RAs, DPP4is, and SUs new-users with established CVD in US Open Claims using PS stratification.** Systematic error distributions are measured by effect estimates and 95% confidence intervals for negative control outcomes, presented before (bottom) and after (top) empirical calibration. Results are shown using PS stratification.

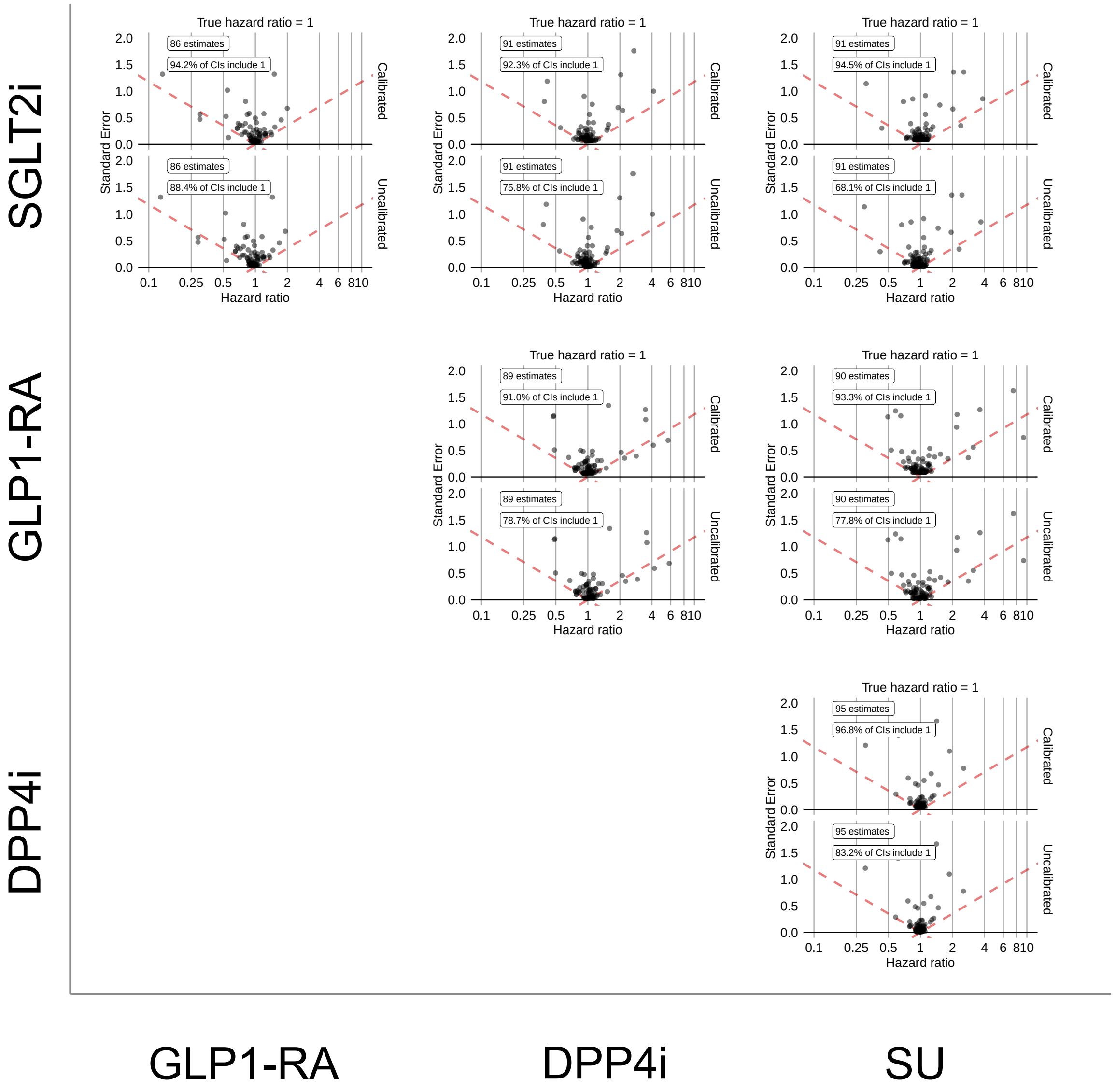


**Supplementary Figure 44: Systematic error distributions comparing SGLT2is, GLP1-RAs, DPP4is, and SUs new-users with established CVD in US Open Claims using PS matching.** Systematic error distributions are measured by effect estimates and 95% confidence intervals for negative control outcomes, presented before (bottom) and after (top) empirical calibration. Results are shown using PS matching.

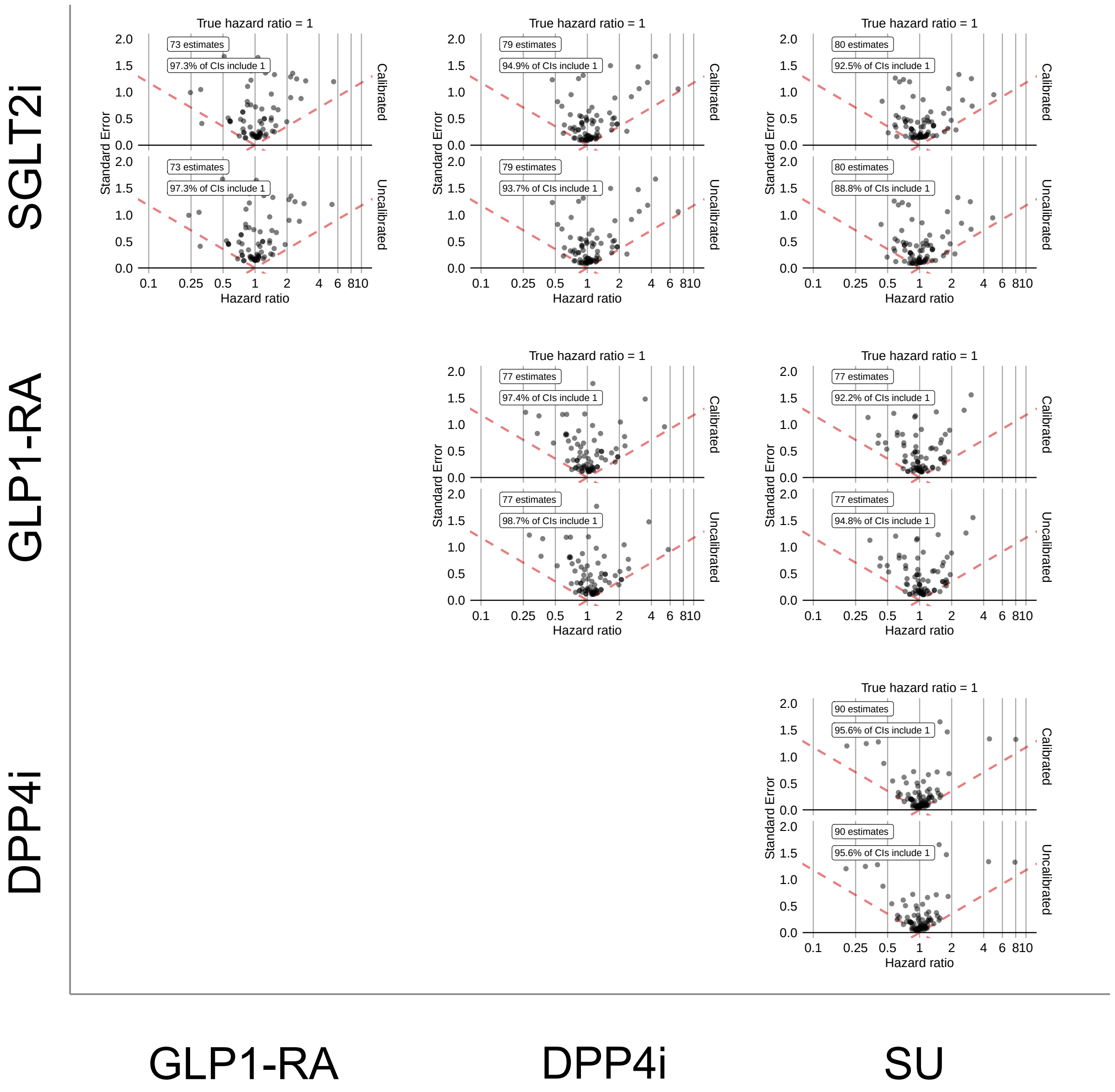


**Supplementary Figure 45: Systematic error distributions comparing SGLT2is, GLP1-RAs, DPP4is, and SUs new-users with established CVD in OptumEHR using PS stratification.** Systematic error distributions are measured by effect estimates and 95% confidence intervals for negative control outcomes, presented before (bottom) and after (top) empirical calibration. Results are shown using PS stratification.

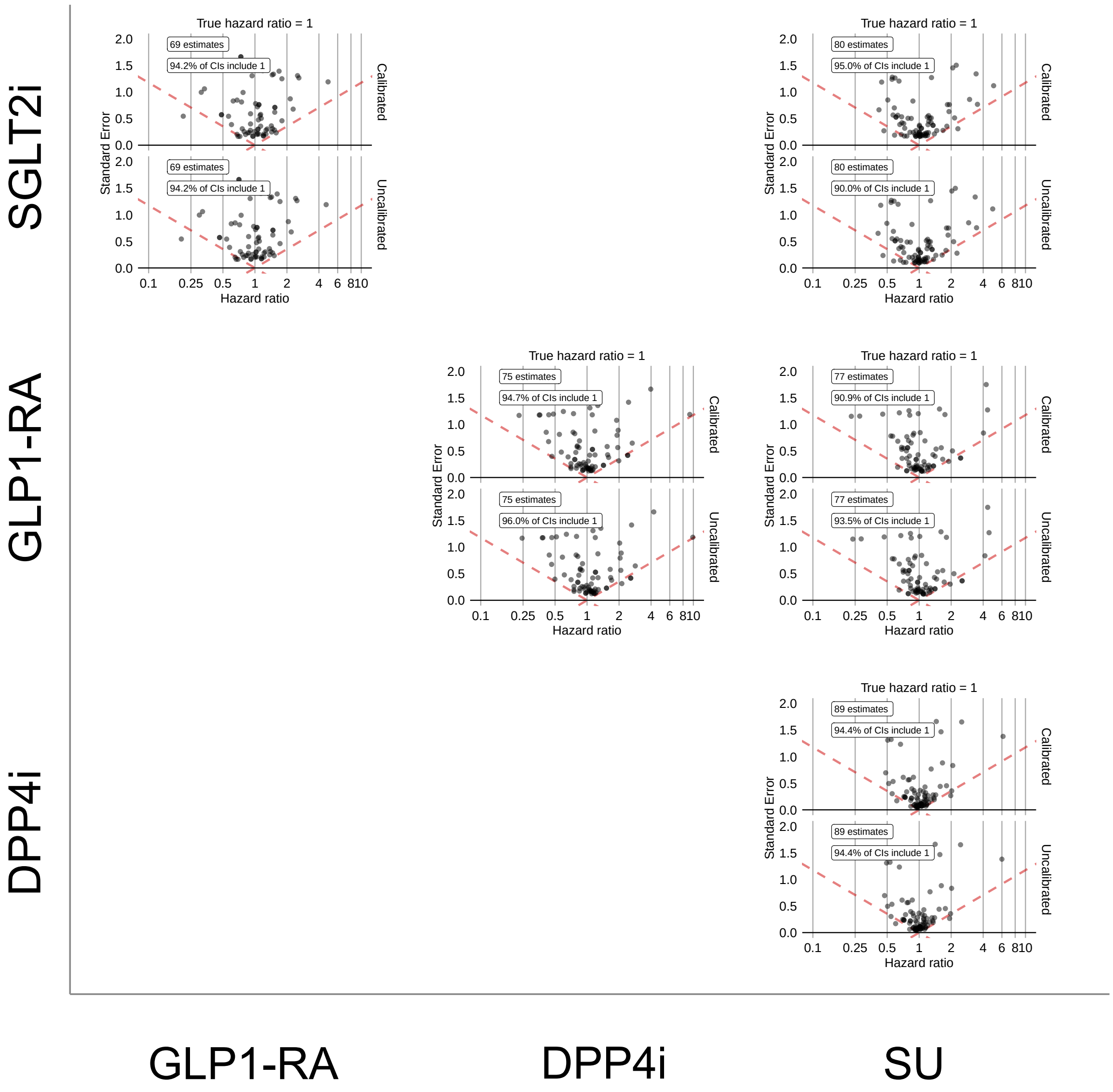


**Supplementary Figure 46: Systematic error distributions comparing SGLT2is, GLP1-RAs, DPP4is, and SUs new-users with established CVD in OptumEHR using PS matching.** Systematic error distributions are measured by effect estimates and 95% confidence intervals for negative control outcomes, presented before (bottom) and after (top) empirical calibration. Results are shown using PS matching.

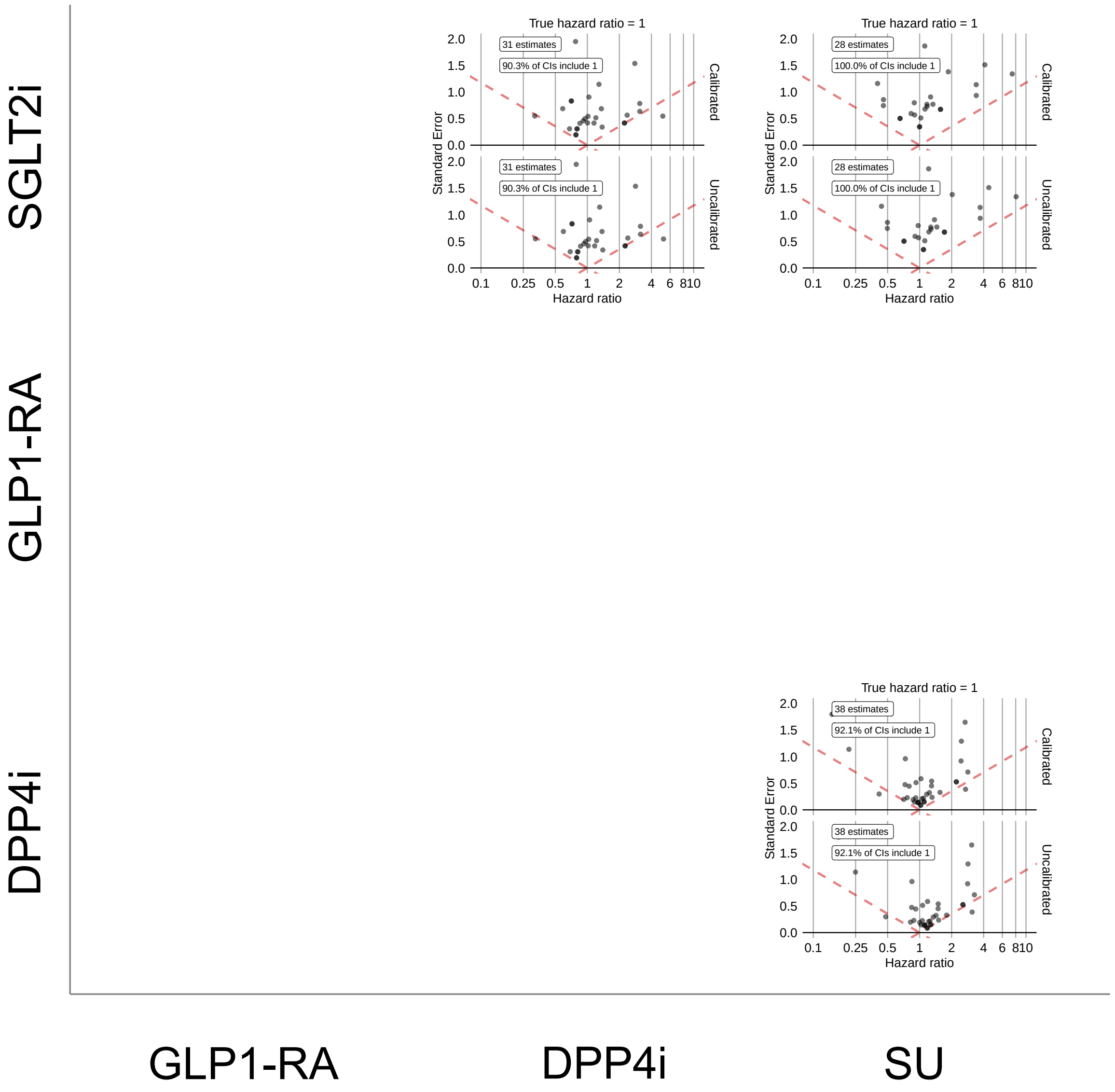


**Supplementary Figure 47: Systematic error distributions comparing SGLT2is, GLP1-RAs, DPP4is, and SUs new-users with established CVD in SIDIAP using PS stratification.** Systematic error distributions are measured by effect estimates and 95% confidence intervals for negative control outcomes, presented before (bottom) and after (top) empirical calibration. Results are shown using PS stratification.

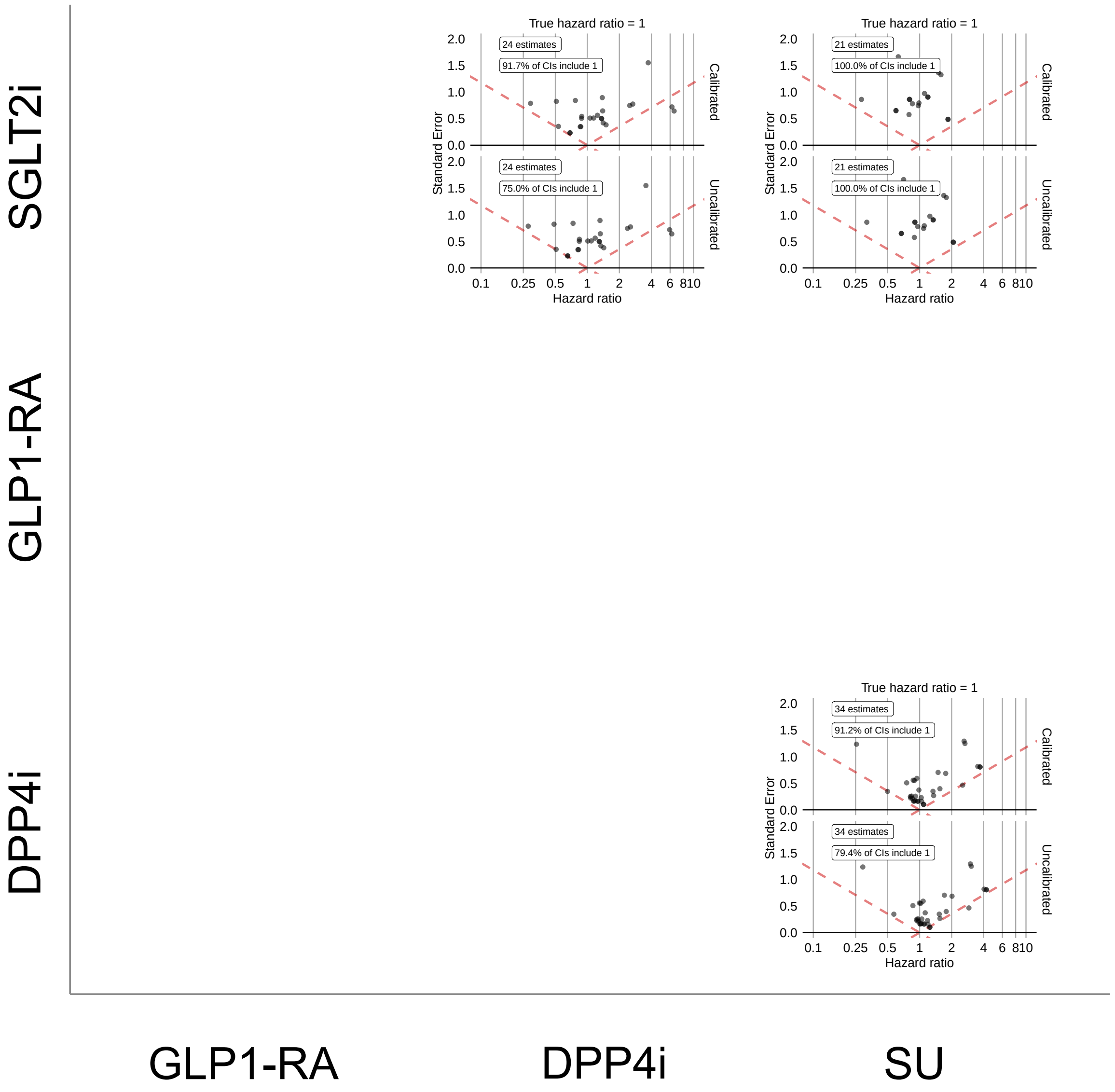


**Supplementary Figure 48: Systematic error distributions comparing SGLT2is, GLP1-RAs, DPP4is, and SUs new-users with established CVD in SIDIAP using PS matching.** Systematic error distributions are measured by effect estimates and 95% confidence intervals for negative control outcomes, presented before (bottom) and after (top) empirical calibration. Results are shown using PS matching.

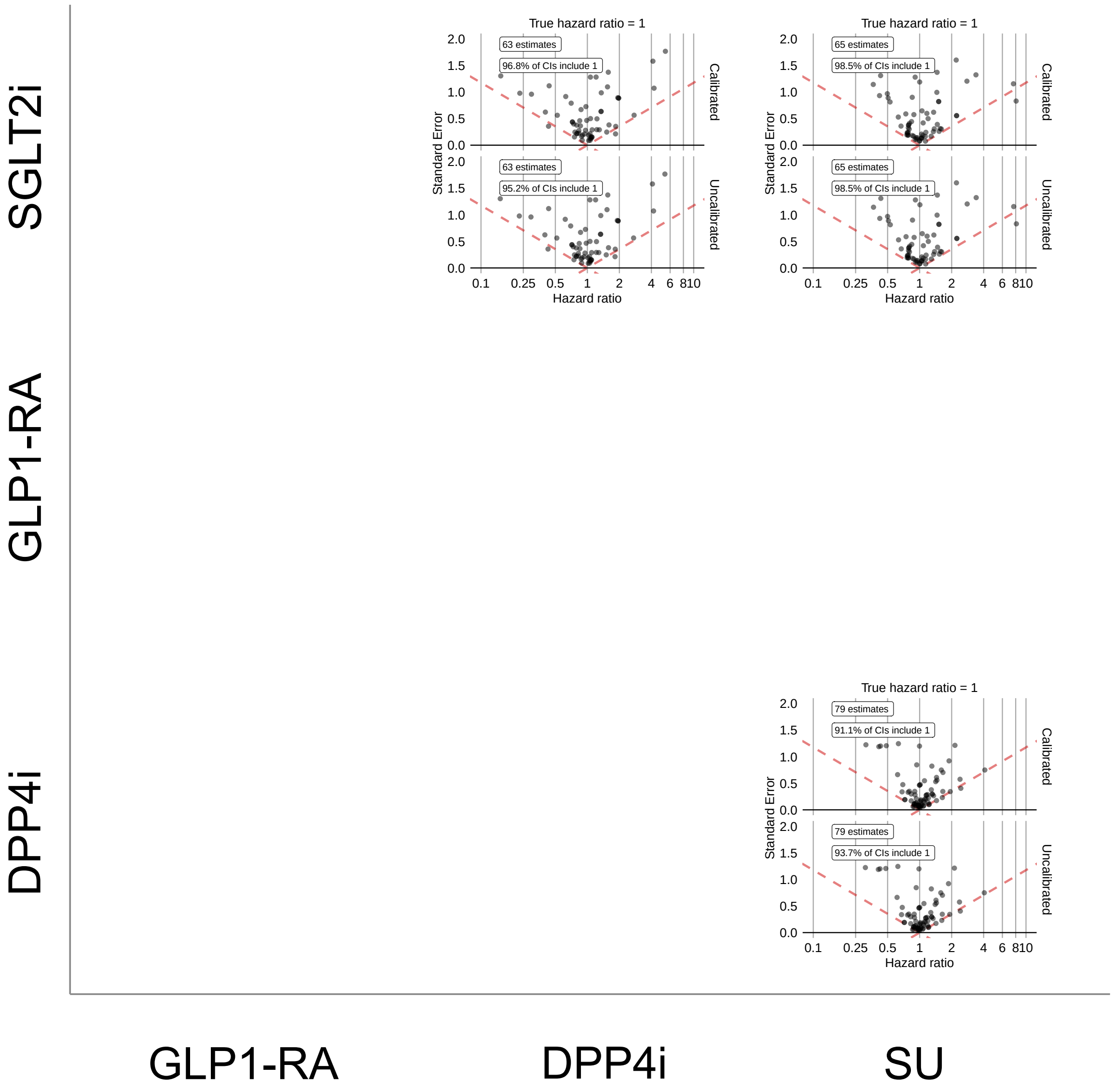


**Supplementary Figure 49: Systematic error distributions comparing SGLT2is, GLP1-RAs, DPP4is, and SUs new-users with established CVD in VA using PS stratification.** Systematic error distributions are measured by effect estimates and 95% confidence intervals for negative control outcomes, presented before (bottom) and after (top) empirical calibration. Results are shown using PS stratification.

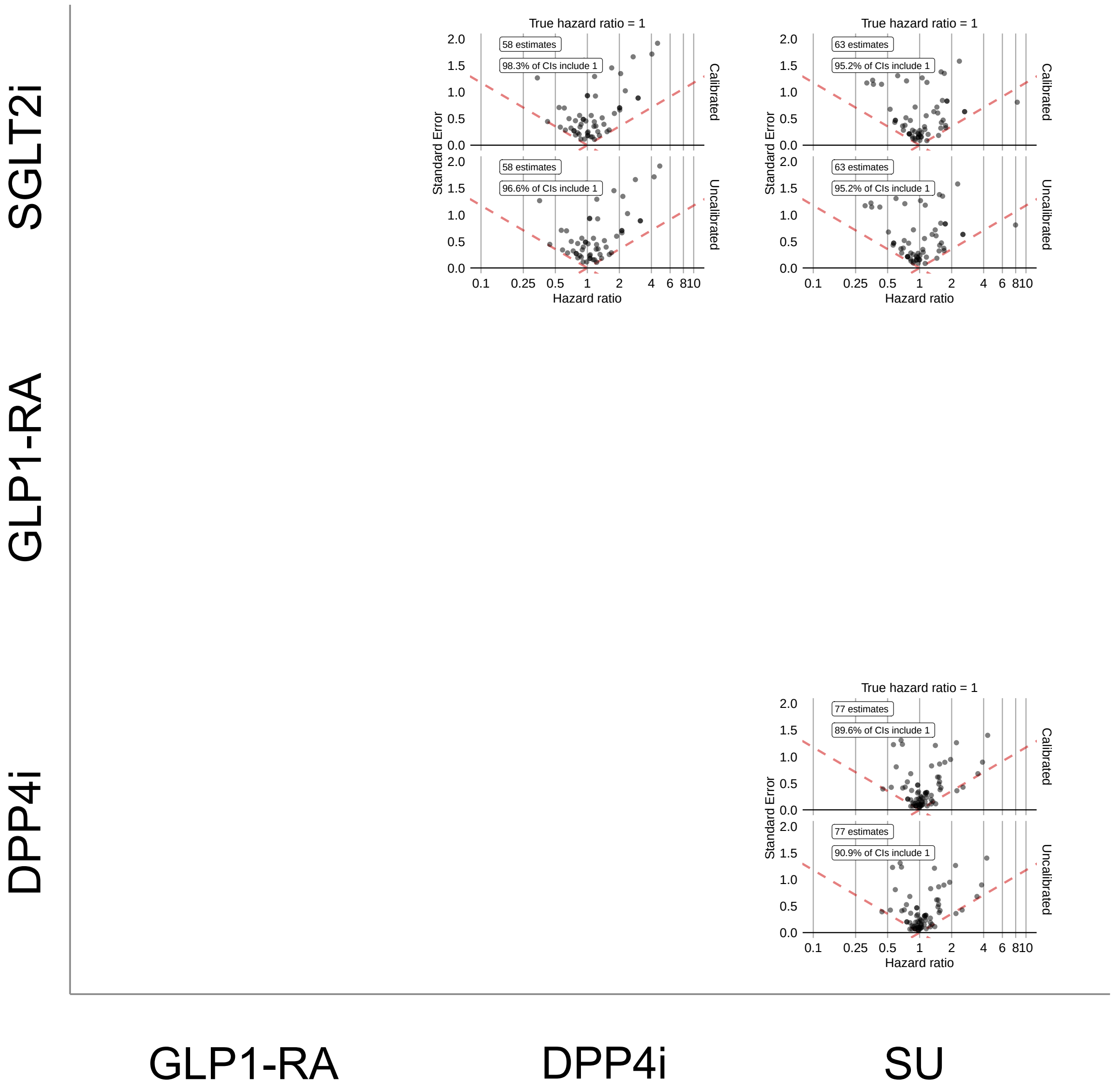


**Supplementary Figure 50: Systematic error distributions comparing SGLT2is, GLP1-RAs, DPP4is, and SUs new-users with established CVD in VA using PS matching.** Systematic error distributions are measured by effect estimates and 95% confidence intervals for negative control outcomes, presented before (bottom) and after (top) empirical calibration. Results are shown using PS matching.

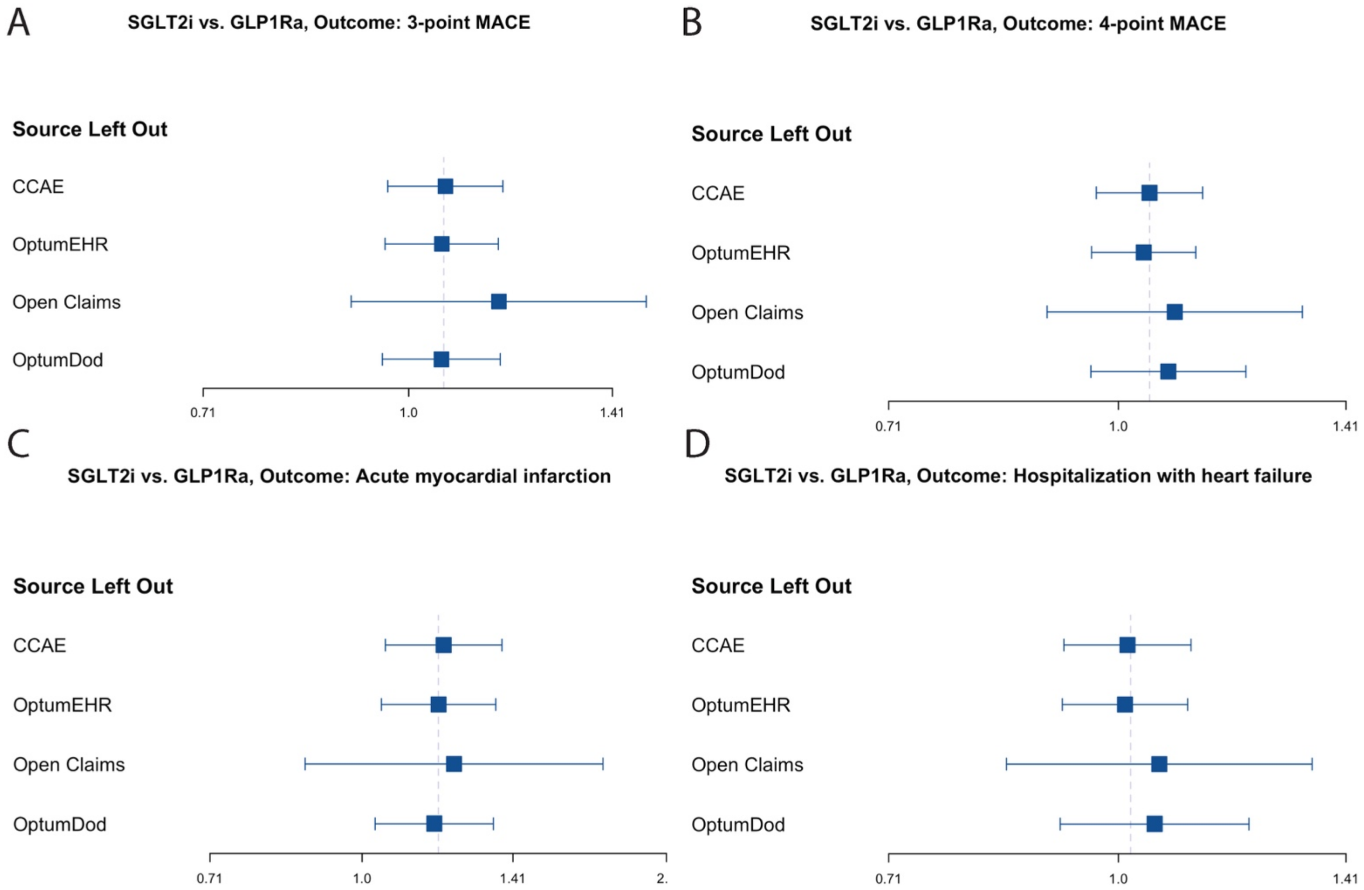


**Supplementary Figure 51: Leave-one-out meta-analysis of calibrated hazard ratio estimates of cardiovascular outcomes for SGLT2i vs GLP1-RA comparison across databases.**

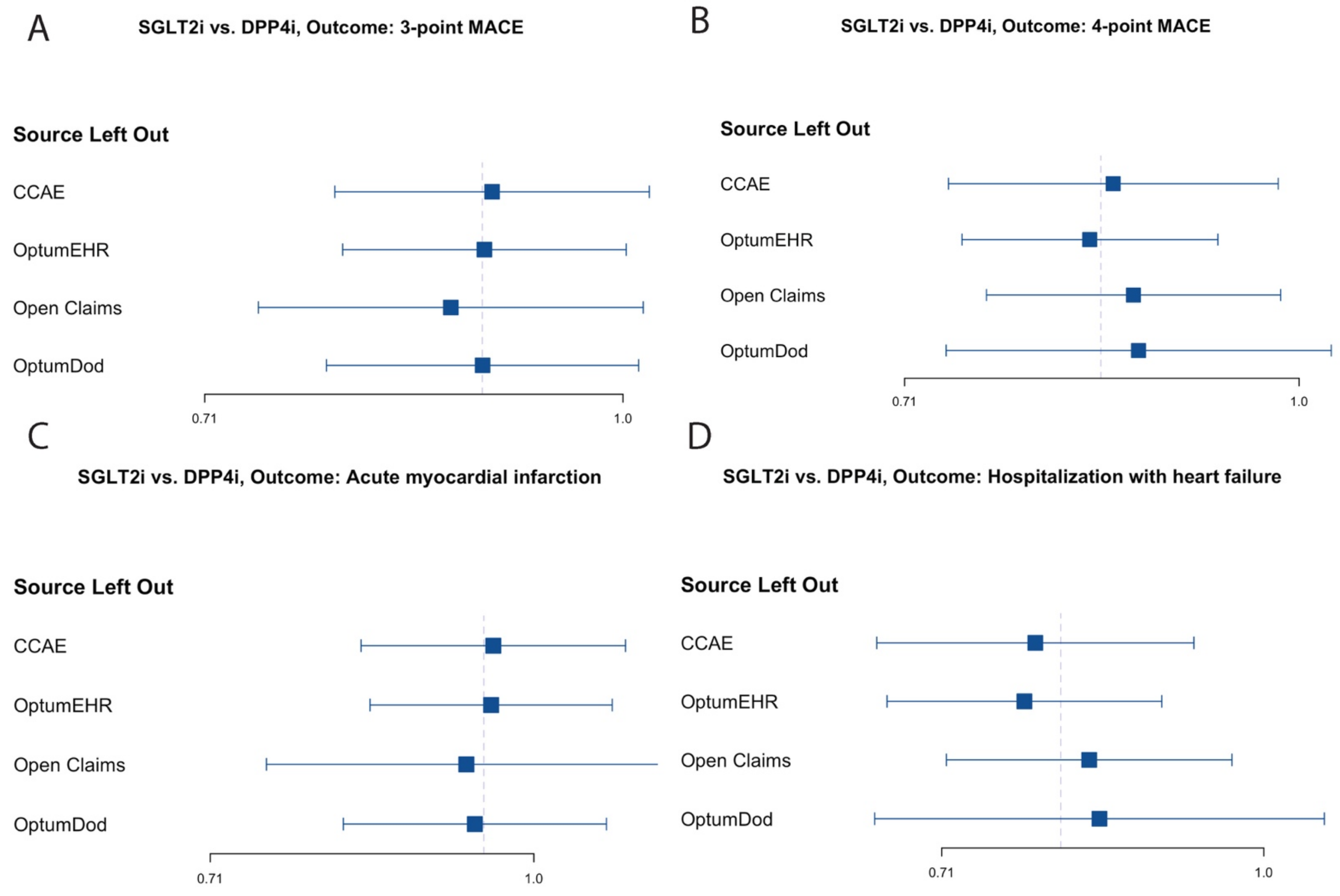


**Supplementary Figure 52: Leave-one-out meta-analysis of calibrated hazard ratio estimates of cardiovascular outcomes for SGLT2i vs DPP4i comparison across databases.**

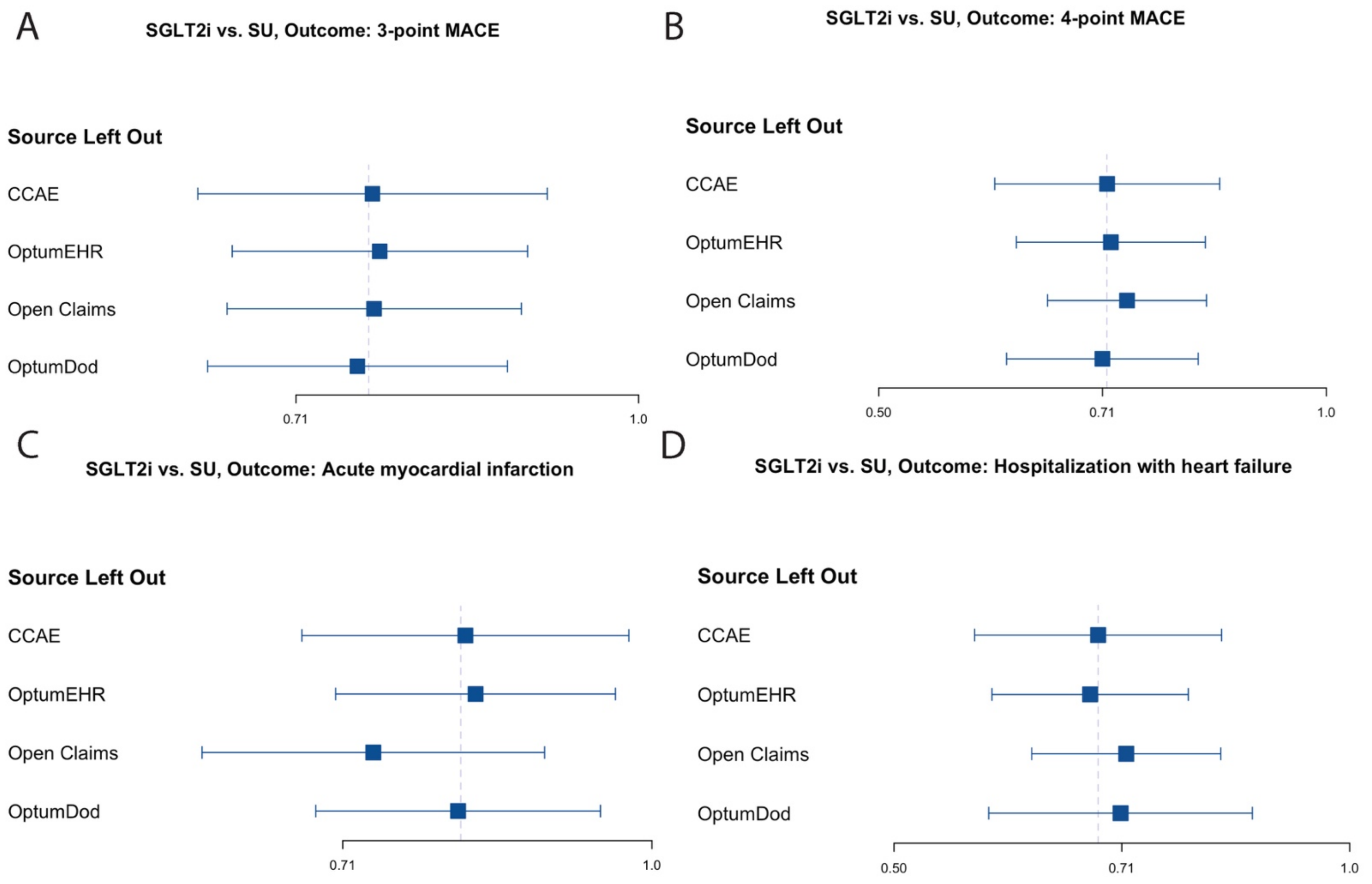


**Supplementary Figure 53: Leave-one-out meta-analysis of calibrated hazard ratio estimates of cardiovascular outcomes for SGLT2i vs SU comparison across databases.**

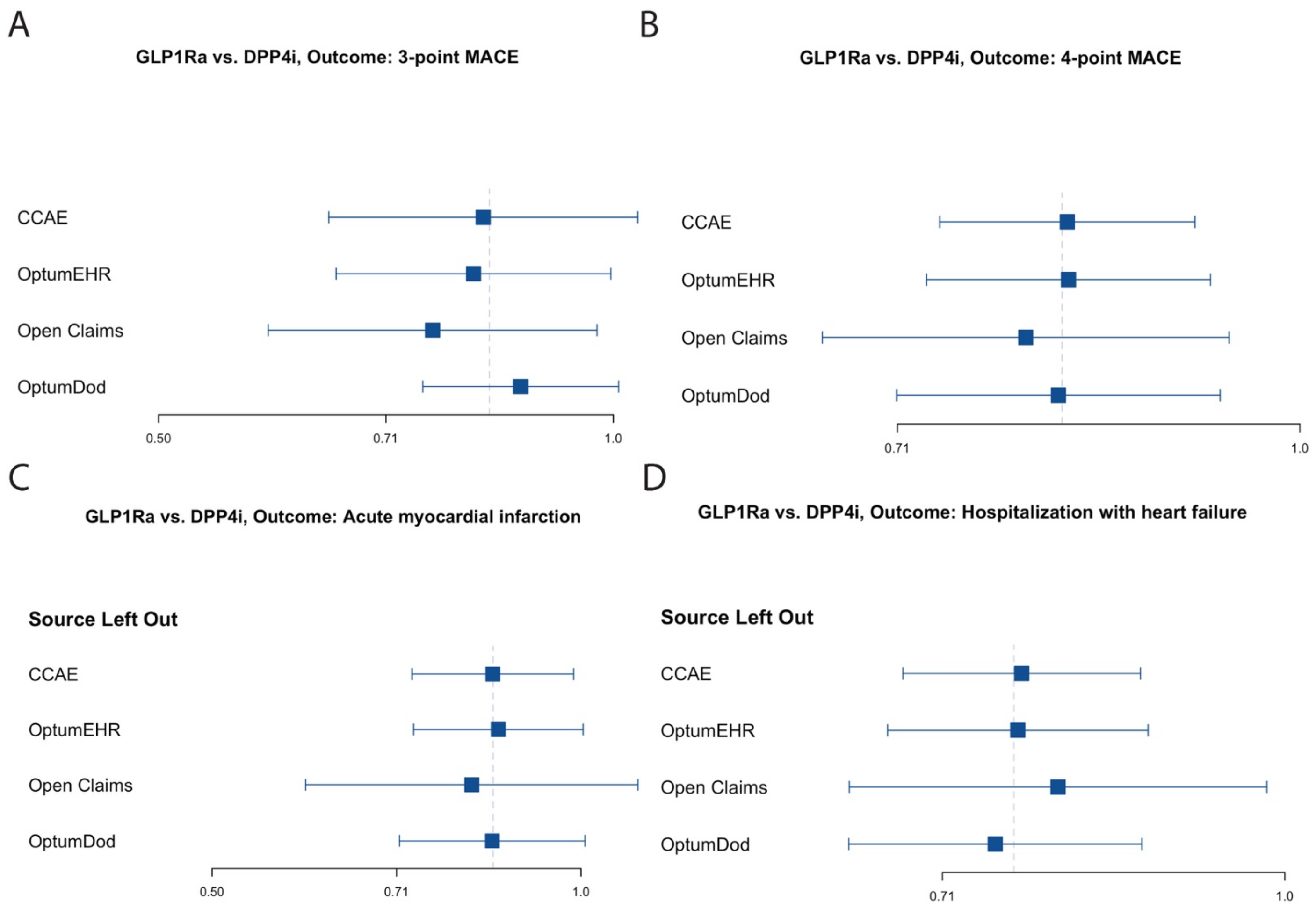


**Supplementary Figure 54: Leave-one-out meta-analysis of calibrated hazard ratio estimates of cardiovascular outcomes for GLP1-RA vs DPP4i comparison across databases.**

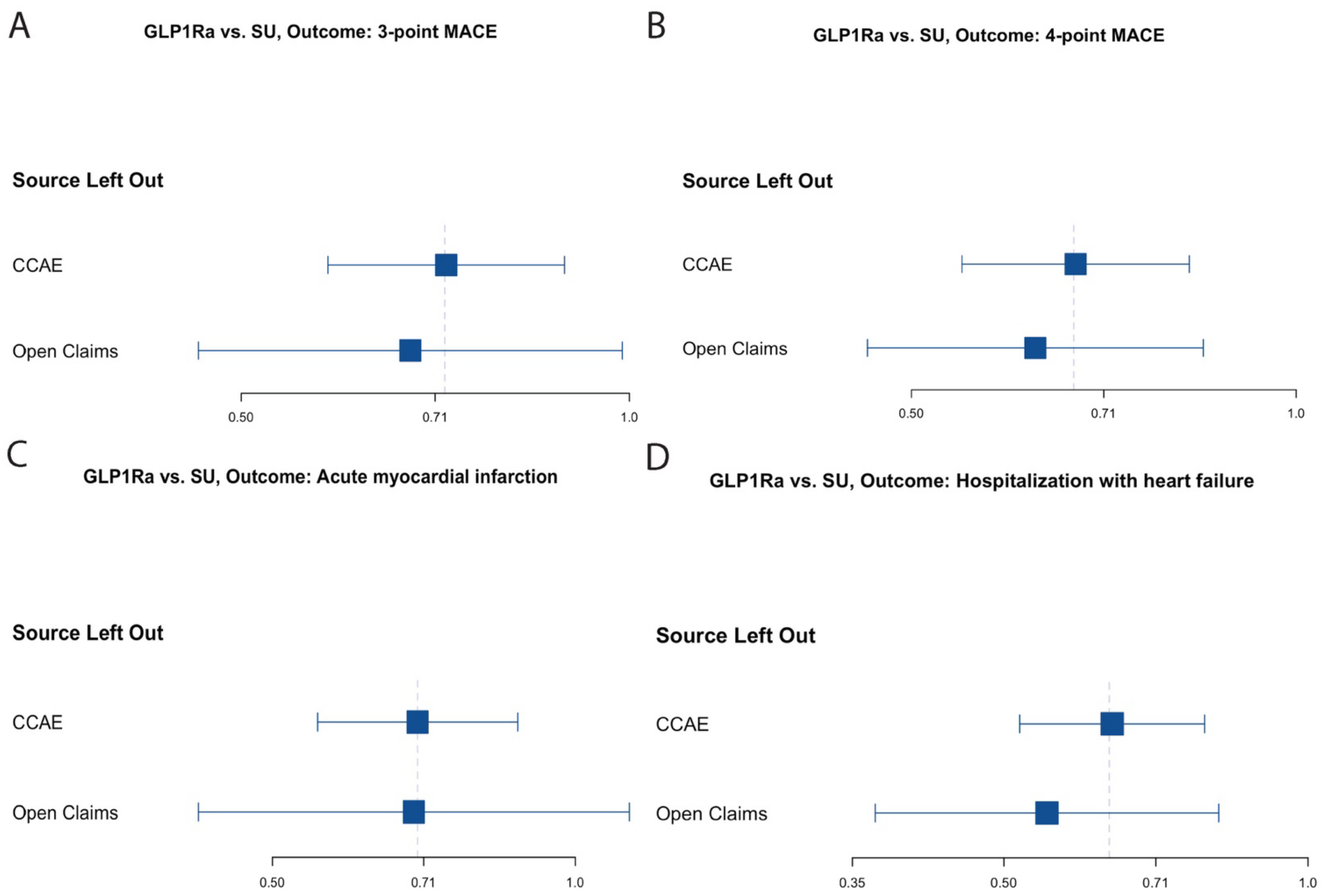


**Supplementary Figure 55: Leave-one-out meta-analysis of calibrated hazard ratio estimates of cardiovascular outcomes for GLP1-RA vs SU comparison across databases.**

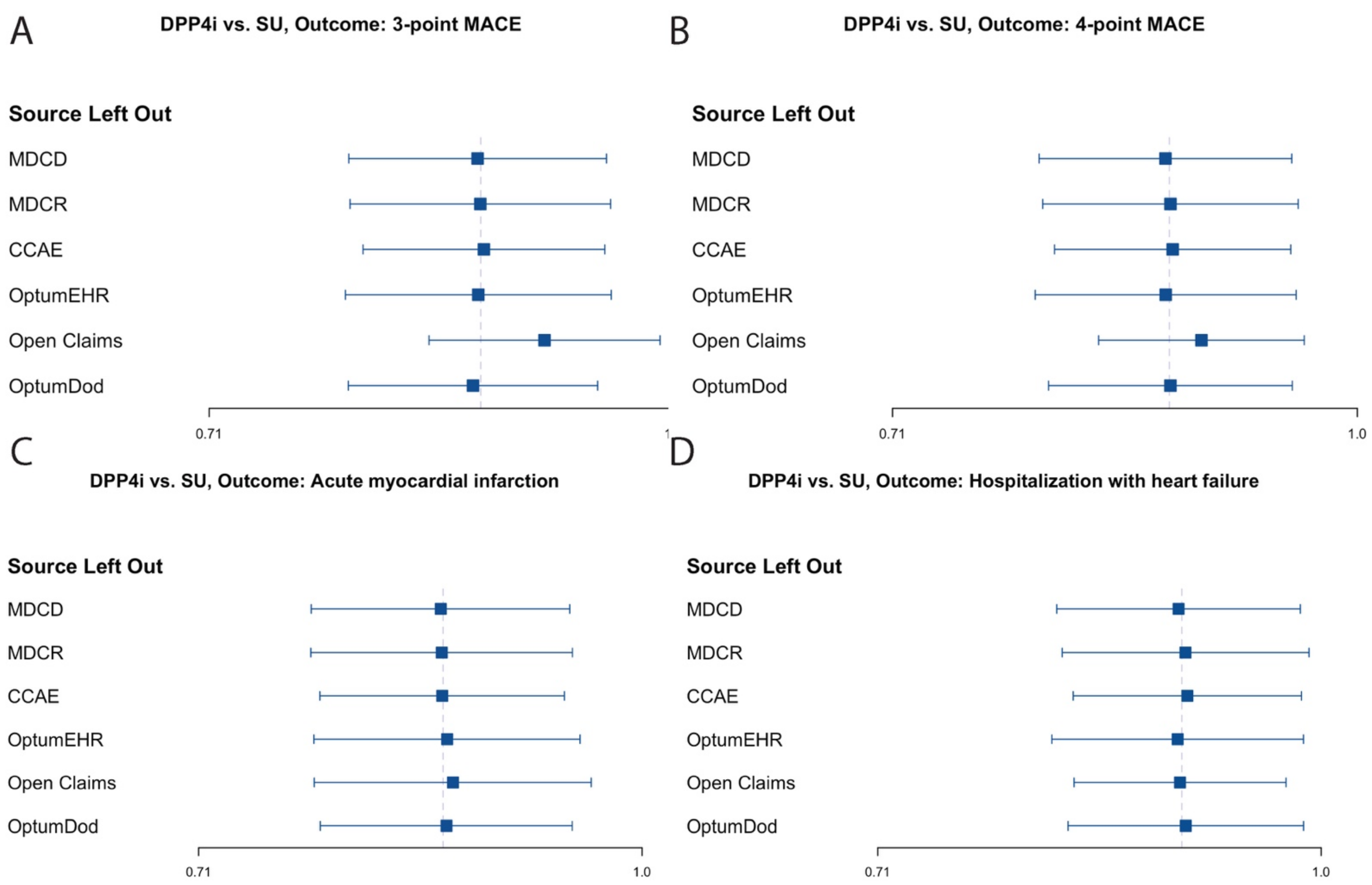


**Supplementary Figure 56: Leave-one-out meta-analysis of calibrated hazard ratio estimates of cardiovascular outcomes for DPP4i vs SU comparison across databases.**
